# Prevalence of long-term effects in individuals diagnosed with COVID-19: an updated living systematic review

**DOI:** 10.1101/2021.06.03.21258317

**Authors:** Francesca Reyes Domingo, Lisa A Waddell, Angela M. Cheung, Curtis L. Cooper, Veronica J. Belcourt, Alexandra M. E. Zuckermann, Tricia Corrin, Rukshanda Ahmad, Laura Boland, Claudie Laprise, Leanne Idzerda, Anam Khan, Kate Morissette, Alejandra Jaramillo Garcia

**Author notes:** Corresponding author: Francesca Reyes Domingo. **Funding Statement:** Funding for the conduct of this review is provided by the Public Health Agency of Canada. **Competing interests:** none.

## Abstract

**Objective:** Post COVID-19 condition refers to persisting or recurring symptoms weeks after acute COVID-19 illness which can significantly impact quality of life and health systems. It is important to understand the manifestation and magnitude of this condition. The objective of this living systematic review is to summarize the prevalence of symptoms and sequelae reported by people ≥4 weeks after COVID-19 diagnosis.

**Design:** Systematic review, meta-analysis and narrative synthesis.

**Data sources:** Embase, Medline, PsychInfo, Cochrane Central and select grey literature up to April 14, 2021.

**Methods:** We adapted a previous search strategy used by the U.K. National Institute for Health and Care Excellence and updated it to search for new literature. Two reviewers screened references independently; one extracted data and assessed risk of bias and certainty of the evidence while another verified them. Prevalence data from laboratory-confirmed populations were meta-analyzed using a random effects model and synthesized separately in the short-term (4-12 weeks) and long-term (>12 weeks) periods after diagnosis. Data from clinically-diagnosed populations were synthesized narratively.

**Results:** Of the 4444 unique citations, 84 observational studies met our inclusion criteria. Over 100 post COVID-19 symptoms and sequelae were reported. Sixty-one percent (95% CI: 44-76%, *low certainty*) and 53% (95% CI: 41-65%, *low certainty*) of laboratory-confirmed individuals reported persistence or presence of one or more symptoms in the short- and long-term periods, respectively. The most prevalent symptoms in both periods included: fatigue, general pain or discomfort, shortness of breath, cognitive impairment and mental health symptoms.

**Conclusions:** A substantial proportion of individuals reported a variety of symptoms ≥4 weeks after COVID-19 diagnosis. Due to gaps in the research base, and the low certainty of the evidence currently available, further research is needed to determine the true burden of post COVID-19 condition in the general population and in specific subgroups.

**PROSPERO registration number:** CRD42021231476.

## Introduction

Severe acute respiratory syndrome-coronavirus-2 (SARS-CoV-2) has resulted in over 230 million cases of COVID-19 and over 4 million deaths worldwide as of September 2021 (1). The typical duration of acute illness is two to six weeks, however, some patients have described debilitating symptoms persisting or recurring for weeks or months after acute illness(2). Affected patients are commonly referred to as post COVID or COVID long-haulers (3–5).

In this review, we used the term “post COVID-19 condition” to describe persistent or recurring symptoms. Other terms used in the literature have included long COVID, post-COVID conditions, chronic COVID syndrome and post-acute sequelae of SARS-CoV-2 infection (PASC) (2,6–9). Definitions of ‘long-term’ have varied from ≥4 to ≥12 weeks after COVID-19 diagnosis (10–13). On October 6, 2021, the World Health Organization published a clinical case definition for post COVID-19 condition, which was developed using a Delphi consensus process, noting that the definition may change as new information on the condition emerges(14). Post COVID-19 condition was defined as symptoms that usually appear three months from onset of COVID-19 symptoms and last for at least two months in individuals with a history of probable or confirmed SARS-CoV-2 infection.

Due, in part, to the lack of a standard case definition for post COVID-19 condition prior to October 2021(15), prevalence estimates have varied from 3% to as high as 80%(16–18). Given the millions of individuals who have been infected with COVID-19, serious ramifications on quality of life, health care utilization, and workforce productivity are anticipated even if the lowest prevalence estimates were found accurate. Improved understanding of the prevalence of post COVID-19 condition, the symptoms and sequelae observed, its effects on COVID-19 survivors, and its resolution over time is important to address this issue. Other reviews have also looked at estimating the prevalence of various symptoms related to post COVID-19 condition (13,19–26); however, none have assessed for certainty in the evidence body of evidence. The objective of this systematic review is to identify and summarize studies reporting the frequency of symptoms, sequelae, and difficulties in conducting usual activities experienced by individuals living with post COVID-19 condition at four weeks or more after initial COVID-19 diagnosis and provide certainty in the findings for select outcomes.

## Methods

We conducted a systematic review that adhered to Cochrane methodology and the Preferred Reporting Items for Systematic Reviews and Meta-Analyses (PRISMA) guidelines (27, 28). Our multidisciplinary team included methodologists and subject matter experts currently treating patients with post COVID-19 condition. The review question and methodology were determined *a priori* (protocol registered in PROSPERO: CRD42021231476) (29). This living review will be periodically updated on a quarterly basis or as resources permit. Detailed methods are provided in Supplementary File 1 – Detailed methodology.

### Information Sources and Search strategy

Our formative search identified a systematic review conducted by the National Institute for Health and Care Excellence (NICE) that examined post COVID-19 condition. We adapted the NICE review search strategy (NICE search dates were January 1^st^ to October 21^st^, 2020) in consultation with a health librarian (12) (see Supplementary File 1 for the detailed search strategy). All studies included in the NICE review and any French articles they excluded were eligible for inclusion. We then updated the search (October 22, 2020 to January 15, 2021 and again from January 16, 2021 to April 14, 2021) using the following databases: Embase, Medline, PsycINFO, and Cochrane Central. We conducted a complementary search for grey literature in January and again in April 2021 and included any relevant literature.

### Eligibility Criteria

We included primary studies in English or French that included 50 or more participants of any age with laboratory-confirmed SARS-CoV-2 infection (“laboratory-confirmed”) or with COVID-19 clinically-diagnosed by a health professional (“clinically-diagnosed”), and reported the prevalence of symptoms or sequelae four or more weeks after COVID-19 diagnosis. For participants who self-reported their laboratory diagnosis for COVID-19, and where proof of laboratory findings was not required for inclusion in the study, then we considered these participants as clinically-diagnosed. We excluded pre-prints, and non-peer reviewed articles and primary studies that recruited participants specifically because they reported experiencing such long-term effects.

“Time since COVID-19 diagnosis” was used synonymously with time since symptom onset, positive laboratory result, or diagnosis by a health professional. We defined short- and long-term outcomes as those measured between four and 12 weeks and those measured more than 12 weeks after COVID-19 diagnosis, respectively.

### Study selection and data collection process

*A priori*, we developed multi-stage (title/abstract and full text) screening questions and data extraction forms that were piloted by all reviewers (Supplementary File 1). Two reviewers screened citations and full texts independently. At the title and abstract screening stage, citations passed to full text screening if they were included by at least one reviewer, but both reviewers had to agree on exclusions. At the full text stage, consensus was required for both inclusion and exclusion. For data extraction, one reviewer extracted data from included studies, which were verified by a second reviewer. At each stage, reviewers resolved conflicts through consensus or consultation with a third reviewer.

### Outcomes

The main outcomes of interest were any symptom, sequelae or outcomes pertaining to difficulties conducting usual activities (i.e. functional outcomes) reported by individuals four or more weeks after a COVID-19 diagnosis. We identified the following key symptoms or sequelae: fatigue, shortness of breath, neurocognitive impairment, pain (in the joints, chest, or muscles), organ damage, dizziness, tachycardia, chest tightness or heaviness, mental health, olfactory and gustatory impairments, and sleeping disturbances. Additional outcomes, such as those from diagnostic imaging or pulmonary function tests, which may often provide abnormal results despite resolution of patient symptoms, were considered as outcomes of interest in the long-term period only.

### Evidence synthesis

Our primary synthesis focused on outcomes in individuals who had laboratory-confirmed COVID-19 to reduce likelihood of including results due to unrelated conditions. We synthesized short- and long-term outcomes separately. When outcomes were reported at multiple time points within a study, we used results from the longest follow-up time point.

Where appropriate, we conducted meta-analyses for outcomes with two or more studies using a random effects model. To explore reasons for high heterogeneity across studies, we determined sub-group analyses *a priori* and considered these for key outcomes. We stratified results by level of care received during the acute stage of COVID-19 infection (i.e., admitted to ICU, hospitalized, non-hospitalized), which was used as a proxy for severity of COVID-19 (i.e., patients with more severe COVID-19 were more likely to require hospitalized care). We performed the analyses using R statistical software version 4.0.4 (30), with package metafor version 2.4-0 (31) and package meta version 4.18-0 (32). The pooled results with 95% confidence intervals are presented in forest plots. We conducted narrative syntheses for the outcomes in the clinically-diagnosed population and did not explore heterogeneity across these studies.

### Assessing risk of bias and certainty in the evidence

We used a modified version of the Joanna Briggs Institute critical appraisal tool for prevalence studies (33) to assess risk of bias. After consulting with the authors of this tool, we omitted questions 3-5 to avoid duplication with the criteria on imprecision and indirectness used to assess the certainty of the evidence. We rated the risk of bias for each outcome separately for questions 6-8 and then categorized questions into three domains (participants [questions 1, 2 and 9], outcome measures [questions 6-7], and statistics [question 8]). Studies that met the criteria in these domains were rated as low risk of bias, those partially met were rated moderate, and those that did not meet the criteria were rated high risk of bias. The criteria and schema used to assess risk of bias are provided in Appendix 4 of Supplementary File 1. One reviewer assessed risk of bias, which was verified by a second reviewer. Reviewers resolved conflicts through consensus or consultation with a third reviewer.

We assessed certainty in the body of evidence for key symptoms or sequelae and the most prevalent outcomes using the Grading of Recommendations Assessment, Development and Evaluation (GRADE) approach (34). In the absence of a formal framework for prevalence and GRADE, and in consultation with GRADE experts, we adapted the GRADE framework for assessment of incidence estimates in the context of prognostic studies(35) as a basis to assess prevalence estimates similar to those used previously by Righy et al.(36)

In the context of prevalence, observational studies may provide robust estimates due to broad eligibility criteria and enrollment of representative populations (35). Thus, the quality of the evidence from observational studies were initially assigned as “high” for all outcomes. The quality of the evidence was then downgraded (to “moderate”, “low”, or “very low”) if there were serious or very serious concerns over any of the following five domains that reduce certainty in the prevalence estimates: risk of bias, inconsistency, indirectness, imprecision, or publication bias. In instances where there were minor concerns with one of these domains, a decision was made to downgrade by half a point instead of a full point. Half-points were then combined across domains to yield a total score (e.g., downgrading by 1.5 points for risk of bias and 0.5 points for indirectness for a total of 2 points from “high” to “low”). In the event that the final rating included a half point (e.g., 1.5 points), we conservatively rounded up (i.e., in this case to downgrade by 2). Due to the nature of prevalence outcomes and design of the application of GRADE criteria to these outcomes, no criteria for upgrading were considered applicable. The specific decision rules that we applied for judgments in relation to the five GRADE domains are described in detail in Appendix 5 of Supplementary File 1.

After piloting the tool, one reviewer assessed risk of bias and graded the evidence while a second verified the assessments. Reviewers resolved conflicts through consensus or consultation with a third reviewer.

### Patient and Public Involvement

We did not involve patients in the conduct of this review.

## Results

### Study Selection

Of the 4444 unique citations yielded in the search, 779 met criteria for full-text screening (Figure 1). The list of studies excluded in full-text screening by reason of exclusion are provided in Supplementary File 2. Eighty-four studies met our inclusion criteria, of which 62 included prevalence data for individuals with laboratory-confirmed COVID-19, 21 included prevalence data for individuals who were clinically-diagnosed with COVID-19, and one included prevalence data on both. (Table 1, Supplementary Table 1)

**Figure 1:**
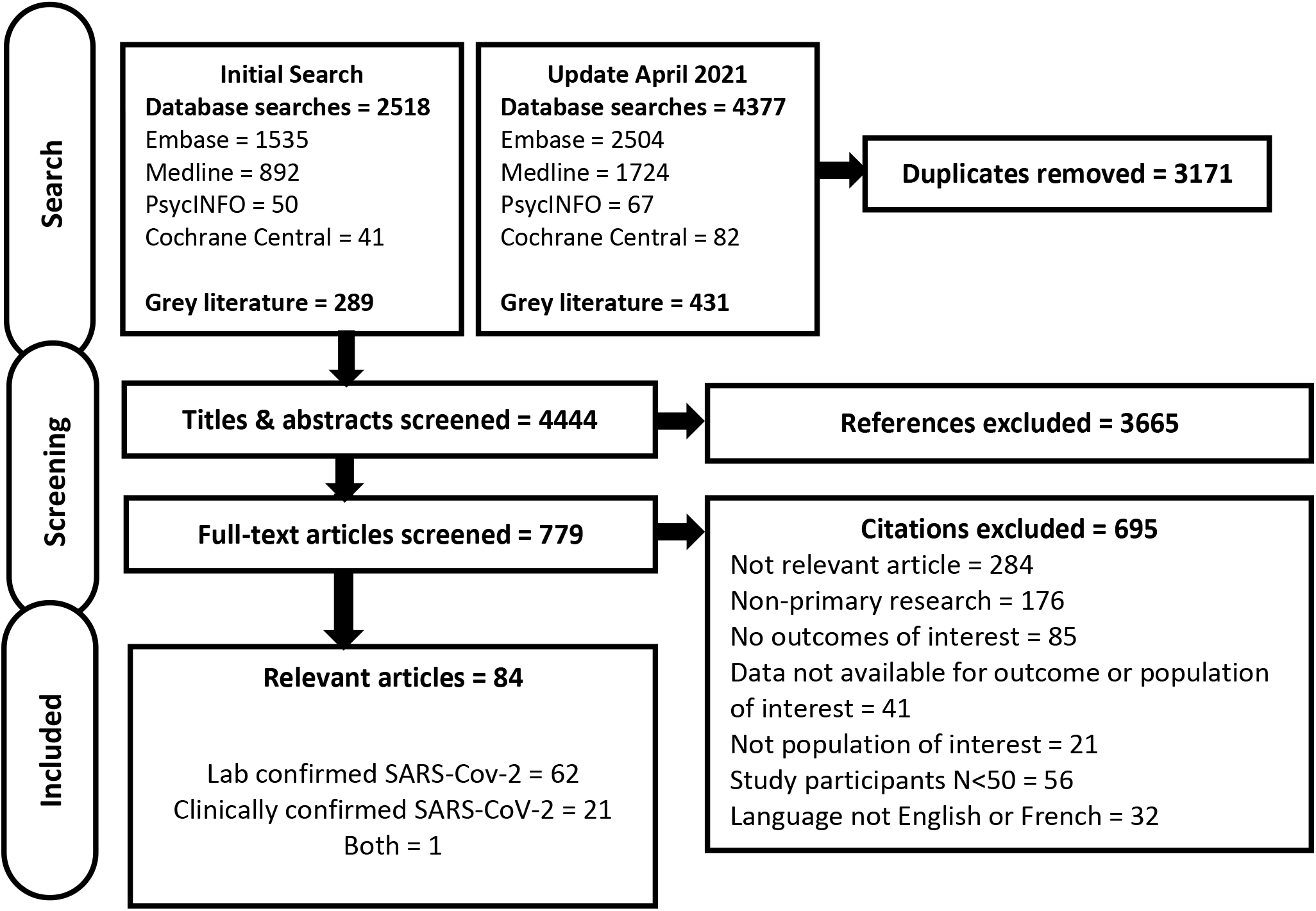
PRISMA flow diagram of articles through the systematic review process

**Table 1:**
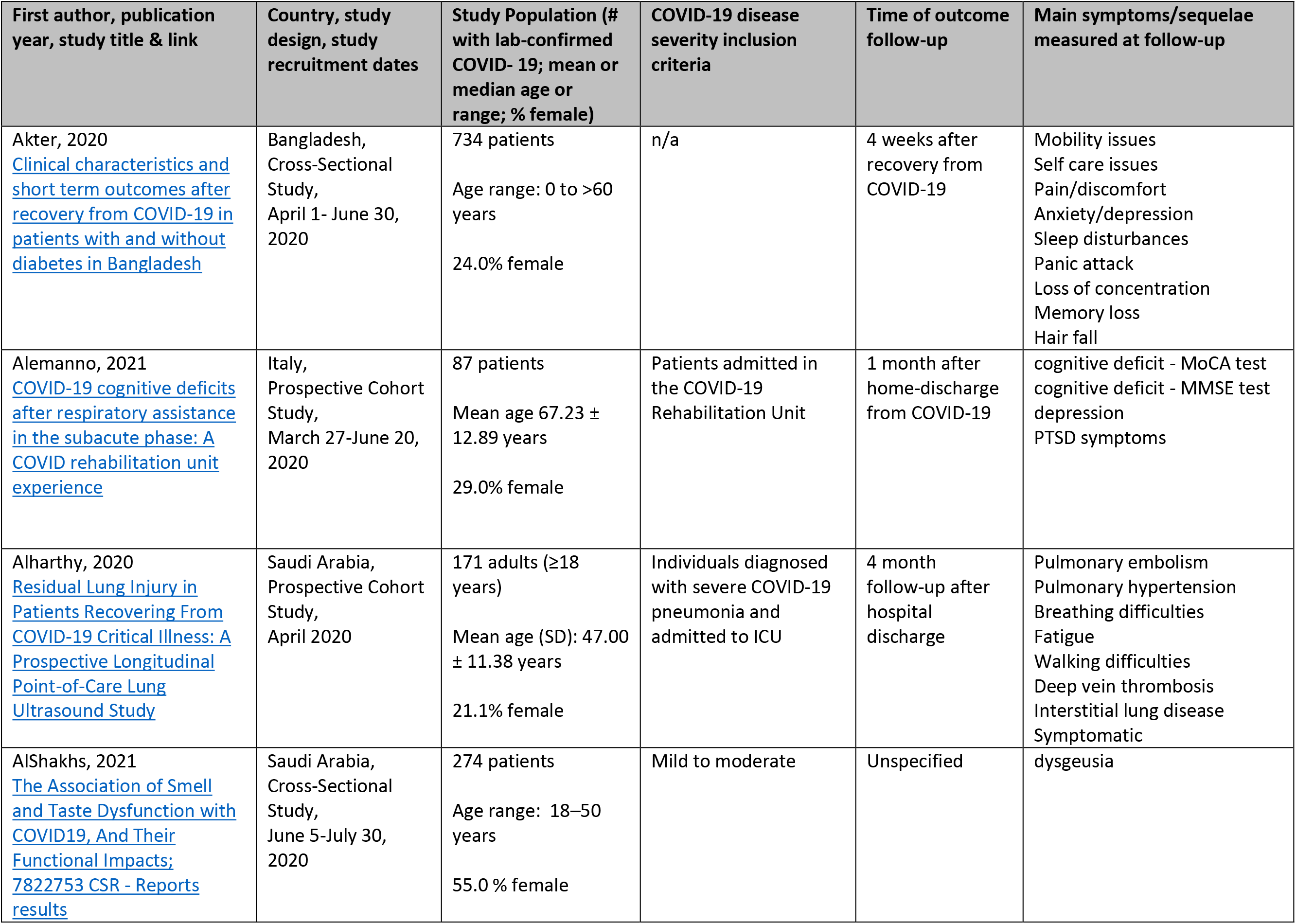
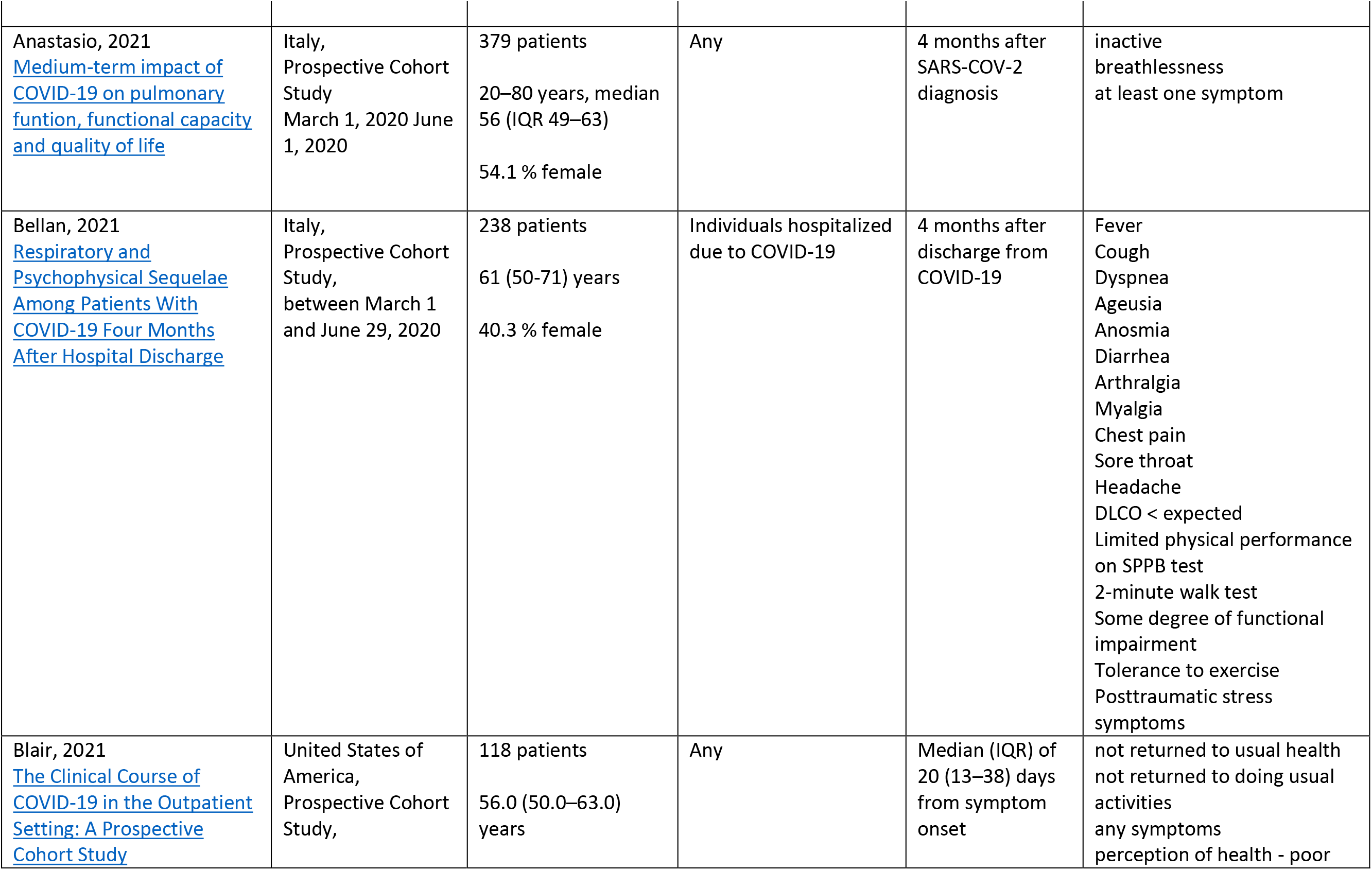
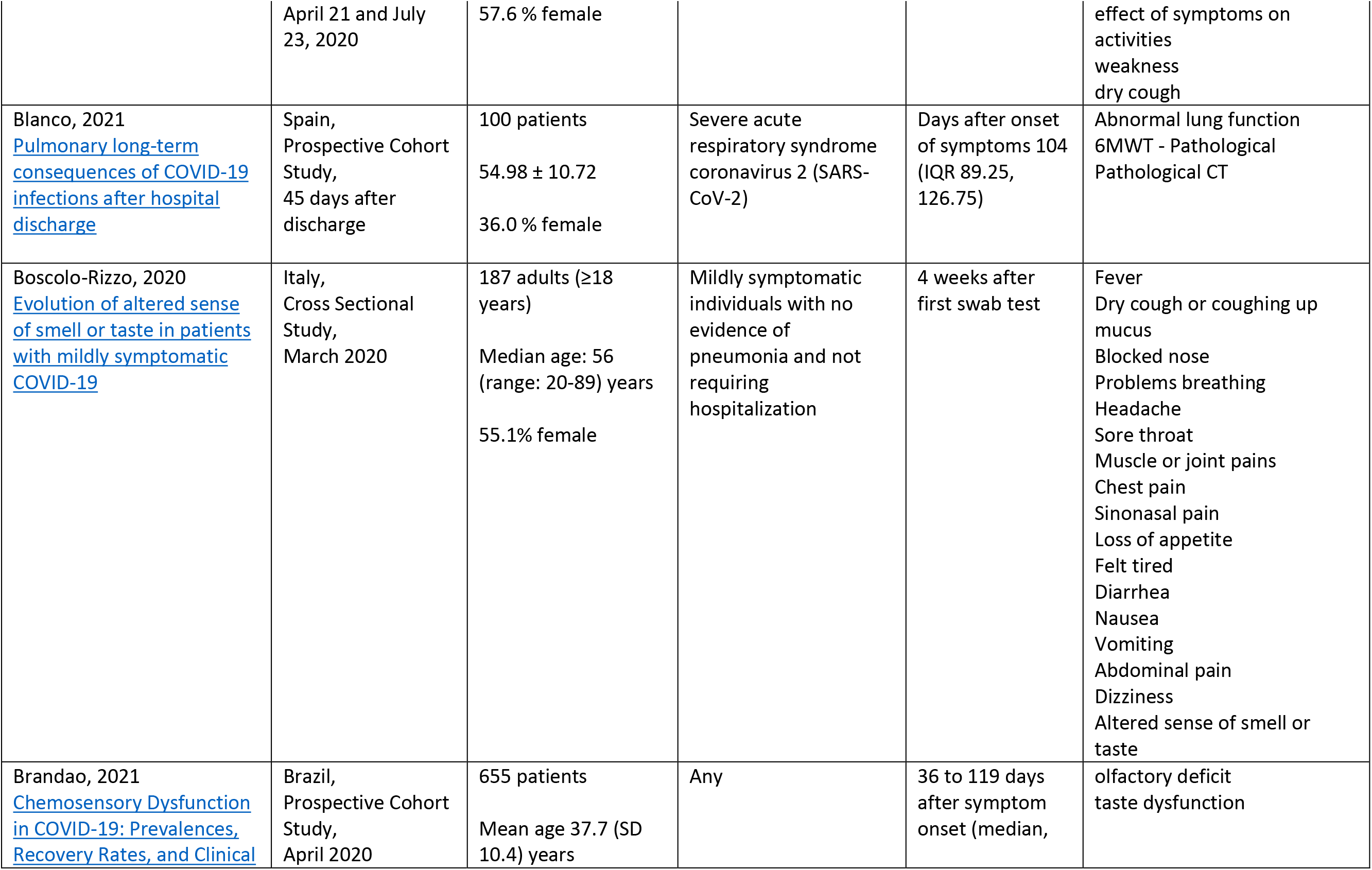
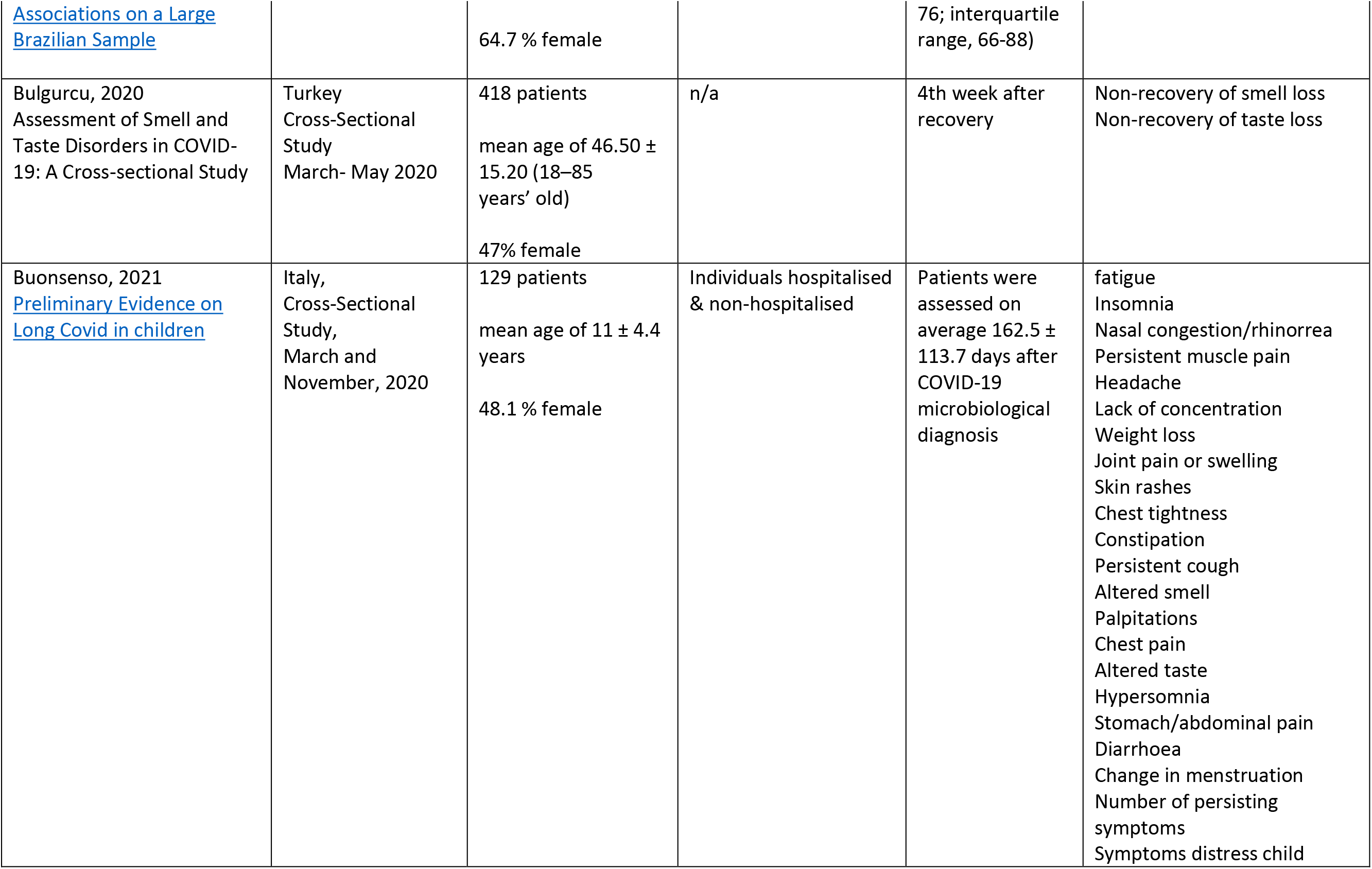
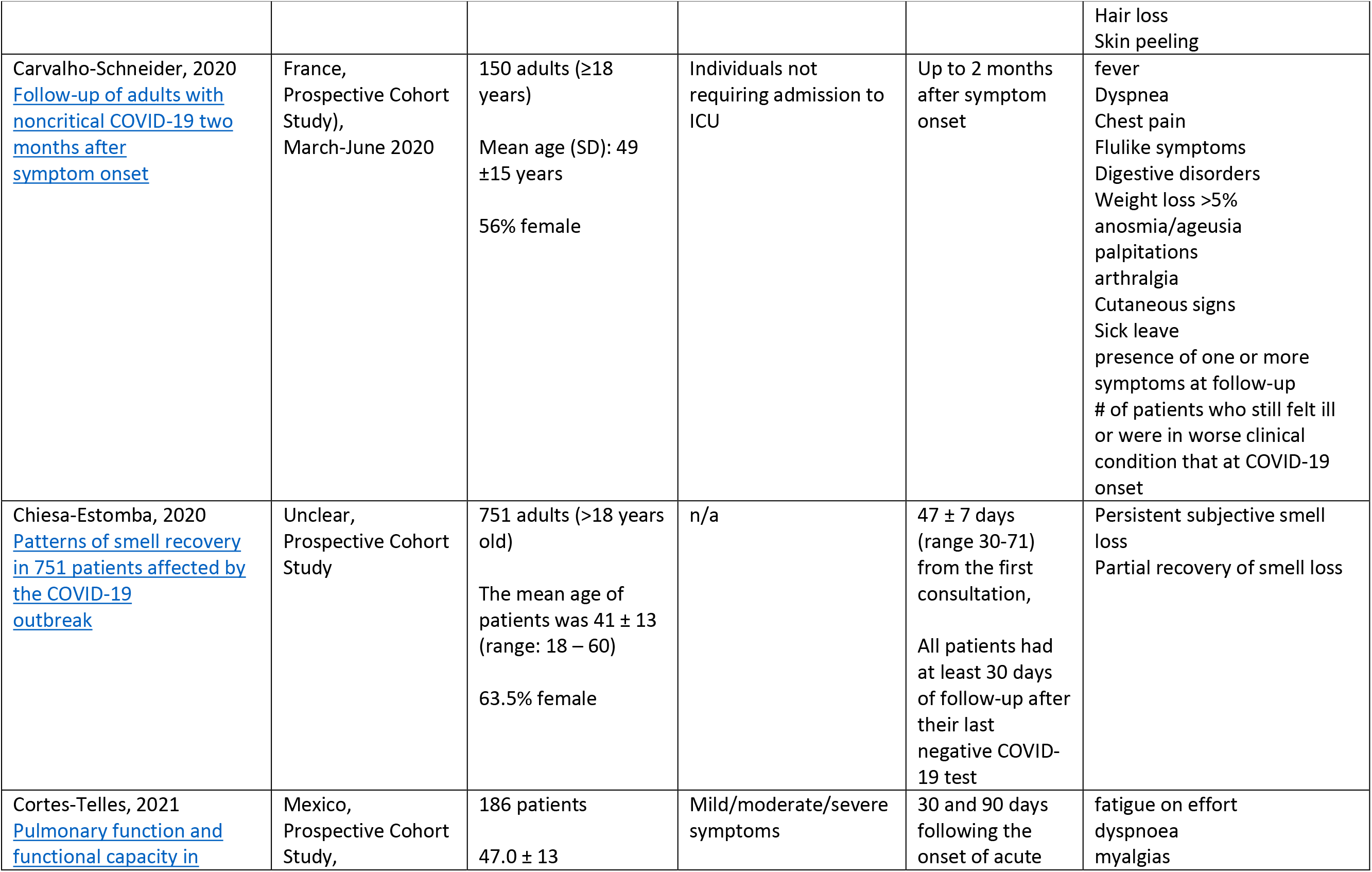
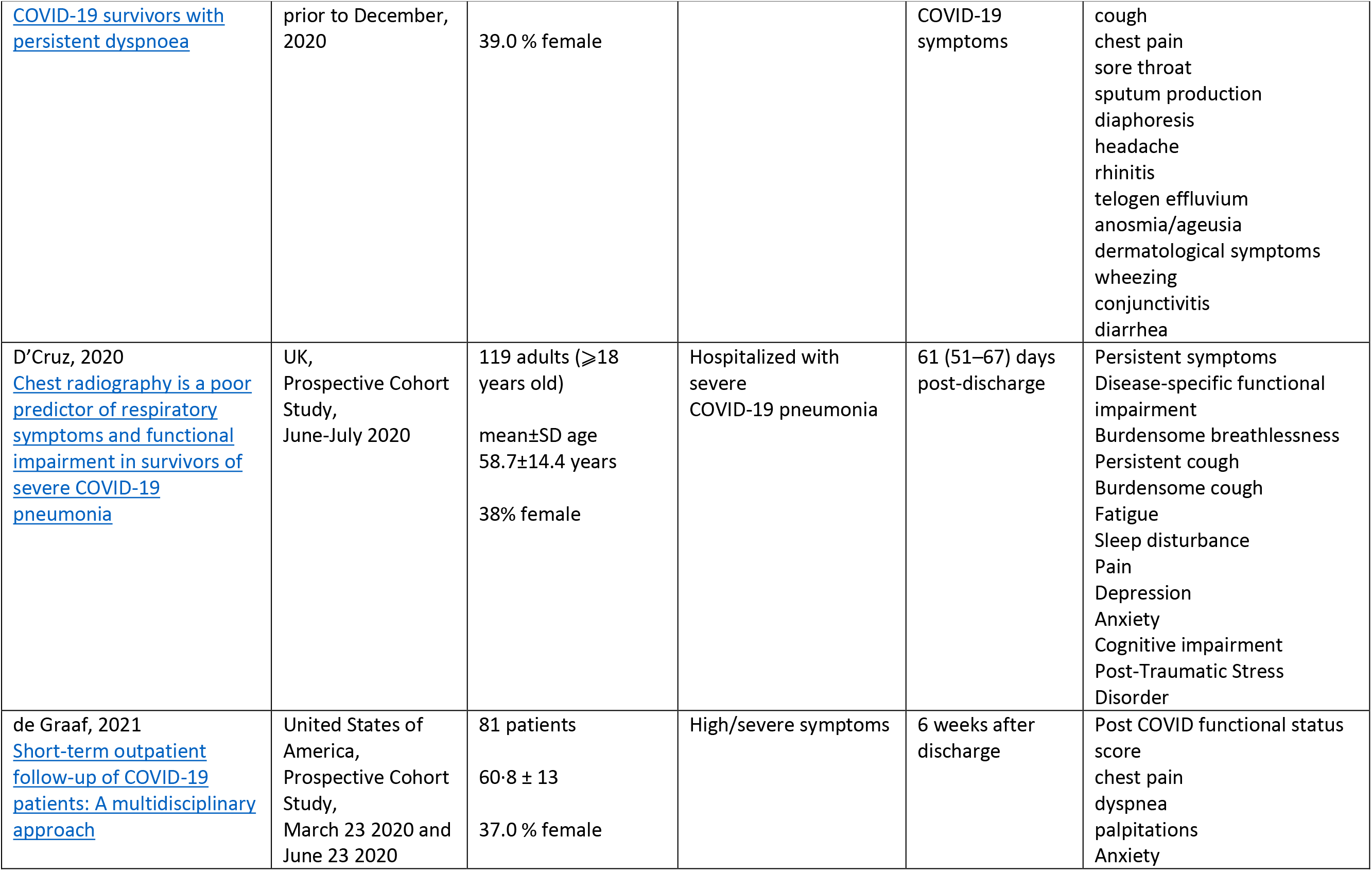
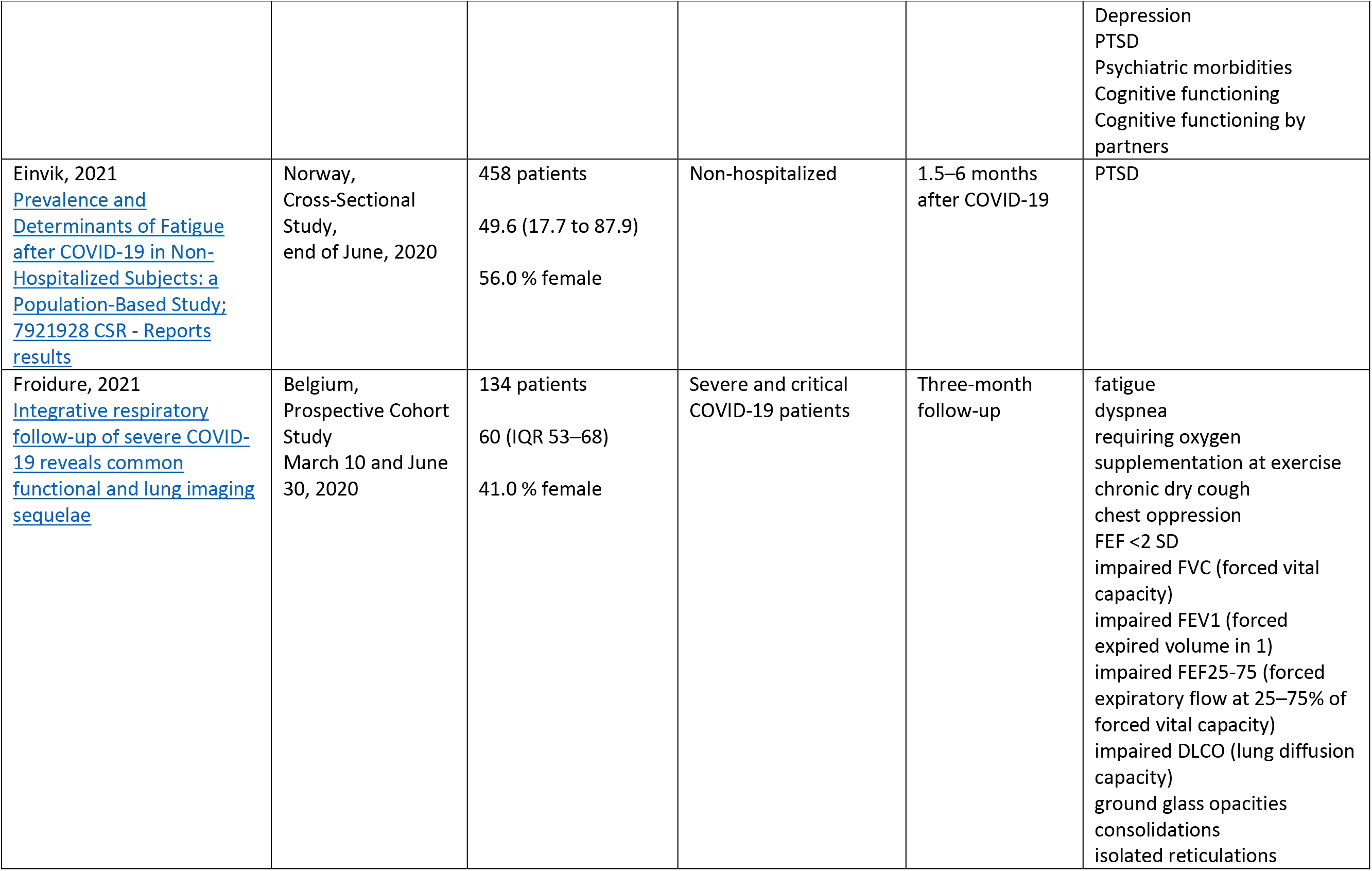
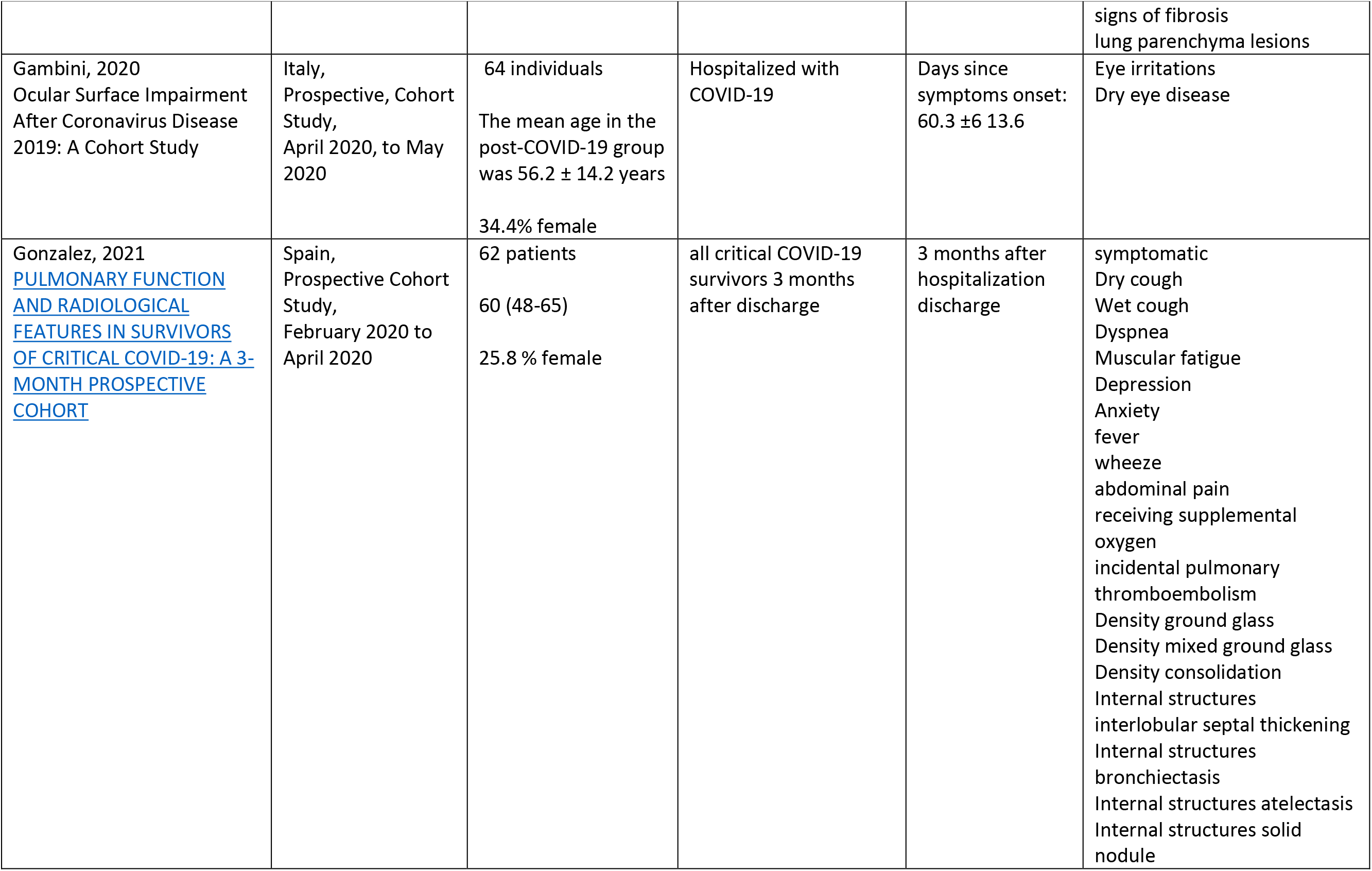
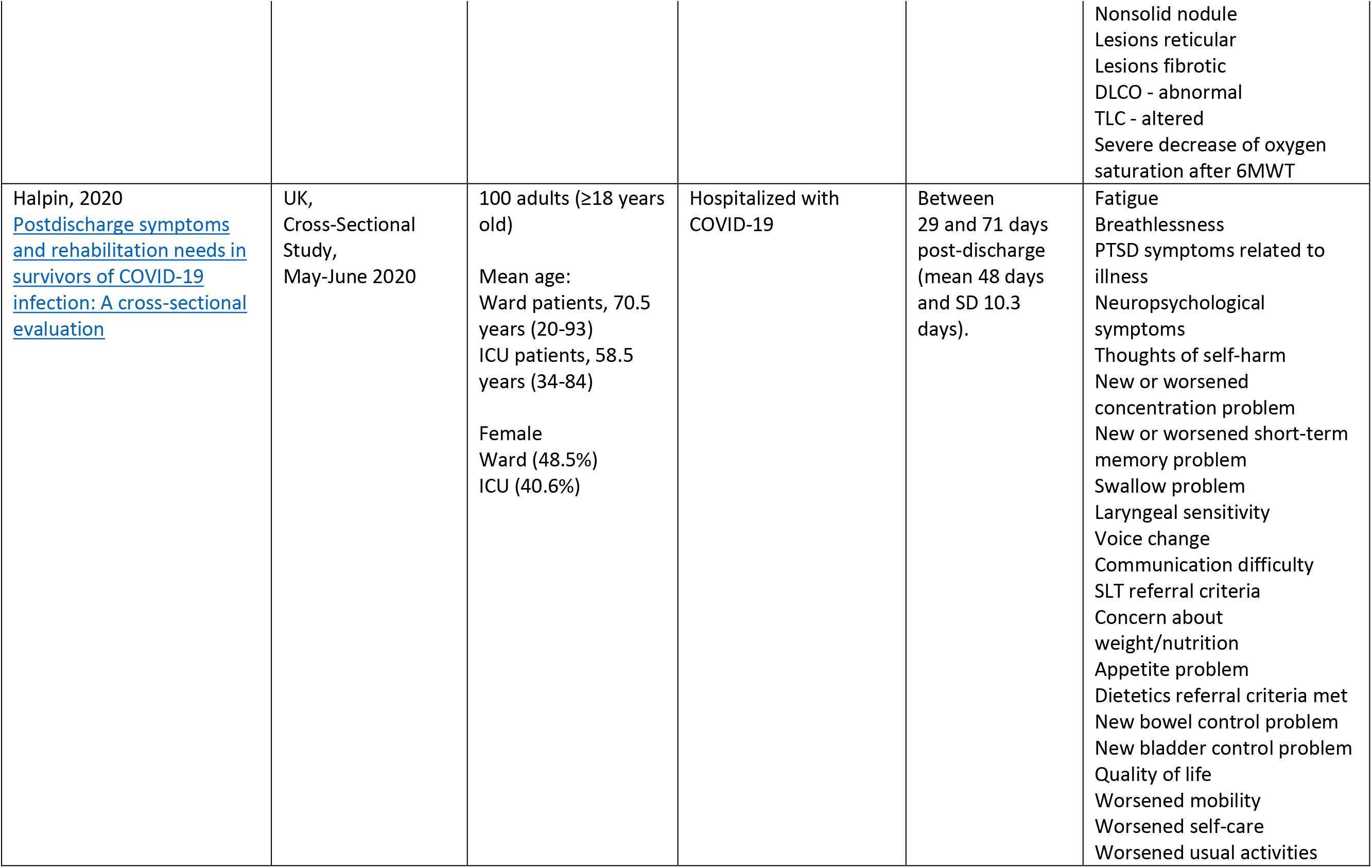
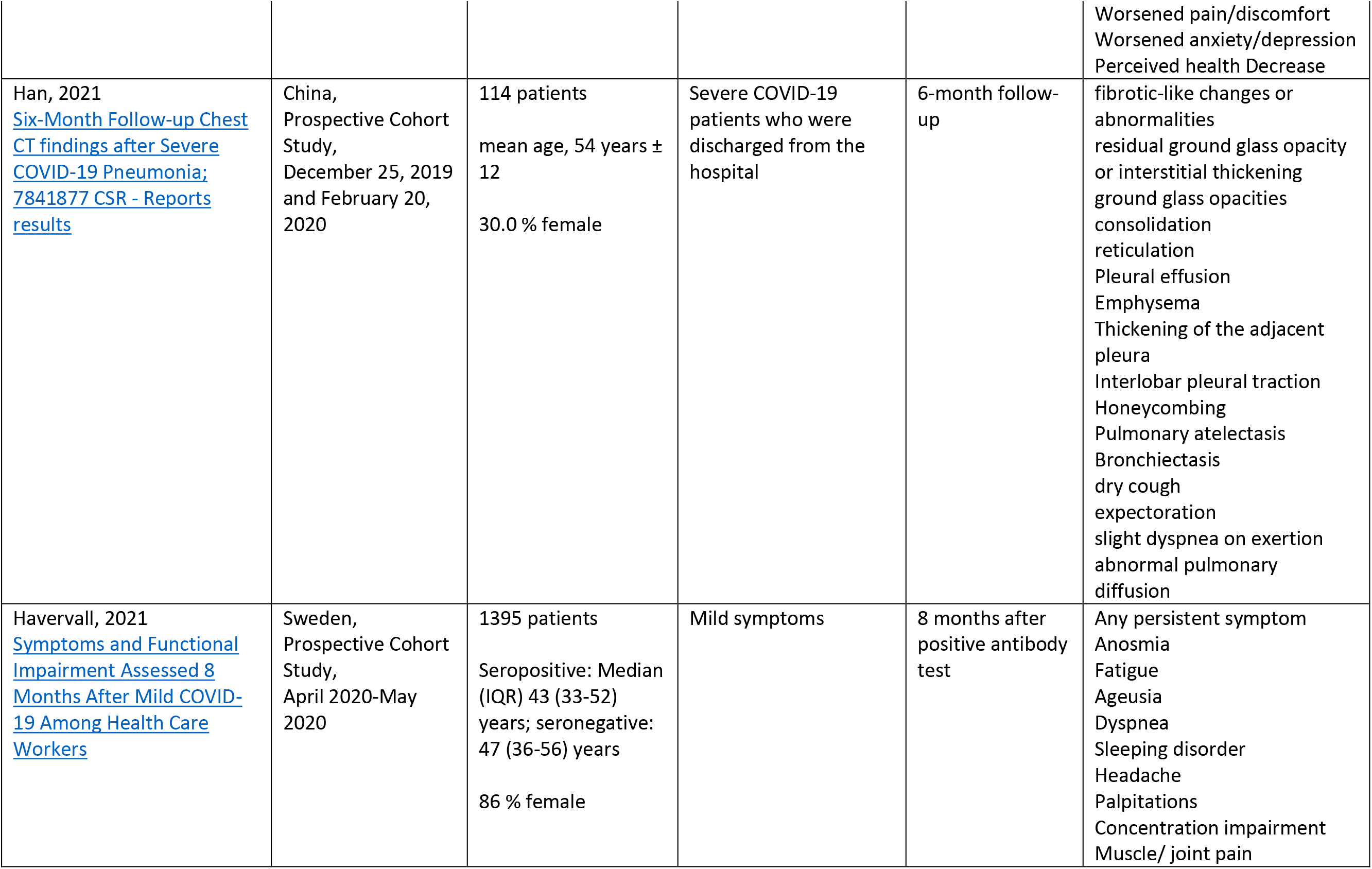
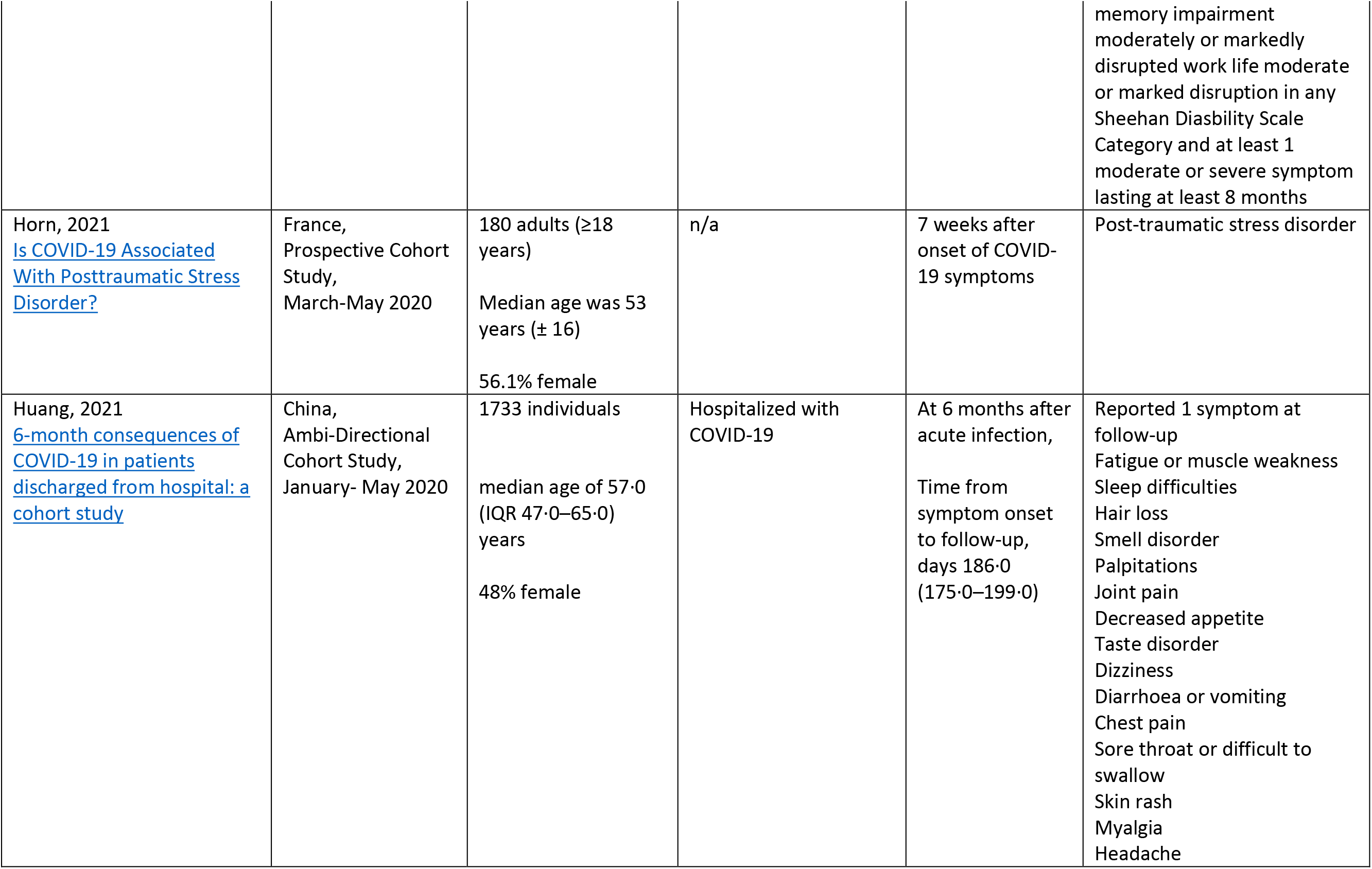
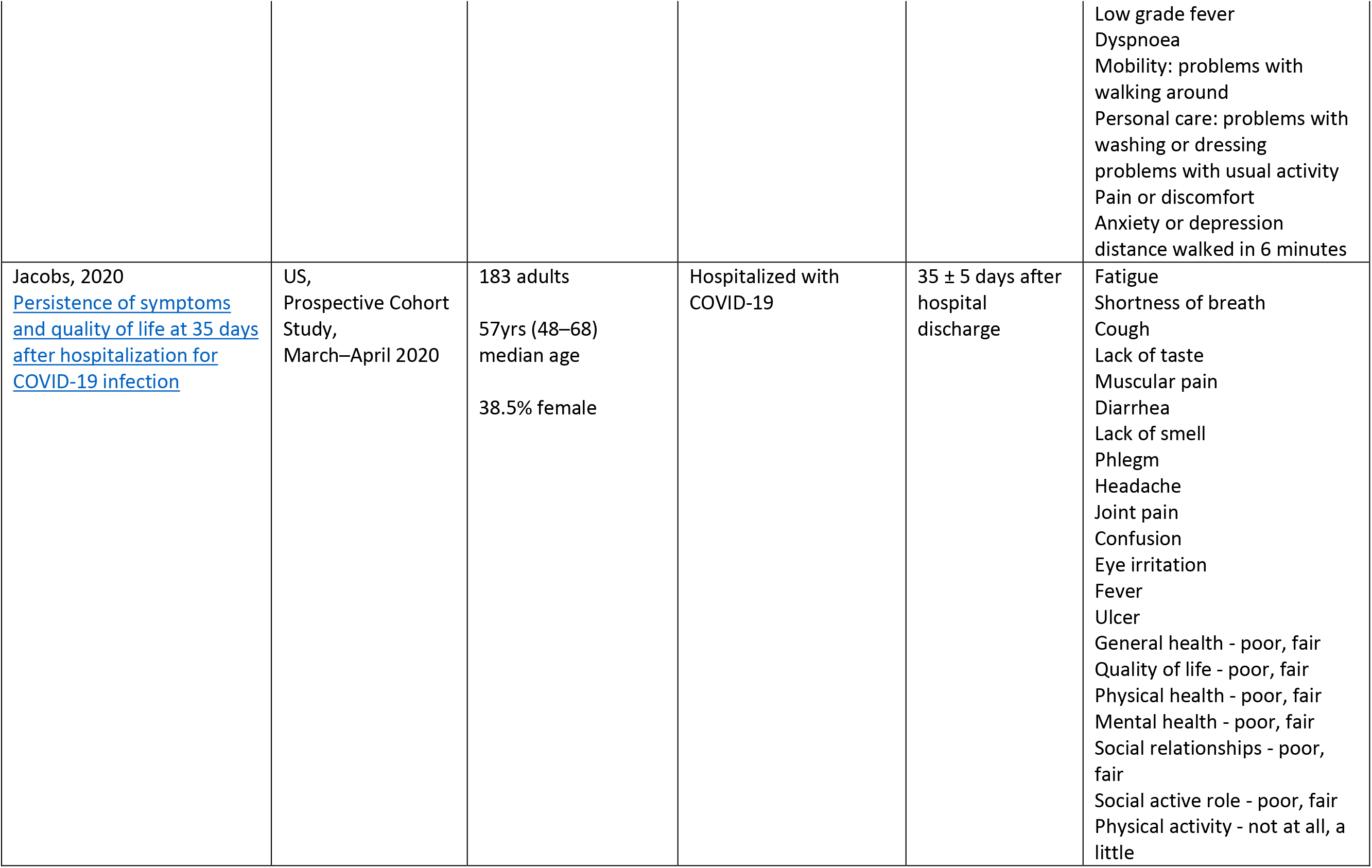
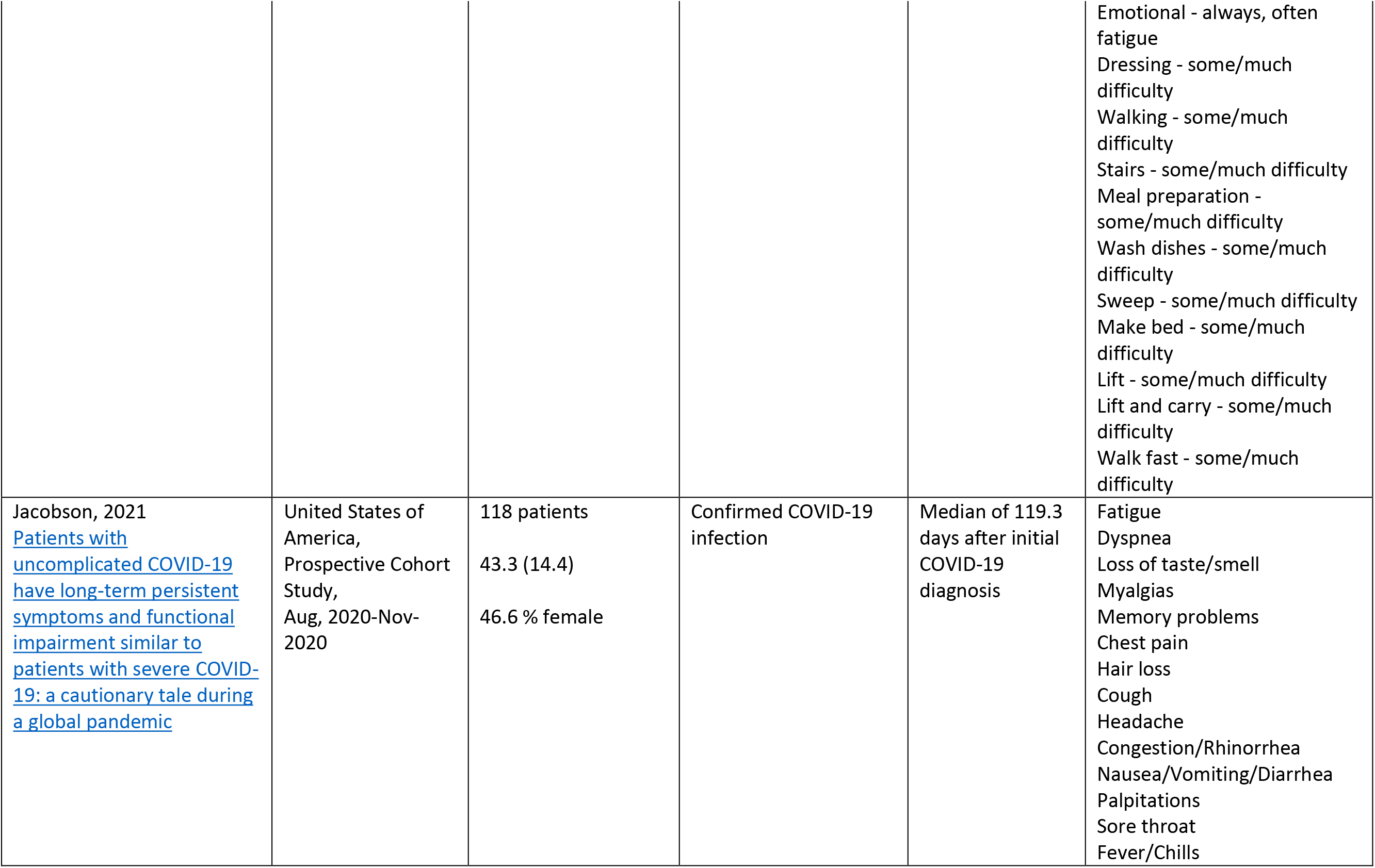
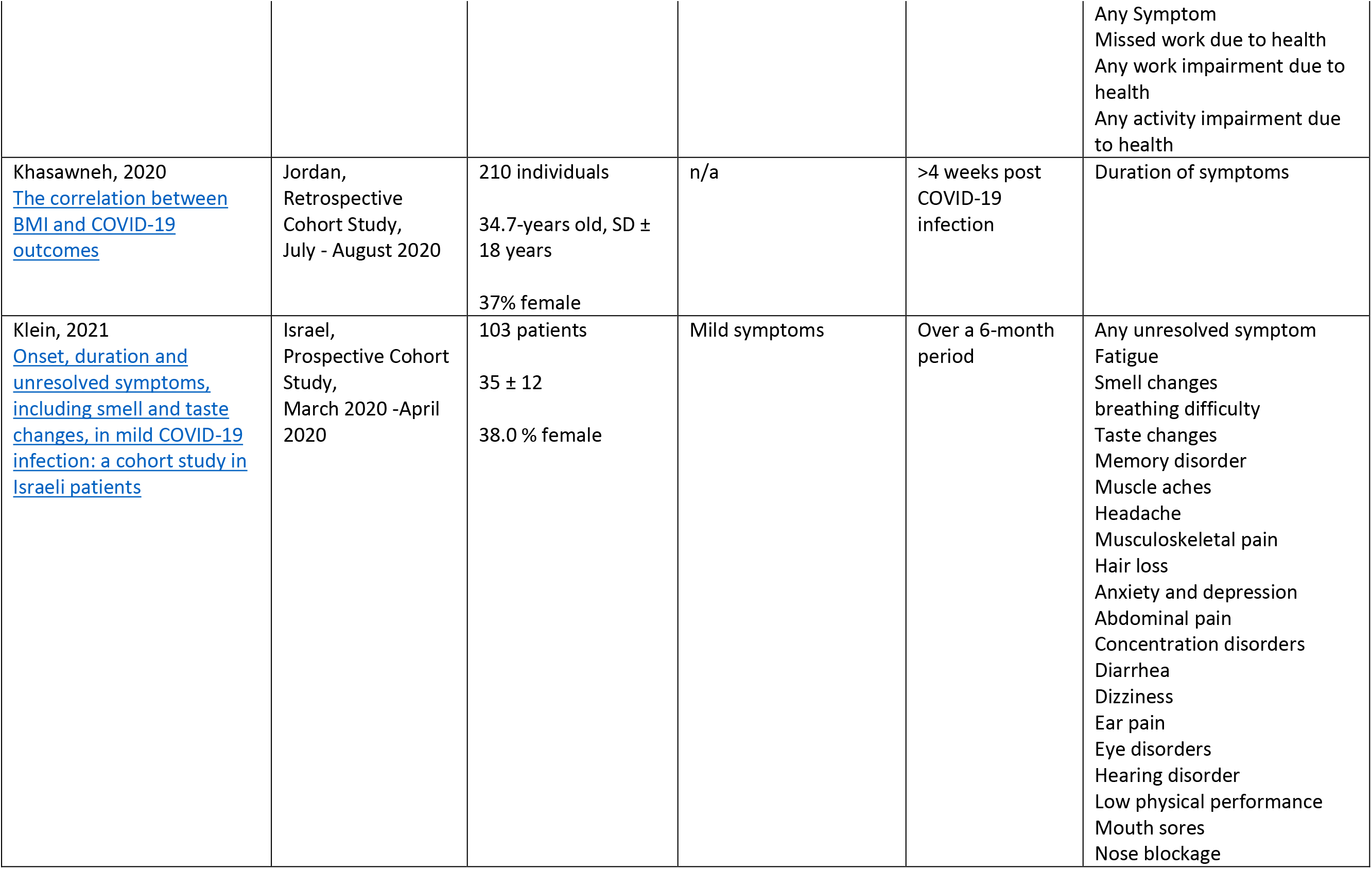
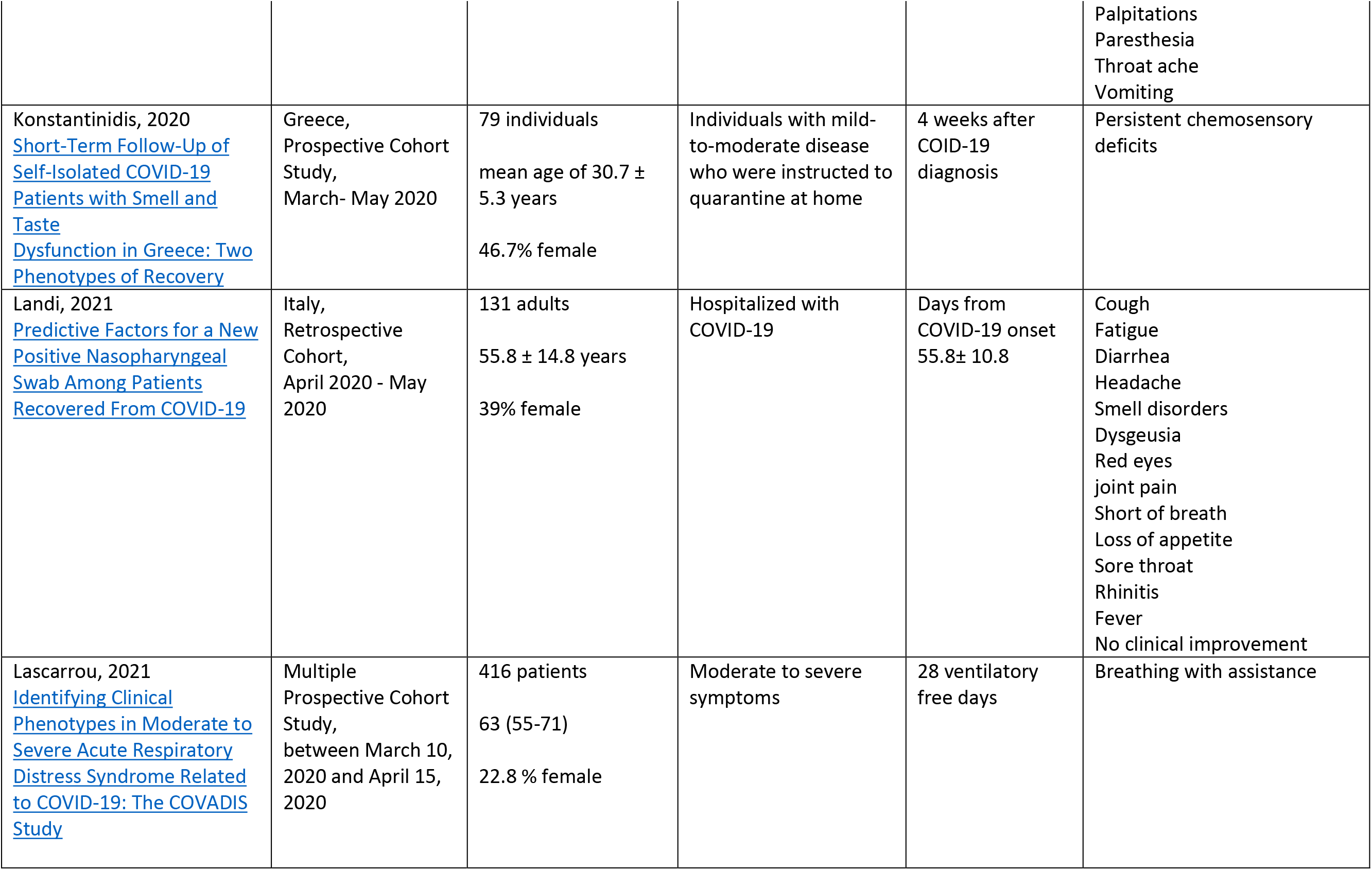
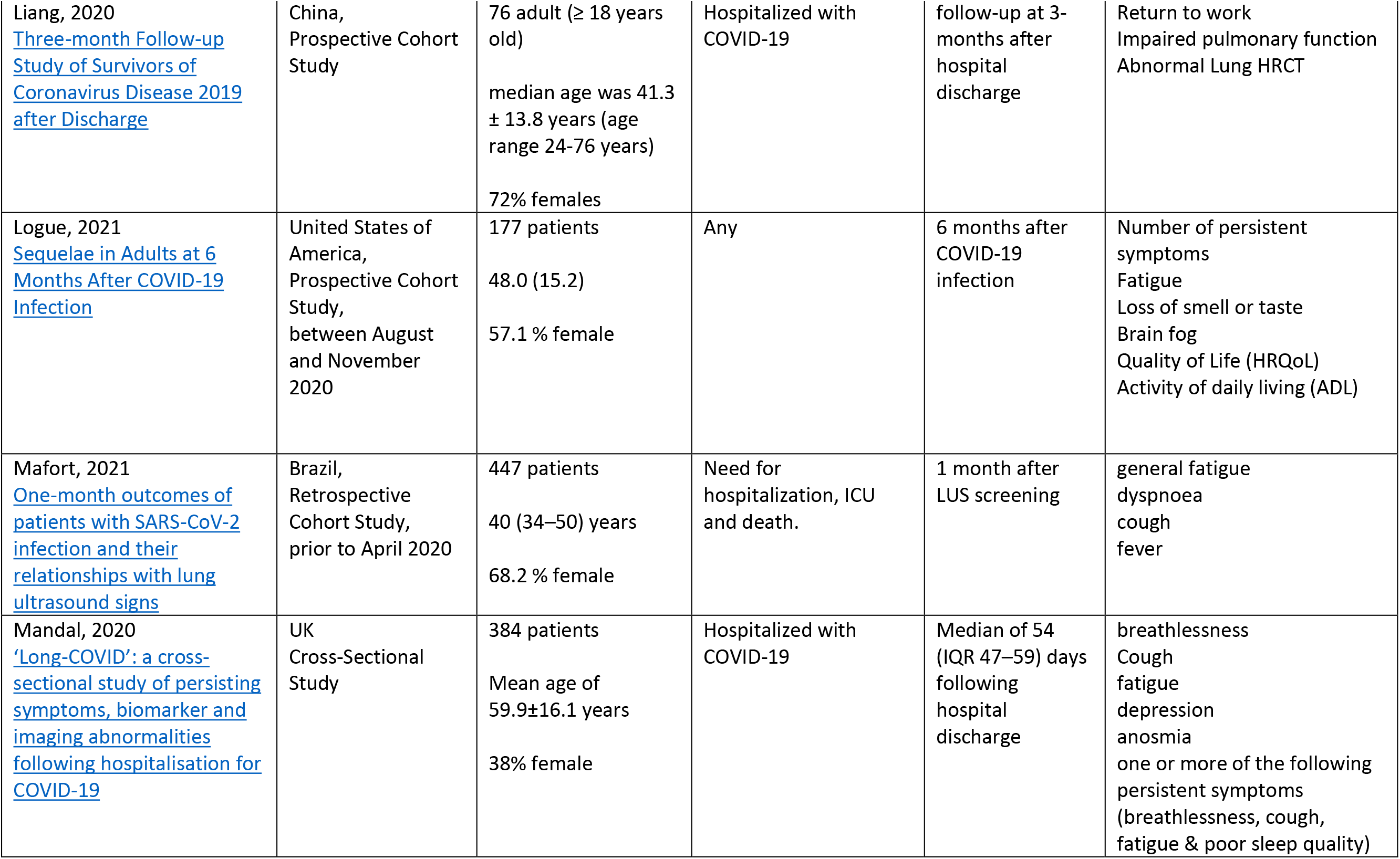
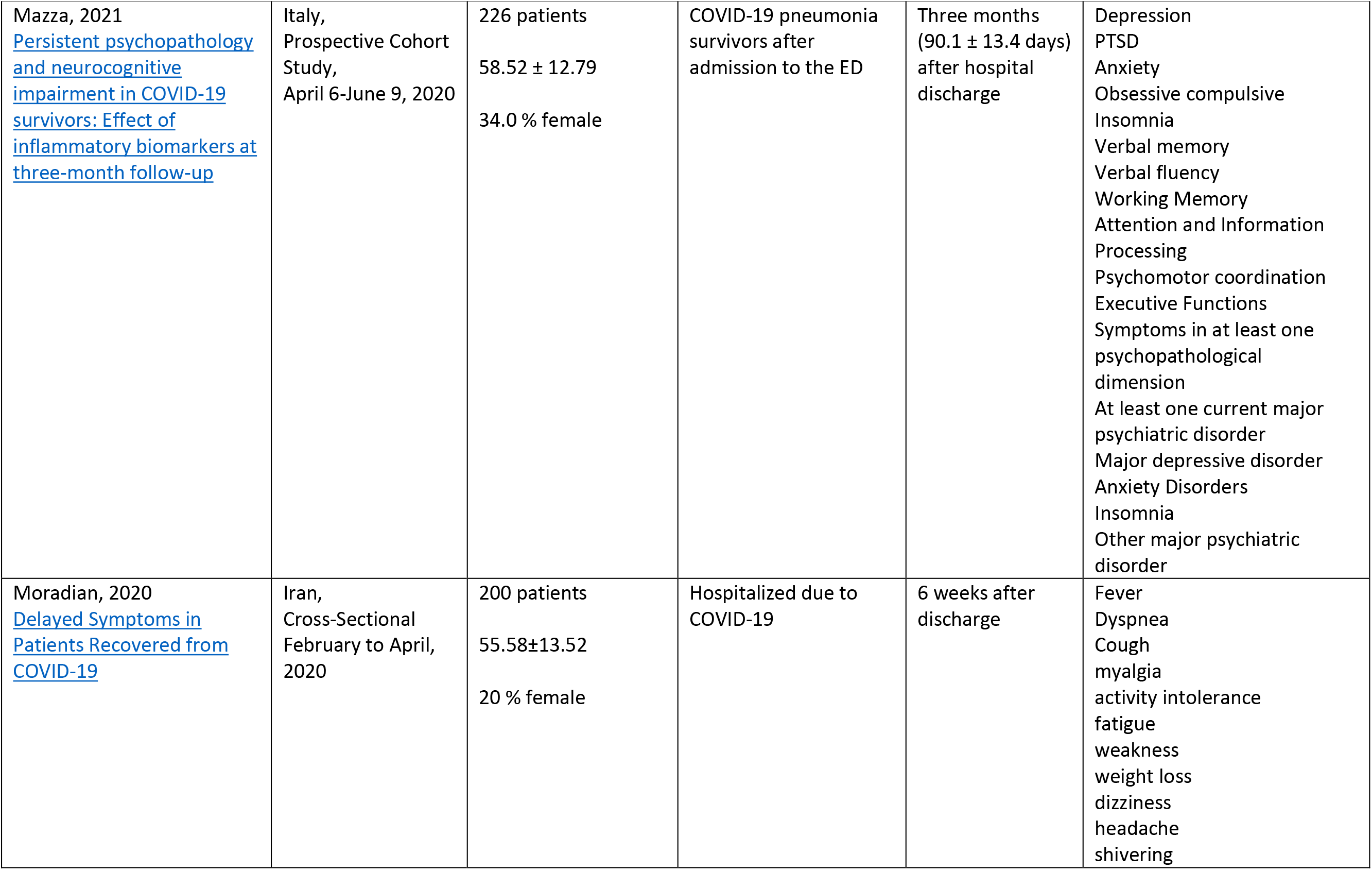
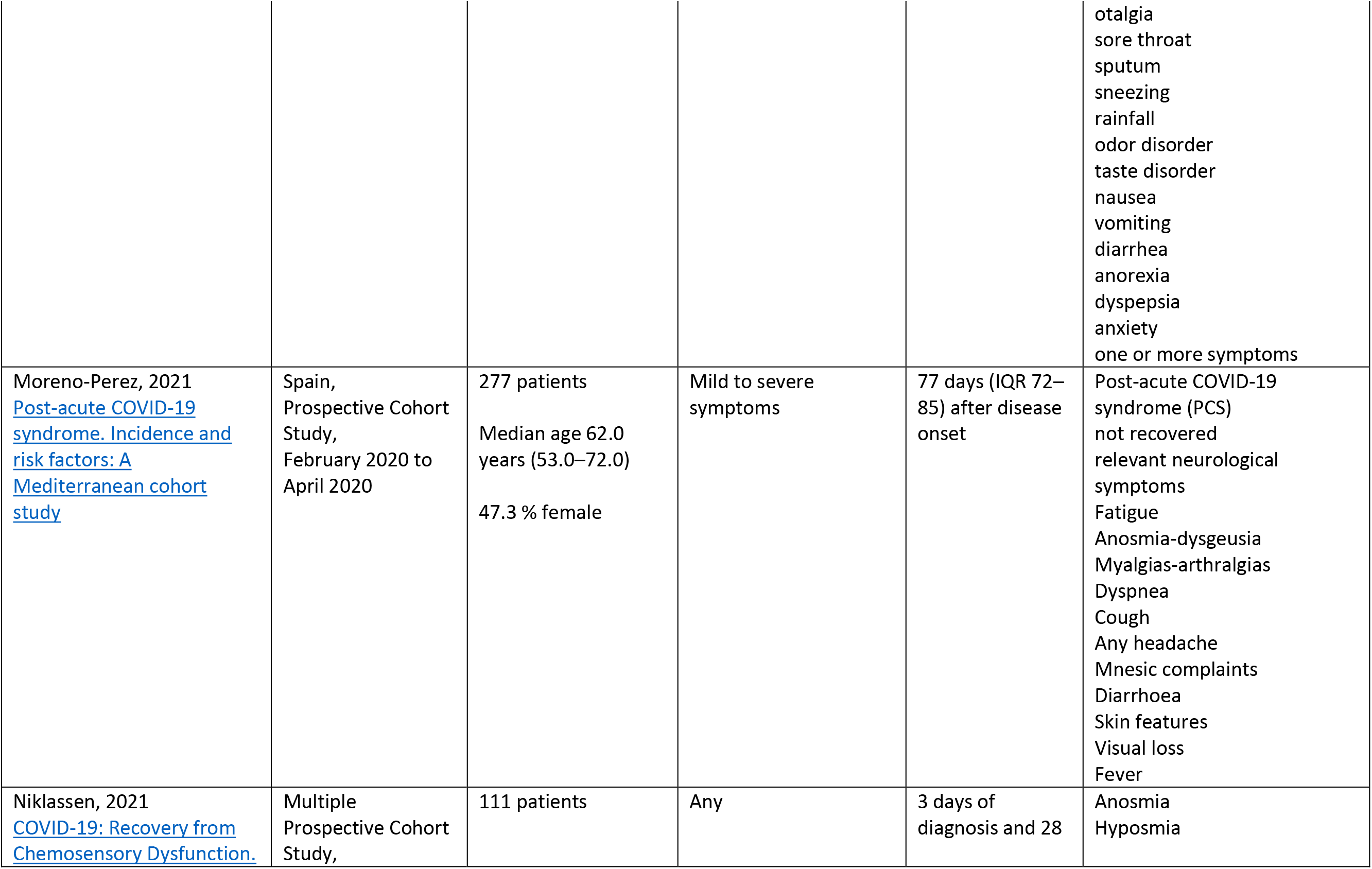
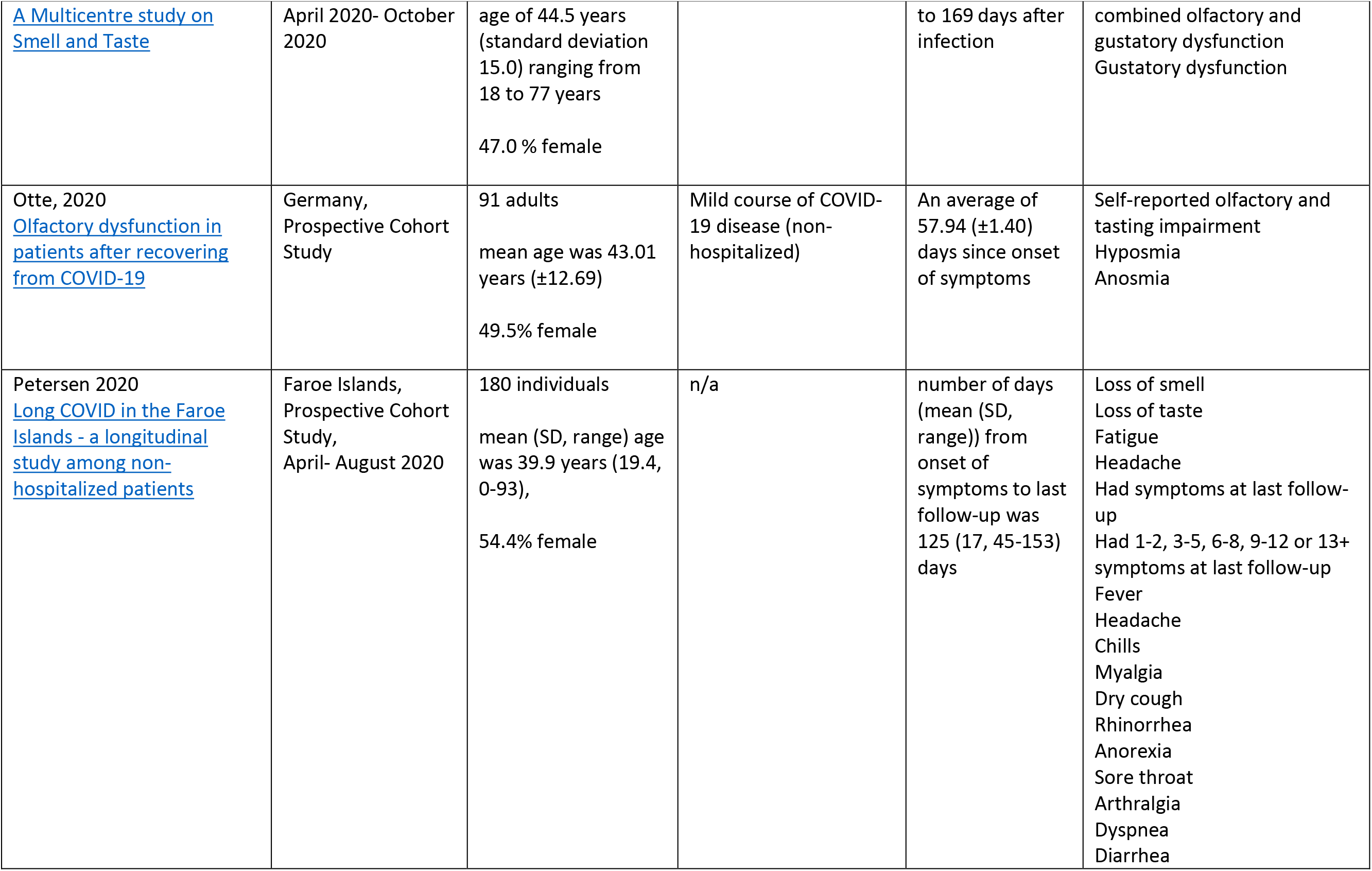
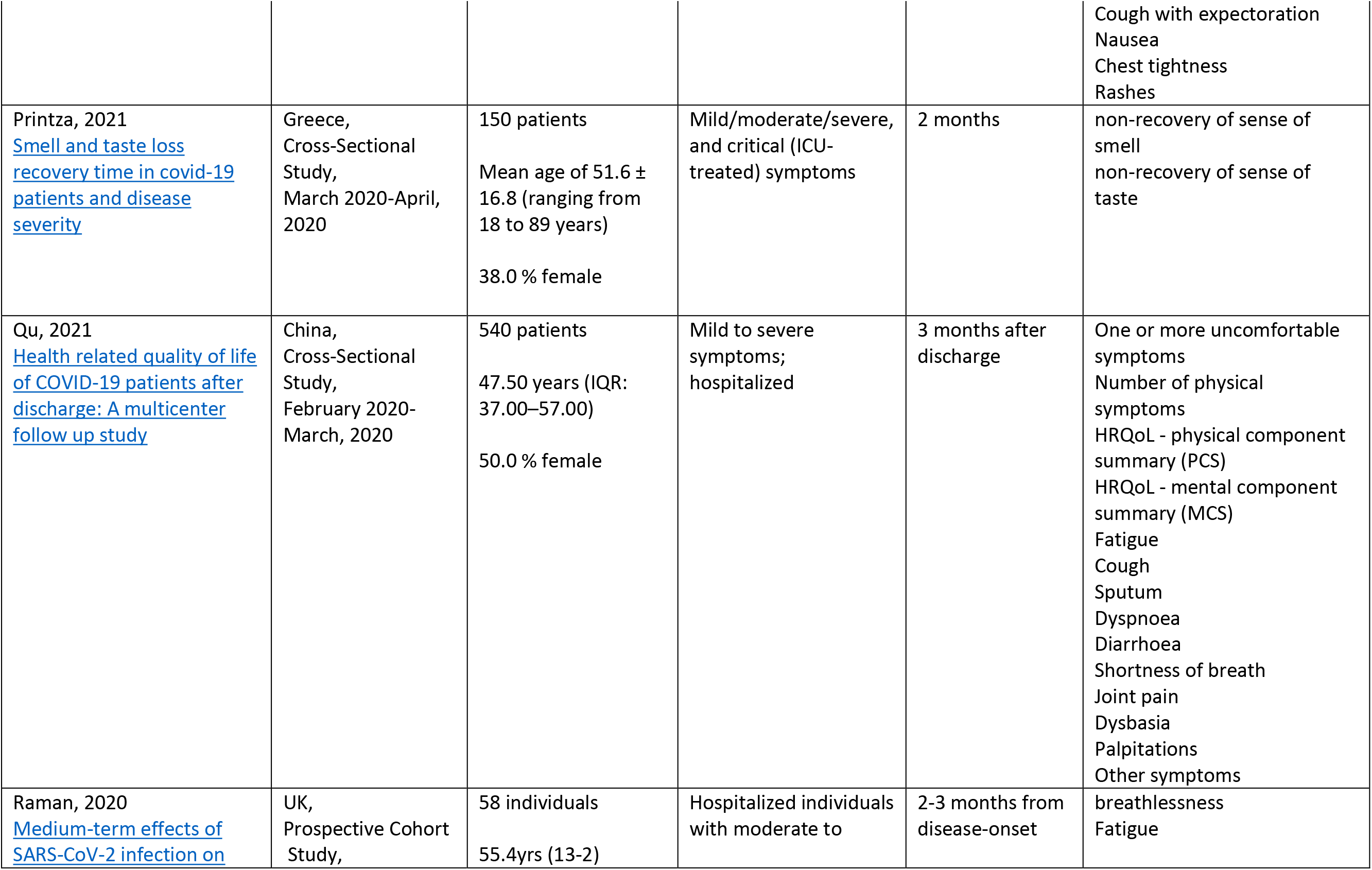
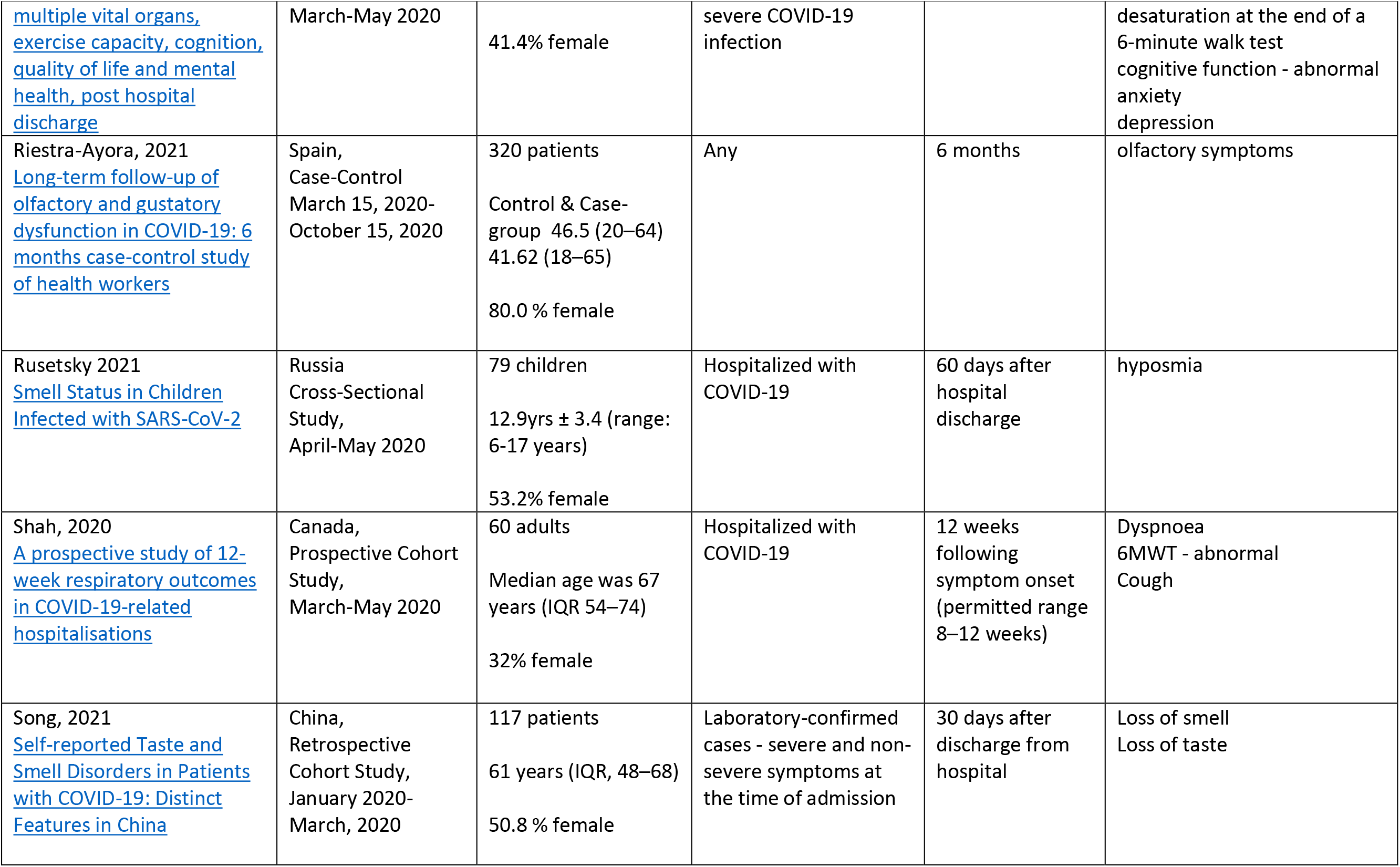
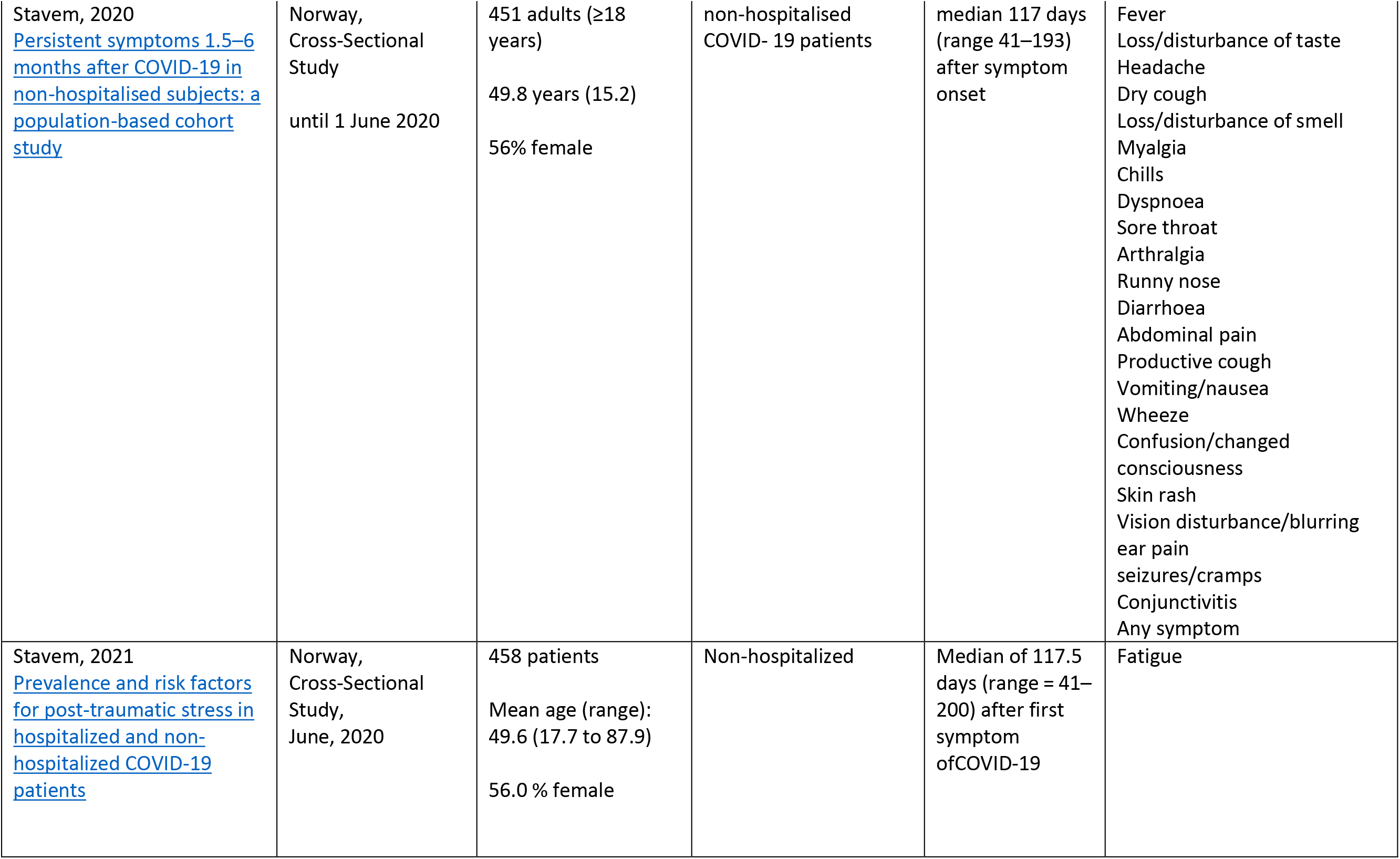
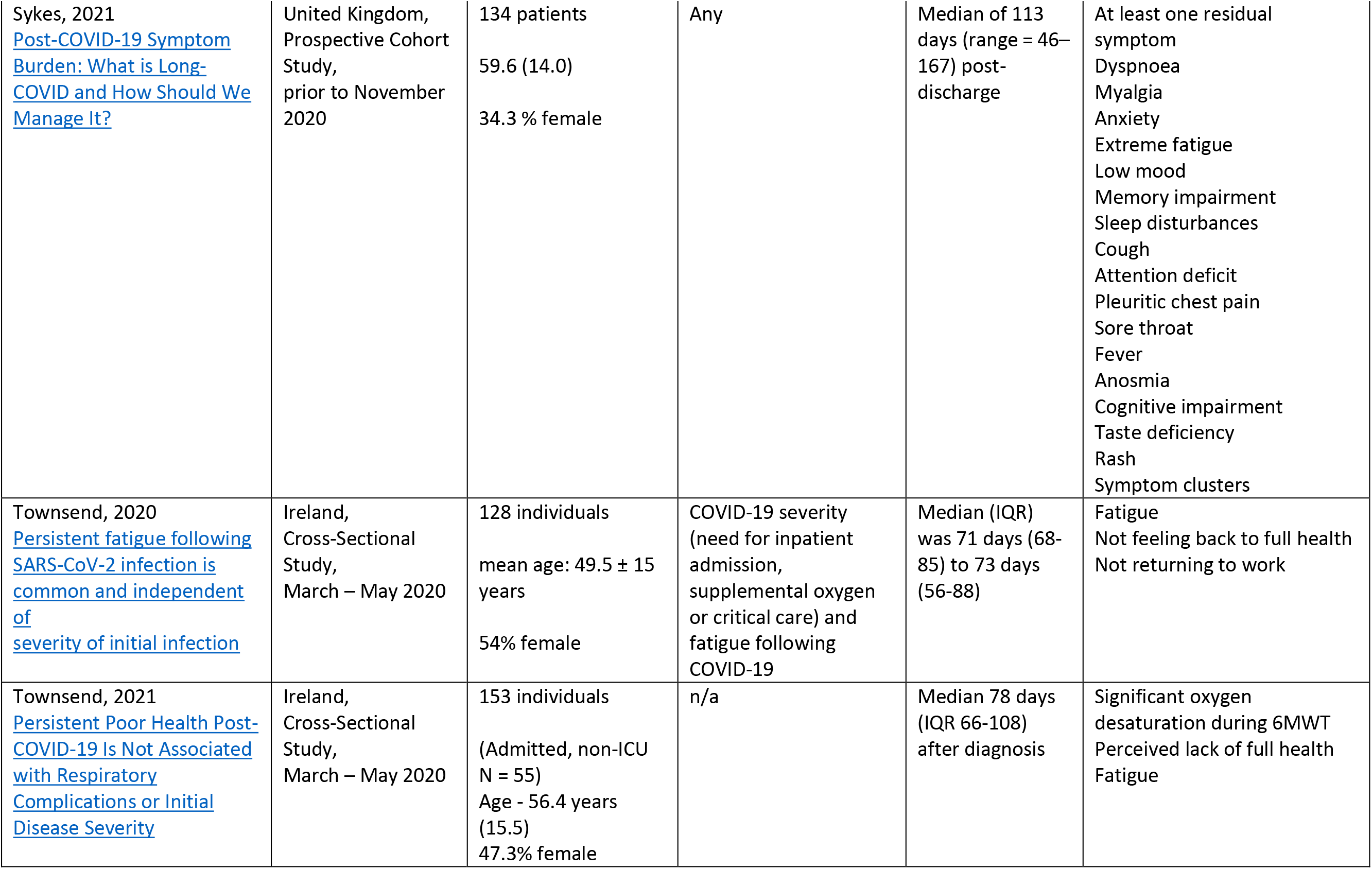
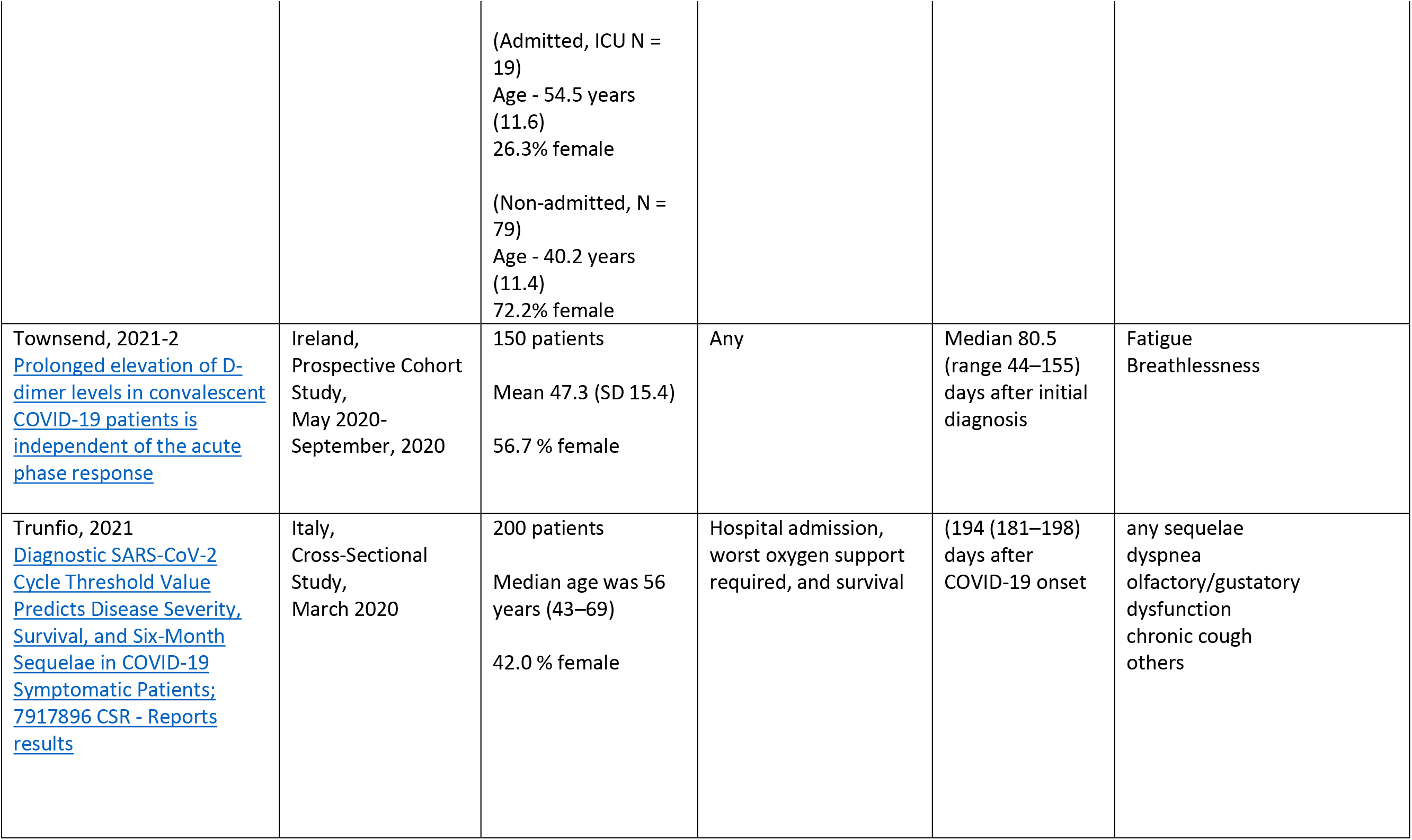
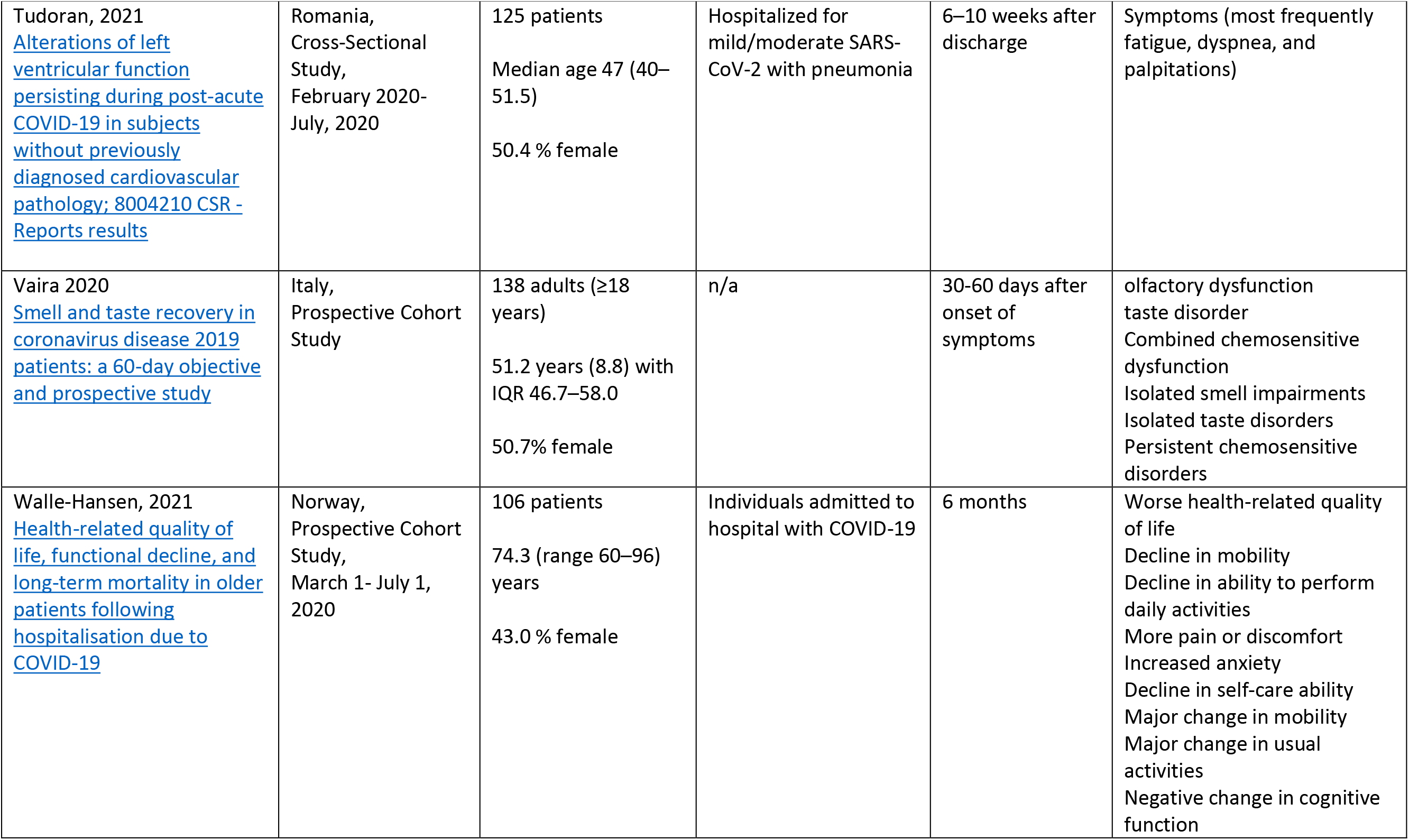
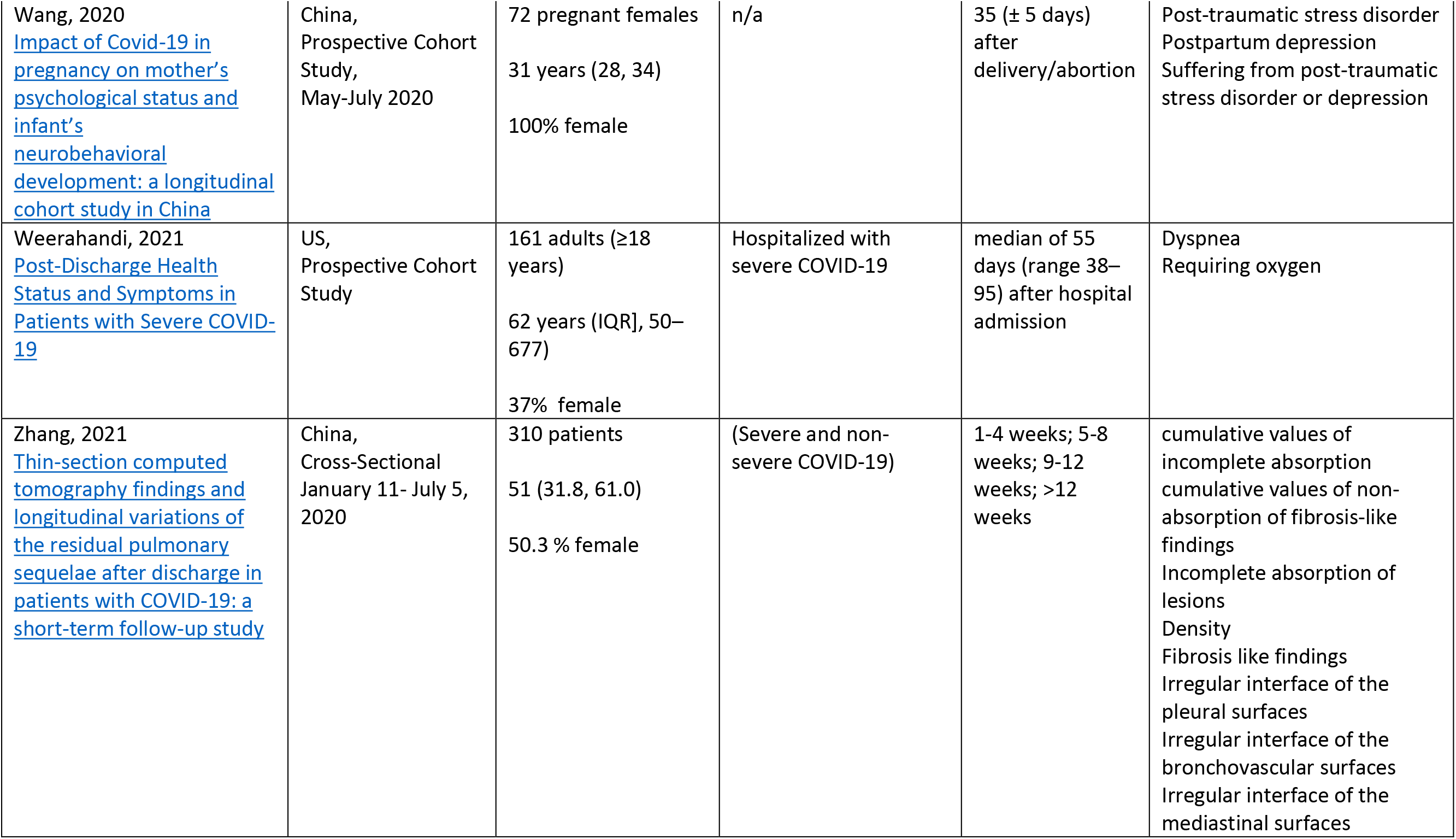
Characteristics of Included Studies with Laboratory-Confirmed Participants

### Prevalence in laboratory-confirmed COVID-19 population (all ages)

#### Study Characteristics

Study characteristics of 63 studies with data from participants with laboratory-confirmed COVID-19 are shown in Table 1. All studies were observational (cohort or cross-sectional) and included between 58 and 1733 individuals, with most studies (42/63) having less than 200 participants. The majority were conducted in Europe (38/63), with the remaining in Asia (8/63), North America (6/63) and others (11/63). More than half of the studies (37/63) only recruited adult participants, 24/63 did not restrict recruitment by age, and 2/63 studies focused on a pediatric sample. Almost half of the studies (27/63) only recruited participants who were hospitalized or admitted to the intensive care unit (ICU) due to COVID-19. Outcome data were reported with mean or median times from COVID-19 diagnosis occurring in the short-term for 56% (35/63), in the long-term for 36% (23/63) and in both the short- and long-term for 8% (5/63) of the studies.

#### Risk of bias and certainty of the evidence

Of the 63 studies, we assessed 45 to be at moderate risk of bias and 18 to be at high risk of bias [Supplementary Table 2]. The most common sources of potential biases were from participant selection (i.e. convenience samples or study population was not representative of the target population) and poor objectivity/validity of outcome measurement (i.e. many outcomes were self-reported or obtained using non-validated measures).

We assessed certainty (or confidence) of the body of evidence for 81 key outcomes and found moderate certainty in 6% (5/81), low certainty in 62% (50/81) and very low certainty in 32% (26/81) of these outcomes. (Table 2, Supplementary Table 3)

**Table 2.**
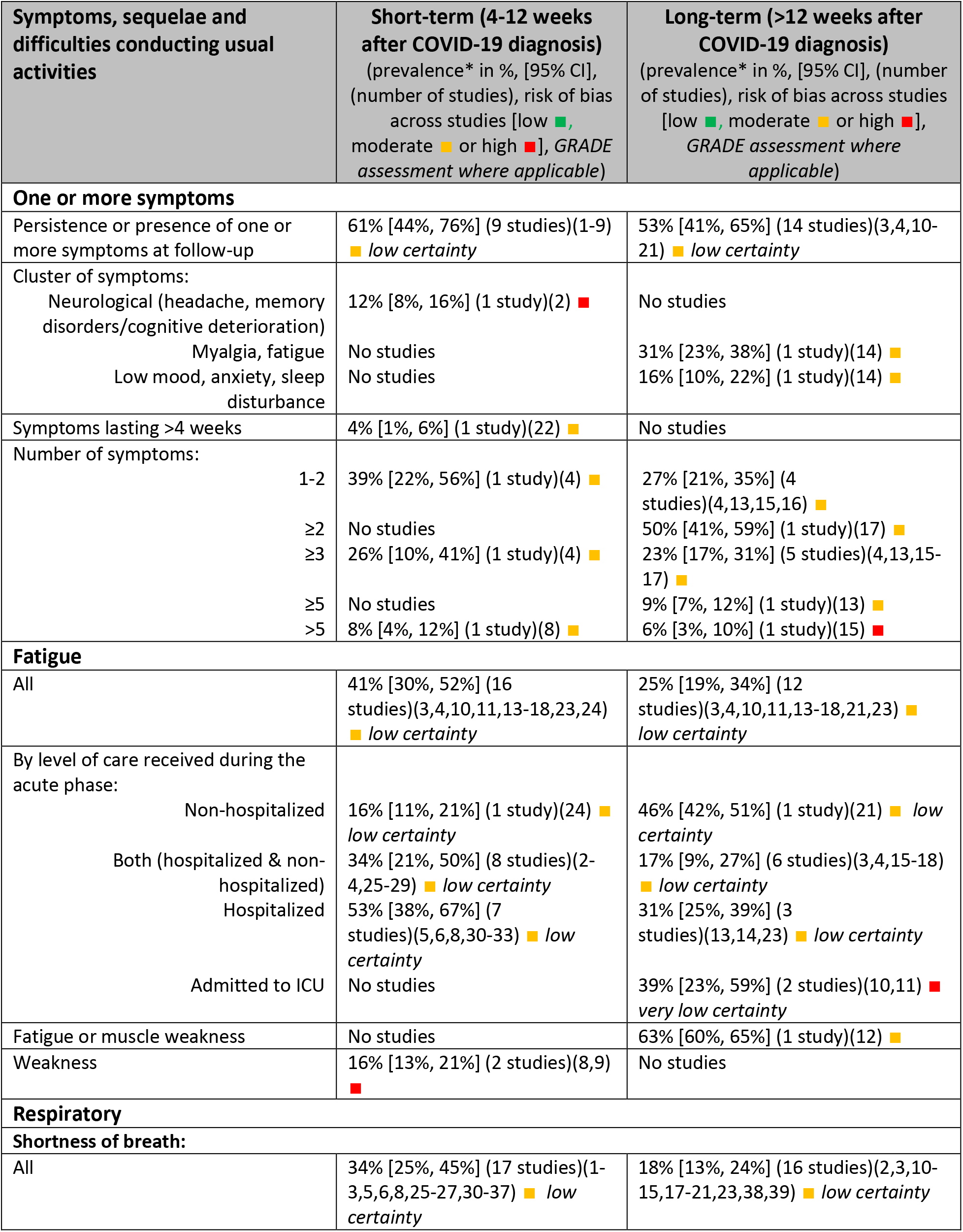
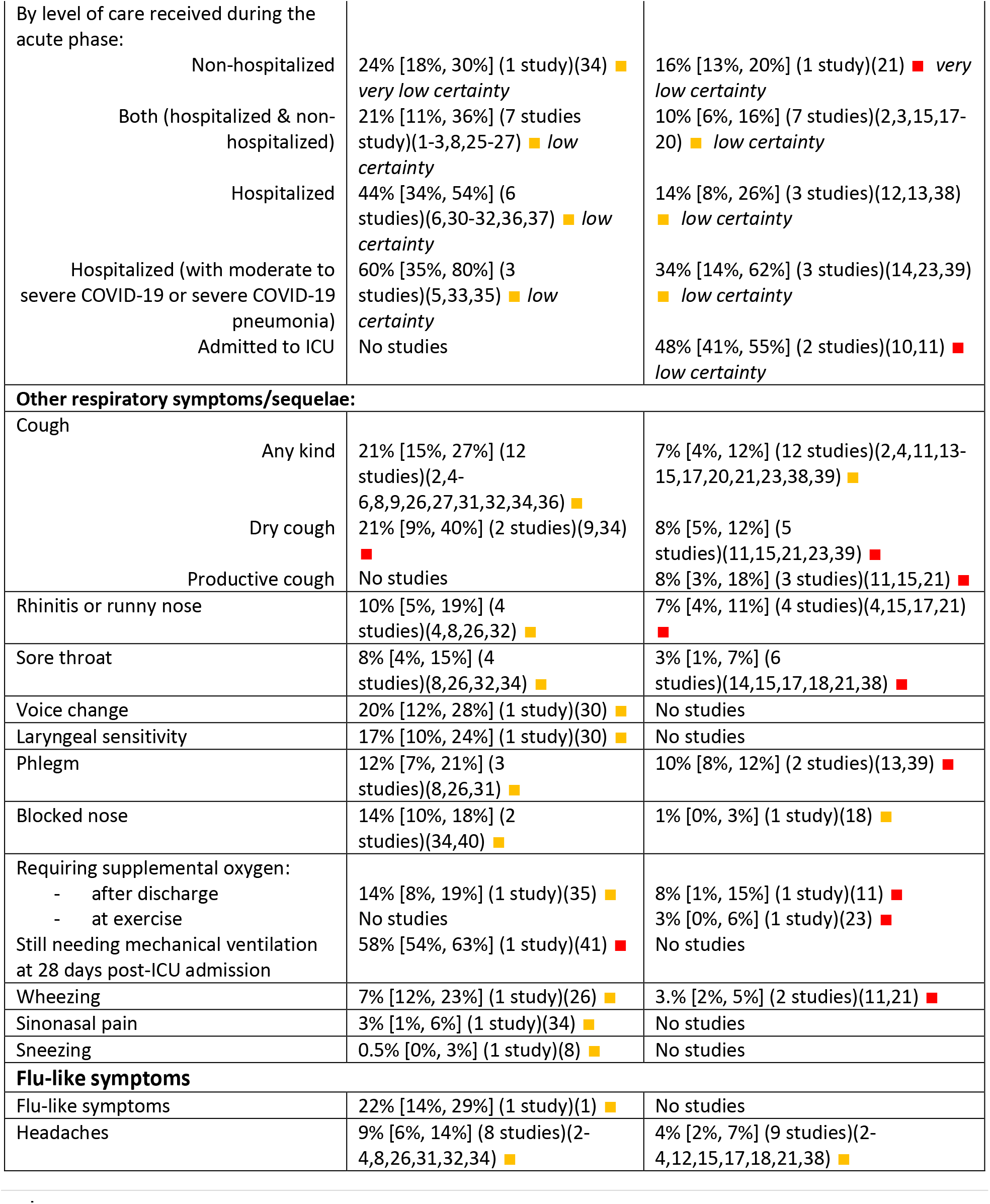
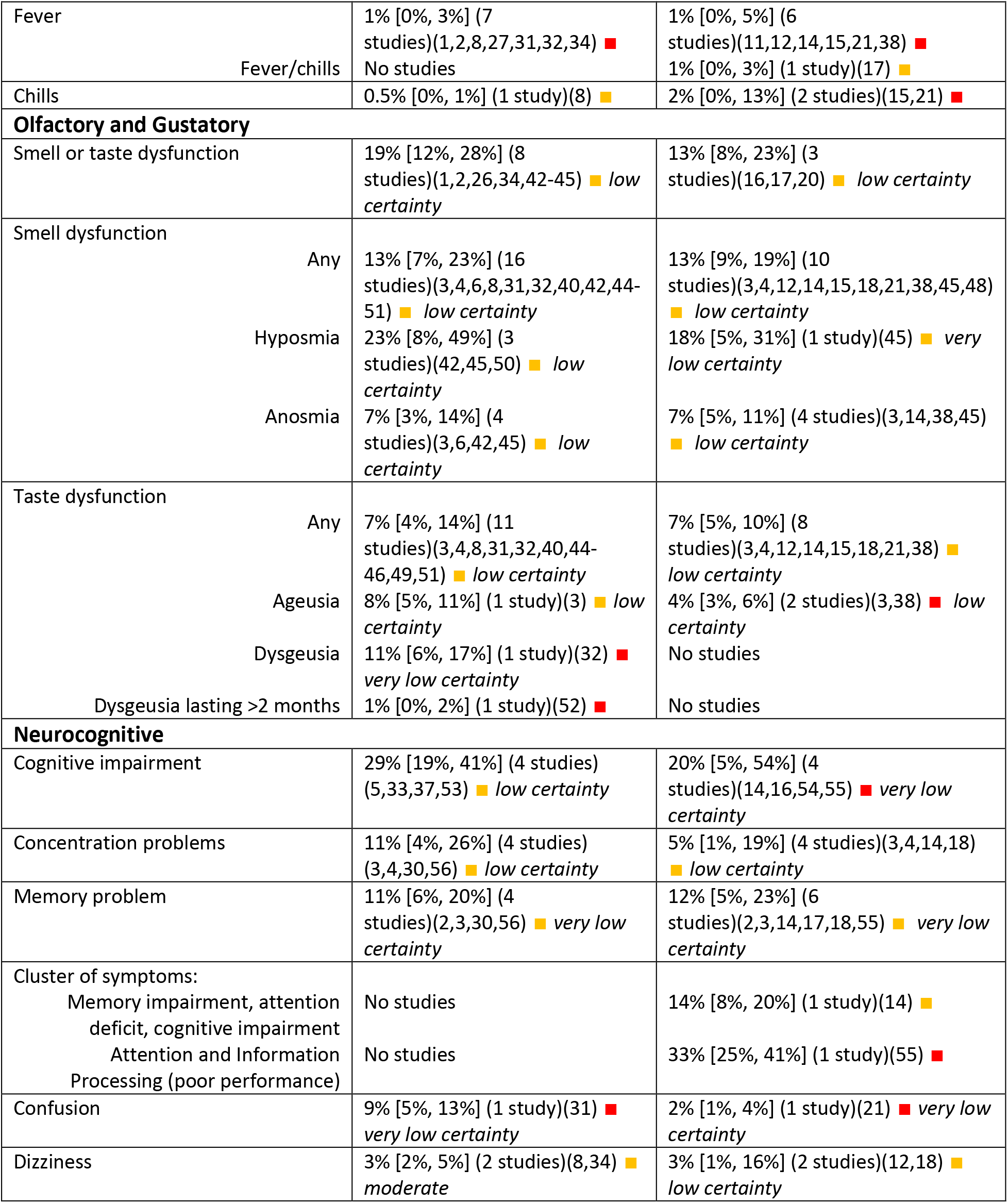
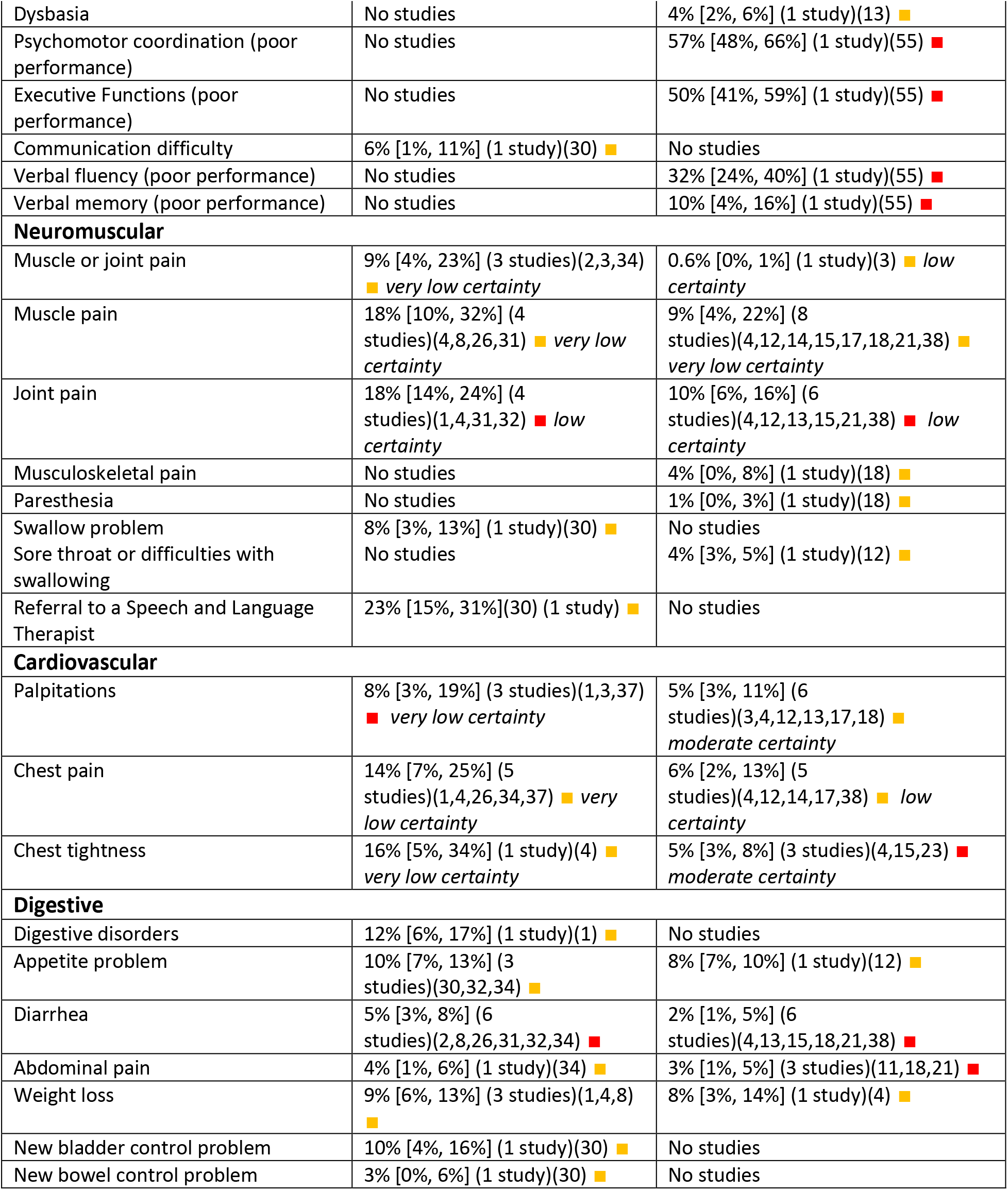
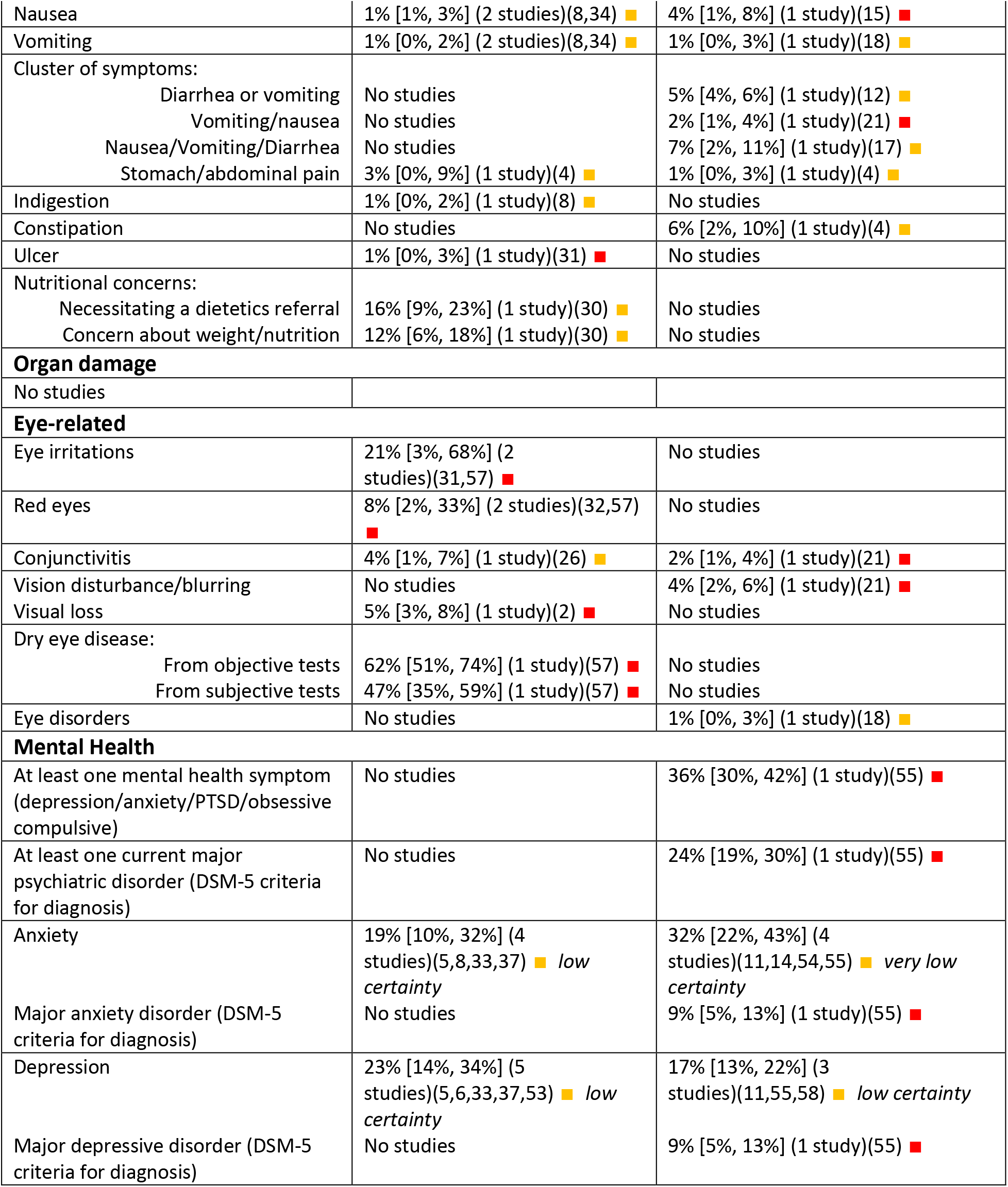
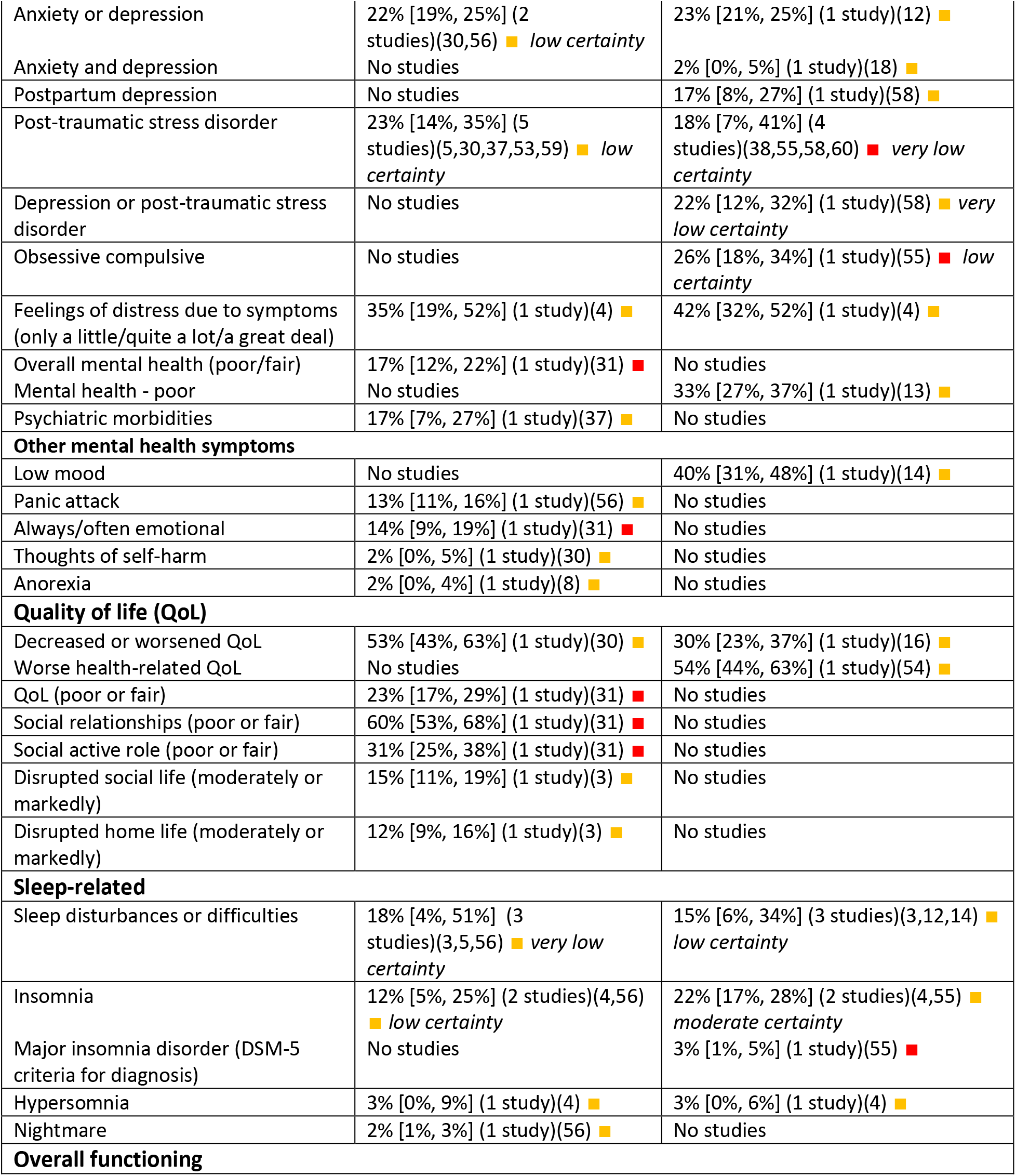
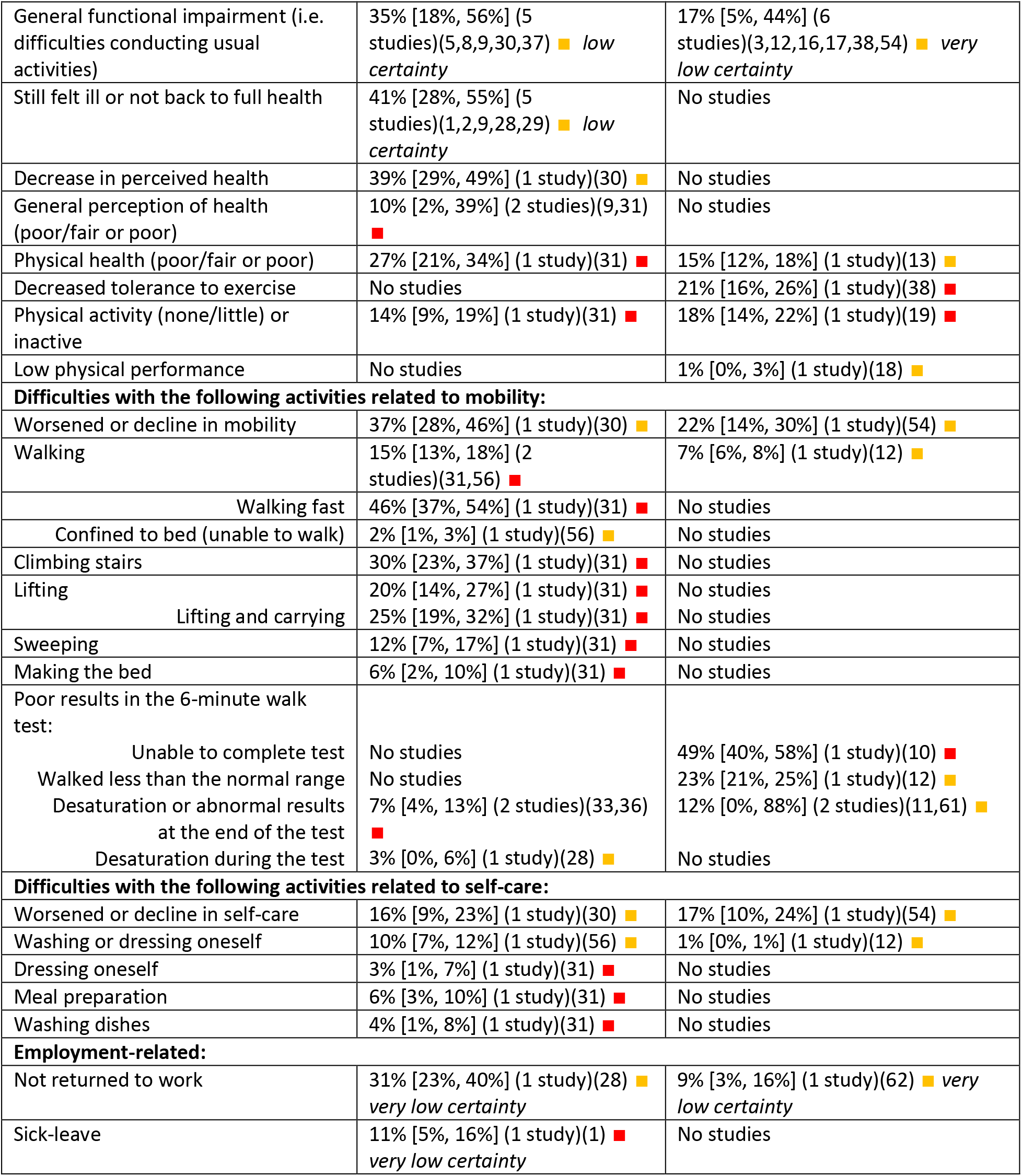
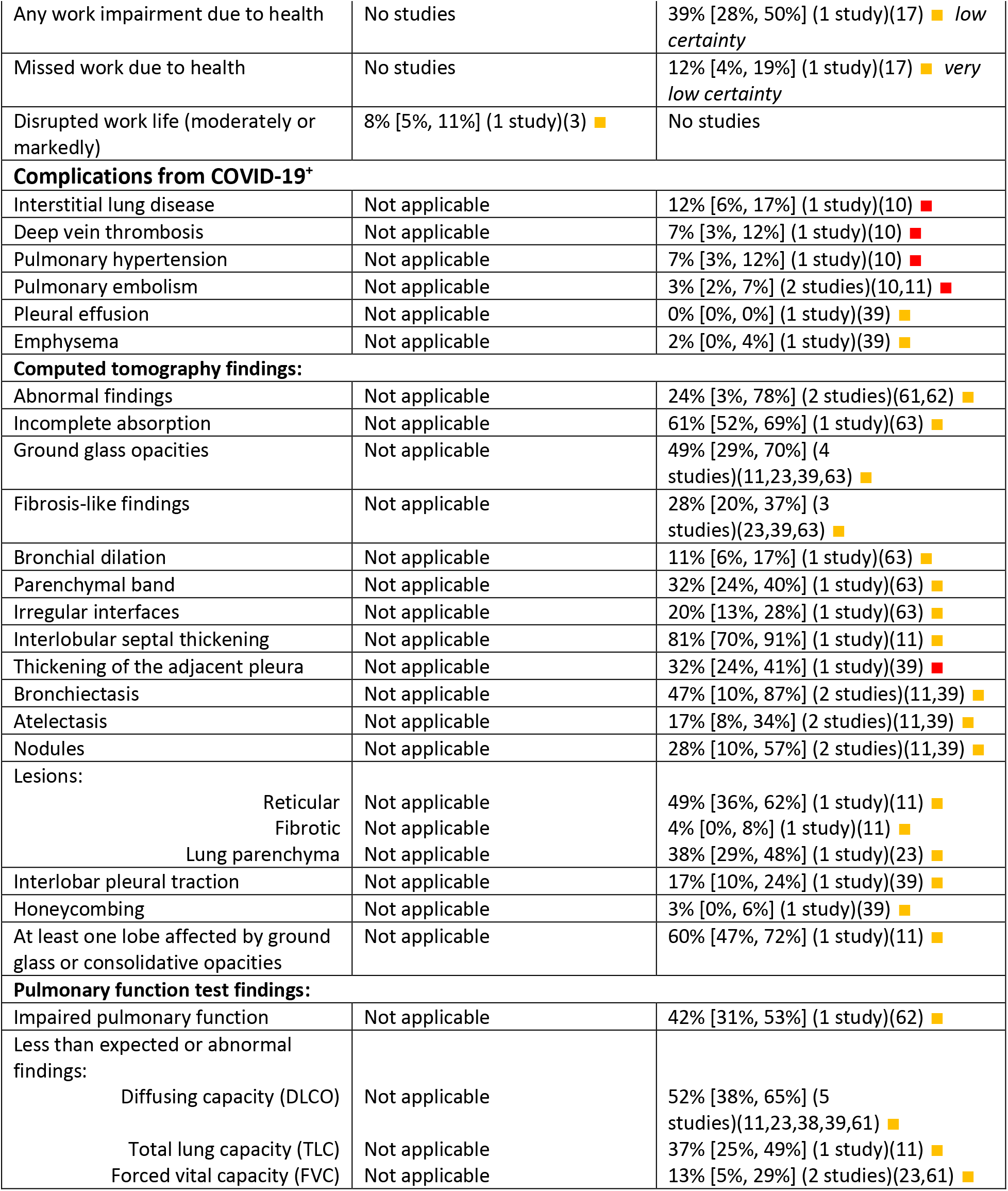
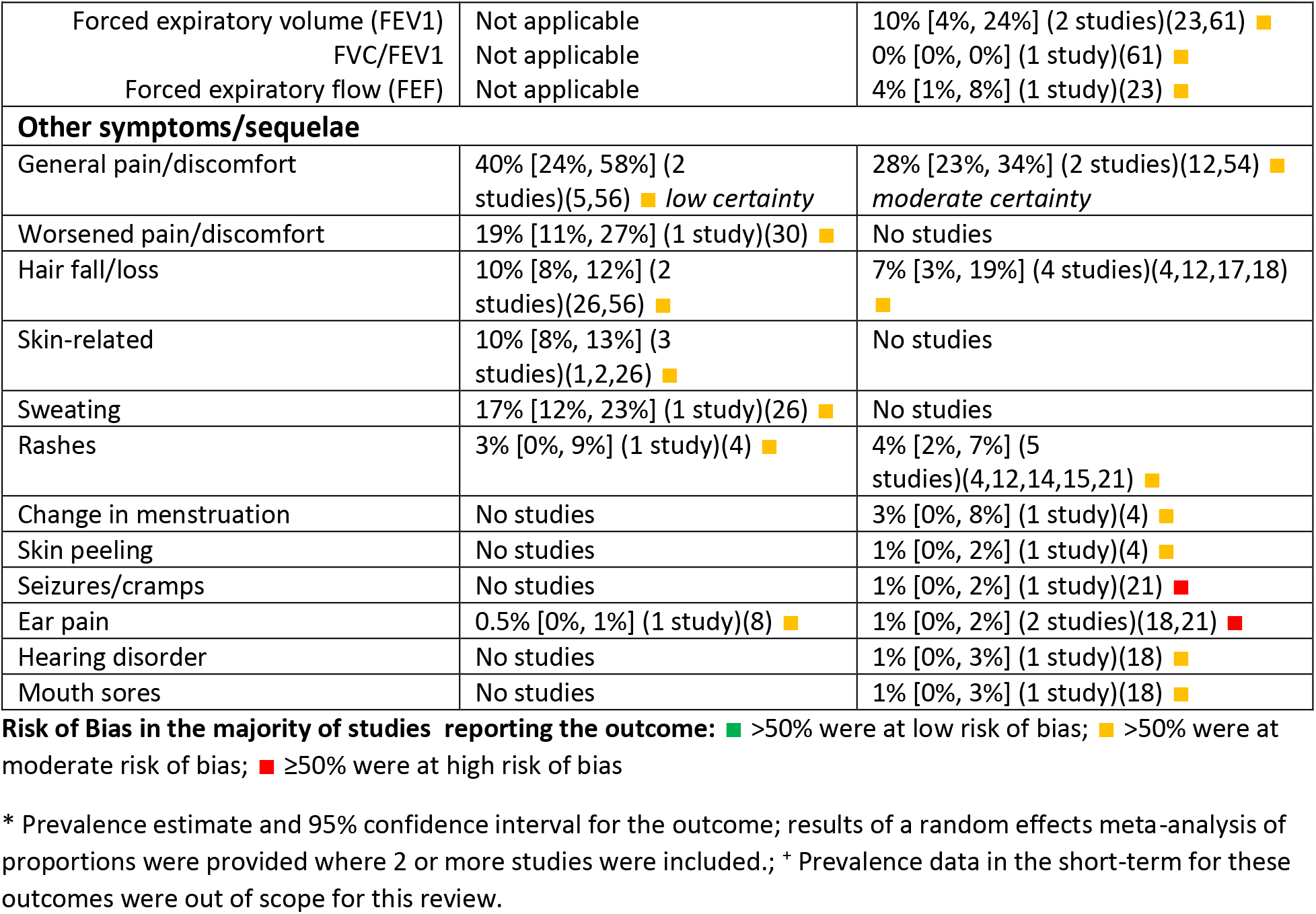
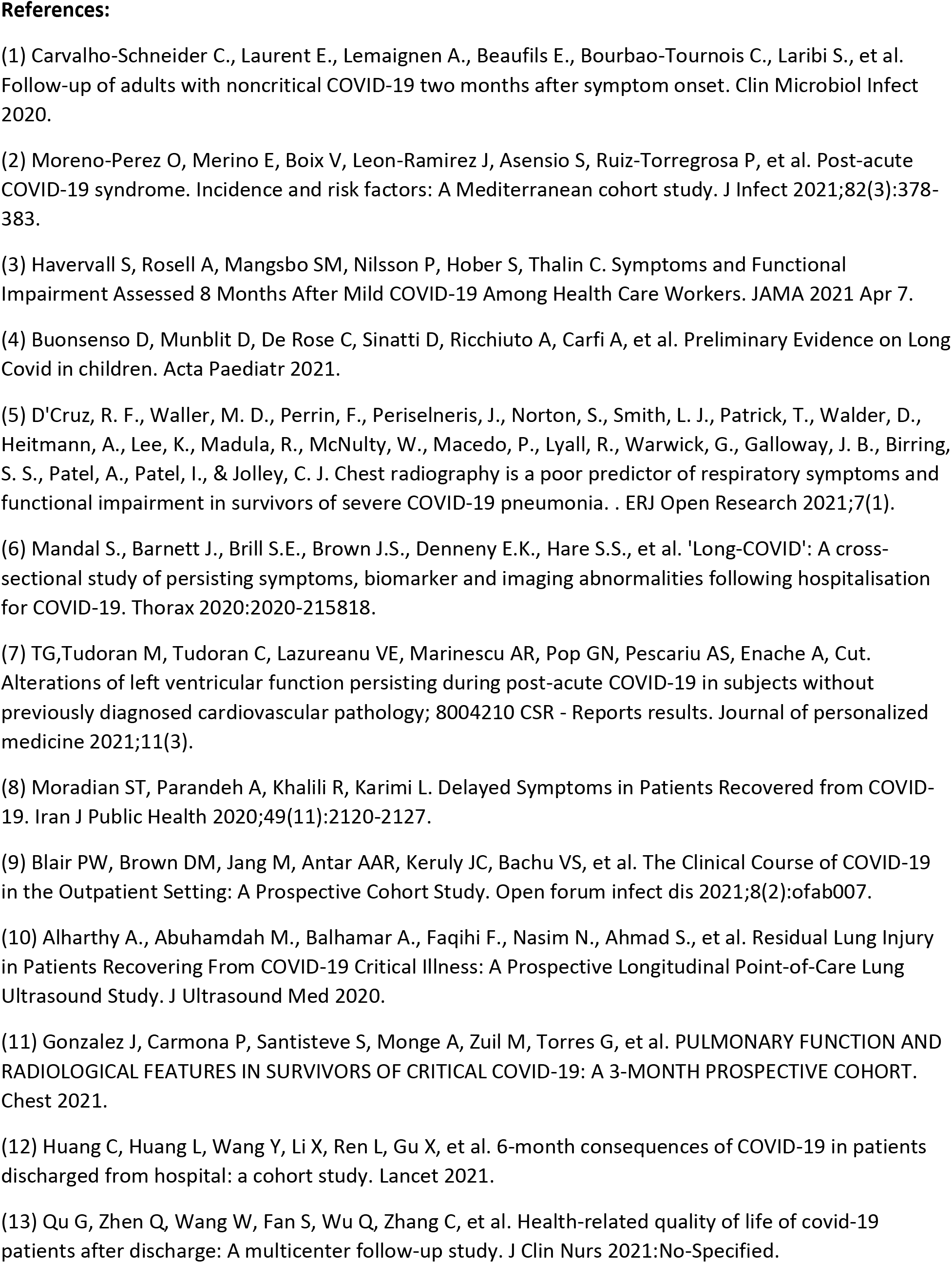
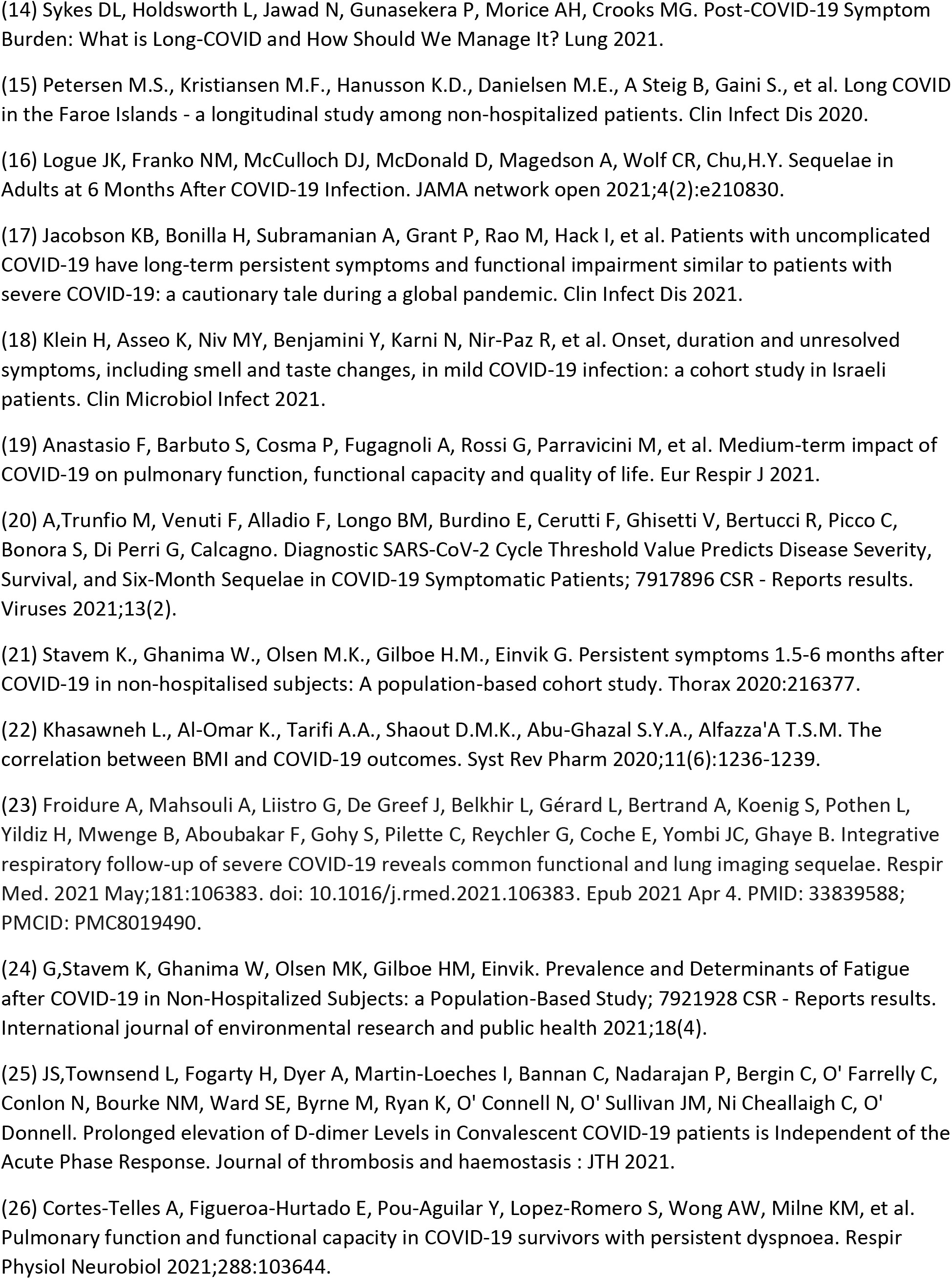
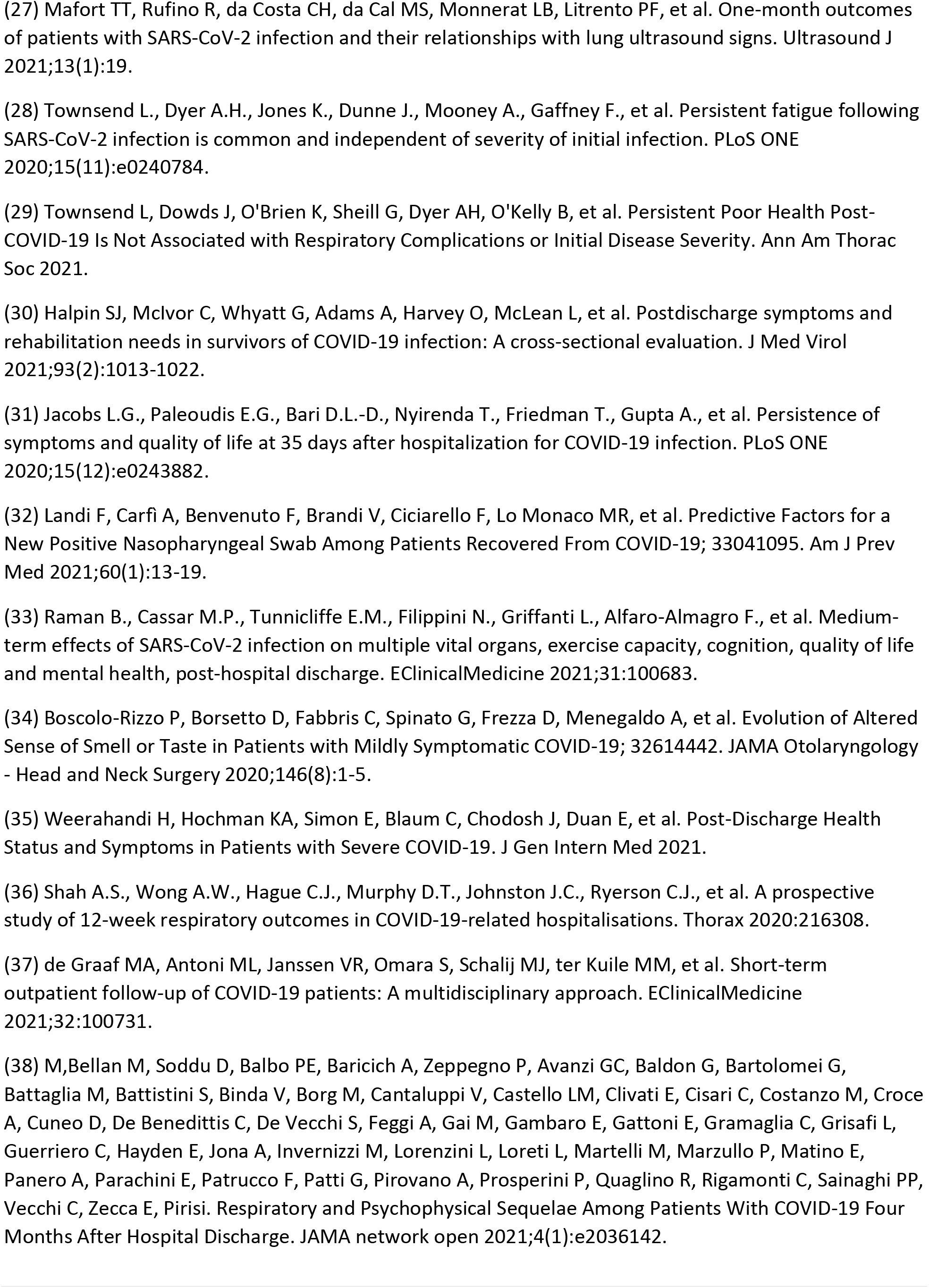
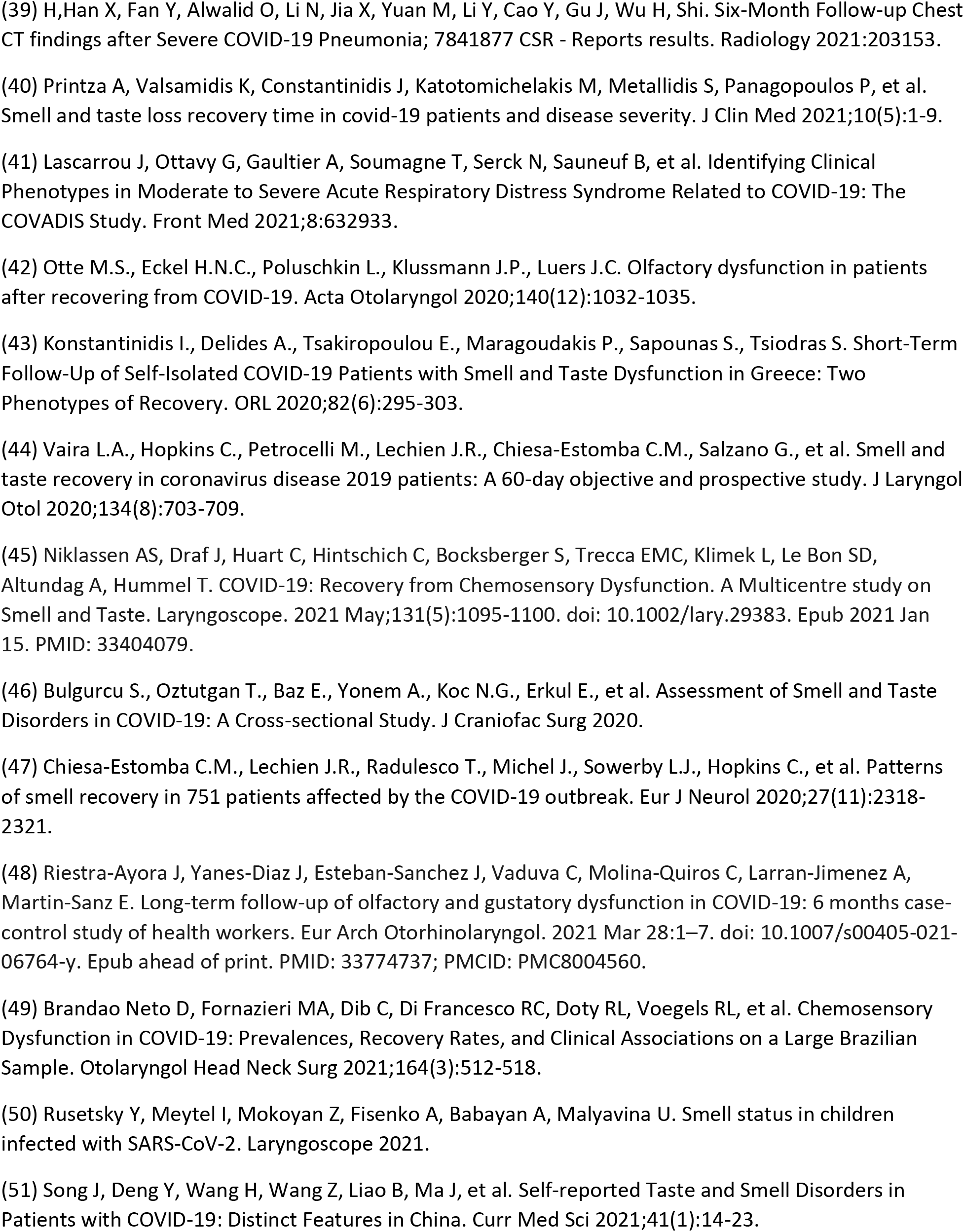
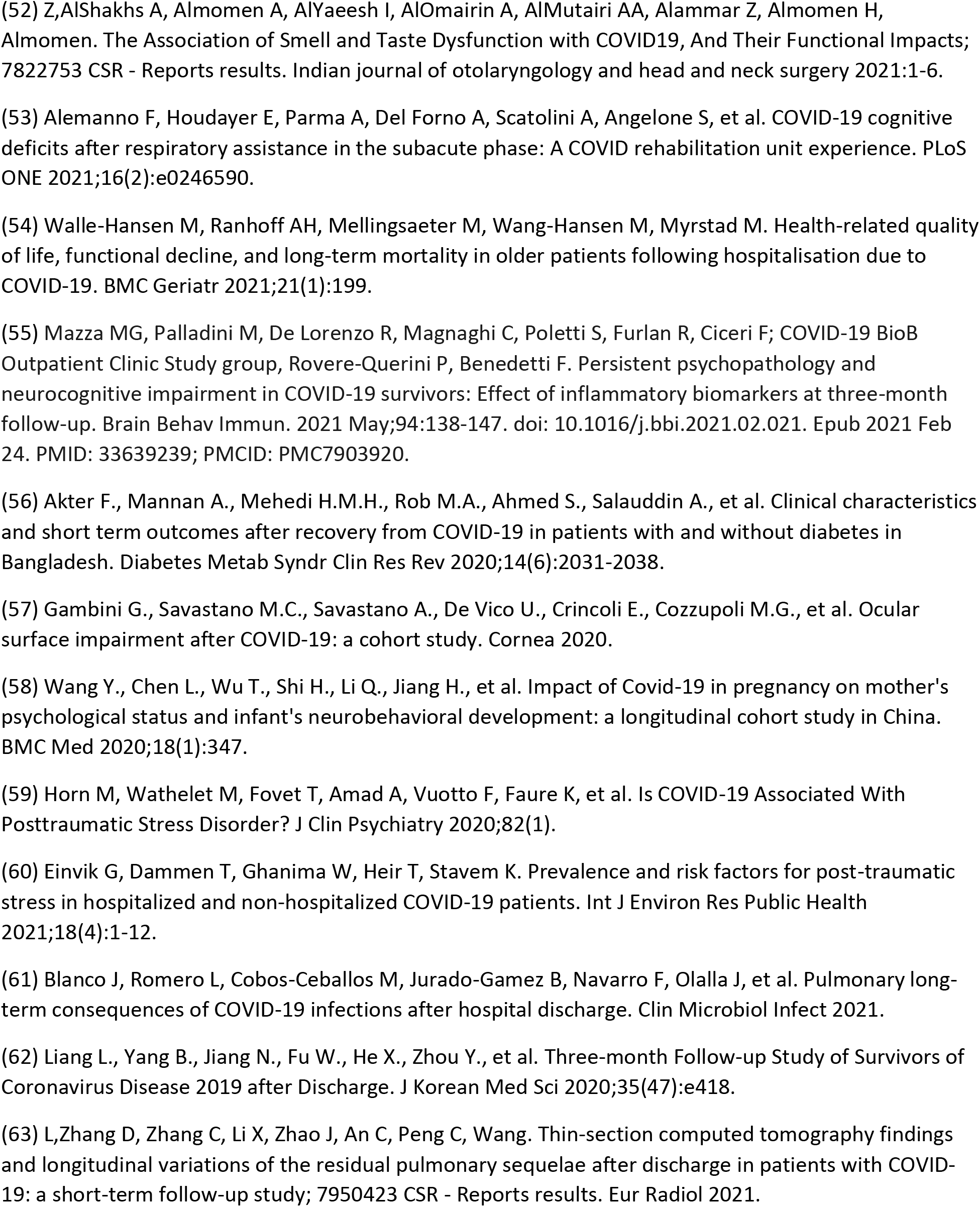
Prevalence of various symptoms, sequelae and difficulties conducting usual activities post-COVID-19 infection in laboratory-confirmed individuals

#### Prevalence of symptoms, sequelae, and difficulties conducting usual activities

Over 100 different outcomes were reported for which the prevalence estimates in the short- and long-term are provided in Table 2.

##### Short-term (4-12 weeks after COVID-19 diagnosis)

Approximately 3 in 5 individuals (61%, 95% CI: 44-76%, *low certainty*) reported the persistence or presence of one or more symptoms in the short-term. The most prevalent symptoms were: fatigue (41%, 95% CI: 30-52%, *low certainty*), general pain or discomfort (40%, 95% CI: 24-58%, *low certainty*), shortness of breath (34%, 95% CI: 25-45%, *low certainty*), cognitive impairment (29%, 95% CI: 19-41%, *low certainty*), depression (23%, 95% CI: 14-34%, *low certainty*), post-traumatic stress disorder (PTSD) (23%, 95% CI: 14-35%, *low certainty*) and hyposmia (23%, 95% CI: 8-49%, *low certainty*). General functional impairment was reported in 35% of individuals (95% CI: 18-56%, *low certainty*) and 41% (95% CI: 28-55%, *low certainty*) reported feeling ill or not back to full health in the short-term.

##### Long-term (>12 weeks after COVID-19 diagnosis)

Approximately half of individuals (53%, 95% CI: 41-65%, *low certainty*) reported persistence or presence of one or more symptoms in the long-term. The most prevalent symptoms were: anxiety (32%, 95% CI: 22-43%, *very low certainty*), general pain or discomfort (28%, 95% CI: 23-34%, *moderate certainty*), fatigue (25%, 95% CI: 19-34%, *low certainty*), insomnia (22%, 95% CI: 17-28%, *moderate certainty*) and cognitive impairment (20%, 95% CI: 5-54%, *very low certainty*). The following symptoms had similar prevalence of 17%-18% (*low to very low certainty*): depression, PTSD and shortness of breath. General functional impairment was reported in 17% (95% CI: 5-44%, *very low certainty*) of individuals in the long-term.

##### Prevalence in children (≤18 years of age)

Only two studies specifically recruited children (<18 years) to determine the prevalence of short- and long-term effects after COVID-19 infection (37, 38). One study (37) reported 58% (95% CI: 50-67%, *low certainty*) of children experienced at least one symptom 162.5 ± 113.7 days after laboratory-confirmed COVID-19 diagnosis, which is similar to the proportion reported for the whole laboratory-confirmed population studied in this review. The most prevalent of these symptoms (ranging from 10-19%, *low certainty*) included: insomnia, nasal congestion or runny nose, fatigue, headache, concentration problems, muscle pain (Table 3). The other study (38) only reported on the prevalence of one outcome in children – hyposmia – and found that none (0%, n=72) experienced hyposmia 2 months after COVID-19 diagnosis.

**Table 3.**
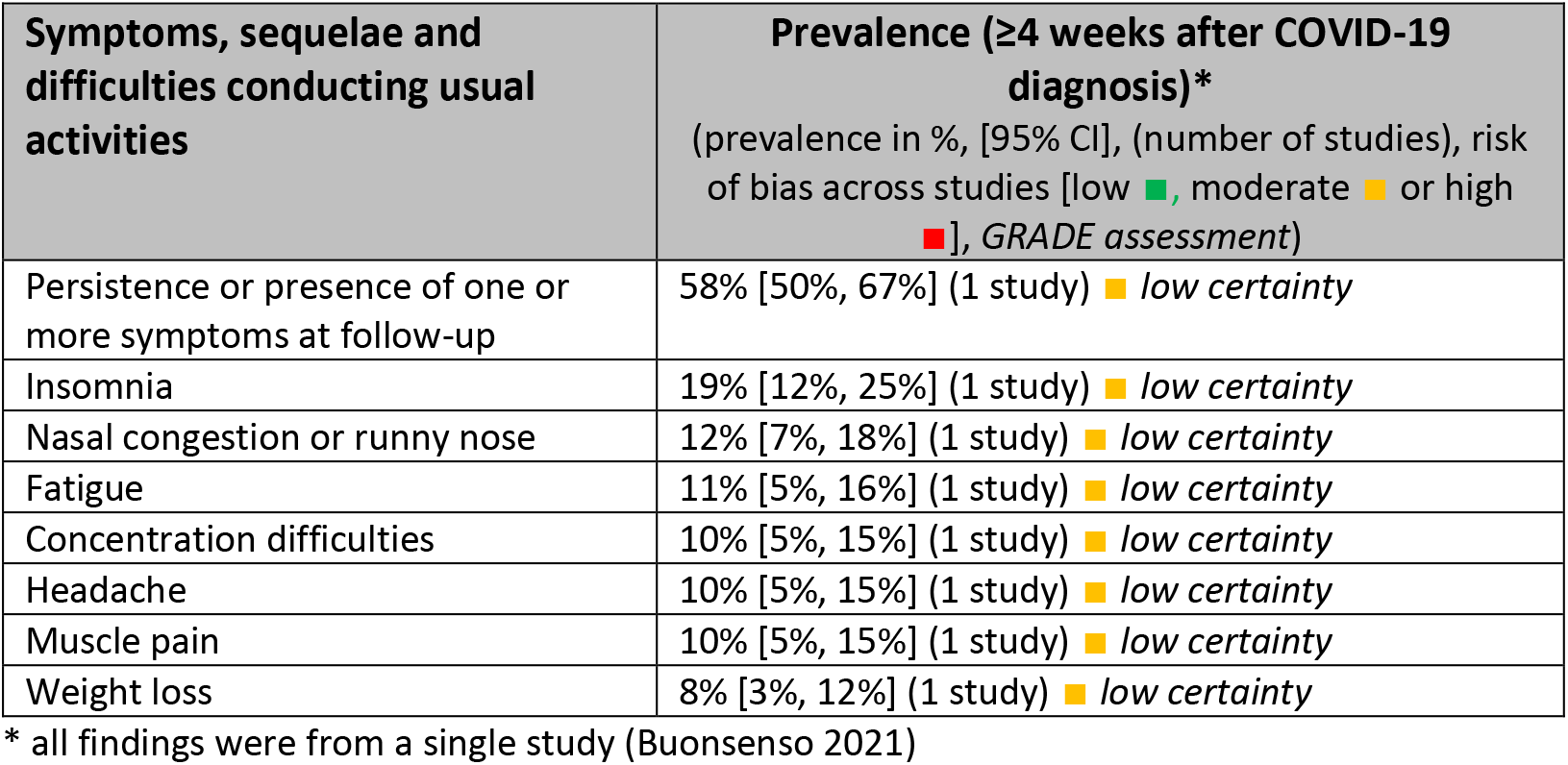
Most prevalent symptoms post-COVID-19 infection in children

### Potential reasons for heterogeneity

Subgroup analyses by level of care received during the acute COVID-19 infection appeared to explain some of the heterogeneity in prevalence of outcomes such as fatigue and shortness of breath (Figure 2, Supplementary File 3). However, we still observed moderate to high heterogeneity within some of the subgroups, particularly among hospitalized populations. Differences in how outcomes were measured (i.e., self-reported versus validated tests) and the thresholds used in each study to indicate an adverse outcome (e.g. binary versus a multi-point scale) may have contributed to differences in prevalence estimates across studies (analyses not shown). In addition, measurement of outcomes at different points or periods of follow-up within the short-or long-term (e.g. outcomes measured at 4, 8 and 12 weeks reported together in the short-term) may have contributed to differences in prevalence estimates across studies; however, there were insufficient data to conduct subgroup analyses at each separate time point.

**Figure 2.**
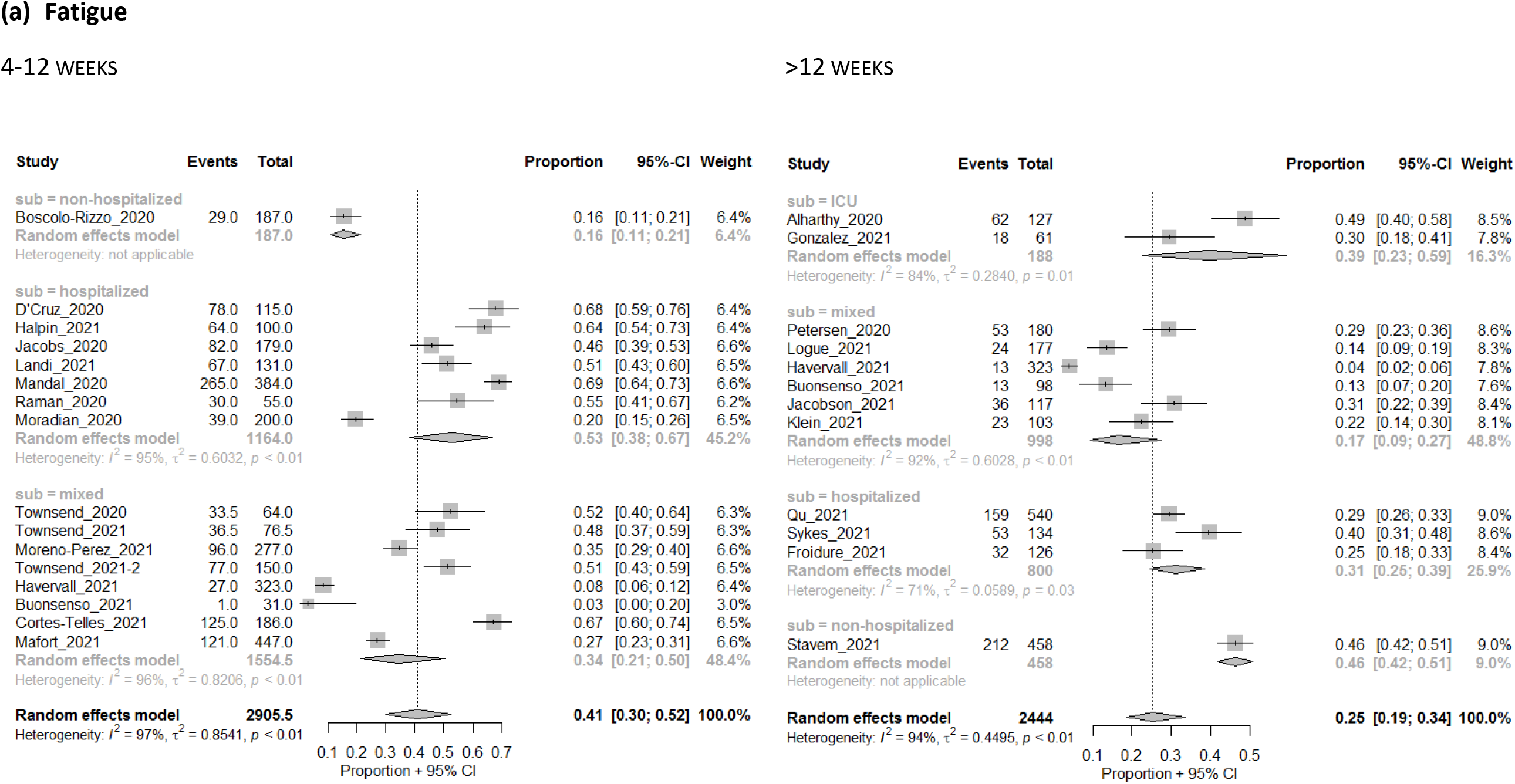
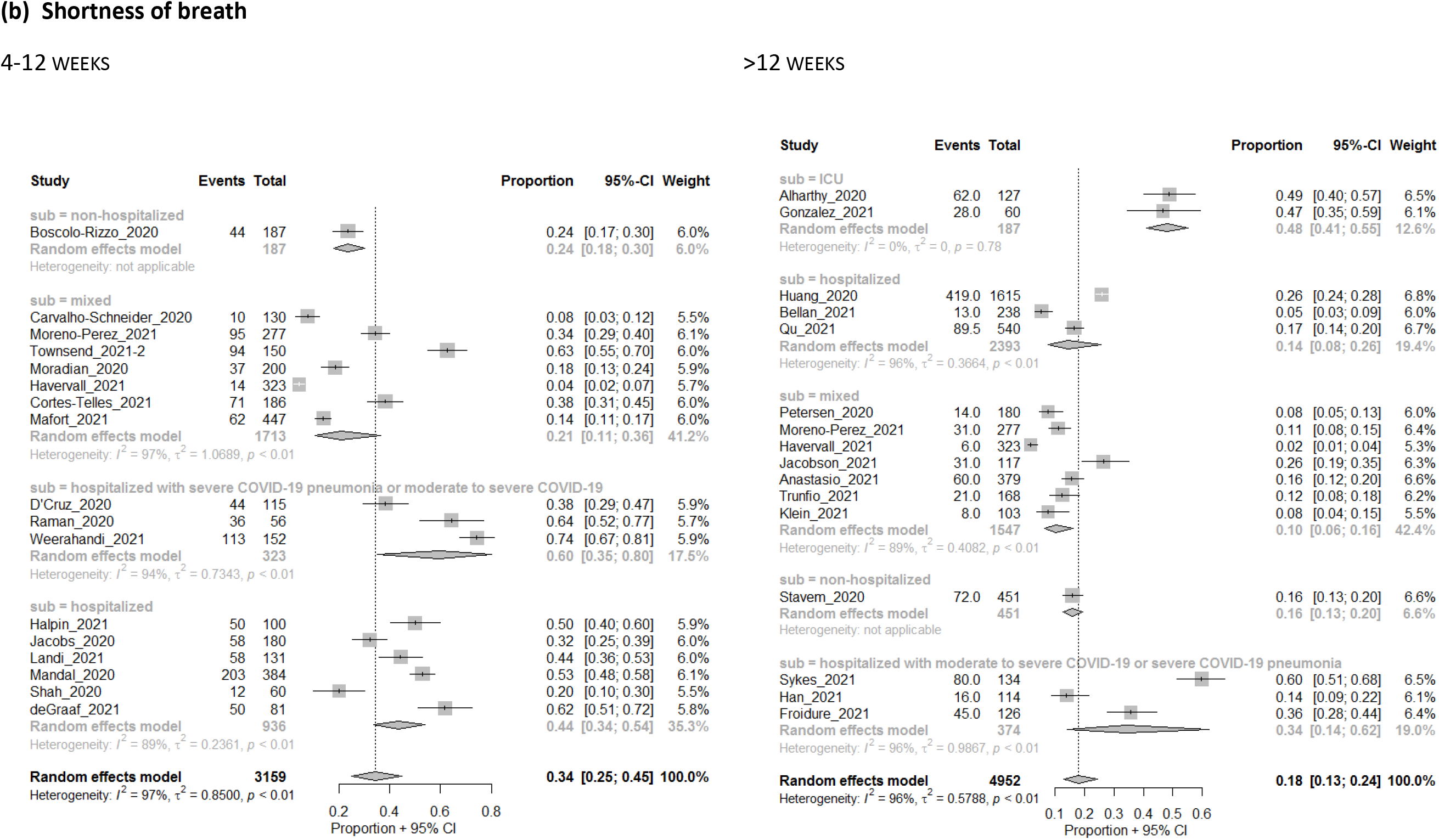
Prevalence (proportion of study sample) of (a) fatigue and (b) shortness of breath, at 4-12 weeks and >12 weeks after COVID-19 diagnosis and by level of care received at the acute stage of COVID-19 infection. Note that level of care may be considered a proxy for severity of COVID-19 (i.e., such that patients with more severe COVID-19 were more likely to require hospitalized care).

### Prevalence in clinically-diagnosed COVID-19 population (all ages)

Characteristics of the 22 included studies with prevalence data for individuals (all ages) who were clinically-diagnosed with COVID-19 are presented in Supplementary Table 1. About half of the studies (12/22) recruited participants who were hospitalized for COVID-19, 64% (14/22) included adult participants only and 55% (12/22) were conducted in Europe. Outcome data were reported with mean or median times from COVID-19 diagnosis occurring in the short-term for 6/22 studies, in the long-term for 13/22 studies and in both the short- and long-term for 3/22 studies. Seventeen of 22 studies were assessed to be at moderate risk of bias while 5/22 were at high risk of bias. [Supplementary Table 2]

The prevalence of various symptoms, sequelae, and complications from COVID-19 in the short- and long-term in the clinically-diagnosed population are presented in Supplementary Table 4.

## Interpretation

Based on data from this systematic review, most laboratory-confirmed COVID-19 patients experienced one or more symptoms in the short-(61%) and long-term (53%) after diagnosis. The most commonly reported symptoms included fatigue, general pain or discomfort, sleep disturbances, shortness of breath, cognitive impairment and mental health symptoms. A large proportion of convalescent COVID-19 patients experienced other mild to severe and debilitating symptoms as well. Consequently, roughly 30% and 10% of individuals were unable to return to work in the short- and long-term following COVID-19 diagnosis, respectively. Because a large proportion of population included in our review represent those who experienced moderate to severe COVID-19 during their acute phase, it is possible that the kinds of symptoms or the frequencies of symptoms reported in those who experienced a milder COVID-19 infection may be different from those reported in our review. In addition, due to limited data available, and low certainty in the existing evidence, clarity around the burden of post COVID-19 condition will require additional research to untangle the sequelae caused directly by COVID-19 infection from those arising from related factors such as extensive hospitalization due to severe illness. The prevalence and complex nature of this condition will require multi-disciplinary approaches in developing appropriate diagnostic models and tools, patient care pathways, and support structures to address the needs of those suffering from post COVID-19 condition.

Several systematic reviews, most of which conducted meta-analyses to arrive at prevalence estimates for the long-term effects of COVID-19, similarly reported two or more of the following symptoms as the most frequently reported: fatigue, shortness of breath, mental health-related symptoms and cognitive impairment (13,19–26). Such similarities in findings across reviews helps advance our understanding of post COVID-19 condition and its burden, and substantiates the need for ongoing support for many COVID-19 survivors. Where prevalence estimates for specific symptoms or sequelae varied widely between the findings from our review and those of others’, we believe that the variability may in part be explained by methodological differences across reviews. For example, the other reviews synthesized both laboratory-confirmed and clinically-diagnosed populations, while we analyzed these populations separately in order to minimize bias in capturing symptoms that may be due to other causes. We wanted to identify all possible symptoms that may be associated with post COVID-19 condition, therefore we did not restrict inclusion of studies in our review, nor our syntheses of the findings, to only select symptoms, whereas other reviews did (21,24,26). Other reviews also excluded studies with less than 100 participants (23, 25) while our review included relevant studies with a sample size of 50 or more; thereby increasing our likelihood of capturing additional relevant studies compared to others. Some reviews included studies with follow-up periods occurring between 2 and 3 weeks post-infection (19-21,25). We excluded such studies in our review in order to minimize the likelihood of capturing symptoms from acute COVID-19 infection. We also thought it was important to report prevalence separately for the short-term and long-term after COVID-19 infection, and to gain insight into any potential differences in what may be observed over time, whereas most reviews combined results from follow-up periods spanning both short and long-terms (13,20–22,24) or only reported on the long-term (23, 26). Similar to the other reviews, we critically assessed studies for risk of bias. However, distinct from other reviews, we used the GRADE approach to provide certainty of the evidence for select outcomes. This information provides clinicians and policy-makers with an understanding of how confident we are in the findings given the limitations of the available data.

Some limitations should be considered including the possible omission of relevant studies. We adapted and updated our search based on the evidence review conducted by NICE in October 2020; the only studies that were eligible for inclusion in our review that were published prior to October 2020 were studies that were included in the NICE review, or French–language studies that were excluded from that review (12). To minimize the potential for missing relevant studies we supplemented with grey-literature searches. Another limitation is the inclusion of only English and French articles that may have introduced a language bias, but this was likely minimal (39). Finally, although we had consulted with GRADE experts on our modified process for assessing evidence on prevalence using the GRADE approach, this process has yet to be validated.

### Evidence gaps and Research Priorities

There were limited data (i.e. estimates based on a single study) or no studies identified for many outcomes in this systematic review. The majority of studies in this review included adults only or individuals who were hospitalized or treated for moderate-to-severe COVID-19; therefore, the prevalence of long-term effects in children (particularly very young children between 0-5 years of age), and in individuals who were asymptomatic or who presented with mild COVID-19 symptoms in the acute stage may not be sufficiently represented in our results. Additionally, investigating how post COVID-19 condition may differ across specific populations (e.g., sex/gender, race, age, underlying conditions) will be important to inform equity considerations for future health program and policy decisions.

Just over half of our included studies (n=43) reported on long-term effects beyond 12 weeks post-diagnosis (laboratory-confirmed or clinically-diagnosed). However, only a few (n= 6) of these included mean follow-up periods beyond 6 months from initial COVID-19 diagnosis. Longer-term follow up is needed to help inform how post COVID-19 condition changes or resolves over time. Many of the included studies had small sample sizes (<200 participants) or were at moderate or high risk of bias due to the selection of participants and use of non-validated outcome measures. The Post-COVID Core Outcome Set (PC-COS) group, along with the World Health Organization, are in the process of creating a core set of outcomes for use in research studies and clinical care for individuals with post COVID-19 condition (40). The use of a standardized approach, such as the one being developed by the PC-COS group for measuring symptoms and other outcomes in individuals with post COVID-19 condition, would help reduce heterogeneity and bias and subsequently increase confidence in the research findings. Given the lack of contemporaneous control groups, it was not possible to determine whether symptoms were due exclusively to COVID-19. Other possible contributing factors could include the presence of pre-existing symptoms or conditions prior to COVID-19 infection, effects of treatment received or effects of being hospitalized or admitted to the ICU, and effects due to the pandemic itself (e.g., barriers to seeking treatment, psychosocial impacts). However, the extensive list of symptoms reported in this review provide a good starting point for further detailed investigations as to which ones are more closely associated with post-infection sequelae versus those related to other factors.

### Health policy implications

Understanding the burden and characteristics of post COVID-19 condition is important in the planning and development of mitigation strategies to support those in need of rehabilitation, medical care and other community resources for recovery after COVID-19 infection. This evidence is expected to support national and international health organizations who are in the process of planning for and developing supportive measures for patients with post COVID-19 condition. Such efforts include developing clinical practice and public health guidelines, innovative patient care pathways, education materials for patients and healthcare professionals, and creating appropriate services and social constructs to support COVID-19 survivors for a full recovery (41). Understanding the burden of post COVID-19 condition will also help inform broader public health measures to mitigate COVID-19 transmission.

## Conclusion

A substantial proportion of individuals reported a variety of symptoms and sequelae more than four weeks after COVID-19 diagnosis. These physical and mental health symptoms have led to difficulties in conducting usual activities and resulted in diminished quality of life among COVID-19 survivors. This review provides a snapshot of symptoms presenting in COVID-19 survivors in the months after diagnosis. The data indicate that many are experiencing post COVID-19 condition, the range and impact of which are broad and will require a multidisciplinary approach to develop appropriate diagnostics, clinical practice and public health guidelines, and patient care pathways. Research on post COVID-19 condition is rapidly being produced and work is on-going to gather evidence which will lead to better and more refined estimates and understanding of the burden of post COVID-19 condition, the social and economic impacts, and resources needed to support a large number of survivors.

### Ethics statements

Ethics approval was not required to conduct this systematic review of previously published literature.

## Data Availability

The data that support the findings of this study are available from the corresponding author, FRD, upon reasonable request.

## Acknowledgements

The authors would like to thank the following individuals: Dr. Adrienne Stevens for providing methodological support and conducting risk of bias assessments; Dr. Adrienne Stevens, Dr. Maicon Falavigna and Dr. Zachary Munn for their guidance in the application of risk of bias using the Joanna Briggs Institute checklist for prevalence studies and modified GRADE for prevalence; Nana Amankwah, Judy Niles and Janet-Leigh Potvin from the Public Health Agency of Canada (PHAC) for helping with screening and data extraction; Gareth Leung from PHAC for helping with screening and risk of bias and GRADE assessments; Dr. Muhammad Mullah from PHAC for providing guidance on the meta-analyses; Linda Gamble from the Health Canada Library for conducting the literature searches.

AM Cheung is partially supported by a Tier 1 Canada Research Chair in Musculoskeletal and Postmenopausal Health as well as the KY and Betty Ho Chair in Integrative Medicine at the University of Toronto.

## Author contributions

<u>Francesca Reyes Domingo</u> – conception & design/methods, screening and data extraction, analysis and interpretation of the data, critical appraisal (risk of bias & GRADE), drafting and revision of the manuscript, final approval of the article

<u>Lisa A Waddell</u> – conception & design/methods, interpretation of the data, drafting and revision of the manuscript, final approval of the article

<u>Angela M Cheung</u> – conception & design/methods, interpretation of the data, revision of the manuscript, final approval of the article

<u>Curtis L Cooper</u> – conception & design/methods, interpretation of the data, revision of the manuscript, final approval of the article

<u>Veronica J Belcourt</u> - design/methods, interpretation of the data, critical appraisal (risk of bias & GRADE), revision of the manuscript, final approval of the article

<u>Alexandra M E Zuckermann</u> - design/methods, screening and data extraction, interpretation of the data, critical appraisal (risk of bias), drafting and revision of the manuscript, final approval of the article

<u>Tricia Corrin</u> – design/methods, screening and data extraction, interpretation of the data, drafting and revision of the manuscript, final approval of the article

<u>Rukshanda Ahmad</u> - design/methods, interpretation of the data, critical appraisal (risk of bias), drafting and revision of the manuscript, final approval of the article

<u>Laura Boland</u> – design/methods, screening, interpretation of the data, critical appraisal (risk of bias & GRADE), revision of the manuscript, final approval of the article

<u>Claudie Laprise</u> – design/methods, screening, interpretation of the data, critical appraisal (risk of bias), revision of the manuscript, final approval of the article

<u>Leanne Idzerda</u> – screening and data extraction, interpretation of the data, critical appraisal (risk of bias), revision of the manuscript, final approval of the article

<u>Anam Khan</u> - screening and data extraction, interpretation of the data, revision of the manuscript, final approval of the article

<u>Kate Morissette</u> - critical appraisal (risk of bias & GRADE), drafting and revision of the manuscript, final approval of the article

<u>Alejandra Jaramillo Garcia</u> – conception & design/methods, interpretation of the data, critical appraisal (risk of bias & GRADE), drafting and revision of the manuscript, final approval of the article

