## Supplementary File 1 for "Prevalence of long-term effects in individuals diagnosed with COVID-19: an updated living systematic review"

#### Supplementary File 1 – Detailed methodology

**Authors:**

Francesca Reyes Domingo MHSc[1], Lisa A Waddell MSc PhD[2], Angela M. Cheung MD FRCPC[3], Curtis L. Cooper MD FRCPC[4ah], Veronica J. Belcourt MSc PhD[1], Alexandra M. E. Zuckermann PhD[1], Tricia Corrin MPH[2], Rukshanda Ahmad MBBS MHA[5], Laura Boland SLP-C PhD[1], Claudie Laprise MSc PhD[6,7], Leanne Idzerda MSc PhD[1], Anam Khan MPH[5], Kate Morissette MSc[1], Alejandra Jaramillo Garcia MSc[1]

**Affiliations:**

[1] Evidence Synthesis and Knowledge Translation Unit, Applied Research Division, Public Health Agency of Canada

[2] Public Health Risk Sciences Division, National Microbiology Laboratory, Public Health Agency of Canada

[3] Department of Medicine, University Health Network and University of Toronto, Canada

[4] Department of Medicine, University of Ottawa; Ottawa Hospital Research Institute

[5] Health Professionals Guidance Unit, Centre for Food-borne, Environmental and Zoonotic Infectious Diseases, Public Health Agency of Canada

[6] Public Health Capacity and Knowledge Management Unit, Quebec Regional Office, Public Health Agency of Canada

[7] Division of Oral Health and Society, Faculty of Dentistry, McGill University

### 1.0 Objective & Research Question

The primary objective of this living systematic review is to systematically identify, review, appraise, and synthesize the existing evidence on the long-term impacts in individuals diagnosed with COVID-19.

**Research Question:** Among those diagnosed with COVID-19, what is the prevalence of long-term effects, including the frequency of symptoms, sequelae and difficulties being able to carry out usual activities, four or more weeks after diagnosis?

### 2.0 Methods

Guidance produced by the Cochrane Methods Group was followed to conduct the systematic review and adhere to the "Preferred Reporting Items for Systematic Reviews and Meta-Analyses" (PRISMA) guidelines (1,2). To minimize potential bias and enhance reliability of the findings, the literature search strategy, review questions, study selection criteria, data collection, risk of bias and certainty of evidence assessment methods, synthesis plan, and approaches for investigating causes of heterogeneity, have been determined *a priori* and included in this protocol. A shortened version of this protocol is registered in PROSPERO: CRD42021231476 (3).

#### 2.1 Formative search

Existing systematic reviews covering the proposed topic were identified. We compared our inclusion criteria for population, outcomes, setting, study types and geography with those from the identified reviews. We determined the existing review by the National Institute for Health and Care Excellence (NICE) on the prevalence of long-term effects of COVID-19 (NG188) (4) could be adapted and updated to meet our evidence needs.

#### 2.2 Review team and stakeholder engagement

A multidisciplinary team with expertise in knowledge synthesis, epidemiology, chronic and infectious diseases, public health, information science and subject matter expertise conducted the review. Key stakeholder consultations, including researchers, front-line clinicians treating patients with long-term effects from COVID-19, and policy-makers, were conducted throughout the review to set and refine the review question, eligibility criteria, and outcomes of interest. All review authors, subject matter experts, and stakeholders have reported any potential conflict of interest prior to agreeing to be part of the review process.

#### 2.3 Search strategy

The search strategy developed by NICE for their evidence review on the prevalence of long-term effects of COVID-19 served as the starting point for this systematic review (4). A NICE librarian had previously peer-reviewed the original search strategy. Our librarian conducted an additional peer-review and concluded the search strategy met our needs.

The NICE search strategy was adapted to capture all relevant literature published after October 22-28, 2020 and included French language articles. Keywords around the concept of human/animal limits were added to capture any unindexed articles. Searches were conducted in the following four bibliographic databases from October 22, 2020 to April 14, 2021: Embase, Medline, PsychInfo, and Cochrane Central. The full search strategy is included in Appendix 1.

A complementary search for additional primary research using keywords from the search strategy was conducted in the following websites: COVID-END (April 18-19, 2021), Centre for Evidence-Based Medicine – Oxford COVID-19 Evidence Service (April 18-19, 2021), CADTH COVID-19 Pandemic Database (April 18-19, 2021), National COVID-19 Clinical Evidence Task Force (April 18-19, 2021), Cochrane COVID Review Bank (April 20, 2021), Cochrane COVID-19 Study Register (October 28, 2020 – April 12, 2021), and BMJ Best Practice COVID-19 – Complications – Post-COVID-19 Syndrome (long COVID) (April 21, 2021). Details on the grey literature search can be found in Appendix 1.

The results of the updated search and all studies in the NICE review and all French articles they had excluded were eligible for inclusion in this review and proceeded to relevance screening.

Results of search strategy can be found in Appendix 6.

After the April 2021 search, monthly literature search updates will be conducted and references routinely screened; however, a review of the new literature and any updates to the synthesis report will be conducted on a quarterly basis or as resources allow.

#### 2.4 Eligibility criteria

All citations identified by the search strategy will be assessed for inclusion in the systematic review.

**Overall inclusion criteria**

Studies will be selected for inclusion in the systematic review based on the following set of predefined selection criteria.

The following inclusion criteria will be applied:

1) Publication date: Studies published after January 1, 2020
2) Countries: All
3) Language: English or French
4) Document type: All primary published research (e.g. journal articles). Pre-print, non peer-reviewed articles, reviews, editorials, and opinion papers were excluded
5) Population: Individuals who were diagnosed with COVID-19 (either lab-confirmed or clinically diagnosed by a physician or healthcare practitioner) and were assessed for long-term effects of COVID-19 at four or more weeks after diagnosis. Populations that were recruited because they reported long-term effects of COVID-19 were excluded
6) Sample size: The study must have included 50 or more participants diagnosed with COVID-19
7) Study design: All primary research study designs were considered eligible, including observational studies, experimental studies, and case series

**PICO criteria details**

Participants/population

Population must meet all of the following inclusion criteria:

1) Individuals who were diagnosed with COVID-19 (laboratory-confirmed or clinically-diagnosed by a physician/health practitioner)
2) Were assessed for long-term effects of COVID-19 at 4 or more weeks after COVID-19 diagnosis

Other exclusion criteria based on study population:

1) Study participants were recruited specifically because they reported long-term effects (including symptoms, sequelae and difficulties carrying out usual activities) 4 or more weeks after COVID-19 diagnosis

Intervention(s), exposure(s)

COVID-19 (SARS-CoV-2) diagnosis

Comparator(s)/control

Not applicable (this is a review on prevalence)

Main outcome(s)

1) The prevalence of long-term effects at 4 weeks or more after COVID-19 diagnosis (lab-confirmed only)
2) The prevalence of key symptoms or clusters of symptoms or sequelae 4 or more weeks after COVID-19 diagnosis (lab-confirmed only)
3) The proportion of individuals reporting difficulties in being able to carry out usual activities (due to both physical and mental health symptoms) 4 or more weeks after COVID-19 diagnosis (lab-confirmed only)

**Key symptoms/sequelae:** will include (but are not limited to): fatigue, shortness of breath/breathlessness, neurocognitive impairment ('brain fog'; memory or loss of concentration issues), pain (in the joints, chest or muscles), organ damage, dizziness, tachycardia, chest tightness or heaviness, olfactory and taste impairments and sleeping disturbances.

**Measures of effect:** prevalence

Additional outcome(s)

1) The prevalence of long-term effects 4 weeks or more after COVID-19 diagnosis (clinically-diagnosed only)
2) The prevalence of symptoms or clusters of symptoms or sequelae 4 or more weeks after COVID-19 diagnosis (clinically-diagnosed only)

3) The proportion of individuals reporting difficulties in being able to carry out usual activities (due to both physical and mental health symptoms) 4 or more weeks after COVID-19 diagnosis (clinically-diagnosed only)

**Measures of effect:** prevalence

#### 2.5 Study selection

The librarian removed duplicates from all potentially relevant articles identified through the searches. The references were then imported to the web-based systematic review program DistillerSR (5) for a second round of duplicate removal. Results of the study selection process were presented within a PRISMA diagram (2).

A title and abstract screening form and a full text screening form were developed *a priori* to incorporate the inclusion and exclusion criteria of the review (Appendix 2). Prior to implementation, the title and abstract form was pre-tested by all reviewers on a random sample of 50 references and the full text form was pretested on 10. The forms were adjusted until there was consensus that it was performing well. Two reviewers independently used these forms to screen titles, abstracts, and full texts for relevance to the review topic and ensure all inclusion criteria were met. For the title and abstract screening, only one reviewer was required to indicate the reference was potentially relevant to move to full-text screening and two reviewers were required to exclude a citation. Conflicts at the full-text screening stage were resolved through consensus. Where required, articles were ordered through the Health Canada Library.

#### 2.6 Data extraction

Two data extraction forms were developed *a priori.* The first form was designed to extract key study characteristics such as study design, country, study recruitment date, and population and can be found in Appendix 3. One reviewer extracted the study characteristics and another verified the extracted information. The second form collected all outcome data and was pre-tested by all reviewers to ensure clarity, consistency, and that all the necessary information to address the research topic was extracted. This form is located in Appendix 3. The forms were adjusted until there was consensus that they were performing well. For each included study, one reviewer extracted the outcome data and a second reviewer verified the extraction. Any discrepancies found by the verifier were discussed and resolved by consensus.

#### 2.8 Data analysis

Descriptive statistics were computed in Microsoft Excel (Microsoft Corporation, Redmond, WA) and results synthesized by time since COVID-19 diagnosis (short-term or 4-12 weeks after, or long-term or 12 weeks or more after).

Where appropriate, meta-analyses were conducted using R statistical software version 4.0.4 (6), with package metaphor version 2.4-0 (7) and package meta version 4.18-0 (8) for outcomes with two or more studies contributing to the outcome. For studies with partially overlapping populations (n=2), data for the same outcomes were still included from both studies; however, the numerator and denominator from each study were halved to account for the overlap. Random effects meta-analysis of proportions were selected based on the assumption of true variation of long-term effects of COVID-19 across studies (9). Heterogeneity was quantified by calculation of Higgins' $I^2$ and $T^2$, an estimate of $\tau^2$, which represents the variance in prevalence across studies (10). Heterogeneity of effect estimates was categorized as 'low to moderate' if $I^2 \leq 60\%$.

To explore reasons for heterogeneity across studies and to inform GRADE assessments, sub-group analyses were considered for select key outcomes if heterogeneity is very high. Potential sub-groups were selected based on clinically meaningful reasons for heterogeneity and for which sub-group data was available.

#### 2.7 Risk-of-bias assessment

A modified version of the Joanna Briggs Institute (JBI) critical appraisal tool for prevalence studies (11) was used to assess for risk of bias for each study (questions 1, 2 and 9) and for each outcome reported by the study (questions 6, 7 and 8). To avoid duplication with the criteria used to assess the certainty of the evidence, questions 3-5 were omitted. The questions were divided into three domains (participants, outcome measures and statistics) and studies were rated as low risk of bias if sufficient criteria were met for all three domains. The details of the schema that were followed and the rationale for decisions can be found in Appendix 4. After piloting the tool by all reviewers, one reviewer assessed the risk of bias for each study- outcome combination and another verified the assessment with disagreements resolved through discussion or consultation with a third reviewer.

#### 2.9 Modified GRADE Assessment

The Grading of Recommendations Assessment, Development and Evaluation (GRADE) methodology was applied to assess the quality of the body of evidence for each outcome (12). In the absence of a formal framework for prevalence and GRADE, we used the GRADE framework for assessment of incidence estimates in the context of prognostic studies (13) as a basis and similar to what others have done (e.g., Righy et al., 2019 (14)), with specific adaptations described herein. After piloting the tool by all reviewers, one reviewer assessed the quality of the body of evidence for each outcome and another verified the assessment with disagreements resolved through discussion or consultation with a third reviewer.

In the context of prevalence, observational studies may provide robust estimates due to broad eligibility criteria and enrollment of representative populations (13). Thus, the quality of the evidence from observational studies were initially assigned as "high" for all outcomes. The quality of the evidence was then downgraded (to "moderate", "low", or "very low") if there were serious or very serious concerns over any of the following five domains that reduce certainty in the prevalence estimates: risk of bias, inconsistency, indirectness, imprecision, or publication bias. In instances where there were minor concerns with one of these domains, a decision was made to downgrade by half a point instead of a full point. Half-points were then combined across domains to yield a total score (e.g., downgrading by 1.5 points for risk of bias and 0.5 points for indirectness for a total of 2 points from "high" to "low"). In the event that the final rating included a half point (e.g., 1.5 points), we conservatively rounded up (i.e., in this case to downgrade by 2). Due to the nature of prevalence outcomes and design of the application of GRADE criteria to these outcomes, no criteria for upgrading were considered applicable. The specific decision rules that were applied for judgments in relation to each of these criteria are described in Appendix 5.

**Risk of Bias**

To assess the risk of bias across studies contributing to an outcome, we considered whether any studies were at moderate or high risk of bias, and whether studies at high risk of bias exerted disproportionate influence on the range of prevalence estimates (i.e., were responsible for either the highest or lowest prevalence estimate in the range of estimates in the case of narrative syntheses) or summary prevalence estimate (i.e., comprised 50% or more of studies contributing to an outcome in the case of meta-analyses).

Specifically, for outcomes in which the findings were summarized narratively (i.e., as median and range of prevalence estimates), we downgraded for risk of bias across studies contributing to each outcome as follows:

- No serious risk of bias:
  - Do not downgrade: all studies contributing to an outcome were considered to be at low risk of bias.
- Serious risk of bias:
  - Downgrade by 0.5 points: all studies contributing to an outcome were considered to be at either low or moderate risk of bias.
  - Downgrade by 1 point: at least one study contributing to an outcome was at high risk of bias, but a study at high risk of bias was not responsible for either the highest or lowest prevalence estimate in the range of estimates for the outcome (i.e., any studies at high risk of bias were within the middle of the range of estimates).
- Very serious risk of bias:
  - Downgrade by 1.5 points: at least one study was at high risk of bias, and a study at high risk of bias was responsible for either the highest or lowest prevalence estimate (or both) in the range of estimates for the outcome.

For outcomes in which findings were summarized by random effects meta-analysis, studies will be weighted similarly in the pooled estimate (Higgins et al., 2021), and thus it will not be relevant whether a study at high risk of bias is responsible for either the lowest or highest prevalence estimate or falls somewhere in the middle of the range of estimates. Therefore, for outcomes in which findings were summarized by meta-analysis, we downgraded for risk of bias across studies contributing to each outcome as follows:

- No serious risk of bias:
  - Do not downgrade: all studies contributing to an outcome were considered to be at low risk of bias.
- Serious risk of bias:
  - Downgrade by 0.5 points: all studies contributing to an outcome were considered to be at either low or moderate risk of bias.

    - Downgrade by 1 point: at least one study contributing to an outcome (but <50% of total studies) was at high risk of bias.
- Very serious risk of bias:
    - Downgrade by 1.5 points: 50% or more of studies contributing to an outcome were at high risk of bias.

**Inconsistency**

Studies with small sample sizes may contribute to inconsistency since they are less precise (i.e. their estimates may be further from the true proportion compared to studies with higher precision) and it is also more difficult to ensure a small sample represents the target population (13). Therefore, the *a priori* decision was made to exclude studies with fewer than 50 participants. Substantial heterogeneity is still expected when synthesizing studies that include studies drawn from different target populations. Therefore, the *a priori* decision was made to not consider measures of statistical heterogeneity (i.e., $I^2$) in assessment of inconsistency for outcomes in which meta-analyses were conducted. Rather, for all outcomes, inconsistency will be evaluated based on variability between prevalence estimates, and whether variability could be explained by *a priori*-hypothesized sources of heterogeneity (i.e., based on subgroup analyses).

If there was only a single study contributing to an outcome, we downgraded by 1 point for serious inconsistency. This is because, given a single study, there is no opportunity to observe heterogeneity across studies and failing to rate down would artificially inflate the certainty in the evidence. This approach has been taken in previous systematic reviews (15,16) which were conducted to inform national-level public health guidelines (17).

For outcomes where there was more than one study contributing, subgroup analyses were conducted to explore heterogeneity where deemed necessary by subject matter experts (SMEs). SMEs were presented with the range of prevalence estimates for each outcome and asked to judge whether the range of estimates for an outcome was expected or acceptable. If the range of estimates is considered expected or acceptable, we did not rate down for inconsistency. If the range of estimates was considered large, subgroup analyses were conducted to explore potential sources of heterogeneity. If heterogeneity could be partially (but not completely) explained by subgroup analyses, we downgrade by 0.5 points; if it could not be explained by subgroup analyses, we downgraded by 1 point.

In summary, downgrades were applied for inconsistency across studies contributing to each outcome as follows:

- No serious inconsistency:
    - Do not downgrade:
        - In the judgment of SMEs, heterogeneity is considered expected or acceptable
        - Heterogeneity can be explained by subgroup analyses
- Serious inconsistency:
    - Downgrade by 0.5 points:
        - Heterogeneity can be partially (but not completely) explained by subgroup analyses
    - Downgrade by 1 point:
        - Only one study contributed to an outcome

- Heterogeneity can not be explained by subgroup analyses

**Indirectness**

To evaluate indirectness, we considered whether the study populations corresponded to the target population of interest for our review (i.e., all individuals who were diagnosed with lab-confirmed COVID-19). We did not consider directness of outcomes (e.g., whether outcomes were measured objectively/directly or indirectly) as this was already considered in our evaluation of risk of bias, and because all outcomes were considered to be directly important to patients. If the study or studies contributing to an outcome include only a subset of our target population, we will downgrade by 0.5 points.

**Imprecision**

To evaluate imprecision, we considered two factors: 1) whether a clinically-important threshold for decision-making (e.g., for decisions around clinical care and management of healthcare resources) would differ if the upper versus lower boundary of the confidence interval, or the highest versus lowest point estimate from individual studies, represented the truth; and 2) whether the optimal information size was met. For the first factor, SMEs were presented with the data and asked to provide their judgments.

If in the judgment of SME's a clinically-important threshold for decision-making was crossed, we would downgrade by 1 point (regardless of whether the optimal information size (OIS) was met).

If in the judgment of SME's a clinically-important threshold for decision-making was not crossed, we then considered the OIS. The OIS was calculated using the formula provided in the JBI tool, using 95% Z score of 1.96 and a precision of 5% [d=0.05]. Using the lowest and highest prevalence estimates (95% CI's for the outcome), we were able to calculate and see if OIS is met. If the OIS was met, we did not downgrade; however, if the OIS was not met, we downgraded by 0.5 points.

**Publication Bias**

Due to the large volume of COVID-related manuscripts being published and the large interest in publishing any findings related to long-COVID, an *a priori* decision was made not to downgrade for risk of publication bias.

### Appendix 1 – Search Strategy

#### Database Search

Database(s): **EBM Reviews - Cochrane Central Register of Controlled Trials** March 2021
Search Strategy:

| # | Searches |
|---|---|
| 1 | (longcovid* or long covid* or longcoronavirus* or longcorona* virus* or long coronavirus* or long corona* virus* or longcoronovirus* or longcorono* virus* or long coronovirus* or long corono* virus* or longcoronavirinae* or longcorona* virinae* or long coronavirinae* or long corona* virinae* or longCov or long Cov or longsars* or long sars* or "long severe acute respiratory syndrome*" or longncov* or long ncov* or longhcov* or long hcov*).ti,ab,kw. |
| 2 | ((long* or endur* or legacy* or slow* or gradual* or protract* or lengthy* or chronic* or persist* or relaps* or remit* or remission* or residual* or delay* or prolong* or extend* or linger* or permanent* or fluctuat* or sequela* or multisystem* or "multi system*" or nonrecover* or "non recover*" or subacute* or "sub acute*" or lasting* or continuous* or continual* or continuing* or postacute* or "post acute*" or postdischarg* or "post discharg*" or postinfect* or "post infect*" or postviral* or "post viral*" or postvirus* or "post virus*") adj1 (covid* or coronavirus* or corona* virus* or coronovirus* or corono* virus* or coronavirinae* or corona* virinae* or Cov or "2019-nCoV*" or 2019nCoV* or "19-nCoV*" or 19nCoV* or nCoV2019* or "nCoV-2019*" or nCoV19* or "nCoV-19*" or "HCoV-19*" or HCoV19* or "HCoV-2019*" or HCoV2019* or "2019 novel*" or Ncov* or "n-cov" or "SARS-CoV-2*" or "SARSCoV-2*" or "SARSCoV2*" or "SARS-CoV2*" or SARSCov19* or "SARS-Cov19*" or "SARSCov-19*" or "SARS-Cov-19*" or SARSCov2019* or "SARSCov2019*" or "SARSCov-2019*" or "SARS-Cov-2019*" or SARS2* or "SARS-2*" or SARScoronavirus2* or "SARS-coronavirus-2*" or "SARScoronavirus 2*" or "SARS coronavirus2*" or SARScoronovirus2* or "SARS-coronovirus-2*" or "SARScoronovirus 2*" or "SARS coronovirus2*" or "severe acute respiratory syndrome*")).ti,ab. |
| 3 | (("long* term*" or longterm* or "long* haul*" or longhaul* or "long* tail*" or longtail* or longduration* or "long duration*" or longlast* or "long last*" or longstanding* or "long standing*" or "medium* term*" or mediumterm*) adj3 (covid* or coronavirus* or corona* virus* or coronovirus* or corono* virus* or coronavirinae* or corona* virinae* or Cov or "2019-nCoV*" or 2019nCoV* or "19-nCoV*" or 19nCoV* or nCoV2019* or "nCoV-2019*" or nCoV19* or "nCoV-19*" or "HCoV-19*" or HCoV19* or "HCoV-2019*" or HCoV2019* or "2019 novel*" or Ncov* or "n-cov" or "SARS-CoV-2*" or "SARSCoV-2*" or "SARSCoV2*" or "SARS-CoV2*" or SARSCov19* or "SARS-Cov19*" or "SARSCov-19*" or "SARS-Cov-19*" or SARSCov2019* or "SARS-Cov2019*" or "SARSCov-2019*" or "SARS-Cov-2019*" or SARS2* or "SARS-2*" or SARScoronavirus2* or "SARS164 37 coronavirus-2*" or "SARScoronavirus 2*" or "SARS coronavirus2*" or SARScoronovirus2* or "SARS-coronovirus-2*" or "SARScoronovirus 2*" or "SARS coronovirus2*" or "severe acute respiratory syndrome*")).ti,ab. |

| | |
|---|---|
| 4 | ((postcovid* or post covid* or postcoronavirus* or postcorona* virus* or post coronavirus* or post corona* virus* or postcoronovirus* or postcorono* virus* or post coronovirus* or post corono* virus* or postcoronavirinae* or postcorona* virinae* or post coronavirinae* or post corona* virinae* or postCov or post Cov or postsars* or post sars* or "post severe acute respiratory syndrome*" or postncov* or post ncov* or posthcov* or post hcov*) adj3 (syndrome* or disorder* or illness* or sickness* or disease* or condition* or symptom* or sign* or prognos* or followup* or "follow up*" or feature* or comorbid* or "co morbid*" or multimorbid* or "multi morbid*" or survivor* or survival* or risk* or care* or convalescen* or recuperat* or aftercare* or ambulatory* or outpatient* or "out patient*")).ti,ab. |
| 5 | ((ongoing* or long* or endur* or legacy* or slow* or gradual* or protract* or lengthy* or chronic* or persist* or relaps* or remit* or remission* or residual* or delay* or prolong* or extend* or linger* or permanent* or fluctuat* or multisystem* or "multi system*" or nonrecover* or "non recover*" or subacute* or "sub acute*" or lasting* or continuous* or continual* or continuing* or postacute* or "post acute*" or postdischarg* or "post discharg*" or postinfect* or "post infect*" or postviral* or "post viral*" or postvirus* or "post virus*" or "medium* term*" or mediumterm*) adj4 (sequela* or illness* or symptom* or sign* or prognos* or rehab* or convalescen* or recuperat* or followup* or "follow up*" or feature*) adj10 (covid* or coronavirus* or corona* virus* or coronovirus* or corono* virus* or coronavirinae* or corona* virinae* or Cov or "2019-nCoV*" or 2019nCoV* or "19- nCoV*" or 19nCoV* or nCoV2019* or "nCoV-2019*" or nCoV19* or "nCoV-19*" or "HCoV-19*" or HCoV19* or "HCoV-2019*" or HCoV2019* or "2019 novel*" or Ncov* or "n-cov" or "SARS-CoV-2*" or "SARSCoV-2*" or "SARSCoV2*" or "SARS-CoV2*" or SARSCov19* or "SARS-Cov19*" or "SARSCov-19*" or "SARS-Cov-19*" or SARSCov2019* or "SARS-Cov2019*" or "SARSCov-2019*" or "SARS-Cov-2019*" or SARS2* or "SARS-2*" or SARScoronavirus2* or "SARS-coronavirus-2*" or "SARScoronavirus 2*" or "SARS coronavirus2*" or SARScoronovirus2* or "SARScoronovirus-2*" or "SARScoronovirus 2*" or "SARS coronovirus2*" or "severe acute respiratory syndrome*")).ti,ab. |
| 6 | ((ongoing* or long* or endur* or legacy* or slow* or gradual* or protract* or lengthy* or chronic* or persist* or relaps* or remit* or remission* or residual* or delay* or prolong* or extend* or linger* or permanent* or fluctuat* or multisystem* or "multi system*" or subacute* or "sub acute*" or lasting* or continuous* or continual* or continuing* or post* or after* or follow* or "medium* term*" or mediumterm*) adj1 recover* adj10 (covid* or coronavirus* or corona* virus* or coronovirus* or corono* virus* or coronavirinae* or corona* virinae* or Cov or "2019-nCoV*" or 2019nCoV* or "19-nCoV*" or 19nCoV* or 128 38 nCoV2019* or "nCoV-2019*" or nCoV19* or "nCoV-19*" or "HCoV-19*" or HCoV19* or "HCoV-2019*" or HCoV2019* or "2019 novel*" or Ncov* or "n-cov" or "SARS-CoV-2*" or "SARSCoV-2*" or "SARSCoV2*" or "SARS-CoV2*" or SARSCov19* or "SARS-Cov19*" or "SARSCov-19*" or "SARS-Cov-19*" or SARSCov2019* or "SARS-Cov2019*" or "SARSCov-2019*" or "SARS-Cov-2019*" or SARS2* or "SARS-2*" or SARScoronavirus2* or "SARS-coronavirus-2*" or "SARScoronavirus 2*" or "SARS coronavirus2*" or SARScoronovirus2* or "SARS-coronovirus-2*" or "SARScoronovirus 2*" or "SARS coronovirus2*" or "severe acute respiratory syndrome*")).ti,ab. |

| | |
|---|---|
| 7 | or/1-6 |
| 8 | exp coronavirus/ |
| 9 | exp Coronavirus infections/ |
| 10 | (covid* or coronavirus* or corona* virus* or coronovirus* or corono* virus* or coronavirinae* or corona* virinae* or Cov or "2019-nCoV*" or 2019nCoV* or "19-nCoV*" or 19nCoV* or nCoV2019* or "nCoV-2019*" or nCoV19* or "nCoV-19*" or "HCoV-19*" or HCoV19* or "HCoV-2019*" or HCoV2019* or "2019 novel*" or Ncov* or "n-cov" or "SARS-CoV-2*" or "SARSCoV-2*" or "SARSCoV2*" or "SARS-CoV2*" or SARSCov19* or "SARS-Cov19*" or "SARSCov-19*" or "SARS-Cov-19*" or SARSCov2019* or "SARSCov2019*" or "SARSCov-2019*" or "SARS-Cov-2019*" or SARS2* or "SARS-2*" or SARScoronavirus2* or "SARS-coronavirus-2*" or "SARScoronavirus 2*" or "SARS coronavirus2*" or SARScoronovirus2* or "SARS-coronovirus-2*" or "SARScoronovirus 2*" or "SARS coronovirus2*" or "severe acute respiratory syndrome*").ti. |
| 11 | or/8-10 |
| 12 | "Recovery of Function"/ |
| 13 | Aftercare/ |
| 14 | rehabilitation/ |
| 15 | Activities of Daily Living/ |
| 16 | Convalescence/ |
| 17 | Long Term Adverse Effects/ |
| 18 | Physical Functional Performance/ |
| 19 | or/12-18 |
| 20 | 11 and 19 |
| 21 | ("long* haul*" or longhaul* or "long* tail*" or longtail* or longduration* or "long duration*" or longlast* or "long last*" or longstanding* or "long standing*").ti,ab. and (8 or 9) |
| 22 | ((recover* or nonrecover*) adj3 function* adj10 (covid* or coronavirus* or corona* virus* or coronovirus* or corono* virus* or coronavirinae* or corona* virinae* or Cov or "2019- 15 39 nCoV*" or 2019nCoV* or "19-nCoV*" or 19nCoV* or nCoV2019* or "nCoV-2019*" or nCoV19* or "nCoV-19*" or "HCoV-19*" or HCoV19* or "HCoV-2019*" or HCoV2019* or "2019 novel*" or Ncov* or "n-cov" or "SARS-CoV-2*" or "SARSCoV-2*" or "SARSCoV2*" or "SARS-CoV2*" or SARSCov19* or "SARS-Cov19*" or "SARSCov-19*" or "SARS-Cov19*" or SARSCov2019* or "SARS-Cov2019*" or "SARSCov-2019*" or "SARS-Cov2019*" or SARS2* or "SARS-2*" or SARScoronavirus2* or "SARS-coronavirus-2*" or "SARScoronavirus 2*" or "SARS coronavirus2*" or SARScoronovirus2* or "SARScoronovirus-2*" or "SARScoronovirus 2*" or "SARS coronovirus2*" or "severe acute respiratory syndrome*")).ti,ab. |

| | |
|---|---|
| 23 | ((postacute* or "post acute*" or postdischarg* or "post discharg*" or postinfect* or "post infect*" or postviral* or "post viral*" or postvirus* or "post virus*" or subacute* or "sub acute*") adj3 (care* or convalescen* or recuperat* or aftercare* or ambulatory* or outpatient* or "out patient*" or survivor* or survival*) adj10 (covid* or coronavirus* or corona* virus* or coronovirus* or corono* virus* or coronavirinae* or corona* virinae* or Cov or "2019-nCoV*" or 2019nCoV* or "19-nCoV*" or 19nCoV* or nCoV2019* or "nCoV2019*" or nCoV19* or "nCoV-19*" or "HCoV-19*" or HCoV19* or "HCoV-2019*" or HCoV2019* or "2019 novel*" or Ncov* or "n-cov" or "SARS-CoV-2*" or "SARSCoV-2*" or "SARSCoV2*" or "SARS-CoV2*" or SARSCov19* or "SARS-Cov19*" or "SARSCov-19*" or "SARS-Cov-19*" or SARSCov2019* or "SARS-Cov2019*" or "SARSCov-2019*" or "SARS-Cov-2019*" or SARS2* or "SARS-2*" or SARScoronavirus2* or "SARScoronavirus-2*" or "SARScoronavirus 2*" or "SARS coronavirus2*" or SARScoronovirus2* or "SARS-coronovirus-2*" or "SARScoronovirus 2*" or "SARS coronovirus2*" or "severe acute respiratory syndrome*")).ti,ab. |
| 24 | ((convalescen* or recuperat* or after* or followup* or "follow up*" or rehab*) adj1 (therap* or care*) adj10 (covid* or coronavirus* or corona* virus* or coronovirus* or corono* virus* or coronavirinae* or corona* virinae* or Cov or "2019-nCoV*" or 2019nCoV* or "19- nCoV*" or 19nCoV* or nCoV2019* or "nCoV-2019*" or nCoV19* or "nCoV-19*" or "HCoV-19*" or HCoV19* or "HCoV-2019*" or HCoV2019* or "2019 novel*" or Ncov* or "n-cov" or "SARS-CoV-2*" or "SARSCoV-2*" or "SARSCoV2*" or "SARS-CoV2*" or SARSCov19* or "SARS-Cov19*" or "SARSCov-19*" or "SARS-Cov-19*" or SARSCov2019* or "SARS-Cov2019*" or "SARSCov-2019*" or "SARS-Cov-2019*" or SARS2* or "SARS-2*" or SARScoronavirus2* or "SARS-coronavirus-2*" or "SARScoronavirus 2*" or "SARS coronavirus2*" or SARScoronovirus2* or "SARScoronovirus-2*" or "SARScoronovirus 2*" or "SARS coronovirus2*" or "severe acute respiratory syndrome*")).ti,ab. |
| 25 | ((ongoing* or endur* or long* or legacy* or slow* or gradual* or protract* or lengthy* or chronic* or persist* or relaps* or remit* or remission* or residual* or delay* or prolong* or extend* or linger* or permanent* or fluctuat* or multisystem* or "multi system*" or 58 40 nonrecover* or "non recover*" or subacute* or "sub acute*" or lasting* or continuous* or continual* or continuing* or postacute* or "post acute*" or postdischarg* or "post discharg*" or postinfect* or "post infect*" or postviral* or "post viral*" or postvirus* or "post virus*" or "medium* term*" or mediumterm* or adverse* or dangerous* or harmful* or indirect* or injurious* or secondary* or side effect* or undesirable* or sequela* or complication* or consequence* or effect* or event* or impact* or outcome* or reaction* or complexit* or aftercare* or impair* or problem* or issue* or rehab* or function* or perform*) adj10 ((daily* or everyday* or day* or normal* or usual*) adj1 (activit* or living* or life* or lives* or job* or work* or employ* or occupation* or hobby* or hobbies* or leisure*)) adj10 (covid* or coronavirus* or corona* virus* or coronovirus* or corono* virus* or coronavirinae* or corona* virinae* or Cov or "2019-nCoV*" or 2019nCoV* or "19- nCoV*" or 19nCoV* or nCoV2019* or "nCoV-2019*" or nCoV19* or "nCoV-19*" or "HCoV-19*" or HCoV19* or "HCoV-2019*" or HCoV2019* or "2019 novel*" or Ncov* or "n-cov" or "SARS-CoV-2*" or "SARSCoV-2*" or "SARSCoV2*" or "SARS-CoV2*" or SARSCov19* or "SARS-Cov19*" or "SARSCov-19*" or "SARS-Cov-19*" or SARSCov2019* or "SARS-Cov2019*" or "SARSCov-2019*" or "SARS-Cov-2019*" or |

| | |
|---|---|
| | SARS2* or "SARS-2*" or SARScoronavirus2* or "SARS-coronavirus-2*" or "SARScoronavirus 2*" or "SARS coronavirus2*" or SARScoronovirus2* or "SARScoronovirus-2*" or "SARScoronovirus 2*" or "SARS coronovirus2*" or "severe acute respiratory syndrome*")).ti,ab. |
| 26 | ((ongoing* or endur* or long* or legacy* or slow* or gradual* or protract* or lengthy* or chronic* or persist* or relaps* or remit* or remission* or residual* or delay* or prolong* or extend* or linger* or permanent* or fluctuat* or multisystem* or "multi system*" or nonrecover* or "non recover*" or subacute* or "sub acute*" or lasting* or continuous* or continual* or continuing* or postacute* or "post acute*" or postdischarg* or "post discharg*" or postinfect* or "post infect*" or postviral* or "post viral*" or postvirus* or "post virus*" or "medium* term*" or mediumterm*) adj3 ((health* or adverse* or dangerous* or harmful* or indirect* or injurious* or secondary* or side* or undesirable* or negative* or damaging* or detriment* or abnormal*) adj1 (sequela* or complication* or consequence* or effect* or event* or impact* or outcome* or reaction* or complexit* or aftercare* or impair* or problem* or issue* or symptom* or disorder*)) adj10 (covid* or coronavirus* or corona* virus* or coronovirus* or corono* virus* or coronavirinae* or corona* virinae* or Cov or "2019-nCoV*" or 2019nCoV* or "19-nCoV*" or 19nCoV* or nCoV2019* or "nCoV2019*" or nCoV19* or "nCoV-19*" or "HCoV-19*" or HCoV19* or "HCoV-2019*" or HCoV2019* or "2019 novel*" or Ncov* or "n-cov" or "SARS-CoV-2*" or "SARSCoV-2*" or "SARSCoV2*" or "SARS-CoV2*" or SARSCov19* or "SARS-Cov19*" or "SARSCov-19*" or "SARS-Cov-19*" or SARSCov2019* or "SARS-Cov2019*" or "SARSCov-2019*" or "SARS-Cov-2019*" or SARS2* or "SARS-2*" or SARScoronavirus2* or "SARS40 41 coronavirus-2*" or "SARScoronavirus 2*" or "SARS coronavirus2*" or SARScoronovirus2* or "SARS-coronovirus-2*" or "SARScoronovirus 2*" or "SARS coronovirus2*" or "severe acute respiratory syndrome*")).ti,ab. |
| 27 | ((ongoing* or endur* or long* or legacy* or slow* or gradual* or protract* or lengthy* or chronic* or persist* or relaps* or remit* or remission* or residual* or delay* or prolong* or extend* or linger* or permanent* or fluctuat* or multisystem* or "multi system*" or nonrecover* or "non recover*" or subacute* or "sub acute*" or lasting* or continuous* or continual* or continuing* or postacute* or "post acute*" or postdischarg* or "post discharg*" or postinfect* or "post infect*" or postviral* or "post viral*" or postvirus* or "post virus*" or "medium* term*" or mediumterm*) adj3 ((physiolog* or neuro* or cardio* or gastro* or musculo* or renal* or kidney* or cognitive* or cognition* or rheumato* or dermatol* or skin* or haematol* or blood* or autonomic* or nervous* or nervous system* or otolaryngol* or laryngol* or otolog* or cerebro* or brain* or vascular* or respirator* or lung* or pulmonary* or psycholog* or mental health* or mental* or psychiatr* or exertion* or debilit* or devitali* or enervat* or drain* or sleep* or weak* or tired* or frail* or sapp* or strength* or confusion* or letharg* or fatigue* or tired* or weariness* or exhaust* or malaise* or pain* or headache* or breathless* or breathing* or myalgia* or delirious* or delirium* or appetite* or muscle* or muscular* or fitness* or memory* or stress* or depress* or anxiety* or emotion* or cough* or fever* or temperatur* or pneumon* or conjunctivit* or throat* or pharyngit* or dyspnea* or dyspnoea* or sick* or nausea* or nauseous* or vomit* or diarrhoea* or diarrhea* or taste* or anosmia* or smell* or olfact* or sweat* or dehydrat* or pyrexi* or nasal* or nose* or mucus* or ear* or hearing* |

| | |
|---|---|
| | or deaf* or "brain fog*" or cardiac* or thoracic* or chest* or ischemic* or ischaemic* or heart* or liver* or hepatic* or immuno* or palpitation* or vertigo* or metabol*) adj1 (sequela* or complication* or consequence* or effect* or event* or impact* or outcome* or reaction* or complexit* or aftercare* or impair* or problem* or issue* or symptom* or disorder* or abnormal*)) adj10 (covid* or coronavirus* or corona* virus* or coronovirus* or corono* virus* or coronavirinae* or corona* virinae* or Cov or "2019-nCoV*" or 2019nCoV* or "19- nCoV*" or 19nCoV* or nCoV2019* or "nCoV-2019*" or nCoV19* or "nCoV-19*" or "HCoV-19*" or HCoV19* or "HCoV-2019*" or HCoV2019* or "2019 novel*" or Ncov* or "n-cov" or "SARS-CoV-2*" or "SARSCoV-2*" or "SARSCoV2*" or "SARS-CoV2*" or SARSCov19* or "SARS-Cov19*" or "SARSCov-19*" or "SARS-Cov-19*" or SARSCov2019* or "SARS-Cov2019*" or "SARSCov-2019*" or "SARS-Cov-2019*" or SARS2* or "SARS-2*" or SARScoronavirus2* or "SARS-coronavirus-2*" or "SARScoronavirus 2*" or "SARS coronavirus2*" or SARScoronovirus2* or "SARScoronovirus-2*" or "SARScoronovirus 2*" or "SARS coronovirus2*" or "severe acute respiratory syndrome*")).ti,ab. |
| 28 | ((physiolog* or neuro* or cardio* or gastro* or musculo* or renal* or kidney* or cognitive* 49 42 or cognition* or rheumato* or dermatol* or skin* or haematol* or blood* or autonomic* or nervous* or nervous system* or otolaryngol* or laryngol* or otolog* or cerebro* or brain* or vascular* or respirator* or lung* or pulmonary* or psycholog* or mental health* or mental* or psychiatr* or exertion* or debilit* or devitali* or enervat* or drain* or sleep* or weak* or tired* or frail* or sapp* or strength* or confusion* or letharg* or fatigue* or tired* or weariness* or exhaust* or malaise* or pain* or headache* or breathless* or breathing* or myalgia* or delirious* or delirium* or appetite* or muscle* or muscular* or fitness* or memory* or stress* or depress* or anxiety* or emotion* or cough* or fever* or temperatur* or pneumon* or conjunctivit* or throat* or pharyngit* or dyspnea* or dyspnoea* or sick* or nausea* or nauseous* or vomit* or diarrhoea* or diarrhea* or taste* or anosmia* or smell* or olfact* or sweat* or dehydrat* or pyrexi* or nasal* or nose* or mucus* or ear* or hearing* or deaf* or "brain fog*" or cardiac* or thoracic* or chest* or ischemic* or ischaemic* or heart* or liver* or hepatic* or immuno* or palpitation* or vertigo* or metabol*) adj3 (postcovid* or post covid* or postcoronavirus* or postcorona* virus* or post coronavirus* or post corona* virus* or postcoronovirus* or postcorono* virus* or post coronovirus* or post corono* virus* or postcoronavirinae* or postcorona* virinae* or post coronavirinae* or post corona* virinae* or postCov or post Cov or postsars* or post sars* or "post severe acute respiratory syndrome*" or postncov* or post ncov* or posthcov* or post hcov*)).ti,ab. |
| 29 | ((physiolog* or neuro* or cardio* or gastro* or musculo* or renal* or kidney* or cognitive* or cognition* or rheumato* or dermatol* or skin* or haematol* or blood* or autonomic* or nervous* or nervous system* or otolaryngol* or laryngol* or otolog* or cerebro* or brain* or vascular* or respirator* or lung* or pulmonary* or psycholog* or mental health* or mental* or psychiatr* or exertion* or debilit* or devitali* or enervat* or drain* or sleep* or weak* or tired* or frail* or sapp* or strength* or confusion* or letharg* or fatigue* or tired* or weariness* or exhaust* or malaise* or pain* or headache* or breathless* or breathing* or myalgia* or delirious* or delirium* or appetite* or muscle* or muscular* or fitness* or memory* or stress* or depress* or anxiety* or |

| | |
|---|---|
| | emotion* or cough* or fever* or temperatur* or pneumon* or conjunctivit* or throat* or pharyngit* or dyspnea* or dyspnoea* or sick* or nausea* or nauseous* or vomit* or diarrhoea* or diarrhea* or taste* or anosmia* or smell* or olfact* or sweat* or dehydrat* or pyrexi* or nasal* or nose* or mucus* or ear* or hearing* or deaf* or "brain fog*" or cardiac* or thoracic* or chest* or ischemic* or ischaemic* or heart* or liver* or hepatic* or immuno* or palpitation* or vertigo* or metabol*) adj1 (sequela* or complication* or consequence* or complexit*) adj10 (covid* or coronavirus* or corona* virus* or coronovirus* or corono* virus* or coronavirinae* or corona* virinae* or Cov or "2019-nCoV*" or 2019nCoV* or "19-nCoV*" or 19nCoV* or nCoV2019* or "nCoV-2019*" or nCoV19* or "nCoV-19*" or "HCoV-19*" or HCoV19* or "HCoV-2019*" or HCoV2019* or "2019 novel*" or Ncov* or "n-cov" or "527 43 SARS-CoV-2*" or "SARSCoV-2*" or "SARSCoV2*" or "SARS-CoV2*" or SARSCov19* or "SARS-Cov19*" or "SARSCov-19*" or "SARS-Cov-19*" or SARSCov2019* or "SARSCov2019*" or "SARSCov-2019*" or "SARS-Cov-2019*" or SARS2* or "SARS-2*" or SARScoronavirus2* or "SARS-coronavirus-2*" or "SARScoronavirus 2*" or "SARS coronavirus2*" or SARScoronovirus2* or "SARS-coronovirus-2*" or "SARScoronovirus 2*" or "SARS coronovirus2*" or "severe acute respiratory syndrome*")).ti,ab. |
| 30 | (((((multi or multiple* or overlap* or cluster* or numerous* or varied* or variety*) adj1 (symptom* or system* or disease* or disorder* or illness* or condition* or syndrome*)) or (multisymptom* or multisystem* or multidisease* or multidisorder* or multiillness* or multicondition* or multisyndrom*)) adj10 (covid* or coronavirus* or corona* virus* or coronovirus* or corono* virus* or coronavirinae* or corona* virinae* or Cov or "2019- nCoV*" or 2019nCoV* or "19-nCoV*" or 19nCoV* or nCoV2019* or "nCoV-2019*" or nCoV19* or "nCoV-19*" or "HCoV-19*" or HCoV19* or "HCoV-2019*" or HCoV2019* or "2019 novel*" or Ncov* or "n-cov" or "SARS-CoV-2*" or "SARSCoV-2*" or "SARSCoV2*" or "SARS-CoV2*" or SARSCov19* or "SARS-Cov19*" or "SARSCov-19*" or "SARS-Cov19*" or SARSCov2019* or "SARS-Cov2019*" or "SARSCov-2019*" or "SARS-Cov2019*" or SARS2* or "SARS-2*" or SARScoronavirus2* or "SARS-coronavirus-2*" or "SARScoronavirus 2*" or "SARS coronavirus2*" or SARScoronovirus2* or "SARScoronovirus-2*" or "SARScoronovirus 2*" or "SARS coronovirus2*" or "severe acute respiratory syndrome*")) not ("Multisystem* inflammatory* syndrome*" or "inflammatory* multisystem* syndrome*")).ti,ab. |
| 31 | or/20-30 |
| 32 | 7 or 31 |
| 33 | (2021* not ("20210101" or "20210102" or "20210103" or "20210104" or "20210105")).up. |
| 34 | 32 and 33 |
| 35 | from 34 keep 1-82 |

Database(s): **Embase** 1974 to 2021 April 13
Search Strategy:

| # | Searches |
|---|---|

| | |
|---|---|
| 1 | (longcovid* or long covid* or longcoronavirus* or longcorona* virus* or long coronavirus* or long corona* virus* or longcoronovirus* or longcorono* virus* or long coronovirus* or long corono* virus* or longcoronavirinae* or longcorona* virinae* or long coronavirinae* or long corona* virinae* or longCov or long Cov or longsars* or long sars* or "long severe acute respiratory syndrome*" or longncov* or long ncov* or longhcov* or long hcov*).ti,ab,kw. |
| 2 | ((long* or endur* or legacy* or slow* or gradual* or protract* or lengthy* or chronic* or persist* or relaps* or remit* or remission* or residual* or delay* or prolong* or extend* or linger* or permanent* or fluctuat* or sequela* or multisystem* or "multi system*" or nonrecover* or "non recover*" or subacute* or "sub acute*" or lasting* or continuous* or continual* or continuing* or postacute* or "post acute*" or postdischarg* or "post discharg*" or postinfect* or "post infect*" or postviral* or "post viral*" or postvirus* or "post virus*") adj1 (covid* or coronavirus* or corona* virus* or coronovirus* or corono* virus* or coronavirinae* or corona* virinae* or Cov or "2019-nCoV*" or 2019nCoV* or "19-nCoV*" or 19nCoV* or nCoV2019* or "nCoV-2019*" or nCoV19* or "nCoV-19*" or "HCoV-19*" or HCoV19* or "HCoV-2019*" or HCoV2019* or "2019 novel*" or Ncov* or "n-cov" or "SARS-CoV-2*" or "SARSCoV-2*" or "SARSCoV2*" or "SARS-CoV2*" or SARSCov19* or "SARS-Cov19*" or "SARSCov-19*" or "SARS-Cov-19*" or SARSCov2019* or "SARSCov2019*" or "SARSCov-2019*" or "SARS-Cov-2019*" or SARS2* or "SARS-2*" or SARScoronavirus2* or "SARS-coronavirus-2*" or "SARScoronavirus 2*" or "SARS coronavirus2*" or SARScoronovirus2* or "SARS-coronovirus-2*" or "SARScoronovirus 2*" or "SARS coronovirus2*" or "severe acute respiratory syndrome*")).ti,ab. |
| 3 | (("long* term*" or longterm* or "long* haul*" or longhaul* or "long* tail*" or longtail* or longduration* or "long duration*" or longlast* or "long last*" or longstanding* or "long standing*" or "medium* term*" or mediumterm*) adj3 (covid* or coronavirus* or corona* virus* or coronovirus* or corono* virus* or coronavirinae* or corona* virinae* or Cov or "2019-nCoV*" or 2019nCoV* or "19-nCoV*" or 19nCoV* or nCoV2019* or "nCoV-2019*" or nCoV19* or "nCoV-19*" or "HCoV-19*" or HCoV19* or "HCoV-2019*" or HCoV2019* or "2019 novel*" or Ncov* or "n-cov" or "SARS-CoV-2*" or "SARSCoV-2*" or "SARSCoV2*" or "SARS-CoV2*" or SARSCov19* or "SARS-Cov19*" or "SARSCov-19*" or "SARS-Cov-19*" or SARSCov2019* or "SARS-Cov2019*" or "SARSCov-2019*" or "SARS-Cov-2019*" or SARS2* or "SARS-2*" or SARScoronavirus2* or "SARS164 37 coronavirus-2*" or "SARScoronavirus 2*" or "SARS coronavirus2*" or SARScoronovirus2* or "SARS-coronovirus-2*" or "SARScoronovirus 2*" or "SARS coronovirus2*" or "severe acute respiratory syndrome*")).ti,ab. |
| 4 | ((postcovid* or post covid* or postcoronavirus* or postcorona* virus* or post coronavirus* or post corona* virus* or postcoronovirus* or postcorono* virus* or post coronovirus* or post corono* virus* or postcoronavirinae* or postcorona* virinae* or post coronavirinae* or post corona* virinae* or postCov or post Cov or postsars* or post sars* or "post severe acute respiratory syndrome*" or postncov* or post ncov* or posthcov* or post hcov*) adj3 (syndrome* or disorder* or illness* or sickness* or disease* or condition* or symptom* or sign* or prognos* or followup* or "follow up*" or feature* or comorbid* or "co morbid*" or multimorbid* or "multi morbid*" or survivor* |

| | |
|---|---|
| | or survival* or risk* or care* or convalescen* or recuperat* or aftercare* or ambulatory* or outpatient* or "out patient*")).ti,ab. |
| 5 | ((ongoing* or long* or endur* or legacy* or slow* or gradual* or protract* or lengthy* or chronic* or persist* or relaps* or remit* or remission* or residual* or delay* or prolong* or extend* or linger* or permanent* or fluctuat* or multisystem* or "multi system*" or nonrecover* or "non recover*" or subacute* or "sub acute*" or lasting* or continuous* or continual* or continuing* or postacute* or "post acute*" or postdischarg* or "post discharg*" or postinfect* or "post infect*" or postviral* or "post viral*" or postvirus* or "post virus*" or "medium* term*" or mediumterm*) adj4 (sequela* or illness* or symptom* or sign* or prognos* or rehab* or convalescen* or recuperat* or followup* or "follow up*" or feature*) adj10 (covid* or coronavirus* or corona* virus* or coronovirus* or corono* virus* or coronavirinae* or corona* virinae* or Cov or "2019-nCoV*" or 2019nCoV* or "19- nCoV*" or 19nCoV* or nCoV2019* or "nCoV-2019*" or nCoV19* or "nCoV-19*" or "HCoV-19*" or HCoV19* or "HCoV-2019*" or HCoV2019* or "2019 novel*" or Ncov* or "n-cov" or "SARS-CoV-2*" or "SARSCoV-2*" or "SARSCoV2*" or "SARS-CoV2*" or SARSCov19* or "SARS-Cov19*" or "SARSCov-19*" or "SARS-Cov-19*" or SARSCov2019* or "SARS-Cov2019*" or "SARSCov-2019*" or "SARS-Cov-2019*" or SARS2* or "SARS-2*" or SARScoronavirus2* or "SARS-coronavirus-2*" or "SARScoronavirus 2*" or "SARS coronavirus2*" or SARScoronovirus2* or "SARScoronovirus-2*" or "SARScoronovirus 2*" or "SARS coronovirus2*" or "severe acute respiratory syndrome*")).ti,ab. |
| 6 | ((ongoing* or long* or endur* or legacy* or slow* or gradual* or protract* or lengthy* or chronic* or persist* or relaps* or remit* or remission* or residual* or delay* or prolong* or extend* or linger* or permanent* or fluctuat* or multisystem* or "multi system*" or subacute* or "sub acute*" or lasting* or continuous* or continual* or continuing* or post* or after* or follow* or "medium* term*" or mediumterm*) adj1 recover* adj10 (covid* or coronavirus* or corona* virus* or coronovirus* or corono* virus* or coronavirinae* or corona* virinae* or Cov or "2019-nCoV*" or 2019nCoV* or "19-nCoV*" or 19nCoV* or 128 38 nCoV2019* or "nCoV-2019*" or nCoV19* or "nCoV-19*" or "HCoV-19*" or HCoV19* or "HCoV-2019*" or HCoV2019* or "2019 novel*" or Ncov* or "n-cov" or "SARS-CoV-2*" or "SARSCoV-2*" or "SARSCoV2*" or "SARS-CoV2*" or SARSCov19* or "SARS-Cov19*" or "SARSCov-19*" or "SARS-Cov-19*" or SARSCov2019* or "SARS-Cov2019*" or "SARSCov-2019*" or "SARS-Cov-2019*" or SARS2* or "SARS-2*" or SARScoronavirus2* or "SARS-coronavirus-2*" or "SARScoronavirus 2*" or "SARS coronavirus2*" or SARScoronovirus2* or "SARS-coronovirus-2*" or "SARScoronovirus 2*" or "SARS coronovirus2*" or "severe acute respiratory syndrome*")).ti,ab. |
| 7 | or/1-6 |
| 8 | exp Coronavirinae/ |
| 9 | exp Coronavirus infection/ |
| 10 | "coronavirus disease 2019"/ or "severe acute respiratory syndrome coronavirus 2"/ |

| | |
|---|---|
| 11 | (covid* or coronavirus* or corona* virus* or coronovirus* or corono* virus* or coronavirinae* or corona* virinae* or Cov or "2019-nCoV*" or 2019nCoV* or "19-nCoV*" or 19nCoV* or nCoV2019* or "nCoV-2019*" or nCoV19* or "nCoV-19*" or "HCoV-19*" or HCoV19* or "HCoV-2019*" or HCoV2019* or "2019 novel*" or Ncov* or "n-cov" or "SARS-CoV-2*" or "SARSCoV-2*" or "SARSCoV2*" or "SARS-CoV2*" or SARSCov19* or "SARS-Cov19*" or "SARSCov-19*" or "SARS-Cov-19*" or SARSCov2019* or "SARSCov2019*" or "SARSCov-2019*" or "SARS-Cov-2019*" or SARS2* or "SARS-2*" or SARScoronavirus2* or "SARS-coronavirus-2*" or "SARScoronavirus 2*" or "SARS coronavirus2*" or SARScoronovirus2* or "SARS-coronovirus-2*" or "SARScoronovirus 2*" or "SARS coronovirus2*" or "severe acute respiratory syndrome*").ti. |
| 12 | or/8-11 |
| 13 | Functional status/ |
| 14 | Aftercare/ |
| 15 | Convalescence/ |
| 16 | rehabilitation/ |
| 17 | Rehabilitation care/ |
| 18 | daily life activity/ |
| 19 | physical performance/ |
| 20 | or/13-19 |
| 21 | 12 and 20 |
| 22 | ("long* haul*" or longhaul* or "long* tail*" or longtail* or longduration* or "long duration*" or longlast* or "long last*" or longstanding* or "long standing*").ti,ab. and (8 or 9 or 10) |
| 23 | ((recover* or nonrecover*) adj3 function* adj10 (covid* or coronavirus* or corona* virus* or coronovirus* or corono* virus* or coronavirinae* or corona* virinae* or Cov or "2019- 15 39 nCoV*" or 2019nCoV* or "19-nCoV*" or 19nCoV* or nCoV2019* or "nCoV-2019*" or nCoV19* or "nCoV-19*" or "HCoV-19*" or HCoV19* or "HCoV-2019*" or HCoV2019* or "2019 novel*" or Ncov* or "n-cov" or "SARS-CoV-2*" or "SARSCoV-2*" or "SARSCoV2*" or "SARS-CoV2*" or SARSCov19* or "SARS-Cov19*" or "SARSCov-19*" or "SARS-Cov19*" or SARSCov2019* or "SARS-Cov2019*" or "SARSCov-2019*" or "SARS-Cov2019*" or SARS2* or "SARS-2*" or SARScoronavirus2* or "SARS-coronavirus-2*" or "SARScoronavirus 2*" or "SARS coronavirus2*" or SARScoronovirus2* or "SARScoronovirus-2*" or "SARScoronovirus 2*" or "SARS coronovirus2*" or "severe acute respiratory syndrome*")).ti,ab. |
| 24 | ((postacute* or "post acute*" or postdischarg* or "post discharg*" or postinfect* or "post infect*" or postviral* or "post viral*" or postvirus* or "post virus*" or subacute* or "sub acute*") adj3 (care* or convalescen* or recuperat* or aftercare* or ambulatory* or outpatient* or "out patient*" or survivor* or survival*) adj10 (covid* or coronavirus* or corona* virus* or coronovirus* or corono* virus* or coronavirinae* or corona* virinae* or Cov or "2019-nCoV*" or 2019nCoV* or "19-nCoV*" or 19nCoV* or nCoV2019* or "nCoV2019*" or nCoV19* or "nCoV- |

| | |
|---|---|
| | 19*" or "HCoV-19*" or HCoV19* or "HCoV-2019*" or HCoV2019* or "2019 novel*" or Ncov* or "n-cov" or "SARS-CoV-2*" or "SARSCoV-2*" or "SARSCoV2*" or "SARS-CoV2*" or SARSCov19* or "SARS-Cov19*" or "SARSCov-19*" or "SARS-Cov-19*" or SARSCov2019* or "SARS-Cov2019*" or "SARSCov-2019*" or "SARS-Cov-2019*" or SARS2* or "SARS-2*" or SARScoronavirus2* or "SARScoronavirus-2*" or "SARScoronavirus 2*" or "SARS coronavirus2*" or SARScoronovirus2* or "SARS-coronovirus-2*" or "SARScoronovirus 2*" or "SARS coronovirus2*" or "severe acute respiratory syndrome*")).ti,ab. |
| 25 | ((convalescen* or recuperat* or after* or followup* or "follow up*" or rehab*) adj1 (therap* or care*) adj10 (covid* or coronavirus* or corona* virus* or coronovirus* or corono* virus* or coronavirinae* or corona* virinae* or Cov or "2019-nCoV*" or 2019nCoV* or "19- nCoV*" or 19nCoV* or nCoV2019* or "nCoV-2019*" or nCoV19* or "nCoV-19*" or "HCoV-19*" or HCoV19* or "HCoV-2019*" or HCoV2019* or "2019 novel*" or Ncov* or "n-cov" or "SARS-CoV-2*" or "SARSCoV-2*" or "SARSCoV2*" or "SARS-CoV2*" or SARSCov19* or "SARS-Cov19*" or "SARSCov-19*" or "SARS-Cov-19*" or SARSCov2019* or "SARS-Cov2019*" or "SARSCov-2019*" or "SARS-Cov-2019*" or SARS2* or "SARS-2*" or SARScoronavirus2* or "SARS-coronavirus-2*" or "SARScoronavirus 2*" or "SARS coronavirus2*" or SARScoronovirus2* or "SARScoronovirus-2*" or "SARScoronovirus 2*" or "SARS coronovirus2*" or "severe acute respiratory syndrome*")).ti,ab. |
| 26 | ((ongoing* or endur* or long* or legacy* or slow* or gradual* or protract* or lengthy* or chronic* or persist* or relaps* or remit* or remission* or residual* or delay* or prolong* or extend* or linger* or permanent* or fluctuat* or multisystem* or "multi system*" or 58 40 nonrecover* or "non recover*" or subacute* or "sub acute*" or lasting* or continuous* or continual* or continuing* or postacute* or "post acute*" or postdischarg* or "post discharg*" or postinfect* or "post infect*" or postviral* or "post viral*" or postvirus* or "post virus*" or "medium* term*" or mediumterm* or adverse* or dangerous* or harmful* or indirect* or injurious* or secondary* or side effect* or undesirable* or sequela* or complication* or consequence* or effect* or event* or impact* or outcome* or reaction* or complexit* or aftercare* or impair* or problem* or issue* or rehab* or function* or perform*) adj10 ((daily* or everyday* or day* or normal* or usual*) adj1 (activit* or living* or life* or lives* or job* or work* or employ* or occupation* or hobby* or hobbies* or leisure*)) adj10 (covid* or coronavirus* or corona* virus* or coronovirus* or corono* virus* or coronavirinae* or corona* virinae* or Cov or "2019-nCoV*" or 2019nCoV* or "19- nCoV*" or 19nCoV* or nCoV2019* or "nCoV-2019*" or nCoV19* or "nCoV-19*" or "HCoV-19*" or HCoV19* or "HCoV-2019*" or HCoV2019* or "2019 novel*" or Ncov* or "n-cov" or "SARS-CoV-2*" or "SARSCoV-2*" or "SARSCoV2*" or "SARS-CoV2*" or SARSCov19* or "SARS-Cov19*" or "SARSCov-19*" or "SARS-Cov-19*" or SARSCov2019* or "SARS-Cov2019*" or "SARSCov-2019*" or "SARS-Cov-2019*" or SARS2* or "SARS-2*" or SARScoronavirus2* or "SARS-coronavirus-2*" or "SARScoronavirus 2*" or "SARS coronavirus2*" or SARScoronovirus2* or "SARScoronovirus-2*" or "SARScoronovirus 2*" or "SARS coronovirus2*" or "severe acute respiratory syndrome*")).ti,ab. |
| 27 | ((ongoing* or endur* or long* or legacy* or slow* or gradual* or protract* or lengthy* or chronic* or persist* or relaps* or remit* or remission* or residual* or delay* or prolong* or extend* or linger* or permanent* or fluctuat* |

| | |
|---|---|
| | or multisystem* or "multi system*" or nonrecover* or "non recover*" or subacute* or "sub acute*" or lasting* or continuous* or continual* or continuing* or postacute* or "post acute*" or postdischarg* or "post discharg*" or postinfect* or "post infect*" or postviral* or "post viral*" or postvirus* or "post virus*" or "medium* term*" or mediumterm*) adj3 ((health* or adverse* or dangerous* or harmful* or indirect* or injurious* or secondary* or side* or undesirable* or negative* or damaging* or detriment* or abnormal*) adj1 (sequela* or complication* or consequence* or effect* or event* or impact* or outcome* or reaction* or complexit* or aftercare* or impair* or problem* or issue* or symptom* or disorder*)) adj10 (covid* or coronavirus* or corona* virus* or coronovirus* or corono* virus* or coronavirinae* or corona* virinae* or Cov or "2019-nCoV*" or 2019nCoV* or "19-nCoV*" or 19nCoV* or nCoV2019* or "nCoV2019*" or nCoV19* or "nCoV-19*" or "HCoV-19*" or HCoV19* or "HCoV-2019*" or HCoV2019* or "2019 novel*" or Ncov* or "n-cov" or "SARS-CoV-2*" or "SARSCoV-2*" or "SARSCoV2*" or "SARS-CoV2*" or SARSCov19* or "SARS-Cov19*" or "SARSCov-19*" or "SARS-Cov-19*" or SARSCov2019* or "SARS-Cov2019*" or "SARSCov-2019*" or "SARS-Cov-2019*" or SARS2* or "SARS-2*" or SARScoronavirus2* or "SARS40 41 coronavirus-2*" or "SARScoronavirus 2*" or "SARS coronavirus2*" or SARScoronovirus2* or "SARS-coronovirus-2*" or "SARScoronovirus 2*" or "SARS coronovirus2*" or "severe acute respiratory syndrome*")).ti,ab. |
| 28 | ((ongoing* or endur* or long* or legacy* or slow* or gradual* or protract* or lengthy* or chronic* or persist* or relaps* or remit* or remission* or residual* or delay* or prolong* or extend* or linger* or permanent* or fluctuat* or multisystem* or "multi system*" or nonrecover* or "non recover*" or subacute* or "sub acute*" or lasting* or continuous* or continual* or continuing* or postacute* or "post acute*" or postdischarg* or "post discharg*" or postinfect* or "post infect*" or postviral* or "post viral*" or postvirus* or "post virus*" or "medium* term*" or mediumterm*) adj3 ((physiolog* or neuro* or cardio* or gastro* or musculo* or renal* or kidney* or cognitive* or cognition* or rheumato* or dermatol* or skin* or haematol* or blood* or autonomic* or nervous* or nervous system* or otolaryngol* or laryngol* or otolog* or cerebro* or brain* or vascular* or respirator* or lung* or pulmonary* or psycholog* or mental health* or mental* or psychiatr* or exertion* or debilit* or devitali* or enervat* or drain* or sleep* or weak* or tired* or frail* or sapp* or strength* or confusion* or letharg* or fatigue* or tired* or weariness* or exhaust* or malaise* or pain* or headache* or breathless* or breathing* or myalgia* or delirious* or delirium* or appetite* or muscle* or muscular* or fitness* or memory* or stress* or depress* or anxiety* or emotion* or cough* or fever* or temperatur* or pneumon* or conjunctivit* or throat* or pharyngit* or dyspnea* or dyspnoea* or sick* or nausea* or nauseous* or vomit* or diarrhoea* or diarrhea* or taste* or anosmia* or smell* or olfact* or sweat* or dehydrat* or pyrexi* or nasal* or nose* or mucus* or ear* or hearing* or deaf* or "brain fog*" or cardiac* or thoracic* or chest* or ischemic* or ischaemic* or heart* or liver* or hepatic* or immuno* or palpitation* or vertigo* or metabol*) adj1 (sequela* or complication* or consequence* or effect* or event* or impact* or outcome* or reaction* or complexit* or aftercare* or impair* or problem* or issue* or symptom* or disorder* or abnormal*)) adj10 (covid* or coronavirus* or corona* virus* or coronovirus* or corono* virus* or coronavirinae* or corona* virinae* or Cov or "2019-nCoV*" or 2019nCoV* or "19- nCoV*" or |

| | |
|---|---|
| | 19nCoV* or nCoV2019* or "nCoV-2019*" or nCoV19* or "nCoV-19*" or "HCoV-19*" or HCoV19* or "HCoV-2019*" or HCoV2019* or "2019 novel*" or Ncov* or "n-cov" or "SARS-CoV-2*" or "SARSCoV-2*" or "SARSCoV2*" or "SARS-CoV2*" or SARSCov19* or "SARS-Cov19*" or "SARSCov-19*" or "SARS-Cov-19*" or SARSCov2019* or "SARS-Cov2019*" or "SARSCov-2019*" or "SARS-Cov-2019*" or SARS2* or "SARS-2*" or SARScoronavirus2* or "SARS-coronavirus-2*" or "SARScoronavirus 2*" or "SARS coronavirus2*" or SARScoronovirus2* or "SARScoronovirus-2*" or "SARScoronovirus 2*" or "SARS coronovirus2*" or "severe acute respiratory syndrome*")).ti,ab. |
| 29 | ((physiolog* or neuro* or cardio* or gastro* or musculo* or renal* or kidney* or cognitive* 49 42 or cognition* or rheumato* or dermatol* or skin* or haematol* or blood* or autonomic* or nervous* or nervous system* or otolaryngol* or laryngol* or otolog* or cerebro* or brain* or vascular* or respirator* or lung* or pulmonary* or psycholog* or mental health* or mental* or psychiatr* or exertion* or debilit* or devitali* or enervat* or drain* or sleep* or weak* or tired* or frail* or sapp* or strength* or confusion* or letharg* or fatigue* or tired* or weariness* or exhaust* or malaise* or pain* or headache* or breathless* or breathing* or myalgia* or delirious* or delirium* or appetite* or muscle* or muscular* or fitness* or memory* or stress* or depress* or anxiety* or emotion* or cough* or fever* or temperatur* or pneumon* or conjunctivit* or throat* or pharyngit* or dyspnea* or dyspnoea* or sick* or nausea* or nauseous* or vomit* or diarrhoea* or diarrhea* or taste* or anosmia* or smell* or olfact* or sweat* or dehydrat* or pyrexi* or nasal* or nose* or mucus* or ear* or hearing* or deaf* or "brain fog*" or cardiac* or thoracic* or chest* or ischemic* or ischaemic* or heart* or liver* or hepatic* or immuno* or palpitation* or vertigo* or metabol*) adj3 (postcovid* or post covid* or postcoronavirus* or postcorona* virus* or post coronavirus* or post corona* virus* or postcoronovirus* or postcorono* virus* or post coronovirus* or post corono* virus* or postcoronavirinae* or postcorona* virinae* or post coronavirinae* or post corona* virinae* or postCov or post Cov or postsars* or post sars* or "post severe acute respiratory syndrome*" or postncov* or post ncov* or posthcov* or post hcov*)).ti,ab. |
| 30 | ((physiolog* or neuro* or cardio* or gastro* or musculo* or renal* or kidney* or cognitive* or cognition* or rheumato* or dermatol* or skin* or haematol* or blood* or autonomic* or nervous* or nervous system* or otolaryngol* or laryngol* or otolog* or cerebro* or brain* or vascular* or respirator* or lung* or pulmonary* or psycholog* or mental health* or mental* or psychiatr* or exertion* or debilit* or devitali* or enervat* or drain* or sleep* or weak* or tired* or frail* or sapp* or strength* or confusion* or letharg* or fatigue* or tired* or weariness* or exhaust* or malaise* or pain* or headache* or breathless* or breathing* or myalgia* or delirious* or delirium* or appetite* or muscle* or muscular* or fitness* or memory* or stress* or depress* or anxiety* or emotion* or cough* or fever* or temperatur* or pneumon* or conjunctivit* or throat* or pharyngit* or dyspnea* or dyspnoea* or sick* or nausea* or nauseous* or vomit* or diarrhoea* or diarrhea* or taste* or anosmia* or smell* or olfact* or sweat* or dehydrat* or pyrexi* or nasal* or nose* or mucus* or ear* or hearing* or deaf* or "brain fog*" or cardiac* or thoracic* or chest* or ischemic* or ischaemic* or heart* or liver* or hepatic* or immuno* or palpitation* or vertigo* or metabol*) adj1 (sequela* or complication* or consequence* or |

| | |
|---|---|
| | complexit*) adj10 (covid* or coronavirus* or corona* virus* or coronovirus* or corono* virus* or coronavirinae* or corona* virinae* or Cov or "2019-nCoV*" or 2019nCoV* or "19-nCoV*" or 19nCoV* or nCoV2019* or "nCoV-2019*" or nCoV19* or "nCoV-19*" or "HCoV-19*" or HCoV19* or "HCoV-2019*" or HCoV2019* or "2019 novel*" or Ncov* or "n-cov" or "527 43 SARS-CoV-2*" or "SARSCoV-2*" or "SARSCoV2*" or "SARS-CoV2*" or SARSCov19* or "SARS-Cov19*" or "SARSCov-19*" or "SARS-Cov-19*" or SARSCov2019* or "SARSCov2019*" or "SARSCov-2019*" or "SARS-Cov-2019*" or SARS2* or "SARS-2*" or SARScoronavirus2* or "SARS-coronavirus-2*" or "SARScoronavirus 2*" or "SARS coronavirus2*" or SARScoronovirus2* or "SARS-coronovirus-2*" or "SARScoronovirus 2*" or "SARS coronovirus2*" or "severe acute respiratory syndrome*")).ti,ab. |
| 31 | (((((multi or multiple* or overlap* or cluster* or numerous* or varied* or variety*) adj1 (symptom* or system* or disease* or disorder* or illness* or condition* or syndrome*)) or (multisymptom* or multisystem* or multidisease* or multidisorder* or multiillness* or multicondition* or multisyndrom*)) adj10 (covid* or coronavirus* or corona* virus* or coronovirus* or corono* virus* or coronavirinae* or corona* virinae* or Cov or "2019- nCoV*" or 2019nCoV* or "19-nCoV*" or 19nCoV* or nCoV2019* or "nCoV-2019*" or nCoV19* or "nCoV-19*" or "HCoV-19*" or HCoV19* or "HCoV-2019*" or HCoV2019* or "2019 novel*" or Ncov* or "n-cov" or "SARS-CoV-2*" or "SARSCoV-2*" or "SARSCoV2*" or "SARS-CoV2*" or SARSCov19* or "SARS-Cov19*" or "SARSCov-19*" or "SARS-Cov19*" or SARSCov2019* or "SARS-Cov2019*" or "SARSCov-2019*" or "SARS-Cov2019*" or SARS2* or "SARS-2*" or SARScoronavirus2* or "SARS-coronavirus-2*" or "SARScoronavirus 2*" or "SARS coronavirus2*" or SARScoronovirus2* or "SARScoronovirus-2*" or "SARScoronovirus 2*" or "SARS coronovirus2*" or "severe acute respiratory syndrome*")) not ("Multisystem* inflammatory* syndrome*" or "inflammatory* multisystem* syndrome*")).ti,ab. |
| 32 | or/21-31 |
| 33 | 7 or 32 |
| 34 | (letter or editorial).pt. |
| 35 | 33 not 34 |
| 36 | nonhuman/ not (human/ and nonhuman/) |
| 37 | 35 not 36 |
| 38 | (nonhuman* or animal* or amphib* or ape or apes or beagle* or bird or birds or boar? or bovine* or canin* or cat or cats or catfish* or cattle or chicken* or chimpanzee* or cow or cows or dog or dogs or equine or ewe? or fish or fishes or feline* or frog or frogs or gastropod* or gerbil* or goat* or "guinea-pig*" or hamster? or hare or hares or horse* or macaque* or mammal* or marmoset* or mice or monkey* or mouse or murine or ostrich* or ovine or pig or piglet* or pigs or porcine or pork* or primate* or rabbit* or rat or rats or rodent? or sheep or sow or sows or swine* or toad? or veterinar* or zebrafish* or zebra fish*).ti. |

| | |
|---|---|
| 39 | (human or humans or adolescen* or baby or babies or boy or boys or child* or elderly or girl or girls* or man or mans or men or mens or patient* or people* or person or persons or senior or seniors or teen* or woman* or women* or volunteer?).mp. |
| 40 | 38 not (38 and 39) |
| 41 | 37 not 40 |
| 42 | (2021* not ("20210101" or "20210102" or "20210103" or "20210104" or "20210105")).dc,dd. |
| 43 | 41 and 42 |

Embase Update 1

Database(s): **Embase** 1974 to 2021 April 13
Search Strategy:

| # | Searches |
|---|---|
| 1 | ((ongoing* or endur* or long* or legacy* or slow* or gradual* or protract* or lengthy* or chronic* or persist* or relaps* or remit* or remission* or residual* or delay* or prolong* or extend* or linger* or permanent* or fluctuat* or multisystem* or "multi system*" or nonrecover* or "non recover*" or subacute* or "sub acute*" or lasting* or continuous* or continual* or continuing* or postacute* or "post acute*" or postdischarg* or "post discharg*" or postinfect* or "post infect*" or postviral* or "post viral*" or postvirus* or "post virus*" or "medium* term*" or mediumterm*) adj3 ((vestibular* or endocrine* or encephalit*) adj1 (sequela* or complication* or consequence* or effect* or event* or impact* or outcome* or reaction* or complexit* or aftercare* or impair* or problem* or issue* or symptom* or disorder* or abnormal*)) adj10 (covid* or coronavirus* or corona* virus* or coronovirus* or corono* virus* or coronavirinae* or corona* virinae* or Cov or "2019-nCoV*" or 2019nCoV* or "19-nCoV*" or 19nCoV* or nCoV2019* or "nCoV-2019*" or nCoV19* or "nCoV-19*" or "HCoV-19*" or HCoV19* or "HCoV-2019*" or HCoV2019* or "2019 novel*" or Ncov* or "ncov" or "SARS-CoV-2*" or "SARSCoV-2*" or "SARSCoV2*" or "SARS-CoV2*" or SARSCov19* or "SARS-Cov19*" or "SARSCov-19*" or "SARS-Cov-19*" or SARSCov2019* or "SARS-Cov2019*" or "SARSCov-2019*" or "SARS-Cov-2019*" or SARS2* or "SARS-2*" or SARScoronavirus2* or "SARS-coronavirus-2*" or "SARScoronavirus 2*" or "SARS coronavirus2*" or SARScoronovirus2* or "SARScoronovirus-2*" or "SARScoronovirus 2*" or "SARS coronovirus2*" or "severe acute respiratory syndrome*")).ti,ab. |
| 2 | ((vestibular* or endocrine* or encephalit*) adj3 (postcovid* or post covid* or postcoronavirus* or postcorona* virus* or post coronavirus* or post corona* virus* or postcoronovirus* or postcorono* virus* or post coronovirus* or post corono* virus* or postcoronavirinae* or postcorona* virinae* or post coronavirinae* or post corona* virinae* or postCov or post Cov or postsars* or post sars* or "post severe acute respiratory syndrome*" or postncov* or post ncov* or posthcov* or post hcov*)).ti,ab. |
| 3 | ((vestibular* or endocrine* or encephalit*) adj1 (sequela* or complication* or consequence* or complexit*) adj10 (covid* or coronavirus* or corona* virus* or coronovirus* or corono* virus* or coronavirinae* or corona* virinae* |

| | |
|---|---|
| | or Cov or "2019-nCoV*" or 2019nCoV* or "19- nCoV*" or 19nCoV* or nCoV2019* or "nCoV-2019*" or nCoV19* or "nCoV-19*" or "HCoV19*" or HCoV19* or "HCoV-2019*" or HCoV2019* or "2019 novel*" or Ncov* or "n-cov" or "SARS-CoV-2*" or "SARSCoV-2*" or "SARSCoV2*" or "SARS-CoV2*" or SARSCov19* or "SARS-Cov19*" or "SARSCov-19*" or "SARS-Cov-19*" or SARSCov2019* or "SARS3 45 Cov2019*" or "SARSCov-2019*" or "SARS-Cov-2019*" or SARS2* or "SARS-2*" or SARScoronavirus2* or "SARS-coronavirus-2*" or "SARScoronavirus 2*" or "SARS coronavirus2*" or SARScoronovirus2* or "SARS-coronovirus-2*" or "SARScoronovirus 2*" or "SARS coronovirus2*" or "severe acute respiratory syndrome*")).ti,ab. |
| 4 | or/1-3 |
| 5 | (2021* not ("20210101" or "20210102" or "20210103" or "20210104" or "20210105")).dc,dd. |
| 6 | 4 and 5 |

Embase TopUp 2

Database(s): **Embase** 1974 to 2021 April 13
Search Strategy:

| # | Searches |
|---|---|
| 1 | ((ongoing* or endur* or long* or legacy* or slow* or gradual* or protract* or lengthy* or chronic* or persist* or relaps* or remit* or remission* or residual* or delay* or prolong* or extend* or linger* or permanent* or fluctuat* or multisystem* or "multi system*" or nonrecover* or "non recover*" or subacute* or "sub acute*" or lasting* or continuous* or continual* or continuing* or postacute* or "post acute*" or postdischarg* or "post discharg*" or postinfect* or "post infect*" or postviral* or "post viral*" or postvirus* or "post virus*" or "medium* term*" or mediumterm*) adj3 ((physical* or cough* or fibrosis* or myocarditis* or Guillain* or barre* or neuralgi* or amyotroph* or thrombo* or clot* or rash* or hive* or urticari* or lymph* or stroke* or TIA or toe* or foot* or feet* or finger* or chilblain* or numb* or inflammat* or inflame* or arthralgi* or eye* or organ or organs or tingl* or sting* or burn* or bladder* or urogenit* or genitourin* or genital* or reproducti* or urinary* or joint* or tachycard* or atrial* or autoimmun* or dysautonomi* or polyneuro* or mast* or mobility* or walking* or ambulation* or energy*) adj1 (sequela* or complication* or consequence* or effect* or event* or impact* or outcome* or reaction* or complexit* or aftercare* or impair* or problem* or issue* or symptom* or disorder* or abnormal*)) adj10 (covid* or coronavirus* or corona* virus* or coronovirus* or corono* virus* or coronavirinae* or corona* virinae* or Cov or "2019-nCoV*" or 2019nCoV* or "19-nCoV*" or 19nCoV* or nCoV2019* or "nCoV-2019*" or nCoV19* or "nCoV-19*" or "HCoV-19*" or HCoV19* or "HCoV-2019*" or HCoV2019* or "2019 novel*" or Ncov* or "n-cov" or "SARSCoV-2*" or "SARSCoV-2*" or "SARSCoV2*" or "SARS-CoV2*" or SARSCov19* or "SARSCov19*" or "SARSCov-19*" or "SARS-Cov-19*" or SARSCov2019* or "SARS-Cov2019*" or "SARSCov-2019*" or "SARS-Cov-2019*" or SARS2* or "SARS-2*" or SARScoronavirus2* or "SARS-coronavirus-2*" or |

| | |
|---|---|
| | "SARScoronavirus 2*" or "SARS coronavirus2*" or SARScoronovirus2* or "SARS-coronovirus-2*" or "SARScoronovirus 2*" or "SARS coronovirus2*" or "severe acute respiratory syndrome*")).ti,ab. |
| 2 | ((physical* or cough* or fibrosis* or myocarditis* or Guillain* or barre* or neuralgi* or amyotroph* or thrombo* or clot* or rash* or hive* or urticari* or lymph* or stroke* or TIA or toe* or foot* or feet* or finger* or chilblain* or numb* or inflammat* or inflame* or "arthralgi* 0 46" or eye* or organ or organs or tingl* or sting* or burn* or bladder* or urogenit* or genitourin* or genital* or reproducti* or urinary* or joint* or tachycard* or atrial* or autoimmun* or dysautonomi* or polyneuro* or mast* or mobility* or walking* or ambulation* or energy*) adj1 (sequela* or complication* or consequence* or effect* or event* or impact* or outcome* or reaction* or complexit* or aftercare* or impair* or problem* or issue* or symptom* or disorder* or abnormal*) adj3 (postcovid* or post covid* or postcoronavirus* or postcorona* virus* or post coronavirus* or post corona* virus* or postcoronovirus* or postcorono* virus* or post coronovirus* or post corono* virus* or postcoronavirinae* or postcorona* virinae* or post coronavirinae* or post corona* virinae* or postCov or post Cov or postsars* or post sars* or "post severe acute respiratory syndrome*" or postncov* or post ncov* or posthcov* or post hcov*)).ti,ab. |
| 3 | ((physical* or cough* or fibrosis* or myocarditis* or Guillain* or barre* or neuralgi* or amyotroph* or thrombo* or clot* or rash* or hive* or urticari* or lymph* or stroke* or TIA or toe* or foot* or feet* or finger* or chilblain* or numb* or inflammat* or inflame* or arthralgi* or eye* or organ or organs or tingl* or sting* or burn* or bladder* or urogenit* or genitourin* or genital* or reproducti* or urinary* or joint* or tachycard* or atrial* or autoimmun* or dysautonomi* or polyneuro* or mast* or mobility* or walking* or ambulation* or energy*) adj1 (sequela* or complication* or consequence* or effect* or event* or impact* or outcome* or reaction* or complexit* or aftercare* or impair* or problem* or issue* or symptom* or disorder* or abnormal*) adj1 (sequela* or complication* or consequence* or complexit*) adj10 (covid* or coronavirus* or corona* virus* or coronovirus* or corono* virus* or coronavirinae* or corona* virinae* or Cov or "2019-nCoV*" or 2019nCoV* or "19- nCoV*" or 19nCoV* or nCoV2019* or "nCoV-2019*" or nCoV19* or "nCoV-19*" or "HCoV19*" or HCoV19* or "HCoV-2019*" or HCoV2019* or "2019 novel*" or Ncov* or "n-cov" or "SARS-CoV-2*" or "SARSCoV-2*" or "SARSCoV2*" or "SARS-CoV2*" or SARSCov19* or "SARS-Cov19*" or "SARSCov-19*" or "SARS-Cov-19*" or SARSCov2019* or "SARSCov2019*" or "SARSCov-2019*" or "SARS-Cov-2019*" or SARS2* or "SARS-2*" or SARScoronavirus2* or "SARS-coronavirus-2*" or "SARScoronavirus 2*" or "SARS coronavirus2*" or SARScoronovirus2* or "SARS-coronovirus-2*" or "SARScoronovirus 2*" or "SARS coronovirus2*" or "severe acute respiratory syndrome*")).ti,ab. |
| 4 | or/1-3 |
| 5 | (letter or editorial).pt. |
| 6 | 4 not 5 |
| 7 | nonhuman/ not (human/ and nonhuman/) |

| | |
|---|---|
| 8 | 6 not 7 |
| 9 | (nonhuman* or animal* or amphib* or ape or apes or beagle* or bird or birds or boar? or bovine* or canin* or cat or cats or catfish* or cattle or chicken* or chimpanzee* or cow or cows or dog or dogs or equine or ewe? or fish or fishes or feline* or frog or frogs or gastropod* or gerbil* or goat* or "guinea-pig*" or hamster? or hare or hares or horse* or macaque* or mammal* or marmoset* or mice or monkey* or mouse or murine or ostrich* or ovine or pig or piglet* or pigs or porcine or pork* or primate* or rabbit* or rat or rats or rodent? or sheep or sow or sows or swine* or toad? or veterinar* or zebrafish* or zebra fish*).ti. |
| 10 | (human or humans or adolescen* or baby or babies or boy or boys or child* or elderly or girl or girls* or man or mans or men or mens or patient* or people* or person or persons or senior or seniors or teen* or woman* or women* or volunteer?).mp. |
| 11 | 9 not (9 and 10) |
| 12 | 8 not 11 |
| 13 | (2021* not ("20210101" or "20210102" or "20210103" or "20210104" or "20210105")).dc,dd. |
| 14 | 12 and 13 |

Medline

Database(s): **Ovid MEDLINE(R) ALL** 1946 to April 13, 2021
Search Strategy:

| # | Searches |
|---|---|
| 1 | (longcovid* or long covid* or longcoronavirus* or longcorona* virus* or long coronavirus* or long corona* virus* or longcoronovirus* or longcorono* virus* or long coronovirus* or long corono* virus* or longcoronavirinae* or longcorona* virinae* or long coronavirinae* or long corona* virinae* or longCov or long Cov or longsars* or long sars* or "long severe acute respiratory syndrome*" or longncov* or long ncov* or longhcov* or long hcov*).ti,ab,kw. |
| 2 | ((long* or endur* or legacy* or slow* or gradual* or protract* or lengthy* or chronic* or persist* or relaps* or remit* or remission* or residual* or delay* or prolong* or extend* or linger* or permanent* or fluctuat* or sequela* or multisystem* or "multi system*" or nonrecover* or "non recover*" or subacute* or "sub acute*" or lasting* or continuous* or continual* or continuing* or postacute* or "post acute*" or postdischarg* or "post discharg*" or postinfect* or "post infect*" or postviral* or "post viral*" or postvirus* or "post virus*") adj1 (covid* or coronavirus* or corona* virus* or coronovirus* or corono* virus* or coronavirinae* or corona* virinae* or Cov or "2019-nCoV*" or 2019nCoV* or "19-nCoV*" or 19nCoV* or nCoV2019* or "nCoV-2019*" or nCoV19* or "nCoV-19*" or "HCoV-19*" or HCoV19* or "HCoV-2019*" or HCoV2019* or "2019 novel*" or Ncov* or "n-cov" or "SARS-CoV-2*" or "SARSCoV-2*" or "SARSCoV2*" or "SARS-CoV2*" or SARSCov19* or "SARS-Cov19*" or "SARSCov-19*" or "SARS-Cov-19*" or SARSCov2019* or "SARSCov2019*" or "SARSCov-2019*" or "SARS-Cov-2019*" or SARS2* or "SARS-2*" or SARScoronavirus2* or "SARS-coronavirus-2*" or |

| | |
|---|---|
| | "SARScoronavirus 2*" or "SARS coronavirus2*" or SARScoronovirus2* or "SARS-coronovirus-2*" or "SARScoronovirus 2*" or "SARS coronovirus2*" or "severe acute respiratory syndrome*")).ti,ab. |
| 3 | (("long* term*" or longterm* or "long* haul*" or longhaul* or "long* tail*" or longtail* or longduration* or "long duration*" or longlast* or "long last*" or longstanding* or "long standing*" or "medium* term*" or mediumterm*) adj3 (covid* or coronavirus* or corona* virus* or coronovirus* or corono* virus* or coronavirinae* or corona* virinae* or Cov or "2019-nCoV*" or 2019nCoV* or "19-nCoV*" or 19nCoV* or nCoV2019* or "nCoV-2019*" or nCoV19* or "nCoV-19*" or "HCoV-19*" or HCoV19* or "HCoV-2019*" or HCoV2019* or "2019 novel*" or Ncov* or "n-cov" or "SARS-CoV-2*" or "SARSCoV-2*" or "SARSCoV2*" or "SARS-CoV2*" or SARSCov19* or "SARS-Cov19*" or "SARSCov-19*" or "SARS-Cov-19*" or SARSCov2019* or "SARS-Cov2019*" or "SARSCov-2019*" or "SARS-Cov-2019*" or SARS2* or "SARS-2*" or SARScoronavirus2* or "SARS164 37 coronavirus-2*" or "SARScoronavirus 2*" or "SARS coronavirus2*" or SARScoronovirus2* or "SARS-coronovirus-2*" or "SARScoronovirus 2*" or "SARS coronovirus2*" or "severe acute respiratory syndrome*")).ti,ab. |
| 4 | ((postcovid* or post covid* or postcoronavirus* or postcorona* virus* or post coronavirus* or post corona* virus* or postcoronovirus* or postcorono* virus* or post coronovirus* or post corono* virus* or postcoronavirinae* or postcorona* virinae* or post coronavirinae* or post corona* virinae* or postCov or post Cov or postsars* or post sars* or "post severe acute respiratory syndrome*" or postncov* or post ncov* or posthcov* or post hcov*) adj3 (syndrome* or disorder* or illness* or sickness* or disease* or condition* or symptom* or sign* or prognos* or followup* or "follow up*" or feature* or comorbid* or "co morbid*" or multimorbid* or "multi morbid*" or survivor* or survival* or risk* or care* or convalescen* or recuperat* or aftercare* or ambulatory* or outpatient* or "out patient*")).ti,ab. |
| 5 | ((ongoing* or long* or endur* or legacy* or slow* or gradual* or protract* or lengthy* or chronic* or persist* or relaps* or remit* or remission* or residual* or delay* or prolong* or extend* or linger* or permanent* or fluctuat* or multisystem* or "multi system*" or nonrecover* or "non recover*" or subacute* or "sub acute*" or lasting* or continuous* or continual* or continuing* or postacute* or "post acute*" or postdischarg* or "post discharg*" or postinfect* or "post infect*" or postviral* or "post viral*" or postvirus* or "post virus*" or "medium* term*" or mediumterm*) adj4 (sequela* or illness* or symptom* or sign* or prognos* or rehab* or convalescen* or recuperat* or followup* or "follow up*" or feature*) adj10 (covid* or coronavirus* or corona* virus* or coronovirus* or corono* virus* or coronavirinae* or corona* virinae* or Cov or "2019-nCoV*" or 2019nCoV* or "19- nCoV*" or 19nCoV* or nCoV2019* or "nCoV-2019*" or nCoV19* or "nCoV-19*" or "HCoV-19*" or HCoV19* or "HCoV-2019*" or HCoV2019* or "2019 novel*" or Ncov* or "n-cov" or "SARS-CoV-2*" or "SARSCoV-2*" or "SARSCoV2*" or "SARS-CoV2*" or SARSCov19* or "SARS-Cov19*" or "SARSCov-19*" or "SARS-Cov-19*" or SARSCov2019* or "SARS-Cov2019*" or "SARSCov-2019*" or "SARS-Cov-2019*" or SARS2* or "SARS-2*" or SARScoronavirus2* or "SARS-coronavirus-2*" or "SARScoronavirus 2*" or "SARS coronavirus2*" or SARScoronovirus2* or "SARScoronovirus-2*" or "SARScoronovirus 2*" or "SARS coronovirus2*" or "severe acute respiratory syndrome*")).ti,ab. |

| | |
|---|---|
| 6 | ((ongoing* or long* or endur* or legacy* or slow* or gradual* or protract* or lengthy* or chronic* or persist* or relaps* or remit* or remission* or residual* or delay* or prolong* or extend* or linger* or permanent* or fluctuat* or multisystem* or "multi system*" or subacute* or "sub acute*" or lasting* or continuous* or continual* or continuing* or post* or after* or follow* or "medium* term*" or mediumterm*) adj1 recover* adj10 (covid* or coronavirus* or corona* virus* or coronovirus* or corono* virus* or coronavirinae* or corona* virinae* or Cov or "2019-nCoV*" or 2019nCoV* or "19-nCoV*" or 19nCoV* or 128 38 nCoV2019* or "nCoV-2019*" or nCoV19* or "nCoV-19*" or "HCoV-19*" or HCoV19* or "HCoV-2019*" or HCoV2019* or "2019 novel*" or Ncov* or "n-cov" or "SARS-CoV-2*" or "SARSCoV-2*" or "SARSCoV2*" or "SARS-CoV2*" or SARSCov19* or "SARS-Cov19*" or "SARSCov-19*" or "SARS-Cov-19*" or SARSCov2019* or "SARS-Cov2019*" or "SARSCov-2019*" or "SARS-Cov-2019*" or SARS2* or "SARS-2*" or SARScoronavirus2* or "SARS-coronavirus-2*" or "SARScoronavirus 2*" or "SARS coronavirus2*" or SARScoronovirus2* or "SARS-coronovirus-2*" or "SARScoronovirus 2*" or "SARS coronovirus2*" or "severe acute respiratory syndrome*")).ti,ab. |
| 7 | or/1-6 |
| 8 | exp coronavirus/ |
| 9 | exp Coronavirus infections/ |
| 10 | (covid* or coronavirus* or corona* virus* or coronovirus* or corono* virus* or coronavirinae* or corona* virinae* or Cov or "2019-nCoV*" or 2019nCoV* or "19-nCoV*" or 19nCoV* or nCoV2019* or "nCoV-2019*" or nCoV19* or "nCoV-19*" or "HCoV-19*" or HCoV19* or "HCoV-2019*" or HCoV2019* or "2019 novel*" or Ncov* or "n-cov" or "SARS-CoV-2*" or "SARSCoV-2*" or "SARSCoV2*" or "SARS-CoV2*" or SARSCov19* or "SARS-Cov19*" or "SARSCov-19*" or "SARS-Cov-19*" or SARSCov2019* or "SARSCov2019*" or "SARSCov-2019*" or "SARS-Cov-2019*" or SARS2* or "SARS-2*" or SARScoronavirus2* or "SARS-coronavirus-2*" or "SARScoronavirus 2*" or "SARS coronavirus2*" or SARScoronovirus2* or "SARS-coronovirus-2*" or "SARScoronovirus 2*" or "SARS coronovirus2*" or "severe acute respiratory syndrome*").ti. |
| 11 | or/8-10 |
| 12 | "Recovery of Function"/ |
| 13 | Aftercare/ |
| 14 | rehabilitation/ |
| 15 | Activities of Daily Living/ |
| 16 | Convalescence/ |
| 17 | Long Term Adverse Effects/ |
| 18 | Physical Functional Performance/ |
| 19 | or/12-18 |
| 20 | 11 and 19 |

| | |
|---|---|
| 21 | ("long* haul*" or longhaul* or "long* tail*" or longtail* or longduration* or "long duration*" or longlast* or "long last*" or longstanding* or "long standing*").ti,ab. and (8 or 9) |
| 22 | ((recover* or nonrecover*) adj3 function* adj10 (covid* or coronavirus* or corona* virus* or coronovirus* or corono* virus* or coronavirinae* or corona* virinae* or Cov or "2019- 15 39 nCoV*" or 2019nCoV* or "19-nCoV*" or 19nCoV* or nCoV2019* or "nCoV-2019*" or nCoV19* or "nCoV-19*" or "HCoV-19*" or HCoV19* or "HCoV-2019*" or HCoV2019* or "2019 novel*" or Ncov* or "n-cov" or "SARS-CoV-2*" or "SARSCoV-2*" or "SARSCoV2*" or "SARS-CoV2*" or SARSCov19* or "SARS-Cov19*" or "SARSCov-19*" or "SARS-Cov19*" or SARSCov2019* or "SARS-Cov2019*" or "SARSCov-2019*" or "SARS-Cov2019*" or SARS2* or "SARS-2*" or SARScoronavirus2* or "SARS-coronavirus-2*" or "SARScoronavirus 2*" or "SARS coronavirus2*" or SARScoronovirus2* or "SARScoronovirus-2*" or "SARScoronovirus 2*" or "SARS coronovirus2*" or "severe acute respiratory syndrome*")).ti,ab. |
| 23 | ((postacute* or "post acute*" or postdischarg* or "post discharg*" or postinfect* or "post infect*" or postviral* or "post viral*" or postvirus* or "post virus*" or subacute* or "sub acute*") adj3 (care* or convalescen* or recuperat* or aftercare* or ambulatory* or outpatient* or "out patient*" or survivor* or survival*) adj10 (covid* or coronavirus* or corona* virus* or coronovirus* or corono* virus* or coronavirinae* or corona* virinae* or Cov or "2019-nCoV*" or 2019nCoV* or "19-nCoV*" or 19nCoV* or nCoV2019* or "nCoV2019*" or nCoV19* or "nCoV-19*" or "HCoV-19*" or HCoV19* or "HCoV-2019*" or HCoV2019* or "2019 novel*" or Ncov* or "n-cov" or "SARS-CoV-2*" or "SARSCoV-2*" or "SARSCoV2*" or "SARS-CoV2*" or SARSCov19* or "SARS-Cov19*" or "SARSCov-19*" or "SARS-Cov-19*" or SARSCov2019* or "SARS-Cov2019*" or "SARSCov-2019*" or "SARS-Cov-2019*" or SARS2* or "SARS-2*" or SARScoronavirus2* or "SARScoronavirus-2*" or "SARScoronavirus 2*" or "SARS coronavirus2*" or SARScoronovirus2* or "SARS-coronovirus-2*" or "SARScoronovirus 2*" or "SARS coronovirus2*" or "severe acute respiratory syndrome*")).ti,ab. |
| 24 | ((convalescen* or recuperat* or after* or followup* or "follow up*" or rehab*) adj1 (therap* or care*) adj10 (covid* or coronavirus* or corona* virus* or coronovirus* or corono* virus* or coronavirinae* or corona* virinae* or Cov or "2019-nCoV*" or 2019nCoV* or "19- nCoV*" or 19nCoV* or nCoV2019* or "nCoV-2019*" or nCoV19* or "nCoV-19*" or "HCoV-19*" or HCoV19* or "HCoV-2019*" or HCoV2019* or "2019 novel*" or Ncov* or "n-cov" or "SARS-CoV-2*" or "SARSCoV-2*" or "SARSCoV2*" or "SARS-CoV2*" or SARSCov19* or "SARS-Cov19*" or "SARSCov-19*" or "SARS-Cov-19*" or SARSCov2019* or "SARS-Cov2019*" or "SARSCov-2019*" or "SARS-Cov-2019*" or SARS2* or "SARS-2*" or SARScoronavirus2* or "SARS-coronavirus-2*" or "SARScoronavirus 2*" or "SARS coronavirus2*" or SARScoronovirus2* or "SARScoronovirus-2*" or "SARScoronovirus 2*" or "SARS coronovirus2*" or "severe acute respiratory syndrome*")).ti,ab. |
| 25 | ((ongoing* or endur* or long* or legacy* or slow* or gradual* or protract* or lengthy* or chronic* or persist* or relaps* or remit* or remission* or residual* or delay* or prolong* or extend* or linger* or permanent* or fluctuat* or multisystem* or "multi system*" or 58 40 nonrecover* or "non recover*" or subacute* or "sub acute*" or lasting* or continuous* or continual* or continuing* or postacute* or "post acute*" or postdischarg* or "post |

| | |
|---|---|
| | discharg*" or postinfect* or "post infect*" or postviral* or "post viral*" or postvirus* or "post virus*" or "medium* term*" or mediumterm* or adverse* or dangerous* or harmful* or indirect* or injurious* or secondary* or side effect* or undesirable* or sequela* or complication* or consequence* or effect* or event* or impact* or outcome* or reaction* or complexit* or aftercare* or impair* or problem* or issue* or rehab* or function* or perform*) adj10 ((daily* or everyday* or day* or normal* or usual*) adj1 (activit* or living* or life* or lives* or job* or work* or employ* or occupation* or hobby* or hobbies* or leisure*)) adj10 (covid* or coronavirus* or corona* virus* or coronovirus* or corono* virus* or coronavirinae* or corona* virinae* or Cov or "2019-nCoV*" or 2019nCoV* or "19- nCoV*" or 19nCoV* or nCoV2019* or "nCoV-2019*" or nCoV19* or "nCoV-19*" or "HCoV-19*" or HCoV19* or "HCoV-2019*" or HCoV2019* or "2019 novel*" or Ncov* or "n-cov" or "SARS-CoV-2*" or "SARSCoV-2*" or "SARSCoV2*" or "SARS-CoV2*" or SARSCov19* or "SARS-Cov19*" or "SARSCov-19*" or "SARS-Cov-19*" or SARSCov2019* or "SARS-Cov2019*" or "SARSCov-2019*" or "SARS-Cov-2019*" or SARS2* or "SARS-2*" or SARScoronavirus2* or "SARS-coronavirus-2*" or "SARScoronavirus 2*" or "SARS coronavirus2*" or SARScoronovirus2* or "SARScoronovirus-2*" or "SARScoronovirus 2*" or "SARS coronovirus2*" or "severe acute respiratory syndrome*")).ti,ab. |
| 26 | ((ongoing* or endur* or long* or legacy* or slow* or gradual* or protract* or lengthy* or chronic* or persist* or relaps* or remit* or remission* or residual* or delay* or prolong* or extend* or linger* or permanent* or fluctuat* or multisystem* or "multi system*" or nonrecover* or "non recover*" or subacute* or "sub acute*" or lasting* or continuous* or continual* or continuing* or postacute* or "post acute*" or postdischarg* or "post discharg*" or postinfect* or "post infect*" or postviral* or "post viral*" or postvirus* or "post virus*" or "medium* term*" or mediumterm*) adj3 ((health* or adverse* or dangerous* or harmful* or indirect* or injurious* or secondary* or side* or undesirable* or negative* or damaging* or detriment* or abnormal*) adj1 (sequela* or complication* or consequence* or effect* or event* or impact* or outcome* or reaction* or complexit* or aftercare* or impair* or problem* or issue* or symptom* or disorder*)) adj10 (covid* or coronavirus* or corona* virus* or coronovirus* or corono* virus* or coronavirinae* or corona* virinae* or Cov or "2019-nCoV*" or 2019nCoV* or "19-nCoV*" or 19nCoV* or nCoV2019* or "nCoV2019*" or nCoV19* or "nCoV-19*" or "HCoV-19*" or HCoV19* or "HCoV-2019*" or HCoV2019* or "2019 novel*" or Ncov* or "n-cov" or "SARS-CoV-2*" or "SARSCoV-2*" or "SARSCoV2*" or "SARS-CoV2*" or SARSCov19* or "SARS-Cov19*" or "SARSCov-19*" or "SARS-Cov-19*" or SARSCov2019* or "SARS-Cov2019*" or "SARSCov-2019*" or "SARS-Cov-2019*" or SARS2* or "SARS-2*" or SARScoronavirus2* or "SARS40 41 coronavirus-2*" or "SARScoronavirus 2*" or "SARS coronavirus2*" or SARScoronovirus2* or "SARS-coronovirus-2*" or "SARScoronovirus 2*" or "SARS coronovirus2*" or "severe acute respiratory syndrome*")).ti,ab. |
| 27 | ((ongoing* or endur* or long* or legacy* or slow* or gradual* or protract* or lengthy* or chronic* or persist* or relaps* or remit* or remission* or residual* or delay* or prolong* or extend* or linger* or permanent* or fluctuat* or multisystem* or "multi system*" or nonrecover* or "non recover*" or subacute* or "sub acute*" or lasting* or continuous* or continual* or continuing* or postacute* or "post acute*" or postdischarg* or "post discharg*" or |

| | |
|---|---|
| | postinfect* or "post infect*" or postviral* or "post viral*" or postvirus* or "post virus*" or "medium* term*" or mediumterm*) adj3 ((physiolog* or neuro* or cardio* or gastro* or musculo* or renal* or kidney* or cognitive* or cognition* or rheumato* or dermatol* or skin* or haematol* or blood* or autonomic* or nervous* or nervous system* or otolaryngol* or laryngol* or otolog* or cerebro* or brain* or vascular* or respirator* or lung* or pulmonary* or psycholog* or mental health* or mental* or psychiatr* or exertion* or debilit* or devitali* or enervat* or drain* or sleep* or weak* or tired* or frail* or sapp* or strength* or confusion* or letharg* or fatigue* or tired* or weariness* or exhaust* or malaise* or pain* or headache* or breathless* or breathing* or myalgia* or delirious* or delirium* or appetite* or muscle* or muscular* or fitness* or memory* or stress* or depress* or anxiety* or emotion* or cough* or fever* or temperatur* or pneumon* or conjunctivit* or throat* or pharyngit* or dyspnea* or dyspnoea* or sick* or nausea* or nauseous* or vomit* or diarrhoea* or diarrhea* or taste* or anosmia* or smell* or olfact* or sweat* or dehydrat* or pyrexi* or nasal* or nose* or mucus* or ear* or hearing* or deaf* or "brain fog*" or cardiac* or thoracic* or chest* or ischemic* or ischaemic* or heart* or liver* or hepatic* or immuno* or palpitation* or vertigo* or metabol*) adj1 (sequela* or complication* or consequence* or effect* or event* or impact* or outcome* or reaction* or complexit* or aftercare* or impair* or problem* or issue* or symptom* or disorder* or abnormal*)) adj10 (covid* or coronavirus* or corona* virus* or coronovirus* or corono* virus* or coronavirinae* or corona* virinae* or Cov or "2019-nCoV*" or 2019nCoV* or "19- nCoV*" or 19nCoV* or nCoV2019* or "nCoV-2019*" or nCoV19* or "nCoV-19*" or "HCoV-19*" or HCoV19* or "HCoV-2019*" or HCoV2019* or "2019 novel*" or Ncov* or "n-cov" or "SARS-CoV-2*" or "SARSCoV-2*" or "SARSCoV2*" or "SARS-CoV2*" or SARSCov19* or "SARS-Cov19*" or "SARSCov-19*" or "SARS-Cov-19*" or SARSCov2019* or "SARS-Cov2019*" or "SARSCov-2019*" or "SARS-Cov-2019*" or SARS2* or "SARS-2*" or SARScoronavirus2* or "SARS-coronavirus-2*" or "SARScoronavirus 2*" or "SARS coronavirus2*" or SARScoronovirus2* or "SARScoronovirus-2*" or "SARScoronovirus 2*" or "SARS coronovirus2*" or "severe acute respiratory syndrome*")).ti,ab. |
| 28 | ((physiolog* or neuro* or cardio* or gastro* or musculo* or renal* or kidney* or cognitive* 49 42 or cognition* or rheumato* or dermatol* or skin* or haematol* or blood* or autonomic* or nervous* or nervous system* or otolaryngol* or laryngol* or otolog* or cerebro* or brain* or vascular* or respirator* or lung* or pulmonary* or psycholog* or mental health* or mental* or psychiatr* or exertion* or debilit* or devitali* or enervat* or drain* or sleep* or weak* or tired* or frail* or sapp* or strength* or confusion* or letharg* or fatigue* or tired* or weariness* or exhaust* or malaise* or pain* or headache* or breathless* or breathing* or myalgia* or delirious* or delirium* or appetite* or muscle* or muscular* or fitness* or memory* or stress* or depress* or anxiety* or emotion* or cough* or fever* or temperatur* or pneumon* or conjunctivit* or throat* or pharyngit* or dyspnea* or dyspnoea* or sick* or nausea* or nauseous* or vomit* or diarrhoea* or diarrhea* or taste* or anosmia* or smell* or olfact* or sweat* or dehydrat* or pyrexi* or nasal* or nose* or mucus* or ear* or hearing* or deaf* or "brain fog*" or cardiac* or thoracic* or chest* or ischemic* or ischaemic* or heart* or liver* or hepatic* or immuno* or palpitation* or vertigo* or metabol*) adj3 (postcovid* or post covid* or postcoronavirus* or |

| | |
|---|---|
| | postcorona* virus* or post coronavirus* or post corona* virus* or postcoronovirus* or postcorono* virus* or post coronovirus* or post corono* virus* or postcoronavirinae* or postcorona* virinae* or post coronavirinae* or post corona* virinae* or postCov or post Cov or postsars* or post sars* or "post severe acute respiratory syndrome*" or postncov* or post ncov* or posthcov* or post hcov*)).ti,ab. |
| 29 | ((physiolog* or neuro* or cardio* or gastro* or musculo* or renal* or kidney* or cognitive* or cognition* or rheumato* or dermatol* or skin* or haematol* or blood* or autonomic* or nervous* or nervous system* or otolaryngol* or laryngol* or otolog* or cerebro* or brain* or vascular* or respirator* or lung* or pulmonary* or psycholog* or mental health* or mental* or psychiatr* or exertion* or debilit* or devitali* or enervat* or drain* or sleep* or weak* or tired* or frail* or sapp* or strength* or confusion* or letharg* or fatigue* or tired* or weariness* or exhaust* or malaise* or pain* or headache* or breathless* or breathing* or myalgia* or delirious* or delirium* or appetite* or muscle* or muscular* or fitness* or memory* or stress* or depress* or anxiety* or emotion* or cough* or fever* or temperatur* or pneumon* or conjunctivit* or throat* or pharyngit* or dyspnea* or dyspnoea* or sick* or nausea* or nauseous* or vomit* or diarrhoea* or diarrhea* or taste* or anosmia* or smell* or olfact* or sweat* or dehydrat* or pyrexi* or nasal* or nose* or mucus* or ear* or hearing* or deaf* or "brain fog*" or cardiac* or thoracic* or chest* or ischemic* or ischaemic* or heart* or liver* or hepatic* or immuno* or palpitation* or vertigo* or metabol*) adj1 (sequela* or complication* or consequence* or complexit*) adj10 (covid* or coronavirus* or corona* virus* or coronovirus* or corono* virus* or coronavirinae* or corona* virinae* or Cov or "2019-nCoV*" or 2019nCoV* or "19-nCoV*" or 19nCoV* or nCoV2019* or "nCoV-2019*" or nCoV19* or "nCoV-19*" or "HCoV-19*" or HCoV19* or "HCoV-2019*" or HCoV2019* or "2019 novel*" or Ncov* or "n-cov" or "527 43 SARS-CoV-2*" or "SARSCoV-2*" or "SARSCoV2*" or "SARS-CoV2*" or SARSCov19* or "SARS-Cov19*" or "SARSCov-19*" or "SARS-Cov-19*" or SARSCov2019* or "SARSCov2019*" or "SARSCov-2019*" or "SARS-Cov-2019*" or SARS2* or "SARS-2*" or SARScoronavirus2* or "SARS-coronavirus-2*" or "SARScoronavirus 2*" or "SARS coronavirus2*" or SARScoronovirus2* or "SARS-coronovirus-2*" or "SARScoronovirus 2*" or "SARS coronovirus2*" or "severe acute respiratory syndrome*")).ti,ab. |
| 30 | (((((multi or multiple* or overlap* or cluster* or numerous* or varied* or variety*) adj1 (symptom* or system* or disease* or disorder* or illness* or condition* or syndrome*)) or (multisymptom* or multisystem* or multidisease* or multidisorder* or multiillness* or multicondition* or multisyndrom*)) adj10 (covid* or coronavirus* or corona* virus* or coronovirus* or corono* virus* or coronavirinae* or corona* virinae* or Cov or "2019- nCoV*" or 2019nCoV* or "19-nCoV*" or 19nCoV* or nCoV2019* or "nCoV-2019*" or nCoV19* or "nCoV-19*" or "HCoV-19*" or HCoV19* or "HCoV-2019*" or HCoV2019* or "2019 novel*" or Ncov* or "n-cov" or "SARS-CoV-2*" or "SARSCoV-2*" or "SARSCoV2*" or "SARS-CoV2*" or SARSCov19* or "SARS-Cov19*" or "SARSCov-19*" or "SARS-Cov19*" or SARSCov2019* or "SARS-Cov2019*" or "SARSCov-2019*" or "SARS-Cov2019*" or SARS2* or "SARS-2*" or SARScoronavirus2* or "SARS-coronavirus-2*" or "SARScoronavirus 2*" or "SARS coronavirus2*" or SARScoronovirus2* or "SARScoronovirus-2*" or |

| | |
|---|---|
| | "SARScoronovirus 2*" or "SARS coronovirus2*" or "severe acute respiratory syndrome*")) not ("Multisystem* inflammatory* syndrome*" or "inflammatory* multisystem* syndrome*")).ti,ab. |
| 31 | or/20-30 |
| 32 | 7 or 31 |
| 33 | (letter or historical article or comment or editorial or news).pt. |
| 34 | 32 not 33 |
| 35 | exp animals/ or (animal* or amphib* or ape or apes or beagle* or bird or birds or boar? or bovine* or canin* or cat or cats or catfish* or cattle or chicken* or chimpanzee* or cow or cows or dog or dogs or equine or ewe? or fish or fishes or feline* or frog or frogs or gastropod* or gerbil* or goat* or "guinea-pig*" or hamster? or hare or hares or horse* or macaque* or mammal* or marmoset* or mice or monkey* or mouse or murine or ostrich* or ovine or pig or piglet* or pigs or porcine or pork* or primate* or rabbit* or rat or rats or rodent? or sheep or sow or sows or swine* or toad? or veterinar* or zebrafish* or zebra fish*).ti. |
| 36 | humans/ or (human or humans or adolescen* or baby or babies or boy or boys or child* or elderly or girl or girls* or man or mans or men or mens or patient* or people* or person or persons or senior or seniors or teen* or woman* or women* or volunteer?).mp. |
| 37 | 35 not (35 and 36) |
| 38 | 34 not 37 |
| 39 | (2021* not ("20210101" or "20210102" or "20210103" or "20210104" or "20210105" or "20210106")).dt. |
| 40 | (2021* not ("20210 1 01" or "2021 01 02" or "2021 01 03" or "2021 01 04" or "2021 01 05" or "2021 01 06")).dt,dp. |
| 41 | 39 or 40 |
| 42 | 38 and 41 |

Medline TopUp 1

Database(s): **Ovid MEDLINE(R) ALL** 1946 to April 13, 2021
Search Strategy:

| # | Searches |
|---|---|
| 1 | ((ongoing* or endur* or long* or legacy* or slow* or gradual* or protract* or lengthy* or chronic* or persist* or relaps* or remit* or remission* or residual* or delay* or prolong* or extend* or linger* or permanent* or fluctuat* or multisystem* or "multi system*" or nonrecover* or "non recover*" or subacute* or "sub acute*" or lasting* or continuous* or continual* or continuing* or postacute* or "post acute*" or postdischarg* or "post discharg*" or postinfect* or "post infect*" or postviral* or "post viral*" or postvirus* or "post virus*" or "medium* term*" or mediumterm*) adj3 ((vestibular* or endocrine* or encephalit*) adj1 (sequela* or complication* or consequence* or effect* or event* or impact* or outcome* or reaction* or complexit* or aftercare* or impair* or problem* or issue* or symptom* or disorder* or abnormal*)) adj10 (covid* or coronavirus* or corona* virus* or coronovirus* or |

| | |
|---|---|
| | corono* virus* or coronavirinae* or corona* virinae* or Cov or "2019-nCoV*" or 2019nCoV* or "19-nCoV*" or 19nCoV* or nCoV2019* or "nCoV-2019*" or nCoV19* or "nCoV-19*" or "HCoV-19*" or HCoV19* or "HCoV-2019*" or HCoV2019* or "2019 novel*" or Ncov* or "ncov" or "SARS-CoV-2*" or "SARSCoV-2*" or "SARSCoV2*" or "SARS-CoV2*" or SARSCov19* or "SARS-Cov19*" or "SARSCov-19*" or "SARS-Cov-19*" or SARSCov2019* or "SARS-Cov2019*" or "SARSCov-2019*" or "SARS-Cov-2019*" or SARS2* or "SARS-2*" or SARScoronavirus2* or "SARS-coronavirus-2*" or "SARScoronavirus 2*" or "SARS coronavirus2*" or SARScoronovirus2* or "SARScoronovirus-2*" or "SARScoronovirus 2*" or "SARS coronovirus2*" or "severe acute respiratory syndrome*")).ti,ab. |
| 2 | ((vestibular* or endocrine* or encephalit*) adj3 (postcovid* or post covid* or postcoronavirus* or postcorona* virus* or post coronavirus* or post corona* virus* or postcoronovirus* or postcorono* virus* or post coronovirus* or post corono* virus* or postcoronavirinae* or postcorona* virinae* or post coronavirinae* or post corona* virinae* or postCov or post Cov or postsars* or post sars* or "post severe acute respiratory syndrome*" or postncov* or post ncov* or posthcov* or post hcov*)).ti,ab. |
| 3 | ((vestibular* or endocrine* or encephalit*) adj1 (sequela* or complication* or consequence* or complexit*) adj10 (covid* or coronavirus* or corona* virus* or coronovirus* or corono* virus* or coronavirinae* or corona* virinae* or Cov or "2019-nCoV*" or 2019nCoV* or "19- nCoV*" or 19nCoV* or nCoV2019* or "nCoV-2019*" or nCoV19* or "nCoV-19*" or "HCoV19*" or HCoV19* or "HCoV-2019*" or HCoV2019* or "2019 novel*" or Ncov* or "n-cov" or "SARS-CoV-2*" or "SARSCoV-2*" or "SARSCoV2*" or "SARS-CoV2*" or SARSCov19* or "SARS-Cov19*" or "SARSCov-19*" or "SARS-Cov-19*" or SARSCov2019* or "SARS3 45 Cov2019*" or "SARSCov-2019*" or "SARS-Cov-2019*" or SARS2* or "SARS-2*" or SARScoronavirus2* or "SARS-coronavirus-2*" or "SARScoronavirus 2*" or "SARS coronavirus2*" or SARScoronovirus2* or "SARS-coronovirus-2*" or "SARScoronovirus 2*" or "SARS coronovirus2*" or "severe acute respiratory syndrome*")).ti,ab. |
| 4 | or/1-3 |
| 5 | (2021* not ("20210101" or "20210102" or "20210103" or "20210104" or "20210105" or "20210106")).dt. |
| 6 | (2021* not ("20210 1 01" or "2021 01 02" or "2021 01 03" or "2021 01 04" or "2021 01 05" or "2021 01 06")).dt,dp. |
| 7 | 5 or 6 |
| 8 | 4 and 7 |

Medline TopUp 2

Database(s): **Ovid MEDLINE(R) ALL** 1946 to April 13, 2021
Search Strategy:

| # | Searches |
|---|---|
| 1 | ((ongoing* or endur* or long* or legacy* or slow* or gradual* or protract* or lengthy* or chronic* or persist* or relaps* or remit* or remission* or residual* or delay* or prolong* or extend* or linger* or permanent* or fluctuat* |

| | |
|---|---|
| | or multisystem* or "multi system*" or nonrecover* or "non recover*" or subacute* or "sub acute*" or lasting* or continuous* or continual* or continuing* or postacute* or "post acute*" or postdischarg* or "post discharg*" or postinfect* or "post infect*" or postviral* or "post viral*" or postvirus* or "post virus*" or "medium* term*" or mediumterm*) adj3 ((physical* or cough* or fibrosis* or myocarditis* or Guillain* or barre* or neuralgi* or amyotroph* or thrombo* or clot* or rash* or hive* or urticari* or lymph* or stroke* or TIA or toe* or foot* or feet* or finger* or chilblain* or numb* or inflammat* or inflame* or arthralgi* or eye* or organ or organs or tingl* or sting* or burn* or bladder* or urogenit* or genitourin* or genital* or reproducti* or urinary* or joint* or tachycard* or atrial* or autoimmun* or dysautonomi* or polyneuro* or mast* or mobility* or walking* or ambulation* or energy*) adj1 (sequela* or complication* or consequence* or effect* or event* or impact* or outcome* or reaction* or complexit* or aftercare* or impair* or problem* or issue* or symptom* or disorder* or abnormal*)) adj10 (covid* or coronavirus* or corona* virus* or coronovirus* or corono* virus* or coronavirinae* or corona* virinae* or Cov or "2019-nCoV*" or 2019nCoV* or "19-nCoV*" or 19nCoV* or nCoV2019* or "nCoV-2019*" or nCoV19* or "nCoV-19*" or "HCoV-19*" or HCoV19* or "HCoV-2019*" or HCoV2019* or "2019 novel*" or Ncov* or "n-cov" or "SARSCoV-2*" or "SARSCoV-2*" or "SARSCoV2*" or "SARS-CoV2*" or SARSCov19* or "SARSCov19*" or "SARSCov-19*" or "SARS-Cov-19*" or SARSCov2019* or "SARS-Cov2019*" or "SARSCov-2019*" or "SARS-Cov-2019*" or SARS2* or "SARS-2*" or SARScoronavirus2* or "SARS-coronavirus-2*" or "SARScoronavirus 2*" or "SARS coronavirus2*" or SARScoronovirus2* or "SARS-coronovirus-2*" or "SARScoronovirus 2*" or "SARS coronovirus2*" or "severe acute respiratory syndrome*")).ti,ab. |
| 2 | ((physical* or cough* or fibrosis* or myocarditis* or Guillain* or barre* or neuralgi* or amyotroph* or thrombo* or clot* or rash* or hive* or urticari* or lymph* or stroke* or TIA or toe* or foot* or feet* or finger* or chilblain* or numb* or inflammat* or inflame* or "arthralgi* 0 46" or eye* or organ or organs or tingl* or sting* or burn* or bladder* or urogenit* or genitourin* or genital* or reproducti* or urinary* or joint* or tachycard* or atrial* or autoimmun* or dysautonomi* or polyneuro* or mast* or mobility* or walking* or ambulation* or energy*) adj1 (sequela* or complication* or consequence* or effect* or event* or impact* or outcome* or reaction* or complexit* or aftercare* or impair* or problem* or issue* or symptom* or disorder* or abnormal*) adj3 (postcovid* or post covid* or postcoronavirus* or postcorona* virus* or post coronavirus* or post corona* virus* or postcoronovirus* or postcorono* virus* or post coronovirus* or post corono* virus* or postcoronavirinae* or postcorona* virinae* or post coronavirinae* or post corona* virinae* or postCov or post Cov or postsars* or post sars* or "post severe acute respiratory syndrome*" or postncov* or post ncov* or posthcov* or post hcov*)).ti,ab. |
| 3 | ((physical* or cough* or fibrosis* or myocarditis* or Guillain* or barre* or neuralgi* or amyotroph* or thrombo* or clot* or rash* or hive* or urticari* or lymph* or stroke* or TIA or toe* or foot* or feet* or finger* or chilblain* or numb* or inflammat* or inflame* or arthralgi* or eye* or organ or organs or tingl* or sting* or burn* or bladder* or urogenit* or genitourin* or genital* or reproducti* or urinary* or joint* or tachycard* or atrial* or autoimmun* or dysautonomi* or polyneuro* or mast* or mobility* or walking* or ambulation* or energy*) adj1 (sequela* or |

| | |
|---|---|
| | complication* or consequence* or effect* or event* or impact* or outcome* or reaction* or complexit* or aftercare* or impair* or problem* or issue* or symptom* or disorder* or abnormal*) adj1 (sequela* or complication* or consequence* or complexit*) adj10 (covid* or coronavirus* or corona* virus* or coronovirus* or corono* virus* or coronavirinae* or corona* virinae* or Cov or "2019-nCoV*" or 2019nCoV* or "19- nCoV*" or 19nCoV* or nCoV2019* or "nCoV-2019*" or nCoV19* or "nCoV-19*" or "HCoV19*" or HCoV19* or "HCoV-2019*" or HCoV2019* or "2019 novel*" or Ncov* or "n-cov" or "SARS-CoV-2*" or "SARSCoV-2*" or "SARSCoV2*" or "SARS-CoV2*" or SARSCov19* or "SARS-Cov19*" or "SARSCov-19*" or "SARS-Cov-19*" or SARSCov2019* or "SARSCov2019*" or "SARSCov-2019*" or "SARS-Cov-2019*" or SARS2* or "SARS-2*" or SARScoronavirus2* or "SARS-coronavirus-2*" or "SARScoronavirus 2*" or "SARS coronavirus2*" or SARScoronovirus2* or "SARS-coronovirus-2*" or "SARScoronovirus 2*" or "SARS coronovirus2*" or "severe acute respiratory syndrome*")).ti,ab. |
| 4 | or/1-3 |
| 5 | (letter or historical article or comment or editorial or news).pt. |
| 6 | 4 not 5 |
| 7 | exp animals/ or (animal* or amphib* or ape or apes or beagle* or bird or birds or boar? or bovine* or canin* or cat or cats or catfish* or cattle or chicken* or chimpanzee* or cow or cows or dog or dogs or equine or ewe? or fish or fishes or feline* or frog or frogs or gastropod* or gerbil* or goat* or "guinea-pig*" or hamster? or hare or hares or horse* or macaque* or mammal* or marmoset* or mice or monkey* or mouse or murine or ostrich* or ovine or pig or piglet* or pigs or porcine or pork* or primate* or rabbit* or rat or rats or rodent? or sheep or sow or sows or swine* or toad? or veterinar* or zebrafish* or zebra fish*).ti. |
| 8 | humans/ or (human or humans or adolescen* or baby or babies or boy or boys or child* or elderly or girl or girls* or man or mans or men or mens or patient* or people* or person or persons or senior or seniors or teen* or woman* or women* or volunteer?).mp. |
| 9 | 7 not (7 and 8) |
| 10 | 6 not 9 |
| 11 | (2021* not ("20210101" or "20210102" or "20210103" or "20210104" or "20210105" or "20210106")).dt. |
| 12 | (2021* not ("20210 1 01" or "2021 01 02" or "2021 01 03" or "2021 01 04" or "2021 01 05" or "2021 01 06")).dp,dt. |
| 13 | or/11-12 |
| 14 | 10 and 13 |

Database(s): **APA PsycInfo** 1806 to April Week 1 2021
Search Strategy:

| # | Searches |
| --- | --- |
| 1 | (longcovid* or long covid* or longcoronavirus* or longcorona* virus* or long coronavirus* or long corona* virus* or longcoronovirus* or longcorono* virus* or long coronovirus* or long corono* virus* or longcoronavirinae* or longcorona* virinae* or long coronavirinae* or long corona* virinae* or longCov or long Cov or longsars* or long sars* or "long severe acute respiratory syndrome*" or longncov* or long ncov* or longhcov* or long hcov*).ti,ab. |
| 2 | ((long* or endur* or legacy* or slow* or gradual* or protract* or lengthy* or chronic* or persist* or relaps* or remit* or remission* or residual* or delay* or prolong* or extend* or linger* or permanent* or fluctuat* or sequela* or multisystem* or "multi system*" or nonrecover* or "non recover*" or subacute* or "sub acute*" or lasting* or continuous* or continual* or continuing* or postacute* or "post acute*" or postdischarg* or "post discharg*" or postinfect* or "post infect*" or postviral* or "post viral*" or postvirus* or "post virus*") adj1 (covid* or coronavirus* or corona* virus* or coronovirus* or corono* virus* or coronavirinae* or corona* virinae* or Cov or "2019-nCoV*" or 2019nCoV* or "19-nCoV*" or 19nCoV* or nCoV2019* or "nCoV-2019*" or nCoV19* or "nCoV-19*" or "HCoV-19*" or HCoV19* or "HCoV-2019*" or HCoV2019* or "2019 novel*" or Ncov* or "n-cov" or "SARS-CoV-2*" or "SARSCoV-2*" or "SARSCoV2*" or "SARS-CoV2*" or SARSCov19* or "SARS-Cov19*" or "SARSCov-19*" or "SARS-Cov-19*" or SARSCov2019* or "SARSCov2019*" or "SARSCov-2019*" or "SARS-Cov-2019*" or SARS2* or "SARS-2*" or SARScoronavirus2* or "SARS-coronavirus-2*" or "SARScoronavirus 2*" or "SARS coronavirus2*" or SARScoronovirus2* or "SARS-coronovirus-2*" or "SARScoronovirus 2*" or "SARS coronovirus2*" or "severe acute respiratory syndrome*")).ti,ab. |
| 3 | (("long* term*" or longterm* or "long* haul*" or longhaul* or "long* tail*" or longtail* or longduration* or "long duration*" or longlast* or "long last*" or longstanding* or "long standing*" or "medium* term*" or mediumterm*) adj3 (covid* or coronavirus* or corona* virus* or coronovirus* or corono* virus* or coronavirinae* or corona* virinae* or Cov or "2019-nCoV*" or 2019nCoV* or "19-nCoV*" or 19nCoV* or nCoV2019* or "nCoV-2019*" or nCoV19* or "nCoV-19*" or "HCoV-19*" or HCoV19* or "HCoV-2019*" or HCoV2019* or "2019 novel*" or Ncov* or "n-cov" or "SARS-CoV-2*" or "SARSCoV-2*" or "SARSCoV2*" or "SARS-CoV2*" or SARSCov19* or "SARS-Cov19*" or "SARSCov-19*" or "SARS-Cov-19*" or SARSCov2019* or "SARS-Cov2019*" or "SARSCov-2019*" or "SARS-Cov-2019*" or SARS2* or "SARS-2*" or SARScoronavirus2* or "SARS164 37 coronavirus-2*" or "SARScoronavirus 2*" or "SARS coronavirus2*" or SARScoronovirus2* or "SARS-coronovirus-2*" or "SARScoronovirus 2*" or "SARS coronovirus2*" or "severe acute respiratory syndrome*")).ti,ab. |
| 4 | ((postcovid* or post covid* or postcoronavirus* or postcorona* virus* or post coronavirus* or post corona* virus* or postcoronovirus* or postcorono* virus* or post coronovirus* or post corono* virus* or postcoronavirinae* or postcorona* virinae* or post coronavirinae* or post corona* virinae* or postCov or post Cov or postsars* or post sars* or "post severe acute respiratory syndrome*" or postncov* or post ncov* or posthcov* or post hcov*) adj3 (syndrome* or disorder* or illness* or sickness* or disease* or condition* or symptom* or sign* or prognos* or followup* or "follow up*" or feature* or comorbid* or "co morbid*" or multimorbid* or "multi morbid*" or survivor* |

| | |
|---|---|
| | or survival* or risk* or care* or convalescen* or recuperat* or aftercare* or ambulatory* or outpatient* or "out patient*")).ti,ab. |
| 5 | ((ongoing* or long* or endur* or legacy* or slow* or gradual* or protract* or lengthy* or chronic* or persist* or relaps* or remit* or remission* or residual* or delay* or prolong* or extend* or linger* or permanent* or fluctuat* or multisystem* or "multi system*" or nonrecover* or "non recover*" or subacute* or "sub acute*" or lasting* or continuous* or continual* or continuing* or postacute* or "post acute*" or postdischarg* or "post discharg*" or postinfect* or "post infect*" or postviral* or "post viral*" or postvirus* or "post virus*" or "medium* term*" or mediumterm*) adj4 (sequela* or illness* or symptom* or sign* or prognos* or rehab* or convalescen* or recuperat* or followup* or "follow up*" or feature*) adj10 (covid* or coronavirus* or corona* virus* or coronovirus* or corono* virus* or coronavirinae* or corona* virinae* or Cov or "2019-nCoV*" or 2019nCoV* or "19- nCoV*" or 19nCoV* or nCoV2019* or "nCoV-2019*" or nCoV19* or "nCoV-19*" or "HCoV-19*" or HCoV19* or "HCoV-2019*" or HCoV2019* or "2019 novel*" or Ncov* or "n-cov" or "SARS-CoV-2*" or "SARSCoV-2*" or "SARSCoV2*" or "SARS-CoV2*" or SARSCov19* or "SARS-Cov19*" or "SARSCov-19*" or "SARS-Cov-19*" or SARSCov2019* or "SARS-Cov2019*" or "SARSCov-2019*" or "SARS-Cov-2019*" or SARS2* or "SARS-2*" or SARScoronavirus2* or "SARS-coronavirus-2*" or "SARScoronavirus 2*" or "SARS coronavirus2*" or SARScoronovirus2* or "SARScoronovirus-2*" or "SARScoronovirus 2*" or "SARS coronovirus2*" or "severe acute respiratory syndrome*")).ti,ab. |
| 6 | ((ongoing* or long* or endur* or legacy* or slow* or gradual* or protract* or lengthy* or chronic* or persist* or relaps* or remit* or remission* or residual* or delay* or prolong* or extend* or linger* or permanent* or fluctuat* or multisystem* or "multi system*" or subacute* or "sub acute*" or lasting* or continuous* or continual* or continuing* or post* or after* or follow* or "medium* term*" or mediumterm*) adj1 recover* adj10 (covid* or coronavirus* or corona* virus* or coronovirus* or corono* virus* or coronavirinae* or corona* virinae* or Cov or "2019-nCoV*" or 2019nCoV* or "19-nCoV*" or 19nCoV* or 128 38 nCoV2019* or "nCoV-2019*" or nCoV19* or "nCoV-19*" or "HCoV-19*" or HCoV19* or "HCoV-2019*" or HCoV2019* or "2019 novel*" or Ncov* or "n-cov" or "SARS-CoV-2*" or "SARSCoV-2*" or "SARSCoV2*" or "SARS-CoV2*" or SARSCov19* or "SARS-Cov19*" or "SARSCov-19*" or "SARS-Cov-19*" or SARSCov2019* or "SARS-Cov2019*" or "SARSCov-2019*" or "SARS-Cov-2019*" or SARS2* or "SARS-2*" or SARScoronavirus2* or "SARS-coronavirus-2*" or "SARScoronavirus 2*" or "SARS coronavirus2*" or SARScoronovirus2* or "SARS-coronovirus-2*" or "SARScoronovirus 2*" or "SARS coronovirus2*" or "severe acute respiratory syndrome*")).ti,ab. |
| 7 | or/1-6 |
| 8 | exp Coronavirus infection/ |
| 9 | (covid* or coronavirus* or corona* virus* or coronovirus* or corono* virus* or coronavirinae* or corona* virinae* or Cov or "2019-nCoV*" or 2019nCoV* or "19-nCoV*" or 19nCoV* or nCoV2019* or "nCoV-2019*" or nCoV19* or "nCoV-19*" or "HCoV-19*" or HCoV19* or "HCoV-2019*" or HCoV2019* or "2019 novel*" or Ncov* or "n-cov" or "SARS-CoV-2*" or "SARSCoV-2*" or "SARSCoV2*" or "SARS-CoV2*" or SARSCov19* or "SARS- |

| | |
|---|---|
| | Cov19*" or "SARSCov-19*" or "SARS-Cov-19*" or SARSCov2019* or "SARSCov2019*" or "SARSCov-2019*" or "SARS-Cov-2019*" or SARS2* or "SARS-2*" or SARScoronavirus2* or "SARS-coronavirus-2*" or "SARScoronavirus 2*" or "SARS coronavirus2*" or SARScoronovirus2* or "SARS-coronovirus-2*" or "SARScoronovirus 2*" or "SARS coronovirus2*" or "severe acute respiratory syndrome*").ti. |
| 10 | 8 or 9 |
| 11 | Functional status/ |
| 12 | exp "Recovery (Disorders)"/ |
| 13 | Aftercare/ |
| 14 | rehabilitation/ |
| 15 | Activities of Daily Living/ |
| 16 | Activity level/ |
| 17 | or/11-16 |
| 18 | 10 and 17 |
| 19 | ("long* haul*" or longhaul* or "long* tail*" or longtail* or longduration* or "long duration*" or longlast* or "long last*" or longstanding* or "long standing*").ti,ab. and 8 |
| 20 | ((recover* or nonrecover*) adj3 function* adj10 (covid* or coronavirus* or corona* virus* or coronovirus* or corono* virus* or coronavirinae* or corona* virinae* or Cov or "2019- 15 39 nCoV*" or 2019nCoV* or "19-nCoV*" or 19nCoV* or nCoV2019* or "nCoV-2019*" or nCoV19* or "nCoV-19*" or "HCoV-19*" or HCoV19* or "HCoV-2019*" or HCoV2019* or "2019 novel*" or Ncov* or "n-cov" or "SARS-CoV-2*" or "SARSCoV-2*" or "SARSCoV2*" or "SARS-CoV2*" or SARSCov19* or "SARS-Cov19*" or "SARSCov-19*" or "SARS-Cov19*" or SARSCov2019* or "SARS-Cov2019*" or "SARSCov-2019*" or "SARS-Cov2019*" or SARS2* or "SARS-2*" or SARScoronavirus2* or "SARS-coronavirus-2*" or "SARScoronavirus 2*" or "SARS coronavirus2*" or SARScoronovirus2* or "SARScoronovirus-2*" or "SARScoronovirus 2*" or "SARS coronovirus2*" or "severe acute respiratory syndrome*")).ti,ab. |
| 21 | ((postacute* or "post acute*" or postdischarg* or "post discharg*" or postinfect* or "post infect*" or postviral* or "post viral*" or postvirus* or "post virus*" or subacute* or "sub acute*") adj3 (care* or convalescen* or recuperat* or aftercare* or ambulatory* or outpatient* or "out patient*" or survivor* or survival*) adj10 (covid* or coronavirus* or corona* virus* or coronovirus* or corono* virus* or coronavirinae* or corona* virinae* or Cov or "2019-nCoV*" or 2019nCoV* or "19-nCoV*" or 19nCoV* or nCoV2019* or "nCoV2019*" or nCoV19* or "nCoV-19*" or "HCoV-19*" or HCoV19* or "HCoV-2019*" or HCoV2019* or "2019 novel*" or Ncov* or "n-cov" or "SARS-CoV-2*" or "SARSCoV-2*" or "SARSCoV2*" or "SARS-CoV2*" or SARSCov19* or "SARS-Cov19*" or "SARSCov-19*" or "SARS-Cov-19*" or SARSCov2019* or "SARS-Cov2019*" or "SARSCov-2019*" or "SARS-Cov-2019*" or SARS2* or "SARS-2*" or SARScoronavirus2* or "SARScoronavirus-2*" or "SARScoronavirus |

| | |
|---|---|
| | 2*" or "SARS coronavirus2*" or SARScoronovirus2* or "SARS-coronovirus-2*" or "SARScoronovirus 2*" or "SARS coronovirus2*" or "severe acute respiratory syndrome*")).ti,ab. |
| 22 | ((convalescen* or recuperat* or after* or followup* or "follow up*" or rehab*) adj1 (therap* or care*) adj10 (covid* or coronavirus* or corona* virus* or coronovirus* or corono* virus* or coronavirinae* or corona* virinae* or Cov or "2019-nCoV*" or 2019nCoV* or "19- nCoV*" or 19nCoV* or nCoV2019* or "nCoV-2019*" or nCoV19* or "nCoV-19*" or "HCoV-19*" or HCoV19* or "HCoV-2019*" or HCoV2019* or "2019 novel*" or Ncov* or "n-cov" or "SARS-CoV-2*" or "SARSCoV-2*" or "SARSCoV2*" or "SARS-CoV2*" or SARSCov19* or "SARS-Cov19*" or "SARSCov-19*" or "SARS-Cov-19*" or SARSCov2019* or "SARS-Cov2019*" or "SARSCov-2019*" or "SARS-Cov-2019*" or SARS2* or "SARS-2*" or SARScoronavirus2* or "SARS-coronavirus-2*" or "SARScoronavirus 2*" or "SARS coronavirus2*" or SARScoronovirus2* or "SARScoronovirus-2*" or "SARScoronovirus 2*" or "SARS coronovirus2*" or "severe acute respiratory syndrome*")).ti,ab. |
| 23 | ((ongoing* or endur* or long* or legacy* or slow* or gradual* or protract* or lengthy* or chronic* or persist* or relaps* or remit* or remission* or residual* or delay* or prolong* or extend* or linger* or permanent* or fluctuat* or multisystem* or "multi system*" or 58 40 nonrecover* or "non recover*" or subacute* or "sub acute*" or lasting* or continuous* or continual* or continuing* or postacute* or "post acute*" or postdischarg* or "post discharg*" or postinfect* or "post infect*" or postviral* or "post viral*" or postvirus* or "post virus*" or "medium* term*" or mediumterm* or adverse* or dangerous* or harmful* or indirect* or injurious* or secondary* or side effect* or undesirable* or sequela* or complication* or consequence* or effect* or event* or impact* or outcome* or reaction* or complexit* or aftercare* or impair* or problem* or issue* or rehab* or function* or perform*) adj10 ((daily* or everyday* or day* or normal* or usual*) adj1 (activit* or living* or life* or lives* or job* or work* or employ* or occupation* or hobby* or hobbies* or leisure*)) adj10 (covid* or coronavirus* or corona* virus* or coronovirus* or corono* virus* or coronavirinae* or corona* virinae* or Cov or "2019-nCoV*" or 2019nCoV* or "19- nCoV*" or 19nCoV* or nCoV2019* or "nCoV-2019*" or nCoV19* or "nCoV-19*" or "HCoV-19*" or HCoV19* or "HCoV-2019*" or HCoV2019* or "2019 novel*" or Ncov* or "n-cov" or "SARS-CoV-2*" or "SARSCoV-2*" or "SARSCoV2*" or "SARS-CoV2*" or SARSCov19* or "SARS-Cov19*" or "SARSCov-19*" or "SARS-Cov-19*" or SARSCov2019* or "SARS-Cov2019*" or "SARSCov-2019*" or "SARS-Cov-2019*" or SARS2* or "SARS-2*" or SARScoronavirus2* or "SARS-coronavirus-2*" or "SARScoronavirus 2*" or "SARS coronavirus2*" or SARScoronovirus2* or "SARScoronovirus-2*" or "SARScoronovirus 2*" or "SARS coronovirus2*" or "severe acute respiratory syndrome*")).ti,ab. |
| 24 | ((ongoing* or endur* or long* or legacy* or slow* or gradual* or protract* or lengthy* or chronic* or persist* or relaps* or remit* or remission* or residual* or delay* or prolong* or extend* or linger* or permanent* or fluctuat* or multisystem* or "multi system*" or nonrecover* or "non recover*" or subacute* or "sub acute*" or lasting* or continuous* or continual* or continuing* or postacute* or "post acute*" or postdischarg* or "post discharg*" or postinfect* or "post infect*" or postviral* or "post viral*" or postvirus* or "post virus*" or "medium* term*" or mediumterm*) adj3 ((health* or adverse* or dangerous* or harmful* or indirect* or injurious* or secondary* or |

| | |
|---|---|
| | side* or undesirable* or negative* or damaging* or detriment* or abnormal*) adj1 (sequela* or complication* or consequence* or effect* or event* or impact* or outcome* or reaction* or complexit* or aftercare* or impair* or problem* or issue* or symptom* or disorder*)) adj10 (covid* or coronavirus* or corona* virus* or coronovirus* or corono* virus* or coronavirinae* or corona* virinae* or Cov or "2019-nCoV*" or 2019nCoV* or "19-nCoV*" or 19nCoV* or nCoV2019* or "nCoV2019*" or nCoV19* or "nCoV-19*" or "HCoV-19*" or HCoV19* or "HCoV-2019*" or HCoV2019* or "2019 novel*" or Ncov* or "n-cov" or "SARS-CoV-2*" or "SARSCoV-2*" or "SARSCoV2*" or "SARS-CoV2*" or SARSCov19* or "SARS-Cov19*" or "SARSCov-19*" or "SARS-Cov-19*" or SARSCov2019* or "SARS-Cov2019*" or "SARSCov-2019*" or "SARS-Cov-2019*" or SARS2* or "SARS-2*" or SARScoronavirus2* or "SARS40 41 coronavirus-2*" or "SARScoronavirus 2*" or "SARS coronavirus2*" or SARScoronovirus2* or "SARS-coronovirus-2*" or "SARScoronovirus 2*" or "SARS coronovirus2*" or "severe acute respiratory syndrome*")).ti,ab. |
| 25 | ((ongoing* or endur* or long* or legacy* or slow* or gradual* or protract* or lengthy* or chronic* or persist* or relaps* or remit* or remission* or residual* or delay* or prolong* or extend* or linger* or permanent* or fluctuat* or multisystem* or "multi system*" or nonrecover* or "non recover*" or subacute* or "sub acute*" or lasting* or continuous* or continual* or continuing* or postacute* or "post acute*" or postdischarg* or "post discharg*" or postinfect* or "post infect*" or postviral* or "post viral*" or postvirus* or "post virus*" or "medium* term*" or mediumterm*) adj3 ((physiolog* or neuro* or cardio* or gastro* or musculo* or renal* or kidney* or cognitive* or cognition* or rheumato* or dermatol* or skin* or haematol* or blood* or autonomic* or nervous* or nervous system* or otolaryngol* or laryngol* or otolog* or cerebro* or brain* or vascular* or respirator* or lung* or pulmonary* or psycholog* or mental health* or mental* or psychiatr* or exertion* or debilit* or devitali* or enervat* or drain* or sleep* or weak* or tired* or frail* or sapp* or strength* or confusion* or letharg* or fatigue* or tired* or weariness* or exhaust* or malaise* or pain* or headache* or breathless* or breathing* or myalgia* or delirious* or delirium* or appetite* or muscle* or muscular* or fitness* or memory* or stress* or depress* or anxiety* or emotion* or cough* or fever* or temperatur* or pneumon* or conjunctivit* or throat* or pharyngit* or dyspnea* or dyspnoea* or sick* or nausea* or nauseous* or vomit* or diarrhoea* or diarrhea* or taste* or anosmia* or smell* or olfact* or sweat* or dehydrat* or pyrexi* or nasal* or nose* or mucus* or ear* or hearing* or deaf* or "brain fog*" or cardiac* or thoracic* or chest* or ischemic* or ischaemic* or heart* or liver* or hepatic* or immuno* or palpitation* or vertigo* or metabol*) adj1 (sequela* or complication* or consequence* or effect* or event* or impact* or outcome* or reaction* or complexit* or aftercare* or impair* or problem* or issue* or symptom* or disorder* or abnormal*)) adj10 (covid* or coronavirus* or corona* virus* or coronovirus* or corono* virus* or coronavirinae* or corona* virinae* or Cov or "2019-nCoV*" or 2019nCoV* or "19- nCoV*" or 19nCoV* or nCoV2019* or "nCoV-2019*" or nCoV19* or "nCoV-19*" or "HCoV-19*" or HCoV19* or "HCoV-2019*" or HCoV2019* or "2019 novel*" or Ncov* or "n-cov" or "SARS-CoV-2*" or "SARSCoV-2*" or "SARSCoV2*" or "SARS-CoV2*" or SARSCov19* or "SARS-Cov19*" or "SARSCov-19*" or "SARS-Cov-19*" or SARSCov2019* or "SARS-Cov2019*" or "SARSCov-2019*" or "SARS-Cov-2019*" or SARS2* or "SARS-2*" or |

| | |
|---|---|
| | SARScoronavirus2* or "SARS-coronavirus-2*" or "SARScoronavirus 2*" or "SARS coronavirus2*" or SARScoronovirus2* or "SARScoronovirus-2*" or "SARScoronovirus 2*" or "SARS coronovirus2*" or "severe acute respiratory syndrome*")).ti,ab. |
| 26 | ((physiolog* or neuro* or cardio* or gastro* or musculo* or renal* or kidney* or cognitive* 49 42 or cognition* or rheumato* or dermatol* or skin* or haematol* or blood* or autonomic* or nervous* or nervous system* or otolaryngol* or laryngol* or otolog* or cerebro* or brain* or vascular* or respirator* or lung* or pulmonary* or psycholog* or mental health* or mental* or psychiatr* or exertion* or debilit* or devitali* or enervat* or drain* or sleep* or weak* or tired* or frail* or sapp* or strength* or confusion* or letharg* or fatigue* or tired* or weariness* or exhaust* or malaise* or pain* or headache* or breathless* or breathing* or myalgia* or delirious* or delirium* or appetite* or muscle* or muscular* or fitness* or memory* or stress* or depress* or anxiety* or emotion* or cough* or fever* or temperatur* or pneumon* or conjunctivit* or throat* or pharyngit* or dyspnea* or dyspnoea* or sick* or nausea* or nauseous* or vomit* or diarrhoea* or diarrhea* or taste* or anosmia* or smell* or olfact* or sweat* or dehydrat* or pyrexi* or nasal* or nose* or mucus* or ear* or hearing* or deaf* or "brain fog*" or cardiac* or thoracic* or chest* or ischemic* or ischaemic* or heart* or liver* or hepatic* or immuno* or palpitation* or vertigo* or metabol*) adj3 (postcovid* or post covid* or postcoronavirus* or postcorona* virus* or post coronavirus* or post corona* virus* or postcoronovirus* or postcorono* virus* or post coronovirus* or post corono* virus* or postcoronavirinae* or postcorona* virinae* or post coronavirinae* or post corona* virinae* or postCov or post Cov or postsars* or post sars* or "post severe acute respiratory syndrome*" or postncov* or post ncov* or posthcov* or post hcov*)).ti,ab. |
| 27 | ((physiolog* or neuro* or cardio* or gastro* or musculo* or renal* or kidney* or cognitive* or cognition* or rheumato* or dermatol* or skin* or haematol* or blood* or autonomic* or nervous* or nervous system* or otolaryngol* or laryngol* or otolog* or cerebro* or brain* or vascular* or respirator* or lung* or pulmonary* or psycholog* or mental health* or mental* or psychiatr* or exertion* or debilit* or devitali* or enervat* or drain* or sleep* or weak* or tired* or frail* or sapp* or strength* or confusion* or letharg* or fatigue* or tired* or weariness* or exhaust* or malaise* or pain* or headache* or breathless* or breathing* or myalgia* or delirious* or delirium* or appetite* or muscle* or muscular* or fitness* or memory* or stress* or depress* or anxiety* or emotion* or cough* or fever* or temperatur* or pneumon* or conjunctivit* or throat* or pharyngit* or dyspnea* or dyspnoea* or sick* or nausea* or nauseous* or vomit* or diarrhoea* or diarrhea* or taste* or anosmia* or smell* or olfact* or sweat* or dehydrat* or pyrexi* or nasal* or nose* or mucus* or ear* or hearing* or deaf* or "brain fog*" or cardiac* or thoracic* or chest* or ischemic* or ischaemic* or heart* or liver* or hepatic* or immuno* or palpitation* or vertigo* or metabol*) adj1 (sequela* or complication* or consequence* or complexit*) adj10 (covid* or coronavirus* or corona* virus* or coronovirus* or corono* virus* or coronavirinae* or corona* virinae* or Cov or "2019-nCoV*" or 2019nCoV* or "19-nCoV*" or 19nCoV* or nCoV2019* or "nCoV-2019*" or nCoV19* or "nCoV-19*" or "HCoV-19*" or HCoV19* or "HCoV-2019*" or HCoV2019* or "2019 novel*" or Ncov* or "n-cov" or "527 43 SARS-CoV-2*" or "SARSCoV-2*" or "SARSCoV2*" or "SARS-CoV2*" or |

| | |
|---|---|
| | SARSCov19* or "SARS-Cov19*" or "SARSCov-19*" or "SARS-Cov-19*" or SARSCov2019* or "SARSCov2019*" or "SARSCov-2019*" or "SARS-Cov-2019*" or SARS2* or "SARS-2*" or SARScoronavirus2* or "SARS-coronavirus-2*" or "SARScoronavirus 2*" or "SARS coronavirus2*" or SARScoronovirus2* or "SARS-coronovirus-2*" or "SARScoronovirus 2*" or "SARS coronovirus2*" or "severe acute respiratory syndrome*")).ti,ab. |
| 28 | (((((multi or multiple* or overlap* or cluster* or numerous* or varied* or variety*) adj1 (symptom* or system* or disease* or disorder* or illness* or condition* or syndrome*)) or (multisymptom* or multisystem* or multidisease* or multidisorder* or multiillness* or multicondition* or multisyndrom*)) adj10 (covid* or coronavirus* or corona* virus* or coronovirus* or corono* virus* or coronavirinae* or corona* virinae* or Cov or "2019- nCoV*" or 2019nCoV* or "19-nCoV*" or 19nCoV* or nCoV2019* or "nCoV-2019*" or nCoV19* or "nCoV-19*" or "HCoV-19*" or HCoV19* or "HCoV-2019*" or HCoV2019* or "2019 novel*" or Ncov* or "n-cov" or "SARS-CoV-2*" or "SARSCoV-2*" or "SARSCoV2*" or "SARS-CoV2*" or SARSCov19* or "SARS-Cov19*" or "SARSCov-19*" or "SARS-Cov19*" or SARSCov2019* or "SARS-Cov2019*" or "SARSCov-2019*" or "SARS-Cov2019*" or SARS2* or "SARS-2*" or SARScoronavirus2* or "SARS-coronavirus-2*" or "SARScoronavirus 2*" or "SARS coronavirus2*" or SARScoronovirus2* or "SARScoronovirus-2*" or "SARScoronovirus 2*" or "SARS coronovirus2*" or "severe acute respiratory syndrome*")) not ("Multisystem* inflammatory* syndrome*" or "inflammatory* multisystem* syndrome*")).ti,ab. |
| 29 | or/18-28 |
| 30 | 7 or 29 |
| 31 | limit 30 to ("column/opinion" or editorial or letter) |
| 32 | 30 not 31 |
| 33 | (2021* not ("20210101" or "20210102" or "20210103" or "20210104" or "20210105")).up. |
| 34 | 32 and 33 |

#### PsychInfo Top Up 1

Database(s): **APA PsycInfo** 1806 to April Week 1 2021
Search Strategy:

| # | Searches |
|---|---|
| 1 | ((ongoing* or endur* or long* or legacy* or slow* or gradual* or protract* or lengthy* or chronic* or persist* or relaps* or remit* or remission* or residual* or delay* or prolong* or extend* or linger* or permanent* or fluctuat* or multisystem* or "multi system*" or nonrecover* or "non recover*" or subacute* or "sub acute*" or lasting* or continuous* or continual* or continuing* or postacute* or "post acute*" or postdischarg* or "post discharg*" or postinfect* or "post infect*" or postviral* or "post viral*" or postvirus* or "post virus*" or "medium* term*" or mediumterm*) adj3 ((vestibular* or endocrine* or encephalit*) adj1 (sequela* or complication* or consequence* |

| | |
|---|---|
| | or effect* or event* or impact* or outcome* or reaction* or complexit* or aftercare* or impair* or problem* or issue* or symptom* or disorder* or abnormal*)) adj10 (covid* or coronavirus* or corona* virus* or coronovirus* or corono* virus* or coronavirinae* or corona* virinae* or Cov or "2019-nCoV*" or 2019nCoV* or "19-nCoV*" or 19nCoV* or nCoV2019* or "nCoV-2019*" or nCoV19* or "nCoV-19*" or "HCoV-19*" or HCoV19* or "HCoV-2019*" or HCoV2019* or "2019 novel*" or Ncov* or "ncov" or "SARS-CoV-2*" or "SARSCoV-2*" or "SARSCoV2*" or "SARS-CoV2*" or SARSCov19* or "SARS-Cov19*" or "SARSCov-19*" or "SARS-Cov-19*" or SARSCov2019* or "SARS-Cov2019*" or "SARSCov-2019*" or "SARS-Cov-2019*" or SARS2* or "SARS-2*" or SARScoronavirus2* or "SARS-coronavirus-2*" or "SARScoronavirus 2*" or "SARS coronavirus2*" or SARScoronovirus2* or "SARScoronovirus-2*" or "SARScoronovirus 2*" or "SARS coronovirus2*" or "severe acute respiratory syndrome*")).ti,ab. |
| 2 | ((vestibular* or endocrine* or encephalit*) adj3 (postcovid* or post covid* or postcoronavirus* or postcorona* virus* or post coronavirus* or post corona* virus* or postcoronovirus* or postcorono* virus* or post coronovirus* or post corono* virus* or postcoronavirinae* or postcorona* virinae* or post coronavirinae* or post corona* virinae* or postCov or post Cov or postsars* or post sars* or "post severe acute respiratory syndrome*" or postncov* or post ncov* or posthcov* or post hcov*)).ti,ab. |
| 3 | ((vestibular* or endocrine* or encephalit*) adj1 (sequela* or complication* or consequence* or complexit*) adj10 (covid* or coronavirus* or corona* virus* or coronovirus* or corono* virus* or coronavirinae* or corona* virinae* or Cov or "2019-nCoV*" or 2019nCoV* or "19- nCoV*" or 19nCoV* or nCoV2019* or "nCoV-2019*" or nCoV19* or "nCoV-19*" or "HCoV19*" or HCoV19* or "HCoV-2019*" or HCoV2019* or "2019 novel*" or Ncov* or "n-cov" or "SARS-CoV-2*" or "SARSCoV-2*" or "SARSCoV2*" or "SARS-CoV2*" or SARSCov19* or "SARS-Cov19*" or "SARSCov-19*" or "SARS-Cov-19*" or SARSCov2019* or "SARS3 45 Cov2019*" or "SARSCov-2019*" or "SARS-Cov-2019*" or SARS2* or "SARS-2*" or SARScoronavirus2* or "SARS-coronavirus-2*" or "SARScoronavirus 2*" or "SARS coronavirus2*" or SARScoronovirus2* or "SARS-coronovirus-2*" or "SARScoronovirus 2*" or "SARS coronovirus2*" or "severe acute respiratory syndrome*")).ti,ab. |
| 4 | or/1-3 |
| 5 | (2021* not ("20210101" or "20210102" or "20210103" or "20210104" or "20210105")).up. |
| 6 | 4 and 5 |

Database(s): **APA PsycInfo** 1806 to April Week 1 2021
Search Strategy:

| # | Searches |
|---|---|
| 1 | ((ongoing* or endur* or long* or legacy* or slow* or gradual* or protract* or lengthy* or chronic* or persist* or relaps* or remit* or remission* or residual* or delay* or prolong* or extend* or linger* or permanent* or fluctuat* or multisystem* or "multi system*" or nonrecover* or "non recover*" or subacute* or "sub acute*" or lasting* or continuous* or continual* or continuing* or postacute* or "post acute*" or postdischarg* or "post discharg*" or postinfect* or "post infect*" or postviral* or "post viral*" or postvirus* or "post virus*" or "medium* term*" or |

| | |
|---|---|
| | mediumterm*) adj3 ((physical* or cough* or fibrosis* or myocarditis* or Guillain* or barre* or neuralgi* or amyotroph* or thrombo* or clot* or rash* or hive* or urticari* or lymph* or stroke* or TIA or toe* or foot* or feet* or finger* or chilblain* or numb* or inflammat* or inflame* or arthralgi* or eye* or organ or organs or tingl* or sting* or burn* or bladder* or urogenit* or genitourin* or genital* or reproducti* or urinary* or joint* or tachycard* or atrial* or autoimmun* or dysautonomi* or polyneuro* or mast* or mobility* or walking* or ambulation* or energy*) adj1 (sequela* or complication* or consequence* or effect* or event* or impact* or outcome* or reaction* or complexit* or aftercare* or impair* or problem* or issue* or symptom* or disorder* or abnormal*)) adj10 (covid* or coronavirus* or corona* virus* or coronovirus* or corono* virus* or coronavirinae* or corona* virinae* or Cov or "2019-nCoV*" or 2019nCoV* or "19-nCoV*" or 19nCoV* or nCoV2019* or "nCoV-2019*" or nCoV19* or "nCoV-19*" or "HCoV-19*" or HCoV19* or "HCoV-2019*" or HCoV2019* or "2019 novel*" or Ncov* or "n-cov" or "SARSCoV-2*" or "SARSCoV-2*" or "SARSCoV2*" or "SARS-CoV2*" or SARSCov19* or "SARSCov19*" or "SARSCov-19*" or "SARS-Cov-19*" or SARSCov2019* or "SARS-Cov2019*" or "SARSCov-2019*" or "SARS-Cov-2019*" or SARS2* or "SARS-2*" or SARScoronavirus2* or "SARS-coronavirus-2*" or "SARScoronavirus 2*" or "SARS coronavirus2*" or SARScoronovirus2* or "SARS-coronovirus-2*" or "SARScoronovirus 2*" or "SARS coronovirus2*" or "severe acute respiratory syndrome*")).ti,ab. |
| 2 | ((physical* or cough* or fibrosis* or myocarditis* or Guillain* or barre* or neuralgi* or amyotroph* or thrombo* or clot* or rash* or hive* or urticari* or lymph* or stroke* or TIA or toe* or foot* or feet* or finger* or chilblain* or numb* or inflammat* or inflame* or "arthralgi* 0 46" or eye* or organ or organs or tingl* or sting* or burn* or bladder* or urogenit* or genitourin* or genital* or reproducti* or urinary* or joint* or tachycard* or atrial* or autoimmun* or dysautonomi* or polyneuro* or mast* or mobility* or walking* or ambulation* or energy*) adj1 (sequela* or complication* or consequence* or effect* or event* or impact* or outcome* or reaction* or complexit* or aftercare* or impair* or problem* or issue* or symptom* or disorder* or abnormal*) adj3 (postcovid* or post covid* or postcoronavirus* or postcorona* virus* or post coronavirus* or post corona* virus* or postcoronovirus* or postcorono* virus* or post coronovirus* or post corono* virus* or postcoronavirinae* or postcorona* virinae* or post coronavirinae* or post corona* virinae* or postCov or post Cov or postsars* or post sars* or "post severe acute respiratory syndrome*" or postncov* or post ncov* or posthcov* or post hcov*)).ti,ab. |
| 3 | ((physical* or cough* or fibrosis* or myocarditis* or Guillain* or barre* or neuralgi* or amyotroph* or thrombo* or clot* or rash* or hive* or urticari* or lymph* or stroke* or TIA or toe* or foot* or feet* or finger* or chilblain* or numb* or inflammat* or inflame* or arthralgi* or eye* or organ or organs or tingl* or sting* or burn* or bladder* or urogenit* or genitourin* or genital* or reproducti* or urinary* or joint* or tachycard* or atrial* or autoimmun* or dysautonomi* or polyneuro* or mast* or mobility* or walking* or ambulation* or energy*) adj1 (sequela* or complication* or consequence* or effect* or event* or impact* or outcome* or reaction* or complexit* or aftercare* or impair* or problem* or issue* or symptom* or disorder* or abnormal*) adj1 (sequela* or complication* or consequence* or complexit*) adj10 (covid* or coronavirus* or corona* virus* or coronovirus* or corono* virus* or coronavirinae* or corona* virinae* or Cov or "2019-nCoV*" or 2019nCoV* or "19- nCoV*" or |

| | |
|---|---|
| | 19nCoV* or nCoV2019* or "nCoV-2019*" or nCoV19* or "nCoV-19*" or "HCoV19*" or HCoV19* or "HCoV-2019*" or HCoV2019* or "2019 novel*" or Ncov* or "n-cov" or "SARS-CoV-2*" or "SARSCoV-2*" or "SARSCoV2*" or "SARS-CoV2*" or SARSCov19* or "SARS-Cov19*" or "SARSCov-19*" or "SARS-Cov-19*" or SARSCov2019* or "SARSCov2019*" or "SARSCov-2019*" or "SARS-Cov-2019*" or SARS2* or "SARS-2*" or SARScoronavirus2* or "SARS-coronavirus-2*" or "SARScoronavirus 2*" or "SARS coronavirus2*" or SARScoronovirus2* or "SARS-coronovirus-2*" or "SARScoronovirus 2*" or "SARS coronovirus2*" or "severe acute respiratory syndrome*")).ti,ab. |
| 4 | or/1-3 |
| 5 | (2021* not ("20210101" or "20210102" or "20210103" or "20210104" or "20210105")).up. |
| 6 | 4 and 5 |

#### Grey Literature

1. **Cochrane COVID-19 Study Register**

URL https://covid-19.cochrane.org/

| | **Search statement** |
|---|---|
| Search 1 | longcovid* or "long covid*" or longcoronavirus* or longcoronavirinae or "long coronavirinae" or longCov or "long Cov" or longsars or "long sars" |
| Search 2 | “long severe acute respiratory syndrome” |
| Search 3 | Sequelae |
| Search 4 | nonrecover* or "non recover*" or subacute* or "sub acute*" or postacute* or "post acute*" or postdischarg* or "post discharg*" or postinfect* or "post infect*" or postviral* or "post viral*" or postvirus* or "post virus*" |
| Search 5 | postcovid* or "post covid*" or postcoronavirus* or postcoronavirinae or "post coronavirinae" or postCov or "post Cov" or postsars or "post sars" |
| Search 6 | “ongoing symptom” or “persistent symptom” |
| Search 7 | "after care" or aftercare |

| | |
|---|---|
| Search 8 | rehabilitation or recuperation or convalescence or recovery AND ongoing |
| Search 9 | "daily activities" |
| Search 10 | "functional status" |

2. **COVID-END**
   https://www.mcmasterforum.org/networks/covid-end
   Look at the inventory of "evidence about clinical management" and "evidence about public health measures" for any syntheses on the topic of long-COVID

3. **CADTH COVID-19 Grey literature resources**
   https://covid.cadth.ca/literature-searching-tools/cadth-covid-19-grey-literature-resources/?utm_source=CONS+List&utm_campaign=7627559cc1-pCODR-pending-08-02-2019-Lonsurf_COPY_01&utm_medium=email&utm_term=0_f3b3313866-7627559cc1-263023725#rapid-reviews
   - Review all articles in the following databases for anything on the topic of long-COVID
     i. Centre for Evidence-based Medicine – Oxford COVID-19 Evidence Service
        https://www.cebm.net/oxford-covid-19-evidence-service/
     ii. CADTH COVID-19 pandemic database
        https://covid.cadth.ca/screening-and-testing/?s=&asp_active=1&p_asid=3&p_asp_data=1&current_page_id=989&qtranslate_lang=0&polylang_lang=en&filters_changed=1&filters_initial=0&asp_gen%5B%5D=title&asp_gen%5B%5D=content&asp_gen%5B%5D=excerpt&customset%5B%5D=post&termset%5Bcategory%5D%5B%5D=27
        - Select "screening and testing" and "treatment" filter categories
     iii. National COVID-19 Clinical Evidence Task Force
        https://covid19evidence.net.au/#clinical-flowcharts
        - Search "living guidelines", "Clinical flowcharts" and "evidence under review" for any articles related to long-COVID. Review the reference lists for any relevant articles.

4. **Cochrane COVID Review bank**
   https://covidreviews.cochrane.org/search/site
   Review all articles in the following sites and obtain citations for anything on the topic of long-COVID

5. **BMJ Best Practice**
   Obtain references used in this page: https://bestpractice.bmj.com/topics/en-gb/3000168

### Appendix 2 - Screening Forms

#### Title and Abstract Screening Form

1) **Does the title and/or abstract include information on people who were evaluated for long-term effects after COVID-19 diagnosis?**
    a) Yes *[include]*
    b) No *[exclude]*
    c) Cannot determine *[include]*

*If [exclude]*, form complete; excluded from review.
*If [include]*, go to next question.

2) **Is the article written in English or French?**
    a. Yes *[include]*
    b. No *[exclude]*
    c. Cannot determine *[include]*

*If [exclude]*, form complete; excluded from review.
*If [include],* go to next question.

3) **Did the study include involve 50+ participants?**
    a. Yes *[include]*
    b. No *[exclude]*
    c. Cannot determine *[include]*

*If [exclude]*, form complete; excluded from review.
*If [include],* go to next question.

4) **Select the type of study/research conducted or type of article:**
    a) RCT study *[include]*
    b) Case-control, Cohort Cross-sectional, longitudinal or case series *[include]*
    c) Review (including narrative, scoping, systematic or rapid review) *[exclude]*
    d) Editorial, commentary or opinion article *[exclude]*
    e) Other *[exclude]*
    f) Cannot determine *[include]*

*If [exclude]*, form complete; excluded from review. However, primary studies from reviews (option c) that are excluded at this stage will be added to the list of citations to be screened after deduplication.

*If [include]*, form complete and include for Full text screening.

#### Full Text Screening Form

1) **Does the article include information on people who were evaluated for outcomes 4 or more weeks (or 28+ days) after COVID-19 diagnosis?**
   a) Yes *[include]*
   b) No *[exclude]*
   c) Cannot determine *[include]* *(only select if article is not in English or French)*

*If [exclude]*, form complete; excluded from review.
*If [include]*, go to next question.

2) **Is the article written in English or French?**
   a) Yes *[include]*
   b) No *[exclude]*

*If [exclude]*, form complete; excluded from review.
*If [include],* go to next question.

3) **Select the type of study/research conducted or type of article:**
   g) Primary published study involving 50+ participants with COVID-19 (e.g. RCT, Case-control, Cohort Cross-sectional, longitudinal or case series) *[include]*
   h) Primary published study involving <50 participants with COVID-19 (e.g. RCT, Case-control, Cohort Cross-sectional, longitudinal, case series or case report) *[exclude]*
   i) Review (narrative, scoping, rapid, or systematic) *[exclude]*
   j) Editorial, commentary or opinion article *[exclude]*
   k) Other (e.g. protocol, abstract, pre-print/non-peer reviewed article, guideline, conference summary) *[exclude]*

*If [exclude]*, form complete; excluded from review.
*If [include],* go to next question.

4) **Did the study collect data on participants with COVID-19 for any outcomes of interest: symptoms, sequelae or difficulties carrying out usual activities?**
   a) Yes *[include]* [add comment box here]
   b) No (e.g. only collected mortality or immunity outcomes) *[exclude]* [add comment box here]

*If [exclude]*, form complete; excluded from review.
*If [include],* go to next question.

   **a) Were the data on any of these outcomes of interest available separately for the period of interest (4 or more weeks, 28+ days)**
   a) Yes, and data was available for 50+ COVID-19 participants *[include]* [add comment box here]
   b) Yes, but data was available for <50 COVID-19 participants *[exclude]* [add comment box here]
   c) No (e.g. data combined with those collected <4 weeks after COVID-19 diagnosis) *[exclude]* [add comment box here]

*If [exclude]*, form complete; excluded from review.
*If [include],* go to next question.

**5) The study DID NOT recruit participants specifically because they reported long-term effects (including symptoms, sequelae and difficulties carrying out usual activities) <u>4 or more weeks (28+ days) after</u> COVID-19 diagnosis.**

a) Yes or not indicated *[include]*
b) No *[exclude]*

*If [exclude]*, form complete; excluded from review.
*If [include],* go to next question.

**6) How was COVID-19 infection in the study participants determined?**

a) Laboratory-confirmed (e.g. NAAT/PCR, point of care test, serology) *[include]*
b) Clinically-diagnosed (*i.e. diagnosis was provided by a physician/health practitioner*) *[include]*
c) Self-diagnosed (*i.e. diagnosis was NOT provided by a physician/health practitioner*) *[exclude]*
d) Two or more of the above *[include]*
e) None of the above *[exclude] [add comment box to indicate method used]*
f) Not reported *[exclude] [add comment box to indicate term used by the study to refer to COVID-19 participants]*

*If [exclude]*, form complete; excluded from review.
*If selected (a) or (b)*, form complete and include for final review.
*If selected (d),* go to next question.

**7) How were data on long-term effects (outcomes/results) reported?** *Select first applicable option in order of priority:*

a) Separately for laboratory-confirmed participants *[include]*
b) Separately for clinically-diagnosed *(I.e. diagnosis was provided by a physician/health practitioner) participants [include]*
c) Combined for laboratory-confirmed and clinically-diagnosed participants only *[include]*
d) Combined with data from self-diagnosed participants (*i.e. diagnosis was NOT provided by a physician/health practitioner*) only *[exclude]*

*If [exclude]*, form complete; excluded from review.
*If [include]*, form complete and include for final review.

### Appendix 3 – Data Extraction Forms

Table 1: Characteristics of Included Studies

| **Study** | **Country, Study design, Dates** | **Study Population** (# with lab-confirmed COVID-19; mean or median age; % female) | **Disease severity** | **Time of outcome assessment since COVID-19 diagnosis** | **Main symptoms/sequelae reported** |
|---|---|---|---|---|---|

Table 2: Data extraction form – main population

| **StudyID** | **Outcome details** | | **Lab-confirmed only** | | **Prevalence** | **Time/period of follow-up** | **Clinically-diagnosed** | **Indicate with an 'X' if outcome data is reported by any of the following (& in brackets a brief explanation of which subgroups the data is available for):** | | | | | | | | | | | **Entered by:** | **Verified by:** | **Comments** |
|---|---|---|---|---|---|---|---|---|---|---|---|---|---|---|---|---|---|---|---|---|---|
| | **Outcome** (indicate name of outcome used in the study) | **Definition used in the study** (indicate if none was provided) | **# with outcome** (numerator) | **# total assessed** (denominator) | **%** | Include additional details if available (i.e. mean/median, range) | Indicate with an "X" if data available for clinically-diagnosed participants as well | **Period of observation or duration** (reported in days or weeks) | **Age group** | **Sex/gender** | **Hospitalization/ICU status** | **Severity of COVID-19 symptoms** | **Pre-existing condition or disability** | **Ethnicity** | **Geography** | **Socioeconomic factors** (e.g. occupation, education, income) | **Behavioural factors** (e.g. smoking status, alcohol use, level of physical activity) | **Other factors** (e.g. type of residence, co-infections, etc.) *please list which ones* | | | |

Table 3: Data extraction form – subgroup

| **StudyID** | **Outcome details** | | **Sub-group details** | | **Lab-confirmed only** | | **Prevalence** | **Time/period of follow-up** | **Clinically-diagnosed** | **Entered by:** | **Verified by:** | **Comments** |
|---|---|---|---|---|---|---|---|---|---|---|---|---|
| | **Outcome** (indicate name of outcome used in the study) | **Definition of outcome used in the study** (indicate if none was provided) | **Sub-group** (indicate name of subgroup used in the study) | **Definition of sub-group used in the study** (indicate if none was provided) | **# with outcome** (numerator) | **# total assessed** (denominator) | **%** | Include additional details if available (i.e. mean/median, range) | Indicate with an "X" if data available for clinically-diagnosed participants as well | | | |

#### Appendix 4 – RoB Criteria

Table 2: Risk of Bias Criteria

| ITEM | GENERAL AREA OF BIAS it addresses or corresponds to | COMMENTS | Examples |
|---|---|---|---|
| 1. Was the sample frame appropriate to address the target population? | Bias in the selection of participants into the study | This applies to the target population of the study (i.e. not the target population for our review).<br>We determine if the sample frame appropriately addresses the study's objectives.<br>e.g. consideration to specific population characteristics in the study (age range, gender, morbidities, and other potential influential factors) | Target population of study: COVID-19 adult patients who were admitted to ICU<br><br>Answer 'No' if the study population included a very large proportion of individuals who were >65 years old |
| 2. Were study participants sampled in an appropriate way? | Bias in the selection of participants into the study | Gold standard would be a census of the entire target population for the study, and if not, it would be at a higher risk of bias. | Target population of study: COVID-19 individuals with mild clinical presentation<br><br>Answer 'No' if the study population only recruited from a population that was assessed (whether outpatient or inpatient) in a hospital.<br><br>Answer 'Yes' if the study also recruited individuals who would not have sought treatment/care due to their mild clinical presentation (i.e. community survey) |
| 3. Was the sample size adequate? | N/A – will be addressed at the level of body of evidence (GRADE) | Addresses imprecision; omit | |

| | | | |
|---|---|---|---|
| 4. Were the study subjects and the setting described in detail? | N/A – will be addressed at the level of body of evidence (GRADE) | Omit. Phrased in relation to reporting but the review team can assess the scope of population & setting in relation to indirectness at the level of the body of evidence. | |
| 5. Was the data analysis conducted with sufficient coverage of the identified sample? | N/A – will be addressed at the level of body of evidence (GRADE) | This item seems to be more about indirectness than missing data. | |
| 6. Were valid methods used for the identification of the outcome? *(a list of the various outcomes reported in the study will be provided)* | Bias in measurement/ classification of condition/outcome | Focus on the validity of the tool used to assess the outcome<br>Just want to clarify language to also include the risk associated with the measurement approach – for example a self-reported tool might have decent psychometric properties (be valid and reliable) but still introduce bias due to the measurement approach (self-report, not objective). | Outcome: fever<br><br>Answer 'No' if outcome was self-reported.<br>But what if observer/self-reports are validated and it is the best we have (e.g., validated pain scales)? See last sentence in the description. This question almost needs one item for measurement approach and one for validated tool. Perhaps ask Adrienne.<br>*Response: there is still a risk of bias in self-reported outcomes despite use of a validated tool. So answer will still be 'No'*<br><br>Answer 'Yes' if a thermometer was used to record the subject's temperature to determine if fever was present or not |
| 7. Was the outcome measured in a standard, reliable | Bias in measurement/ classification of outcome: | This pertains to how the outcome was measured across observers (inter-rater reliability) and across participants (i.e. same tool was | Outcome: self-reported fatigue<br><br>Answer 'No' if some participants were asked |

| | | | |
|---|---|---|---|
| way for all participants?<br>*(a list of the various outcomes reported in the study will be provided)* | (1) for inter-rater reliability and (2) for consistency of measurement across participants | used to measure the outcome for ALL participants)<br>Assesses bias of the assessor and how standardized and reliable the methods are across participants.<br><br>*Example of "presumed" low risk of bias:*<br>*If study administered the exact same survey to all participants (no option for deviations from script/methods), then there should be no suspicion of high risk of bias for inter-rater reliability*<br>*<u>BUT</u>, this is also dependent on the outcome of interest (i.e. outcomes that require judgement from the outcome assessor introduces bias and therefore require additional information on inter-rater reliability from the study)* | to rate their level of fatigue from 0-5 and others were asked a Y/N question for presence of fatigue.<br><br>Answer 'Yes' if the same tool to measure fatigue outcome was used for all participants, and the study reported that staff were trained in the use of the tool and had good consistency |
| 8. Was the data for the outcome reported clearly and completely? (i.e. includes a numerator and denominator)<br>*(a list of the various outcomes and how the data was reported in the study will be provided)* | Focus on bias as it relates to handling missing data or selective reporting/analysis? | Otherwise, instructions are about reporting and completeness of information. | Primary outcome~~s~~ as indicated in the study methods: presence of fatigue<br><br>Answer 'No' if study reported percentages only (no numerator or denominator) **OR** indicated in their methods that they imputed missing data |
| 9. Was the response rate adequate, and if not, was the low response rate managed appropriately? | Bias due to missing data | If response rate was low, did the study compare the characteristics (i.e. especially in terms of socio-demographic characteristics) of the final study population with those that dropped out or refused to participate in the survey?<br><br>Could the outcomes be biased based on differences between those who responded versus those who did not. | Answer 'Yes' if non-participation was low (i.e. proportion of refusals or dropouts were small)<br><br>Answer 'Yes' if despite a high non-participation rate, the study found that the characteristics of the non-participating population and the study population were very similar |

**Schema for assessing Overall Risk of Bias – PER STUDY**

(a) Overall RoB assessment for Q6, Q7 & Q8 per study

- Study-level RoB assessments were asked for Q1, Q2 & Q9 in the JBI RoB tool but outcome-specific assessments were made for Q6, Q7 & Q8
- We will still report on the outcome-specific assessments per study, BUT, we need an overall study assessment for these questions.
- Here is a proposed approach, per question, and applicable only to Q6-8:

| **RoB assessments for each outcome-specific question** | **Answer for Q6, Q7 or Q8** | **Example:** Study is reporting on 10 outcomes |
|---|---|---|
| Yes response for all outcomes | "Yes" | Yes responses for Q6 for all 10 outcomes = "Yes" in Q6 for entire study |
| Mix of Yes & No responses:<br>- "Yes" was reported in **≥50%** of the outcomes<br><br>- "No" was reported in **>50%** of the outcomes | <br>"Yes"<br><br>"No" | <br>Yes responses for Q6 for 6/10 outcomes = "Yes" in Q6 for entire study<br><br>No responses for Q6 for 6/10 outcomes = "No" in Q6 for entire study |
| No response for all outcomes | "No" | No responses for Q6 for all 10 outcomes = "No" in Q6 for entire study |

(b) Overall RoB for each study – Qualitative assessment

- We need to assign a rating (high, moderate, low) for RoB per study overall
- We are adopting the qualitative assessment rating used by Maicon:
  - The JBI checklist was divided into 3 domains:
    - Participants (Q1, 2, 4 & 9)
    - Outcome measurement (Q6 & 7)
    - Statistics (Q3, 5 & 8)
  - A study was rated as having high quality when the methods were appropriate in all 3 domains.
- Because not much guidance as provided on how each domain was assessed as appropriate, we have to develop our own schema for rating. Also, we excluded Q3-5 as they were not assessing RoB.
- Here is a proposed approach:

**Step 1: Assess if RoB criteria per domain met**

| **Domain** | **Criterion NOT MET for domain** (i.e. high RoB for domain) | **Criterion PARTIALLY MET for domain** | **Criterion MET for domain** (i.e. low RoB for domain) |
|---|---|---|---|

| | | | |
|---|---|---|---|
| Participants (Q1, 2 & 9) | No for all 3 questions<br>OR<br>any other combination | Yes for Q1 and 2 but not Q9 | Yes for all 3 questions |
| Outcome measurement (Q6 & 7) | No for both questions | Yes for 1 question and No for the other | Yes for both questions |
| Statistics (Q8) | No | | Yes |

**Step 2: Assign an overall RoB assessment for the study**

| Overall RoB for study | Results of domain assessments (Step 1) | Comments |
|---|---|---|
| Low | Criteria <u>MET</u>:<br>- in ALL 3 domains<br>OR<br>- in 2 of 3 domains (& 3rd domain was partially met) | Cannot have high RoB in ANY domain |
| Moderate | High RoB in any single domain, with low/moderate RoB in the other two domains | Cannot have more than 1 domain be high RoB |
| High | Criteria <u>NOT MET</u>:<br>- in ALL 3 domains<br>OR<br>- in 2 of 3 domains | High RoB in 2+ domains |

### Appendix 5 – GRADE Assessment Decision Rules

Table 3: Summary of Decision Rules: GRADE Assessments for Certainty of Evidence from Prevalence Studies

| Domain | Judgment | Scoring | Criteria |
|---|---|---|---|
| **Risk of Bias** | No serious risk of bias | 0 | • All studies were considered at low ROB |
| | Serious risk of bias | –0.5 points | • All studies were considered at low/moderate ROB |
| | | –1 point | *If data synthesized narratively:*<br>• ≥1 study was at high ROB, but a study at high ROB *was not* responsible for either the highest or lowest prevalence estimate in the range of estimates for the outcome<br>*If data synthesized by random effects MA:*<br>• ≥1 study (but <50% of total studies) was at high ROB |
| | Very serious risk of bias | –1.5 points | *If data synthesized narratively:*<br>• ≥1 study was at high ROB, and a study at high ROB *was* responsible for either the highest or lowest prevalence estimate in the range of estimates for the outcome<br>*If data synthesized by random effects MA:*<br>• 50% or more of studies contributing to an outcome were at high ROB |
| **Inconsistency**[a] | No serious inconsistency | 0 | • In the judgment of SMEs, heterogeneity was considered expected or acceptable<br>• Heterogeneity could be explained by *a priori*-determined subgroup analyses |
| | Serious inconsistency | –0.5 points | • Heterogeneity could be partially (but not completely) explained by *a priori*-determined subgroup analyses |
| | | –1 point | • Only one study contributed to an outcome<br>• Heterogeneity could not be explained by *a priori*-determined subgroup analyses |
| **Indirectness** | No serious indirectness | 0 | • The study populations corresponded to the target population of interest for our review |
| | Serious indirectness | –0.5 points | • The study/studies contributing to an outcome included only a subset of the target population of interest for our review |
| **Imprecision** | No serious imprecision | 0 | *If data synthesized narratively:*<br>• In the judgment of SMEs, the data were sufficiently precise such that decision-making[b] would not differ if the highest versus lowest point estimate from individual studies represented the truth, *and* the OIS was met<br>*If data synthesized by random effects MA:* |

| | | | |
|---|---|---|---|
| | | | • In the judgment of SMEs, the data were sufficiently precise such that decision-making would not differ if the upper or lower boundary of the CI represented the truth, *and* the OIS was met |
| | Serious imprecision | –0.5 points | *If data synthesized narratively:*<br>• In the judgment of SMEs, the data were sufficiently precise such that decision-making would not differ if the highest versus lowest point estimate from individual studies represented the truth, but the OIS *was not* met<br>*If data synthesized by random effects MA:*<br>• In the judgment of SMEs, the data were sufficiently precise such that decision-making would not differ if the upper or lower boundary of the CI represented the truth, but the OIS *was not* met |
| | | –1 point | *If data synthesized narratively:*<br>• In the judgment of SMEs, decision-making *would* differ if the highest versus lowest point estimate from individual studies represented the truth<br>*If data synthesized by random effects MA:*<br>• In the judgment of SMEs, decision-making *would* differ if the upper or lower boundary of the CI represented the truth |
| **Publication Bias** | No serious risk of publication bias | 0 | Due to the large volume of COVID-19-related manuscripts being published and the large interest in publishing any findings related to long-COVID, an *a priori* decision was made not to downgrade for risk of publication bias |

CI = confidence interval; COVID-19 = coronavirus disease 2019; GRADE = Grading of Recommendations Assessment, Development and Evaluation; long-COVID = long-term effects of COVID-19; MA = meta-analysis; OIS = optimal information size; ROB = risk of bias; SME = subject matter expert.

**Notes:**

- The quality of the prevalence evidence from studies of all designs was initially assigned as "high" for all outcomes (Iorio et al. 2014).
- We did not consider upgrading the quality of the evidence given that upgrading is only appropriate when there is no cause to downgrade, and the quality of evidence was initially assigned as "high".
- Half-points were combined across domains to yield a total score; if the final scoring included a half-point, we conservatively rounded up.
- Final scoring for the overall certainty of evidence was as follows: –1 point = "moderate"; –1.5 or –2 points = "low"; –2.5 points or more = "very low".

[a] $I^2$ values were not considered because high $I^2$ values are expected in a MA of prevalence studies that include heterogeneous populations.
[b] In this context, potential decisions pertained to whether to invest in additional research and/or surveillance for an outcome.

### Appendix 6 – Search Strategy Implementation

**Summary**

| | January 15, 2021 | April 14, 2021 |
|---|---|---|
| Embase | 1442 | 2343 |
| Embase TopUp1 | 1 | 1 |
| Embase TopUp2 | 92 | 160 |
| Medline | 838 | 1581 |
| Medline TopUp1 | 0 | 2 |
| Medline TopUp2 | 54 | 141 |
| PsychInfo | 50 | 67 |
| PsychInfo TopUp1 | 0 | 0 |
| PsychInfo TopUp2 | 0 | 0 |
| Cochrane Central | 41 | 82 |
| Grey Literature | 289 | |
| Total | 2805 | |
| **Total after deduplication** | **2142** | |

| **Grey Literature** | January 2021 | April 2021 |
|---|---|---|
| Cochrane COVID Review Bank | 0 | 0 |
| BMJ Best Practice | 7 | 6 |
| Cochrane COVID-19 Study Register | 246 | 422 |
| COVID-END | 2 | 0 |
| Centre for Evidence-based Medicine – Oxford COVID-19 Evidence Service | 0 | 0 |
| CADTH COVID-19 pandemic database | 0 | 2 |
| National COVID-19 Clinical Evidence Task Force | 5 | 1 |

| | | |
|---|---|---|
| Appendix 1 #2 | 21 | Not applicable |
| Appendix 1 #3 | 3 | Not applicable |
| French language | 5 | Not applicable |
| **Total** | **289** | **431** |

(5) Partners E. DistillerSR.

(6) R CT. R Studio. 2013.

(7) Viechtbauer W. Conducting Meta-Analyses in R with the metafor Package. Journal of Statistical Software 2010;36(3).

(8) Schwarzer G. General Package for Meta-Analysis. 2021; Available at: https://cran.r-project.org/web/packages/meta/meta.pdf. Accessed April, 2021.

(9) Barendregt JJ, Doi SA, Lee YY, Norman RE, Vos T. Meta-analysis of prevalence. J Epidemiol Community Health 2013 11;67(11):974-978.

(10) Higgins JPT. Measuring inconsistency in meta-analyses. BMJ 2003 09;327(7414):557-560.

(11) Munn Z, Moola S, Lisy K, Riitano D, Tufanaru C. Chapter 5: Systematic reviews of prevalence and incidence. In: Aromataris E, Munn Z, editors. : JBI; 2017.
