## Supplementary Files 2 to 4 for "Prevalence of long-term effects in individuals diagnosed with COVID-19: an updated living systematic review"

**Authors:**

Francesca Reyes Domingo MHSc[1], Lisa A Waddell MSc PhD[2], Angela M. Cheung MD FRCPC[3], Curtis L. Cooper MD FRCPC[4ah], Veronica J. Belcourt MSc PhD[1], Alexandra M. E. Zuckermann PhD[1], Tricia Corrin MPH[2], Rukshanda Ahmad MBBS MHA[5], Laura Boland SLP-C PhD[1], Claudie Laprise MSc PhD[6,7], Leanne Idzerda MSc PhD[1], Anam Khan MPH[5], Kate Morissette MSc[1], Alejandra Jaramillo Garcia MSc[1]

**Affiliations:**

[1] Evidence Synthesis and Knowledge Translation Unit, Applied Research Division, Public Health Agency of Canada

[2] Public Health Risk Sciences Division, National Microbiology Laboratory, Public Health Agency of Canada

[3] Department of Medicine, University Health Network and University of Toronto, Canada

[4] Department of Medicine, University of Ottawa; Ottawa Hospital Research Institute

[5] Health Professionals Guidance Unit, Centre for Food-borne, Environmental and Zoonotic Infectious Diseases, Public Health Agency of Canada

[6] Public Health Capacity and Knowledge Management Unit, Quebec Regional Office, Public Health Agency of Canada

[7] Division of Oral Health and Society, Faculty of Dentistry, McGill University

Runny nose ... 111
Any cough ... 112
Dry cough ... 113
Productive cough ... 113
Sore throat ... 113
Phlegm ... 114
Wheezing ... 114
Influenza-like illness ... 115
Headaches ... 115
Chills ... 115
Fever ... 115
Olfactory and Taste Impairments ... 116
Any Smell dysfunction ... 116
Anosmia ... 116
Any Taste dysfunction ... 117
Ageusia ... 117
Any Smell or Taste dysfunction ... 118
Neuromuscular ... 118
Joint pain ... 118
Muscle pain ... 119
Neurocognitive ... 120
Cognitive impairment ... 120
Concentration problems ... 120
Memory problems ... 120
Dizziness ... 121
Cardiovascular ... 121
Chest pain ... 121
Palpitations ... 122
Chest tightness ... 122
Digestive ... 123
Diarrhea ... 123
Abdominal pain ... 124
Mental ... 124
Anxiety ... 124
Depression ... 124

### Supplementary File 2 – Excluded studies

**Not relevant article = 284**

| | |
|---|---|
| 1 | Quincozes-Santos A., Rosa R.L., Tureta E.F., Bobermin L.D., Berger M., Guimaraes J.A., Santi L., Beys-da-Silva W.O.. COVID-19 impacts the expression of molecular markers associated with neuropsychiatric disorders. Brain, Behavior, and Immunity - Health. 2021; 11(#issue#): 100196. doi: http://dx.doi.org/10.1016/j.bbih.2020.100196. Available at: https://www.journals.elsevier.com/brain-behavior-and-immunity-health/; http://ovidsp.ovid.com/ovidweb.cgi?T=JS&PAGE=reference&D=emedx&NEWS=N&AN=2010527885 |
| 2 | Jianrong L., Lyu H., Yuan X., An Y., Yi X., Wei X., Yi H., Yang Y.. Late Breaking Abstract - Multidisciplinary medical team in severe COVID-19 treatment. European Respiratory Journal. 2020; 56(#issue#): #pages#. doi: http://dx.doi.org/10.1183/13993003.congress-2020.333. Available at: https://erj.ersjournals.com/content/56/suppl_64/333; http://ovidsp.ovid.com/ovidweb.cgi?T=JS&PAGE=reference&D=emedx&NEWS=N&AN=633805683 |
| 3 | Ruberto F., Chistolini A., Curreli M., Frati G., Marullo A.G.M., Biondi-Zoccai G., Mancone M., Sciarretta S., Miraldi F., Alessandri F., Ceccarelli G., Barone F., Santoro C., Alvaro D., Pugliese F., Albante A., Antonelli G., Araimo-Morselli F., Auricchio D., Bilotta F., Brisciani G.B.-Z.M., Bruno K., Cappannoli A., Cardinale V., Celli P., Consolo S., Croce C., Crocitti B., D'Ettorre G., De Daniela L., De Lazzaro F., Fedele F., Galardo G., Giannetti L., Ianni S., Imperiale C., Maestrini V., Magnanimi E., Maldarelli F., Martelli S., Mastroianni C., Messina T., Pattelli E., Pecorari F., Perrella S., Piazzolla M., Portieri M., Pulcinelli F.M., Ratini F., Ricci C., Santopietro P., Tellan G., Titi L., Tordiglione P., Tosi A., Trigilia F.. Von Willebrand factor with increased binding capacity is associated with reduced platelet aggregation but enhanced agglutination in COVID-19 patients: another COVID-19 paradox?. Journal of thrombosis and thrombolysis. 2021; #volume#(#issue#): #pages#. doi: http://dx.doi.org/10.1007/s11239-020-02339-6. Available at: http://www.wkap.nl/journalhome.htm/0929-5305; http://ovidsp.ovid.com/ovidweb.cgi?T=JS&PAGE=reference&D=emedx&NEWS=N&AN=2007711838 |
| 4 | Sirin D.A., Ozcelik F.. The relationship between COVID-19 and the dental damage stage determined by radiological examination. Oral Radiology. 2021; #volume#(#issue#): #pages#. doi: http://dx.doi.org/10.1007/s11282-020-00497-0. Available at: http://www.springer.com/journal/11282; http://ovidsp.ovid.com/ovidweb.cgi?T=JS&PAGE=reference&D=emedx&NEWS=N&AN=2007711822 |
| 5 | Aggarwal A., Jain A., Jain P., Sangha M., Benias P.C., Trindade A.J.. SARS-CoV-2 Impact on Screening Colonoscopy: Implications for Postpandemic Recovery. The American Journal of Gastroenterology. 2020; #volume#(#issue#): #pages#. doi: http://dx.doi.org/10.14309/ajg.0000000000001097. Available at: http://ovidsp.ovid.com/ovidweb.cgi?T=JS&PAGE=reference&D=emedx&NEWS=N&AN=633805962 |
| 6 | Sharma S.K., Rahat I.. A pathophysiological review on understanding multiorgan effects of COVID-19. European Journal of Molecular and Clinical Medicine. 2020; 7(6): 1037. doi: #DOI#. Available at: https://ejmcm.com/pdf_3640_c22912585aaeb978fee1567d41e339b9.html; http://ovidsp.ovid.com/ovidweb.cgi?T=JS&PAGE=reference&D=emedx&NEWS=N&AN=2010507759 |
| 7 | McMullen K.M., Smith B.A., Rebmann T.. Impact of SARS-CoV-2 on hospital acquired infection rates in the United States: Predictions and early results. American Journal of Infection Control. 2020; 48(11): 1409. doi: http://dx.doi.org/10.1016/j.ajic.2020.06.209. Available at: http://www.journals.elsevier.com/ajic-american-journal-of-infection-control/; http://ovidsp.ovid.com/ovidweb.cgi?T=JS&PAGE=reference&D=emexc&NEWS=N&AN=2007179735 |
| 8 | Donnelly S.C.. Post COVID Syndrome (PCS) and healthcare workers: Who cares for the carers?. QJM. 2020; 113(9): 611. doi: http://dx.doi.org/10.1093/qjmed/hcaa248. Available at: http://qjmed.oxfordjournals.org/; http://ovidsp.ovid.com/ovidweb.cgi?T=JS&PAGE=reference&D=emexc&NEWS=N&AN=2010200962 |
| 9 | de Havenon A., Ney J.P., Callaghan B., Delic A., Hohmann S., Shippey E., Esper G.J., Stulberg E., Tirschwell D., Frontera J., Yaghi S., Anadani M., Majersik J.J.. Impact of COVID-19 on Outcomes in Ischemic Stroke |

| | |
|---|---|
| | Patients in the United States. Journal of Stroke and Cerebrovascular Diseases. 2021; 30(2): 105535. doi: http://dx.doi.org/10.1016/j.jstrokecerebrovasdis.2020.105535. Available at: http://www.elsevier.com/inca/publications/store/6/2/3/3/6/9/index.htt; http://ovidsp.ovid.com/ovidweb.cgi?T=JS&PAGE=reference&D=emexc&NEWS=N&AN=2010294079 |
| 10 | Gordon W.J., Henderson D., DeSharone A., Fisher H.N., Judge J., Levine D.M., MacLean L., Sousa D., Su M.Y., Boxer R.. Remote Patient Monitoring Program for Hospital Discharged COVID-19 Patients. Applied clinical informatics. 2020; 11(5): 792. doi: http://dx.doi.org/10.1055/s-0040-1721039. Available at: http://ovidsp.ovid.com/ovidweb.cgi?T=JS&PAGE=reference&D=emexc&NEWS=N&AN=633535728 |
| 11 | Bao Y., Ling Y., Chen Y.-Y., Tian D., Zhao G.-P., Zhang X.-H., Hong H., Li Y., Su B., Lu H.-Z., Xu J., Wang Y.. Dynamic Anti-Spike Protein Antibody Profiles in COVID-19 Patients. International journal of infectious diseases : IJID : official publication of the International Society for Infectious Diseases. 2020; #volume#(#issue#): #pages#. doi: http://dx.doi.org/10.1016/j.ijid.2020.12.014. Available at: http://ovidsp.ovid.com/ovidweb.cgi?T=JS&PAGE=reference&D=emexc&NEWS=N&AN=633786683 |
| 12 | Zhu S., Gao Q., Yang L., Yang Y., Xia W., Cai X., Hui Y., Zhu D., Zhang Y., Zhang G., Wu S., Wang Y., Zhou Z., Liu H., Zhang C., Zhang B., Yang J., Feng M., Ni Z., Chen B., Du C., He H., Qu Y., Wei Q., He C., Reinhardt J.D.. Prevalence and risk factors of disability and anxiety in a retrospective cohort of 432 survivors of Coronavirus Disease-2019 (Covid-19) from China. PLoS ONE. 2020; 15(12): e0243883. doi: http://dx.doi.org/10.1371/journal.pone.0243883. Available at: https://journals.plos.org/plosone/article/file?id=10.1371/journal.pone.0243883&type=printable; http://ovidsp.ovid.com/ovidweb.cgi?T=JS&PAGE=reference&D=emexc&NEWS=N&AN=2010476143 |
| 13 | Lyons D., Frampton M., Naqvi S., Donohoe D., Adams G., Glynn K.. Fallout from the COVID-19 pandemic-Should we prepare for a tsunami of post viral depression?. Irish Journal of Psychological Medicine. 2020; 37(4): 295. doi: http://dx.doi.org/10.1017/ipm.2020.40. Available at: http://journals.cambridge.org/action/displayBackIssues?jid=IPM) Content List URL; http://ovidsp.ovid.com/ovidweb.cgi?T=JS&PAGE=reference&D=emexc&NEWS=N&AN=631875286 |
| 14 | Kunal S., Sharma S.M., Sharma S.K., Gautam D., Bhatia H., Mahla H., Sharma S., Bhandari S.. Cardiovascular complications and its impact on outcomes in COVID-19. Indian heart journal. 2020; 72(6): 593. doi: http://dx.doi.org/10.1016/j.ihj.2020.10.005. Available at: http://www.journals.elsevier.com/indian-heart-journal/; http://ovidsp.ovid.com/ovidweb.cgi?T=JS&PAGE=reference&D=emexc&NEWS=N&AN=2010052640 |
| 15 | Dotson S., Ciarocco S., Koh K.A.. Disaster psychiatry and homelessness: creating a mental health COVID-19 response. The Lancet Psychiatry. 2020; 7(12): 1006. doi: http://dx.doi.org/10.1016/S2215-0366%2820%2930343-6. Available at: https://www.journals.elsevier.com/the-lancet-psychiatry; http://ovidsp.ovid.com/ovidweb.cgi?T=JS&PAGE=reference&D=emexc&NEWS=N&AN=2007636478 |
| 16 | Nafakhi H., Alareedh M., Al-Buthabhak K., Shaghee F., Nafakhi A., Kasim S.. Predictors of adverse in-hospital outcome and recovery in patients with diabetes mellitus and COVID-19 pneumonia in Iraq. Diabetes and Metabolic Syndrome: Clinical Research and Reviews. 2021; 15(1): 33. doi: http://dx.doi.org/10.1016/j.dsx.2020.12.014. Available at: http://www.journals.elsevier.com/diabetes-and-metabolic-syndrome-clinical-research-and-reviews/; http://ovidsp.ovid.com/ovidweb.cgi?T=JS&PAGE=reference&D=emexc&NEWS=N&AN=2010257161 |
| 17 | Rinaldi M., Madaro F., Righetti G., Scarafino A., Infantino R., Chetta N., Covella B., Mastroianni F.. Two-step analysis of the COVID19 pandemic:has the virus reduced its lethality?. Italian Journal of Medicine. 2020; 14(#issue#): 116. doi: #DOI#. Available at: https://www.italjmed.org/index.php/ijm/article/view/itjm.2020.s2/1309; http://ovidsp.ovid.com/ovidweb.cgi?T=JS&PAGE=reference&D=emexc&NEWS=N&AN=633718876 |
| 18 | Lagana N., Cerutti S., Rotiroti G., Capra R., Porta C., Mazzone A., Cei M., Evangelista I., Mumoli N.. Myocarditis in patients with COVID-19: A multicenter caseseries. Italian Journal of Medicine. 2020; 14(#issue#): 113. doi: #DOI#. Available at: https://www.italjmed.org/index.php/ijm/article/view/itjm.2020.s2/1309; http://ovidsp.ovid.com/ovidweb.cgi?T=JS&PAGE=reference&D=emexc&NEWS=N&AN=633718534 |

| | |
|---|---|
| 19 | Ciarambino T., Giannico O.V., Rotunno S., Fiorentini A., Cipriani R., Campagna G., Straface G., Pistella E., Pietrantonio F., Lorenzi F.. Covid-19 differences by gender: An experience byCovid- Hospital Lazio. Italian Journal of Medicine. 2020; 14(#issue#): 111. doi: #DOI#. Available at: https://www.italjmed.org/index.php/ijm/article/view/itjm.2020.s2/1309; http://ovidsp.ovid.com/ovidweb.cgi?T=JS&PAGE=reference&D=emexc&NEWS=N&AN=633718258 |
| 20 | Calles A., Alva M., Aparicio I., Soto J., Gutierrez N., Bringas M., Escudero V., Collado R., Galera M., Alvarez R.. Impact of COVID-19 in continuity ofcancer treatment for lung cancer patients. Clinical Cancer Research. 2020; 26(18): #pages#. doi: http://dx.doi.org/10.1158/1557-3265.COVID-19-PO-021. Available at: https://clincancerres.aacrjournals.org/content/26/18_Supplement/PO-021; http://ovidsp.ovid.com/ovidweb.cgi?T=JS&PAGE=reference&D=emexc&NEWS=N&AN=633717637 |
| 21 | Garcez F.B., Aliberti M.J.R., Poco P.C.E., Hiratsuka M., Takahashi S.D.F., Coelho V.A., Salotto D.B., Moreira M.L.V., Jacob-Filho W., Avelino-Silva T.J.. Delirium and Adverse Outcomes in Hospitalized Patients with COVID-19. Journal of the American Geriatrics Society. 2020; 68(11): 2440. doi: http://dx.doi.org/10.1111/jgs.16803. Available at: http://onlinelibrary.wiley.com/journal/10.1111/(ISSN)1532-5415; http://ovidsp.ovid.com/ovidweb.cgi?T=JS&PAGE=reference&D=emexc&NEWS=N&AN=2006074370 |
| 22 | Venter C., Bezuidenhout J.A., Laubscher G.J., Lourens P.J., Steenkamp J., Kell D.B., Pretorius E.. Erythrocyte, platelet, serum ferritin, and p-selectin pathophysiology implicated in severe hypercoagulation and vascular complications in COVID-19. International Journal of Molecular Sciences. 2020; 21(21): 1. doi: http://dx.doi.org/10.3390/ijms21218234. Available at: https://www.mdpi.com/1422-0067/21/21/8234/pdf; http://ovidsp.ovid.com/ovidweb.cgi?T=JS&PAGE=reference&D=emexc&NEWS=N&AN=2005417497 |
| 23 | Pinto M., Gimigliano F., De Simone S., Costa M., Bianchi A.A.M., Iolascon G.. Post-acute covid-19 rehabilitation network proposal: From intensive to extensive and home-based it supported services. International Journal of Environmental Research and Public Health. 2020; 17(24): 1. doi: http://dx.doi.org/10.3390/ijerph17249335. Available at: https://www.mdpi.com/1660-4601/17/24/9335/pdf; http://ovidsp.ovid.com/ovidweb.cgi?T=JS&PAGE=reference&D=emexc&NEWS=N&AN=2005616604 |
| 24 | Rha M.-S., Jeong H.W., Ko J.-H., Choi S.J., Seo I.-H., Lee J.S., Sa M., Kim A.R., Joo E.-J., Ahn J.Y., Kim J.H., Song K.-H., Kim E.S., Oh D.H., Ahn M.Y., Choi H.K., Jeon J.H., Choi J.-P., Kim H.B., Kim Y.K., Park S.-H., Choi W.S., Choi J.Y., Peck K.R., Shin E.-C.. PD-1-Expressing SARS-CoV-2-Specific CD8+ T Cells Are Not Exhausted, but Functional in Patients with COVID-19. Immunity. 2020; #volume#(#issue#): #pages#. doi: http://dx.doi.org/10.1016/j.immuni.2020.12.002. Available at: http://www.immunity.com; http://ovidsp.ovid.com/ovidweb.cgi?T=JS&PAGE=reference&D=emexc&NEWS=N&AN=2010403890 |
| 25 | Abrishami M., Emamverdian Z., Shoeibi N., Omidtabrizi A., Daneshvar R., Saeidi Rezvani T., Saeedian N., Eslami S., Mazloumi M., Sadda S., Sarraf D.. Optical coherence tomography angiography analysis of the retina in patients recovered from COVID-19: a case-control study. Canadian Journal of Ophthalmology. 2020; #volume#(#issue#): #pages#. doi: http://dx.doi.org/10.1016/j.jcjo.2020.11.006. Available at: http://www.sciencedirect.com/science/journal/00084182; http://ovidsp.ovid.com/ovidweb.cgi?T=JS&PAGE=reference&D=emexc&NEWS=N&AN=2010293657 |
| 26 | Rashid A.. Yonder: Long Covid, mental health video consultations, overactive bladder syndrome, and haematological malignancies. British Journal of General Practice. 2020; 70(701): 601. doi: http://dx.doi.org/10.3399/BJGP20X713765. Available at: http://bjgp.org/lookup/doi/10.3399/bjgp20X713765; http://ovidsp.ovid.com/ovidweb.cgi?T=JS&PAGE=reference&D=emexc&NEWS=N&AN=2010355065 |
| 27 | El Sayed S., Shokry D., Gomaa S.M.. Post-COVID-19 fatigue and anhedonia: A cross-sectional study and their correlation to post-recovery period. Neuropsychopharmacology Reports. 2020; #volume#(#issue#): #pages#. doi: http://dx.doi.org/10.1002/npr2.12154. Available at: http://onlinelibrary.wiley.com/journal/10.1002/(ISSN)2574-173X; http://ovidsp.ovid.com/ovidweb.cgi?T=JS&PAGE=reference&D=emexc&NEWS=N&AN=2007620941 |

| | |
|---|---|
| 28 | Hasichaolu, Zhang X., Li X., Li D.. Circulating cytokines and lymphocyte subsets in patients who have recovered from covid-19. BioMed Research International. 2020; 2020(#issue#): 7570981. doi: http://dx.doi.org/10.1155/2020/7570981. Available at: http://www.hindawi.com/journals/biomed/; http://ovidsp.ovid.com/ovidweb.cgi?T=JS&PAGE=reference&D=emexc&NEWS=N&AN=2010321523 |
| 29 | Loomba R.S., Villarreal E.G., Farias J.S., Bronicki R.A., Flores S.. Pediatric intensive care unit admissions for COVID-19: Insights using state-level data. International Journal of Pediatrics (United Kingdom). 2020; 2020(#issue#): 9680905. doi: http://dx.doi.org/10.1155/2020/9680905. Available at: http://www.hindawi.com/journals/ijpedi/; http://ovidsp.ovid.com/ovidweb.cgi?T=JS&PAGE=reference&D=emexc&NEWS=N&AN=2010321290 |
| 30 | Huang D., Yang H., Yu H., Wang T., Yao R., Liang Z.. A novel risk score to predict cardiovascular complications in patients with coronavirus disease 2019 (COVID-19): A retrospective, multicenter, observational study. Immunity, Inflammation and Disease. 2020; 8(4): 638. doi: http://dx.doi.org/10.1002/iid3.353. Available at: http://onlinelibrary.wiley.com/journal/10.1002/(ISSN)2050-4527; http://ovidsp.ovid.com/ovidweb.cgi?T=JS&PAGE=reference&D=emexc&NEWS=N&AN=2006776308 |
| 31 | Tale S., Ghosh S., Meitei S.P., Kolli M., Garbhapu A.K., Pudi S.. Post-COVID-19 pneumonia pulmonary fibrosis. QJM. 2020; 113(11): 837. doi: http://dx.doi.org/10.1093/qjmed/hcaa255. Available at: http://qjmed.oxfordjournals.org/; http://ovidsp.ovid.com/ovidweb.cgi?T=JS&PAGE=reference&D=emexc&NEWS=N&AN=2010312022 |
| 32 | Wang Z., Lorenzi J.C.C., Muecksch F., Finkin S., Viant C., Gaebler C., Cipolla M., Hoffmann H.-H., Oliveira T.Y., Oren D.A., Ramos V., Nogueira L., Michailidis E., Robbiani D.F., Gazumyan A., Rice C.M., Hatziioannou T., Bieniasz P.D., Caskey M., Nussenzweig M.C.. Enhanced SARS-CoV-2 neutralization by dimeric IgA. Science translational medicine. 2020; #volume#(#issue#): #pages#. doi: http://dx.doi.org/10.1126/scitranslmed.abf1555. Available at: http://ovidsp.ovid.com/ovidweb.cgi?T=JS&PAGE=reference&D=emexc&NEWS=N&AN=633651572 |
| 33 | Li L., Zhang S., He B., Chen X., Wang S., Zhao Q.. Risk factors and electrocardiogram characteristics for mortality in critical inpatients with COVID-19. Clinical cardiology. 2020; 43(12): 1624. doi: http://dx.doi.org/10.1002/clc.23492. Available at: http://onlinelibrary.wiley.com/journal/10.1002/(ISSN)1932-8737; http://ovidsp.ovid.com/ovidweb.cgi?T=JS&PAGE=reference&D=emexc&NEWS=N&AN=2007041057 |
| 34 | Cicco S., Vacca A., Cittadini A., Marra A.M.. Long-term follow-up may be useful in coronavirus disease 2019 survivors to prevent chronic complications. Infection and Chemotherapy. 2020; 52(3): 407. doi: http://dx.doi.org/10.3947/ic.2020.52.3.407. Available at: https://www.icjournal.org/Synapse/Data/PDFData/0086IC/ic-52-407.pdf; http://ovidsp.ovid.com/ovidweb.cgi?T=JS&PAGE=reference&D=emexc&NEWS=N&AN=2008412244 |
| 35 | Romero M.M., Cespedes A.A., Sahuquillo M.T.T., Zamora E.B.C., Ballesteros C.C., Alfaro V.S.-F., Bru R.L., Utiel M.L., Cifuentes S.C., Longobardo L.M.P., Romero A.M., Carmona L.P., Garcia B.G., Fernandez-Rius A.P., Corcoles R.A.C., Garcia B.R., Rizos L.R., Jurado P.M.S., Ortiz M.L., Nunez P.A., Garcia A.N., Garcia M.F.R., Molina R.G., De Dios Estrella Cazalla J., Moreno J.O., Abizanda P.. COVID-19 outbreak in long-term care facilities from Spain. Many lessons to learn. PLoS ONE. 2020; 15(10): e0241030. doi: http://dx.doi.org/10.1371/journal.pone.0241030. Available at: https://journals.plos.org/plosone/article/file?id=10.1371/journal.pone.0241030&type=printable; http://ovidsp.ovid.com/ovidweb.cgi?T=JS&PAGE=reference&D=emexc&NEWS=N&AN=2008446836 |
| 36 | Cusack D.A.. COVID-19 pandemic: Coroner's database of death inquiries with clinical epidemiology and total and excess mortality analyses in the District of Kildare March to June 2020. Journal of Forensic and Legal Medicine. 2020; 76(#issue#): 102072. doi: http://dx.doi.org/10.1016/j.jflm.2020.102072. Available at: http://www.elsevier.com; http://ovidsp.ovid.com/ovidweb.cgi?T=JS&PAGE=reference&D=emexc&NEWS=N&AN=2008456173 |
| 37 | Rebora P., Rozzini R., Bianchetti A., Blangiardo P., Marchegiani A., Piazzoli A., Mazzeo F., Cesaroni G., Chizzoli A., Guerini F., Bonfanti P., Morandi A., Faraci B., Gentile S., Bna C., Savelli G., Citerio G., Valsecchi M.G., Mazzola P., Bellelli G.. Delirium in Patients with SARS-CoV-2 Infection: A Multicenter Study. Journal |

| | |
|---|---|
| | of the American Geriatrics Society. 2020; #volume#(#issue#): #pages#. doi: http://dx.doi.org/10.1111/jgs.16969. Available at: http://onlinelibrary.wiley.com/journal/10.1111/(ISSN)1532-5415; http://ovidsp.ovid.com/ovidweb.cgi?T=JS&PAGE=reference&D=emexc&NEWS=N&AN=2007544847 |
| 38 | Liu Y., Zhang X.. Persistent coronavirus infection of progenitor oligodendrocytes. The Nidoviruses: Toward Control of SARS and other Nidovirus Diseases; Advances in Experimental Medicine and Biology. 2006; 581(#issue#): 379. doi: http://dx.doi.org/10.1007/978-0-387-33012-9_67. Available at: http://www.springer.com/series/5584; http://ovidsp.ovid.com/ovidweb.cgi?T=JS&PAGE=reference&D=emexc&NEWS=N&AN=354064223 |
| 39 | Vahedi E., Ghanei M., Ghazvini A., Azadi H., Izadi M., Panahi Y., Fathi S., Salesi M., Saadat S.H., Ghazale A.H., Rezapour M., Mozafari A., Zand N., Parsaei M.R., Ranjkesh M.H., Jafari R., Movaseghi F., Darabi E.. The clinical value of two combination regimens in the Management of Patients Suffering from Covid-19 pneumonia: a single centered, retrospective, observational study. DARU, Journal of Pharmaceutical Sciences. 2020; 28(2): 507. doi: http://dx.doi.org/10.1007/s40199-020-00353-w. Available at: https://rd.springer.com/journal/40199; http://ovidsp.ovid.com/ovidweb.cgi?T=JS&PAGE=reference&D=emexc&NEWS=N&AN=2005273673 |
| 40 | Padalabalanarayanan S., Hanumanthu V.S., Sen B.. Association of State Stay-at-Home Orders and State-Level African American Population with COVID-19 Case Rates. JAMA Network Open. 2020; 3(10): e2026010. doi: http://dx.doi.org/10.1001/jamanetworkopen.2020.26010. Available at: https://jamanetwork.com/journals/jamanetworkopen; http://ovidsp.ovid.com/ovidweb.cgi?T=JS&PAGE=reference&D=emexc&NEWS=N&AN=633197463 |
| 41 | Maghool F., Valiani A., Safari T., Emami M.H., Mohammadzadeh S.. Gastrointestinal and renal complications in SARS-CoV-2-infected patients: Role of immune system. Scandinavian Journal of Immunology. 2020; #volume#(#issue#): #pages#. doi: http://dx.doi.org/10.1111/sji.12999. Available at: http://onlinelibrary.wiley.com/journal/10.1111/(ISSN)1365-3083; http://ovidsp.ovid.com/ovidweb.cgi?T=JS&PAGE=reference&D=emexc&NEWS=N&AN=2007543323 |
| 42 | Xu K., Chen Y., Yuan J., Yi P., Ding C., Wu W., Li Y., Ni Q., Zou R., Li X., Xu M., Zhang Y., Zhao H., Zhang X., Su J., Lang G., Liu J., Wu X., Guo Y., Tao J., Shi D., Yu L., Cao Q., Ruan B., Liu L., Wang Z., Xu Y., Liu Y., Sheng J., Li L.. Factors associated with prolonged viral RNA shedding in patients with coronavirus disease 2019 (COVID-19). Clinical Infectious Diseases. 2020; 71(15): 799. doi: http://dx.doi.org/10.1093/cid/ciaa351. Available at: http://cid.oxfordjournals.org/content/by/year; http://ovidsp.ovid.com/ovidweb.cgi?T=JS&PAGE=reference&D=emexc&NEWS=N&AN=2008415127 |
| 43 | Habel J.R., Nguyen T.H.O., van de Sandt C.E., Juno J.A., Chaurasia P., Wragg K., Koutsakos M., Hensen L., Jia X., Chua B., Zhang W., Tan H.-X., Flanagan K.L., Doolan D.L., Torresi J., Chen W., Wakim L.M., Cheng A.C., Doherty P.C., Petersen J., Rossjohn J., Wheatley A.K., Kent S.J., Rowntree L.C., Kedzierska K.. Suboptimal SARS-CoV-2-specific CD8+ T cell response associated with the prominent HLA-A*02:01 phenotype. Proceedings of the National Academy of Sciences of the United States of America. 2020; 117(39): 24384. doi: http://dx.doi.org/10.1073/pnas.2015486117. Available at: https://www.pnas.org/content/pnas/117/39/24384.full.pdf; http://ovidsp.ovid.com/ovidweb.cgi?T=JS&PAGE=reference&D=emexc&NEWS=N&AN=2008030595 |
| 44 | Griffin D.E.. Are T cells helpful for COVID-19: The relationship between response and risk. Journal of Clinical Investigation. 2020; 130(12): 6222. doi: http://dx.doi.org/10.1172/JCI142081. Available at: https://www.jci.org/articles/view/142081/pdf; http://ovidsp.ovid.com/ovidweb.cgi?T=JS&PAGE=reference&D=emexc&NEWS=N&AN=2010243240 |
| 45 | Roberts P., Wertheimer J., Park E., Nuno M., Riggs R.. Identification of Functional Limitations and Discharge Destination in COVID-19 Patients. Archives of Physical Medicine and Rehabilitation. 2020; #volume#(#issue#): #pages#. doi: http://dx.doi.org/10.1016/j.apmr.2020.11.005. Available at: http://ovidsp.ovid.com/ovidweb.cgi?T=JS&PAGE=reference&D=emexc&NEWS=N&AN=633607203 |
| 46 | Dawson C., Capewell R., Ellis S., Matthews S., Adamson S., Wood M., Fitch L., Reid K., Shaw M., Wheeler J., Pracy P., Nankivell P., Sharma N.. Dysphagia presentation and management following coronavirus disease 2019: an acute care tertiary centre experience. Journal of Laryngology and Otology. 2020; |

| | |
|---|---|
| | #volume#(#issue#): #pages#. doi: http://dx.doi.org/10.1017/S0022215120002443. Available at: http://journals.cambridge.org/action/displayJournal?jid=JLO; http://ovidsp.ovid.com/ovidweb.cgi?T=JS&PAGE=reference&D=emexc&NEWS=N&AN=633601667 |
| 47 | Brenner E.J., Pigneur B., Focht G., Zhang X., Ungaro R.C., Colombel J.-F., Turner D., Kappelman M.D., Ruemmele F.M.. Benign Evolution of SARS-Cov2 Infections in Children With Inflammatory Bowel Disease: Results From Two International Databases. Clinical Gastroenterology and Hepatology. 2020; #volume#(#issue#): #pages#. doi: http://dx.doi.org/10.1016/j.cgh.2020.10.010. Available at: http://www.elsevier.com/inca/publications/store/6/7/2/7/4/3/index.htt; http://ovidsp.ovid.com/ovidweb.cgi?T=JS&PAGE=reference&D=emexc&NEWS=N&AN=2010262370 |
| 48 | Takkar A., Singla M.. The Neurological Aspects of COVID-19: Do Women Respond Differently. Indian Journal of Cardiovascular Disease in Women - WINCARS. 2020; 5(3): 264. doi: http://dx.doi.org/10.1055/s-0040-1716134. Available at: https://www.thieme.com/books-main/surgery/product/4443-indian-journal-of-cardiovascular-disease-in-women-wincars; http://ovidsp.ovid.com/ovidweb.cgi?T=JS&PAGE=reference&D=emexc&NEWS=N&AN=632860319 |
| 49 | Schmulson M., Ghoshal U.C., Barbara G.. Managing the Inevitable Surge of Post-COVID-19 Functional Gastrointestinal Disorders. The American Journal of Gastroenterology. 2020; #volume#(#issue#): #pages#. doi: http://dx.doi.org/10.14309/ajg.0000000000001062. Available at: http://ovidsp.ovid.com/ovidweb.cgi?T=JS&PAGE=reference&D=emexc&NEWS=N&AN=633598939 |
| 50 | Couzin-Frankel J.. How an immunologist pivoted to tackle COVID-19. Science. 2020; 370(6516): 514. doi: http://dx.doi.org/10.1126/SCIENCE.370.6516.514. Available at: https://www.sciencemag.org/lookup/doi/10.1126/science.370.6516.514; http://ovidsp.ovid.com/ovidweb.cgi?T=JS&PAGE=reference&D=emexc&NEWS=N&AN=2010115006 |
| 51 | Liu M., Maxwell C.J., Armstrong P., Schwandt M., Moser A., McGregor M.J., Bronskill S.E., Dhalla I.A.. COVID-19 in long-term care homes in Ontario and British Columbia. CMAJ. 2020; 192(47): E1540. doi: http://dx.doi.org/10.1503/cmaj.201860. Available at: https://www.cmaj.ca/content/192/47/E1540; http://ovidsp.ovid.com/ovidweb.cgi?T=JS&PAGE=reference&D=emexc&NEWS=N&AN=2010096962 |
| 52 | De Wilton A., Kilich E., Chaudhry Z., Bell L.C.K., Gahir J., Cadman J., Lever R.A., Logan S.A.. Delayed healthcare seeking and prolonged illness in healthcare workers during the COVID-19 pandemic: A single-centre observational study. BMJ Open. 2020; 10(11): e040216. doi: http://dx.doi.org/10.1136/bmjopen-2020-040216. Available at: http://bmjopen.bmj.com/content/early/by/section; http://ovidsp.ovid.com/ovidweb.cgi?T=JS&PAGE=reference&D=emexc&NEWS=N&AN=633518091 |
| 53 | Robecchi Majnardi A., Malfitano C., Tesio L.. COVID-19 pandemic: why time-dependent rehabilitation is forgotten. International journal of rehabilitation research.Internationale Zeitschrift fur Rehabilitationsforschung.Revue internationale de recherches de readaptation. 2020; #volume#(#issue#): #pages#. doi: http://dx.doi.org/10.1097/MRR.0000000000000448. Available at: http://ovidsp.ovid.com/ovidweb.cgi?T=JS&PAGE=reference&D=emexc&NEWS=N&AN=633557489 |
| 54 | Lin C.-H., Yang C.-Y., Wang M., Ou S.-C., Lo C.-Y., Tsai T.-L., Wu H.-Y.. Effects of coronavirus persistence on the genome structure and subsequent gene expression, pathogenicity and adaptation capability. Cells. 2020; 9(10): 1. doi: http://dx.doi.org/10.3390/cells9102322. Available at: https://www.mdpi.com/2073-4409/9/10/2322/pdf; http://ovidsp.ovid.com/ovidweb.cgi?T=JS&PAGE=reference&D=emexc&NEWS=N&AN=2005248914 |
| 55 | Valizadeh H., Abdolmohammadi-vahid S., Danshina S., Ziya Gencer M., Ammari A., Sadeghi A., Roshangar L., Aslani S., Esmaeilzadeh A., Ghaebi M., Valizadeh S., Ahmadi M.. Nano-curcumin therapy, a promising method in modulating inflammatory cytokines in COVID-19 patients. International immunopharmacology. 2020; 89(#issue#): 107088. doi: http://dx.doi.org/10.1016/j.intimp.2020.107088. Available at: http://www.elsevier.com/locate/intimp; http://ovidsp.ovid.com/ovidweb.cgi?T=JS&PAGE=reference&D=emexc&NEWS=N&AN=2008366882 |
| 56 | Boyle E.C., Haverich A.. Microvasculature dysfunction as the common thread between atherosclerosis, Kawasaki disease, and severe acute respiratory syndrome coronavirus 2 (SARS-CoV-2)-associated multi-system inflammatory syndrome in children. European journal of cardio-thoracic surgery : official journal of the European Association for Cardio-thoracic Surgery. 2020; 58(6): 1109. doi: |

| | |
|---|---|
| | http://dx.doi.org/10.1093/ejcts/ezaa367. Available at: http://ovidsp.ovid.com/ovidweb.cgi?T=JS&PAGE=reference&D=emexc&NEWS=N&AN=633548673 |
| 57 | Tincati C., Cannizzo E.S., Giacomelli M., Badolato R., d'Arminio Monforte A., Marchetti G.. Heightened Circulating Interferon-Inducible Chemokines, and Activated Pro-Cytolytic Th1-Cell Phenotype Features Covid-19 Aggravation in the Second Week of Illness. Frontiers in Immunology. 2020; 11(#issue#): 580987. doi: http://dx.doi.org/10.3389/fimmu.2020.580987. Available at: https://www.frontiersin.org/journals/immunology#; http://ovidsp.ovid.com/ovidweb.cgi?T=JS&PAGE=reference&D=emexc&NEWS=N&AN=633280379 |
| 58 | Shah S.J., Barish P.N., Prasad P.A., Kistler A., Neff N., Kamm J., Li L.M., Chiu C.Y., Babik J.M., Fang M.C., Abe-Jones Y., Alipanah N., Alvarez F.N., Botvinnik O.B., Castaneda G., Dadasovich R.M., Davis J., Deng X., DeRisi J.L., Detweiler A.M., Federman S., Haliburton J., Hao S., Kerkhoff A.D., Kumar G.R., Malcolm K.B., Mann S.A., Martinez S., Mary R.K., Mick E., Mwakibete L., Najafi N., Peluso M.J., Phelps M., Pisco A.O., Ratnasiri K., Rubio L.A., Sellas A., Sherwood K.D., Sheu J., Spottiswoode N., Tan M., Yu G., Kangelaris K.N., Langelier C.. Clinical features, diagnostics, and outcomes of patients presenting with acute respiratory illness: A retrospective cohort study of patients with and without COVID-19. EClinicalMedicine. 2020; 27(#issue#): 100518. doi: http://dx.doi.org/10.1016/j.eclinm.2020.100518. Available at: http://www.thelancet.com/journals/eclinm/issue/current; http://ovidsp.ovid.com/ovidweb.cgi?T=JS&PAGE=reference&D=emexc&NEWS=N&AN=2007593804 |
| 59 | Kupferschmidt K.. Global trial eliminates drugs, pivots to new ones. Science. 2020; 370(6515): 388. doi: http://dx.doi.org/10.1126/science.370.6515.388. Available at: https://science.sciencemag.org/content/370/6515/388; http://ovidsp.ovid.com/ovidweb.cgi?T=JS&PAGE=reference&D=emexc&NEWS=N&AN=2010074469 |
| 60 | Ayers J.C.. My last drop. Science. 2020; 370(6514): 374. doi: http://dx.doi.org/10.1126/science.370.6514.374. Available at: https://science.sciencemag.org/content/370/6514/374; http://ovidsp.ovid.com/ovidweb.cgi?T=JS&PAGE=reference&D=emexc&NEWS=N&AN=2010070226 |
| 61 | Chiappini E., Licari A., Motisi M.A., Manti S., Marseglia G.L., Galli L., Lionetti P.. Gastrointestinal involvement in children with SARS-COV-2 infection: An overview for the pediatrician. Pediatric Allergy and Immunology. 2020; 31(#issue#): 92. doi: http://dx.doi.org/10.1111/pai.13373. Available at: http://onlinelibrary.wiley.com/journal/10.1111/(ISSN)1399-3038; http://ovidsp.ovid.com/ovidweb.cgi?T=JS&PAGE=reference&D=emexc&NEWS=N&AN=2007369711 |
| 62 | Bekelis K., Missios S., Ahmad J., Labropoulos N., Schirmer C.M., Calnan D.R., Skinner J., MacKenzie T.A.. Ischemic Stroke Occurs Less Frequently in Patients with COVID-19: A Multicenter Cross-Sectional Study. Stroke. 2020; #volume#(#issue#): 3570. doi: http://dx.doi.org/10.1161/STROKEAHA.120.031217. Available at: https://www.ahajournals.org/journal/str; http://ovidsp.ovid.com/ovidweb.cgi?T=JS&PAGE=reference&D=emexc&NEWS=N&AN=633516225 |
| 63 | Uygun O., Ertas M., Ekizoglu E., Bolay H., Ozge A., Kocasoy Orhan E., Cagatay A.A., Baykan B.. Headache characteristics in COVID-19 pandemic-a survey study. Journal of Headache and Pain. 2020; 21(1): 121. doi: http://dx.doi.org/10.1186/s10194-020-01188-1. Available at: https://thejournalofheadacheandpain.biomedcentral.com/; http://ovidsp.ovid.com/ovidweb.cgi?T=JS&PAGE=reference&D=emexc&NEWS=N&AN=2007010137 |
| 64 | Kamal M., Abo Omirah M., Hussein A., Saeed H.. Assessment and characterisation of post-COVID-19 manifestations. International journal of clinical practice. 2020; #volume#(#issue#): #pages#. doi: http://dx.doi.org/10.1111/ijcp.13746. Available at: http://onlinelibrary.wiley.com/journal/10.1111/(ISSN)1742-1241; http://ovidsp.ovid.com/ovidweb.cgi?T=JS&PAGE=reference&D=emexc&NEWS=N&AN=2007417284 |
| 65 | Madariaga M.L.L., Guthmiller J.J., Schrantz S., Jansen M.O., Christensen C., Kumar M., Prochaska M., Wool G., Durkin-Celauro A., Oh W.H., Trockman L., Vigneswaran J., Keskey R., Shaw D.G., Dugan H., Zheng N.-Y., Cobb M., Utset H., Wang J., Stovicek O., Bethel C., Matushek S., Giurcanu M., Beavis K.G., di Sabato D., Meltzer D., Ferguson M.K., Kress J.P., Shanmugarajah K., Matthews J.B., Fung J.F., Wilson P.C., Alverdy J.C., Donington J.S.. Clinical predictors of donor antibody titre and correlation with recipient antibody |

| | |
|---|---|
| | response in a COVID-19 convalescent plasma clinical trial. Journal of internal medicine. 2020; #volume#(#issue#): #pages#. doi: http://dx.doi.org/10.1111/joim.13185. Available at: http://onlinelibrary.wiley.com/journal/10.1111/(ISSN)1365-2796; http://ovidsp.ovid.com/ovidweb.cgi?T=JS&PAGE=reference&D=emexc&NEWS=N&AN=2007393354 |
| 66 | Park S., Elliott J., Berlin A., Hamer-Hunt J., Haines A.. Strengthening the UK primary care response to covid-19. The BMJ. 2020; 370(#issue#): m3691. doi: http://dx.doi.org/10.1136/bmj.m3691. Available at: http://www.bmj.com/; http://ovidsp.ovid.com/ovidweb.cgi?T=JS&PAGE=reference&D=emexc&NEWS=N&AN=632938899 |
| 67 | Mitrani R.D., Dabas N., Goldberger J.J.. COVID-19 cardiac injury: Implications for long-term surveillance and outcomes in survivors. Heart Rhythm. 2020; 17(11): 1984. doi: http://dx.doi.org/10.1016/j.hrthm.2020.06.026. Available at: http://www.elsevier.com/inca/publications/store/7/0/2/3/3/3/index.htt; http://ovidsp.ovid.com/ovidweb.cgi?T=JS&PAGE=reference&D=emexc&NEWS=N&AN=2007501131 |
| 68 | Ganz-Lord F., Segal K.R., Rinke M.L.. Title: Covid-19 symptoms, duration, and prevalence among healthcare workers in the New York metropolitan area. Infection Control and Hospital Epidemiology. 2020; #volume#(#issue#): #pages#. doi: http://dx.doi.org/10.1017/ice.2020.1334. Available at: http://www.journals.uchicago.edu/ICHE/home.html; http://ovidsp.ovid.com/ovidweb.cgi?T=JS&PAGE=reference&D=emexc&NEWS=N&AN=633483569 |
| 69 | Palermo G., Tommasini L., Baldacci F., Del Prete E., Siciliano G., Ceravolo R.. Impact of Coronavirus Disease 2019 Pandemic on Cognition in Parkinson's Disease. Movement Disorders. 2020; 35(10): 1717. doi: http://dx.doi.org/10.1002/mds.28254. Available at: http://onlinelibrary.wiley.com/journal/10.1002/(ISSN)1531-8257; http://ovidsp.ovid.com/ovidweb.cgi?T=JS&PAGE=reference&D=emexc&NEWS=N&AN=2006096229 |
| 70 | Li Y., Li M., Wang M., Zhou Y., Chang J., Xian Y., Wang D., Mao L., Jin H., Hu B.. Acute cerebrovascular disease following COVID-19: a single center, retrospective, observational study. Stroke and Vascular Neurology. 2020; 5(3): 279. doi: http://dx.doi.org/10.1136/svn-2020-000431. Available at: http://svn.bmj.com; http://ovidsp.ovid.com/ovidweb.cgi?T=JS&PAGE=reference&D=emexc&NEWS=N&AN=632392214 |
| 71 | Hansen C.B., Jarlhelt I., Perez-Alos L., Hummelshoj Landsy L., Loftager M., Rosbjerg A., Helgstrand C., Bjelke J.R., Egebjerg T., Jardine J.G., Svaerke Jorgensen C., Iversen K., Bayarri-Olmos R., Garred P., Skjoedt M.-O.. SARS-CoV-2 Antibody Responses Are Correlated to Disease Severity in COVID-19 Convalescent Individuals. Journal of immunology (Baltimore, Md.: 1950). 2020; #volume#(#issue#): #pages#. doi: http://dx.doi.org/10.4049/jimmunol.2000898. Available at: http://ovidsp.ovid.com/ovidweb.cgi?T=JS&PAGE=reference&D=emexc&NEWS=N&AN=633467064 |
| 72 | Qayed E., Deshpande A.R., Elmunzer B.J.. Low incidence of severe gastrointestinal complications in COVID-19 patients admitted to the intensive care unit: a large, multi-center study. Gastroenterology. 2020; #volume#(#issue#): #pages#. doi: http://dx.doi.org/10.1053/j.gastro.2020.11.009. Available at: http://ovidsp.ovid.com/ovidweb.cgi?T=JS&PAGE=reference&D=emexc&NEWS=N&AN=633420006 |
| 73 | Metkus T.S., Sokoll L.J., Barth A.S., Czarny M.J., Hays A.G., Lowenstein C.J., Michos E.D., Nolley E.P., Post W.S., Resar J.R., Thiemann D.R., Trost J.C., Hasan R.K.. Myocardial Injury in Severe COVID-19 Compared to Non-COVID Acute Respiratory Distress Syndrome. Circulation. 2020; #volume#(#issue#): #pages#. doi: http://dx.doi.org/10.1161/CIRCULATIONAHA.120.050543. Available at: http://ovidsp.ovid.com/ovidweb.cgi?T=JS&PAGE=reference&D=emexc&NEWS=N&AN=633418188 |
| 74 | Vahia V.N., Shah A.B.. COVID-19 pandemic and mental health care of older adults in India. International Psychogeriatrics. 2020; 32(10): 1125. doi: http://dx.doi.org/10.1017/S1041610220001441. Available at: http://journals.cambridge.org/action/displayJournal?jid=IPG; http://ovidsp.ovid.com/ovidweb.cgi?T=JS&PAGE=reference&D=emexc&NEWS=N&AN=632753527 |
| 75 | Schulien I., Kemming J., Oberhardt V., Wild K., Seidel L.M., Killmer S., Sagar, Daul F., Salvat Lago M., Decker A., Luxenburger H., Binder B., Bettinger D., Sogukpinar O., Rieg S., Panning M., Huzly D., Schwemmle M., Kochs G., Waller C.F., Nieters A., Duerschmied D., Emmerich F., Mei H.E., Schulz A.R., Llewellyn-Lacey S., Price D.A., Boettler T., Bengsch B., Thimme R., Hofmann M., Neumann-Haefelin C.. |

| | |
|---|---|
| | Characterization of pre-existing and induced SARS-CoV-2-specific CD8+ T cells. Nature medicine. 2020; #volume#(#issue#): #pages#. doi: http://dx.doi.org/10.1038/s41591-020-01143-2. Available at: http://www.nature.com/nm/index.html; http://ovidsp.ovid.com/ovidweb.cgi?T=JS&PAGE=reference&D=emexc&NEWS=N&AN=2007250614 |
| 76 | Forni R., Besana T., Amitrano A., Voinea C., Ogna A.. Ventilatory weaning and early rehabilitation in COVID-19-related acute respiratory distress syndrome: the experience at Locarno hospital, canton of Ticino, Switzerland. Swiss medical weekly. 2020; 150(#issue#): w20397. doi: http://dx.doi.org/10.4414/smw.2020.20397. Available at: http://ovidsp.ovid.com/ovidweb.cgi?T=JS&PAGE=reference&D=emexc&NEWS=N&AN=633410500 |
| 77 | Cobb N.L., Sathe N.A., Duan K.I., Seitz K.P., Thau M.R., Sung C.C., Morrell E.D., Mikacenic C., Kim H.N., Liles W.C., Luks A.M., Town J., Pipavath S., Wurfel M.M., Hough C.L., West T.E., Bhatraju P.K.. Comparison of Clinical Features and Outcomes in Critically Ill Patients Hospitalized with COVID-19 versus Influenza. Annals of the American Thoracic Society. 2020; #volume#(#issue#): #pages#. doi: http://dx.doi.org/10.1513/AnnalsATS.202007-805OC. Available at: http://ovidsp.ovid.com/ovidweb.cgi?T=JS&PAGE=reference&D=emexc&NEWS=N&AN=633412338 |
| 78 | Paneroni M., Simonelli C., Saleri M., Bertacchini L., Venturelli M., Troosters T., Ambrosino N., Vitacca M.. Muscle strength and physical performance in patients without previous disabilities recovering from COVID-19 pneumonia. American journal of physical medicine & rehabilitation. 2020; #volume#(#issue#): #pages#. doi: http://dx.doi.org/10.1097/PHM.0000000000001641. Available at: http://ovidsp.ovid.com/ovidweb.cgi?T=JS&PAGE=reference&D=emexc&NEWS=N&AN=633412193 |
| 79 | Brotons P., Launes C., Buetas E., Fumado V., Henares D., de Sevilla M.F., Redin A., Fuente-Soro L., Cuadras D., Mele M., Jou C., Millat P., Jordan I., Garcia-Garcia J.J., Bassat Q., Munoz-Almagro C.. Susceptibility to Sars-COV-2 Infection Among Children And Adults: A Seroprevalence Study of Family Households in the Barcelona Metropolitan Region, Spain. Clinical infectious diseases : an official publication of the Infectious Diseases Society of America. 2020; #volume#(#issue#): #pages#. doi: http://dx.doi.org/10.1093/cid/ciaa1721. Available at: http://ovidsp.ovid.com/ovidweb.cgi?T=JS&PAGE=reference&D=emexc&NEWS=N&AN=633410392 |
| 80 | Fragoso G., Sciutto E.. GK1: An Alternative Treatment to Control the Respiratory Complications During COVID19. Archives of Medical Research. 2020; #volume#(#issue#): #pages#. doi: http://dx.doi.org/10.1016/j.arcmed.2020.10.019. Available at: http://www.elsevier.com/locate/arcmed; http://ovidsp.ovid.com/ovidweb.cgi?T=JS&PAGE=reference&D=emexc&NEWS=N&AN=2008500956 |
| 81 | Michie S., West R., Harvey N.. The concept of "fatigue" in tackling covid-19. The BMJ. 2020; 371(#issue#): m4171. doi: http://dx.doi.org/10.1136/bmj.m4171. Available at: http://www.bmj.com/; http://ovidsp.ovid.com/ovidweb.cgi?T=JS&PAGE=reference&D=emexc&NEWS=N&AN=633310774 |
| 82 | Iob E., Steptoe A., Fancourt D.. Abuse, self-harm and suicidal ideation in the UK during the COVID-19 pandemic. British Journal of Psychiatry. 2020; 217(4): 543. doi: http://dx.doi.org/10.1192/bjp.2020.130. Available at: https://www.cambridge.org/core/journals/the-british-journal-of-psychiatry/all-issues; http://ovidsp.ovid.com/ovidweb.cgi?T=JS&PAGE=reference&D=emexc&NEWS=N&AN=633039418 |
| 83 | Damodaran S., Joshi S.S., Kumar, V. S., Natarajan P., Patangi S.O., Kumaran T.. COVID Convalescence-A Boon or Bane in Cardiac Surgery?: A "Second Hit" Hypothesis. Journal of cardiothoracic and vascular anesthesia. 2020; #volume#(#issue#): #pages#. doi: http://dx.doi.org/10.1053/j.jvca.2020.10.021. Available at: http://www.journals.elsevier.com/journal-of-cardiothoracic-and-vascular-anesthesia/; http://ovidsp.ovid.com/ovidweb.cgi?T=JS&PAGE=reference&D=emexc&NEWS=N&AN=2008466887 |
| 84 | Savastano A., Crincoli E., Savastano M.C., Younis S., Gambini G., De Vico U., Cozzupoli G.M., Culiersi C., Rizzo S.. Peripapillary retinal vascular involvement in early post-covid-19 patients. Journal of Clinical Medicine. 2020; 9(9): 1. doi: http://dx.doi.org/10.3390/jcm9092895. Available at: https://www.mdpi.com/2077-0383/9/9/2895/pdf; http://ovidsp.ovid.com/ovidweb.cgi?T=JS&PAGE=reference&D=emexc&NEWS=N&AN=2005040320 |
| 85 | Akhter N., Ahmad S., Alzahrani F.A., Dar S.A., Wahid M., Haque S., Bhatia K., Almalki S.S.R., Alharbi R.A., Sindi A.A.-A.. Impact of COVID-19 on the cerebrovascular system and the prevention of RBC lysis. European review for medical and pharmacological sciences. 2020; 24(19): 10267. doi: |

| | |
|---|---|
| | http://dx.doi.org/10.26355/eurrev_202010_23251. Available at: https://www.europeanreview.org/article/23251; http://ovidsp.ovid.com/ovidweb.cgi?T=JS&PAGE=reference&D=emexc&NEWS=N&AN=2008451966 |
| 86 | Shiers S., Ray P.R., Wangzhou A., Sankaranarayanan I., Tatsui C.E., Rhines L.D., Li Y., Uhelski M.L., Dougherty P.M., Price T.J.. ACE2 and SCARF expression in human dorsal root ganglion nociceptors: implications for SARS-CoV-2 virus neurological effects. Pain. 2020; 161(11): 2494. doi: http://dx.doi.org/10.1097/j.pain.0000000000002051. Available at: http://ovidsp.ovid.com/ovidweb.cgi?T=JS&PAGE=reference&D=emexc&NEWS=N&AN=632676270 |
| 87 | Tauben D.J., Langford D.J., Sturgeon J.A., Rundell S.D., Towle C., Bockman C., Nicholas M.. Optimizing telehealth pain care after COVID-19. Pain. 2020; 161(11): 2437. doi: http://dx.doi.org/10.1097/j.pain.0000000000002048. Available at: http://ovidsp.ovid.com/ovidweb.cgi?T=JS&PAGE=reference&D=emexc&NEWS=N&AN=632675786 |
| 88 | Shi W., Liu X., Cao Q., Ma P., Le W., Xie L., Ye J., Wen W., Tang H., Su W., Zheng Y., Liu Y.. High-dimensional single-cell analysis reveals the immune characteristics of COVID-19. American journal of physiology.Lung cellular and molecular physiology. 2020; #volume#(#issue#): #pages#. doi: http://dx.doi.org/10.1152/ajplung.00355.2020. Available at: http://ovidsp.ovid.com/ovidweb.cgi?T=JS&PAGE=reference&D=emexc&NEWS=N&AN=633343306 |
| 89 | Zhao H.-M., Xie Y.-X., Wang C.. Recommendations for respiratory rehabilitation in adults with coronavirus disease 2019. Chinese medical journal. 2020; 133(13): 1595. doi: http://dx.doi.org/10.1097/CM9.0000000000000848. Available at: https://journals.lww.com/cmj/pages/default.aspx; http://ovidsp.ovid.com/ovidweb.cgi?T=JS&PAGE=reference&D=emexc&NEWS=N&AN=633299476 |
| 90 | Fathi F., Sami R., Mozafarpoor S., Hafezi H., Motedayyen H., Arefnezhad R., Eskandari N.. Immune system changes during COVID-19 recovery play key role in determining disease severity. International Journal of Immunopathology and Pharmacology. 2020; 34(#issue#): 2058738420966497. doi: http://dx.doi.org/10.1177/2058738420966497. Available at: http://ovidsp.ovid.com/ovidweb.cgi?T=JS&PAGE=reference&D=emexc&NEWS=N&AN=633214555 |
| 91 | Files J.K., Boppana S., Perez M.D., Sarkar S., Lowman K.E., Qin K., Sterrett S., Carlin E., Bansal A., Sabbaj S., Long D.M., Kutsch O., Kobie J., Goepfert P., Erdmann N.. Sustained cellular immune dysregulation in individuals recovering from SARS-CoV-2 infection. The Journal of clinical investigation. 2020; #volume#(#issue#): #pages#. doi: http://dx.doi.org/10.1172/JCI140491. Available at: http://ovidsp.ovid.com/ovidweb.cgi?T=JS&PAGE=reference&D=emexc&NEWS=N&AN=633299589 |
| 92 | Azizi S.A., Azizi S.-A.. Neurological injuries in COVID-19 patients: direct viral invasion or a bystander injury after infection of epithelial/endothelial cells. Journal of neurovirology. 2020; 26(5): 631. doi: http://dx.doi.org/10.1007/s13365-020-00903-7. Available at: https://rd.springer.com/journal/13365; http://ovidsp.ovid.com/ovidweb.cgi?T=JS&PAGE=reference&D=emexc&NEWS=N&AN=2006064524 |
| 93 | An J., Liao X., Xiao T., Qian S., Yuan J., Ye H., Qi F., Shen C., Wang L., Cheng X., Li N., Wang F., Chen J., Li G., Cai Q., Liu Y., Wang Y., Zhang F., Fu Y., He Q., Tan X., Liu L., Zhang Z.. Clinical characteristics of recovered COVID-19 patients with re-detectable positive RNA test. Annals of Translational Medicine. 2020; 8(17): 1084. doi: http://dx.doi.org/10.21037/atm-20-5602. Available at: http://atm.amegroups.com/; http://ovidsp.ovid.com/ovidweb.cgi?T=JS&PAGE=reference&D=emexc&NEWS=N&AN=632916643 |
| 94 | Singh N.K.. Letter to the Editor: Myopia Epidemic Post-Coronavirus Disease 2019. Optometry and vision science : official publication of the American Academy of Optometry. 2020; 97(10): 911. doi: http://dx.doi.org/10.1097/OPX.0000000000001591. Available at: http://ovidsp.ovid.com/ovidweb.cgi?T=JS&PAGE=reference&D=emexc&NEWS=N&AN=633287257 |
| 95 | Ashraf O., Young M., Malik K.J., Cheema T.. Systemic Complications of COVID-19. Critical Care Nursing Quarterly. 2020; 43(4): 390. doi: http://dx.doi.org/10.1097/CNQ.0000000000000324. Available at: http://journals.lww.com/ccnq/pages/default.aspx; http://ovidsp.ovid.com/ovidweb.cgi?T=JS&PAGE=reference&D=emexc&NEWS=N&AN=632999788 |

| | |
|---|---|
| 96 | Poletti V., Capozzolo A.. Respiratory Rehabilitation in the COVID-19 Era. Respiration. 2020; 99(6): 461. doi: http://dx.doi.org/10.1159/000509558. Available at: http://www.karger.com/journals/res/res_jh.htm; http://ovidsp.ovid.com/ovidweb.cgi?T=JS&PAGE=reference&D=emexc&NEWS=N&AN=632410113 |
| 97 | Ramos-Rincon J.-M., Buonaiuto V., Ricci M., Martin-Carmona J., Paredes-Ruiz D., Calderon-Moreno M., Rubio-Rivas M., Beato-Perez J.-L., Arnalich-Fernandez F., Monge-Monge D., Vargas-Nunez J.-A., Acebes-Repiso G., Mendez-Bailon M., Perales-Fraile I., Garcia-Garcia G.-M., Guisado-Vasco P., Abdelhady-Kishta A., Pascual-Perez M.-L.-R., Rodriguez-Fernandez-Viagas C., Montano-Martinez A., Lopez-Ruiz A., Gonzalez-Juarez M.-J., Perez-Garcia C., Casas-Rojo J.-M., Gomez-Huelgas R.. Clinical Characteristics and Risk Factors for Mortality in Very Old Patients Hospitalized with COVID-19 in Spain. The journals of gerontology.Series A, Biological sciences and medical sciences. 2020; #volume#(#issue#): #pages#. doi: http://dx.doi.org/10.1093/gerona/glaa243. Available at: http://ovidsp.ovid.com/ovidweb.cgi?T=JS&PAGE=reference&D=emexc&NEWS=N&AN=633270660 |
| 98 | Fisher M.. Cardiovascular disease and cardiovascular outcomes in COVID-19. Practical Diabetes. 2020; 37(5): #pages#. doi: http://dx.doi.org/10.1002/pdi.2294. Available at: http://onlinelibrary.wiley.com/journal/10.1002/(ISSN)1528-252X; http://ovidsp.ovid.com/ovidweb.cgi?T=JS&PAGE=reference&D=emexc&NEWS=N&AN=2006889469 |
| 99 | Cates J., Lucero-Obusan C., Dahl R.M., Schirmer P., Garg S., Oda G., Hall A.J., Langley G., Havers F.P., Holodniy M., Cardemil C.V.. Risk for In-Hospital Complications Associated with COVID-19 and Influenza - Veterans Health Administration, United States, October 1, 2018-May 31, 2020. MMWR.Morbidity and mortality weekly report. 2020; 69(42): 1528. doi: http://dx.doi.org/10.15585/mmwr.mm6942e3. Available at: http://ovidsp.ovid.com/ovidweb.cgi?T=JS&PAGE=reference&D=emexc&NEWS=N&AN=633242270 |
| 100 | Ferroli P., Villa C., Ciuffi A., Gubertini G., Broggi M.. Long lasting hypercoagulability after subclinical COVID-19. Journal of thrombosis and thrombolysis. 2020; 50(4): 822. doi: http://dx.doi.org/10.1007/s11239-020-02215-3. Available at: http://www.wkap.nl/journalhome.htm/0929-5305; http://ovidsp.ovid.com/ovidweb.cgi?T=JS&PAGE=reference&D=emexc&NEWS=N&AN=2005781983 |
| 101 | Kihira S., Schefflein J., Pawha P., Rigney B., Delman B.N., Xuan D., Fifi J., Doshi A.H., Belani P.. Neurovascular complications that can be seen in COVID-19 patients. Clinical imaging. 2021; 69(#issue#): 280. doi: http://dx.doi.org/10.1016/j.clinimag.2020.09.011. Available at: http://www.elsevier.com/locate/clinimag; http://ovidsp.ovid.com/ovidweb.cgi?T=JS&PAGE=reference&D=emexc&NEWS=N&AN=2007994418 |
| 102 | De Lorenzo R., Conte C., Lanzani C., Benedetti F., Roveri L., Mazza M.G., Brioni E., Giacalone G., Canti V., Sofia V., D'Amico M., Di Napoli D., Ambrosio A., Scarpellini P., Castagna A., Landoni G., Zangrillo A., Bosi E., Tresoldi M., Ciceri F., Rovere-Querini P.. Residual clinical damage after COVID-19: A retrospective and prospective observational cohort study. PLoS ONE. 2020; 15(10): e0239570. doi: http://dx.doi.org/10.1371/journal.pone.0239570. Available at: https://journals.plos.org/plosone/article/file?id=10.1371/journal.pone.0239570&type=printable; http://ovidsp.ovid.com/ovidweb.cgi?T=JS&PAGE=reference&D=emexc&NEWS=N&AN=2008347658 |
| 103 | Yang R., Mei H., Zheng T., Fu Q., Buka S., Yao X., Tang Z., Zhang X., Qiu L., Zhang Y., Zhou J., Cao J., Wang Y., Zhou A.. Pregnant women with COVID-19 and risk of adverse birth outcomes and maternal-fetal vertical transmission: a population-based cohort study in Wuhan, China. BMC Medicine. 2020; 18(1): 330. doi: http://dx.doi.org/10.1186/s12916-020-01798-1. Available at: http://www.biomedcentral.com/bmcmed/; http://ovidsp.ovid.com/ovidweb.cgi?T=JS&PAGE=reference&D=emexc&NEWS=N&AN=2007019093 |
| 104 | Hartman W.R., Hess A.S., Connor J.P.. Persistent viral RNA shedding after COVID-19 symptom resolution in older convalescent plasma donors. Transfusion. 2020; 60(10): 2189. doi: http://dx.doi.org/10.1111/trf.15927. Available at: http://onlinelibrary.wiley.com/journal/10.1111/(ISSN)1537-2995; http://ovidsp.ovid.com/ovidweb.cgi?T=JS&PAGE=reference&D=emexc&NEWS=N&AN=2005416011 |
| 105 | Innes S., Skyllberg E., Fielden H., Khan A., Cheng D., Kaiser R., Ainley A.. Late Breaking Abstract - Thromboembolic complications in COVTD-19 patients admitted to two large East London hospitals. |

| | |
|---|---|
| | European Respiratory Journal. 2020; 56(#issue#): #pages#. doi: http://dx.doi.org/10.1183/13993003.congress-2020.4039. Available at: https://erj.ersjournals.com/content/56/suppl_64/4039; http://ovidsp.ovid.com/ovidweb.cgi?T=JS&PAGE=reference&D=emedx&NEWS=N&AN=633804008 |
| 106 | Schiaffino M.T., Di Natale M., Garcia-Martinez E., Navarro J., Munoz-Blanco J.L., Demelo-Rodriguez P., Sanchez-Mateos P.. Immunoserologic Detection and Diagnostic Relevance of Cross-Reactive Autoantibodies in Coronavirus Disease 2019 Patients. Journal of Infectious Diseases. 2020; 222(9): 1439. doi: http://dx.doi.org/10.1093/infdis/jiaa485. Available at: http://jid.oxfordjournals.org/content/current; http://ovidsp.ovid.com/ovidweb.cgi?T=JS&PAGE=reference&D=emexc&NEWS=N&AN=2010130122 |
| 107 | Varatharaj, Aravinthan, Thomas, Naomi, Ellul, Mark A., Davies, Nicholas W. S., Pollak, Thomas A., Tenorio, Elizabeth L., Sultan, Mustafa, Easton, Ava, Breen, Gerome, Zandi, Michael, Coles, Jonathan P., Manji, Hadi, Al-Shahi Salman, Rustam, Menon, David K., Nicholson, Timothy R., Benjamin, Laura A., Carson, Alan, Smith, Craig, Turner, Martin R., Solomon, Tom, Kneen, Rachel, Pett, Sarah L., Galea, Ian, Thomas, Rhys H., Michael, Benedict D.. "Neurological and neuropsychiatric complications of COVID-19 in 153 patients: A UK-wide surveillance study": Correction. The Lancet Psychiatry. 2020; 7(10): e64. doi: http://dx.doi.org/10.1016/S2215-0366%2820%2930320-5; http://dx.doi.org/10.1016/S2215-0366%2820%2930320-5. Available at: http://ovidsp.ovid.com/ovidweb.cgi?T=JS&PAGE=reference&D=psyc17&NEWS=N&AN=2020-71085-030 |
| 108 | Varatharaj, Aravinthan, Thomas, Naomi, Ellul, Mark A., Davies, Nicholas W. S., Pollak, Thomas A., Tenorio, Elizabeth L., Sultan, Mustafa, Easton, Ava, Breen, Gerome, Zandi, Michael, Coles, Jonathan P., Manji, Hadi, Al-Shahi Salman, Rustam, Menon, David K., Nicholson, Timothy R., Benjamin, Laura A., Carson, Alan, Smith, Craig, Turner, Martin R., Solomon, Tom, Kneen, Rachel, Pett, Sarah L., Galea, Ian, Thomas, Rhys H., Michael, Benedict D.. Neurological and neuropsychiatric complications of COVID-19 in 153 patients: A UK-wide surveillance study. The Lancet Psychiatry. 2020; 7(10): 875. doi: http://dx.doi.org/10.1016/S2215-0366%2820%2930287-X; http://dx.doi.org/10.1016/S2215-0366%2820%2930287-X. Available at: http://ovidsp.ovid.com/ovidweb.cgi?T=JS&PAGE=reference&D=psyc17&NEWS=N&AN=2020-71085-023 |
| 109 | Avula, Akshay, Nalleballe, Krishna, Toom, Sudhamshi, Siddamreddy, Suman, Gurala, Dhineshreddy, Katyal, Nakul, Maddika, Srikanth, Polavarapu, Abhishek D., Sharma, Rohan, Onteddu, Sanjeeva. Incidence of Thrombotic Events and Outcomes in COVID-19 Patients Admitted to Intensive Care Units. Cureus. 2020; 12(10): e11079. doi: https://dx.doi.org/10.7759/cureus.11079. Available at: http://ovidsp.ovid.com/ovidweb.cgi?T=JS&PAGE=reference&D=prem6&NEWS=N&AN=33224673 |
| 110 | Sahin, Ayca Sultan, Irak, Kader, Kaya, Ebru, Ozkan, Sureyya, Baskan, Fikret. A thrombotic complication of COVID-19: cerebral ischaemia in the intensive care unit. Archives of medical sciences.Atherosclerotic diseases. 2020; 5(#issue#): e224. doi: https://dx.doi.org/10.5114/amsad.2020.98920. Available at: http://ovidsp.ovid.com/ovidweb.cgi?T=JS&PAGE=reference&D=prem&NEWS=N&AN=33305059 |
| 111 | Victorino, Daniella Balduino, Guimaraes-Marques, Marcia, Nejm, Mariana, Scorza, Fulvio Alexandre, Scorza, Carla Alessandra. COVID-19 and stroke: Red flags for secondary movement disorders?. eNeurologicalSci. 2020; 21(#issue#): 100289. doi: https://dx.doi.org/10.1016/j.ensci.2020.100289. Available at: http://ovidsp.ovid.com/ovidweb.cgi?T=JS&PAGE=reference&D=prem6&NEWS=N&AN=33200103 |
| 112 | Briguglio, Matteo, Porta, Mauro, Zuffada, Francesca, Bona, Alberto R., Crespi, Tiziano, Pino, Fabio, Perazzo, Paolo, Mazzocchi, Marco, Giorgino, Riccardo, De Angelis, Giuseppe, Ielasi, Alfonso, De Blasio, Giuseppe, Turiel, Maurizio. SARS-CoV-2 Aiming for the Heart: A Multicenter Italian Perspective About Cardiovascular Issues in COVID-19. Frontiers in physiology. 2020; 11(#issue#): 571367. doi: https://dx.doi.org/10.3389/fphys.2020.571367. Available at: http://ovidsp.ovid.com/ovidweb.cgi?T=JS&PAGE=reference&D=prem6&NEWS=N&AN=33240098 |
| 113 | Brucki, Sonia M. D., Corazza, Luiza A., de Queiroz, Apolonio,P., Barros, Maraysa P., Tatsch, Joao F. S., Riso, Ivy L., Batista, Nardiel A., Manfroi, Gregori, Sawada, Luis A., Batista, Luana L. R., Baldocchi, Meire Argentoni, de Freitas, Julian,L., Aidar, Mariana B., Rocha, Maria S. G.. Neurological complications in COVID-19 patients from Latin America. Brain : a journal of neurology. 2020; #volume#(#issue#): #pages#. |

| | |
|---|---|
| | doi: https://dx.doi.org/10.1093/brain/awaa440. Available at: http://ovidsp.ovid.com/ovidweb.cgi?T=JS&PAGE=reference&D=medp&NEWS=N&AN=33347532 |
| 114 | Connor-Schuler, Randi, Wong, A. I., Shah, Anand, Fiza, Babar, Lyle, Melissa, Ramonell, Richard, Hockstein, Maxwell, Chang, George, Markham, David, Searles, Charles, McDaniel, Michael, Baer, Jefferson, Powell, Cindy, Daniels, Lisa. Experience With Cardiology-Oriented Outcomes in Critically Ill Patients With Coronavirus Disease 2019. Critical care explorations. 2020; 2(12): e0288. doi: https://dx.doi.org/10.1097/CCE.0000000000000288. Available at: http://ovidsp.ovid.com/ovidweb.cgi?T=JS&PAGE=reference&D=prem&NEWS=N&AN=33294847 |
| 115 | Filippetti, Laura, Pace, Nathalie, Marie, Pierre-Yves. Cardiac Involvement After Recovering From COVID-19. JAMA cardiology. 2020; #volume#(#issue#): #pages#. doi: https://dx.doi.org/10.1001/jamacardio.2020.5279. Available at: http://ovidsp.ovid.com/ovidweb.cgi?T=JS&PAGE=reference&D=medp&NEWS=N&AN=33112937 |
| 116 | Fisher, Kiva A., Olson, Samantha M., Tenforde, Mark W., Self, Wesley H., Wu, Michael, Lindsell, Christopher J., Shapiro, Nathan I., Files, D. C., Gibbs, Kevin W., Erickson, Heidi L., Prekker, Matthew E., Steingrub, Jay S., Exline, Matthew C., Henning, Daniel J., Wilson, Jennifer G., Brown, Samuel M., Peltan, Ithan D., Rice, Todd W., Hager, David N., Ginde, Adit A., Talbot, H. K., Casey, Jonathan D., Grijalva, Carlos G., Flannery, Brendan, Patel, Manish M., Feldstein, Leora R.. Symptoms and recovery among adult outpatients with and without COVID-19 at 11 healthcare facilities-July 2020, United States. Influenza and other respiratory viruses. 2021; #volume#(#issue#): #pages#. doi: https://dx.doi.org/10.1111/irv.12832. Available at: http://ovidsp.ovid.com/ovidweb.cgi?T=JS&PAGE=reference&D=medp&NEWS=N&AN=33405338 |
| 117 | Ganz-Lord, Fran, Segal, Kathryn R., Rinke, Michael L.. COVID-19 symptoms, duration, and prevalence among healthcare workers in the New York metropolitan area. Infection control and hospital epidemiology. 2020; #volume#(#issue#): 1. doi: https://dx.doi.org/10.1017/ice.2020.1334. Available at: http://ovidsp.ovid.com/ovidweb.cgi?T=JS&PAGE=reference&D=medp&NEWS=N&AN=33213542 |
| 118 | Gold, Jeffrey E., Baumgartl, William H., Okyay, Ramazan A., Licht, Warren E., Fidel, Paul L.,Jr, Noverr, Mairi C., Tilley, Larry P., Hurley, David J., Rada, Balazs, Ashford, John W.. Analysis of Measles-Mumps-Rubella (MMR) Titers of Recovered COVID-19 Patients. mBio. 2020; 11(6): #pages#. doi: https://dx.doi.org/10.1128/mBio.02628-20. Available at: http://ovidsp.ovid.com/ovidweb.cgi?T=JS&PAGE=reference&D=medl&NEWS=N&AN=33219096 |
| 119 | Heming, Michael, Li, Xiaolin, Rauber, Saskia, Mausberg, Anne K., Borsch, Anna-Lena, Hartlehnert, Maike, Singhal, Arpita, Lu, I-N, Fleischer, Michael, Szepanowski, Fabian, Witzke, Oliver, Brenner, Thorsten, Dittmer, Ulf, Yosef, Nir, Kleinschnitz, Christoph, Wiendl, Heinz, Stettner, Mark, Meyer Zu Horste, Gerd. Neurological Manifestations of COVID-19 Feature T Cell Exhaustion and Dedifferentiated Monocytes in Cerebrospinal Fluid. Immunity. 2020; #volume#(#issue#): #pages#. doi: https://dx.doi.org/10.1016/j.immuni.2020.12.011. Available at: http://ovidsp.ovid.com/ovidweb.cgi?T=JS&PAGE=reference&D=medp&NEWS=N&AN=33382973 |
| 120 | Lim, Shir Lynn, Woo, Kai Lee, Lim, Eleanor, Ng, Faclin, Chan, Mark Y., Gandhi, Mihir. Impact of COVID-19 on health-related quality of life in patients with cardiovascular disease: a multi-ethnic Asian study. Health and quality of life outcomes. 2020; 18(1): 387. doi: https://dx.doi.org/10.1186/s12955-020-01640-5. Available at: http://ovidsp.ovid.com/ovidweb.cgi?T=JS&PAGE=reference&D=medl&NEWS=N&AN=33317547 |
| 121 | Loftus, Patricia A., Roland, Lauren T., Gurrola, Jose G.,2nd, Cheung, Steven W., Chang, Jolie L.. Temporal Profile of Olfactory Dysfunction in COVID-19. OTO open. 2020; 4(4): 2473974X20978133. doi: https://dx.doi.org/10.1177/2473974X20978133. Available at: http://ovidsp.ovid.com/ovidweb.cgi?T=JS&PAGE=reference&D=prem&NEWS=N&AN=33330830 |
| 122 | Makvandi, Shayan, Alibrahim, Omar, Abdul-Aziz, Rabheh, Abdul-Fattah Sallam, Mohammad, McGreevy, Megan. A Rare Presentation of Multi-System Inflammatory Disease in Children Associated With Severe Acute Respiratory Syndrome Coronavirus 2 (SARS-CoV-2). Cureus. 2020; 12(10): e10892. doi: https://dx.doi.org/10.7759/cureus.10892. Available at: http://ovidsp.ovid.com/ovidweb.cgi?T=JS&PAGE=reference&D=prem6&NEWS=N&AN=33194461 |

| | |
|---|---|
| 123 | McCarthy, Cameron G., Wilczynski, Stephanie, Wenceslau, Camilla F., Webb, R. C.. A new storm on the horizon in COVID-19: Bradykinin-induced vascular complications. Vascular pharmacology. 2020; #volume#(#issue#): 106826. doi: https://dx.doi.org/10.1016/j.vph.2020.106826. Available at: http://ovidsp.ovid.com/ovidweb.cgi?T=JS&PAGE=reference&D=medp&NEWS=N&AN=33358968 |
| 124 | Muller, Jan, Oberhoffer, Renate, Brudy, Leon, Ewert, And Peter. COVID-19 and paediatric patient involvement (cardiovascular aspects). European heart journal supplements : journal of the European Society of Cardiology. 2020; 22(#issue#): P19. doi: https://dx.doi.org/10.1093/eurheartj/suaa168. Available at: http://ovidsp.ovid.com/ovidweb.cgi?T=JS&PAGE=reference&D=prem&NEWS=N&AN=33390865 |
| 125 | Nie, Lei, Dai, Kai, Wu, Jiang, Zhou, Xia, Hu, Junjun, Zhang, Chao, Zhan, Yan, Song, Yu, Fan, Wen, Hu, Zhimin, Yang, Hongshan, Yang, Qiong, Wu, Dongde, Li, Fajiu, Li, Daoyuan, Nie, Rui. Clinical characteristics and risk factors for in-hospital mortality of lung cancer patients with COVID-19: A multicenter, retrospective, cohort study. Thoracic cancer. 2021; 12(1): 57. doi: https://dx.doi.org/10.1111/1759-7714.13710. Available at: http://ovidsp.ovid.com/ovidweb.cgi?T=JS&PAGE=reference&D=prem&NEWS=N&AN=33142039 |
| 126 | Prendki, Virginie, Tau, Noam, Avni, Tomer, Falcone, Marco, Huttner, Angela, Kaiser, Laurent, Paul, Mical, Leibovici-Weissmann, Yaara, Yahav, Dafna, ESCMID Study Group for Infections in the Elderly,(ESGIE). A systematic review assessing the under-representation of elderly adults in COVID-19 trials. BMC geriatrics. 2020; 20(1): 538. doi: https://dx.doi.org/10.1186/s12877-020-01954-5. Available at: http://ovidsp.ovid.com/ovidweb.cgi?T=JS&PAGE=reference&D=medl&NEWS=N&AN=33342426 |
| 127 | Ribeiro, Bruno Niemeyer de Freitas, Marchiori, Edson. Facial palsy as a neurological complication of SARS-CoV-2. Arquivos de Neuro-Psiquiatria. 2020; 78(10): 667. doi: https://dx.doi.org/10.1590/0004-282X20200127. Available at: http://ovidsp.ovid.com/ovidweb.cgi?T=JS&PAGE=reference&D=medl&NEWS=N&AN=33111851 |
| 128 | Rubel, Kolin, Sharma, Dhruv, Campiti, Vincent, Yedlicka, Grace, Burgin, Sarah J., Illing, Elisa A., Kroenke, Kurt, Ting, Jonathan Y.. COVID-19 Status Differentially Affects Olfaction: A Prospective Case-Control Study. OTO open. 2020; 4(4): 2473974X20970176. doi: https://dx.doi.org/10.1177/2473974X20970176. Available at: http://ovidsp.ovid.com/ovidweb.cgi?T=JS&PAGE=reference&D=prem6&NEWS=N&AN=33225199 |
| 129 | Sivalingam, Thivya, Inusa, Baba, Doyle, Pat, Oteng-Ntim, Eugene. COVID-19 and the pulmonary complications of sickle cell disease. EJHaem. 2020; 1(2): 545. doi: https://dx.doi.org/10.1002/jha2.105. Available at: http://ovidsp.ovid.com/ovidweb.cgi?T=JS&PAGE=reference&D=prem6&NEWS=N&AN=33230510 |
| 130 | Uddin, T., Islam, M. T., Rahman, M. A.. COVID-19 Rehabilitation Response in Bangladesh: Ongoing Efforts and Future Directives. Mymensingh medical journal : MMJ. 2021; 30(1): 3. doi: #DOI#. Available at: http://ovidsp.ovid.com/ovidweb.cgi?T=JS&PAGE=reference&D=prem&NEWS=N&AN=33397843 |
| 131 | Vink, Mark, Vink-Niese, Alexandra. Could Cognitive Behavioural Therapy Be an Effective Treatment for Long COVID and Post COVID-19 Fatigue Syndrome? Lessons from the Qure Study for Q-Fever Fatigue Syndrome. Healthcare (Basel, Switzerland). 2020; 8(4): #pages#. doi: https://dx.doi.org/10.3390/healthcare8040552. Available at: http://ovidsp.ovid.com/ovidweb.cgi?T=JS&PAGE=reference&D=prem&NEWS=N&AN=33322316 |
| 132 | Ward, Alex E., Tadross, Daniel, Wells, Fiona, Majkowski, Lawrence, Naveed, Umna, Jeyapalan, Rathan, Partridge, David G., Madan, Suvira, Blundell, Chris M.. The impact of COVID-19 on morbidity and mortality in neck of femur fracture patients: a prospective case-control cohort study. Bone & joint open. 2020; 1(11): 669. doi: https://dx.doi.org/10.1302/2633-1462.111.BJO-2020-0141.R1. Available at: http://ovidsp.ovid.com/ovidweb.cgi?T=JS&PAGE=reference&D=prem&NEWS=N&AN=33263106 |
| 133 | Chen D.E., Tay E.K., Tan P.L., Su P., Koh M.M., Ong P.L., Chew E., Ng Y.S.. The Impact of COVID-19 Pandemic on Rehabilitation in Singapore. Annals of the Academy of Medicine, Singapore. 2020; 49(11): 925. doi: http://dx.doi.org/10.47102/annals-acadmedsg.2020297. Available at: http://ovidsp.ovid.com/ovidweb.cgi?T=JS&PAGE=reference&D=emedx&NEWS=N&AN=633903873 |

| | |
|---|---|
| 134 | Vena A., Taramasso L., Di Biagio A., Mikulska M., Dentone C., De Maria A., Magnasco L., Nicolini L.A., Bruzzone B., Icardi G., Orsi A., Pelosi P., Ball L., Battaglini D., Brunetti I., Loconte M., Patroniti N.A., Robba C., Bavastro M., Cerchiaro M., Giacobbe D.R., Schiavetti I., Berruti M., Bassetti M., Alessandrini A., Camera M., Delfino E., Dodi F., Ferrazin A., Mazzarello G., Nicolini L., Toscanini F., Balletto E., Portunato F., Schenone E., Rosseti N., Baldi F., Briano F., Dettori S., Labate L., Mirabella M., Pincino R., Russo C., Sarteschi G., Sepulcri C., Tutino S., Pontremoli R., Beccati V., Casciaro S., Casu M., Gavaudan F., Ghinatti M., Gualco E., Leoncini G., Pitto P., Salam K.. Prevalence and Clinical Significance of Persistent Viral Shedding in Hospitalized Adult Patients with SARS-CoV-2 Infection: A Prospective Observational Study. Infectious Diseases and Therapy. 2021; #volume#(#issue#): #pages#. doi: http://dx.doi.org/10.1007/s40121-020-00381-8. Available at: http://www.springer.com/springer+healthcare/journal/40121; http://ovidsp.ovid.com/ovidweb.cgi?T=JS&PAGE=reference&D=emedx&NEWS=N&AN=2010142566 |
| 135 | Ghaffari M., Ansari H., Beladimoghadam N., Aghamiri S.H., Haghighi M., Nabavi M., Mansouri B., Mehrpour M., Assarzadegan F., Hesami O., Sedaghat M., Farahbakhsh M., Lima B.S.. Neurological features and outcome in COVID-19: dementia can predict severe disease. Journal of neurovirology. 2021; #volume#(#issue#): #pages#. doi: http://dx.doi.org/10.1007/s13365-020-00918-0. Available at: https://rd.springer.com/journal/13365; http://ovidsp.ovid.com/ovidweb.cgi?T=JS&PAGE=reference&D=emedx&NEWS=N&AN=2010142560 |
| 136 | Collaborative C.O.V.I.D.S.. Preoperative nasopharyngeal swab testing and postoperative pulmonary complications in patients undergoing elective surgery during the SARS-CoV-2 pandemic. European Journal of Surgical Oncology. 2021; 47(1): e4. doi: http://dx.doi.org/10.1016/j.ejso.2020.11.021. Available at: http://ovidsp.ovid.com/ovidweb.cgi?T=JS&PAGE=reference&D=emexc&NEWS=N&AN=2010442509 |
| 137 | Girardi M., Casolo A., Nuccio S., Gattoni C., Capelli C.. Detraining Effects Prevention: A New Rising Challenge for Athletes. Frontiers in Physiology. 2020; 11(#issue#): 588784. doi: http://dx.doi.org/10.3389/fphys.2020.588784. Available at: http://www.frontiersin.org/Physiology/archive/; http://ovidsp.ovid.com/ovidweb.cgi?T=JS&PAGE=reference&D=emexc&NEWS=N&AN=633243335 |
| 138 | Baiocchi G., Aguiar S., Duprat J.P., Coimbra F.J.F., Makdissi F.B., Vartanian J.G., Zequi S.D.C., Gross J.L., Nakagawa S.A., Yazbek G., Diniz T.P., Goncalves B.T., Zurstrassen C.E., Campos H.G.D.A., Joaquim E.H.G., Franca e Silva I.A., Kowalski L.P.. Early postoperative outcomes among patients with delayed surgeries after preoperative positive test for SARS-CoV-2: A case-control study from a single institution. Journal of surgical oncology. 2021; #volume#(#issue#): #pages#. doi: http://dx.doi.org/10.1002/jso.26377. Available at: http://onlinelibrary.wiley.com/journal/10.1002/(ISSN)1096-9098; http://ovidsp.ovid.com/ovidweb.cgi?T=JS&PAGE=reference&D=emexc&NEWS=N&AN=2010153379 |
| 139 | Khodjieva D.T., Kazakov B.Sh., Kadirovna K.D., Kadirovich K.N.. Clinical and neurological factors in the formation of an individual predisposition to COVID-associated ischemic stroke. European Journal of Molecular and Clinical Medicine. 2020; 7(7): 5776. doi: #DOI#. Available at: https://ejmcm.com/pdf_5920_342ba0bbdbf4014fe3c4236d9c721c2d.html; http://ovidsp.ovid.com/ovidweb.cgi?T=JS&PAGE=reference&D=emexc&NEWS=N&AN=2010559014 |
| 140 | Parolari A., di Mauro M., Bonalumi G., Barili F., Garatti A., Carretta G., Donato D., Pagano D., Gerosa G.. Safety for all: Coronavirus disease 2019 pandemic and cardiac surgery: A roadmap to 'phase' 2. European Journal of Cardio-thoracic Surgery. 2020; 58(2): 213. doi: http://dx.doi.org/10.1093/ejcts/ezaa187. Available at: http://ejcts.oxfordjournals.org/; http://ovidsp.ovid.com/ovidweb.cgi?T=JS&PAGE=reference&D=emexc&NEWS=N&AN=633234052 |
| 141 | Beltran-Garcia J., Osca-Verdegal R., Pallardo F.V., Ferreres J., Rodriguez M., Mulet S., Ferrando-Sanchez C., Carbonell N., Garcia-Gimenez J.L.. Sepsis and Coronavirus Disease 2019: Common Features and Anti-Inflammatory Therapeutic Approaches. Critical Care Medicine. 2020; 48(12): 1841. doi: http://dx.doi.org/10.1097/CCM.0000000000004625. Available at: http://ovidsp.ovid.com/ovidweb.cgi?T=JS&PAGE=reference&D=emexc&NEWS=N&AN=632674858 |
| 142 | Pal G.K., Nanda N., Renugasundari M., Pal P., Pachegaonkar U.. Acute effects of prone asanas and pal's pranayama on myalgia, headache, psychological stress and respiratory problems in the covid-19 patients |

| | |
|---|---|
| | in the recovery phase. Biomedicine (India). 2020; 40(4): 526. doi: http://dx.doi.org/10.51248/.v40i4.334. Available at: https://biomedicineonline.org/index.php/home/article/download/334/138; http://ovidsp.ovid.com/ovidweb.cgi?T=JS&PAGE=reference&D=emexc&NEWS=N&AN=2005731065 |
| 143 | Ashok Kumar A., Vellapandi. A review stdy on covid 19 and diarrhoea. European Journal of Molecular and Clinical Medicine. 2020; 7(7): 5652. doi: #DOI#. Available at: https://ejmcm.com/pdf_5863_76efec332f5e0e1ad85473965b1f4629.html; http://ovidsp.ovid.com/ovidweb.cgi?T=JS&PAGE=reference&D=emexc&NEWS=N&AN=2010558997 |
| 144 | Alimov A., Rakhimov R., Alimov U., Azizov A.. Treatment of patients with asymptomatic and mild case of covid-19 infection and its prevention in outpatient basis by using infrared resonance therapy. European Journal of Molecular and Clinical Medicine. 2020; 7(3): 5356. doi: #DOI#. Available at: https://ejmcm.com/pdf_5429_d4d56e3fb6f51a0455a686d8eb2b92ff.html; http://ovidsp.ovid.com/ovidweb.cgi?T=JS&PAGE=reference&D=emexc&NEWS=N&AN=2010558866 |
| 145 | Udwadia Z.F., Vora A., Tripathi A.R., Malu K.N., Lange C., Sara Raju R.. COVID-19 -Tuberculosis interactions: When dark forces collide. Indian Journal of Tuberculosis. 2020; 67(4): S155. doi: http://dx.doi.org/10.1016/j.ijtb.2020.07.003. Available at: http://tbassnindia.org/Archives.html; http://ovidsp.ovid.com/ovidweb.cgi?T=JS&PAGE=reference&D=emexc&NEWS=N&AN=2007328028 |
| 146 | Cui Y., Zhang L., Gao Q., Hou Y., Tian G.. Clinical Characteristics of COVID-19 Elderly Patients and Their Home-based Care after Treatment. Basic and Clinical Pharmacology and Toxicology. 2020; 127(#issue#): 260. doi: http://dx.doi.org/10.1111/bcpt.13461. Available at: http://ovidsp.ovid.com/ovidweb.cgi?T=JS&PAGE=reference&D=emexc&NEWS=N&AN=633830588 |
| 147 | McWilliams D., Weblin J., Hodson J., Veenith T., Whitehouse T., Snelson C.. Rehabilitation Levels in Patients with COVID-19 Admitted to Intensive Care Requiring Invasive Ventilation An Observational Study. Annals of the American Thoracic Society. 2021; 18(1): 122. doi: http://dx.doi.org/10.1513/AnnalsATS.202005-560OC. Available at: https://www.atsjournals.org/doi/pdf/10.1513/AnnalsATS.202005-560OC; http://ovidsp.ovid.com/ovidweb.cgi?T=JS&PAGE=reference&D=emexc&NEWS=N&AN=2010547487 |
| 148 | Mandora, Elena, Comini, Laura, Olivares, Adriana, Fracassi, Michela, Grazia Cadei, Maria, Paneroni, Mara, Marchina, Lucia, Suruniuc, Adrian, Luisa, Alberto, Scalvini, Simonetta, Corica, Giacomo, Vitacca, Michele. Patients recovering from COVID-19 pneumonia in sub-acute care exhibit severe frailty: Role of the nurse assessment. Journal of Clinical Nursing. 2021; #volume#(#issue#): #pages#. doi: https://dx.doi.org/10.1111/jocn.15637. Available at: http://ovidsp.ovid.com/ovidweb.cgi?T=JS&PAGE=reference&D=medp&NEWS=N&AN=33434372 |
| 149 | Muhammad, Sajjad, Fischer, Igor, Naderi, Soheil, Faghih Jouibari, Morteza, Abdolreza, Sheikhrezaei, Karimialavijeh, Ehsan, Aslzadeh, Sara, Mashayekhi, Mahsa, Zojaji, Mohaddeseh, Kahlert, Ulf Dietrich, Hanggi, Daniel. Systemic Inflammatory Index Is a Novel Predictor of Intubation Requirement and Mortality after SARS-CoV-2 Infection. Pathogens (Basel, Switzerland). 2021; 10(1): #pages#. doi: https://dx.doi.org/10.3390/pathogens10010058. Available at: http://ovidsp.ovid.com/ovidweb.cgi?T=JS&PAGE=reference&D=prem&NEWS=N&AN=33440649 |
| 150 | Gupta, Sandeep, Kaushik, Ashlesha, Kest, Helen, Charles, Alexandra K., De Bruin, William, Colletti, Mario, Goldberg, David. Severe Enteritis as the Sole Manifestation of Novel Coronavirus Disease 2019 (COVID-19) in Adolescent Patients. Case reports in infectious diseases. 2020; 2020(#issue#): 8823622. doi: https://dx.doi.org/10.1155/2020/8823622. Available at: http://ovidsp.ovid.com/ovidweb.cgi?T=JS&PAGE=reference&D=prem&NEWS=N&AN=33425408 |
| 151 | Feitosa, Marley Ribeiro, Parra, Rogerio Serafim, de Camargo, Hugo Parra, Ferreira, Sandro da Costa, Troncon, Luiz Ernesto de Almeida, da Rocha, Jose,Joaquim Ribeiro, Feres, Omar. COVID-19 quarantine measures are associated with negative social impacts and compromised follow-up care in patients with inflammatory bowel disease in Brazil. Annals of gastroenterology. 2021; 34(1): 39. doi: https://dx.doi.org/10.20524/aog.2020.0558. Available at: http://ovidsp.ovid.com/ovidweb.cgi?T=JS&PAGE=reference&D=prem&NEWS=N&AN=33414620 |
| 152 | Bolton KL, Koh Y, Foote MB, Im H, Jee J, Sun CH, Safonov A, Ptashkin R, Moon JH, Lee JY, Jung J, Kang CK, Song K-H, Choe PG, Park WB, Kim HB, Oh M-d, Song H, Kim S, Patel M, Derkach A, Gedvilaite E, Tkachuk |

| | |
|---|---|
| | KA, Braunstein LZ, Gao T, Papaemmanuil E, Babady NE, Pessin MS, Kamboj M, Diaz LA, Ladanyi M, Rauh MJ, Natarajan P, Machiela MJ, Awadalla P, Joseph V, Offit K, Norton L, Berger MF, Levine RL, Kim ES, Kim NJ, Zehir,A.. Clonal hematopoiesis is associated with risk of severe Covid-19 (preprint). Medrxiv. 2020; #volume#(#issue#): 2020.11.25.20233163. doi: 10.1101/2020.11.25.20233163. Available at: http://medrxiv.org/content/early/2020/11/27/2020.11.25.20233163.abstract |
| 153 | Butt AA, Kartha A, Asaad N, Azad AM, Bertollini R, Abou-Samra,A.B.. Impact of COVID-19 upon changes in emergency room visits with chest pain of possible cardiac origin; 7672168 CSR - Reports results. BMC research notes. 2020; 13(1): 539. doi: 10.1186/s13104-020-05381-y. Available at: https://pubmed.ncbi.nlm.nih.gov/33208182 |
| 154 | Di Pietro DA, Luisa A, Vitacca,M.. Patients recovering from COVID-19 pneumonia at a sub-acute admissions unit exhibit profound muscular weakness: a case series. European journal of physical and rehabilitation medicine. 2020; #volume#(#issue#): #pages#. doi: 10.23736/S1973-9087.20.06582-X. Available at: https://pubmed.ncbi.nlm.nih.gov/33215906 CSR - Reports results |
| 155 | S,Malhotra A, Sturgill M, Whitley-Williams P, Lee YH, Esochaghi C, Rajasekhar H, Olson B, Gaur. Pediatric COVID-19 and Appendicitis: a Gut Reaction to SARS-CoV-2?. Pediatric infectious disease journal. 2020; #volume#(#issue#): #pages#. doi: 10.1097/INF.0000000000002998. Available at: https://pubmed.ncbi.nlm.nih.gov/33298761 |
| 156 | Silva AB, Freschi G, Carrera RV, Astolfi RH, Berti FF, Gattas N, Meller,A.. COVID-19 pandemic impact on clinical outcomes of patients with obstructive pyelonephritis; 7680071 CSR - Reports results. International urology and nephrology. 2020; #volume#(#issue#): #pages#. doi: 10.1007/s11255-020-02708-3. Available at: https://pubmed.ncbi.nlm.nih.gov/33219920 |
| 157 | V,Lenehan P, Ramudu E, Venkatakrishnan AJ, Horo J, Badley AD, Morice WG, Soundararajan. Longitudinal lab test analysis confirms pre-existing anemia as a severe risk factor for post-viral clearance hospitalization in COVID-19 patients (preprint). Medrxiv. 2020; #volume#(#issue#): 2020.12.02.20242958. doi: 10.1101/2020.12.02.20242958. Available at: http://medrxiv.org/content/early/2020/12/04/2020.12.02.20242958.abstract |
| 158 | Roth, C., Berat, N., Schnider, A., Graf, C. E.. Feedback on post COVID-19 rehabilitation; 32852173. Revue Medicale Suisse. 2020; 16(703): 1507. doi: #DOI#. Available at: https://www.scopus.com/inward/record.uri?eid=2-s2.0-85089980795&partnerID=40&md5=bafb9bd8a88773e67fa9f76e340b7557 |
| 159 | EUCTR2020-002518-42-ES. EVALUATING THE EFFECT OF PIRFENIDONE COMPARED WITH PLACEBO IN PULMONARY FIBROSIS POST-COVID 19. A RANDOMIZED PHASE-II CLINICAL TRIAL TO EVALUATE THE EFFECT OF PIRFENIDONE COMPARED WITH PLACEBO IN PULMONARY FIBROSIS POST-COVID 19. 2020///; #volume#(#issue#): #pages#. doi: #DOI#. Available at: http://ovidsp.ovid.com/ovidweb.cgi?T=JS&PAGE=reference&D=cctr&NEWS=N&AN=CN-02240595 |
| 160 | .. NewsCAP: Autoantibody reactivity implicated in 'long' COVID-19.. AJN, American Journal of Nursing. 2021/03//; 121(3): 17. doi: 10.1097/01.NAJ.0000737260.41769.52. Available at: http://ovidsp.ovid.com/ovidweb.cgi?T=JS&PAGE=reference&D=emexb&NEWS=N&AN=634382917,https://journals.lww.com/10.1097/01.NAJ.0000737260.41769.52 |
| 161 | Cennamo, Gilda, Triassi, Maria, Reibaldi, Michele, Montorio, Daniela, D'Andrea, Luca, Fallico, Matteo. Optical coherence tomography angiography features in post COVID-19 pneumonia patients: a pilot study. American Journal of Ophthalmology. 2021///; #volume#(#issue#): #pages#. doi: http://dx.doi.org/10.1016/j.ajo.2021.03.015. Available at: http://ovidsp.ovid.com/ovidweb.cgi?T=JS&PAGE=reference&D=emedx&NEWS=N&AN=634689964 |
| 162 | Pandian, Vinciya, Brigham, Emily P, Parker, Ann M, Brodsky, Martin B, Needham, Dale M, Hillel, Alexander T, Levy, Joshua M, Rassekh, Christopher H, Lalwani, Anil K, Brenner, Michael J. COVID-19 survivorship: How otolaryngologist-head and neck surgeons can restore quality of life after critical illness. American Journal of Otolaryngology - Head and Neck Medicine and Surgery. 2021///; 42(3): 102917. doi: http://dx.doi.org/10.1016/j.amjoto.2021.102917. Available at: http://www.elsevier.com/inca/publications/store/6/2/3/1/1/9/index.htt,http://ovidsp.ovid.com/ovidweb.cgi?T=JS&PAGE=reference&D=emexb&NEWS=N&AN=2010877001 |

| | |
|---|---|
| 163 | Leigh, Alexandra E, McCall, Jonathan, Burke, Rebecca V, Rome, Robin, Raines, Amanda M. Predictors of Functional Dependence After COVID-19: A Retrospective Examination Among Veterans. American journal of physical medicine & rehabilitation. 2021///; 100(1): 34. doi: http://dx.doi.org/10.1097/PHM.0000000000001614. Available at: http://ovidsp.ovid.com/ovidweb.cgi?T=JS&PAGE=reference&D=emexb&NEWS=N&AN=633163206 |
| 164 | Landi, Francesco, Carfi, Angelo, Benvenuto, Francesca, Brandi, Vincenzo, Ciciarello, Francesca, Lo Monaco, Maria Rita, Martone, Anna Maria, Napolitano, Carmen, Pagano, Francesco, Paglionico, Annamaria, Petricca, Luca, Rocchi, Sara, Rota, Elisabetta, Salerno, Andrea, Tritto, Marcello, Gremese, Elisa, Bernabei, Roberto. Predictive Factors for a New Positive Nasopharyngeal Swab Among Patients Recovered From COVID-19. American Journal of Preventive Medicine. 2021///; 60(1): 13. doi: http://dx.doi.org/10.1016/j.amepre.2020.08.014. Available at: http://www.elsevier.com/locate/amepre,http://ovidsp.ovid.com/ovidweb.cgi?T=JS&PAGE=reference&D=emexb&NEWS=N&AN=2008023468 |
| 165 | . Timing of surgery following SARS-CoV-2 infection: an international prospective cohort study. Anaesthesia. 2021/03//; #volume#(#issue#): anae.15458. doi: 10.1111/anae.15458. Available at: http://onlinelibrary.wiley.com/journal/10.1111/(ISSN)1365-2044,http://ovidsp.ovid.com/ovidweb.cgi?T=JS&PAGE=reference&D=emexb&NEWS=N&AN=2010728143, https://onlinelibrary.wiley.com/doi/10.1111/anae.15458 |
| 166 | G Cai J, Sun W, Jing W, Wang Y, Gamber M, Fan L, He, Zhou M. Does Post-COVID-19 symptoms exist? A longitudinal study of COVID-19 sequelae in Wenzhou, China; 7934689 CSR - Reports results. Annales Medico-Psychologiques. 2021///; #volume#(#issue#): #pages#. doi: 10.1016/j.amp.2021.03.003. Available at: https://pubmed.ncbi.nlm.nih.gov/33688094 |
| 167 | Zhou, Min, Cai, Jing, Sun, Wenjie, Wu, Jing, Wang, Yidong, Gamber, Michelle, Fan, Lingyan, He, Guiqing. Do post-COVID-19 symptoms exist? A longitudinal study of COVID-19 sequelae in Wenzhou, China. Annales Medico-Psychologiques. 2021///; #volume#(#issue#): #pages#. doi: http://dx.doi.org/10.1016/j.amp.2021.03.003. Available at: http://www.elsevier.com,http://ovidsp.ovid.com/ovidweb.cgi?T=JS&PAGE=reference&D=emexb&NEWS=N&AN=2011418859 |
| 168 | Graham, Edith L, Clark, Jeffrey R, Orban, Zachary S, Lim, Patrick H, Szymanski, April L, Taylor, Carolyn, DiBiase, Rebecca M, Jia, Dan Tong, Balabanov, Roumen, Ho, Sam U, Batra, Ayush, Liotta, Eric M, Koralnik, Igor J. Persistent neurologic symptoms and cognitive dysfunction in non-hospitalized Covid-19 "long haulers". Annals of Clinical and Translational Neurology. 2021///; #volume#(#issue#): #pages#. doi: http://dx.doi.org/10.1002/acn3.51350. Available at: http://onlinelibrary.wiley.com/journal/10.1002/(ISSN)2328-9503,http://ovidsp.ovid.com/ovidweb.cgi?T=JS&PAGE=reference&D=emexb&NEWS=N&AN=2010983360 |
| 169 | Mao, Jian, Dai, Rong, Du, Rui-Chan, Zhu, Yan, Shui, Li-Ping, Luo, Xiao-Hua. Hematologic changes predict clinical outcome in recovered patients with COVID-19. Annals of Hematology. 2021///; 100(3): 675. doi: https://dx.doi.org/10.1007/s00277-021-04426-x. Available at: http://ovidsp.ovid.com/ovidweb.cgi?T=JS&PAGE=reference&D=medl&NEWS=N&AN=33523290 |
| 170 | Mohapatra, Abhisekh. Comment on "Acute Thrombosis of Lower Limbs Arteries in the Acute Phase and After Recovery From COVID19 Infection". Annals of Surgery. 2021///; 273(4): e161. doi: http://dx.doi.org/10.1097/SLA.0000000000004732. Available at: http://ovidsp.ovid.com/ovidweb.cgi?T=JS&PAGE=reference&D=emexb&NEWS=N&AN=634406628 |
| 171 | Taylor, Stephanie Parks, Rios, Aleta, Kowalkowski, Marc A. Translating Post-Sepsis Care to Post-COVID Care: The Case for a Virtual Recovery Program. Annals of the American Thoracic Society. 2021///; #volume#(#issue#): #pages#. doi: http://dx.doi.org/10.1513/AnnalsATS.202006-649IP. Available at: http://ovidsp.ovid.com/ovidweb.cgi?T=JS&PAGE=reference&D=emexb&NEWS=N&AN=634257327 |
| 172 | group Forman J, Brighouse J, Sobande A, McIntosh A, Watterson C, Boot E, Montgomery F, Gilmour I, Tan J, Fogarty MJ, Gomez X, Cheung R, Lillie J, Shivamurthy V, Handforth J, Miller O, PIMS-TS study, Broad J. Post-COVID-19 paediatric inflammatory multisystem syndrome: association of ethnicity, key worker and socioeconomic status with risk and severity; 7977079 CSR - Reports results. Archives of Disease in |

| | |
|---|---|
| | Childhood. 2021///; #volume#(#issue#): #pages#. doi: 10.1136/archdischild-2020-320388. Available at: https://pubmed.ncbi.nlm.nih.gov/33727312 |
| 173 | Leite, Victor Figueiredo, Rampim, Danielle Bianchini, Jorge, Valeria Conceicao, Correia de Lima do Carmo, Maria, Cezarino, Leandro Goncalves, da Rocha, Cleber Nunes, Esper, Rodrigo Barbosa. Persistent symptoms and disability after COVID-19 hospitalization: data from a comprehensive telerehabilitation program. Archives of Physical Medicine and Rehabilitation. 2021///; #volume#(#issue#): #pages#. doi: http://dx.doi.org/10.1016/j.apmr.2021.03.001. Available at: http://ovidsp.ovid.com/ovidweb.cgi?T=JS&PAGE=reference&D=emexb&NEWS=N&AN=634563205 |
| 174 | Paneroni, Mara, Bertacchini, Laura, Simonelli, Carla, Vitacca, Michele, Vogiatzis, Ioannis. Predictors of Low Physical Function in Patients With COVID-19 With Acute Respiratory Failure Admitted to a Subacute Unit. Archives of Physical Medicine and Rehabilitation. 2021///; #volume#(#issue#): #pages#. doi: http://dx.doi.org/10.1016/j.apmr.2020.12.021. Available at: http://www.elsevier.com/inca/publications/store/6/2/3/3/5/4/index.htt,http://ovidsp.ovid.com/ovidweb.cgi?T=JS&PAGE=reference&D=emexb&NEWS=N&AN=2011097654 |
| 175 | Wiertz, Carolina M H, Vints, Wouter A J, Maas, Geert J C M, Rasquin, Sascha M C, van Horn Y., Yvette, Dremmen, Martijn P M, Hemmen, Bena, Verbunt, Jeanine A. COVID-19: patient characteristics in the first phase of post-intensive care rehabilitation. Archives of rehabilitation research and clinical translation. 2021///; #volume#(#issue#): 100108. doi: https://dx.doi.org/10.1016/j.arrct.2021.100108. Available at: http://ovidsp.ovid.com/ovidweb.cgi?T=JS&PAGE=reference&D=medp&NEWS=N&AN=33558860 |
| 176 | Lau HM Jones AY, Lee EW, Siu EH, Hui,D.S., Ng G Y. A randomised controlled trial of the effectiveness of an exercise training program in patients recovering from severe acute respiratory syndrome. Australian journal of physiotherapy. 2005///; 51(4): 213. doi: #DOI#. Available at: http://ovidsp.ovid.com/ovidweb.cgi?T=JS&PAGE=reference&D=cctr&NEWS=N&AN=CN-00553264 |
| 177 | Theoharides, Theoharis C, Cholevas, Christos, Polyzoidis, Konstantinos, Politis, Antonios. Long-COVID syndrome-associated brain fog and chemofog: Luteolin to the rescue. BioFactors (Oxford, England). 2021///; #volume#(#issue#): #pages#. doi: https://dx.doi.org/10.1002/biof.1726. Available at: http://ovidsp.ovid.com/ovidweb.cgi?T=JS&PAGE=reference&D=medp&NEWS=N&AN=33847020 |
| 178 | Donegani, Maria Isabella, Miceli, Alberto, Pardini, Matteo, Bauckneht, Matteo, Chiola, Silvia, Pennone, Michele, Marini, Cecilia, Massa, Federico, Raffa, Stefano, Ferrarazzo, Giulia, Arnaldi, Dario, Sambuceti, Gianmario, Nobili, Flavio, Morbelli, Silvia. Brain Metabolic Correlates of Persistent Olfactory Dysfunction after SARS-Cov2 Infection. Biomedicines. 2021///; 9(3): #pages#. doi: https://dx.doi.org/10.3390/biomedicines9030287. Available at: http://ovidsp.ovid.com/ovidweb.cgi?T=JS&PAGE=reference&D=pmnm&NEWS=N&AN=33808956 |
| 179 | Camille, Simard, Koolian, Maral, Escobar, Diana, Blostein, Mark, Lipes, Jed, Avram, Adelina Teona, Kerzner, Ryan, Mantzanis, Helen, Aguilar, Mateo Porres, Tagalakis, Vicky. Thrombotic complications in a canadian population of critically iii patients with COVID-19. Blood. 2020///; 136(#issue#): 41. doi: http://dx.doi.org/10.1182/blood-2020-141996. Available at: https://ashpublications.org/blood/article/136/Supplement 1/41/470677/Thrombotic-Complications-in-a-Canadian-Population,http://ovidsp.ovid.com/ovidweb.cgi?T=JS&PAGE=reference&D=emexb&NEWS=N&AN=634311270 |
| 180 | O Waizman E, Tzur I, Yusupov A, Shapira Y, Gorelik, Plotnikov G. The prognostic role of functional dependency in older inpatients with COVID-19; 8011046 CSR - Reports results. BMC geriatrics. 2021///; 21(1): 219. doi: 10.1186/s12877-021-02158-1. Available at: https://pubmed.ncbi.nlm.nih.gov/33789578 |
| 181 | Firman, Chloe, Makaronidis, Janine, Magee, Cormac G, Mok, Jessica, Balogun, Nyaladzi, Carnemolla, Alisia, Batterham, Rachel L, Lechner, Matt. Distorted chemosensory perception and female sex associate with persistent smell and/or taste loss in people with SARS-CoV-2 antibodies: a community based cohort study investigating clinical course and resolution of acute smell and/or taste loss in people with and without SARS-CoV-2 antibodies in London, UK. BMC Infectious Diseases. 2021///; 21(1): 221. doi: http://dx.doi.org/10.1186/s12879-021-05927-w. Available at: |

| | |
|---|---|
| | http://www.biomedcentral.com/bmcinfectdis/,http://ovidsp.ovid.com/ovidweb.cgi?T=JS&PAGE=reference&D=emexb&NEWS=N&AN=2010583278 |
| 182 | Rong, Yan, Wang, Fei, Tian, Jinfei, Liang, Xinhua, Wang, Jing, Li, Xiaoli, Zhang, Dandan, Liu, Jing, Zeng, Huadong, Zhou, Yang, Shi, Yi. Clinical and CT features of mild-to-moderate COVID-19 cases after two sequential negative nucleic acid testing results: a retrospective analysis. BMC infectious diseases. 2021///; 21(1): 333. doi: https://dx.doi.org/10.1186/s12879-021-06013-x. Available at: http://ovidsp.ovid.com/ovidweb.cgi?T=JS&PAGE=reference&D=mesx&NEWS=N&AN=33832444 |
| 183 | Kumar MA Arul,J.N., Krishnaswamy M. Post COVID-19 sequelae: venous thromboembolism complicated by lower GI bleed; 7843205 CSR - Reports results. BMJ case reports. 2021///; 14(1): #pages#. doi: 10.1136/bcr-2020-241059. Available at: https://pubmed.ncbi.nlm.nih.gov/33504542 |
| 184 | O'Keefe, James B, Tong, Elizabeth J, O'Keefe, Ghazala D, Tong, David C. Description of symptom course in a telemedicine monitoring clinic for acute symptomatic covid-19: A retrospective cohort study. BMJ Open. 2021///; 11(3): e044154. doi: http://dx.doi.org/10.1136/bmjopen-2020-044154. Available at: http://bmjopen.bmj.com/content/early/by/section,http://ovidsp.ovid.com/ovidweb.cgi?T=JS&PAGE=reference&D=emexb&NEWS=N&AN=634387252 |
| 185 | Wang, Yuanyuan, Mao, Kaiping, Xu, Wenying, Li, Zuomin, Shao, Hui, Zhang, Rui. Clinical study of pulmonary CT lesions and associated bronchiectasis in 115 convalescent patients with novel coronavirus pneumonia (COVID-19) in China. Canadian journal of physiology and pharmacology. 2021///; 99(3): 328. doi: http://dx.doi.org/10.1139/cjpp-2020-0522. Available at: http://www.nrc.ca/cgi-bin/cisti/journals/rp/rp_desy_e?cjpp,http://ovidsp.ovid.com/ovidweb.cgi?T=JS&PAGE=reference&D=emexb&NEWS=N&AN=634568120 |
| 186 | . COVID-19 Prolonged in Patients with Cancer. Cancer Discovery. 2021/02//; #volume#(#issue#): #pages#. doi: 10.1158/2159-8290.CD-NB2021-0319. Available at: http://ovidsp.ovid.com/ovidweb.cgi?T=JS&PAGE=reference&D=medp&NEWS=N&AN=33637485,http://cancerdiscovery.aacrjournals.org/lookup/doi/10.1158/2159-8290.CD-NB2021-0319 |
| 187 | Ciniselli, Chiara Maura, Verderio, Paolo, Micali, Arianna, De Cecco, Loris, Notti, Paola, Sinno, Valentina, Luison, Elena, Daidone, Maria Grazia, Figini, Mariangela, Melani, Cecilia C, Apolone, Giovanni. Sars-cov-2 serology monitoring of a cancer center staff in the pandemic most infected italian region. Cancers. 2021///; 13(5): 1. doi: http://dx.doi.org/10.3390/cancers13051035. Available at: https://www.mdpi.com/2072-6694/13/5/1035/pdf,http://ovidsp.ovid.com/ovidweb.cgi?T=JS&PAGE=reference&D=emexb&NEWS=N&AN=2006124974 |
| 188 | Filipiak, Krzysztof J, Gasecka, Aleksandra, Pruc, Michal, Kukula, Katarzyna, Gilis-Malinowska, Natasza, Jaguszewski, Milosz J, Szarpak, Lukasz. Post-COVID-19 heart syndrome. Cardiology journal. 2021///; #volume#(#issue#): #pages#. doi: http://dx.doi.org/10.5603/CJ.a2021.0028. Available at: http://ovidsp.ovid.com/ovidweb.cgi?T=JS&PAGE=reference&D=emexb&NEWS=N&AN=634439231 |
| 189 | Fatakdawala, Mariya, Pang, Jason, Patel, Ritesh, Thannoun, Tariq, Grable, Cullen, Damaraju, Sarita, Mawji, Shamim B, Folivi, Messan, Chauhan, Siddharth, Powell, Christopher, Shabbir, Muhammad A, Lyubarova, Radmila, Damaraju, Srikanth, Palaskas, Nicolas L, Deswal, Anita M, Taegtmeyer, Heinrich, Garcia-Sayan, Enrique, Koutroumpakis, Efstratios. Electrocardiographic Findings and Their Associations with Adverse Outcomes in Hospitalized Patients with COVID-19. Circulation. 2020///; 142(#issue#): #pages#. doi: http://dx.doi.org/10.1161/circ.142.suppl_3.16107. Available at: http://ovidsp.ovid.com/ovidweb.cgi?T=JS&PAGE=reference&D=emexb&NEWS=N&AN=634379658 |
| 190 | Xie, Mingxing, Sun, Wei, Zhang, Yanting, Wu, Chun, Xie, Yuji, Zhang, Li, Taub, Cynthia C. Incremental Prognostic Value of Biventricular Longitudinal Strain and Hs-tni in Covid-19 Patients. Circulation. 2020///; 142(#issue#): #pages#. doi: http://dx.doi.org/10.1161/circ.142.suppl_3.15561. Available at: http://ovidsp.ovid.com/ovidweb.cgi?T=JS&PAGE=reference&D=emexb&NEWS=N&AN=634380460 |
| 191 | Hesse, Elisabeth M, Paulin, Heather N, Datta, Deblina, Katz, Lee S, Talwar, Amish, Chang, Gregory, Galang, Romeo R, Wong, Karen K, Stevens, Valerie, Kim, Kaylee, Iskander, John, Mitruka, Kiren, Lee, James T, Harcourt, Jennifer L, Tamin, Azaibi, Thornburg, Natalie J, Tong, Suxiang, Zhou, Bin, Queen, Krista, Drobeniuc, Jan, Folster, Jennifer M, Sexton, D J, Ramachandran, Sumathi, Browne, Hannah. Clinical and |

| | |
|---|---|
| | Laboratory Findings in Patients with Potential SARS-CoV-2 Reinfection, May-July 2020. Clinical infectious diseases : an official publication of the Infectious Diseases Society of America. 2021///; #volume#(#issue#): #pages#. doi: http://dx.doi.org/10.1093/cid/ciab148. Available at: http://ovidsp.ovid.com/ovidweb.cgi?T=JS&PAGE=reference&D=emexb&NEWS=N&AN=634334323 |
| 192 | Sheehan, Megan M, Reddy, Anita J, Rothberg, Michael B. Reinfection Rates among Patients who Previously Tested Positive for COVID-19: a Retrospective Cohort Study. Clinical infectious diseases : an official publication of the Infectious Diseases Society of America. 2021///; #volume#(#issue#): #pages#. doi: http://dx.doi.org/10.1093/cid/ciab234. Available at: http://ovidsp.ovid.com/ovidweb.cgi?T=JS&PAGE=reference&D=emexb&NEWS=N&AN=634571078 |
| 193 | Ladds, Emma, Rushforth, Alex, Wieringa, Sietse, Husain, Laiba, Greenhalgh, Trisha, Taylor, Sharon, Rayner, Clare. Erratum: Developing services for long COVID: lessons from a study of wounded healers. Clinical medicine (London, England). 2021///; 21(2): 160. doi: http://dx.doi.org/10.7861/clinmed.err.21.2. Available at: http://ovidsp.ovid.com/ovidweb.cgi?T=JS&PAGE=reference&D=emexb&NEWS=N&AN=634662656 |
| 194 | Bulfone, Luca, Da Porto, Andrea, Colussi, Gianluca, Brosolo, Gabriele, Catena, Cristiana, Peghin, Maddalena, Tascini, Carlo, Patruno, Vincenzo, Sechi, Leonardo A. Short-term cardiac outcome in survivors of COVID-19: a systematic study after hospital discharge. Clinical Research in Cardiology. 2021///; #volume#(#issue#): #pages#. doi: http://dx.doi.org/10.1007/s00392-020-01800-z. Available at: https://link.springer.com/journal/392,http://ovidsp.ovid.com/ovidweb.cgi?T=JS&PAGE=reference&D=emexb&NEWS=N&AN=2010236049 |
| 195 | N Barbieri C, Fontana M, Scelfo C, Castagnetti C, Ghidoni G, Ruggiero P, Livrieri F, Piro R, Ghidorsi L, Montanari G, Gibellini G, Casalini E, Falco F, Catellani C, Facciolongo, Menzella F. Effectiveness of noninvasive ventilation in COVID-19 related-acute respiratory distress syndrome. Clinical respiratory journal. 2021///; #volume#(#issue#): #pages#. doi: 10.1111/crj.13361. Available at: https://pubmed.ncbi.nlm.nih.gov/33728822 |
| 196 | Sieiro-Santos, Cristiana, Lopez-Castro, Jose. Post-coronavirus disease syndrome and disseminated microthrombosis: the role of the von Willebrand factor and antiphospholipid antibodies. Clinics (Sao Paulo, Brazil). 2021///; 76(#issue#): e2784. doi: http://dx.doi.org/10.6061/clinics/2021/e2784. Available at: http://ovidsp.ovid.com/ovidweb.cgi?T=JS&PAGE=reference&D=emedx&NEWS=N&AN=634694256 |
| 197 | Zettersten, Erik, Bell, Max, Jaderling, Gabriella, Martensson, Johan, Engerstrom, Lars, Larsson, Emma, Block, Linda. Long-term outcome after intensive care for COVID-19: differences between men and women-a nationwide cohort study. Critical Care. 2021///; 25(1): 86. doi: http://dx.doi.org/10.1186/s13054-021-03511-x. Available at: http://ccforum.com/content/17,http://ovidsp.ovid.com/ovidweb.cgi?T=JS&PAGE=reference&D=emexb&NEWS=N&AN=2010583119 |
| 198 | DS Chaudhry R, Rai S, Mitra S, Pradhan S, Sunder A, Nag, Ray A. Prolonged Oxygen Therapy Post COVID-19 Infection: factors Leading to the Risk of Poor Outcome; 7899261 CSR - Reports results. Cureus. 2021///; 13(2): e13357. doi: 10.7759/cureus.13357. Available at: https://pubmed.ncbi.nlm.nih.gov/33633916 |
| 199 | R Berwanger O, Koch GG, Martinez F, Mukhtar O, Verma S, Chopra V, Javaheri A, Ambery P, Gasparyan SB, Buenconsejo J, Sjostrom CD, Langkilde AM, Oscarsson J, Esterline, Kosiborod M. Effects of dapagliflozin on prevention of major clinical events and recovery in patients with respiratory failure because of COVID-19: design and rationale for the DARE-19 study. Diabetes, obesity & metabolism. 2020///; #volume#(#issue#): #pages#. doi: #DOI#. Available at: http://ovidsp.ovid.com/ovidweb.cgi?T=JS&PAGE=reference&D=cctr&NEWS=N&AN=CN-02243753 |
| 200 | Parry, Arshed Hussain, Wani, Abdul Haseeb, Jehangir, Majid, Shah, Naveed Nazir. Medium-term chest computed tomography (CT) follow-up of COVID-19 pneumonia patients after recovery to assess the rate of resolution and determine the potential predictors of persistent lung changes. Egyptian Journal of Radiology and Nuclear Medicine. 2021///; 52(1): 55. doi: http://dx.doi.org/10.1186/s43055-021-00434-z. Available at: https://ejrnm.springeropen.com/,http://ovidsp.ovid.com/ovidweb.cgi?T=JS&PAGE=reference&D=emexb&NEWS=N&AN=2010504575 |

| | |
|---|---|
| 201 | Fan, B F, Lee, Y, Lui, L M W, McIntyre, R S, Liao, Y H, Zhang, H M, Guo, L, Wang, W X, Li, W Y, Gong, M Q, Lu, C Y, Li, L J. The impact of COVID-19 on subthreshold depressive symptoms: A longitudinal study. Epidemiology and Psychiatric Sciences. 2021///; #volume#(#issue#): #pages#. doi: http://dx.doi.org/10.1017/S2045796021000044. Available at: http://journals.cambridge.org/action/displayJournal?jid=EPS,http://ovidsp.ovid.com/ovidweb.cgi?T=JS&PAGE=reference&D=emexb&NEWS=N&AN=634210957 |
| 202 | Danoun, Omar A, Zillgitt, Andrew, Hill, Chloe, Harris, David, Rath, Subhendu, Osman, Gamaleldin, Affan, Muhammad, Schultz, Lonni, Zutshi, Deepti, Marawar, Rohit, Syed, Maryam J, Wasade, Vibhangini S. Outcomes of seizures, status epilepticus, and EEG findings in critically ill patient with COVID-19. Epilepsy and Behavior. 2021///; 118(#issue#): 107923. doi: http://dx.doi.org/10.1016/j.yebeh.2021.107923. Available at: http://www.elsevier.com/inca/publications/store/6/2/2/8/2/2/index.htt,http://ovidsp.ovid.com/ovidweb.cgi?T=JS&PAGE=reference&D=emexb&NEWS=N&AN=2011463907 |
| 203 | Fernandez-Rius, Ana, GarcAa, Borja Gil, Corcoles, Ruben Alcantud, Carmona, Laura Plaza, GarcAa, Belen Roldan, Ballesteros, Cristina Gomez, Romero, Marta Mas, Cespedes, Almudena AvendanEo, Utiel, Melisa Lopez, Cifuentes, Sara Celaya, GarcAa, Alicia Nogueron, Molina, Rafael GarcAa, Soler, Pedro Abizanda, Longobardo, Luz MarAa PenEa, Moreno, Juan Oliva. Epidemiological features of residents in long-term care facilities (LTCF) during the outbreak of Covid-19 in Spain. European Geriatric Medicine. 2020///; 11(#issue#): S9. doi: http://dx.doi.org/10.1007/s41999-020-00428-6. Available at: http://ovidsp.ovid.com/ovidweb.cgi?T=JS&PAGE=reference&D=emexb&NEWS=N&AN=634448438 |
| 204 | Palanques Pastor, T, Megias Vericat, J, Poveda Andres, J L, Montestinos, P, Martinez, P, Cornago Navascues, J, Lopez Lorenzo, J L, Rodriguez, G, Cano, I, Arnan Sangerman, M. Impact of SARS-CoV-2 infection in acute myeloid leukaemia patients: Experience of the pethema registry. European Journal of Hospital Pharmacy. 2021///; 28(#issue#): A58. doi: http://dx.doi.org/10.1136/ejhpharm-2021-eahpconf.119. Available at: http://ovidsp.ovid.com/ovidweb.cgi?T=JS&PAGE=reference&D=emedx&NEWS=N&AN=634720280 |
| 205 | Del Brutto, Oscar H, Wu, Shasha, Issa, Naoum P, Mera, Robertino M, Costa, Aldo F, Recalde, Bettsy Y. Cognitive decline among individuals with history of mild symptomatic SARS-CoV-2 infection: A longitudinal prospective study nested to a population cohort. European Journal of Neurology. 2021///; #volume#(#issue#): #pages#. doi: http://dx.doi.org/10.1111/ene.14775. Available at: http://onlinelibrary.wiley.com/journal/10.1111/(ISSN)1468-1331,http://ovidsp.ovid.com/ovidweb.cgi?T=JS&PAGE=reference&D=emexb&NEWS=N&AN=2010611122 |
| 206 | Nanavati, Ruchi, Mascarenhas, Dwayne, Goyal, Medha, Haribalakrishna, Anitha, Nataraj, Gita. A single-center observational study on clinical features and outcomes of 21 SARS-CoV-2-infected neonates from India. European journal of pediatrics. 2021///; #volume#(#issue#): #pages#. doi: http://dx.doi.org/10.1007/s00431-021-03967-7. Available at: https://rd.springer.com/journal/431,http://ovidsp.ovid.com/ovidweb.cgi?T=JS&PAGE=reference&D=emexb&NEWS=N&AN=2010389555 |
| 207 | Jiang, Mingshan, Mu, Jingxi, Shen, Silan, Zhang, Hu. COVID-19 With Preexisting Hypercoagulability Digestive Disease. Frontiers in Medicine. 2020///; 7(#issue#): 587350. doi: http://dx.doi.org/10.3389/fmed.2020.587350. Available at: http://journal.frontiersin.org/journal/medicine,http://ovidsp.ovid.com/ovidweb.cgi?T=JS&PAGE=reference&D=emexb&NEWS=N&AN=633990670 |
| 208 | Saburi, Ehsan, Abazari, Mohammad Foad, Hassannia, Hadi, Mansour, Reyhaneh Nassiri, Eshaghi-Gorji, Reza, Gheibi, Mobina, Rahmati, Mohammad, Enderami, Seyed Ehsan. The use of mesenchymal stem cells in the process of treatment and tissue regeneration after recovery in patients with Covid-19. Gene. 2021///; 777(#issue#): 145471. doi: http://dx.doi.org/10.1016/j.gene.2021.145471. Available at: http://www.elsevier.com/locate/gene,http://ovidsp.ovid.com/ovidweb.cgi?T=JS&PAGE=reference&D=emexb&NEWS=N&AN=2010974421 |
| 209 | Gomez-Mesa, Juan, Galindo-Coral, Stephania, Montes, Maria Claudia, Alarco, Walter, Barisani, Jose Luis, Magana, Antonio, Perna, Eduardo R, Romero, Alexander, Speranza, Mario, Mendoza, Ivan, Wyss, |

| | |
|---|---|
| | Fernando. Latin-American Registry of Cardiovascular Disease and COVID-19: Rationale and Design of the CARDIO COVID 19-20 Registry. Global heart. 2021///; 16(1): 14. doi: https://dx.doi.org/10.5334/gh.925. Available at: http://ovidsp.ovid.com/ovidweb.cgi?T=JS&PAGE=reference&D=prem&NEWS=N&AN=33833938 |
| 210 | Savastano, Maria Cristina, Gambini, Gloria, Cozzupoli, Grazia Maria, Crincoli, Emanuele, Savastano, Alfonso, De Vico, Umberto, Culiersi, Carola, Falsini, Benedetto, Minnella, Angelo Maria, Martelli, Francesco, Landi, Francesco, Pagano, Francesco Cosimo, Rizzo, Stanislao. Retinal capillary involvement in early post-COVID-19 patients: a healthy controlled study. Graefe's archive for clinical and experimental ophthalmology = Albrecht von Graefes Archiv fur klinische und experimentelle Ophthalmologie. 2021///; #volume#(#issue#): #pages#. doi: http://dx.doi.org/10.1007/s00417-020-05070-3. Available at: http://ovidsp.ovid.com/ovidweb.cgi?T=JS&PAGE=reference&D=emexb&NEWS=N&AN=634157489 |
| 211 | NCT04801940. HElping Alleviate the Longer-term Consequences of COVID-19 (HEAL-COVID). HElping Alleviate the Longer-term Consequences of COVID-19 (HEAL-COVID): a National Platform Trial. 2021///; #volume#(#issue#): #pages#. doi: #DOI#. Available at: http://ovidsp.ovid.com/ovidweb.cgi?T=JS&PAGE=reference&D=cctr&NEWS=N&AN=CN-02249995 |
| 212 | Abdel-Hamid, Hoda, Magdy, Sally, Rizk, Hoda Ibrahim. Occurrence of pulmonary residuals as one of the sequelae of COVID-19 and it's predictors among moderate and severe cases. Indian Journal of Tuberculosis. 2021///; #volume#(#issue#): #pages#. doi: http://dx.doi.org/10.1016/j.ijtb.2021.01.006. Available at: http://tbassnindia.org/Archives.html,http://ovidsp.ovid.com/ovidweb.cgi?T=JS&PAGE=reference&D=emexb&NEWS=N&AN=2011027169 |
| 213 | Garritsen, Anja, Scholzen, Anja, van den Nieuwenhof, Daan W A, Smits, Anke P F, Datema, E S, van Galen, Luc S, Kouwijzer, Milou L C E. Two-tiered SARS-CoV-2 seroconversion screening in the Netherlands and stability of nucleocapsid, spike protein domain 1 and neutralizing antibodies. Infectious Diseases. 2021///; #volume#(#issue#): #pages#. doi: http://dx.doi.org/10.1080/23744235.2021.1893378. Available at: http://www.tandfonline.com/loi/infd20#.VrgcOLdf1Fo,http://ovidsp.ovid.com/ovidweb.cgi?T=JS&PAGE=reference&D=emexb&NEWS=N&AN=2010718156 |
| 214 | Zarrougui, W, Bejaoui, N, Sedrati, F, Elbeki, N, Lambiotte, F. Decline in functional status after ICU discharge of COVID-19 patients. Intensive Care Medicine Experimental. 2020///; 8(#issue#): #pages#. doi: http://dx.doi.org/10.1186/s40635-020-00354-8. Available at: http://ovidsp.ovid.com/ovidweb.cgi?T=JS&PAGE=reference&D=emexb&NEWS=N&AN=634008301 |
| 215 | Haghighi Aski, Behzad, Manafi Anari, Ali, Abolhasan Choobdar, Farhad, Zareh Mahmoudabadi, Ramin, Sakhaie, Maryam. Cardiac abnormalities due to multisystem inflammatory syndrome temporally associated with Covid-19 among children: A systematic review and meta-analysis. International journal of cardiology.Heart & vasculature. 2021///; #volume#(#issue#): 100764. doi: https://dx.doi.org/10.1016/j.ijcha.2021.100764. Available at: http://ovidsp.ovid.com/ovidweb.cgi?T=JS&PAGE=reference&D=medp&NEWS=N&AN=33778151 |
| 216 | Gharebaghi, Naser, Nejadrahim, Rahim, Mousavi, Jalil, Farshid, Saman, Boroofeh, Behdad, Dindarian, Sina, Mohammadi, Sedra. Evaluation of epidemiology, clinical features, prognosis, diagnosis and treatment outcomes of patients with COVID-19 in West Azerbaijan Province. International journal of clinical practice. 2021///; #volume#(#issue#): #pages#. doi: http://dx.doi.org/10.1111/ijcp.14108. Available at: http://onlinelibrary.wiley.com/journal/10.1111/(ISSN)1742-1241,http://ovidsp.ovid.com/ovidweb.cgi?T=JS&PAGE=reference&D=emexb&NEWS=N&AN=2010875691 |
| 217 | Mancini, Luca, Sacrini, Francesco, Narcisi, Alessandra, Valenti, Mario, Pavia, Giulia, Gargiulo, Luigi, Facheris, Paola, Borroni, Riccardo G, Costanzo, Antonio, Nucca, Ombretta. Impact of delay in follow-up due to COVID-19 pandemic on skin cancer progression: a real-life experience from an Italian hub hospital. International journal of dermatology. 2021///; #volume#(#issue#): #pages#. doi: http://dx.doi.org/10.1111/ijd.15501. Available at: http://onlinelibrary.wiley.com/journal/10.1111/(ISSN)1365-4632,http://ovidsp.ovid.com/ovidweb.cgi?T=JS&PAGE=reference&D=emexb&NEWS=N&AN=2010693363 |

| | |
|---|---|
| 218 | Ferdous, Most Zannatul, Islam, Ummay Soumayia, Islam, Md Saiful, Mosaddek, Abu Syed Md, Potenza, Marc N, Pardhan, Shahina. Treatment, persistent symptoms, and depression in people infected with covid-19 in bangladesh. International Journal of Environmental Research and Public Health. 2021///; 18(4): 1. doi: http://dx.doi.org/10.3390/ijerph18041453. Available at: https://www.mdpi.com/1660-4601/18/4/1453/pdf,http://ovidsp.ovid.com/ovidweb.cgi?T=JS&PAGE=reference&D=emexb&NEWS=N&AN=2005957992 |
| 219 | Kocak, Orhan, Kocak, Omer Erdem, Younis, Mustafa Z. The psychological consequences of COVID-19 fear and the moderator effects of individuals' underlying illness and witnessing infected friends and family. International Journal of Environmental Research and Public Health. 2021///; 18(4): 1. doi: http://dx.doi.org/10.3390/ijerph18041836. Available at: https://www.mdpi.com/1660-4601/18/4/1836/pdf,http://ovidsp.ovid.com/ovidweb.cgi?T=JS&PAGE=reference&D=emexb&NEWS=N&AN=2006018548 |
| 220 | Schoendorf, Sabine, Spielmanns, Marc, Pekacka-Egli, Anna, Windisch, Wolfram, Hermann, Matthias. Effects of a comprehensive pulmonary rehabilitation in severe post-covid-19 patients. International Journal of Environmental Research and Public Health. 2021///; 18(5): 1. doi: http://dx.doi.org/10.3390/ijerph18052695. Available at: https://www.mdpi.com/1660-4601/18/5/2581/pdf,http://ovidsp.ovid.com/ovidweb.cgi?T=JS&PAGE=reference&D=emexb&NEWS=N&AN=2006148326 |
| 221 | Tomar, Balvir S, Singh, Mahaveer, Nathiya, Deepak, Sharma, Abhishek, Sharma, Eshan, Bareth, Hemant, Suman, Supriya, Ruparelia, Darshan Purshotambhai, Patel, Jaykumar Bharatbhai, Gajera, Vikrant Kantilal, NIMS COVID 19, Investigator Group. Prevalence of Symptoms in Patients Discharged from COVID Care Facility of NIMS Hospital: Is RT PCR Negativity Truly Reflecting Recovery? A Single-Centre Observational Study. International journal of general medicine. 2021///; 14(#issue#): 1069. doi: https://dx.doi.org/10.2147/IJGM.S295499. Available at: http://ovidsp.ovid.com/ovidweb.cgi?T=JS&PAGE=reference&D=pmnm&NEWS=N&AN=33790636 |
| 222 | Zhang, Jintao, Xu, Jiawei, Ning, Kang, Pan, Yun, Dong, Liang, Zhou, Shengyu, Liu, Tian, Zhao, Jiping, Wang, Chunting, Wang, Ximing, Zhang, Wei. The characteristics of 527 discharged COVID-19 patients undergoing long-term follow-up in China. International Journal of Infectious Diseases. 2021///; 104(#issue#): 685. doi: http://dx.doi.org/10.1016/j.ijid.2021.01.064. Available at: https://www.journals.elsevier.com/international-journal-of-infectious-diseases,http://ovidsp.ovid.com/ovidweb.cgi?T=JS&PAGE=reference&D=emexb&NEWS=N&AN=2011091979 |
| 223 | An, Ya-Wen, Yuan, Bo, Wang, Jian-Chun, Wang, Cheng, Liu, Ting-Ting, Song, Shuo, Liu, Han-Qing. Clinical characteristics and impacts of traditional Chinese medicine treatment on the convalescents of COVID-19. International journal of medical sciences. 2021///; 18(3): 646. doi: https://dx.doi.org/10.7150/ijms.52664. Available at: http://ovidsp.ovid.com/ovidweb.cgi?T=JS&PAGE=reference&D=medl&NEWS=N&AN=33437199 |
| 224 | Haqberdiev, Jasur, Tadjiev, Umid, Salokhiddinov, Marufjon, Kurbanova, Bernara, Magzumov, Nodir, Ashirov, Bekzod. Radiological characteristics of lung-related complications in covid patients. International Journal of Pharmaceutical Research. 2021///; 13(2): 828. doi: http://dx.doi.org/10.31838/ijpr/2021.13.02.134. Available at: http://www.ijpronline.com/ViewArticleDetail.aspx?ID=20632,http://ovidsp.ovid.com/ovidweb.cgi?T=JS&PAGE=reference&D=emexb&NEWS=N&AN=2005985693 |
| 225 | Martinez MW Bloom OJ, Green G, DiFiori JP, Solomon G, Phelan D, Kim JH, Meeuwisse W, Sills AK, Rowe D, Bogoch II, Smith PT, Baggish AL, Putukian M, Engel,D.J., Tucker A M. Prevalence of Inflammatory Heart Disease Among Professional Athletes With Prior COVID-19 Infection Who Received Systematic Return-to-Play Cardiac Screening. JAMA cardiology. 2021///; #volume#(#issue#): #pages#. doi: 10.1001/jamacardio.2021.0565. Available at: https://pubmed.ncbi.nlm.nih.gov/33662103 |
| 226 | Udelson, James E, Rowin, Ethan J, Curtis, Michael A. Return to play for athletes after coronavirus disease 2019 infection-Making high-stakes recommendations as data evolve. JAMA Cardiology. 2021///; 6(2): 136. doi: http://dx.doi.org/10.1001/jamacardio.2020.5896. Available at: |

| | |
|---|---|
| | http://jamanetwork.com/journals/jamacardiology,http://ovidsp.ovid.com/ovidweb.cgi?T=JS&PAGE=reference&D=emexb&NEWS=N&AN=633333695 |
| 227 | Barocas, Joshua A, Gai, Mam Jarra, White, Laura F, Faretra, Deanna, Sachs, Kerry, Komaromy, Miriam. Implementation of a Recuperation Unit and Hospitalization Rates Among People Experiencing Homelessness With COVID-19. JAMA network open. 2021///; 4(3): e212826. doi: https://dx.doi.org/10.1001/jamanetworkopen.2021.2826. Available at: http://ovidsp.ovid.com/ovidweb.cgi?T=JS&PAGE=reference&D=medl&NEWS=N&AN=33688963 |
| 228 | Mahil, Satveer K, Tsakok, Teresa, Meynell, Freya, Moorhead, Lucy, Smith, Catherine H, Capon, Francesca, Dand, Nick, Mason, Kayleigh J, Yiu, Zenas Z N, Warren, Richard B, Griffiths, Christopher E M, Coker, Bola, McAteer, Helen, Mackenzie, Teena, Rossi, Maria Teresa, Rivera, Raquel, Mahe, Emmanuel, Carugno, Andrea, Magnano, Michela, Rech, Giulia, Balogh, Esther A, Feldman, Steven R, De La Cruz, Claudia, Choon, Siew Eng, Naldi, Luigi, Lambert, Jo, Spuls, Phyllis, Jullien, Denis, Bachelez, Herve, McMahon, Devon E, Freeman, Esther E, Gisondi, Paolo, Puig, Luis, Di Meglio, Paola, Barker, Jonathan N, Langan, Sinead M, Shah, Aadarsh, Barea, Alberto, Romero-Mate, Alberto, Singapore, Alekya, Paolino, Alexandra, Mwale, Alice, Morales Callaghan, Ana Maria, Martinez, Ana, DeCrescenzo, Andrew, Pink, Andrew E, Jones, Ann, Sergeant, Ann, Essex, Annette, Bewley, Anthony, Makrygeorgou, Areti, van Huizen, Astrid, Perez-Suarez, Beatriz, Farida, Benhadou, Clareus, Birgitta Wilson, Prims, Carla Tubau, Davis, Carrie, Quinlan, Catherine, Maybury, Catriona, Cesar, Gonzalez A, Barclay, Charlotte, Greco, Claudio, Brassard, Danielle, Cummings, Deanna, Kolli, Deepti, Descamps, Vincent, Genao, Diana Ruiz, Carras, Efrossini, Hawryluk, Elena, Martinez-Garcia, Eliseo, Klujszo, Elzbieta, Dwyer, Emily, Toni, Emmanuel, Sonkoly, Eniko, Loayza, Enrique, Dauden, Esteban, Valenzuela, Fernando, Popov, Georgi, King, Georgie, Celine, Girard, Aparicio, Gloria, Johnston, Graham A, Cardozo, Gustavo Anibal, Pearson, Ian, Yanguas, Ignacio, Weisman, Jamie, Carolan, Jennifer E, Hughes, Jenny, Ortiz-Salvador, Jose, Carrascosa, Jose-Manuel, Schwartz, Joseph J, Jackson, Karina, Kerisit, Kathryn G, Wu, Keith, Asfour, Leila, de Graaf, Leontien, Lesort, Cecile, Meuleman, Lieve, Eidsmo, Liv, Skov, Lone, Gribben, Lorraine, Rustin, Malcolm, Velasco, Manel, Panchal, Manisha, Lakhan, Manpreet, Franco, Manuel D, Svensson, Marie-Louise, Vandaele, Mark, Marovt, Maruska, Zargari, Omid, De Caso, Pablo, Varela, Paulo, Jenkin, Peter, Phan, Celine, Hampton, Philip, Goldsmith, Portia, Bak, Rachel, Speeckaert, Reinhart, Romiti, Ricardo, Woolf, Richard, Mercado-Seda, Rogelio, Khatun, Rohima, Ceovic, Romana, Taberner, Rosa, Cohen, Russell W, Stefanescu, Simina, Kirk, Sarah, Reeken, Saskia, Ayob, Shanti, Perez-Barrio, Silvia, Piaserico, Stefano, Hoey, Susannah, Torres, Tiago, Talme, Toomas, Desai, Trupti V, van Geest, Adrienne J, King, Victoria, Di Lernia, Vito, Koreja, Zahira, Hasab, Vito Zeeshaan. Factors associated with adverse COVID-19 outcomes in patients with psoriasis-insights from a global registry-based study. Journal of Allergy and Clinical Immunology. 2021///; 147(1): 60. doi: http://dx.doi.org/10.1016/j.jaci.2020.10.007. Available at: http://www.elsevier.com/inca/publications/store/6/2/3/3/6/8/index.htt,http://ovidsp.ovid.com/ovidweb.cgi?T=JS&PAGE=reference&D=emexb&NEWS=N&AN=2010066119 |
| 229 | Guan, Wei-jie, Zhong, Nan-shan, Liang, Wen-hua, He, Jian-xing, Shi, Ying, Gan, Lan-xia, Wang, Hai-bo. Chronic Respiratory Diseases and the Outcomes of COVID-19: A Nationwide Retrospective Cohort Study of 39,420 Cases. Journal of Allergy and Clinical Immunology: In Practice. 2021///; #volume#(#issue#): #pages#. doi: http://dx.doi.org/10.1016/j.jaip.2021.02.041. Available at: http://www.elsevier.com/journals/the-journal-of-allergy-and-clinical-immunology-in-practice/2213-2198,http://ovidsp.ovid.com/ovidweb.cgi?T=JS&PAGE=reference&D=emexb&NEWS=N&AN=2011437657 |
| 230 | UlHaq, Zia, Shahzad, Muhammad, Khattak, Maria Ishaq, Fazid, Sheraz, Ullah, Naeem, Shireen, Akhtar, Ulhaq, Naeem, Izhar, Asif, Farooq, Umer, Darwesh, Nizam Muhammad, Asim, Muhammad, Gohar, MianGul Ali, Ashraf, Amber, Ashraf, Saadia, Khattak, Saad Ishaq, Akbar, Shahzad, Amir, Azizul Hasan, Fawad, Muhammad, Amanullah, Amir, Khan, Muhammad Ismail, Siddiq, Muhammad. Clinical Characteristics, Mortality and Associated risk factors in COVID-19 patients reported in ten major hospitals of Khyber Pakhtunkhwa, Pakistan. Journal of Ayub Medical College, Abbottabad : JAMC. 2020///; 32 1)(4): S633. doi: #DOI#. Available at: http://ovidsp.ovid.com/ovidweb.cgi?T=JS&PAGE=reference&D=emexb&NEWS=N&AN=634638749 |

| | |
|---|---|
| 231 | Cormican, Daniel S, Winter, Daniel, McHugh, Stephen, Sonny, Abraham, Crowley, Jerome, Yu, Raymond, Barrack, Frank, Nunez-Gil, Ivan, Ramakrishna, Harish. Severe Acute Respiratory Syndrome Coronavirus-2 Cardiovascular Complications: Implications for Cardiothoracic Anesthesiology. Journal of cardiothoracic and vascular anesthesia. 2021///; 35(3): 932. doi: https://dx.doi.org/10.1053/j.jvca.2020.05.035. Available at: http://ovidsp.ovid.com/ovidweb.cgi?T=JS&PAGE=reference&D=medl&NEWS=N&AN=32601001 |
| 232 | McKemey, Emily, Baranskaya, Aliaksandra, Gompertz, Simon, Dosanjh, Davinder, Madathil, Shyam, Shields, Adrian M, Faustini, Sian E, Richter, Alex G, Hill, Harriet J, Stamataki, Zania. Erratum: Correction to: Resolution of Persistent COVID-19 After Convalescent Plasma in a Patient with B Cell Aplasia (Journal of clinical immunology (2021)). Journal of clinical immunology. 2021///; #volume#(#issue#): #pages#. doi: http://dx.doi.org/10.1007/s10875-021-01029-z. Available at: http://ovidsp.ovid.com/ovidweb.cgi?T=JS&PAGE=reference&D=emedx&NEWS=N&AN=634694004 |
| 233 | Roseboom, Tessa J, Ozanne, Susan E, Godfrey, Keith M, Barker, Mary, Fall, Caroline H D, Isasi, Carmen R, Itoh, Hiroaki, Simmons, Rebecca, Bansal, Amita, Plosch, Torsten, Sloboda, Deb M, Matthews, Stephen G, Poston, Lucilla, Hanson, Mark A. Unheard, unseen and unprotected: DOHaD council's call for action to protect the younger generation from the long-term effects of COVID-19. Journal of developmental origins of health and disease. 2021///; 12(1): 3. doi: http://dx.doi.org/10.1017/S2040174420000847. Available at: http://ovidsp.ovid.com/ovidweb.cgi?T=JS&PAGE=reference&D=emexb&NEWS=N&AN=632955366 |
| 234 | Hughes, Thomas, Pietropaolo, Amelia, Archer, Matthew, Davis, Tania, Tear, Loretta, Somani, Bhaskar K. Lessons Learnt (Clinical Outcomes and Cost Savings) from Virtual Stone Clinic and Their Application in the Era Post-COVID-19: Prospective Outcomes over a 6-Year Period from a University Teaching Hospital. Journal of endourology. 2021///; 35(2): 200. doi: https://dx.doi.org/10.1089/end.2020.0708. Available at: http://ovidsp.ovid.com/ovidweb.cgi?T=JS&PAGE=reference&D=medl&NEWS=N&AN=32731751 |
| 235 | Aslam, M I, Minhas, A, Ghorbani, A, Jani, V, Hsu, S, Sharma, K, Hays, A, Gilotra, N A, Shade, J, Cihakova, D. Natriuretic Peptide Levels and Clinical Outcomes among Patients Hospitalized with COVID-19 Infection. Journal of Heart and Lung Transplantation. 2021///; 40(4): S210. doi: http://dx.doi.org/10.1016/j.healun.2021.01.606. Available at: http://ovidsp.ovid.com/ovidweb.cgi?T=JS&PAGE=reference&D=emexb&NEWS=N&AN=2011430585 |
| 236 | Pakhchanian, H, Patel, V A, Raiker, R. 342 Clinical outcomes in COVID-19 patients with Atopic dermatitis. Journal of Investigative Dermatology. 2021///; 141(5): S60. doi: http://dx.doi.org/10.1016/j.jid.2021.02.364. Available at: http://ovidsp.ovid.com/ovidweb.cgi?T=JS&PAGE=reference&D=emedx&NEWS=N&AN=2011607989 |
| 237 | Pakhchanian, H, Patel, V A, Raiker, R. 254 COVID-19 related outcomes in psoriasis and psoriasis arthritis patients. Journal of Investigative Dermatology. 2021///; 141(5): S45. doi: http://dx.doi.org/10.1016/j.jid.2021.02.276. Available at: http://ovidsp.ovid.com/ovidweb.cgi?T=JS&PAGE=reference&D=emedx&NEWS=N&AN=2011607891 |
| 238 | Ali, Kameran Mohammed, Ali, Ayad M, Tawfeeq, Hassan Mohammad, Figueredo, Grazziela, Rostam, Hassan Muhammad. Hypoalbuminemia in Patients Following Their Recovery from Severe Coronavirus Disease 2019. Journal of medical virology. 2021///; #volume#(#issue#): #pages#. doi: https://dx.doi.org/10.1002/jmv.27002. Available at: http://ovidsp.ovid.com/ovidweb.cgi?T=JS&PAGE=reference&D=medp&NEWS=N&AN=33830538 |
| 239 | Decourcey, Danielle D. A Breather during the Time of Breathlessness. Journal of palliative medicine. 2021///; 24(2): 289. doi: http://dx.doi.org/10.1089/jpm.2020.0410. Available at: http://www.liebertonline.com/jpm,http://ovidsp.ovid.com/ovidweb.cgi?T=JS&PAGE=reference&D=emexb&NEWS=N&AN=634101165 |
| 240 | Huguet, Nathalie, Angier, Heather, Marino, Miguel, DeVoe, Jennifer, Schmidt, Teresa, Larson, Annie, O'Malley, Jean, Hoopes, Megan. Prevalence of pre-existing conditions among community health center patients with COVID-19: Implications for the patient protection and affordable care act. Journal of the American Board of Family Medicine. 2021///; 34(#issue#): S247. doi: http://dx.doi.org/10.3122/JABFM.2021.S1.200571. Available at: http://www.jabfm.org/lookup/doi/10.3122/jabfm.2021.S1.200571,http://ovidsp.ovid.com/ovidweb.cgi?T=JS&PAGE=reference&D=emexb&NEWS=N&AN=2011504216 |

| | |
|---|---|
| 241 | Greco, Giada Ida, Trevisan, Caterina, Zatti, Giancarlo, Pozza, Martino Dalla, Lazzarin, Martina, Haxhiaj, Labjona, Sergi, Giuseppe, Noale, Marianna, Maggi, Stefania, Ramon, Roberto, Imoscopi, Alessandra, Bellon, Stefano. Increase in Frailty in Nursing Home Survivors of Coronavirus Disease 2019: Comparison With Noninfected Residents. Journal of the American Medical Directors Association. 2021///; #volume#(#issue#): #pages#. doi: http://dx.doi.org/10.1016/j.jamda.2021.02.019. Available at: http://ovidsp.ovid.com/ovidweb.cgi?T=JS&PAGE=reference&D=emexb&NEWS=N&AN=634649908 |
| 242 | Pant, Pankaj, Bam, Niraj, Das, Santa Kumar, Joshi, Aishana, Basnet, Babin, Shrestha, Bibek Man, Bista, Navindra Raj. Prevalence of functional limitation in COVID-19 recovered patients using the post COVID-19 functional status scale. Journal of the Nepal Medical Association. 2021///; 59(233): 7. doi: http://dx.doi.org/10.31729/jnma.5980. Available at: https://www.jnma.com.np/jnma/index.php/jnma/article/view/5980/3354,http://ovidsp.ovid.com/ovidweb.cgi?T=JS&PAGE=reference&D=emexb&NEWS=N&AN=2005968058 |
| 243 | Garcia-Azorin, David, Aguirre, Maria Elena Erro, Monco, Juan Carlos Garcia, Guijarro-Castro, Cristina, Delgado, Fernando Romero, Andres, Jose Miguel Lainez, Ezpeleta, David, Fernandez, Alejandra Collia, Horga, Alejandro, Etessam, Jesus Porta, Davila, Lucia Galan, Gonzalez, Nuria, Prieto, Ana Barragan, del Villar Igea, Ana, Vilar Ventura, Rosa M, Nunez, Ana Urbanos, Serrano, Angel Aledo, Garcia, Araceli, Menendez, Beatriz Martinez, Gomez, Jose Balseiro, Recasens, Bernat Bertran, Serrano, Blanca Serrano, de la Esperanza, Blanca Talavera, de la Hoz Fuenmayor Fernandez, Carlos Pablo de, Perez Martinez, David A, Juste, Carlos Tejero, Iniguez, Cristina, Bueno, Jose Carlos Roche, Atienza, Saida, Martin, Carmen Valderrama, Lopez, Jose Maria Barrios, Garcia, Cristina Fernandez, Llamas, Esteban Pena, Ochoa, Marta, Garcia, Daniel Macias, Raquel Lamas, Leire Ainz, Rodriguez, Manuel Medina, Santacruz, Debora M Cerdan, Caballero, Lorena, Robert, Misericordia Floriach, Gomez, Elisabet Franquet, Puiggros, Elsa, Alvarez, Eric Freire, Villareal, Estibaliz, Diaz, Eva Fernandez, Burguillos, Fernando Morejon, Beitia, Garazi Agirre, Abildua, Maria Jose Abenza, Gutierrez-Gutierrez, Gerardo, Monteiro, Guilherme Carvalho, Ygual, Guillermo Cervera, Hernandez, Guillermo, Rubio, Guillermo, Alonso, Hortenia, Froiz, Iago Payo, Garcia, Iago Rego, Penas, Inmaculada Redondo, Perez Sanchez, Javier R, Mas, Miguel, Garcia, Javier Tejada, Alvarez, Javier Villacieros, Dominguez, Jessica Fernandez, Equiza, Jon, Zabaleta, Maite Martinez, Pascual, Jorge Millan, Usieto, Jose Antonio Olivan, Reyes, Jose Antonio, Velasquez, Jose Luis Camacho, Lezama, Lidia Binela Lara, de Antonio Alberto Rodriguez, Luis, Torres, M A, Moragues, Maria Dolores, Garcia, Maria Fuensanta Valero, Salas, Teresa Mateos, Platas, Montserrat Gonzalez, Hernandez, Maria, Rabasa, Maria, Santos, Maria Rico, Usero, Maria, Zurdo, Martin, Maneiro, Miren, Muriana, Nafria, Noelia Gonzalez, Esteve, Oriol Barrachina, del Saz Saucedo, Pablo, Rocio, Navas, Sandra Rodriguez, Fernandez, Santiago Fernandez, Borja, Sergio, Navarro, Silvia Gil, Garcia, Solange Kapetanovic, Garcia, Susana, Palasi, Toni, Cantero, Victoriano Romero. Neurological presentations of COVID-19: Findings from the Spanish Society of Neurology neuroCOVID-19 registry. Journal of the neurological sciences. 2021///; 423(#issue#): 117283. doi: http://dx.doi.org/10.1016/j.jns.2020.117283. Available at: http://www.elsevier.com/locate/jns,http://ovidsp.ovid.com/ovidweb.cgi?T=JS&PAGE=reference&D=emexb&NEWS=N&AN=2011111702 |
| 244 | Koh, Jasmine Shimin, De Silva, Deidre Anne, Quek, Amy May Lin, Chiew, Hui Jin, Tu, Tian Ming, Seet, Christopher Ying Hao, Hoe, Rebecca Hui Min, Saini, Monica, Hui, Andrew Che-Fai, Angon, Jasmyn, Ker, Justin Ruixin, Yong, Ming Hui, Goh, Yihui, Yu, Wai-Yung, Lim, Tchoyoson Choie Cheio, Tan, Benjamin Yong Qiang, Ng, Kay Wei Ping, Yeo, Leonard Leong Litt, Pang, Yu Zhi, Prakash, Kumar M, Ahmad, Aftab, Thomas, Terrence, Lye, David Chien Boon, Tan, Kevin, Umapathi, Thirugnanam. Neurology of COVID-19 in Singapore. Journal of the neurological sciences. 2020///; 418(#issue#): #pages#. doi: http://dx.doi.org/10.1016/j.jns.2020.117118; http://dx.doi.org/10.1016/j.jns.2020.117118. Available at: http://ovidsp.ovid.com/ovidweb.cgi?T=JS&PAGE=reference&D=psyc17&NEWS=N&AN=2020-82179-001 |
| 245 | Xu, Jiaxuan, Liang, Zhenyu, Jian, Wenhua, Tang, Guoyan, Zhang, Dongying, Zheng, Jinping, Li, Shiyue, Li, Jianyu, Qian, Yuanxin, Mo, Xiaoneng, Liu, Jinxin. Changes of quantitative CT-based airway wall dimensions in patients with COVID-19 during early recovery. Journal of Thoracic Disease. 2021///; 13(3): 1517. doi: http://dx.doi.org/10.21037/jtd-20-2790. Available at: |

| | |
|---|---|
| | https://jtd.amegroups.com/article/view/49370/pdf,http://ovidsp.ovid.com/ovidweb.cgi?T=JS&PAGE=reference&D=emexb&NEWS=N&AN=2011548609 |
| 246 | Gaspardone, Carlo, Preda, Alberto, Romagnolo, Davide, Meloni, Carlo, Maranta, Francesco, Cianflone, Domenico, Brugliera, Luigia, Castellazzi, Paola, Tettamanti, Andrea, Iannaccone, Sandro, Conte, Caterina, Secchi, Antonio. Lung Ultrasound in COVID-19 A Role Beyond the Acute Phase?. Journal of ultrasound in medicine : official journal of the American Institute of Ultrasound in Medicine. 2021///; 40(3): 503. doi: http://dx.doi.org/10.1002/jum.15425. Available at: http://ovidsp.ovid.com/ovidweb.cgi?T=JS&PAGE=reference&D=emexb&NEWS=N&AN=632561355 |
| 247 | Cantarella, Giovanna, Pignataro, Lorenzo, Alde, Mirko, Berardino, Federica Di, Barozzi, Stefania, Zanetti, Diego, Consonni, Dario, Zuccotti, Gianvincenzo, Bertoli, Simona, Battezzati, Alberto. Prevalence of Dysphonia in Non hospitalized Patients with COVID-19 in Lombardy, the Italian Epicenter of the Pandemic. Journal of Voice. 2021///; #volume#(#issue#): #pages#. doi: http://dx.doi.org/10.1016/j.jvoice.2021.03.009. Available at: http://www.elsevier.com/wps/find/journaldescription.cws_home/672710/description#description,http://ovidsp.ovid.com/ovidweb.cgi?T=JS&PAGE=reference&D=emexb&NEWS=N&AN=2011471750 |
| 248 | Huang, Shujie, Zhuang, Weitao, Wang, Dongya, Zha, Lulu, Xu, Xi, Li, Xiangdong, Shi, Qiuling, Wang, Xin Shelley, Qiao, Guibin. Persistent somatic symptom burden and sleep disturbance in patients with COVID-19 during hospitalization and after discharge: A prospective cohort study. Medical Science Monitor. 2021///; 27(#issue#): e930447. doi: http://dx.doi.org/10.12659/MSM.930447. Available at: https://www.medscimonit.com/abstract/index/idArt/930447,http://ovidsp.ovid.com/ovidweb.cgi?T=JS&PAGE=reference&D=emexb&NEWS=N&AN=2011271002 |
| 249 | Omezli, M-M, Torul, D. Evaluation of the xerostomia, taste and smell impairments after Covid-19. Medicina oral, patologia oral y cirugia bucal. 2021///; #volume#(#issue#): #pages#. doi: http://dx.doi.org/10.4317/medoral.24510. Available at: http://ovidsp.ovid.com/ovidweb.cgi?T=JS&PAGE=reference&D=emexb&NEWS=N&AN=634677277 |
| 250 | Capdevila-Reniu A Prieto-Gonzalez S, Ventosa H, Ladino A, Naval J, Rodriguez-Nunez O, Cesar Milisenda J, Moreno-Lozano PJ, Soriano A, Bosch X, Lopez-Soto A, Hospital Clinic COVID-19,Research Group, Pellice M. Clinical characteristics and outcome of patients aged over 80 years with covid-19; 7909132 CSR - Reports results. Medicine. 2021///; 100(8): e24750. doi: 10.1097/MD.0000000000024750. Available at: https://pubmed.ncbi.nlm.nih.gov/33663089 |
| 251 | KGC Mescia F, Turner L, Hanson A, Kotagiri P, Dunmore BJ, Ruffieux H, De Sa A, Huhn O, Wills MR, Baker S, Doffinger R, Dougan G, Elmer A, Goodfellow IG, Gupta RK, Hosmillo M, Hunter K, Kingston N, Lehner PJ, Matheson NJ, Nicholson JK, Petrunkina AM, Richardson S, Saunders C, Thaventhiran JED, Toonen EJM, Weekes MP, Toshner M, Hess C, Bradley JR, Lyons PA, Smith, Bergamaschi L. Early immune pathology and persistent dysregulation characterise severe COVID-19 (preprint). Medrxiv. 2021///; #volume#(#issue#): 2021.01.11.20248765. doi: 10.1101/2021.01.11.20248765. Available at: http://medrxiv.org/content/early/2021/01/15/2021.01.11.20248765.abstract |
| 252 | Etemadifar, Masoud, Aghababaee, Ali, Rayani, Milad, Sedaghat, Nahad, Abhari, Amir Parsa, Khorvash, Reza, Nouri, Hosein, Kargaran, Parisa K, Maracy, Mohammad Reza, Ganjalikhani-Hakemi, Mazdak, Salari, Mehri. COVID-19 and the Risk of Relapse in Multiple Sclerosis Patients: A Fight with No Bystander Effect?. Multiple Sclerosis and Related Disorders. 2021///; 51(#issue#): 102915. doi: http://dx.doi.org/10.1016/j.msard.2021.102915. Available at: http://www.elsevier.com/wps/find/journaldescription.cws_home/725776/description#description,http://ovidsp.ovid.com/ovidweb.cgi?T=JS&PAGE=reference&D=emedx&NEWS=N&AN=2011539096 |
| 253 | Subbaraman, Nidhi. US health agency will invest billion to investigate 'long COVID'. Nature. 2021///; 591(7850): 356. doi: http://dx.doi.org/10.1038/d41586-021-00586-y. Available at: http://ovidsp.ovid.com/ovidweb.cgi?T=JS&PAGE=reference&D=emexb&NEWS=N&AN=634516178 |
| 254 | Neeland, Melanie R, Dohle, Kate, Mulholland, Kim, Sutton, Philip, Saffery, Richard, Bannister, Samantha, Curtis, Nigel, Steer, Andrew C, Burgner, David P, Clifford, Vanessa, Crawford, Nigel W, Tosif, Shidan. Innate cell profiles during the acute and convalescent phase of SARS-CoV-2 infection in children. Nature Communications. 2021///; 12(1): 1084. doi: http://dx.doi.org/10.1038/s41467-021-21414-x. Available at: |

| | |
|---|---|
| | http://www.nature.com/ncomms/index.html,http://ovidsp.ovid.com/ovidweb.cgi?T=JS&PAGE=reference&D=emexb&NEWS=N&AN=2010517841 |
| 255 | NCT04397510. Nebulized Heparin for the Treatment of COVID-19 Induced Lung Injury. Nebulized Heparin vs.Placebo for the Treatment of COVID-19 Induced Lung Injury. 2020///; #volume#(#issue#): #pages#. doi: #DOI#. Available at: http://ovidsp.ovid.com/ovidweb.cgi?T=JS&PAGE=reference&D=cctr&NEWS=N&AN=CN-02124825 |
| 256 | Liu, Fuyou, Chen, Wei, Sun, Lin. Covid-19 and kidney injury: Caution on novel coronavirus associated pulmonary-renal syndrome. Nephrology. 2020///; 25(#issue#): 10. doi: http://dx.doi.org/10.1111/nep.13771. Available at: http://ovidsp.ovid.com/ovidweb.cgi?T=JS&PAGE=reference&D=emexb&NEWS=N&AN=633928342 |
| 257 | Ermis, Ummehan, Rust, Marcus Immanuel, Bungenberg, Julia, Costa, Ana, Tauber, Simone C, Dreher, Michael, Balfanz, Paul, Marx, Gernot, Wiesmann, Martin, Reetz, Kathrin, Schulz, Jorg B. Neurological symptoms in COVID-19: a cross-sectional monocentric study of hospitalized patients. Neurological Research and Practice. 2021///; 3(1): 17. doi: http://dx.doi.org/10.1186/s42466-021-00116-1. Available at: https://neurolrespract.biomedcentral.com/,http://ovidsp.ovid.com/ovidweb.cgi?T=JS&PAGE=reference&D=emexb&NEWS=N&AN=2010771657 |
| 258 | Pasquini, Jacopo, Silani, Vincenzo, Ticozzi, Nicola, Maremmani, Carlo, Salvadori, Stefano. Testing olfactory dysfunction in acute and recovered COVID-19 patients: a single center study in Italy. Neurological Sciences. 2021///; #volume#(#issue#): #pages#. doi: http://dx.doi.org/10.1007/s10072-021-05200-7. Available at: https://link.springer.com/journal/10072,http://ovidsp.ovid.com/ovidweb.cgi?T=JS&PAGE=reference&D=emexb&NEWS=N&AN=2010936428 |
| 259 | Anand, Pria, Zhou, Lan, Bhadelia, Nahid, Hamer, Davidson H, Greer, David M, Cervantes-Arslanian, Anna. Neurologic Findings Among Inpatients With COVID-19 at a Safety-net US Hospital. Neurology.Clinical practice. 2021///; 11(2): e83. doi: https://dx.doi.org/10.1212/CPJ.0000000000001031. Available at: http://ovidsp.ovid.com/ovidweb.cgi?T=JS&PAGE=reference&D=pmnm&NEWS=N&AN=33842075 |
| 260 | Ferrario, Silvia Rossi, Cerutti, Paola, Sacco, Daniela, Panzeri, Anna. The psychological experience and intervention in post-acute COVID-19 inpatients. Neuropsychiatric Disease and Treatment. 2021///; 17(#issue#): 413. doi: http://dx.doi.org/10.2147/NDT.S283558. Available at: https://www.dovepress.com/getfile.php?fileID=66459,http://ovidsp.ovid.com/ovidweb.cgi?T=JS&PAGE=reference&D=emexb&NEWS=N&AN=2006044191 |
| 261 | Jaywant, Abhishek, Vanderlind, W M, Alexopoulos, George S, Fridman, Chaya B, Perlis, Roy H, Gunning, Faith M. Frequency and profile of objective cognitive deficits in hospitalized patients recovering from covid-19. Neuropsychopharmacology. 2021///; #volume#(#issue#): No. doi: http://dx.doi.org/10.1038/s41386-021-00978-8; http://dx.doi.org/10.1038/s41386-021-00978-8. Available at: http://ovidsp.ovid.com/ovidweb.cgi?T=JS&PAGE=reference&D=psyc17&NEWS=N&AN=2021-30194-001 |
| 262 | Vollbracht, Claudia, Kraft, Karin. Feasibility of vitamin c in the treatment of post viral fatigue with focus on long covid, based on a systematic review of iv vitamin c on fatigue. Nutrients. 2021///; 13(4): 1154. doi: http://dx.doi.org/10.3390/nu13041154. Available at: https://www.mdpi.com/2072-6643/13/4/1154/pdf,http://ovidsp.ovid.com/ovidweb.cgi?T=JS&PAGE=reference&D=emexb&NEWS=N&AN=2006866543 |
| 263 | Abrishami, Mojtaba, Daneshvar, Ramin, Emamverdian, Zahra, Tohidinezhad, Fariba, Eslami, Saeid. Optic Nerve Head Parameters and Peripapillary Retinal Nerve Fiber Layer Thickness in Patients with Coronavirus Disease 2019. Ocular immunology and inflammation. 2021///; #volume#(#issue#): #pages#. doi: http://dx.doi.org/10.1080/09273948.2020.1850800. Available at: http://www.tandfonline.com/loi/ioii20,http://ovidsp.ovid.com/ovidweb.cgi?T=JS&PAGE=reference&D=emexb&NEWS=N&AN=2010533799 |
| 264 | C Wheen P, Armstrong R, Kumar R, McMahon A, White M, Sheehy N, McMahon G, Murphy RT, Daly, Cronin M. CT coronary angiography and COVID-19: inpatient use in acute chest pain service; 7976674 CSR |

| | |
|---|---|
| | - Reports results. Open heart. 2021///; 8(1): #pages#. doi: 10.1136/openhrt-2020-001548. Available at: https://pubmed.ncbi.nlm.nih.gov/33731419 |
| 265 | Fuggle NR Gill C, Patel A, Medeiros A, Mlotek AS, Pierroz DD, Halbout P, Harvey NC, Reginster JY, Cooper C, Greenspan,S.L., Singer A. Correction to: how has COVID-19 affected the treatment of osteoporosis? An IOF-NOF-ESCEO global survey; 7942214 CSR - Reports results. Osteoporosis International. 2021///; #volume#(#issue#): #pages#. doi: 10.1007/s00198-021-05905-7. Available at: https://pubmed.ncbi.nlm.nih.gov/33751152 |
| 266 | NCT04782700. P-Co-Li (Pulmonary Covid-19 Study). P-CO-Li (Pulmonary Covid-19 Longterm Intervention Study). 2021///; #volume#(#issue#): #pages#. doi: #DOI#. Available at: http://ovidsp.ovid.com/ovidweb.cgi?T=JS&PAGE=reference&D=cctr&NEWS=N&AN=CN-02249562 |
| 267 | Yilmaz, Kamil, Gozupirinccioglu, Ayfer, Aktar, Fesih, Akin, Alper, Karabel, Musemma, Sen, Velat, Yolbas, Ilyas, Uzel, Veysiye Hulya. Evaluation of the novel coronavirus disease in Turkish children: Preliminary outcomes. Pediatric pulmonology. 2020///; 55(12): 3587. doi: http://dx.doi.org/10.1002/ppul.25095. Available at: http://onlinelibrary.wiley.com/journal/10.1002/(ISSN)1099-0496,http://ovidsp.ovid.com/ovidweb.cgi?T=JS&PAGE=reference&D=emexb&NEWS=N&AN=2006889799 |
| 268 | Johnson, Joshua K, Lapin, Brittany, Green, Karen, Stilphen, Mary. Frequency of Physical Therapist Intervention Is Associated With Mobility Status and Disposition at Hospital Discharge for Patients With COVID-19. Physical Therapy. 2021///; 101(1): #pages#. doi: http://dx.doi.org/10.1093/ptj/pzaa181. Available at: http://ovidsp.ovid.com/ovidweb.cgi?T=JS&PAGE=reference&D=emexb&NEWS=N&AN=633036245 |
| 269 | Mahmud, Reaz, Rahman, Md Mujibur, Rassel, Mohammad Aftab, Monayem, Farhana Binte, Sayeed, S K J, Islam, Md Shahidul, Islam, Mohammed Monirul. Post-COVID-19 syndrome among symptomatic COVID-19 patients: A prospective cohort study in a tertiary care center of Bangladesh. PloS one. 2021///; 16(4): e0249644. doi: https://dx.doi.org/10.1371/journal.pone.0249644. Available at: http://ovidsp.ovid.com/ovidweb.cgi?T=JS&PAGE=reference&D=prem&NEWS=N&AN=33831043 |
| 270 | Marion, Elodie, Gardes, Sophie, Gerbier-Colomban, Solweig, Kuczewski, Elisabetta, Vanhems, Philippe, Gustin, Marie-Paule, Elias, Christelle, Henaff, Laetitia, Dananche, Cedric, Grisi, Beatrice, Khanafer, Nagham, Hilliquin, Delphine, Amour, Selilah, Saadatian-Elahi, Mitra, Escuret, Vanessa, Lina, Bruno, Argaud, Laurent, Aubrun, Frederic, Bonnefoy, Marc, Bouscambert-Duchamp, Maude, Chapurlat, Roland, Chassard, Dominique, Chidiac, Christian, Chuzeville, Michel, Confavreux, Cyrille, Couraud, Sebastien, Devouassoux, Gilles, Durieu, Isabelle, Fessy, Michel, Gaujard, Sylvain, Gaymard, Alexandre, Hot, Arnaud, Gaujard, Geraldine Martin, Morelon, Emmanuel, Piriou, Vincent, Potinet, Veronique, Richard, Jean-Christophe, Rimmele, Thomas, Seve, Pascal, Sigal, Alain, Tazarourte, Karim. Factors associated with admission to intensive care units in COVID-19 patients in Lyon-France. PLoS ONE. 2021///; 16(1): e0243709. doi: http://dx.doi.org/10.1371/journal.pone.0243709. Available at: https://journals.plos.org/plosone/article/file?id=10.1371/journal.pone.0243709&type=printable,http://ovidsp.ovid.com/ovidweb.cgi?T=JS&PAGE=reference&D=emexb&NEWS=N&AN=2010910528 |
| 271 | Almyroudi, Maria Panagiota, Dimopoulos, George. Incidence, pathogenesis and management of thromboembolic complications in severe COVID-19. Pneumon. 2020///; 33(4): 1. doi: #DOI#. Available at: http://www.pneumon.org/assets/files/789/file826_782.pdf,http://ovidsp.ovid.com/ovidweb.cgi?T=JS&PAGE=reference&D=emexb&NEWS=N&AN=2006128138 |
| 272 | Jowett, Sally, Shevlin, Mark, Hyland, Philip, Karatzias, Thanos. Posttraumatic Stress Disorder and Persistent Somatic Symptoms During the COVID-19 Epidemic: The Role of Sense of Threat. Psychosomatic medicine. 2020///; #volume#(#issue#): #pages#. doi: https://dx.doi.org/10.1097/PSY.0000000000000890. Available at: http://ovidsp.ovid.com/ovidweb.cgi?T=JS&PAGE=reference&D=medp&NEWS=N&AN=33818055 |
| 273 | Bekesi, Dora, Eory, Ajandek, Eory, Ajandok, Rozsa, Sandor. Physical Exercise as a Resilience Factor to Mitigate COVID-Related Allostatic Overload. Psychotherapy and psychosomatics. 2021///; #volume#(#issue#): #pages#. doi: http://dx.doi.org/10.1159/000514331. Available at: http://www.karger.com/journals/pps/pps_jh.htm,http://ovidsp.ovid.com/ovidweb.cgi?T=JS&PAGE=reference&D=emexb&NEWS=N&AN=634493827 |

| | |
|---|---|
| 274 | D Migliori GB, Centis R, Cherubino F, Facchetti C, Feci D, Palmiotto G, Pignatti P, Saderi L, Sotgiu G, Spanevello A, Zappa M, Visca, Zampogna E. Functional impairment during post-acute COVID-19 phase: preliminary finding in 56 patients. Pulmonology. 2021///; #volume#(#issue#): #pages#. doi: 10.1016/j.pulmoe.2020.12.008. Available at: https://pubmed.ncbi.nlm.nih.gov/33454281 CSR - Reports results |
| 275 | ISRCTN11466448. A randomized trial to assess the effects of physical and mental health rehabilitation for people recovering from COVID-19. Rehabilitation Exercise and psycholoGical support After covid-19 InfectioN (REGAIN): a multi-centre randomized controlled trial. 2020///; #volume#(#issue#): #pages#. doi: #DOI#. Available at: http://ovidsp.ovid.com/ovidweb.cgi?T=JS&PAGE=reference&D=cctr&NEWS=N&AN=CN-02239337 |
| 276 | Stals, Mam, Grootenboers, Mjjh, van Guldener, C, Kaptein, Fhj, Braken, Sje, Chen, Q, Chu, G, van Driel E.M., Iglesias Del Sol, A, de Jonge, E, Kant, K M, Pals, F, Toorop, Mma, Cannegieter, S C, Klok, F A, Huisman, M V, Dutch COVID, Thrombosis Coalition (DCTC). Risk of thrombotic complications in influenza versus COVID-19 hospitalized patients. Research and practice in thrombosis and haemostasis. 2021///; #volume#(#issue#): #pages#. doi: https://dx.doi.org/10.1002/rth2.12496. Available at: http://ovidsp.ovid.com/ovidweb.cgi?T=JS&PAGE=reference&D=medp&NEWS=N&AN=33821230 |
| 277 | Laporte, Jean-Charles, Bellocq, Agnes. Therapeutic education for people with chronic respiratory disease. Revue de l'infirmiere. 2021///; 70(269): 20. doi: http://dx.doi.org/10.1016/j.revinf.2021.01.007. Available at: http://ovidsp.ovid.com/ovidweb.cgi?T=JS&PAGE=reference&D=emexb&NEWS=N&AN=634623387 |
| 278 | Schenker, Hannah M, Hagen, Melanie, Simon, David, Schett, Georg, Manger, Bernhard. Reactive arthritis and cutaneous vasculitis after SARS-CoV-2 infection. Rheumatology (United Kingdom). 2021///; 60(1): 479. doi: http://dx.doi.org/10.1093/rheumatology/keaa689. Available at: http://rheumatology.oxfordjournals.org/,http://ovidsp.ovid.com/ovidweb.cgi?T=JS&PAGE=reference&D=emexb&NEWS=N&AN=2010881459 |
| 279 | Ramani, Santhoshini Leela, Samet, Jonathan, Deshmukh, Swati, Franz, Colin K, Hsieh, Christine, Nguyen, Cuong V, Horbinski, Craig. Musculoskeletal involvement of COVID-19: review of imaging. Skeletal radiology. 2021///; #volume#(#issue#): #pages#. doi: http://dx.doi.org/10.1007/s00256-021-03734-7. Available at: http://link.springer.de/link/service/journals/00256/index.htm,http://ovidsp.ovid.com/ovidweb.cgi?T=JS&PAGE=reference&D=emexb&NEWS=N&AN=2010521334 |
| 280 | Xu, Fei, Hu, Xiaowen, Wang, Xixin, Zhang, Kai, Shi, Yudong, Xia, Lei, Liu, Huanzhong, Yang, Yanguo. Depression and insomnia in COVID-19 survivors: a cross-sectional survey from Chinese rehabilitation centers in Anhui province. Sleep medicine. 2021///; #volume#(#issue#): #pages#. doi: http://dx.doi.org/10.1016/j.sleep.2021.02.002. Available at: http://www.elsevier.com/inca/publications/store/6/2/0/2/8/2,http://ovidsp.ovid.com/ovidweb.cgi?T=JS&PAGE=reference&D=emexb&NEWS=N&AN=2011110430 |
| 281 | Zhou, Yuan, Cen, Lusha. COVID-19 and Perioperative Management Strategies for Gastrointestinal Surgery: An Experience From Jiaxing, China. The American Surgeon. 2021///; #volume#(#issue#): 3134821995087. doi: http://dx.doi.org/10.1177/0003134821995087. Available at: http://ovidsp.ovid.com/ovidweb.cgi?T=JS&PAGE=reference&D=emexb&NEWS=N&AN=634356584 |
| 282 | Wise, Jacqui. Long covid: WHO calls on countries to offer patients more rehabilitation. The BMJ. 2021///; 372(#issue#): n405. doi: http://dx.doi.org/10.1136/bmj.n405. Available at: http://www.bmj.com/,http://ovidsp.ovid.com/ovidweb.cgi?T=JS&PAGE=reference&D=emexb&NEWS=N&AN=634162352 |
| 283 | Libby, Peter, Siddiqi, Hasan K, Ridker, Paul M. COVID-19 - A vascular disease. Trends in cardiovascular medicine. 2021///; 31(1): 1. doi: http://dx.doi.org/10.1016/j.tcm.2020.10.005. Available at: http://www.elsevier.com/locate/tcm,http://ovidsp.ovid.com/ovidweb.cgi?T=JS&PAGE=reference&D=emexb&NEWS=N&AN=2008354554 |
| 284 | Xiaofang, Gao, Chunxia, Ma, Qian, Zhao, Yujie, Ma, Xiaoqi, Wang, Wei, Zhou, Xiwei, Zheng, Xiangyuan, Cao, Jianhua, Wei, Guixia, Zhao, Lin, Xu, Tao, Feng. Clinical efficacy and safety of different antiviral regimens in patients with coronavirus disease 2019. Zhonghua Wei Zhong Bing Ji Jiu Yi Xue. 2021///; |

| | |
|---|---|
| | 32(12): 1423. doi: http://dx.doi.org/10.3760/cma.j.cn121430-20201019-00679. Available at: http://www.cccm-em120.com/weizhongbingguokan/32/1423.pdf,http://ovidsp.ovid.com/ovidweb.cgi?T=JS&PAGE=reference&D=emexb&NEWS=N&AN=2010972962 |

**Non-primary research or type of article not of interest = 176**

| | |
|---|---|
| 1 | Greenhalgh T., Knight M.. Long COVID: A Primer for Family Physicians. American Family Physician. 2020; 102(12): 716. doi: #DOI#. Available at: http://ovidsp.ovid.com/ovidweb.cgi?T=JS&PAGE=reference&D=emedx&NEWS=N&AN=633815375 |
| 2 | Tavassoly O., Safavi F., Tavassoly I.. Seeding Brain Protein Aggregation by SARS-CoV-2 as a Possible Long-Term Complication of COVID-19 Infection. ACS Chemical Neuroscience. 2020; 11(22): 3704. doi: http://dx.doi.org/10.1021/acschemneuro.0c00676. Available at: http://pubs.acs.org/journal/acncdm; http://ovidsp.ovid.com/ovidweb.cgi?T=JS&PAGE=reference&D=emedx&NEWS=N&AN=2010118914 |
| 3 | Bernardi E., Plazzi F., Ferravante A., Valentini I., Mussoni A., Mascetti S., Agli L.A.L.. Late Breaking Abstract - Exercise rehabilitation in telemedicine for COVID-19 patients. European Respiratory Journal. 2020; 56(#issue#): #pages#. doi: http://dx.doi.org/10.1183/13993003.congress-2020.882. Available at: https://erj.ersjournals.com/content/56/suppl_64/882; http://ovidsp.ovid.com/ovidweb.cgi?T=JS&PAGE=reference&D=emedx&NEWS=N&AN=633805144 |
| 4 | Yelin D., Margalit I., Yahav D., Runold M., Bruchfeld J.. Long COVID-19 - it's not over until?. Clinical microbiology and infection : the official publication of the European Society of Clinical Microbiology and Infectious Diseases. 2020; #volume#(#issue#): #pages#. doi: http://dx.doi.org/10.1016/j.cmi.2020.12.001. Available at: http://ovidsp.ovid.com/ovidweb.cgi?T=JS&PAGE=reference&D=emedx&NEWS=N&AN=633807286 |
| 5 | Hassett C.E., Gedansky A., Migdady I., Bhimraj A., Uchino K., Cho S.-M.. Neurologic complications of COVID-19. Cleveland Clinic journal of medicine. 2020; 87(12): 729. doi: http://dx.doi.org/10.3949/ccjm.87a.ccc058. Available at: http://ovidsp.ovid.com/ovidweb.cgi?T=JS&PAGE=reference&D=emexc&NEWS=N&AN=632710845 |
| 6 | Nath A.. Long-Haul COVID. Neurology. 2020; 95(13): 559. doi: http://dx.doi.org/10.1212/WNL.0000000000010640. Available at: http://ovidsp.ovid.com/ovidweb.cgi?T=JS&PAGE=reference&D=emexc&NEWS=N&AN=632603954 |
| 7 | Mangion K., Morrow A., Bagot C., Bayes H., Blyth K.G., Church C., Corcoran D., Delles C., Gillespie L., Grieve D., Ho A., Kean S., Lang N.N., Lennie V., Lowe D.J., Kellman P., Macfarlane P.W., McConnachie A., Roditi G., Sykes R., Touyz R.M., Sattar N., Wereski R., Wright S., Berry C.. The Chief Scientist Office Cardiovascular and Pulmonary Imaging in SARS Coronavirus disease-19 (CISCO-19) study. Cardiovascular research. 2020; 116(14): 2185. doi: http://dx.doi.org/10.1093/cvr/cvaa209. Available at: http://ovidsp.ovid.com/ovidweb.cgi?T=JS&PAGE=reference&D=emexc&NEWS=N&AN=632437024 |
| 8 | Ptacek R., Ptackova H., Martin A., Stefano G.B.. Psychiatric manifestations of COVID-19 and their social significance. Medical Science Monitor. 2020; 26(#issue#): e930340. doi: http://dx.doi.org/10.12659/MSM.930340. Available at: https://www.medscimonit.com/abstract/index/idArt/930340; http://ovidsp.ovid.com/ovidweb.cgi?T=JS&PAGE=reference&D=emexc&NEWS=N&AN=2010426547 |
| 9 | Gorna R., MacDermott N., Rayner C., O'Hara M., Evans S., Agyen L., Nutland W., Rogers N., Hastie C.. Long COVID guidelines need to reflect lived experience. The Lancet. 2020; #volume#(#issue#): #pages#. doi: http://dx.doi.org/10.1016/S0140-6736%2820%2932705-7. Available at: http://www.journals.elsevier.com/the-lancet/; http://ovidsp.ovid.com/ovidweb.cgi?T=JS&PAGE=reference&D=emexc&NEWS=N&AN=2010448906 |
| 10 | Davido B., Seang S., Tubiana R., de Truchis P.. Post-COVID-19 chronic symptoms: a postinfectious entity?. Clinical Microbiology and Infection. 2020; 26(11): 1448. doi: http://dx.doi.org/10.1016/j.cmi.2020.07.028. Available at: https://www.journals.elsevier.com/clinical-microbiology-and-infection; http://ovidsp.ovid.com/ovidweb.cgi?T=JS&PAGE=reference&D=emexc&NEWS=N&AN=2007467198 |
| 11 | Colombo A., Pavan L., Baiardini R., Cerutti S., Conte L., Evangelista I., Rotiroti G., Capra R., Porta C., Olivetti M., Migliorisi C., Mazzone A., Mumoli N.. An outpatient follow-up for post-COVID-19 patients. Italian Journal of Medicine. 2020; 14(#issue#): 109. doi: #DOI#. Available at: |

| | |
|---|---|
| | https://www.italjmed.org/index.php/ijm/article/view/itjm.2020.s2/1309; http://ovidsp.ovid.com/ovidweb.cgi?T=JS&PAGE=reference&D=emexc&NEWS=N&AN=633718057 |
| 12 | Karlsson A.C., Humbert M., Buggert M.. The known unknowns of T cell immunity to COVID-19. Science Immunology. 2020; 5(53): abe8063. doi: http://dx.doi.org/10.1126/SCIIMMUNOL.ABE8063. Available at: https://immunology.sciencemag.org/lookup/doi/10.1126/sciimmunol.abe8063; http://ovidsp.ovid.com/ovidweb.cgi?T=JS&PAGE=reference&D=emexc&NEWS=N&AN=2010391368 |
| 13 | Shurovi B.. P.789 Neurological manifestations of intensive care patients with COVID-19 in a South-London Hospital: a single centre observational study. European Neuropsychopharmacology. 2020; 40(#issue#): S447. doi: http://dx.doi.org/10.1016/j.euroneuro.2020.09.582. Available at: http://ovidsp.ovid.com/ovidweb.cgi?T=JS&PAGE=reference&D=emexc&NEWS=N&AN=2008991377 |
| 14 | Siegelman J.N.. Reflections of a COVID-19 Long Hauler. JAMA - Journal of the American Medical Association. 2020; 324(20): 2031. doi: http://dx.doi.org/10.1001/jama.2020.22130. Available at: http://jama.jamanetwork.com/journal.aspx; http://ovidsp.ovid.com/ovidweb.cgi?T=JS&PAGE=reference&D=emexc&NEWS=N&AN=633410003 |
| 15 | Ye L., Sun P., Wang T.. Acupuncture strategies to tackle post covid-19 psychological and neuropsychiatric disorders. Journal of Chinese Medicine. 2020; 2020(124): 11. doi: #DOI#. Available at: http://www.jcm.co.uk/contact-us/; http://ovidsp.ovid.com/ovidweb.cgi?T=JS&PAGE=reference&D=emexc&NEWS=N&AN=2005285429 |
| 16 | Salehi S., Reddy S., Gholamrezanezhad A.. Long-term Pulmonary Consequences of Coronavirus Disease 2019 (COVID-19): What We Know and What to Expect. Journal of thoracic imaging. 2020; 35(4): W87. doi: http://dx.doi.org/10.1097/RTI.0000000000000534. Available at: http://journals.lww.com/thoracicimaging; http://ovidsp.ovid.com/ovidweb.cgi?T=JS&PAGE=reference&D=emexc&NEWS=N&AN=633266282 |
| 17 | Anwar M.M., Badawi A.M., Eltablawy N.A.. Can the coronavirus infection penetrates the brain resulting in sudden anosmia followed by severe neurological disorders?. eNeurologicalSci. 2020; 21(#issue#): 100290. doi: http://dx.doi.org/10.1016/j.ensci.2020.100290. Available at: http://www.journals.elsevier.com/eneurologicalsci/; http://ovidsp.ovid.com/ovidweb.cgi?T=JS&PAGE=reference&D=emexc&NEWS=N&AN=2010055251 |
| 18 | Levy J.M.. Treatment Recommendations for Persistent Smell and Taste Dysfunction following COVID-19 - The Coming Deluge. JAMA Otolaryngology - Head and Neck Surgery. 2020; 146(8): 733. doi: http://dx.doi.org/10.1001/jamaoto.2020.1378. Available at: http://archotol.jamanetwork.com/issues.aspx; http://ovidsp.ovid.com/ovidweb.cgi?T=JS&PAGE=reference&D=emexc&NEWS=N&AN=632409695 |
| 19 | Pilloni G., Bikson M., Badran B.W., George M.S., Kautz S.A., Okano A.H., Baptista A.F., Charvet L.E.. Update on the Use of Transcranial Electrical Brain Stimulation to Manage Acute and Chronic COVID-19 Symptoms. Frontiers in Human Neuroscience. 2020; 14(#issue#): 595567. doi: http://dx.doi.org/10.3389/fnhum.2020.595567. Available at: https://www.frontiersin.org/journals/human-neuroscience; http://ovidsp.ovid.com/ovidweb.cgi?T=JS&PAGE=reference&D=emexc&NEWS=N&AN=633491496 |
| 20 | Kazama I.. Stabilizing mast cells by commonly used drugs: A novel therapeutic target to relieve post-COVID syndrome?. Drug Discoveries and Therapeutics. 2020; 14(5): 259. doi: http://dx.doi.org/10.5582/DDT.2020.03095. Available at: https://www.jstage.jst.go.jp/article/ddt/14/5/14_2020.03095/_article; http://ovidsp.ovid.com/ovidweb.cgi?T=JS&PAGE=reference&D=emexc&NEWS=N&AN=2010106726 |
| 21 | Beghi E., Helbok R., Crean M., Chou S.H.-Y., McNett M., Moro E., Bassetti C.. The EAN COVID-19 registry (ENERGY): An international instrument for surveillance of neurological complications in patients with COVID-19. European journal of neurology. 2020; #volume#(#issue#): #pages#. doi: http://dx.doi.org/10.1111/ene.14652. Available at: http://ovidsp.ovid.com/ovidweb.cgi?T=JS&PAGE=reference&D=emexc&NEWS=N&AN=633489350 |
| 22 | Murphy P.. Individuals With Less Severe Manifestations of SARS-CoV-2 Infection May Not Develop Long-Lasting Humoral Immunity. American Journal of Clinical Pathology. 2020; #volume#(#issue#): #pages#. |

| | |
|---|---|
| | doi: http://dx.doi.org/10.1093/ajcp/aqaa233. Available at: http://ovidsp.ovid.com/ovidweb.cgi?T=JS&PAGE=reference&D=emexc&NEWS=N&AN=633488469 |
| 23 | Kayani B., Onochie E., Patil V., Begum F., Cuthbert R., Ferguson D., Bhamra J.S., Sharma A., Bates P., Haddad F.S.. Infographic: The effects of COVID-19 on perioperative morbidity and mortality in patients with hip fractures. Bone and Joint Journal. 2020; 102(10): 1279. doi: http://dx.doi.org/10.1302/0301-620X.102B10.BJJ-2020-1774. Available at: https://online.boneandjoint.org.uk/doi/10.1302/0301-620X.102B10.BJJ-2020-1774; http://ovidsp.ovid.com/ovidweb.cgi?T=JS&PAGE=reference&D=emexc&NEWS=N&AN=2008329971 |
| 24 | Rubin R.. As Their Numbers Grow, COVID-19 "Long Haulers" Stump Experts. JAMA - Journal of the American Medical Association. 2020; 324(14): 1381. doi: http://dx.doi.org/10.1001/jama.2020.17709. Available at: http://jama.jamanetwork.com/journal.aspx; http://ovidsp.ovid.com/ovidweb.cgi?T=JS&PAGE=reference&D=emexc&NEWS=N&AN=633080971 |
| 25 | Boscolo-Rizzo P., Polesel J., Spinato G., Menegaldo A., Fabbris C., Calvanese L., Borsetto D., Hopkins C.. Predominance of an altered sense of smell or taste among long-lasting symptoms in patients with mildly symptomatic COVID-19. Rhinology. 2020; 58(5): 524. doi: http://dx.doi.org/10.4193/Rhin20.263. Available at: http://ovidsp.ovid.com/ovidweb.cgi?T=JS&PAGE=reference&D=emexc&NEWS=N&AN=632398434 |
| 26 | Honigsbaum M., Krishnan L.. Taking pandemic sequelae seriously: from the Russian influenza to COVID-19 long-haulers. The Lancet. 2020; 396(10260): 1389. doi: http://dx.doi.org/10.1016/S0140-6736%2820%2932134-6. Available at: http://www.journals.elsevier.com/the-lancet/; http://ovidsp.ovid.com/ovidweb.cgi?T=JS&PAGE=reference&D=emexc&NEWS=N&AN=2008414197 |
| 27 | Ashton J.. Long COVID - What doesn't kill you may not make you stronger. Journal of the Royal Society of Medicine. 2020; 113(11): 466. doi: http://dx.doi.org/10.1177/0141076820971225. Available at: http://www.uk.sagepub.com/journals/Journal202191; http://ovidsp.ovid.com/ovidweb.cgi?T=JS&PAGE=reference&D=emexc&NEWS=N&AN=2007244559 |
| 28 | Warner J.L., Rubinstein S., Grivas P., Choueiri T.K., Kuderer N.M., Shah D., Rivera D.R., Gupta S., Bilen M.A., Halfdanarson T.R., Doroshow D.B., Wehbe F., Shah S., Shyr Y., Lopes G., Painter C., Lyman G.H., Thompson M.A., Peters S., Rini B.I.. Clinical impact of COVID-19 on patients with cancer: Data from the COVID-19 and Cancer Consortium (CCC19). Journal of Clinical Oncology. 2020; 38(18): #pages#. doi: http://dx.doi.org/10.1200/JCO.2020.38.18_suppl.LBA110. Available at: https://ascopubs.org/doi/abs/10.1200/JCO.2020.38.18_suppl.LBA110; http://ovidsp.ovid.com/ovidweb.cgi?T=JS&PAGE=reference&D=emexc&NEWS=N&AN=633377617 |
| 29 | Ma K., Wang X., Feng S., Xia X., Zhang H., Rahaman A., Dong Z., Lu Y., Li X., Zhou X., Zhao H., Wang Y., Wang S., Baloch Z.. From the perspective of Traditional Chinese Medicine: Treatment of mental disorders in COVID-19 survivors. Biomedicine and Pharmacotherapy. 2020; 132(#issue#): 110810. doi: http://dx.doi.org/10.1016/j.biopha.2020.110810. Available at: https://www.journals.elsevier.com/biomedicine-and-pharmacotherapy; http://ovidsp.ovid.com/ovidweb.cgi?T=JS&PAGE=reference&D=emexc&NEWS=N&AN=2008031667 |
| 30 | Klitzman R.L.. Needs to Prepare for "Post-COVID-19 Syndrome". The American journal of bioethics : AJOB. 2020; 20(11): 4. doi: http://dx.doi.org/10.1080/15265161.2020.1820755. Available at: http://ovidsp.ovid.com/ovidweb.cgi?T=JS&PAGE=reference&D=emexc&NEWS=N&AN=633270472 |
| 31 | Paglionico A., Varriano V., Tolusso B., Alivernini S., Petricca L., Natalello G., Gigante L., Bosello S.L., Martone A.M., Landi F., Gremese E.. Loss of self-tolerance in SARS-COV-2 infection: Immunological assessment of a convalescent cohort. Annals of the Rheumatic Diseases. 2020; 79(#issue#): 209. doi: http://dx.doi.org/10.1136/annrheumdis-2020-eular.6862. Available at: http://ovidsp.ovid.com/ovidweb.cgi?T=JS&PAGE=reference&D=emexc&NEWS=N&AN=633304621 |
| 32 | Baig A.M.. Chronic COVID Syndrome: Need for an appropriate medical terminology for Long-COVID and COVID Long-Haulers. Journal of medical virology. 2020; #volume#(#issue#): #pages#. doi: http://dx.doi.org/10.1002/jmv.26624. Available at: http://ovidsp.ovid.com/ovidweb.cgi?T=JS&PAGE=reference&D=emexc&NEWS=N&AN=633258451 |

| | |
|---|---|
| 33 | Jiang D.H., McCoy R.G.. Planning for the Post-COVID Syndrome: How Payers Can Mitigate Long-Term Complications of the Pandemic. Journal of General Internal Medicine. 2020; 35(10): 3036. doi: http://dx.doi.org/10.1007/s11606-020-06042-3. Available at: http://www.springerlink.com/content/120414/; http://ovidsp.ovid.com/ovidweb.cgi?T=JS&PAGE=reference&D=emexc&NEWS=N&AN=2005654215 |
| 34 | Rosovsky R., Li A., Desai A., Shah D., Fu J., Gulati S., Zon R., Thompson M., Grivas P., De Lima Lopes G., Shyr Y., Warner J., Khaki A., Choueiri T., Peters S., Rivera D., Rini B., Lyman G., Connors J., Kuderer N.. Venous thrombotic complications in cancer patients with SARS-CoV- 2 Infection: Report from the COVID-19 and Cancer Consortium (CCC19) Registry Analysis. Research and Practice in Thrombosis and Haemostasis. 2020; 4(#issue#): 10. doi: http://dx.doi.org/10.1002/rth2.12413. Available at: http://ovidsp.ovid.com/ovidweb.cgi?T=JS&PAGE=reference&D=emexc&NEWS=N&AN=633611310 |
| 35 | Stals M.A.M., Grootenboers M.J.J.H., van Guldener C., Kaptein F.H.J., Braken S.J.E., Chen Q., Chu G., Van Driel E.M., Iglesias Del Sol A., Pals F., Toorop M.M.A., Cannegieter S.C., Klok F.A., Huisman M.V.. Higher incidence of thrombotic complications in hospitalized patients with SARS-COV- 2 virus versus influenza virus infections. Research and Practice in Thrombosis and Haemostasis. 2020; 4(#issue#): 3. doi: http://dx.doi.org/10.1002/rth2.12413. Available at: http://ovidsp.ovid.com/ovidweb.cgi?T=JS&PAGE=reference&D=emexc&NEWS=N&AN=633611231 |
| 36 | Borg, Kristian, Stam, Henk J.. Rehabilitation of post-Covid - 19 syndrome - once again a call for action!. Journal of Rehabilitation Medicine. 2021; 53(1): jrm00132. doi: https://dx.doi.org/10.2340/16501977-2783. Available at: http://ovidsp.ovid.com/ovidweb.cgi?T=JS&PAGE=reference&D=prem&NEWS=N&AN=33385238 |
| 37 | Malek, Lukasz A.. Cardiac Involvement After Recovering From COVID-19. JAMA cardiology. 2020; #volume#(#issue#): #pages#. doi: https://dx.doi.org/10.1001/jamacardio.2020.5276. Available at: http://ovidsp.ovid.com/ovidweb.cgi?T=JS&PAGE=reference&D=medp&NEWS=N&AN=33112368 |
| 38 | Rolfe, Robert J., Smith, Colin M., Wolfe, Cameron R.. The Emerging Chronic Sequelae of COVID-19 and Implications for North Carolina. North Carolina medical journal. 2021; 82(1): 75. doi: https://dx.doi.org/10.18043/ncm.82.1.75. Available at: http://ovidsp.ovid.com/ovidweb.cgi?T=JS&PAGE=reference&D=prem&NEWS=N&AN=33397764 |
| 39 | Soriano, Vicente, Ganado-Pinilla, Pilar, Sanchez-Santos, Miguel, Barreiro, Pablo. Unveiling Long COVID-19 Disease. AIDS reviews. 2020; 22(4): 227. doi: https://dx.doi.org/10.24875/AIDSRev.M20000039. Available at: http://ovidsp.ovid.com/ovidweb.cgi?T=JS&PAGE=reference&D=medp&NEWS=N&AN=33401287 |
| 40 | Tan, Eng Hooi, Sena, Anthony G., Prats-Uribe, Albert, You, Seng Chan, Ahmed, Waheed-Ul-Rahman, Kostka, Kristin, Reich, Christian, Duvall, Scott L., Lynch, Kristine E., Matheny, Michael E., Duarte-Salles, Talita, Bertolin, Sergio Fernandez, Hripcsak, George, Natarajan, Karthik, Falconer, Thomas, Spotnitz, Matthew, Ostropolets, Anna, Blacketer, Clair, Alshammari, Thamir M., Alghoul, Heba, Alser, Osaid, Lane, Jennifer C. E., Dawoud, Dalia M., Shah, Karishma, Yang, Yue, Zhang, Lin, Areia, Carlos, Golozar, Asieh, Relcade, Martina, Casajust, Paula, Jonnagaddala, Jitendra, Subbian, Vignesh, Vizcaya, David, Lai, Lana Yh, Nyberg, Fredrik, Morales, Daniel R., Posada, Jose D., Shah, Nigam H., Gong, Mengchun, Vivekanantham, Arani, Abend, Aaron, Minty, Evan P., Suchard, Marc, Rijnbeek, Peter, Ryan, Patrick B., Prieto-Alhambra, Daniel. Characteristics, outcomes, and mortality amongst 133,589 patients with prevalent autoimmune diseases diagnosed with, and 48,418 hospitalised for COVID-19: a multinational distributed network cohort analysis. medRxiv : the preprint server for health sciences. 2020; #volume#(#issue#): #pages#. doi: https://dx.doi.org/10.1101/2020.11.24.20236802. Available at: http://ovidsp.ovid.com/ovidweb.cgi?T=JS&PAGE=reference&D=prem&NEWS=N&AN=33269355 |
| 41 | van Gassel, Rob,J.J., Bels, Julia L. M., Raafs, Anne, van Bussel, Bas,C.T., van de Poll, Marcel,C.G., Simons, Sami O., van der Meer, Lieke,W.L., Gietema, Hester A., Posthuma, Rein, van Santen, Susanne. High Prevalence of Pulmonary Sequelae at 3 Months After Hospital Discharge in Mechanically Ventilated COVID-19 Survivors. American journal of respiratory and critical care medicine. 2020; #volume#(#issue#): #pages#. doi: https://dx.doi.org/10.1164/rccm.202010-3823LE. Available at: http://ovidsp.ovid.com/ovidweb.cgi?T=JS&PAGE=reference&D=medp&NEWS=N&AN=33326353 |

| | |
|---|---|
| 42 | Walsh-Messinger, Julie, Manis, Hannah, Vrabec, Alison, Sizemore, Jenna, Bishof, Karyn, Debidda, Marcella, Malaspina, Dolores, Greenspan, Noah. The Kids Are Not Alright: A Preliminary Report of Post-COVID Syndrome in University Students. medRxiv : the preprint server for health sciences. 2020; #volume#(#issue#): #pages#. doi: https://dx.doi.org/10.1101/2020.11.24.20238261. Available at: http://ovidsp.ovid.com/ovidweb.cgi?T=JS&PAGE=reference&D=prem&NEWS=N&AN=33269366 |
| 43 | Manzo C., Serra-Mestres J., Isetta M., Castagna A.. Could COVID-19 anosmia and olfactory dysfunction trigger an increased risk of future dementia in patients with ApoE4?. Medical hypotheses. 2021; 147(#issue#): 110479. doi: http://dx.doi.org/10.1016/j.mehy.2020.110479. Available at: https://www.journals.elsevier.com/medical-hypotheses; http://ovidsp.ovid.com/ovidweb.cgi?T=JS&PAGE=reference&D=emexc&NEWS=N&AN=2010576366 |
| 44 | Mu M.. Effect on Novel Corona-Virus Pneumonia Patients' Rehabilitation Training of Tibetan Folk Music. Basic and Clinical Pharmacology and Toxicology. 2020; 127(#issue#): 267. doi: http://dx.doi.org/10.1111/bcpt.13461. Available at: http://ovidsp.ovid.com/ovidweb.cgi?T=JS&PAGE=reference&D=emexc&NEWS=N&AN=633830803 |
| 45 | You Z.. Effect of Aerobics on Rehabilitation Training of New Coronavirus Pneumonia Patients. Basic and Clinical Pharmacology and Toxicology. 2020; 127(#issue#): 272. doi: http://dx.doi.org/10.1111/bcpt.13461. Available at: http://ovidsp.ovid.com/ovidweb.cgi?T=JS&PAGE=reference&D=emexc&NEWS=N&AN=633830096 |
| 46 | Brigham, Emily, O'Toole, Jacqueline, Kim, Soo Yeon, Friedman, Michael, Daly, Laura, Kaplin, Adam, Swarthout, Meghan, Hasselfeld, Brian, Lantz-Garnish, Melissa, Vannorsdall, Tracy, Agranovich, Anna, Raju, Sarath, Parker, Ann. The Johns Hopkins Post-Acute COVID-19 Team (PACT): A Multidisciplinary, Collaborative, Ambulatory Framework Supporting COVID-19 Survivors. The American Journal of Medicine. 2021; #volume#(#issue#): #pages#. doi: https://dx.doi.org/10.1016/j.amjmed.2020.12.009. Available at: http://ovidsp.ovid.com/ovidweb.cgi?T=JS&PAGE=reference&D=medp&NEWS=N&AN=33444589 |
| 47 | Cortinovis, Monica, Perico, Norberto, Remuzzi, Giuseppe. Long-term follow-up of recovered patients with COVID-19. Lancet (London, England). 2021; #volume#(#issue#): #pages#. doi: https://dx.doi.org/10.1016/S0140-6736(21)00039-8. Available at: http://ovidsp.ovid.com/ovidweb.cgi?T=JS&PAGE=reference&D=medp&NEWS=N&AN=33428868 |
| 48 | Salman, David, Vishnubala, Dane, Le Feuvre, Peter, Beaney, Thomas, Korgaonkar, Jonathan, Majeed, Azeem, McGregor, Alison H.. Returning to physical activity after covid-19. BMJ (Clinical research ed.). 2021; 372(#issue#): m4721. doi: https://dx.doi.org/10.1136/bmj.m4721. Available at: http://ovidsp.ovid.com/ovidweb.cgi?T=JS&PAGE=reference&D=mesx&NEWS=N&AN=33419740 |
| 49 | AD,Soundararajan V, Awasthi S, Martin JT, Akama-Garren E, Puranik A, Liukasemsarn S, Venkatakrishnan AJ, Horo JC, Hurt RT, Williams AW, Gores GJ, Halamka J, Badley. Augmented curation of clinical notes of COVID-19 and influenza patients reveals that long-term neuropsychiatric and coagulopathic symptoms are more associated with COVID-19 (preprint). Medrxiv. 2021; #volume#(#issue#): 2021.01.03.20248997. doi: 10.1101/2021.01.03.20248997. Available at: http://medrxiv.org/content/early/2021/01/05/2021.01.03.20248997.abstract |
| 50 | Burges Watson DL, Campbell M, Hopkins C, Smith B, Kelly C, Deary,V.. Altered Smell and Taste: anosmia, parosmia and the long impact of Covid-19 (preprint). Medrxiv. 2020; #volume#(#issue#): 2020.11.26.20239152. doi: 10.1101/2020.11.26.20239152. Available at: http://medrxiv.org/content/early/2020/11/30/2020.11.26.20239152.abstract |
| 51 | Fan BE, Umapathi T, Chua K, Chia YW, Wong SW, Tan GWL, Chandrasekar S, Lum YH, Vasoo S, Dalan,R.. Delayed catastrophic thrombotic events in young and asymptomatic post COVID-19 patients; 7648538 CSR - Reports results. Journal of thrombosis and thrombolysis. 2020; #volume#(#issue#): #pages#. doi: 10.1007/s11239-020-02332-z. Available at: https://pubmed.ncbi.nlm.nih.gov/33159640 |
| 52 | Klein MN, Wang EW, Zimand P, Beauchamp H, Donis C, Ward MD, Martinez-Hernandez A, Tabatabai A, Baddley JW, Bloch EM, Mullins KE, Fontaine,M.J.. Kinetics of SARS-CoV-2 antibody responses pre- and post- COVID-19 convalescent plasma transfusion in patients with severe respiratory failure: an observational case-control study (preprint). Medrxiv. 2020; #volume#(#issue#): 2020.12.10.20247007. |

| | |
|---|---|
| | doi: 10.1101/2020.12.10.20247007. Available at: http://medrxiv.org/content/early/2020/12/11/2020.12.10.20247007.abstract |
| 53 | N,Juarez J, Smith C, Loo G, Nestor. 298 Impact of Presenting Vital Signs on Outcomes of Patients Hospitalized with Coronavirus. Annals of Emergency Medicine. 2020; 76(4): S115. doi: 10.1016/j.annemergmed.2020.09.312. Available at: https://www.embase.com/search/results?subaction=viewrecord&id=L2008409721&from=export |
| 54 | S,Bouslama B, Pierret C, Khelfaoui F, Bellanne-Chantelot C, Donadieu J, Heritier. Post-COVID-19 severe neutropenia. Pediatric blood & cancer. 2020; #volume#(#issue#): e28866. doi: 10.1002/pbc.28866. Available at: https://pubmed.ncbi.nlm.nih.gov/33355971 CSR - Reports results |
| 55 | Sudre CH, Murray B, Varsavsky T, Graham MS, Penfold RS, Bowyer RCE, Capdevila Pujol J, Klaser K, Antonelli M, Canas LS, Molteni E, Modat M, Cardoso MJ, May A, Ganesh S, Davies R, Nguyen LH, Drew DA, Astley CM, Joshi AD, Merino J, Tsereteli N, Fall T, Gomez MF, Duncan E, Menni C, Williams FMK, Franks PW, Chan AT, Wolf J, Ourselin S, Spector T, Steves,C.J.. Attributes and predictors of Long-COVID: analysis of COVID cases and their symptoms collected by the Covid Symptoms Study App (preprint). Medrxiv. 2020; #volume#(#issue#): 2020.10.19.20214494. doi: 10.1101/2020.10.19.20214494. Available at: http://medrxiv.org/content/early/2020/10/21/2020.10.19.20214494.abstract |
| 56 | T,Taboada M, Carinena A, Moreno E, Rodriguez N, Dominguez MJ, Casal A, Riveiro V, Diaz-Vieito M, Valdes L, Alvarez J, Seoane-Pillado. Post-COVID-19 functional status six-months after hospitalization. Journal of infection. 2020; #volume#(#issue#): #pages#. doi: 10.1016/j.jinf.2020.12.022. Available at: https://pubmed.ncbi.nlm.nih.gov/33373650 CSR - Reports results |
| 57 | T,Taboada M, Moreno E, Carinena A, Rey T, Pita-Romero R, Leal S, Sanduende Y, Rodriguez A, Nieto C, Vilas E, Ochoa M, Cid M, Seoane-Pillado. Quality of life, functional status, and persistent symptoms after intensive care of COVID-19 patients. British journal of anaesthesia. 2020; #volume#(#issue#): #pages#. doi: 10.1016/j.bja.2020.12.007. Available at: https://pubmed.ncbi.nlm.nih.gov/33413976 CSR - Reports results |
| 58 | Chopra, V., Flanders, S. A., O'Malley, M., Malani, A. N., Prescott, H. C.. Sixty-Day Outcomes Among Patients Hospitalized With COVID-19. Annals of Internal Medicine. 2020; #volume#(#issue#): #pages#. doi: 10.7326/M20-5661 [doi]. Available at: #URL# |
| 59 | National Institute for Health and,Care Excellence. COVID-19 rapid guideline: managing the long-term effects of COVID-19. #journal#. 2020; 2021(#issue#): #pages#. doi: #DOI#. Available at: https://www.nice.org.uk/guidance/ng188 |
| 60 | Nau, J. -Y. COVID-19: What do we know today about its various sequelae?; 32833371. Revue Medicale Suisse. 2020; 16(701): 1466. doi: #DOI#. Available at: https://www.scopus.com/inward/record.uri?eid=2-s2.0-85090072662&partnerID=40&md5=cd80c2ec0d9407fefdc6279d1d5cbfbb |
| 61 | Delafontaine, A., Ditcharles, S., Hussein, T., Hoffschir, M., Plantefève, G., Michon, D.. Physiotherapy and COVID-19: A major public health role to short, medium and long terms in the patient's rehabilitation process. Kinesitherapie. 2020; 20(223): 11. doi: 10.1016/j.kine.2020.05.005. Available at: https://www.scopus.com/inward/record.uri?eid=2-s2.0-85088677591&doi=10.1016%2fj.kine.2020.05.005&partnerID=40&md5=e2de04c959bff9ee3fee6ed9a780dfdf |
| 62 | Grácio, S., Koçer, S.. Rehabilitation is crucial for severe COVID-19 survivors; 32496707. Revue Medicale Suisse. 2020; 16(696): 1170. doi: #DOI#. Available at: https://www.scopus.com/inward/record.uri?eid=2-s2.0-85086008274&partnerID=40&md5=f05a1138513c5a0b6308bb6ce2b36aed |
| 63 | Cirulli, Elizabeth T., Schiabor Barrett, Kelly M., Riffle, Stephen, Bolze, Alexandre, Neveux, Iva, Dabe, Shaun, Grzymski, Joseph J., Lu, James T., Washington, Nicole L.. Long-term COVID-19 symptoms in a large unselected population. medRxiv. 2020; #volume#(#issue#): 2020.10.07.20208702. doi: 10.1101/2020.10.07.20208702. Available at: http://medrxiv.org/content/early/2020/12/01/2020.10.07.20208702.abstract |
| 64 | Eiros, Rocio, Barreiro-Perez, Manuel, Martin-Garcia, Ana, Almeida, Julia, Villacorta, Eduardo, Perez-Pons, Alba, Merchan, Soraya, Torres-Valle, Alba, Pablo, Clara Sánchez, González-Calle, David, Perez-Escurza, Oihane, Toranzo, Inés, Díaz-Pelaez, Elena, Fuentes-Herrero, Blanca, Macías-Alvarez, Laura, Oliva-Ariza, |

| | |
|---|---|
| | Guillermo, Lecrevisse, Quentin, Fluxa, Rafael, Bravo-Grande, Jose, Orfao, Alberto, Sanchez, Pedro L.. Pericarditis and myocarditis long after SARS-CoV-2 infection: a cross-sectional descriptive study in health-care workers. medRxiv. 2020; #volume#(#issue#): 2020.07.12.20151316. doi: 10.1101/2020.07.12.20151316. Available at: http://medrxiv.org/content/early/2020/07/14/2020.07.12.20151316.abstract |
| 65 | Poyraz, Burç Çağrı, Poyraz, Cana Aksoy, Olğun, Yeşim, Gürel, Özge, Alkan, Sena, Özdemir, Yusuf Emre, İnanç Balkan, İlker, Karaali, R. ıdvan. Psychiatric morbidity and protracted symptoms in recovered COVID-19 patients. medRxiv. 2020; #volume#(#issue#): 2020.10.07.20208249. doi: 10.1101/2020.10.07.20208249. Available at: http://medrxiv.org/content/early/2020/10/09/2020.10.07.20208249.abstract |
| 66 | Santis, Lucía Valiente-De, Pérez-Camacho, Inés, Sobrino, Beatriz, González, Gracia Eugenia, Ruíz-Mesa, Juan Diego, Plata, Antonio, Márquez-Gómez, Ignacio, Delgado-Fernández, Marcial, Castaño, Manuel, Oñate, Francisco, Orihuela, Francisco, Palop, Begoña, María Reguera, José. Clinical and immunoserological status 12 weeks after infection with COVID-19: prospective observational study. medRxiv. 2020; #volume#(#issue#): 2020.10.06.20206060. doi: 10.1101/2020.10.06.20206060. Available at: http://medrxiv.org/content/early/2020/10/08/2020.10.06.20206060.abstract |
| 67 | Dennis, Andrea, Wamil, Malgorzata, Kapur, Sandeep, Alberts, Johann, Badley, Andrew D., Decker, Gustav Anton, Rizza, Stacey A., Banerjee, Rajarshi, Banerjee, Amitava, ,. Multi-organ impairment in low-risk individuals with long COVID. medRxiv. 2020; #volume#(#issue#): 2020.10.14.20212555. doi: 10.1101/2020.10.14.20212555. Available at: http://medrxiv.org/content/early/2020/10/16/2020.10.14.20212555.abstract |
| 68 | Klein, Hadar, Asseo, Kim, Karni, Noam, Benjamini, Yuval, Nir-Paz, Ran, Muszkat, Mordechai, Israel, Sarah, Niv, Masha Y.. Onset, duration, and persistence of taste and smell changes and other COVID-19 symptoms: longitudinal study in Israeli patients. medRxiv. 2020; #volume#(#issue#): 2020.09.25.20201343. doi: 10.1101/2020.09.25.20201343. Available at: http://medrxiv.org/content/early/2020/10/17/2020.09.25.20201343.abstract |
| 69 | Davis, Hannah E., Assaf, Gina S., McCorkell, Lisa, Wei, Hannah, Low, Ryan J., Re'em, Yochai, Redfield, Signe, Austin, Jared P., Akrami, Athena. Characterizing Long COVID in an International Cohort: 7 Months of Symptoms and Their Impact. medRxiv. 2020; #volume#(#issue#): 2020.12.24.20248802. doi: 10.1101/2020.12.24.20248802. Available at: http://medrxiv.org/content/early/2020/12/27/2020.12.24.20248802.abstract |
| 70 | Assaf, G., Davis, H.. An Analysis of the Prolonged COVID-19 Symptoms Survey by Patient-Led Research Team. #journal#. 2020; #volume#(#issue#): #pages#. doi: #DOI#. Available at: https://patientresearchcovid19.com/research/report-1/ |
| 71 | National COVID-19 Clinical Evidence Task Force. CARE OF PEOPLE WHO EXPERIENCE SYMPTOMS POST ACUTE COVID-19. #journal#. 2020; #volume#(#issue#): #pages#. doi: #DOI#. Available at: https://covid19evidence.net.au/wp-content/uploads/FLOWCHART-11-CARE-OF-PEOPLE-WHO-EXPERIENCE-SYMPTOMS-POST-ACUTE-COVID19.pdf?=201218-55302 |
| 72 | Prevention, Centers for Disease Control and. Post-COVID conditions: information for healthcare providers. 2021. #journal#. #year#; #volume#(#issue#): #pages#. doi: #DOI#. Available at: https://www.cdc.gov/coronavirus/2019-ncov/hcp/clinical-care/post-covid-conditions.html |
| 73 | Newfoundland and Labrador Centre for Applied Health Research, Memorial University of Newfoundland. What are the medium-term and long-term health impacts of COVID-19 on infection survivors?. #journal#. #year#; #volume#(#issue#): #pages#. doi: #DOI#. Available at: https://www.nlcahr.mun.ca/CHRSP/COVIDQRRMedandLongtermeffects.pdf |
| 74 | NCT04678830. COVID-19 Long-Haulers Study. A Phase 2, Randomized, Double Blind, Placebo Controlled Study to Evaluate the Efficacy and Safety of Leronlimab in Patients Experiencing Prolonged Coronavirus Disease 2019 (COVID-19) Symptoms. 2020///; #volume#(#issue#): #pages#. doi: #DOI#. Available at: http://ovidsp.ovid.com/ovidweb.cgi?T=JS&PAGE=reference&D=cctr&NEWS=N&AN=CN-02208412 |
| 75 | ACTRN12621000031864. Home-based virtual rehabililtation for people with persistent symptoms after COVID-19 disease (Long COVID-19). A randomized controlled trial investigating the effect of home-based, |

| | |
|---|---|
| | virtual supervised pulmonary rehabilitation program on the exercise capacity of patients with persistent symptoms after COVID-19 disease: a substudy of the ADAPT COVID-19 study. 2021///; #volume#(#issue#): #pages#. doi: #DOI#. Available at: http://ovidsp.ovid.com/ovidweb.cgi?T=JS&PAGE=reference&D=cctr&NEWS=N&AN=CN-02238256 |
| 76 | NCT04705831. Study to Evaluate the Benefit of RUCONEST in Improving Neurological Symptoms in Post COVID-19 Infection. A Randomized, Double Blind, Placebo Controlled, Cross-Over, Proof-of-Concept Study to Evaluate the Benefit of RUCONEST (C1 Esterase Inhibitor Recombinant]) in Improving Neurological Symptoms in Post-SARS-CoV-2 Infection. 2021///; #volume#(#issue#): #pages#. doi: #DOI#. Available at: http://ovidsp.ovid.com/ovidweb.cgi?T=JS&PAGE=reference&D=cctr&NEWS=N&AN=CN-02233908 |
| 77 | Sari, Cenk, Simsek, Ersin Cagri, Ekmekci, Cenk, Kucukukur, Murat, Ozdogan, Oner, Kose, Sukran. Evaluation of endothelial function patient infected with SARS-CoV-2 virus. Anatolian Journal of Cardiology. 2020///; 24(#issue#): 70. doi: #DOI#. Available at: https://jag.journalagent.com/anatoljcardiol/pdfs/AJC_24_50_1_139.pdf,http://ovidsp.ovid.com/ovidweb.cgi?T=JS&PAGE=reference&D=emexb&NEWS=N&AN=634645690 |
| 78 | Vasconcello-Castillo, Luis, Torres-Castro, Rodrigo, Vilaro, Jordi, Torralba, Yolanda, Peroy-Badal, Renata. The Relation between Persistent Poor Health Post-COVID-19 and Respiratory Complications or Initial Disease Severity. Annals of the American Thoracic Society. 2021///; #volume#(#issue#): #pages#. doi: http://dx.doi.org/10.1513/AnnalsATS.202101-045LE. Available at: http://ovidsp.ovid.com/ovidweb.cgi?T=JS&PAGE=reference&D=emexb&NEWS=N&AN=634332727 |
| 79 | CTRI/2020/11/029188. Quality of life in post covid affected patients. Assessment of quality of life in patients who have recovered from SARS -COV 2 infection. 2020///; #volume#(#issue#): #pages#. doi: #DOI#. Available at: http://ovidsp.ovid.com/ovidweb.cgi?T=JS&PAGE=reference&D=cctr&NEWS=N&AN=CN-02237633 |
| 80 | Wainwright, Laurence. Lessons from psychiatry for treating post-acute Covid-19 fatigue. Australasian psychiatry : bulletin of Royal Australian and New Zealand College of Psychiatrists. 2021///; #volume#(#issue#): 10398562211003598. doi: https://dx.doi.org/10.1177/10398562211003598. Available at: http://ovidsp.ovid.com/ovidweb.cgi?T=JS&PAGE=reference&D=medp&NEWS=N&AN=33818173 |
| 81 | Wijeratne, Tissa, Crewther, Sheila. COVID-19 and long-term neurological problems: Challenges ahead with Post-COVID-19 Neurological Syndrome. Australian journal of general practice. 2021///; 50(#issue#): #pages#. doi: http://dx.doi.org/10.31128/AJGP-COVID-43. Available at: http://ovidsp.ovid.com/ovidweb.cgi?T=JS&PAGE=reference&D=emexb&NEWS=N&AN=634213201 |
| 82 | Kostov, Kosta V. Ongoing covid-19 syndrome and post-covid-19 syndrome: Long-term symptoms and residual changes after sars-cov-2 infection. Biomedical Reviews. 2020///; 31(#issue#): 117. doi: http://dx.doi.org/10.14748/bmr.v31.7711. Available at: https://journals.mu-varna.bg/index.php/bmr/article/download/7711/6806,http://ovidsp.ovid.com/ovidweb.cgi?T=JS&PAGE=reference&D=emedx&NEWS=N&AN=2006886878 |
| 83 | Torjesen, Ingrid. Covid-19: Middle aged women face greater risk of debilitating long term symptoms. BMJ (Clinical research ed.). 2021///; 372(#issue#): #pages#. doi: http://dx.doi.org/10.1136/bmj.n829. Available at: http://ovidsp.ovid.com/ovidweb.cgi?T=JS&PAGE=reference&D=emexb&NEWS=N&AN=634665775 |
| 84 | JT Cevik M, Jesudason E, Lim WS, Rello J, Amuasi J, Bozza F, Palmieri C, Munblit D, Holter JC, Kildal AB, Reyes LF, Russell CD, Ho A, Turtle L, Drake TM, Beltrame A, Hann K, Bangura IR, Fowler R, Lakoh S, Berry C, Lowe DJ, McPeake J, Hashmi M, Dyrhol-Riise AM, Donohue C, Plotkin D, Hardwick H, Elkheir N, Lone NI, Docherty A, Harrison E, Baille JK, Carson G, Semple MG, Scott, Sigfrid L. What is the recovery rate and risk of long-term consequences following a diagnosis of COVID-19? A harmonised, global longitudinal observational study protocol; 7948153 CSR - Reports results. BMJ open. 2021///; 11(3): e043887. doi: 10.1136/bmjopen-2020-043887. Available at: https://pubmed.ncbi.nlm.nih.gov/33692181 |
| 85 | NCT04764981. Olfactory Training for Olfactory Dysfunction After Coronavirus Disease - 19 (COVID-19). Clinical Outcomes of Olfactory Training for Treatment of Olfactory Dysfunction After COVID-19. 2021///; |

| | |
|---|---|
| | #volume#(#issue#): #pages#. doi: #DOI#. Available at: http://ovidsp.ovid.com/ovidweb.cgi?T=JS&PAGE=reference&D=cctr&NEWS=N&AN=CN-02249160 |
| 86 | Murray, Terry. Unpacking "long COVID". CMAJ : Canadian Medical Association journal = journal de l'Association medicale canadienne. 2021///; 193(9): E318. doi: http://dx.doi.org/10.1503/cmaj.1095923. Available at: http://ovidsp.ovid.com/ovidweb.cgi?T=JS&PAGE=reference&D=emexb&NEWS=N&AN=634449654 |
| 87 | NCT04695704. Montelukast Clinical Trial in Mild-moderate Respiratory Symptoms in Patients With Long-COVID (ESPERANZA COVID). Double-blind Randomized Clinical Trial, Placebo-controlled to Assess the Efficacy of Montelukast in Mild-moderate Respiratory Symptoms in Patients With Long-COVID: E-SPERANZA COVID PROJECT. 2021///; #volume#(#issue#): #pages#. doi: #DOI#. Available at: http://ovidsp.ovid.com/ovidweb.cgi?T=JS&PAGE=reference&D=cctr&NEWS=N&AN=CN-02233650 |
| 88 | Iqbal, Fahad M, Lam, Kyle, Sounderajah, Viknesh, Elkin, Sarah, Ashrafian, Hutan, Darzi, Ara. Understanding the survivorship burden of long COVID. EClinicalMedicine. 2021///; 33(#issue#): 100767. doi: http://dx.doi.org/10.1016/j.eclinm.2021.100767. Available at: http://www.thelancet.com/journals/eclinm/issue/current,http://ovidsp.ovid.com/ovidweb.cgi?T=JS&PAGE=reference&D=emexb&NEWS=N&AN=2011127975 |
| 89 | NCT04795557. Efficacy of Adaptogens in Patients With Long COVID-19. Effect of ADAPT232 Supplementation on Recovery of Patients in Rehabilitation Period in Long COVID-19: a Randomized, Double-blind, Placebo-controlled Trial. 2021///; #volume#(#issue#): #pages#. doi: #DOI#. Available at: http://ovidsp.ovid.com/ovidweb.cgi?T=JS&PAGE=reference&D=cctr&NEWS=N&AN=CN-02249877 |
| 90 | CTRI/2020/12/029476. A clinical trial to study the effect of herbal medicine on enhancing immune function in COVID recovery patients. Effect of purified standard extract of Withania somnifera, Phyllanthus emblica and Shilajeet in SARS-CoV-2 recovery patients. 2020///; #volume#(#issue#): #pages#. doi: #DOI#. Available at: http://ovidsp.ovid.com/ovidweb.cgi?T=JS&PAGE=reference&D=cctr&NEWS=N&AN=CN-02240599 |
| 91 | NCT04791072. Tele-coaching in Patients With COVID-19. Effectiveness of Pulmonary Rehabilitation With Tele-coaching in Patients With COVID-19. 2021///; #volume#(#issue#): #pages#. doi: #DOI#. Available at: http://ovidsp.ovid.com/ovidweb.cgi?T=JS&PAGE=reference&D=cctr&NEWS=N&AN=CN-02249789 |
| 92 | Aldcroft, Daniel, Fearon, Anna, George, Ryan, Malloupa, Eleni, Ng, Cassandra. Assessing the impact of covid-19 and frailty in older people. European Geriatric Medicine. 2020///; 11(#issue#): S88. doi: http://dx.doi.org/10.1007/s41999-020-00428-6. Available at: http://ovidsp.ovid.com/ovidweb.cgi?T=JS&PAGE=reference&D=emexb&NEWS=N&AN=634448270 |
| 93 | Kotovskaya, Natalia, Belokrylov, Igor, Rozanov, Alexander, Manevich, Tatiana, Ostapenko, Valentina, Schedrina, Anna, Izyumov, Andrey, Eruslanova, Ksenia, Balaeva, Madina, Kotovskaya, Yulia, Tkacheva, Olga, Deryugina, Natalia. Age and intensity of psychopathological symptoms in COVID-19 Survivals. European Geriatric Medicine. 2020///; 11(#issue#): S260. doi: http://dx.doi.org/10.1007/s41999-020-00428-6. Available at: http://ovidsp.ovid.com/ovidweb.cgi?T=JS&PAGE=reference&D=emexb&NEWS=N&AN=634448464 |
| 94 | McElroy, Morven, Quinn, Terry, McCarthy, Carol, Waddell, Greg, Cameron, Ewen, McIlroy, Kathyrn, McGovern, Aine. Experience of COVID-19 in a UK older adult rehabilitation hospital, An observational case series. European Geriatric Medicine. 2020///; 11(#issue#): S56. doi: http://dx.doi.org/10.1007/s41999-020-00428-6. Available at: http://ovidsp.ovid.com/ovidweb.cgi?T=JS&PAGE=reference&D=emexb&NEWS=N&AN=634448196 |
| 95 | Medetalibeyoglu, Alpay, Catma, Yunus, Senkal, Naci, Kose, Murat, Tukek, Tufan, Bahat, Gulistan, Karan, Mehmet Akif, Oren, Meryem Merve, Caparali, Emine Bilge, Bayrakdar, Sena, Basaran, Seniha, Erelel, Mustafa. Follow-up of patients after acute COVID-19: Results on persistent symptoms and control imaging findings. European Geriatric Medicine. 2020///; 11(#issue#): S35. doi: http://dx.doi.org/10.1007/s41999-020-00428-6. Available at: http://ovidsp.ovid.com/ovidweb.cgi?T=JS&PAGE=reference&D=emexb&NEWS=N&AN=634448823 |
| 96 | Shodiyorovich, Kazakov Bekzod, Tadjiyevna, Khodjiyeva Dilbar. Clinical rehabilitation of patients with cerebral stroke caused by a stroke with COVID-19. European Journal of Molecular and Clinical Medicine. |

| | |
|---|---|
| | 2021///; 8(1): 1446. doi: #DOI#. Available at: https://ejmcm.com/article_7281_e9cff8a959c95d1b6b4a8c67c34c2801.pdf,http://ovidsp.ovid.com/ovidweb.cgi?T=JS&PAGE=reference&D=emexb&NEWS=N&AN=2011070035 |
| 97 | Lapp, Charles W, John, Joseph F. Managing COVID-19 post viral Fatigue Syndrome. Fatigue: Biomedicine, Health and Behavior. 2021///; #volume#(#issue#): #pages#. doi: http://dx.doi.org/10.1080/21641846.2021.1890347. Available at: http://www.tandfonline.com/toc/rftg20/current,http://ovidsp.ovid.com/ovidweb.cgi?T=JS&PAGE=reference&D=emexb&NEWS=N&AN=2010554485 |
| 98 | Kumar, Sanjay, Veldhuis, Alfred, Malhotra, Tina. Neuropsychiatric and Cognitive Sequelae of COVID-19. Frontiers in psychology. 2021///; 12(#issue#): 577529. doi: https://dx.doi.org/10.3389/fpsyg.2021.577529. Available at: http://ovidsp.ovid.com/ovidweb.cgi?T=JS&PAGE=reference&D=pmnm&NEWS=N&AN=33737894 |
| 99 | Catalfamo, Collin J, Heslin, Kelly M, Shilen, Alexandra, Khan, Sana M, Hunsaker, Josh R, Austhof, Erika, Farland, Leslie V, Hoskinson, Joshua, Harris, Robin B, Chen, Zhao, Klimentidis, Yann C, Bell, Melanie L, Ernst, Kacey C, Pogreba-Brown, Kristen, Barraza, Leila, Cordova-Marks, Felina, Jacobs, Elizabeth T, Garcia-Filion, Pamela, Jehn, Megan, Kohler, Lindsay N, Lutrick, Karen. Design of the Arizona CoVHORT: A Population-Based COVID-19 Cohort. Frontiers in public health. 2021///; 9(#issue#): 620060. doi: http://dx.doi.org/10.3389/fpubh.2021.620060. Available at: http://ovidsp.ovid.com/ovidweb.cgi?T=JS&PAGE=reference&D=emexb&NEWS=N&AN=634436480 |
| 100 | Clavario, Piero, Barbara, Cristina, Porcile, Annalisa, Russo, Carmelo, Bottaro, Luigi, Caltabellotta, Marta, De Marzo, Vincenzo, Lotti, Roberta, Bona, Roberta Della, Canepa, Marco, Ameri, Pietro, Porto, Italo, Hautala, Arto. Prospective cardiopulmonary stress testing evaluation of non-severe COVID-19 patients at three months after hospital discharge. Giornale italiano di cardiologia. 2020///; 21(12): e80. doi: #DOI#. Available at: https://www.giornaledicardiologia.it/r.php?v=3492&a=34754&l=342561&f=allegati/03492_0_12/fulltext/01 Abstract SIC_2020 (e3-e177).pdf,http://ovidsp.ovid.com/ovidweb.cgi?T=JS&PAGE=reference&D=emexb&NEWS=N&AN=634463348 |
| 101 | Yan, Wudan. Understanding the Long-Term Impacts of COVID-19 in Survivors. IEEE Pulse. 2021///; 12(1): 19. doi: http://dx.doi.org/10.1109/MPULS.2021.3052616. Available at: http://pulse.embs.org/pastissues.html,http://ovidsp.ovid.com/ovidweb.cgi?T=JS&PAGE=reference&D=emexb&NEWS=N&AN=634317073 |
| 102 | NCT04678700. Implementation of a Respiratory Physiotherapy Program in Post COVID-19 Patients Through Tele-assistance. Implementation of a Respiratory Physiotherapy Program in Post COVID-19 Patients Through Tele-assistance. 2020///; #volume#(#issue#): #pages#. doi: #DOI#. Available at: http://ovidsp.ovid.com/ovidweb.cgi?T=JS&PAGE=reference&D=cctr&NEWS=N&AN=CN-02209522 |
| 103 | Prakash, Jay, Bhattacharya, Pradip K, Priye, Shio, Kumar, Naveen. Post-COVID-19 Pulmonary Fibrosis: A Lifesaving Challenge. Indian journal of critical care medicine : peer-reviewed, official publication of Indian Society of Critical Care Medicine. 2021///; 25(1): 104. doi: https://dx.doi.org/10.5005/jp-journals-10071-23709. Available at: http://ovidsp.ovid.com/ovidweb.cgi?T=JS&PAGE=reference&D=pmnm&NEWS=N&AN=33603312 |
| 104 | Kumar, Ankit, Dawra, Saurabh. Thromboembolic complications following recovery from COVID-19 disease: A case series. Indian Journal of Hematology and Blood Transfusion. 2020///; 36(1): S207. doi: http://dx.doi.org/10.1007/s12288-020-01384-8. Available at: http://ovidsp.ovid.com/ovidweb.cgi?T=JS&PAGE=reference&D=emexb&NEWS=N&AN=634127745 |
| 105 | Bueno, M, De Alba, M, Leon, R, Rodriguez, J, Lopez, C, Duenas, J M, Torres, E, Bautista, D, Robles, J C, De La Fuente-Martos, C. Comparison of clinical features and prognosis factors in patients with acute respiratory syndrome related to SARS-COV2 versus other respiratory virus. Intensive Care Medicine Experimental. 2020///; 8(#issue#): #pages#. doi: http://dx.doi.org/10.1186/s40635-020-00354-8. Available at: http://ovidsp.ovid.com/ovidweb.cgi?T=JS&PAGE=reference&D=emexb&NEWS=N&AN=634009545 |

| | |
|---|---|
| 106 | Pastora-Bernal, Jose, Estebanez-Perez, Maria, Martin-Valero, Rocio, Molina-Torres, Guadalupe, Garcia-Lopez, Francisco, Sobrino-Sanchez, Raquel. Telerehabilitation intervention in patients with covid-19 after hospital discharge to improve functional capacity and quality of life. Study protocol for a multicenter randomized clinical trial. International Journal of Environmental Research and Public Health. 2021///; 18(6): 1. doi: http://dx.doi.org/10.3390/ijerph18062924. Available at: https://www.mdpi.com/1660-4601/18/6/2924/pdf,http://ovidsp.ovid.com/ovidweb.cgi?T=JS&PAGE=reference&D=emexb&NEWS=N&AN=2006723946 |
| 107 | Del Rio, Carlos, Collins, Lauren F, Malani, Preeti. Long-term Health Consequences of COVID-19. JAMA - Journal of the American Medical Association. 2020///; 324(17): 1723. doi: http://dx.doi.org/10.1001/jama.2020.19719. Available at: http://jama.jamanetwork.com/journal.aspx,http://ovidsp.ovid.com/ovidweb.cgi?T=JS&PAGE=reference&D=emexb&NEWS=N&AN=633179266 |
| 108 | Rossavik, Eevar B, Iftikhar, Zain, Hussain, Iftikhar. A Survey of Respiratory Symptoms Reported in Patients Following Hospitalization with COVID-19 Infection. Journal of Allergy and Clinical Immunology. 2021///; 147(2): AB243. doi: http://dx.doi.org/10.1016/j.jaci.2020.12.033. Available at: http://ovidsp.ovid.com/ovidweb.cgi?T=JS&PAGE=reference&D=emexb&NEWS=N&AN=2010823554 |
| 109 | Kurihara, C, Manerikar, A, Kandula, V, Bharat, A. Clinical Characteristics and Outcomes of Extracorporeal Membrane Oxygenation Support in COVID-19: Retrospective Study of Single Center Experience. Journal of Heart and Lung Transplantation. 2021///; 40(4): S398. doi: http://dx.doi.org/10.1016/j.healun.2021.01.1121. Available at: http://ovidsp.ovid.com/ovidweb.cgi?T=JS&PAGE=reference&D=emexb&NEWS=N&AN=2011430639 |
| 110 | Garcia-Pachon E Soler-Sempere MJ, Zamora-Molina L, Baeza-Martinez C, Ruiz-Alcaraz S, Padilla-Navas,I., Grau-Delgado J. Low prevalence of post-COVID-19 syndrome in patients with asthma; 8017946 CSR - Reports results. Journal of infection. 2021///; #volume#(#issue#): #pages#. doi: 10.1016/j.jinf.2021.03.023. Available at: https://pubmed.ncbi.nlm.nih.gov/33819552 |
| 111 | Meagher, Timothy. Long COVID - An Early Perspective. Journal of insurance medicine (New York, N.Y.). 2021///; #volume#(#issue#): #pages#. doi: http://dx.doi.org/10.17849/insm-49-1-1-5.1. Available at: http://ovidsp.ovid.com/ovidweb.cgi?T=JS&PAGE=reference&D=emedx&NEWS=N&AN=634692770 |
| 112 | Dinapoli, Loredana, Camardese, Giovanni, Ferrarese, Daniele, Belella, Daniela, Chieffo, Daniela. "A Lion Sleeping Next to Me": Preliminary Considerations on the Psychological Consequences of Surviving COVID-19. Journal of the Academy of Consultation-Liaison Psychiatry. 2021///; #volume#(#issue#): #pages#. doi: https://dx.doi.org/10.1016/j.jaclp.2020.12.003. Available at: http://ovidsp.ovid.com/ovidweb.cgi?T=JS&PAGE=reference&D=medp&NEWS=N&AN=33681862 |
| 113 | Sancak, Baris, Agirbas, Urun Ozer, Kilic, Cenk. Long COVID and Its Psychiatric Aspects. Journal of the Academy of Consultation-Liaison Psychiatry. 2021///; #volume#(#issue#): #pages#. doi: https://dx.doi.org/10.1016/j.jaclp.2021.03.003. Available at: http://ovidsp.ovid.com/ovidweb.cgi?T=JS&PAGE=reference&D=medp&NEWS=N&AN=33817685 |
| 114 | C Wijeratne, Wijeratne T. Clinical utility of serial systemic immune inflammation indices (SSIIi) in the context of post covid-19 neurological syndrome (PCNS); 7895697 CSR - Reports results. Journal of the neurological sciences. 2021///; 423(#issue#): 117356. doi: 10.1016/j.jns.2021.117356. Available at: https://pubmed.ncbi.nlm.nih.gov/33636659 |
| 115 | Da Silva Lameira, Fernanda, Shahmirzadi, Maryam Sadough, Heide, Richard Vander, Fox, Sharon. Pulmonary pathology in long-haul COVID-19 at autopsy. Laboratory Investigation. 2021///; 101(#issue#): 12. doi: http://dx.doi.org/10.1038/s41374-021-00548-y. Available at: http://ovidsp.ovid.com/ovidweb.cgi?T=JS&PAGE=reference&D=emedx&NEWS=N&AN=634717397 |
| 116 | McCaddon, Andrew, Regland, Bjorn. COVID-19: A methyl-group assault?. Medical hypotheses. 2021///; 149(#issue#): 110543. doi: http://dx.doi.org/10.1016/j.mehy.2021.110543. Available at: https://www.journals.elsevier.com/medical-hypotheses,http://ovidsp.ovid.com/ovidweb.cgi?T=JS&PAGE=reference&D=emexb&NEWS=N&AN=2011173341 |

| | |
|---|---|
| 117 | Bieksiene, Kristina, Zaveckiene, Jurgita, Malakauskas, Kestutis, Vaguliene, Neringa, Zemaitis, Marius, Miliauskas, Skaidrius. Post COVID-19 Organizing Pneumonia: The Right Time to Interfere. Medicina (Kaunas, Lithuania). 2021///; 57(3): #pages#. doi: https://dx.doi.org/10.3390/medicina57030283. Available at: http://ovidsp.ovid.com/ovidweb.cgi?T=JS&PAGE=reference&D=mesx&NEWS=N&AN=33803690 |
| 118 | Rosales-Castillo, Antonio, Garcia de Los Rios, Carlos, Mediavilla Garcia, Juan Diego. Persistent symptoms after acute COVID-19 infection: importance of follow-up. Medicina clinica (English ed.). 2021///; 156(1): 35. doi: https://dx.doi.org/10.1016/j.medcle.2020.08.003. Available at: http://ovidsp.ovid.com/ovidweb.cgi?T=JS&PAGE=reference&D=pmnm&NEWS=N&AN=33521308 |
| 119 | A Khunti K, Nafilyan V, Maddox T, Humberstone B, Diamond SI, Banerjee, Ayoubkhani D. Epidemiology of post-COVID syndrome following hospitalisation with coronavirus: a retrospective cohort study (preprint). Medrxiv. 2021///; #volume#(#issue#): 2021.01.15.21249885. doi: 10.1101/2021.01.15.21249885. Available at: http://medrxiv.org/content/early/2021/01/15/2021.01.15.21249885.abstract |
| 120 | A Wilcox JA, Babady NE, McMillen T, Vachha BA, Halpern NA, Dhawan V, Rosenblum M, Iacobuzio-Donahue CA, Avila EK, Santomasso B, Boire, Remsik J. Inflammatory leptomeningeal cytokines mediate delayed COVID-19 encephalopathy (preprint). Medrxiv. 2020///; #volume#(#issue#): 2020.09.15.20195511. doi: 10.1101/2020.09.15.20195511. Available at: https://pubmed.ncbi.nlm.nih.gov/32995805 |
| 121 | Chioh FWJ Young B, Wu K-X, Siau A, Krishnan S, Chan Y-H, Teo LLY, Gao F, Tan RS, Zhong L, Koh ASM, Tan SY, Tambyah PA, Renia L, Ng LFP, Lye DCB, Cheung,C., Fong S-W. Convalescent COVID-19 patients are susceptible to endothelial dysfunction due to persistent immune activation (preprint). Medrxiv. 2020///; #volume#(#issue#): 2020.11.16.20232835. doi: 10.1101/2020.11.16.20232835. Available at: http://medrxiv.org/content/early/2020/11/18/2020.11.16.20232835.abstract |
| 122 | Davis HE McCorkell L, Wei H, Low RJ, Re'em Y, Redfield S, Austin JP, Akrami,A., Assaf G S. Characterizing Long COVID in an International Cohort: 7 Months of Symptoms and Their Impact (preprint). Medrxiv. 2020///; #volume#(#issue#): 2020.12.24.20248802. doi: 10.1101/2020.12.24.20248802. Available at: http://medrxiv.org/content/early/2020/12/27/2020.12.24.20248802.abstract |
| 123 | Garcia-Abellan J Fernandez-Gonzalez M, Garcia JA, Agullo V, Andreo M, Ruiz S, Galiana A, Gutierrez F, Masia,M., Padilla S. Long-term clinical, virological and immunological outcomes in patients hospitalized for COVID-19: antibody response predicts long COVID (preprint). Medrxiv. 2021///; #volume#(#issue#): 2021.03.08.21253124. doi: 10.1101/2021.03.08.21253124. Available at: http://medrxiv.org/content/early/2021/03/08/2021.03.08.21253124.abstract |
| 124 | Hasan ATMH Khan N, Munna NH, Choton WR, Arefin MK, Khan MAAZ, Mansur M, Hassan R, Siddiqui N, Arefin MS, Afroje N, Islam,M.S., Islam M S. Assessment of Post SARS CoV 2 Fatigue among Physicians Working in COVID Designated Hospitals in Dhaka, Bangladesh (preprint). Medrxiv. 2021///; #volume#(#issue#): 2021.02.08.21251352. doi: 10.1101/2021.02.08.21251352. Available at: http://medrxiv.org/content/early/2021/02/10/2021.02.08.21251352.abstract |
| 125 | Hwang CE Christle JW, Froelicher V, Wheeler MT, Moneghetti,K.J., Kussman A. Findings from Cardiovascular Evaluation of NCAA Division I Collegiate Student-Athletes after Asymptomatic or Mildly Symptomatic SARS-CoV-2 Infection (preprint). Medrxiv. 2021///; #volume#(#issue#): 2021.01.07.21249407. doi: 10.1101/2021.01.07.21249407. Available at: http://medrxiv.org/content/early/2021/01/08/2021.01.07.21249407.abstract |
| 126 | JT Cevik M, Jesudason E, Lim WS, Rello J, Amuasi JH, Bozza F, Palmieri C, Munblit D, Holter JC, Kildal AB, Russell CD, Ho A, Turtle L, Drake TM, Beltrame A, Hann K, Bangura IR, Fowler R, Lakoh S, Berry C, Lowe DJ, McPeake J, Hashmi M, Dyrhol-Riise AM, Donohue C, Plotkin DR, Hardwick H, Elkheir N, Lone N, Docherty AB, Harrison EM, Baille KJ, Carson G, Semple MG, Scott, Sigfrid L. What is the recovery rate and risk of long-term consequences following a diagnosis of COVID-19? - A harmonised, global longitudinal observational study (preprint). Medrxiv. 2020///; #volume#(#issue#): 2020.08.26.20180950. doi: 10.1101/2020.08.26.20180950. Available at: http://medrxiv.org/content/early/2020/09/01/2020.08.26.20180950.abstract |

| | |
|---|---|
| 127 | NA Gurdasani D, Hara ME, Hastie C, Roderick P, Yao G, Alwan, Ziauddeen N. Characteristics of Long Covid: findings from a social media survey (preprint). Medrxiv. 2021///; #volume#(#issue#): 2021.03.21.21253968. doi: 10.1101/2021.03.21.21253968. Available at: http://medrxiv.org/content/early/2021/03/27/2021.03.21.21253968.abstract |
| 128 | Tabacof L Wood J, Cortes M, Kontorovich A, McCarthy D, Rizk D, Nasr L, Breyman E, Mohammadi N, Kellner C, Putrino,D., Tosto-Mancuso J. Post-acute COVID-19 syndrome negatively impacts health and wellbeing despite less severe acute infection (preprint). Medrxiv. 2020///; #volume#(#issue#): 2020.11.04.20226126. doi: 10.1101/2020.11.04.20226126. Available at: http://medrxiv.org/content/early/2020/11/06/2020.11.04.20226126.abstract |
| 129 | U Banasik K, Pedersen OB, Nissen I, Cantwell L, Schwinn M, Tulstrup M, Westergaard D, Ullum H, Brunak S, Tommerup N, Feenstra B, Geller F, Ostrowski SR, Groenbaek K, Nielsen CH, Nielsen SD, Feldt-Rasmussen, Bliddal S. Acute and persistent symptoms in non-hospitalized PCR-confirmed COVID-19 patients (preprint). Medrxiv. 2021///; #volume#(#issue#): 2021.01.22.21249945. doi: 10.1101/2021.01.22.21249945. Available at: http://medrxiv.org/content/early/2021/01/25/2021.01.22.21249945.abstract |
| 130 | V Venkatakrishnan AJ, Ramudu E, Kirkup C, Puranik A, Kayal N, Berner G, Anand A, Barve R, O'Horo JC, Badley AD, Soundararajan, Pawlowski C. Pre-existing conditions are associated with long-COVID patients' hospitalization, despite confirmed clearance of SARS-CoV-2 virus (preprint). Medrxiv. 2020///; #volume#(#issue#): 2020.10.28.20221655. doi: 10.1101/2020.10.28.20221655. Available at: http://medrxiv.org/content/early/2020/10/30/2020.10.28.20221655.abstract |
| 131 | Chun, Hyung J, Coutavas, Elias, Pine, Alexander, Lee, Alfred I, Yu, Vanessa, Shallow, Marcus, Giovacchini, Coral X, Mathews, Anne, Stephenson, Brian, Que, Loretta G, Lee, Patty J, Kraft, Bryan D. Immuno-fibrotic drivers of impaired lung function in post-COVID-19 syndrome. medRxiv : the preprint server for health sciences. 2021///; #volume#(#issue#): #pages#. doi: https://dx.doi.org/10.1101/2021.01.31.21250870. Available at: http://ovidsp.ovid.com/ovidweb.cgi?T=JS&PAGE=reference&D=pmnm&NEWS=N&AN=33564789 |
| 132 | Huang, Yong, Pinto, Melissa D, Borelli, Jessica L, Mehrabadi, Milad Asgari, Abrihim, Heather, Dutt, Nikil, Lambert, Natalie, Nurmi, Erika L, Chakraborty, Rana, Rahmani, Amir M, Downs, Charles A. COVID Symptoms, Symptom Clusters, and Predictors for Becoming a Long-Hauler: Looking for Clarity in the Haze of the Pandemic. medRxiv : the preprint server for health sciences. 2021///; #volume#(#issue#): #pages#. doi: https://dx.doi.org/10.1101/2021.03.03.21252086. Available at: http://ovidsp.ovid.com/ovidweb.cgi?T=JS&PAGE=reference&D=pmnm&NEWS=N&AN=33688670 |
| 133 | Peluso, Michael J, Deitchman, Amelia N, Torres, Leonel, Iyer, Nikita S, Nixon, Christopher C, Munter, Sadie E, Donatelli, Joanna, Thanh, Cassandra, Takahashi, Saki, Hakim, Jill, Turcios, Keirstinne, Janson, Owen, Hoh, Rebecca, Tai, Viva, Hernandez, Yanel, Fehrman, Emily, Spinelli, Matthew A, Gandhi, Monica, Trinh, Lan, Wrin, Terri, Petropoulos, Christos J, Aweeka, Francesca T, Rodriguez-Barraquer, Isabel, Kelly, J D, Martin, Jeffrey N, Deeks, Steven G, Greenhouse, Bryan, Rutishauser, Rachel L, Henrich, Timothy J. Long-Term SARS-CoV-2-Specific Immune and Inflammatory Responses Across a Clinically Diverse Cohort of Individuals Recovering from COVID-19. medRxiv : the preprint server for health sciences. 2021///; #volume#(#issue#): #pages#. doi: https://dx.doi.org/10.1101/2021.02.26.21252308. Available at: http://ovidsp.ovid.com/ovidweb.cgi?T=JS&PAGE=reference&D=pmnm&NEWS=N&AN=33688685 |
| 134 | Peluso, Michael J, Kelly, J D, Lu, Scott, Goldberg, Sarah A, Davidson, Michelle C, Mathur, Sujata, Durstenfeld, Matthew S, Spinelli, Matthew A, Hoh, Rebecca, Tai, Viva, Fehrman, Emily A, Torres, Leonel, Hernandez, Yanel, Williams, Meghann C, Arreguin, Mireya I, Bautista, Jennifer A, Ngo, Lynn H, Deswal, Monika, Munter, Sadie E, Martinez, Enrique O, Anglin, Khamal A, Romero, Mariela D, Tavs, Jacqueline, Rugart, Paulina R, Chen, Jessica Y, Sans, Hannah M, Murray, Victoria W, Ellis, Payton K, Donohue, Kevin C, Massachi, Jonathan A, Weiss, Jacob O, Mehdi, Irum, Pineda-Ramirez, Jesus, Tang, Alex F, Wenger, Megan, Assenzio, Melissa, Yuan, Yan, Krone, Melissa, Rutishauser, Rachel L, Rodriguez-Barraquer, Isabel, Greenhouse, Bryan, Sauceda, John A, Gandhi, Monica, Hsue, Priscilla Y, Henrich, Timothy J, Deeks, Steven G, Martin, Jeffrey N. Rapid implementation of a cohort for the study of post-acute sequelae of SARS-CoV-2 infection/COVID-19. medRxiv : the preprint server for health sciences. 2021///; #volume#(#issue#): |

| | |
|---|---|
| | #pages#. doi: https://dx.doi.org/10.1101/2021.03.11.21252311. Available at: http://ovidsp.ovid.com/ovidweb.cgi?T=JS&PAGE=reference&D=pmnm&NEWS=N&AN=33758895 |
| 135 | Perlis, Roy H, Green, Jon, Santillana, Mauricio, Lazer, David, Ognyanova, Katherine, Simonson, Matthew, Baum, Matthew A, Quintana, Alexi, Chwe, Hanyu, Druckman, James, Volpe, John Della, Lin, Jennifer. Persistence of symptoms up to 10 months following acute COVID-19 illness. medRxiv : the preprint server for health sciences. 2021///; #volume#(#issue#): #pages#. doi: https://dx.doi.org/10.1101/2021.03.07.21253072. Available at: http://ovidsp.ovid.com/ovidweb.cgi?T=JS&PAGE=reference&D=pmnm&NEWS=N&AN=33758896 |
| 136 | Perlis, Roy H, Santillana, Mauricio, Ognyanova, Katherine, Green, Jon, Druckman, James N, Lazer, David, Baum, Matthew A. Comparison of post-COVID depression and major depressive disorder. medRxiv : the preprint server for health sciences. 2021///; #volume#(#issue#): #pages#. doi: https://dx.doi.org/10.1101/2021.03.26.21254425. Available at: http://ovidsp.ovid.com/ovidweb.cgi?T=JS&PAGE=reference&D=pmnm&NEWS=N&AN=33821286 |
| 137 | Bougakov, Dmitri, Podell, Kenneth, Goldberg, Elkhonon. Multiple Neuroinvasive Pathways in COVID-19. Molecular neurobiology. 2021///; 58(2): 564. doi: https://dx.doi.org/10.1007/s12035-020-02152-5. Available at: http://ovidsp.ovid.com/ovidweb.cgi?T=JS&PAGE=reference&D=medl&NEWS=N&AN=32990925 |
| 138 | Graham, Flora. Daily briefing: What the science says about long COVID. Nature. 2021///; #volume#(#issue#): #pages#. doi: http://dx.doi.org/10.1038/d41586-021-00856-9. Available at: http://ovidsp.ovid.com/ovidweb.cgi?T=JS&PAGE=reference&D=emedx&NEWS=N&AN=634693647 |
| 139 | Kandarini, Yenny, Wardani, Ni Wayan Sri, Mahadita, Gede Wira, Widiana, I G, Ayu, Nyoman Paramita, Somia, I K. Profile of kidney disease patients in covid-19 isolation wards denpasar-baliindonesia. Nephrology. 2020///; 25(#issue#): 8. doi: http://dx.doi.org/10.1111/nep.13771. Available at: http://ovidsp.ovid.com/ovidweb.cgi?T=JS&PAGE=reference&D=emexb&NEWS=N&AN=633928332 |
| 140 | Gusev, E I, Martynov, M Y, Boyko, A N, Voznyuk, I A, Latsh, N Y, Sivertseva, S A, Spirin, N N, Shamalov, N A. The novel coronavirus infection (COVID-19) and nervous system involvement: Mechanisms of neurological disorders, clinical manifestations, and the organization of neurological care. Neuroscience and behavioral physiology. 2021///; 51(2): 147. doi: http://dx.doi.org/10.1007/s11055-021-01050-0; http://dx.doi.org/10.1007/s11055-021-01050-0. Available at: http://ovidsp.ovid.com/ovidweb.cgi?T=JS&PAGE=reference&D=psyc17&NEWS=N&AN=2021-21658-003 |
| 141 | Thomson, Helen. Children with long covid. New scientist (1971). 2021///; 249(3323): 10. doi: https://dx.doi.org/10.1016/S0262-4079(21)00303-1. Available at: http://ovidsp.ovid.com/ovidweb.cgi?T=JS&PAGE=reference&D=pmnm&NEWS=N&AN=33686318 |
| 142 | Wilson, Clare. Vaccines may help clear up long-term covid-19 symptoms. New scientist (1971). 2021///; 249(3325): 9. doi: https://dx.doi.org/10.1016/S0262-4079(21)00396-1. Available at: http://ovidsp.ovid.com/ovidweb.cgi?T=JS&PAGE=reference&D=pmnm&NEWS=N&AN=33746325 |
| 143 | Hayes, James P. Considering the long-term respiratory effects of Covid-19. Occupational medicine (Oxford, England). 2021///; #volume#(#issue#): #pages#. doi: http://dx.doi.org/10.1093/occmed/kqaa224. Available at: http://ovidsp.ovid.com/ovidweb.cgi?T=JS&PAGE=reference&D=emexb&NEWS=N&AN=634090894 |
| 144 | Rayner, Clare, Campbell, Richard. Long Covid Implications for the workplace. Occupational medicine (Oxford, England). 2021///; #volume#(#issue#): #pages#. doi: https://dx.doi.org/10.1093/occmed/kqab042. Available at: http://ovidsp.ovid.com/ovidweb.cgi?T=JS&PAGE=reference&D=medp&NEWS=N&AN=33822179 |
| 145 | Al-Jahdhami, Issa, Al-Naamani, Khalid, Al-Mawali, Adhra. The post-acute COVID-19 syndrome (Long COVID). Oman Medical Journal. 2021///; 36(1): 1. doi: http://dx.doi.org/10.5001/omj.2021.91. Available at: http://www.omjournal.org/articleDetails.aspx?coType=2&aId=2837,http://ovidsp.ovid.com/ovidweb.cgi?T=JS&PAGE=reference&D=emexb&NEWS=N&AN=2005922047 |
| 146 | Suarez-Robles, Miguel, Iguaran-Bermudez, Maria, Garcia-Klepizg, Jose, Mendez-Bailon, Manuel, Lorenzo-Villalba, Noel. Ninety days post-hospitalization evaluation of residual covid-19 symptoms through a |

| | |
|---|---|
| | phone call check list. Pan African Medical Journal. 2020///; 37(#issue#): 1. doi: http://dx.doi.org/10.11604/pamj.2020.37.289.27110. Available at: https://www.panafrican-med-journal.com/content/article/37/289/pdf/289.pdf,http://ovidsp.ovid.com/ovidweb.cgi?T=JS&PAGE=reference&D=emexb&NEWS=N&AN=2005940933 |
| 147 | X Huang L, Tang X, Zhang Y, Zhou, Zhang C. Pulmonary sequelae of pediatric patients after discharge for COVID-19: an observational study. Pediatric pulmonology. 2021///; #volume#(#issue#): #pages#. doi: 10.1002/ppul.25239. Available at: https://pubmed.ncbi.nlm.nih.gov/33559979 CSR - Reports results |
| 148 | NCT04725110. Direct Topical Lung T3 Treatment to Improve Outcome & Sequelae of COVID-19 Acute Respiratory Distress Syndrome. Phase II Trial of Direct Topical Lung T3 Treatment to Improve Outcome & Sequelae of COVID-19 ARDS - A Multi-Site, Randomized, Double-blinded, Placebo-Controlled Clinical Trial. 2021///; #volume#(#issue#): #pages#. doi: #DOI#. Available at: http://ovidsp.ovid.com/ovidweb.cgi?T=JS&PAGE=reference&D=cctr&NEWS=N&AN=CN-02218030 |
| 149 | Hameed, Farah, Palatulan, Eugene, Sood, Vandana, Gellhorn, Alfred, Jaywant, Abhishek, Said, Rami, Lau, Corinna, Layton, Aimee. Outcomes of a COVID-19 recovery program for patients hospitalized with SARS-CoV-2 infection in New York City: A prospective cohort study. PM and R. 2021///; #volume#(#issue#): #pages#. doi: http://dx.doi.org/10.1002/pmrj.12578. Available at: https://onlinelibrary.wiley.com/journal/19341563,http://ovidsp.ovid.com/ovidweb.cgi?T=JS&PAGE=reference&D=emexb&NEWS=N&AN=2010860382 |
| 150 | Verduzco-Gutierrez, Monica, Rydberg, Leslie, Sullivan, Melba Nicholson, Mukherjee, Debjani. In This for the Long Haul: Ethics, COVID-19, and Rehabilitation. PM and R. 2021///; 13(3): 325. doi: http://dx.doi.org/10.1002/pmrj.12554. Available at: https://onlinelibrary.wiley.com/journal/19341563,http://ovidsp.ovid.com/ovidweb.cgi?T=JS&PAGE=reference&D=emexb&NEWS=N&AN=2010537789 |
| 151 | Bendayan, M, Boitrelle, F. What could cause the long-term effects of COVID-19 on sperm parameters and male fertility?. QJM : monthly journal of the Association of Physicians. 2021///; #volume#(#issue#): #pages#. doi: http://dx.doi.org/10.1093/qjmed/hcab028. Available at: http://ovidsp.ovid.com/ovidweb.cgi?T=JS&PAGE=reference&D=emexb&NEWS=N&AN=634245916 |
| 152 | Sher, Leo. Post-COVID syndrome and suicide risk. QJM : monthly journal of the Association of Physicians. 2021///; #volume#(#issue#): #pages#. doi: http://dx.doi.org/10.1093/qjmed/hcab007. Available at: http://ovidsp.ovid.com/ovidweb.cgi?T=JS&PAGE=reference&D=emexb&NEWS=N&AN=634102808 |
| 153 | Abe, Toshikazu. Timing of quarantine discontinuation of a COVID-19 long-hauler. QJM: An International Journal of Medicine. 2021/03//; #volume#(#issue#): #pages#. doi: 10.1093/qjmed/hcab050. Available at: http://ovidsp.ovid.com/ovidweb.cgi?T=JS&PAGE=reference&D=emexb&NEWS=N&AN=634550804,https://academic.oup.com/qjmed/advance-article/doi/10.1093/qjmed/hcab050/6166660 |
| 154 | NCT04797871. Resistance Training and Clinical Status in Patients With Post Discharge Symptoms After Covid-19. Resistance Training Intervention on the Clinical Status in Patients With Post Discharge Symptoms After Covid-19: the "EXER-COVID Study". 2021///; #volume#(#issue#): #pages#. doi: #DOI#. Available at: http://ovidsp.ovid.com/ovidweb.cgi?T=JS&PAGE=reference&D=cctr&NEWS=N&AN=CN-02237211 |
| 155 | Altmann, Daniel M. Children and the return to school: How much should we worry about covid-19 and long covid?. The BMJ. 2021///; 372(#issue#): n701. doi: http://dx.doi.org/10.1136/bmj.n701. Available at: http://www.bmj.com/,http://ovidsp.ovid.com/ovidweb.cgi?T=JS&PAGE=reference&D=emexb&NEWS=N&AN=634494719 |
| 156 | Altmann, Daniel M, Boyton, Rosemary J. Decoding the unknowns in long covid. The BMJ. 2021///; 372(#issue#): 372. doi: http://dx.doi.org/10.1136/bmj.n132. Available at: http://www.bmj.com/,http://ovidsp.ovid.com/ovidweb.cgi?T=JS&PAGE=reference&D=emexb&NEWS=N&AN=634109111 |
| 157 | Shah, Waqaar, Hillman, Toby, Playford, E D, Hishmeh, Lyth. Managing the long term effects of covid-19: Summary of NICE, SIGN, and RCGP rapid guideline. The BMJ. 2021///; 372(#issue#): n136. doi: http://dx.doi.org/10.1136/bmj.n136. Available at: |

| | |
|---|---|
| | http://www.bmj.com/,http://ovidsp.ovid.com/ovidweb.cgi?T=JS&PAGE=reference&D=emexb&NEWS=N&AN=633979511 |
| 158 | McMahon, Devon E, Desai, Seemal R, Fox, Lindy P, Gallman, Antonia E, Cyster, Jason G, Fassett, Marlys S, Hruza, George J, Rosenbach, Misha, Lipoff, Jules B, French, Lars E, Lim, Henry, Freeman, Esther E. Long COVID in the skin: a registry analysis of COVID-19 dermatological duration. The Lancet Infectious Diseases. 2021///; 21(3): 313. doi: http://dx.doi.org/10.1016/S1473-3099%2820%2930986-5. Available at: http://www.journals.elsevier.com/the-lancet-infectious-diseases,http://ovidsp.ovid.com/ovidweb.cgi?T=JS&PAGE=reference&D=emexb&NEWS=N&AN=2010850002 |
| 159 | Weng J Li J, Shen L, Zhu L, Liang Y, Lin X, Jiao N, Cheng S, Huang Y, Zou Y, Yan G, Zhu R, Lan,P., Li Y. Gastrointestinal sequelae 90 days after discharge for COVID-19; 7943402 CSR - Reports results. The lancet.Gastroenterology & hepatology. 2021///; #volume#(#issue#): #pages#. doi: 10.1016/S2468-1253(21)00076-5. Available at: https://pubmed.ncbi.nlm.nih.gov/33711290 |
| 160 | Kirby, Tony. Young person with long COVID after mild disease. The Lancet.Respiratory medicine. 2021///; #volume#(#issue#): #pages#. doi: http://dx.doi.org/10.1016/S2213-2600%2821%2900123-5. Available at: http://ovidsp.ovid.com/ovidweb.cgi?T=JS&PAGE=reference&D=emexb&NEWS=N&AN=634502667 |
| 161 | Kirby, Tony. COVID-19 survivor experiencing long-term symptoms. The Lancet.Respiratory medicine. 2021///; #volume#(#issue#): #pages#. doi: http://dx.doi.org/10.1016/S2213-2600%2821%2900092-8. Available at: http://ovidsp.ovid.com/ovidweb.cgi?T=JS&PAGE=reference&D=emexb&NEWS=N&AN=634292081 |
| 162 | Cheung, C M, Manoharan, B, Shafiq, M, Li, K Y, Anderson, S, Capuano, E, Farrugia, A, Peacock, J, Wadsworth, K, Yung, B. Early results of radiological follow-up of non-ITU inpatients with COVID-19 pneumonia in a large UK district general hospital. Thorax. 2021///; 76(#issue#): A185. doi: http://dx.doi.org/10.1136/thorax-2020-BTSabstracts.321. Available at: http://ovidsp.ovid.com/ovidweb.cgi?T=JS&PAGE=reference&D=emexb&NEWS=N&AN=634543314 |
| 163 | Cox, E, Abed El Khaleq, Y, Storrar, W, Bond, L. Chest radiograph features of the COVID-19 infection: Comparison of the initial and follow-up changes. Thorax. 2021///; 76(#issue#): A90. doi: http://dx.doi.org/10.1136/thorax-2020-BTSabstracts.155. Available at: http://ovidsp.ovid.com/ovidweb.cgi?T=JS&PAGE=reference&D=emexb&NEWS=N&AN=634543031 |
| 164 | Creamer, A W, Alaee, S, Iftikhar, H, Ahmed, F, Steer, H, Sharp, C. Clinico-radiological recovery following severe COVID-19 pneumonia. Thorax. 2021///; 76(#issue#): A185. doi: http://dx.doi.org/10.1136/thorax-2020-BTSabstracts.320. Available at: http://ovidsp.ovid.com/ovidweb.cgi?T=JS&PAGE=reference&D=emexb&NEWS=N&AN=634543291 |
| 165 | Daynes, E, Gerlis, C, Briggs-Price, S, Singh, S J, Jones, P W. Using the copd assessment test as a tool to assess symptoms in covid recovery. Thorax. 2021///; 76(#issue#): A181. doi: http://dx.doi.org/10.1136/thorax-2020-BTSabstracts.315. Available at: http://ovidsp.ovid.com/ovidweb.cgi?T=JS&PAGE=reference&D=emexb&NEWS=N&AN=634543219 |
| 166 | D'Cruz, R F, Patel, A, Perrin, F, Waller, M, Periselneris, J, Byrne, A, Mathew, S, Choudhury, M, Smith, L -J, Madula, R, Patrick, T, Walder, D, Lee, K, McNulty, W, Macedo, P, Heitmann, A, Lyall, R, Warwick, G, Birring, S, Patel, I, Jolley, C J, Norton, S, Galloway, J. Clinical, radiological, functional and psychological characteristics of severe COVID-19 pneumonia survivors: A prospective observational cohort study. Thorax. 2021///; 76(#issue#): A34. doi: http://dx.doi.org/10.1136/thorax-2020-BTSabstracts.60. Available at: http://ovidsp.ovid.com/ovidweb.cgi?T=JS&PAGE=reference&D=emexb&NEWS=N&AN=634543114 |
| 167 | Elder, H, Prasad, A, Burns, G P. Experiences from post COVID-19 clinic in a tertiary centre. Thorax. 2021///; 76(#issue#): A186. doi: http://dx.doi.org/10.1136/thorax-2020-BTSabstracts.322. Available at: http://ovidsp.ovid.com/ovidweb.cgi?T=JS&PAGE=reference&D=emexb&NEWS=N&AN=634543331 |
| 168 | Jain, N G, Saigal, A, Naidu, S B, Shah, A J, Brill, S E, Jarvis, H, Barnett, J, Hare, S, Hurst, J R, Lipman, M, Mandal, S. Do persistent chest radiograph changes correlate with ongoing respiratory symptoms in patients recovering from covid-19 pneumonia?. Thorax. 2021///; 76(#issue#): A89. doi: http://dx.doi.org/10.1136/thorax-2020-BTSabstracts.154. Available at: http://ovidsp.ovid.com/ovidweb.cgi?T=JS&PAGE=reference&D=emexb&NEWS=N&AN=634543014 |

| | |
|---|---|
| 169 | Owles, H L B, Padmanaban, V, Thacker, M, Hussein, I, Mallia, P, Russell, G, Kon, O M, Elkin, S L. Feasibility and usage of one minute sit-to-stand test, as a measure of recovery in post-acute covid 19 patients, following hospital discharge. Thorax. 2021///; 76(#issue#): A35. doi: http://dx.doi.org/10.1136/thorax-2020-BTSabstracts.61. Available at: http://ovidsp.ovid.com/ovidweb.cgi?T=JS&PAGE=reference&D=emexb&NEWS=N&AN=634543138 |
| 170 | Patel, G, Dadey, E, Gosal, E, O'Neill, T, Skyllberg, E W, Calderwood, C J, Cheng, D O, Ainley, A. Clinical characteristics, mortality and short term follow up of patients admitted with covid-19 in a North East London NHS Trust: A retrospective analysis. Thorax. 2021///; 76(#issue#): A5. doi: http://dx.doi.org/10.1136/thorax-2020-BTSabstracts.10. Available at: http://ovidsp.ovid.com/ovidweb.cgi?T=JS&PAGE=reference&D=emexb&NEWS=N&AN=634543156 |
| 171 | Pearmain, L, Avram, C, Yioe, V, Webb, P, Margaritopoulos, G A, Rivera-Ortega, P, Chaudhuri, N, Blaikley, J F. Patient symptoms following discharge from hospital after COVID-19 pneumonia. Thorax. 2021///; 76(#issue#): A180. doi: http://dx.doi.org/10.1136/thorax-2020-BTSabstracts.313. Available at: http://ovidsp.ovid.com/ovidweb.cgi?T=JS&PAGE=reference&D=emexb&NEWS=N&AN=634543185 |
| 172 | Saigal, A, Naidu, S B, Shah, A J, Brill, S E, Jarvis, H, Goldring, J G, Hurst, J R, Lipman, M, Mandal, S. 'Long-COVID': The need for multi-disciplinary working. Thorax. 2021///; 76(#issue#): A33. doi: http://dx.doi.org/10.1136/thorax-2020-BTSabstracts.59. Available at: http://ovidsp.ovid.com/ovidweb.cgi?T=JS&PAGE=reference&D=emexb&NEWS=N&AN=634543096 |
| 173 | Saigal, A, Shah, A J, Naidu, S B, Brown, J, Goldring, J G, Sood, T, Lipman, M, Hurst, J R, Mandal, S. Ward vs. Emergency department discharge in patients with COVID-19: Does it make a difference to symptom burden and radiological severity at follow up?. Thorax. 2021///; 76(#issue#): A182. doi: http://dx.doi.org/10.1136/thorax-2020-BTSabstracts.316. Available at: http://ovidsp.ovid.com/ovidweb.cgi?T=JS&PAGE=reference&D=emexb&NEWS=N&AN=634543236 |
| 174 | Wendel, Silvano, Fontao-Wendel, Rita, Fachini, Roberta, Candelaria, Gabriela, Scuracchio, Patricia, Achkar, Ruth, Machado, Rafael Rahal Guaragna, Araujo, Danielle, Soares, Camila Pereira, Durigon, Edison. Kinetics of neutralizing antibodies (nAb) SARS-CoV-2 and anti-nucleocapsid (NP) antibodies in convalescent plasma donors. Transfusion. 2020///; 60(#issue#): 290A. doi: http://dx.doi.org/10.1111/trf.16084. Available at: http://ovidsp.ovid.com/ovidweb.cgi?T=JS&PAGE=reference&D=emexb&NEWS=N&AN=633926924 |
| 175 | Hawkins, Robert B, Mehaffey, J H, Charles, Eric J, Mannem, Hannah C, Roeser, Mark. Lung Transplantation for Severe Post-Coronavirus Disease 2019 Respiratory Failure. Transplantation. 2021///; #volume#(#issue#): #pages#. doi: http://dx.doi.org/10.1097/TP.0000000000003706. Available at: http://ovidsp.ovid.com/ovidweb.cgi?T=JS&PAGE=reference&D=emexb&NEWS=N&AN=634349048 |
| 176 | Vos, Robin, Ceulemans, Laurens J. Bracing for the Next Wave on the Long Haul: Lung Transplantation for Post-COVID-19 Respiratory Failure. Transplantation. 2021///; #volume#(#issue#): #pages#. doi: http://dx.doi.org/10.1097/TP.0000000000003707. Available at: http://ovidsp.ovid.com/ovidweb.cgi?T=JS&PAGE=reference&D=emexb&NEWS=N&AN=634348027 |

**No outcomes of interest = 85**

| | |
|---|---|
| 1 | Schwarzkopf S., Krawczyk A., Knop D., Klump H., Heinold A., Heinemann F.M., Thummler L., Temme C., Breyer M., Witzke O., Dittmer U., Lenz V., Horn P.A., Lindemann M.. Cellular immunity in COVID-19 convalescents with PCR-confirmed infection but with undetectable SARS-CoV-2-Specific IgG. Emerging Infectious Diseases. 2021; 27(1): 122. doi: http://dx.doi.org/10.3201/eid2701.203772. Available at: http://wwwnc.cdc.gov//eid/article/27/1/20-3772_article; http://ovidsp.ovid.com/ovidweb.cgi?T=JS&PAGE=reference&D=emedx&NEWS=N&AN=2010520286 |
| 2 | Carsetti R., Zaffina S., Piano Mortari E., Terreri S., Corrente F., Capponi C., Palomba P., Mirabella M., Cascioli S., Palange P., Cuccaro I., Milito C., Zumla A., Maeurer M., Camisa V., Vinci M.R., Santoro A., Cimini E., Marchioni L., Nicastri E., Palmieri F., Agrati C., Ippolito G., Porzio O., Concato C., Onetti Muda |

| | |
|---|---|
| | A., Raponi M., Quintarelli C., Quinti I., Locatelli F.. Different Innate and Adaptive Immune Responses to SARS-CoV-2 Infection of Asymptomatic, Mild, and Severe Cases. Frontiers in Immunology. 2020; 11(#issue#): 610300. doi: http://dx.doi.org/10.3389/fimmu.2020.610300. Available at: https://www.frontiersin.org/journals/immunology#; http://ovidsp.ovid.com/ovidweb.cgi?T=JS&PAGE=reference&D=emedx&NEWS=N&AN=633772896 |
| 3 | Sancho J., Ferrer S., Lahosa C., Posadas T., Bures E., Banuls P., Fernandez-Presa L., Royo P., Blasco M.L., Signes-Costa J.. Tracheostomy in patients with COVID-19: predictors and clinical features. European Archives of Oto-Rhino-Laryngology. 2021; #volume#(#issue#): #pages#. doi: http://dx.doi.org/10.1007/s00405-020-06555-x. Available at: http://link.springer.de/link/service/journals/00405/index.htm; http://ovidsp.ovid.com/ovidweb.cgi?T=JS&PAGE=reference&D=emedx&NEWS=N&AN=2007708987 |
| 4 | Lal B.K., Prasad N.K., Englum B.R., Turner D.J., Siddiqui T., Carlin M.M., Lake R., Sorkin J.D.. Periprocedural complications in patients with SARS-CoV-2 infection compared to those without infection: A nationwide propensity-matched analysis. American Journal of Surgery. 2020; #volume#(#issue#): #pages#. doi: http://dx.doi.org/10.1016/j.amjsurg.2020.12.024. Available at: http://www.elsevier.com/locate/amjsurg; http://ovidsp.ovid.com/ovidweb.cgi?T=JS&PAGE=reference&D=emedx&NEWS=N&AN=2010511337 |
| 5 | Glasbey J.C., Nepogodiev D., Simoes J.F.F., Omar O., Li E., Venn M.L., Abou Chaar M.K., Capizzi V., Chaudhry D., Desai A., Edwards J.G., Evans J.P., Fiore M., Videria J.F., Ford S.J., Ganly I., Griffiths E.A., Gujjuri R.R., Kolias A.G., Kaafarani H.M.A., Minaya-Bravo A., McKay S.C., Mohan H.M., Roberts K.J., Miguel-Mendez C.S., Pockney P., Shaw R., Smart N.J., Stewart G.D., Sundar S., Vidya R., Bhangu A.A.. Elective cancer surgery in COVID-19-Free surgical pathways during the SARS-cov-2 pandemic: An international, multicenter, comparative cohort study. Journal of Clinical Oncology. 2021; 39(1): 66. doi: http://dx.doi.org/10.1200/JCO.20.01933. Available at: https://ascopubs.org/doi/pdf/10.1200/JCO.20.01933; http://ovidsp.ovid.com/ovidweb.cgi?T=JS&PAGE=reference&D=emedx&NEWS=N&AN=2010489767 |
| 6 | Zhang J., Wu Q., Liu Z., Wang Q., Wu J., Hu Y., Bai T., Xie T., Huang M., Wu T., Peng D., Huang W., Jin K., Niu L., Guo W., Luo D., Lei D., Wu Z., Li G., Huang R., Lin Y., Xie X., He S., Deng Y., Liu J., Li W., Lu Z., Chen H., Zeng T., Luo Q., Li Y.-P., Wang Y., Liu W., Qu X.. Spike-specific circulating T follicular helper cell and cross-neutralizing antibody responses in COVID-19-convalescent individuals. Nature Microbiology. 2021; 6(1): 51. doi: http://dx.doi.org/10.1038/s41564-020-00824-5. Available at: http://www.nature.com/nmicrobiol/; http://ovidsp.ovid.com/ovidweb.cgi?T=JS&PAGE=reference&D=emexc&NEWS=N&AN=2007301168 |
| 7 | Paek J.H., Kim Y., Park W.Y., Jin K., Hyun M., Lee J.Y., Kim H.A., Kwon Y.S., Park J.S., Han S.. Severe acute kidney injury in COVID-19 patients is associated with in-hospital mortality. PLoS ONE. 2020; 15(12): e0243528. doi: http://dx.doi.org/10.1371/journal.pone.0243528. Available at: https://journals.plos.org/plosone/article/file?id=10.1371/journal.pone.0243528&type=printable; http://ovidsp.ovid.com/ovidweb.cgi?T=JS&PAGE=reference&D=emexc&NEWS=N&AN=2010390987 |
| 8 | Jackel M., Rilinger J., Lang C.N., Zotzmann V., Kaier K., Stachon P., Biever P.M., Wengenmayer T., Duerschmied D., Bode C., Staudacher D.L., Supady A.. Outcome of acute respiratory distress syndrome requiring extracorporeal membrane oxygenation in Covid-19 or influenza: A single-center registry study. Artificial Organs. 2020; #volume#(#issue#): #pages#. doi: http://dx.doi.org/10.1111/aor.13865. Available at: http://onlinelibrary.wiley.com/journal/10.1111/(ISSN)1525-1594; http://ovidsp.ovid.com/ovidweb.cgi?T=JS&PAGE=reference&D=emexc&NEWS=N&AN=2007627000 |
| 9 | Brunetta E., Folci M., Bottazzi B., De Santis M., Gritti G., Protti A., Mapelli S.N., Bonovas S., Piovani D., Leone R., My I., Zanon V., Spata G., Bacci M., Supino D., Carnevale S., Sironi M., Davoudian S., Peano C., Landi F., Di Marco F., Raimondi F., Gianatti A., Angelini C., Rambaldi A., Garlanda C., Ciccarelli M., Cecconi M., Mantovani A.. Macrophage expression and prognostic significance of the long pentraxin PTX3 in COVID-19. Nature immunology. 2021; 22(1): 19. doi: http://dx.doi.org/10.1038/s41590-020-00832-x. Available at: http://www.nature.com/ni; http://ovidsp.ovid.com/ovidweb.cgi?T=JS&PAGE=reference&D=emexc&NEWS=N&AN=2007324547 |

| | |
|---|---|
| 10 | Rutten J.J.S., van Loon A.M., van Kooten J., van Buul L.W., Joling K.J., Smalbrugge M., Hertogh C.M.P.M.. Clinical Suspicion of COVID-19 in Nursing Home Residents: Symptoms and Mortality Risk Factors. Journal of the American Medical Directors Association. 2020; 21(12): 1791. doi: http://dx.doi.org/10.1016/j.jamda.2020.10.034. Available at: https://www.journals.elsevier.com/journal-of-the-american-medical-directors-association; http://ovidsp.ovid.com/ovidweb.cgi?T=JS&PAGE=reference&D=emexc&NEWS=N&AN=2010121682 |
| 11 | Kratzer B., Trapin D., Ettel P., Kormoczi U., Rottal A., Tuppy F., Feichter M., Gattinger P., Borochova K., Dorofeeva Y., Tulaeva I., Weber M., Grabmeier-Pfistershammer K., Tauber P.A., Gerdov M., Muhl B., Perkmann T., Fae I., Wenda S., Fuhrer H., Henning R., Valenta R., Pickl W.F.. Immunological imprint of COVID-19 on human peripheral blood leukocyte populations. Allergy: European Journal of Allergy and Clinical Immunology. 2020; #volume#(#issue#): #pages#. doi: http://dx.doi.org/10.1111/all.14647. Available at: http://onlinelibrary.wiley.com/journal/10.1111/(ISSN)1398-9995; http://ovidsp.ovid.com/ovidweb.cgi?T=JS&PAGE=reference&D=emexc&NEWS=N&AN=2007491468 |
| 12 | Bianchi A., Mazzoni L.N., Busoni S., Pinna N., Albanesi M., Cavigli E., Cozzi D., Poggesi A., Miele V., Fainardi E., Gadda D.. Assessment of cerebrovascular disease with computed tomography in COVID-19 patients: correlation of a novel specific visual score with increased mortality risk. La Radiologia medica. 2020; #volume#(#issue#): #pages#. doi: http://dx.doi.org/10.1007/s11547-020-01313-9. Available at: http://ovidsp.ovid.com/ovidweb.cgi?T=JS&PAGE=reference&D=emexc&NEWS=N&AN=633548631 |
| 13 | Rydyznski Moderbacher C., Ramirez S.I., Dan J.M., Grifoni A., Hastie K.M., Weiskopf D., Belanger S., Abbott R.K., Choi J., Kato Y., Crotty E.G., Kim C., Rawlings S.A., Mateus J., Tse L.P.V., Frazier A., Baric R., Peters B., Greenbaum J., Ollmann Saphire E., Smith D.M., Sette A., Crotty S.. Antigen-Specific Adaptive Immunity to SARS-CoV-2 in Acute COVID-19 and Associations with Age and Disease Severity. Cell. 2020; 183(4): 996. doi: http://dx.doi.org/10.1016/j.cell.2020.09.038. Available at: http://www.cell.com; http://ovidsp.ovid.com/ovidweb.cgi?T=JS&PAGE=reference&D=emexc&NEWS=N&AN=2007987135 |
| 14 | Li K., Huang B., Wu M., Zhong A., Li L., Cai Y., Wu L., Zhu M., Li J., Wang Z., Wu W., Li W., Bosco B., Gan Z., Qiao Q., Wu J., Wang Q., Wang S., Xia X.. Dynamic changes in anti-SARS-CoV-2 antibodies during SARS-CoV-2 infection and recovery from COVID-19. Nature Communications. 2020; 11(1): 6044. doi: http://dx.doi.org/10.1038/s41467-020-19943-y. Available at: http://www.nature.com/ncomms/index.html; http://ovidsp.ovid.com/ovidweb.cgi?T=JS&PAGE=reference&D=emexc&NEWS=N&AN=2007425376 |
| 15 | Tehrani S., Killander A., Astrand P., Jakobsson J., Gille-Johnson P.. Risk factors for death in adult COVID-19 patients: Frailty predicts fatal outcome in older patients. International Journal of Infectious Diseases. 2021; 102(#issue#): 415. doi: http://dx.doi.org/10.1016/j.ijid.2020.10.071. Available at: https://www.journals.elsevier.com/international-journal-of-infectious-diseases; http://ovidsp.ovid.com/ovidweb.cgi?T=JS&PAGE=reference&D=emexc&NEWS=N&AN=2010138004 |
| 16 | Dobi A., Seteyen A.-L.S., Rakoto M.L., Lebeau G., Vagner D., Frumence E., Giry C., Septembre-Malaterre A., Raffray L., Gasque P.. Serological surveillance of COVID-19 hospitalized patients in Reunion Island (France) revealed that specific immunoglobulin G are rapidly vanishing in severe cases. Journal of Clinical Medicine. 2020; 9(12): 1. doi: http://dx.doi.org/10.3390/jcm9123847. Available at: https://www.mdpi.com/2077-0383/9/12/3847/pdf; http://ovidsp.ovid.com/ovidweb.cgi?T=JS&PAGE=reference&D=emexc&NEWS=N&AN=2005501436 |
| 17 | Liu C., Yu X., Gao C., Zhang L., Zhai H., Hu Y., Liu E., Wang Q., Gao Y., Wei D., Zhang D., Han Y., Zhang X.. Characterization of antibody responses to SARS-CoV-2 in convalescent COVID-19 patients. Journal of medical virology. 2020; #volume#(#issue#): #pages#. doi: http://dx.doi.org/10.1002/jmv.26646. Available at: http://onlinelibrary.wiley.com/journal/10.1002/(ISSN)1096-9071; http://ovidsp.ovid.com/ovidweb.cgi?T=JS&PAGE=reference&D=emexc&NEWS=N&AN=2007413338 |
| 18 | Seow J., Graham C., Merrick B., Acors S., Pickering S., Steel K.J.A., Hemmings O., O'Byrne A., Kouphou N., Galao R.P., Betancor G., Wilson H.D., Signell A.W., Winstone H., Kerridge C., Huettner I., Jimenez-Guardeno J.M., Lista M.J., Temperton N., Snell L.B., Bisnauthsing K., Moore A., Green A., Martinez L., Stokes B., Honey J., Izquierdo-Barras A., Arbane G., Patel A., Tan M.K.I., O'Connell L., O'Hara G., MacMahon E., Douthwaite S., Nebbia G., Batra R., Martinez-Nunez R., Shankar-Hari M., Edgeworth J.D., |

| | |
|---|---|
| | Neil S.J.D., Malim M.H., Doores K.J.. Longitudinal observation and decline of neutralizing antibody responses in the three months following SARS-CoV-2 infection in humans. Nature Microbiology. 2020; 5(12): 1598. doi: http://dx.doi.org/10.1038/s41564-020-00813-8. Available at: http://www.nature.com/nmicrobiol/; http://ovidsp.ovid.com/ovidweb.cgi?T=JS&PAGE=reference&D=emexc&NEWS=N&AN=2007071211 |
| 19 | Lu Y., Li Y., Wang Y., Luo J., Yu W.. Nucleic acid and antibody assay results in Chinese patients with coronavirus disease 2019 (COVID-19). International immunopharmacology. 2020; 89(#issue#): 107089. doi: http://dx.doi.org/10.1016/j.intimp.2020.107089. Available at: http://www.elsevier.com/locate/intimp; http://ovidsp.ovid.com/ovidweb.cgi?T=JS&PAGE=reference&D=emexc&NEWS=N&AN=2008043275 |
| 20 | Neumann-Podczaska A., Chojnicki M., Karbowski L.M., Al-Saad S.R., Hashmi A.A., Chudek J., Tobis S., Kropinska S., Mozer-Lisewska I., Suwalska A., Tykarski A., Wieczorowska-Tobis K.. Clinical characteristics and survival analysis in a small sample of older COVID-19 patients with defined 60-day outcome. International Journal of Environmental Research and Public Health. 2020; 17(22): 1. doi: http://dx.doi.org/10.3390/ijerph17228362. Available at: https://www.mdpi.com/1660-4601/17/22/8362/pdf; http://ovidsp.ovid.com/ovidweb.cgi?T=JS&PAGE=reference&D=emexc&NEWS=N&AN=2005444855 |
| 21 | Lavery A.M., Preston L.E., Ko J.Y., Chevinsky J.R., DeSisto C.L., Pennington A.F., Kompaniyets L., Datta S.D., Click E.S., Golden T., Goodman A.B., Mac Kenzie W.R., Boehmer T.K., Gundlapalli A.V.. Characteristics of Hospitalized COVID-19 Patients Discharged and Experiencing Same-Hospital Readmission - United States, March-August 2020. MMWR.Morbidity and mortality weekly report. 2020; 69(45): 1695. doi: http://dx.doi.org/10.15585/mmwr.mm6945e2. Available at: http://ovidsp.ovid.com/ovidweb.cgi?T=JS&PAGE=reference&D=emexc&NEWS=N&AN=633410424 |
| 22 | Stevenson J.D., Evans S., Morris G., Tillman R., Abudu A., Jeys L., Parry M.. Mortality of high-risk orthopaedic oncology patients during the COVID-19 pandemic: A prospective cohort study. Journal of surgical oncology. 2020; 122(6): 1027. doi: http://dx.doi.org/10.1002/jso.26127. Available at: http://onlinelibrary.wiley.com/journal/10.1002/(ISSN)1096-9098; http://ovidsp.ovid.com/ovidweb.cgi?T=JS&PAGE=reference&D=emexc&NEWS=N&AN=2005572439 |
| 23 | Schub D., Klemis V., Schneitler S., Mihm J., Lepper P.M., Wilkens H., Bals R., Eichler H., Gartner B.C., Becker S.L., Sester U., Sester M., Schmidt T.. High levels of SARS-CoV-2-specific T cells with restricted functionality in severe courses of COVID-19. JCI Insight. 2020; 5(20): e142167. doi: http://dx.doi.org/10.1172/jci.insight.142167. Available at: https://insight.jci.org/articles/view/142167/pdf; http://ovidsp.ovid.com/ovidweb.cgi?T=JS&PAGE=reference&D=emexc&NEWS=N&AN=2008503323 |
| 24 | Pizzini A., Aichner M., Sahanic S., Bohm A., Egger A., Hoermann G., Kurz K., Widmann G., Bellmann-Weiler R., Weiss G., Tancevski I., Sonnweber T., Loffler-Ragg J.. Impact of vitamin d deficiency on covid-19-a prospective analysis from the covild registry. Nutrients. 2020; 12(9): 1. doi: http://dx.doi.org/10.3390/nu12092775. Available at: https://www.mdpi.com/2072-6643/12/9/2775/pdf; http://ovidsp.ovid.com/ovidweb.cgi?T=JS&PAGE=reference&D=emexc&NEWS=N&AN=2005060950 |
| 25 | Muecksch F., Wise H., Batchelor B., Squires M., Semple E., Richardson C., McGuire J., Clearly S., Furrie E., Greig N., Hay G., Templeton K., Lorenzi J.C.C., Hatziioannou T., Jenks S., Bieniasz P.D.. Longitudinal analysis of serology and neutralizing antibody levels in COVID19 convalescents. The Journal of infectious diseases. 2020; #volume#(#issue#): #pages#. doi: http://dx.doi.org/10.1093/infdis/jiaa659. Available at: http://ovidsp.ovid.com/ovidweb.cgi?T=JS&PAGE=reference&D=emexc&NEWS=N&AN=633331572 |
| 26 | Isho B., Abe K.T., Zuo M., Jamal A.J., Rathod B., Wang J.H., Li Z., Chao G., Rojas O.L., Bang Y.M., Pu A., Christie-Holmes N., Gervais C., Ceccarelli D., Samavarchi-Tehrani P., Guvenc F., Budylowski P., Li A., Paterson A., Yun Y.F., Marin L.M., Caldwell L., Wrana J.L., Colwill K., Sicheri F., Mubareka S., Gray-Owen S.D., Drews S.J., Siqueira W.L., Barrios-Rodiles M., Ostrowski M., Rini J.M., Durocher Y., McGeer A.J., Gommerman J.L., Gingras A.-C.. Persistence of serum and saliva antibody responses to SARS-CoV-2 spike antigens in COVID-19 patients. Science Immunology. 2020; 5(52): eabe5511. doi: http://dx.doi.org/10.1126/sciimmunol.abe5511. Available at: |

| | |
|---|---|
| | https://immunology.sciencemag.org/content/immunology/5/52/eabe5511.full.pdf; http://ovidsp.ovid.com/ovidweb.cgi?T=JS&PAGE=reference&D=emexc&NEWS=N&AN=2008369708 |
| 27 | Munoz P., Galar A., Catalan P., Valerio M., Aldamiz-Echevarria T., Colliga C., Bouza E.. The first 100 cases of covid-19 in a hospital in madrid with a 2-month follow-up. Revista Espanola de Quimioterapia. 2020; 33(5): 369. doi: http://dx.doi.org/10.37201/req/072.2020. Available at: https://seq.es/wp-content/uploads/2020/07/munoz30jul2020.pdf; http://ovidsp.ovid.com/ovidweb.cgi?T=JS&PAGE=reference&D=emexc&NEWS=N&AN=2005105356 |
| 28 | Di Nisio M., Potere N., Candeloro M., Spacone A., Pieramati L., Ferrandu G., Rizzo G., La Vella M., Di Carlo S., Cibelli D., Parruti G., Levi M., Porreca E.. Interleukin-6 receptor blockade with subcutaneous tocilizumab improves coagulation activity in patients with COVID-19. European journal of internal medicine. 2020; #volume#(#issue#): #pages#. doi: http://dx.doi.org/10.1016/j.ejim.2020.10.020. Available at: http://www.elsevier.com/locate/ejim; http://ovidsp.ovid.com/ovidweb.cgi?T=JS&PAGE=reference&D=emexc&NEWS=N&AN=2008477037 |
| 29 | Jonker, Pascal K. C., van der Plas, Willemijn,Y., Steinkamp, Pieter J., Poelstra, Ralph, Emous, Marloes, van der Meij, Wout, Thunnissen, Floris, Bierman, Wouter F. W., Struys, Michel M. R. F., de Reuver, Philip,R., de Vries, Jean-Paul,P.M., Kruijff, Schelto, Dutch Surgical COVID-19, Research Collaborative. Perioperative SARS-CoV-2 infections increase mortality, pulmonary complications, and thromboembolic events: A Dutch, multicenter, matched-cohort clinical study. Surgery. 2020; #volume#(#issue#): #pages#. doi: https://dx.doi.org/10.1016/j.surg.2020.09.022. Available at: http://ovidsp.ovid.com/ovidweb.cgi?T=JS&PAGE=reference&D=medp&NEWS=N&AN=33158548 |
| 30 | Agarwal, Vineet, Venkatakrishnan, A. J., Puranik, Arjun, Kirkup, Christian, Lopez-Marquez, Agustin, Challener, Douglas W., Theel, Elitza S., O'Horo, John,C., Binnicker, Matthew J., Kremers, Walter K., Faubion, William A.,Jr, Badley, Andrew D., Williams, Amy W., Gores, Gregory J., Halamka, John D., Morice, William G.,2nd, Soundararajan, Venky. Long-term SARS-CoV-2 RNA shedding and its temporal association to IgG seropositivity. Cell death discovery. 2020; 6(1): 138. doi: https://dx.doi.org/10.1038/s41420-020-00375-y. Available at: http://ovidsp.ovid.com/ovidweb.cgi?T=JS&PAGE=reference&D=prem&NEWS=N&AN=33298894 |
| 31 | De Luca, Maurizio, Sartori, Alberto, Vitiello, Antonio, Piatto, Giacomo, Noaro, Giulia, Olmi, Stefano, Foschi, Diego, De Re, Luca, Zappa, Marco, Sarro, Giuliano, Rivolta, Umberto, Giraudo, Giorgio, Borghi, Felice, Pozzo, Gabriele, Sorisio, Vincenzo, Pignata, Giusto, Greco, Paola Antonella, Sisti, Valerio, Campagnaro, Tommaso, Guglielmi, Alfredo, Andreuccetti, Jacopo, Di Leo, Alberto, Lauro, Enrico, Ricci, Francesco, Musella, Mario, Zizzo, Maurizio, Bonacini, Stefano, Podda, Mauro, Pisanu, Adolfo, Coletta, Pietro, Guerrieri, Mario, Caracino, Valerio, Basti, Massimo, Pilone, Vincenzo, Raffaelli, Marco, Oragano, Luigi. Complications and mortality in a cohort of patients undergoing emergency and elective surgery with perioperative SARS-CoV-2 infection: an Italian multicenter study. Teachings of Phase 1 to be brought in Phase 2 pandemic. Updates in surgery. 2021; #volume#(#issue#): #pages#. doi: https://dx.doi.org/10.1007/s13304-020-00909-0. Available at: http://ovidsp.ovid.com/ovidweb.cgi?T=JS&PAGE=reference&D=medp&NEWS=N&AN=33389672 |
| 32 | Kang, Seok Hui, Kim, Sang Won, Kim, A. Y., Cho, Kyu Hyang, Park, Jong Won, Do, Jun Young. Association between Chronic Kidney Disease or Acute Kidney Injury and Clinical Outcomes in COVID-19 Patients. Journal of Korean medical science. 2020; 35(50): e434. doi: https://dx.doi.org/10.3346/jkms.2020.35.e434. Available at: http://ovidsp.ovid.com/ovidweb.cgi?T=JS&PAGE=reference&D=mesx&NEWS=N&AN=33372426 |
| 33 | Recalde-Zamacona, B., Tomas-Velazquez, A., Campo, A., Satrustegui-Alzugaray, B., Fernandez-Alonso, M., Inigo, M., Rodriguez-Mateos, M., Di Frisco, M., Felgueroso, C., Berto, J., Marin-Oto, M., Alcaide, A. B., Zulueta, J. J., Seijo, L., Landecho, M. F.. Chronic rhinosinusitis is associated with prolonged SARS-CoV2 RNA shedding in upper respiratory tract samples: A case-control study. Journal of internal medicine. 2020; #volume#(#issue#): #pages#. doi: https://dx.doi.org/10.1111/joim.13237. Available at: http://ovidsp.ovid.com/ovidweb.cgi?T=JS&PAGE=reference&D=medp&NEWS=N&AN=33372300 |
| 34 | Zhong, Lingshan, Zhang, Shuo, Wang, Jigang, Zhao, Xinqian, Wang, Kai, Ding, Wenlong, Xing, Zhiheng, Shen, Jun. Analysis of Chest CT Results of Coronavirus Disease 2019 (COVID-19) Patients at First Follow- |

| | |
|---|---|
| | Up. Canadian respiratory journal. 2020; 2020(#issue#): 5328267. doi: https://dx.doi.org/10.1155/2020/5328267. Available at: http://ovidsp.ovid.com/ovidweb.cgi?T=JS&PAGE=reference&D=medl&NEWS=N&AN=33204376 |
| 35 | Cangiano B., Fatti L.M., Danesi L., Gazzano G., Croci M., Vitale G., Gilardini L., Bonadonna S., Chiodini I., Caparello C.F., Conti A., Persani L., Stramba-Badiale M., Bonomi M.. Mortality in an italian nursing home during COVID-19 pandemic: correlation with gender, age, ADL, vitamin D supplementation, and limitations of the diagnostic tests. Aging. 2020; 12(#issue#): #pages#. doi: http://dx.doi.org/10.18632/aging.202307. Available at: http://ovidsp.ovid.com/ovidweb.cgi?T=JS&PAGE=reference&D=emexc&NEWS=N&AN=633876745 |
| 36 | Cai G., Gao Y., Zeng S., Yu Y., Liu X., Liu D., Wang Y., Yu R., Desai A., Li C., Gao Q.. Immunological alternation in COVID-19 patients with cancer and its implications on mortality. OncoImmunology. 2021; 10(1): 1854424. doi: http://dx.doi.org/10.1080/2162402X.2020.1854424. Available at: http://www.tandfonline.com/loi/koni20; http://ovidsp.ovid.com/ovidweb.cgi?T=JS&PAGE=reference&D=emexc&NEWS=N&AN=2007784965 |
| 37 | Legros, Vincent, Denolly, Solene, Vogrig, Manon, Boson, Bertrand, Siret, Eglantine, Rigaill, Josselin, Pillet, Sylvie, Grattard, Florence, Gonzalo, Sylvie, Verhoeven, Paul, Allatif, Omran, Berthelot, Philippe, Pelissier, Carole, Thiery, Guillaume, Botelho-Nevers, Elisabeth, Millet, Guillaume, Morel, Jerome, Paul, Stephane, Walzer, Thierry, Cosset, Francois-Loic, Bourlet, Thomas, Pozzetto, Bruno. A longitudinal study of SARS-CoV-2-infected patients reveals a high correlation between neutralizing antibodies and COVID-19 severity. Cellular & molecular immunology. 2021; #volume#(#issue#): #pages#. doi: https://dx.doi.org/10.1038/s41423-020-00588-2. Available at: http://ovidsp.ovid.com/ovidweb.cgi?T=JS&PAGE=reference&D=medp&NEWS=N&AN=33408342 |
| 38 | Sasisekharan, Varun, Pentakota, Niharika, Jayaraman, Akila, Tharakaraman, Kannan, Wogan, Gerald N., Narayanasami, Uma. Orthogonal immunoassays for IgG antibodies to SARS-CoV-2 antigens reveal that immune response lasts beyond 4 mo post illness onset. Proceedings of the National Academy of Sciences of the United States of America. 2021; 118(5): #pages#. doi: https://dx.doi.org/10.1073/pnas.2021615118. Available at: http://ovidsp.ovid.com/ovidweb.cgi?T=JS&PAGE=reference&D=prem&NEWS=N&AN=33446512 |
| 39 | Gil-Rodrigo A, Llorens P, Luque-Hernandez MJ, Martinez-Buendia C, Ramos-Rincon,J.M.. Lung Ultrasound Integration in Assessment of Patients with Noncritical COVID-19. Journal of ultrasound in medicine. 2021; #volume#(#issue#): #pages#. doi: 10.1002/jum.15613. Available at: https://pubmed.ncbi.nlm.nih.gov/33426645 |
| 40 | Haase, Nicolai, Plovsing, Ronni, Christensen, Steffen, Poulsen, Lone Musaeus, Brochner, Anne Craveiro, Rasmussen, Bodil Steen, Helleberg, Marie, Jensen, Jens Ulrik Staehr, Andersen, Lars Peter Kloster, Siegel, Hanna, Ibsen, Michael, Jorgensen, Vibeke, Winding, Robert, Iversen, Susanne, Pedersen, Henrik Planck, Madsen, Jacob, Solling, Christoffer, Garcia, Ricardo Sanchez, Michelsen, Jens, Mohr, Thomas, Mannering, Anne, Espelund, Ulrick Skipper, Bundgaard, Helle, Kirkegaard, Lynge, Smitt, Margit, Buck, David Levarett, Ribergaard, Niels-Erik, Pedersen, Helle Scharling, Christensen, Birgitte Viebaek, Perner, Anders. Characteristics, interventions, and longer term outcomes of COVID-19 ICU patients in Denmark-A nationwide, observational study. Acta Anaesthesiologica Scandinavica. 2021///; 65(1): 68. doi: https://dx.doi.org/10.1111/aas.13701. Available at: http://ovidsp.ovid.com/ovidweb.cgi?T=JS&PAGE=reference&D=medl&NEWS=N&AN=32929715 |
| 41 | Giannis, Dimitrios, Allen, Steven L, Davidson, Anne, Marder, Galina S, Flint, Sarah, Garrett, David, Thakur, Richa, Snyder, Matthew, Ghiuzeli, Cristina, McGinn, Thomas G, Davidson, Karina W, Qiu, Michael, Barish, Matthew, Spyropoulos, Alex C. Thromboembolic outcomes of hospitalized COVID-19 patients in the 90-day post-discharge period: early data from the northwell core-19 registry. Blood. 2020///; 136(#issue#): 33. doi: http://dx.doi.org/10.1182/blood-2020-141901. Available at: https://ashpublications.org/blood/article/136/Supplement 1/33/470152/Thromboembolic-Outcomes-of-Hospitalized-COVID-19,http://ovidsp.ovid.com/ovidweb.cgi?T=JS&PAGE=reference&D=emexb&NEWS=N&AN=634310839 |

| | |
|---|---|
| 42 | R Tremblay T, Fournier MJ, Drouin M, Beaudoin-Bussieres G, Prevost J, Lewin A, Begin P, Finzi A, Bazin, Perreault J. Waning of SARS-CoV-2 RBD antibodies in longitudinal convalescent plasma samples within 4 months after symptom onset. Blood. 2020///; 136(22): 2588. doi: #DOI#. Available at: http://ovidsp.ovid.com/ovidweb.cgi?T=JS&PAGE=reference&D=cctr&NEWS=N&AN=CN-02208719 |
| 43 | Qian, Fang, Gao, Guiju, Song, Yangzi, Xu, Yanli, Wang, Aibin, Wang, Sa, Zhao, Tianwei, Chen, Zhihai, Zhang, Fujie, Hao, Yiwei, Chen, Meiling, Ma, Xiaoyang, Guo, Xiaodi. Specific dynamic variations in the peripheral blood lymphocyte subsets in COVID-19 and severe influenza A patients: a retrospective observational study. BMC Infectious Diseases. 2020///; 20(1): 910. doi: http://dx.doi.org/10.1186/s12879-020-05637-9. Available at: http://www.biomedcentral.com/bmcinfectdis/,http://ovidsp.ovid.com/ovidweb.cgi?T=JS&PAGE=reference&D=emexb&NEWS=N&AN=2007476342 |
| 44 | Li, Li, Gou, Chun-yan, Wang, Xiao-jun, Li, Hong-ying, Li, Hong-yan, Li, Xiu-hui, Li, Xue-mei, Song, Wen-yan. Effects of Chinese Medicine on Symptoms, Syndrome Evolution, and Lung Inflammation Absorption in COVID-19 Convalescent Patients during 84-Day Follow-up after Hospital Discharge: A Prospective Cohort and Nested Case-Control Study. Chinese Journal of Integrative Medicine. 2021///; 27(4): 245. doi: http://dx.doi.org/10.1007/s11655-021-3328-3. Available at: http://www.springerlink.com/content/1993-0402,http://ovidsp.ovid.com/ovidweb.cgi?T=JS&PAGE=reference&D=emexb&NEWS=N&AN=2010353683 |
| 45 | Wang, Xiaoli, Pan, Yang, Wang, Quanyi, Guo, Xianghua, Feng, Yingmei, Xin, Qianqian, Hu, Yaling, Li, Jing, Chu, Yanhui. Neutralizing antibody responses to severe acute respiratory syndrome coronavirus 2 in coronavirus disease 2019 inpatients and convalescent patients. Clinical Infectious Diseases. 2020///; 71(10): 2688. doi: http://dx.doi.org/10.1093/cid/ciaa721. Available at: http://cid.oxfordjournals.org/content/by/year,http://ovidsp.ovid.com/ovidweb.cgi?T=JS&PAGE=reference&D=emexb&NEWS=N&AN=2010715407 |
| 46 | Cox, Stuart, James, Tim, Jeffery, Katie, Lumley, Sheila F, O'Donnell, Denise, Howarth, Alison, Hatch, Stephanie B, Cornall, Richard J, Jones, E Y, Stuart, David I, Screaton, Gavin, Conlon, Christopher P, Wei, Jia, Stoesser, Nicole E, Matthews, Philippa C, Hoosdally, Sarah, Crook, Derrick W, Walker, A S, Peto, Tim E A, Marsden, Brian D, Peck, Liam J, Ritter, Thomas G, de Toledo, Zoe, Ebner, Daniel, Pouwels, Koen B, Walker, Timothy M, Eyre, David W. The duration, dynamics and determinants of SARS-CoV-2 antibody responses in individual healthcare workers. Clinical infectious diseases : an official publication of the Infectious Diseases Society of America. 2021///; #volume#(#issue#): #pages#. doi: http://dx.doi.org/10.1093/cid/ciab004. Available at: http://ovidsp.ovid.com/ovidweb.cgi?T=JS&PAGE=reference&D=emexb&NEWS=N&AN=633942603 |
| 47 | Cassaniti, Irene, Percivalle, Elena, Bergami, Federica, Piralla, Antonio, Comolli, Giuditta, Bruno, Raffaele, Vecchia, Marco, Sambo, Margherita, Colaneri, Marta, Zuccaro, Valentina, Benazzo, Marco, Robotti, Carlo, Calastri, Anna, Maiorano, Eugenia, Ferrari, Alessandro, Cambie, Giuseppe, Baldanti, Fausto. SARS-CoV-2 specific T cell immunity in COVID-19 convalescent patients and unexposed controls measured by ex-vivo ELISpot assay. Clinical microbiology and infection : the official publication of the European Society of Clinical Microbiology and Infectious Diseases. 2021///; #volume#(#issue#): #pages#. doi: https://dx.doi.org/10.1016/j.cmi.2021.03.010. Available at: http://ovidsp.ovid.com/ovidweb.cgi?T=JS&PAGE=reference&D=medp&NEWS=N&AN=33813122 |
| 48 | Ansari, Sheeba, Memon, Mubeen, Kumar, Ratan, Memon, Sidra, Memon, Muhammad Khizar. Do COVID-19 Antibodies Provide Long-Term Protection?. Cureus. 2021///; 13(1): e12441. doi: https://dx.doi.org/10.7759/cureus.12441. Available at: http://ovidsp.ovid.com/ovidweb.cgi?T=JS&PAGE=reference&D=pmnm&NEWS=N&AN=33552760 |
| 49 | Vibholm, Line K, Nielsen, Stine S F, Frattari, Giacomo S, Olesen, Rikke, Andersen, Rebecca, Monrad, Ida, Hojen, Jesper F, Schleimann, Mariane H, Pahus, Marie H, Andersen, Anna H F, Thomsen, Michelle M, Konrad, Christina V, Andersen, Sidsel D, Gunst, Jesper D, Ostergaard, Lars, Sogaard, Ole S, Tolstrup, Martin. SARS-CoV-2 persistence is associated with antigen-specific CD8 T-cell responses. EBioMedicine. 2021///; 64(#issue#): 103230. doi: http://dx.doi.org/10.1016/j.ebiom.2021.103230. Available at: http://www.journals.elsevier.com/ebiomedicine/,http://ovidsp.ovid.com/ovidweb.cgi?T=JS&PAGE=reference&D=emexb&NEWS=N&AN=2010850754 |

| | |
|---|---|
| 50 | Caro-Codon, Juan, Rey, Juan R, Iniesta, Angel M, Rosillo, Sandra O, Castrejon-Castrejon, Sergio, Rodriguez-Sotelo, Laura, Garcia-Veas, Jose, Marco, Irene, Martinez, Luis A, Martin-Polo, Lorena, Merino, Carlos, Martinez-Cossiani, Marcel, Lopez-de-Sa, Esteban, Merino, Jose L, Lip, Gregory Y H, Buno, Antonio, Gonzalez-Valle, Luis, Herrero, Alicia. Prediction of thromboembolic events and mortality by the CHADS2 and the CHA2DS2-VASc in COVID-19. Europace : European pacing, arrhythmias, and cardiac electrophysiology : journal of the working groups on cardiac pacing, arrhythmias, and cardiac cellular electrophysiology of the European Society of Cardiology. 2021///; #volume#(#issue#): #pages#. doi: http://dx.doi.org/10.1093/europace/euab015. Available at: http://ovidsp.ovid.com/ovidweb.cgi?T=JS&PAGE=reference&D=emexb&NEWS=N&AN=634247270 |
| 51 | Schlickeiser, Stephan, Schwarz, Tatjana, Steiner, Sophie, Wittke, Kirsten, Al Besher, Nabeel, Meyer, Oliver, Kalus, Ulrich, Prus, Axel, Kurth, Florian, Zoller, Thomas, Witzenrath, Martin, Sander, Leif Erik, Muller, Marcel A, Scheibenbogen, Carmen, Volk, Hans-Dieter, Drosten, Christian, Corman, Victor M, Hanitsch, Leif G. Disease Severity, Fever, Age, and Sex Correlate With SARS-CoV-2 Neutralizing Antibody Responses. Frontiers in immunology. 2020///; 11(#issue#): 628971. doi: https://dx.doi.org/10.3389/fimmu.2020.628971. Available at: http://ovidsp.ovid.com/ovidweb.cgi?T=JS&PAGE=reference&D=medl&NEWS=N&AN=33584731 |
| 52 | Huemer, Florian, Rinnerthaler, Gabriel, Jorg, Benedikt, Morre, Patrick, Stegbuchner, Birgit, Proksch, Elisabeth, Fleimisch, Stefanie, Oberkofler, Hannes, Kremser, Iris, Greil, Richard, Egle, Alexander. Results of a hospitalization policy of asymptomatic and pre-symptomatic COVID-19-positive long-term care facility residents in the province of Salzburg-a report from the AGMT COVID-19 Registry. GeroScience. 2021///; #volume#(#issue#): #pages#. doi: https://dx.doi.org/10.1007/s11357-021-00352-y. Available at: http://ovidsp.ovid.com/ovidweb.cgi?T=JS&PAGE=reference&D=medp&NEWS=N&AN=33837912 |
| 53 | Huang, Chen Lu, Fei, Ling, Li, WeiXia, Xu, Wei, Xie, Xu Dong, Li, Qiang, Chen, Liang. A Novel Prediction Model for Long-Term SARS-CoV-2 RNA Shedding in Non-Severe Adult Hospitalized Patients with COVID-19: A Retrospective Cohort Study. Infectious Diseases and Therapy. 2021///; #volume#(#issue#): #pages#. doi: http://dx.doi.org/10.1007/s40121-021-00437-3. Available at: http://www.springer.com/springer+healthcare/journal/40121,http://ovidsp.ovid.com/ovidweb.cgi?T=JS&PAGE=reference&D=emexb&NEWS=N&AN=2011030261 |
| 54 | Shen, Yinfeng, Ba, Yuanming, Hu, Yaling, Wang, Linqun, Li, Weinan. Relationship between the dynamic changes of serum 2019-nCoV IgM/IgG and patient immunity after 6 month hospital discharge. Inflammation Research. 2021///; 70(2): 241. doi: http://dx.doi.org/10.1007/s00011-020-01429-8. Available at: http://www.springerlink.com/content/1023-3830,http://ovidsp.ovid.com/ovidweb.cgi?T=JS&PAGE=reference&D=emexb&NEWS=N&AN=2007708998 |
| 55 | Hong, Lu-Xiao, Liu, Lian, Lin, Aifen, Yan, Wei-Hua. Risk factors for SARS-CoV-2 re-positivity in COVID-19 patients after discharge. International immunopharmacology. 2021///; 95(#issue#): 107579. doi: http://dx.doi.org/10.1016/j.intimp.2021.107579. Available at: http://www.elsevier.com/locate/intimp,http://ovidsp.ovid.com/ovidweb.cgi?T=JS&PAGE=reference&D=emexb&NEWS=N&AN=2011440800 |
| 56 | McDonough, A, McFeely, A, Connolly, E, Dolphin, H, O'Keeffe, A, Mello, S, Fallon, Aoife, Coughlan, T, Kennelly, S P. Hospitalised older people with covid-19: One month outcomes. Irish medical journal. 2021///; 114(2): P273. doi: #DOI#. Available at: http://imj.ie/advancing-professional-healthcare-by-the-use-of-mentorship/,http://ovidsp.ovid.com/ovidweb.cgi?T=JS&PAGE=reference&D=emexb&NEWS=N&AN=2006874224 |
| 57 | Mehta, Hemalkumar B, Li, Shuang, Goodwin, James S. Risk Factors Associated With SARS-CoV-2 Infections, Hospitalization, and Mortality Among US Nursing Home Residents. JAMA network open. 2021///; 4(3): e216315. doi: http://dx.doi.org/10.1001/jamanetworkopen.2021.6315. Available at: http://ovidsp.ovid.com/ovidweb.cgi?T=JS&PAGE=reference&D=emedx&NEWS=N&AN=634693823 |
| 58 | Donato, Michele L, Park, Steven, Baker, Melissa, Korngold, Robert, Morawski, Alison, Geng, Xue, Tan, Ming, Ip, Andrew, Goldberg, Stuart, Rowley, Scott, Chow, Kar, Brown, Emily, Zenreich, Joshua, McKiernan, Phyllis, Buttner, Kathryn, Ullrich, Anna, Long, Laura, Feinman, Rena, Ricourt, Andrea, Kemp, |

| | |
|---|---|
| | Marlo, Vendivil, Mariefel, Suh, Hyung, Balani, Bindu, Cicogna, Cristina, Sebti, Rani, Al-Khan, Abdulla, Sperber, Steven, Desai, Samit, Fanning, Stacey, Arad, Danit, Go, Ronaldo, Tam, Elizabeth, Rose, Keith, Sadikot, Sean, Siegel, David, Gutierrez, Martin, Feldman, Tatyana, Goy, Andre, Pecora, Andrew, Biran, Noa, Leslie, Lori, Gillio, Alfred, Timmapuri, Sarah, Boonstra, Michele, Singer, Sam, Kaur, Sukhdeep, Richards, Ernest, Perlin, David S. Clinical and laboratory evaluation of patients with SARS-CoV-2 pneumonia treated with high-titer convalescent plasma. JCI insight. 2021///; 6(6): #pages#. doi: https://dx.doi.org/10.1172/jci.insight.143196. Available at: http://ovidsp.ovid.com/ovidweb.cgi?T=JS&PAGE=reference&D=mesx&NEWS=N&AN=33571168 |
| 59 | McGuffin, Sarah, Campbell, Victoria L, McClurkan, Christopher L, Jing, Lichen, Zamora, Danniel, Boonyaratanakornkit, Jim, Gernsheimer, Terry B, Morishima, Chihiro, Selke, Stacy, Chaudhary, Anu, Das, Marie K, Fink, Susan L, Bryan, Andrew, Greninger, Alex L, Jerome, Keith R, Wener, Mark H, Shapiro, Adrienne E, Gross, Robin, Liang, Janie, Postnikova, Elena, Mazur, Steven, Holbrook, Michael R, Lukin, Vladimir V, Wald, Anna, Koelle, David M. Clinical, laboratory, and temporal predictors of neutralizing antibodies against SARS-CoV-2 among COVID-19 convalescent plasma donor candidates. Journal of Clinical Investigation. 2021///; 131(3): e144930. doi: http://dx.doi.org/10.1172/JCI144930. Available at: https://www.jci.org/articles/view/144930/pdf,http://ovidsp.ovid.com/ovidweb.cgi?T=JS&PAGE=reference&D=emexb&NEWS=N&AN=2010889502 |
| 60 | Tudoran, Cristina, Tudoran, Mariana, Lazureanu, Voichita Elena, Marinescu, Adelina Raluca, Pop, Gheorghe Nicusor, Pescariu, Alexandru Silvius, Cut, Talida Georgiana, Enache, Alexandra. Evidence of pulmonary hypertension after SARS-CoV-2 infection in subjects without previous significant cardiovascular pathology. Journal of Clinical Medicine. 2021///; 10(2): 1. doi: http://dx.doi.org/10.3390/jcm10020199. Available at: https://www.mdpi.com/2077-0383/10/2/199/pdf,http://ovidsp.ovid.com/ovidweb.cgi?T=JS&PAGE=reference&D=emexb&NEWS=N&AN=2005786816 |
| 61 | Van Elslande, Jan, Ailliet, Scott, Oyaert, Matthijs, Vandendriessche, Stien, Van Ranst, Marc, Andre, Emmanuel, Lagrou, Katrien, Lorent, Natalie, Vande Weygaerde, Yannick, Vermeersch, Pieter. Longitudinal follow-up of IgG anti-nucleocapsid antibodies in SARS-CoV-2 infected patients up to eight months after infection. Journal of Clinical Virology. 2021///; 136(#issue#): 104765. doi: http://dx.doi.org/10.1016/j.jcv.2021.104765. Available at: http://www.elsevier.com/inca/publications/store/5/2/4/0/6/2,http://ovidsp.ovid.com/ovidweb.cgi?T=JS&PAGE=reference&D=emexb&NEWS=N&AN=2011111587 |
| 62 | Saab, Faysal G, Brook, Rachel, Chiang, Jeffrey N, Adamson, Paul C, Fulcher, Jennifer A, Goodman-Meza, David, Halperin, Eran, Manuel, Vladimir. Discharge Clinical Characteristics and Post-Discharge Events in Patients with Severe COVID-19: A Descriptive Case Series. Journal of General Internal Medicine. 2021///; #volume#(#issue#): #pages#. doi: http://dx.doi.org/10.1007/s11606-020-06494-7. Available at: http://www.springerlink.com/content/120414/,http://ovidsp.ovid.com/ovidweb.cgi?T=JS&PAGE=reference&D=emexb&NEWS=N&AN=2010352817 |
| 63 | Kutsuna, Satoshi, Kinoshita, Noriko, Terada, Mari, Miyazato, Yusuke, Nakamoto, Takato, Suzuki, Tetsuya, Saito, Sho, Kanda, Kohei, Ohmagari, Norio, Asai, Yusuke, Endo, Mio, Matsunaga, Akihiro, Kenji, Maeda, Takasaki, Jin, Hojo, Masayuki, Ishizaka, Yukihito. Factors associated with anti-SARS-CoV-2 IgG antibody production in patients convalescing from COVID-19. Journal of Infection and Chemotherapy. 2021///; #volume#(#issue#): #pages#. doi: http://dx.doi.org/10.1016/j.jiac.2021.01.006. Available at: http://www.journals.elsevier.com/journal-of-infection-and-chemotherapy/,http://ovidsp.ovid.com/ovidweb.cgi?T=JS&PAGE=reference&D=emexb&NEWS=N&AN=2010866519 |
| 64 | Anichini, Gabriele, Terrosi, Chiara, Gori Savellini, Gianni, Prathyumnan, Shibily, Gandolfo, Claudia, Cusi, Maria Grazia, Fabrizi, Simonetta, Miceli, Giovanni Battista, Franchi, Federico. Antibody response to SARS-CoV-2 in infected patients with different clinical outcome. Journal of medical virology. 2021///; 93(4): 2548. doi: http://dx.doi.org/10.1002/jmv.26789. Available at: http://onlinelibrary.wiley.com/journal/10.1002/(ISSN)1096-9071,http://ovidsp.ovid.com/ovidweb.cgi?T=JS&PAGE=reference&D=emexb&NEWS=N&AN=2010264496 |

| | |
|---|---|
| 65 | Clarke CL Dhutia A, Gan J, Edwards C, Prout V, Lightstone L, Parker E, Marchesin F, Griffith M, Charif R, Pickard G, Cox A, McClure M, Tedder R, Randell P, Greathead L, Guckian M, McAdoo SP, Kelleher P, Willicombe,M., Prendecki M. Longevity of SARS-CoV-2 immune responses in hemodialysis patients and protection against reinfection; 7992297 CSR - Reports results. Kidney international. 2021///; #volume#(#issue#): #pages#. doi: 10.1016/j.kint.2021.03.009. Available at: https://pubmed.ncbi.nlm.nih.gov/33774082 |
| 66 | Shuwa, Halima A, Shaw, Tovah N, Knight, Sean B, Wemyss, Kelly, McClure, Flora A, Pearmain, Laurence, Prise, Ian, Jagger, Christopher, Morgan, David J, Khan, Saba, Brand, Oliver, Mann, Elizabeth R, Ustianowski, Andrew, Bakerly, Nawar Diar, Dark, Paul, Brightling, Christopher E, Brij, Seema, CIRCO, Felton, Timothy, Simpson, Angela, Grainger, John R, Hussell, Tracy, Konkel, Joanne E, Menon, Madhvi. Alterations in T and B cell function persist in convalescent COVID-19 patients. Med (New York, N.Y.). 2021///; #volume#(#issue#): #pages#. doi: https://dx.doi.org/10.1016/j.medj.2021.03.013. Available at: http://ovidsp.ovid.com/ovidweb.cgi?T=JS&PAGE=reference&D=medp&NEWS=N&AN=33821250 |
| 67 | Valentini, Ricardo, Fernandez, Jose, Riveros, Dardo, Solimano, Jorge, Laviano, Julia, Fornillo, Florencia, Maymo, Daniela, Gotta, Daniel, Dupont, Juan, Palizas, Fernando, Saul, Pablo, Medina, Juan, Falasco, Viviana, Dupont, Maria Laura, Martinez, Alfredo, Bonvehi, Pablo. Convalescent plasma as a therapy for severe covid-19 pneumonia. Medicina (Argentina). 2020///; 80(#issue#): 9. doi: #DOI#. Available at: https://www.medicinabuenosaires.com/indices-de-2020/volumen-80-ano-2020-s-6-indice/plasma/,http://ovidsp.ovid.com/ovidweb.cgi?T=JS&PAGE=reference&D=emexb&NEWS=N&AN=2006727217 |
| 68 | Antar, Annukka A R, Yu, Tong, Pisanic, Nora, Azamfirei, Razvan, Tornheim, Jeffrey A, Brown, Diane M, Kruczynski, Kate, Hardick, Justin P, Sewell, Thelio, Jang, Minyoung, Church, Taylor, Walch, Samantha N, Reuland, Carolyn, Bachu, Vismaya S, Littlefield, Kirsten, Park, Han-Sol, Ursin, Rebecca L, Ganesan, Abhinaya, Kusemiju, Oyinkansola, Barnaba, Brittany, Charles, Curtisha, Prizzi, Michelle, Johnstone, Jaylynn R, Payton, Christine, Dai, Weiwei, Fuchs, Joelle, Massaccesi, Guido, Armstrong, Derek T, Townsend, Jennifer L, Keller, Sara C, Demko, Zoe O, Hu, Chen, Wang, Mei-Cheng, Sauer, Lauren M, Mostafa, Heba H, Keruly, Jeanne C, Mehta, Shruti H, Klein, Sabra L, Cox, Andrea L, Pekosz, Andrew, Heaney, Christopher D, Thomas, David L, Blair, Paul W, Manabe, Yukari C. Delayed rise of oral fluid antibodies, elevated BMI, and absence of early fever correlate with longer time to SARS-CoV-2 RNA clearance in an longitudinally sampled cohort of COVID-19 outpatients. medRxiv : the preprint server for health sciences. 2021///; #volume#(#issue#): #pages#. doi: https://dx.doi.org/10.1101/2021.03.02.21252420. Available at: http://ovidsp.ovid.com/ovidweb.cgi?T=JS&PAGE=reference&D=pmnm&NEWS=N&AN=33688688 |
| 69 | Hachim, Asmaa, Gu, Haogao, Kavian, Otared, Kwan, Mike Yw, Chan, Wai-Hung, Yau, Yat Sun, Chiu, Susan S, Tsang, Owen Ty, Hui, David Sc, Ma, Fionn, Lau, Eric Hy, Cheng, Samuel Ms, Poon, Leo Lm, Peiris, Js Malik, Valkenburg, Sophie A, Kavian, Niloufar. The SARS-CoV-2 antibody landscape is lower in magnitude for structural proteins, diversified for accessory proteins and stable long-term in children. medRxiv : the preprint server for health sciences. 2021///; #volume#(#issue#): #pages#. doi: https://dx.doi.org/10.1101/2021.01.03.21249180. Available at: http://ovidsp.ovid.com/ovidweb.cgi?T=JS&PAGE=reference&D=pmnm&NEWS=N&AN=33655259 |
| 70 | Favresse, Julien, Eucher, Christine, Elsen, Marc, Gillot, Constant, Van Eeckhoudt, Sandrine, Dogne, Jean-Michel, Douxfils, Jonathan. Persistence of Anti-SARS-CoV-2 Antibodies Depends on the Analytical Kit: A Report for Up to 10 Months after Infection. Microorganisms. 2021///; 9(3): #pages#. doi: https://dx.doi.org/10.3390/microorganisms9030556. Available at: http://ovidsp.ovid.com/ovidweb.cgi?T=JS&PAGE=reference&D=pmnm&NEWS=N&AN=33800489 |
| 71 | Juno, Jennifer A, Selva, Kevin J, Tan, Hyon-Xhi, Lee, Wen Shi, Wragg, Kathleen M, Davis, Samantha K, Mordant, Francesca L, Chung, Amy W, Wheatley, Adam K, Kelly, Hannah G, Esterbauer, Robyn, Wang, Jing J, Reynaldi, Arnold, Khoury, David S, Cromer, Deborah, Davenport, Miles P, Kent, Helen E, Kent, Stephen J, Schlub, Timothy E, Gordon, David L, Subbarao, Kanta, Gordon, Tom P. Evolution of immune responses to SARS-CoV-2 in mild-moderate COVID-19. Nature Communications. 2021///; 12(1): 1162. doi: http://dx.doi.org/10.1038/s41467-021-21444-5. Available at: |

| | |
|---|---|
| | http://www.nature.com/ncomms/index.html,http://ovidsp.ovid.com/ovidweb.cgi?T=JS&PAGE=reference&D=emexb&NEWS=N&AN=2010534570 |
| 72 | Wu, Jun, Liang, Boyun, Wang, Hua, Yang, Xiaoli, Wang, Baoju, Liu, Jia, Yang, Xuecheng, Li, Wei, Zhu, Bin, Zhou, Wenqing, Wang, Huan, Li, Sumeng, Lu, Sihong, Yang, Dongliang, Zheng, Xin, Chen, Cunrong, Fang, Yaohui, Shen, Shu, Deng, Fei, Chen, Liangkai, Chen, Qi, Wu, Yang, Liu, Di, Li, Huadong, Krawczyk, Adalbert, Lu, Mengji, Dittmer, Ulf, Trilling, Mirko. SARS-CoV-2 infection induces sustained humoral immune responses in convalescent patients following symptomatic COVID-19. Nature Communications. 2021///; 12(1): 1813. doi: http://dx.doi.org/10.1038/s41467-021-22034-1. Available at: http://www.nature.com/ncomms/index.html,http://ovidsp.ovid.com/ovidweb.cgi?T=JS&PAGE=reference&D=emexb&NEWS=N&AN=2010874161 |
| 73 | Ewasiuk, Victoria, Gregory, Louise, Chen, Zhong, Fluck, David S, Akhtar, Zaki, Gallagher, Mark M, Leung, Lisa W M, Elbatran, Ahmed I, Madden, Brendan, Breathnach, Aodhan, Yap, Yee Guan, Sharma, Sumeet. Prolonged QT predicts prognosis in COVID-19. Pacing and clinical electrophysiology : PACE. 2021///; #volume#(#issue#): #pages#. doi: http://dx.doi.org/10.1111/pace.14232. Available at: http://ovidsp.ovid.com/ovidweb.cgi?T=JS&PAGE=reference&D=emedx&NEWS=N&AN=634697120 |
| 74 | Asiman, Ikhra, Latif, Ansar, Ansar, Anila, Shafique, Muhammad Nadeem, Zia, Muhammad Qasim, Saleem, Nida. How Covid-19 pandemic effects dialysis patients in sialkot district - A cohort study. Pakistan Journal of Medical and Health Sciences. 2020///; 14(4): 728. doi: #DOI#. Available at: https://pjmhsonline.com/published-issues/2020-issues/oct-dec-2020?product_id=9301,http://ovidsp.ovid.com/ovidweb.cgi?T=JS&PAGE=reference&D=emexb&NEWS=N&AN=2010623541 |
| 75 | De Vito, Andrea, Fiore, Vito, Princic, Elija, Geremia, Nicholas, Napodano, Catello Mario Panu, Muredda, Alberto Augusto, Maida, Ivana, Babudieri, Sergio, Madeddu, Giordano. Predictors of infection, symptoms development, and mortality in people with SARS-CoV-2 living in retirement nursing homes. PLoS ONE. 2021///; 16(3): e0248009. doi: http://dx.doi.org/10.1371/journal.pone.0248009. Available at: https://journals.plos.org/plosone/article/file?id=10.1371/journal.pone.0248009&type=printable,http://ovidsp.ovid.com/ovidweb.cgi?T=JS&PAGE=reference&D=emexb&NEWS=N&AN=2011442920 |
| 76 | Khairallah, Pascale, Toma, Katherine, Verduzco, Hector Alvarado, Daniel, Emily, Douglas, Denzil, Moses, Andrew A, Peleg, Yonatan, Starakiewicz, Piotr, Shah, Vaqar H, King, Kristen L, Li, Miah T, Kim, Daniel W, Stevens, Jacob S, Robbins-Juarez, Shelief, Yu, Kathleen, Qian, Long, Krzysztof, Kiryluk, Dube, Geoffrey, Siddall, Eric, Shirazian, Shayan, Nickolas, Thomas L, Rao, Maya K, Barasch, Jonathan M, Valeri, Anthony M, Radhakrishnan, Jai, Gharavi, Ali G, Husain, S A, Mohan, Sumit, Cummings, Matthew J, O'Donnell, Max R, Zucker, Jason, Natarajan, Karthik, Perotte, Adler, Tsapepas, Demetra. High rate of renal recovery in survivors of COVID-19 associated acute renal failure requiring renal replacement therapy. PLoS ONE. 2020///; 15(12): e0244131. doi: http://dx.doi.org/10.1371/journal.pone.0244131. Available at: https://journals.plos.org/plosone/article/file?id=10.1371/journal.pone.0244131&type=printable,http://ovidsp.ovid.com/ovidweb.cgi?T=JS&PAGE=reference&D=emexb&NEWS=N&AN=2010581881 |
| 77 | Rachfalski, David, Bauer, Jens, Rieth, Jonas, Bilich, Tatjana, Nelde, Annika, Heitmann, Jonas S, Maringer, Yacine, Marklin, Melanie, Roerden, Malte, Wacker, Marcel, Horber, Sebastian, Stevanovic, Stefan, Rammensee, Hans-Georg, Salih, Helmut R, Walz, Juliane S. T cell and antibody kinetics delineate SARS-CoV-2 peptides mediating long-term immune responses in COVID-19 convalescent individuals. Science translational medicine. 2021///; #volume#(#issue#): #pages#. doi: http://dx.doi.org/10.1126/scitranslmed.abf7517. Available at: http://ovidsp.ovid.com/ovidweb.cgi?T=JS&PAGE=reference&D=emexb&NEWS=N&AN=634577178 |
| 78 | Blain, Maxime, Xu, Sheng, Amalou, Amel, Kassin, Michael T, Turkbey, Evrim B, Varble, Nicole, Harmon, Stephanie, Turkbey, Baris, Yang, Dong, Xu, Ziyue, Roth, Holger, Xu, Daguang, Flores, Mona, Sun, Kaiyun, Kadri, Sameer, Patella, Francesca, Cariati, Maurizio, Scarabelli, Alice, Stellato, Elvira, Ierardi, Anna Maria, Carrafiello, Gianpaolo, An, Peng, Wood, Bradford J. Generalized chest CT and lab curves throughout the course of COVID-19. Scientific reports. 2021///; 11(1): 6940. doi: http://dx.doi.org/10.1038/s41598-021-85694-5. Available at: http://ovidsp.ovid.com/ovidweb.cgi?T=JS&PAGE=reference&D=emexb&NEWS=N&AN=634665132 |

| | |
|---|---|
| 79 | Lee, Hye Kyung, Smith, Harold E, Hennighausen, Lothar, Pipperger, Lisa, Volland, Andre, von Laer, Dorothee, Furth, Priscilla A, Kang, Keunsoo, Knabl, Ludwig, Bellmann, Romuald, Bernhard, Christina, Kaiser, Norbert, Ganzer, Hannes, Strohle, Mathias, Walser, Andreas. Immune transcriptomes of highly exposed SARS-CoV-2 asymptomatic seropositive versus seronegative individuals from the Ischgl community. Scientific reports. 2021///; 11(1): 4243. doi: http://dx.doi.org/10.1038/s41598-021-83110-6. Available at: http://ovidsp.ovid.com/ovidweb.cgi?T=JS&PAGE=reference&D=emexb&NEWS=N&AN=634354474 |
| 80 | Khoo, Bernard, Tan, Tricia, Clarke, Sophie A, Patel, Bijal, Modi, Manish, Alexander, Emma C, Mills, Edouard G, Phylactou, Maria, Eng, Pei Chia, Thurston, Layla, Meeran, Karim, Comninos, Alexander N, Abbara, Ali, Dhillo, Waljit S. Thyroid Function Before, During, and After COVID-19. The Journal of clinical endocrinology and metabolism. 2021///; 106(2): e803. doi: http://dx.doi.org/10.1210/clinem/dgaa830. Available at: http://ovidsp.ovid.com/ovidweb.cgi?T=JS&PAGE=reference&D=emexb&NEWS=N&AN=633410512 |
| 81 | Racine-Brzostek, Sabrina, Yang, He S, Chadburn, Amy, Ma, Lucy, Sweeney, Jacob, Zhong, Elaine, Yee, Jim, Cushing, Melissa M, Zhao, Zhen, Jack, Gwendolyne A, Chen, Zhengming, Ketas, Thomas J, Francomano, Erik, Klasse, P J, Moore, John P, McDonough, Kathleen A, Girardin, Roxanne C, Dupuis Ii, Alan P, Payne, Anne F. Post Convalescent SARS-CoV-2 IgG and Neutralizing Antibodies Are Elevated in Individuals with Poor Metabolic Health. The Journal of clinical endocrinology and metabolism. 2021///; #volume#(#issue#): #pages#. doi: http://dx.doi.org/10.1210/clinem/dgab004. Available at: http://ovidsp.ovid.com/ovidweb.cgi?T=JS&PAGE=reference&D=emexb&NEWS=N&AN=634170327 |
| 82 | Qin, Yuanyuan, Li, Jia, Zhang, Guiling, Wu, Di, Zhou, Yiran, Zhu, Wenzhen, Wu, Jinfeng, Cai, Aoling, Chen, Tao, Zheng, Ning, Xu, Fuqiang, Ning, Qin, Manyande, Anne, Wang, Jie. Long-term micro-structure and cerebral blood flow changes in patients recovered from COVID-19 without neurological manifestations. The Journal of clinical investigation. 2021///; #volume#(#issue#): #pages#. doi: http://dx.doi.org/10.1172/JCI147329. Available at: http://ovidsp.ovid.com/ovidweb.cgi?T=JS&PAGE=reference&D=emexb&NEWS=N&AN=634406770 |
| 83 | Kaptein, F.H.J., Stals, M.A.M., Grootenboers, M., Braken, S.J.E., Burggraaf, J.L.I., van Bussel, B.C.T., Cannegieter, S.C., ten Cate, H., Endeman, H., Gommers, D.A.M.P.J., van Guldener, C., de Jonge, E., Juffermans, N.P., Kant, K.M., Kevenaar, M.E., Koster, S., Kroft, L.J.M., Kruip, M.J.H.A., Leentjens, J., Marechal, C., Soei, Y.L., Tjepkema, L., Visser, C., Klok, F.A., Huisman, M.V.. Incidence of thrombotic complications and overall survival in hospitalized patients with COVID-19 in the second and first wave. Thrombosis Research. 2021/03//; 199(#issue#): 143. doi: 10.1016/j.thromres.2020.12.019. Available at: http://www.elsevier.com/locate/thromres,http://ovidsp.ovid.com/ovidweb.cgi?T=JS&PAGE=reference&D=emexb&NEWS=N&AN=2010858142,https://linkinghub.elsevier.com/retrieve/pii/S0049384820306757 |
| 84 | Kute, Vivek B, Ray, Deepak S, Yadav, Dinesh Kumar, Pathak, Vivek, Bhalla, Anil K, Godara, Suraj, Kumar, Anil, Guleria, Sandeep, Khullar, Dinesh, Thukral, Sharmila, Mondal, Rabi Ranjan Sow, Jain, Manish, Jha, Pranaw Kumar, Hegde, Umapati, Abi, Abraham M, Dalal, Sonal, Patel, H Imanshu, Bahadur, Madan M, Shingare, Ashay, Sharma, Ashish, Sharma, Raj Kumar, Anandh, Urmila, Gulati, Sanjeev, Gumber, Manoj, Siddini, Vishwanath, Deshpande, Rushi, Kaswan, Kamal, Varyani, Umesh, Kakde, Shailesh, Kenwar, Deepesh B, Meshram, Hari Shankar, Kher, Vijay. A Multicenter Cohort Study From India of 75 Kidney Transplants in Recipients Recovered After COVID-19. Transplantation. 2021///; #volume#(#issue#): #pages#. doi: http://dx.doi.org/10.1097/TP.0000000000003740. Available at: http://ovidsp.ovid.com/ovidweb.cgi?T=JS&PAGE=reference&D=emexb&NEWS=N&AN=634578794 |
| 85 | Wang, Dianbing, Hou, Baidong, Li, Min, Cui, Mengmeng, Zheng, Yang, Zhang, Qing, Zhou, Xiaoli, Ye, Zhiqin, Chen, Xiaomin, Deng, Lihua, Sun, Xinyi, Zhang, Qian, Li, Yong, Cheng, Yegang, Cao, Shanshan, Cui, Jing, Zhou, Juan, Ali, Ashaq, Shao, Nan, Men, Dong, Zhang, Xian-En, Li, Ke, Yang, Qinfang, Wang, Hui, Lei, Yake, Yu, Bo, Tong, Xiaolin. Sustainability of SARS-CoV-2 Induced Humoral Immune Responses in COVID-19 Patients from Hospitalization to Convalescence Over Six Months. Virologica Sinica. 2021///; #volume#(#issue#): #pages#. doi: http://dx.doi.org/10.1007/s12250-021-00360-4. Available at: http://ovidsp.ovid.com/ovidweb.cgi?T=JS&PAGE=reference&D=emexb&NEWS=N&AN=634475446 |

**Data not available for outcome or population of interest = 41**

| | |
|---|---|
| 1 | Taquet M., Luciano S., Geddes J.R., Harrison P.J.. Bidirectional associations between COVID-19 and psychiatric disorder: retrospective cohort studies of 62 354 COVID-19 cases in the USA. The Lancet Psychiatry. 2020; #volume#(#issue#): #pages#. doi: http://dx.doi.org/10.1016/S2215-0366%2820%2930462-4. Available at: https://www.journals.elsevier.com/the-lancet-psychiatry; http://ovidsp.ovid.com/ovidweb.cgi?T=JS&PAGE=reference&D=emexc&NEWS=N&AN=2010119196 |
| 2 | Poyraz B.C., Poyraz C.A., Olgun Y., Gurel O., Alkan S., Ozdemir Y.E., Balkan I.I., Karaali R.. Psychiatric morbidity and protracted symptoms after COVID-19. Psychiatry research. 2021; 295(#issue#): 113604. doi: http://dx.doi.org/10.1016/j.psychres.2020.113604. Available at: http://www.elsevier.com/locate/psychres; http://ovidsp.ovid.com/ovidweb.cgi?T=JS&PAGE=reference&D=emexc&NEWS=N&AN=2010254203 |
| 3 | Liu H.-Q., Yuan B., An Y.-W., Chen K.-J., Hu Q., Hu X.-P., Zhao J., Dong Y., Chen Y.-X., Li W.-X., Sun C.-Q., Wang J.-C., Wang C., Song S.. Clinical characteristics and follow-up analysis of 324 discharged covid-19 patients in shenzhen during the recovery period. International Journal of Medical Sciences. 2021; 18(2): 347. doi: http://dx.doi.org/10.7150/ijms.50873. Available at: https://www.medsci.org/v18p0347.pdf; http://ovidsp.ovid.com/ovidweb.cgi?T=JS&PAGE=reference&D=emexc&NEWS=N&AN=2005528461 |
| 4 | Galvan-Tejada C.E., Herrera-Garcia C.F., Godina-Gonzalez S., Villagrana-Banuelos K.E., Amaro J.D.L., Herrera-Garcia K., Rodriguez-Quinones C., Zanella-Calzada L.A., Ramirez-Barranco J., Ruiz de Avila J.L., Reyes-Escobedo F., Celaya-Padilla J.M., Galvan-Tejada J.I., Gamboa-Rosales H., Martinez-Acuna M., Cervantes-Villagrana A., Rivas-Santiago B., Gonzalez-Curiel I.E.. Persistence of covid-19 symptoms after recovery in mexican population. International Journal of Environmental Research and Public Health. 2020; 17(24): 1. doi: http://dx.doi.org/10.3390/ijerph17249367. Available at: https://www.mdpi.com/1660-4601/17/24/9367/pdf; http://ovidsp.ovid.com/ovidweb.cgi?T=JS&PAGE=reference&D=emexc&NEWS=N&AN=2005610752 |
| 5 | Battaglini D., Santori G., Chandraptham K., Iannuzzi F., Bastianello M., Tarantino F., Ball L., Giacobbe D.R., Vena A., Bassetti M., Inglese M., Uccelli A., Rocco P.R.M., Patroniti N., Brunetti I., Pelosi P., Robba C.. Neurological Complications and Noninvasive Multimodal Neuromonitoring in Critically Ill Mechanically Ventilated COVID-19 Patients. Frontiers in Neurology. 2020; 11(#issue#): 602114. doi: http://dx.doi.org/10.3389/fneur.2020.602114. Available at: http://www.frontiersin.org/Neurology; http://ovidsp.ovid.com/ovidweb.cgi?T=JS&PAGE=reference&D=emexc&NEWS=N&AN=633619485 |
| 6 | Liotta E.M., Batra A., Clark J.R., Shlobin N.A., Hoffman S.C., Orban Z.S., Koralnik I.J.. Frequent neurologic manifestations and encephalopathy-associated morbidity in Covid-19 patients. Annals of Clinical and Translational Neurology. 2020; 7(11): 2221. doi: http://dx.doi.org/10.1002/acn3.51210. Available at: http://onlinelibrary.wiley.com/journal/10.1002/(ISSN)2328-9503; http://ovidsp.ovid.com/ovidweb.cgi?T=JS&PAGE=reference&D=emexc&NEWS=N&AN=2006876225 |
| 7 | Yan C.H., Prajapati D.P., Ritter M.L., DeConde A.S.. Persistent Smell Loss Following Undetectable SARS-CoV-2. Otolaryngology - Head and Neck Surgery (United States). 2020; 163(5): 923. doi: http://dx.doi.org/10.1177/0194599820934769. Available at: http://oto.sagepub.com/content/by/year; http://ovidsp.ovid.com/ovidweb.cgi?T=JS&PAGE=reference&D=emexc&NEWS=N&AN=2005188853 |
| 8 | Yang Z.L., Chen C., Huang L., Zhou S.C., Hu Y.N., Xia L.M., Li Y.. Fibrotic Changes Depicted by Thin-Section CT in Patients With COVID-19 at the Early Recovery Stage: Preliminary Experience. Frontiers in Medicine. 2020; 7(#issue#): 605088. doi: http://dx.doi.org/10.3389/fmed.2020.605088. Available at: http://journal.frontiersin.org/journal/medicine; http://ovidsp.ovid.com/ovidweb.cgi?T=JS&PAGE=reference&D=emexc&NEWS=N&AN=633608151 |
| 9 | Oh T.K., Park H.Y., Song I.-A.. Risk of psychological sequelae among coronavirus disease-2019 survivors: A nationwide cohort study in South Korea. Depression and anxiety. 2020; #volume#(#issue#): #pages#. doi: http://dx.doi.org/10.1002/da.23124. Available at: |

| | |
|---|---|
| | http://onlinelibrary.wiley.com/journal/10.1002/(ISSN)1520-6394; http://ovidsp.ovid.com/ovidweb.cgi?T=JS&PAGE=reference&D=emexc&NEWS=N&AN=2007543208 |
| 10 | Mizrahi B., Shilo S., Rossman H., Kalkstein N., Marcus K., Barer Y., Keshet A., Shamir-Stein N., Shalev V., Zohar A.E., Chodick G., Segal E.. Longitudinal symptom dynamics of COVID-19 infection. Nature Communications. 2020; 11(1): 6208. doi: http://dx.doi.org/10.1038/s41467-020-20053-y. Available at: http://www.nature.com/ncomms/index.html; http://ovidsp.ovid.com/ovidweb.cgi?T=JS&PAGE=reference&D=emexc&NEWS=N&AN=2007505815 |
| 11 | Puntmann V.O., Carerj M.L., Wieters I., Fahim M., Arendt C., Hoffmann J., Shchendrygina A., Escher F., Vasa-Nicotera M., Zeiher A.M., Vehreschild M., Nagel E.. Outcomes of Cardiovascular Magnetic Resonance Imaging in Patients Recently Recovered from Coronavirus Disease 2019 (COVID-19). JAMA Cardiology. 2020; 5(11): 1265. doi: http://dx.doi.org/10.1001/jamacardio.2020.3557. Available at: http://jamanetwork.com/journals/jamacardiology; http://ovidsp.ovid.com/ovidweb.cgi?T=JS&PAGE=reference&D=emexc&NEWS=N&AN=632543321 |
| 12 | Klironomos S., Tzortzakakis A., Kits A., Ohberg C., Kollia E., Ahoromazdae A., Almqvist H., Aspelin A., Martin H., Ouellette R., Al-Saadi J., Hasselberg M., Haghgou M., Pedersen M., Petersson S., Finnsson J., Lundberg J., Delgado A.F., Granberg T.. Nervous system involvement in coronavirus disease 2019: Results from a retrospective consecutive neuroimaging cohort. Radiology. 2020; 297(3): E324. doi: http://dx.doi.org/10.1148/radiol.2020202791. Available at: https://pubs.rsna.org/doi/pdf/10.1148/radiol.2020202791; http://ovidsp.ovid.com/ovidweb.cgi?T=JS&PAGE=reference&D=emexc&NEWS=N&AN=2010076661 |
| 13 | Zhang S., Liu L., Yang B., Li R., Luo J., Huang J., Long Y., Huang Y., Zhou J., Zha Y., Zhang X.. Clinical characteristics of 134 convalescent patients with COVID-19 in Guizhou, China. Respiratory Research. 2020; 21(1): 314. doi: http://dx.doi.org/10.1186/s12931-020-01580-0. Available at: http://respiratory-research.com/home/; http://ovidsp.ovid.com/ovidweb.cgi?T=JS&PAGE=reference&D=emexc&NEWS=N&AN=2007391223 |
| 14 | Needham E., Newcombe V., Michell A., Thornton R., Grainger A., Anwar F., Warburton E., Menon D., Trivedi M., Sawcer S.. Mononeuritis multiplex: an unexpectedly frequent feature of severe COVID-19. Journal of neurology. 2020; #volume#(#issue#): #pages#. doi: http://dx.doi.org/10.1007/s00415-020-10321-8. Available at: https://link.springer.com/journal/415; http://ovidsp.ovid.com/ovidweb.cgi?T=JS&PAGE=reference&D=emexc&NEWS=N&AN=2007390999 |
| 15 | Altundag A., Saatci O., Sanli D.E.T., Duz O.A., Sanli A.N., Olmuscelik O., Temirbekov D., Kandemirli S.G., Karaaltin A.B.. The temporal course of COVID-19 anosmia and relation to other clinical symptoms. European Archives of Oto-Rhino-Laryngology. 2020; #volume#(#issue#): #pages#. doi: http://dx.doi.org/10.1007/s00405-020-06496-5. Available at: http://link.springer.de/link/service/journals/00405/index.htm; http://ovidsp.ovid.com/ovidweb.cgi?T=JS&PAGE=reference&D=emexc&NEWS=N&AN=2007383590 |
| 16 | Janiri D., Kotzalidis G.D., Giuseppin G., Molinaro M., Modica M., Montanari S., Terenzi B., Carfi A., Landi F., Sani G.. Psychological Distress After Covid-19 Recovery: Reciprocal Effects With Temperament and Emotional Dysregulation. An Exploratory Study of Patients Over 60 Years of Age Assessed in a Post-acute Care Service. Frontiers in Psychiatry. 2020; 11(#issue#): 590135. doi: http://dx.doi.org/10.3389/fpsyt.2020.590135. Available at: http://www.frontiersin.org/Psychiatry; http://ovidsp.ovid.com/ovidweb.cgi?T=JS&PAGE=reference&D=emexc&NEWS=N&AN=633491468 |
| 17 | Bowles K.H., McDonald M., Barron Y., Kennedy E., O'Connor M., Mikkelsen M.. Surviving COVID-19 After Hospital Discharge: Symptom, Functional, and Adverse Outcomes of Home Health Recipients. Annals of Internal Medicine. 2020; #volume#(#issue#): #pages#. doi: http://dx.doi.org/10.7326/M20-5206. Available at: http://ovidsp.ovid.com/ovidweb.cgi?T=JS&PAGE=reference&D=emexc&NEWS=N&AN=633519948 |
| 18 | Xia L., Chen J., Friedemann T., Yang Z., Ling Y., Liu X., Lu S., Li T., Song Z., Huang W., Lu Y., Schroder S., Lu H.. The course of mild and moderate COVID-19 infections-the unexpected long-lasting challenge. Open Forum Infectious Diseases. 2020; 7(9): 1. doi: http://dx.doi.org/10.1093/ofid/ofaa286. Available at: |

| | |
|---|---|
| | https://academic.oup.com/ofid; http://ovidsp.ovid.com/ovidweb.cgi?T=JS&PAGE=reference&D=emexc&NEWS=N&AN=2010099406 |
| 19 | Daynes E., Gerlis C., Briggs-Price S., Jones P., Singh S.J.. COPD assessment test for the evaluation of COVID-19 symptoms. Thorax. 2020; #volume#(#issue#): #pages#. doi: http://dx.doi.org/10.1136/thoraxjnl-2020-215916. Available at: http://thorax.bmj.com/; http://ovidsp.ovid.com/ovidweb.cgi?T=JS&PAGE=reference&D=emexc&NEWS=N&AN=633377309 |
| 20 | Caronna E., Ballve A., Llaurado A., Gallardo V.J., Maria Ariton D., Lallana S., Maza S.L., Gadea M.O., Quibus L., Restrepo J.L., Rodrigo-Gisbert M., Vilaseca A., Gonzalez M.H., Gallo M.M., Alpuente A., Torres-Ferrus M., Borrell R.P., Alvarez-Sabin J., Pozo-Rosich P.. Headache: A striking prodromal and persistent symptom, predictive of COVID-19 clinical evolution. Cephalalgia. 2020; 40(13): 1410. doi: http://dx.doi.org/10.1177/0333102420965157. Available at: http://cep.sagepub.com/; http://ovidsp.ovid.com/ovidweb.cgi?T=JS&PAGE=reference&D=emexc&NEWS=N&AN=2007163174 |
| 21 | Meini S., Suardi L.R., Busoni M., Roberts A.T., Fortini A.. Olfactory and gustatory dysfunctions in 100 patients hospitalized for COVID-19: sex differences and recovery time in real-life. European Archives of Oto-Rhino-Laryngology. 2020; 277(12): 3519. doi: http://dx.doi.org/10.1007/s00405-020-06102-8. Available at: http://link.springer.de/link/service/journals/00405/index.htm; http://ovidsp.ovid.com/ovidweb.cgi?T=JS&PAGE=reference&D=emexc&NEWS=N&AN=2005148004 |
| 22 | Mazza, Mario Gennaro, De Lorenzo, Rebecca, Conte, Caterina, Poletti, Sara, Vai, Benedetta, Bollettini, Irene, Melloni, Elisa Maria Teresa, Furlan, Roberto, Ciceri, Fabio, Rovere-Querini, Patrizia, Benedetti, Francesco. Anxiety and depression in COVID-19 survivors: Role of inflammatory and clinical predictors. Brain, behavior, and immunity. 2020; 89(#issue#): 594. doi: http://dx.doi.org/10.1016/j.bbi.2020.07.037; http://dx.doi.org/10.1016/j.bbi.2020.07.037. Available at: http://ovidsp.ovid.com/ovidweb.cgi?T=JS&PAGE=reference&D=psyc17&NEWS=N&AN=2020-77227-064 |
| 23 | Piazza, Gregory, Campia, Umberto, Hurwitz, Shelley, Snyder, Julia E., Rizzo, Samantha M., Pfeferman, Mariana B., Morrison, Ruth B., Leiva, Orly, Fanikos, John, Nauffal, Victor, Almarzooq, Zaid, Goldhaber, Samuel Z.. Registry of Arterial and Venous Thromboembolic Complications in Patients With COVID-19. Journal of the American College of Cardiology. 2020; 76(18): 2060. doi: https://dx.doi.org/10.1016/j.jacc.2020.08.070. Available at: http://ovidsp.ovid.com/ovidweb.cgi?T=JS&PAGE=reference&D=medl&NEWS=N&AN=33121712 |
| 24 | Loerinc, Leah B., Scheel, Amy M., Evans, Sean T., Shabto, Julie M., O'Keefe, Ghazala,A., O'Keefe, James,B.. Discharge characteristics and care transitions of hospitalized patients with COVID-19. Healthcare (Amsterdam, Netherlands). 2020; 9(1): 100512. doi: https://dx.doi.org/10.1016/j.hjdsi.2020.100512. Available at: http://ovidsp.ovid.com/ovidweb.cgi?T=JS&PAGE=reference&D=medp&NEWS=N&AN=33383393 |
| 25 | Starekova, Jitka, Bluemke, David A., Bradham, William S., Eckhardt, Lee L., Grist, Thomas M., Kusmirek, Joanna E., Purtell, Christopher S., Schiebler, Mark L., Reeder, Scott B.. Evaluation for Myocarditis in Competitive Student Athletes Recovering From Coronavirus Disease 2019 With Cardiac Magnetic Resonance Imaging. JAMA cardiology. 2021; #volume#(#issue#): #pages#. doi: https://dx.doi.org/10.1001/jamacardio.2020.7444. Available at: http://ovidsp.ovid.com/ovidweb.cgi?T=JS&PAGE=reference&D=medp&NEWS=N&AN=33443537 |
| 26 | Arab-Zozani M, Hashemi F, Safari H, Yousefi M, Ameri,H.. Health-Related Quality of Life and its Associated Factors in COVID-19 Patients; 7577388 CSR - Reports results. Osong public health and research perspectives. 2020; 11(5): 296. doi: 10.24171/j.phrp.2020.11.5.05. Available at: https://pubmed.ncbi.nlm.nih.gov/33117634 |
| 27 | J,Wang Y, Jin C, Wu CC, Zhao H, Liang T, Liu Z, Jian Z, Li R, Wang Z, Li F, Zhou J, Cai S, Liu Y, Li H, Liang Y, Tian C, Yang. Organizing pneumonia of COVID-19: time-dependent evolution and outcome in CT findings. PloS one. 2020; 15(11): e0240347. doi: 10.1371/journal.pone.0240347. Available at: https://pubmed.ncbi.nlm.nih.gov/33175876 |
| 28 | P,Horvath L, Lim JWJ, Taylor JW, Saief T, Stuart R, Rimmer J, Michael. Smell and taste loss in COVID-19 patients: assessment outcomes in a Victorian population. Acta Oto-Laryngologica. 2020; |

| | |
|---|---|
| | #volume#(#issue#): 1. doi: 10.1080/00016489.2020.1855366. Available at: https://pubmed.ncbi.nlm.nih.gov/33307905 |
| 29 | Paderno, A., Mattavelli, D., Rampinelli, V., Grammatica, A., Raffetti, E., Tomasoni, M., Gualtieri, T., Taboni, S., Zorzi, S., Del Bon, F., Lombardi, D., Deganello, A., Redaelli De Zinis, L. O., Schreiber, A.. Olfactory and Gustatory Outcomes in COVID-19: A Prospective Evaluation in Nonhospitalized Subjects; 32600175. Otolaryngology - Head and Neck Surgery (United States). 2020; 163(6): 1144. doi: 10.1177/0194599820939538. Available at: #URL# |
| 30 | Ercalık T Gencer-Atalay K, Akgun C, Ozdemir HM, Kuran,B., Ayyıldız A. Symptoms in COVID-19. American journal of physical medicine & rehabilitation. 2021///; #volume#(#issue#): #pages#. doi: 10.1097/PHM.0000000000001699. Available at: https://pubmed.ncbi.nlm.nih.gov/33480608 |
| 31 | Giannis, Dimitrios, Allen, Steven, Tsang, James, Flint, Sarah, Pinhasov, Tamir, Williams, Stephanie, Tan, Gary, Thakur, Richa, Leung, Christian, Snyder, Matthew Salvatore, Bhatia, Chirag, Garrett, David, Cotte, Christina, Isaacs, Shelby, Gugerty, Emma, Davidson, Anne, Marder, Galina S, Schnitzer, Austin, Goldberg, Bradley, McGinn, Thomas, Davidson, Karina, Barish, Matthew A, Qiu, Michael, Zhang, Meng, Goldin, Mark, Matsagkas, Miltiadis, Arnaoutoglou, Eleni Markos, Spyropoulos, Alex C, Northwell Health, Covid-Research Consortium. Post-Discharge Thromboembolic Outcomes and Mortality of Hospitalized COVID-19 Patients: The CORE-19 Registry. Blood. 2021///; #volume#(#issue#): #pages#. doi: https://dx.doi.org/10.1182/blood.2020010529. Available at: http://ovidsp.ovid.com/ovidweb.cgi?T=JS&PAGE=reference&D=medp&NEWS=N&AN=33824972 |
| 32 | Ayoubkhani, Daniel, Khunti, Kamlesh, Nafilyan, Vahe, Maddox, Thomas, Humberstone, Ben, Diamond, Ian, Banerjee, Amitava. Post-covid syndrome in individuals admitted to hospital with covid-19: retrospective cohort study. BMJ (Clinical research ed.). 2021///; 372(#issue#): n693. doi: https://dx.doi.org/10.1136/bmj.n693. Available at: http://ovidsp.ovid.com/ovidweb.cgi?T=JS&PAGE=reference&D=mesx&NEWS=N&AN=33789877 |
| 33 | Iqbal, Ayman, Iqbal, Kinza, Arshad Ali, Shajeea, Azim, Dua, Farid, Eisha, Baig, Mirza D, Bin Arif, Taha, Raza, Mohammad. The COVID-19 Sequelae: A Cross-Sectional Evaluation of Post-recovery Symptoms and the Need for Rehabilitation of COVID-19 Survivors. Cureus. 2021///; 13(2): e13080. doi: https://dx.doi.org/10.7759/cureus.13080. Available at: http://ovidsp.ovid.com/ovidweb.cgi?T=JS&PAGE=reference&D=pmnm&NEWS=N&AN=33680620 |
| 34 | Demelo-Rodriguez, Pablo, Ordieres-Ortega, Lucia, del Toro-Cervera, Jorge, Galeano-Valle, Francisco, Ji, Zichen, de Miguel-Diez, Javier, Alvarez-Sala-Walther, Luis A. Long-term follow-up of patients with venous thromboembolism and COVID-19: Analysis of risk factors for death and major bleeding. European journal of haematology. 2021///; #volume#(#issue#): #pages#. doi: http://dx.doi.org/10.1111/ejh.13603. Available at: http://onlinelibrary.wiley.com/journal/10.1111/(ISSN)1600-0609,http://ovidsp.ovid.com/ovidweb.cgi?T=JS&PAGE=reference&D=emexb&NEWS=N&AN=2010665940 |
| 35 | S Jarmul JA, Shaheen AW, Meaux D, Long T, Saccoccio D, Moll, Eswaran H. Vascular thromboembolic events following COVID-19 hospital discharge: incidence and risk factors; 7938613 CSR - Reports results. Research and practice in thrombosis and haemostasis. 2021///; 5(2): 292. doi: 10.1002/rth2.12485. Available at: https://pubmed.ncbi.nlm.nih.gov/33733027 |
| 36 | Bonazza, Federica, Borghi, Lidia, di San Marco, Eugenia Cao, Piscopo, Kyrie, Vegni, Elena, Bai, Francesca, d'Arminio Monforte, Antonella. Psychological outcomes after hospitalization for COVID-19: Data from a multidisciplinary follow-up screening program for recovered patients. Research in Psychotherapy: Psychopathology, Process and Outcome. 2020///; 23(3): 247. doi: http://dx.doi.org/10.4081/ripppo.2020.491. Available at: https://www.researchinpsychotherapy.org/index.php/rpsy/article/view/491,http://ovidsp.ovid.com/ovidweb.cgi?T=JS&PAGE=reference&D=emexb&NEWS=N&AN=2010898364 |
| 37 | Zampogna, Elisabetta, Paneroni, Mara, Vitacca, Michele, Belli, Stefano, Bellanti, Maria Teresa, Aliani, Maria, Gandolfo, Alessandra, Visca, Dina, Ambrosino, Nicolino. Pulmonary Rehabilitation in Patients Recovering from COVID-19. Respiration. 2021///; #volume#(#issue#): 1. doi: http://dx.doi.org/10.1159/000514387. Available at: |

| | |
|---|---|
| | http://www.karger.com/journals/res/res_jh.htm,http://ovidsp.ovid.com/ovidweb.cgi?T=JS&PAGE=reference&D=emexb&NEWS=N&AN=634658579 |
| 38 | Asare, Smart, Sandio, Aubin, Arhin, Abraham, Palla, Mohan, Ayers, Eric, Elangwe, Paul, Ihezie, Nkechi. COVID-19 Symptoms Can Prolong Seasonal Flu Infection Symptoms and Recovery Times Among Obese African American Female Population. SN Comprehensive Clinical Medicine. 2021///; 3(3): 777. doi: http://dx.doi.org/10.1007/s42399-021-00823-5. Available at: https://link.springer.com/journal/volumesAndIssues/42399,http://ovidsp.ovid.com/ovidweb.cgi?T=JS&PAGE=reference&D=emexb&NEWS=N&AN=2010580328 |
| 39 | PJ Geddes JR, Husain M, Luciano S, Harrison, Taquet M. 6-month neurological and psychiatric outcomes in 236 379 survivors of COVID-19: a retrospective cohort study using electronic health records; 8023694 CSR - Reports results. The lancet.Psychiatry. 2021///; #volume#(#issue#): #pages#. doi: 10.1016/S2215-0366(21)00084-5. Available at: https://pubmed.ncbi.nlm.nih.gov/33836148 |
| 40 | Afshar Y., Gaw S.L., Flaherman V.J., Chambers B.D., Krakow D., Berghella V., Shamshirsaz A.A., Boatin A.A., Aldrovandi G., Greiner A., Riley L., Boscardin W.J., Jamieson D.J., Jacoby V.L.. Clinical Presentation of Coronavirus Disease 2019 (COVID-19) in Pregnant and Recently Pregnant People. Obstetrics and gynecology. 2020; 136(6): 1117. doi: http://dx.doi.org/10.1097/AOG.0000000000004178. Available at: http://ovidsp.ovid.com/ovidweb.cgi?T=JS&PAGE=reference&D=emexc&NEWS=N&AN=633135763 |
| 41 | Gaber, T A-Z K, Ashish, A, Unsworth, A. Persistent post-covid symptoms in healthcare workers. Occupational medicine (Oxford, England). 2021///; #volume#(#issue#): #pages#. doi: https://dx.doi.org/10.1093/occmed/kqab043. Available at: http://ovidsp.ovid.com/ovidweb.cgi?T=JS&PAGE=reference&D=medp&NEWS=N&AN=33830208 |

**Not population of interest = 21**

| | |
|---|---|
| 1 | Hopkins C., Surda P., Vaira L.A., Lechien J.R., Safarian M., Saussez S., Kumar N.. Six month follow-up of self-reported loss of smell during the COVID-19 pandemic. Rhinology. 2020; #volume#(#issue#): #pages#. doi: http://dx.doi.org/10.4193/Rhin20.544. Available at: http://ovidsp.ovid.com/ovidweb.cgi?T=JS&PAGE=reference&D=emedx&NEWS=N&AN=633808187 |
| 2 | Sonnweber T., Sahanic S., Pizzini A., Luger A., Schwabl C., Sonnweber B., Kurz K., Koppelstatter S., Haschka D., Petzer V., Boehm A., Aichner M., Tymoszuk P., Lener D., Theurl M., Lorsbach-Kohler A., Tancevski A., Schapfl A., Schaber M., Hilbe R., Nairz M., Puchner B., Huttenberger D., Tschurtschenthaler C., Asshoff M., Peer A., Hartig F., Bellmann R., Joannidis M., Gollmann-Tepekoylu C., Holfeld J., Feuchtner G., Egger A., Hoermann G., Schroll A., Fritsche G., Wildner S., Bellmann-Weiler R., Kirchmair R., Helbok R., Prosch H., Rieder D., Trajanoski Z., Kronenberg F., Woll E., Weiss G., Widmann G., Loffler-Ragg J., Tancevski I.. Cardiopulmonary recovery after COVID-19 - an observational prospective multi-center trial. The European respiratory journal. 2020; #volume#(#issue#): #pages#. doi: http://dx.doi.org/10.1183/13993003.03481-2020. Available at: http://ovidsp.ovid.com/ovidweb.cgi?T=JS&PAGE=reference&D=emexc&NEWS=N&AN=633749840 |
| 3 | Goertz Y.M.J., Van Herck M., Delbressine J.M., Vaes A.W., Meys R., Machado F.V.C., Houben-Wilke S., Burtin C., Posthuma R., Franssen F.M.E., van Loon N., Hajian B., Spies Y., Vijlbrief H., van 't Hul A.J., Janssen D.J.A., Spruit M.A.. Persistent symptoms 3 months after a SARS-CoV-2 infection: The post-COVID-19 syndrome?. ERJ Open Research. 2020; 6(4): 1. doi: http://dx.doi.org/10.1183/23120541.00542-2020. Available at: https://openres.ersjournals.com/content/erjor/6/4/00542-2020.full.pdf; http://ovidsp.ovid.com/ovidweb.cgi?T=JS&PAGE=reference&D=emexc&NEWS=N&AN=2005402902 |
| 4 | Tabatabaei S.M.H., Rajebi H., Moghaddas F., Ghasemiadl M., Talari H.. Chest CT in COVID-19 pneumonia: what are the findings in mid-term follow-up?. Emergency Radiology. 2020; 27(6): 711. doi: http://dx.doi.org/10.1007/s10140-020-01869-z. Available at: http://link.springer.de/link/service/journals/10140/index.htm; http://ovidsp.ovid.com/ovidweb.cgi?T=JS&PAGE=reference&D=emexc&NEWS=N&AN=2007175334 |

| | |
|---|---|
| 5 | Vaes A.W., Machado F.V.C., Meys R., Delbressine J.M., Goertz Y.M.J., Van Herck M., Houben-Wilke S., Franssen F.M.E., Vijlbrief H., Spies Y., Van 't Hul A.J., Burtin C., Janssen D.J.A., Spruit M.A.. Care dependency in non-hospitalized patients with COVID-19. Journal of Clinical Medicine. 2020; 9(9): 1. doi: http://dx.doi.org/10.3390/jcm9092946. Available at: https://www.mdpi.com/2077-0383/9/9/2946/pdf; http://ovidsp.ovid.com/ovidweb.cgi?T=JS&PAGE=reference&D=emexc&NEWS=N&AN=2005064026 |
| 6 | Ladds, Emma, Rushforth, Alex, Wieringa, Sietse, Taylor, Sharon, Rayner, Clare, Husain, Laiba, Greenhalgh, Trisha. Persistent symptoms after Covid-19: qualitative study of 114 "long Covid" patients and draft quality principles for services. BMC health services research. 2020; 20(1): 1144. doi: https://dx.doi.org/10.1186/s12913-020-06001-y. Available at: http://ovidsp.ovid.com/ovidweb.cgi?T=JS&PAGE=reference&D=medl&NEWS=N&AN=33342437 |
| 7 | Abdelalim A.A., Mohamady A.A., Elsayed R.A., Elawady M.A., Ghallab A.F.. Corticosteroid nasal spray for recovery of smell sensation in COVID-19 patients: A randomized controlled trial. American Journal of Otolaryngology - Head and Neck Medicine and Surgery. 2021; 42(2): 102884. doi: http://dx.doi.org/10.1016/j.amjoto.2020.102884. Available at: http://www.elsevier.com/inca/publications/store/6/2/3/1/1/9/index.htt; http://ovidsp.ovid.com/ovidweb.cgi?T=JS&PAGE=reference&D=emexc&NEWS=N&AN=2010584528 |
| 8 | Lanham, David, Roe, Jennifer, Chauhan, Alisha, Evans, Rebecca, Hillman, Toby, Logan, Sarah, Heightman, Melissa. COVID-19 emergency department discharges: an outcome study. Clinical medicine (London, England). 2021; #volume#(#issue#): #pages#. doi: https://dx.doi.org/10.7861/clinmed.2020-0817. Available at: http://ovidsp.ovid.com/ovidweb.cgi?T=JS&PAGE=reference&D=medp&NEWS=N&AN=33419864 |
| 9 | Myall KJ, Mukherjee B, Castanheira AM, Lam JL, Benedetti G, Mak SM, Preston R, Thillai M, Dewar A, Molyneaux PL, West,A.G.. Persistent Post-COVID-19 Inflammatory Interstitial Lung Disease: an Observational Study of Corticosteroid Treatment. Annals of the American Thoracic Society. 2021; #volume#(#issue#): #pages#. doi: 10.1513/AnnalsATS.202008-1002OC. Available at: https://pubmed.ncbi.nlm.nih.gov/33433263 |
| 10 | Group Luczak C, Seiler F, Monaury J, Martini A, Ward AB, Gracies JM, Motavasseli D, Covid Rehabilitation Study, Piquet V. Do Patients with Covid-19 Benefit from Rehabilitation? Functional outcomes of the first 100 patients in a Covid-19 rehabilitation unit; 7857995 CSR - Reports results. Archives of Physical Medicine and Rehabilitation. 2021///; #volume#(#issue#): #pages#. doi: 10.1016/j.apmr.2021.01.069. Available at: https://pubmed.ncbi.nlm.nih.gov/33548208 |
| 11 | Dennis, Andrea, Banerjee, Rajarshi, Wamil, Malgorzata, Alberts, Johann, Oben, Jude, Cuthbertson, Daniel J, Wootton, Dan, Crooks, Michael, Gabbay, Mark, Brady, Michael, Hishmeh, Lyth, Attree, Emily, Heightman, Melissa, Banerjee, Amitava. Multiorgan impairment in low-risk individuals with post-COVID-19 syndrome: A prospective, community-based study. BMJ Open. 2021///; 11(3): e048391. doi: http://dx.doi.org/10.1136/bmjopen-2020-048391. Available at: http://bmjopen.bmj.com/content/early/by/section,http://ovidsp.ovid.com/ovidweb.cgi?T=JS&PAGE=reference&D=emexb&NEWS=N&AN=634635621 |
| 12 | Remy-Jardin, Martine, Felloni, Paul, Faivre, Jean-Baptiste, Remy, Jacques, Duthoit, Louise, Fry, Stephanie, Bautin, Nathalie, Chenivesse, Cecile, Perez, Thierry, Duhamel, Alain. Assessment of pulmonary arterial circulation 3 months after hospitalization for SARS-CoV-2 pneumonia: Dual-energy CT (DECT) angiographic study in 55 patients. EClinicalMedicine. 2021///; 34(#issue#): 100778. doi: http://dx.doi.org/10.1016/j.eclinm.2021.100778. Available at: http://www.thelancet.com/journals/eclinm/issue/current,http://ovidsp.ovid.com/ovidweb.cgi?T=JS&PAGE=reference&D=emexb&NEWS=N&AN=2011531220 |
| 13 | Elkholi, Safaa Mostafa Ali, Abdelwahab, Mohamed Kamal, Abdelhafeez, Marwa. Impact of the smell loss on the quality of life and adopted coping strategies in COVID-19 patients. European Archives of Oto-Rhino-Laryngology. 2021///; #volume#(#issue#): #pages#. doi: http://dx.doi.org/10.1007/s00405-020-06575-7. Available at: http://link.springer.de/link/service/journals/00405/index.htm,http://ovidsp.ovid.com/ovidweb.cgi?T=JS&PAGE=reference&D=emexb&NEWS=N&AN=2010203497 |

| | |
|---|---|
| 14 | Le Bon, Serge-Daniel, Pisarski, Nathalie, Prunier, Lea, Lechien, Jerome R, Horoi, Mihaela, Konopnicki, Deborah. Efficacy and safety of oral corticosteroids and olfactory training in the management of COVID-19-related loss of smell. European Archives of Oto-Rhino-Laryngology. 2021///; #volume#(#issue#): #pages#. doi: http://dx.doi.org/10.1007/s00405-020-06520-8. Available at: http://link.springer.de/link/service/journals/00405/index.htm,http://ovidsp.ovid.com/ovidweb.cgi?T=JS&PAGE=reference&D=emexb&NEWS=N&AN=2010146553 |
| 15 | Delbressine, Jeannet M, Vaes, Anouk W, Houben-Wilke, Sarah, Meys, Roy, Goertz, Yvonne M J, Posthuma, Rein, Franssen, Frits M E, Machado, Felipe V C, Pitta, Fabio, van Herck, Maarten, Burtin, Chris, Spruit, Martijn A, Boon, Gudula J A M, Klok, Frederikus A, Barco, Stefano, van 't Hul, Alex, Spies, Yvonne, Vijlbrief, Herman, Rezek, Spencer A, Janssen, Daisy J A, Siegerink, Bob. Construct validity of the Post-COVID-19 Functional Status Scale in adult subjects with COVID-19. Health and Quality of Life Outcomes. 2021///; 19(1): 40. doi: http://dx.doi.org/10.1186/s12955-021-01691-2. Available at: http://www.hqlo.com/home/,http://ovidsp.ovid.com/ovidweb.cgi?T=JS&PAGE=reference&D=emexb&NEWS=N&AN=2010356913 |
| 16 | Ozer, Pelin Karaca, Govdeli, Elif Ayduk, Baykiz, Derya, Karaayvaz, Ekrem Bilal, Medetalibeyoglu, Alpay, Catma, Yunus, Elitok, Ali, Cagatay, Atahan, Umman, Berrin, Oncul, Aytac, Tukek, Tufan, Bugra, Zehra. Impairment of right ventricular longitudinal strain associated with severity of pneumonia in patients recovered from COVID-19. The international journal of cardiovascular imaging. 2021///; #volume#(#issue#): #pages#. doi: https://dx.doi.org/10.1007/s10554-021-02214-2. Available at: http://ovidsp.ovid.com/ovidweb.cgi?T=JS&PAGE=reference&D=medp&NEWS=N&AN=33839981 |
| 17 | Hall, Jocelin, Myall, Katherine, Lam, Jodie L, Mason, Thomas, Mukherjee, Bhashkar, West, Alex, Dewar, Amy. Identifying patients at risk of post-discharge complications related to COVID-19 infection. Thorax. 2021///; 76(4): 408. doi: http://dx.doi.org/10.1136/thoraxjnl-2020-215861. Available at: http://thorax.bmj.com/,http://ovidsp.ovid.com/ovidweb.cgi?T=JS&PAGE=reference&D=emexb&NEWS=N&AN=634494738 |
| 18 | Baricich, Alessio, Borg, Margherita B, Cuneo, Daria, Cisari, Carlo, Cadario, Emanuela, Azzolina, Danila, Balbo, Piero E, Bellan, Mattia, Pirisi, Mario, Zeppegno, Patrizia. Midterm functional sequelae and implications in rehabilitation after COVID19. A cross-sectional study. European journal of physical and rehabilitation medicine. 2021///; #volume#(#issue#): #pages#. doi: http://dx.doi.org/10.23736/S1973-9087.21.06699-5. Available at: http://ovidsp.ovid.com/ovidweb.cgi?T=JS&PAGE=reference&D=emexb&NEWS=N&AN=634256137 |
| 19 | Crisafulli, Ernesto, Gabbiani, Daniele, Magnani, Giulia, Busti, Fabiana, Sartori, Giulia, Girelli, Domenico, Dorelli, Gianluigi, Senna, Gianenrico. Residual lung function impairment is associated with hyperventilation in patients recovered from hospitalised covid-19: A cross-sectional study. Journal of Clinical Medicine. 2021///; 10(5): 1. doi: http://dx.doi.org/10.3390/jcm10051036. Available at: https://www.mdpi.com/2077-0383/10/5/1036/pdf,http://ovidsp.ovid.com/ovidweb.cgi?T=JS&PAGE=reference&D=emexb&NEWS=N&AN=2006134462 |
| 20 | Capelli, M, Gatti, P. Anosmia in the first coronavirus disease 2019 outbreak in Europe: Functional recovery after eight months. Journal of Laryngology and Otology. 2021///; 135(3): 224. doi: http://dx.doi.org/10.1017/S0022215121000670. Available at: http://journals.cambridge.org/action/displayJournal?jid=JLO,http://ovidsp.ovid.com/ovidweb.cgi?T=JS&PAGE=reference&D=emexb&NEWS=N&AN=634362302 |
| 21 | Smet, Jelle, Stylemans, Dimitri, Hanon, Shane, Ilsen, Bart, Verbanck, Sylvia, Vanderhelst, Eef. Clinical status and lung function 10 weeks after severe SARS-CoV-2 infection. Respiratory medicine. 2021///; 176(#issue#): 106276. doi: https://dx.doi.org/10.1016/j.rmed.2020.106276. Available at: http://ovidsp.ovid.com/ovidweb.cgi?T=JS&PAGE=reference&D=medl&NEWS=N&AN=33278758 |

**Study participants N<50 = 56**

| | |
|---|---|
| 1 | Vlachou M., Drebes A., Candilio L., Weeraman D., Mir N., Murch N., Davies N., Coghlan J.G.. Pulmonary thrombosis in Covid-19: before, during and after hospital admission. Journal of thrombosis and thrombolysis. 2021; #volume#(#issue#): #pages#. doi: http://dx.doi.org/10.1007/s11239-020-02370-7. Available at: http://www.wkap.nl/journalhome.htm/0929-5305; http://ovidsp.ovid.com/ovidweb.cgi?T=JS&PAGE=reference&D=emedx&NEWS=N&AN=2007708762 |
| 2 | Rafique N.B., Lal S., Mohammed Y.A., Aglan A.I.. The journey of a COVID-19 patient from admission to recovery and learning to breathe again. Anaesthesia, Pain and Intensive Care. 2020; 24(5): 558. doi: http://dx.doi.org/10.35975/APIC.V24I5.1365. Available at: http://apicareonline.com/index.php/APIC/article/view/1365; http://ovidsp.ovid.com/ovidweb.cgi?T=JS&PAGE=reference&D=emexc&NEWS=N&AN=2008509611 |
| 3 | Zheng H.-Y., Xu M., Yang C.-X., Tian R.-R., Zhang M., Li J.-J., Wang X.-C., Ding Z.-L., Li G.-M., Li X.-L., He Y.-Q., Dong X.-Q., Yao Y.-G., Zheng Y.-T.. Longitudinal transcriptome analyses show robust T cell immunity during recovery from COVID-19. Signal Transduction and Targeted Therapy. 2020; 5(1): 294. doi: http://dx.doi.org/10.1038/s41392-020-00457-4. Available at: http://www.nature.com/sigtrans; http://ovidsp.ovid.com/ovidweb.cgi?T=JS&PAGE=reference&D=emexc&NEWS=N&AN=2007668890 |
| 4 | Lineburg K.E., Srihari S., Altaf M., Swaminathan S., Panikkar A., Raju J., Crooks P., Ambalathingal G.R., Martins J.P., Matthews K.K., Neller M.A., Khanna R., Smith C.. Rapid detection of SARS-CoV-2-specific memory T-cell immunity in recovered COVID-19 cases. Clinical and Translational Immunology. 2020; 9(12): e1219. doi: http://dx.doi.org/10.1002/cti2.1219. Available at: https://onlinelibrary.wiley.com/loi/20500068; http://ovidsp.ovid.com/ovidweb.cgi?T=JS&PAGE=reference&D=emexc&NEWS=N&AN=2007667073 |
| 5 | Ortelli P., Ferrazzoli D., Sebastianelli L., Engl M., Romanello R., Nardone R., Bonini I., Koch G., Saltuari L., Quartarone A., Oliviero A., Kofler M., Versace V.. Neuropsychological and neurophysiological correlates of fatigue in post-acute patients with neurological manifestations of COVID-19: Insights into a challenging symptom. Journal of the neurological sciences. 2021; 420(#issue#): 117271. doi: http://dx.doi.org/10.1016/j.jns.2020.117271. Available at: http://www.elsevier.com/locate/jns; http://ovidsp.ovid.com/ovidweb.cgi?T=JS&PAGE=reference&D=emexc&NEWS=N&AN=2010409542 |
| 6 | Stahl K., Brasen J.H., Hoeper M.M., David S.. Direct evidence of SARS-CoV-2 in gut endothelium. Intensive care medicine. 2020; 46(11): 2081. doi: http://dx.doi.org/10.1007/s00134-020-06237-6. Available at: http://link.springer.de/link/service/journals/00134/index.htm; http://ovidsp.ovid.com/ovidweb.cgi?T=JS&PAGE=reference&D=emexc&NEWS=N&AN=2006128801 |
| 7 | Yasuda H., Tsukune Y., Watanabe N., Sugimoto K., Uchimura A., Tateyama M., Miyashita Y., Ochi Y., Komatsu N.. Persistent COVID-19 Pneumonia and Failure to Develop Anti-SARS-CoV-2 Antibodies During Rituximab Maintenance Therapy for Follicular Lymphoma. Clinical Lymphoma, Myeloma and Leukemia. 2020; 20(11): 774. doi: http://dx.doi.org/10.1016/j.clml.2020.08.017. Available at: http://www.sciencedirect.com/science/journal/21522650; http://ovidsp.ovid.com/ovidweb.cgi?T=JS&PAGE=reference&D=emexc&NEWS=N&AN=2007818876 |
| 8 | Dani M., Dirksen A., Taraborrelli P., Torocastro M., Panagopoulos D., Sutton R., Lim P.B.. Autonomic dysfunction in 'long COVID': rationale, physiology and management strategies. Clinical medicine (London, England). 2020; #volume#(#issue#): #pages#. doi: http://dx.doi.org/10.7861/clinmed.2020-0896. Available at: http://ovidsp.ovid.com/ovidweb.cgi?T=JS&PAGE=reference&D=emexc&NEWS=N&AN=633538217 |
| 9 | Cellai M., O'Keefe J.B.. Characterization of prolonged COVID-19 symptoms in an outpatient telemedicine clinic. Open Forum Infectious Diseases. 2020; 7(10): 1. doi: http://dx.doi.org/10.1093/ofid/ofaa420. Available at: https://academic.oup.com/ofid; http://ovidsp.ovid.com/ovidweb.cgi?T=JS&PAGE=reference&D=emexc&NEWS=N&AN=2010099734 |
| 10 | Elarabi A.M., Albakri M.M., Babiker M., Hussein M., Ahmed M., Mazhar R., Raza T.. A series of unfortunate events: Mind the windpipe. Clinical Case Reports. 2020; #volume#(#issue#): #pages#. doi: http://dx.doi.org/10.1002/ccr3.3484. Available at: http://onlinelibrary.wiley.com/journal/10.1002/(ISSN)2050-0904; http://ovidsp.ovid.com/ovidweb.cgi?T=JS&PAGE=reference&D=emexc&NEWS=N&AN=2007396073 |

| | |
|---|---|
| 11 | Law J.C., Koh W.H., Budylowski P., Lin J., Yue F., Abe K.T., Rathod B., Girard M., Li Z., Rini J.M., Mubareka S., McGeer A., Chan A.K., Gingras A.-C., Watts T.H., A Ostrowski, M.. Systematic Examination of Antigen-Specific Recall T Cell Responses to SARS-CoV-2 versus Influenza Virus Reveals a Distinct Inflammatory Profile. Journal of immunology (Baltimore, Md.: 1950). 2020; #volume#(#issue#): #pages#. doi: http://dx.doi.org/10.4049/jimmunol.2001067. Available at: http://ovidsp.ovid.com/ovidweb.cgi?T=JS&PAGE=reference&D=emexc&NEWS=N&AN=633464861 |
| 12 | Klocperk A., Bloomfield M., Parackova Z., Zentsova I., Vrabcova P., Balko J., Meseznikov G., Mendez L.F.C., Grandcourtova A., Sipek J., Tulach M., Zamecnik J., Vymazal T., Sediva A.. Complex immunometabolic profiling reveals the activation of cellular immunity and biliary lesions in patients with severe COVID-19. Journal of Clinical Medicine. 2020; 9(9): 1. doi: http://dx.doi.org/10.3390/jcm9093000. Available at: https://www.mdpi.com/2077-0383/9/9/3000/pdf; http://ovidsp.ovid.com/ovidweb.cgi?T=JS&PAGE=reference&D=emexc&NEWS=N&AN=2005087012 |
| 13 | Polistina G.E., Simioli F., Imitazione P., Lanza M., Annunziata A., Fiorentino G.. Different presentation of pulmonary parenchymal disruption in COVID-19 pneumonia. Case series of Sub-Intensive Care Unit in Naples, Italy. Monaldi Archives for Chest Disease. 2020; 90(4): 572. doi: http://dx.doi.org/10.4081/MONALDI.2020.1482. Available at: https://www.monaldi-archives.org/index.php/macd/article/view/1482; http://ovidsp.ovid.com/ovidweb.cgi?T=JS&PAGE=reference&D=emexc&NEWS=N&AN=2008009208 |
| 14 | Bezerra, Thamara Manoela, Feitosa, Sthefane Gomes, Carneiro, Diego Thiers Oliveira, Costa, Fabio Wildson Gurgel, Pires, Fabio Ramoa, Pereira, Karuza Maria Alves. Oral lesions in COVID-19 infection: Is long-term follow-up important in the affected patients?. Oral diseases. 2020; #volume#(#issue#): #pages#. doi: https://dx.doi.org/10.1111/odi.13705. Available at: http://ovidsp.ovid.com/ovidweb.cgi?T=JS&PAGE=reference&D=medp&NEWS=N&AN=33135278 |
| 15 | Doykov, Ivan, Hallqvist, Jenny, Gilmour, Kimberly C., Grandjean, Louis, Mills, Kevin, Heywood, Wendy E.. 'The long tail of Covid-19' - The detection of a prolonged inflammatory response after a SARS-CoV-2 infection in asymptomatic and mildly affected patients. F1000Research. 2020; 9(#issue#): 1349. doi: https://dx.doi.org/10.12688/f1000research.27287.1. Available at: http://ovidsp.ovid.com/ovidweb.cgi?T=JS&PAGE=reference&D=prem&NEWS=N&AN=33391730 |
| 16 | Olds, Hailey, Liu, Jesse, Luk, Kevin, Lim, Henry W., Ozog, David, Rambhatla, Pranita V.. Telogen Effluvium Associated With COVID-19 Infection. Dermatologic therapy. 2021; #volume#(#issue#): #pages#. doi: https://dx.doi.org/10.1111/dth.14761. Available at: http://ovidsp.ovid.com/ovidweb.cgi?T=JS&PAGE=reference&D=medp&NEWS=N&AN=33405302 |
| 17 | Piper-Vallillo, Andrew, Mooradian, Meghan J., Meador, Catherine B., Yeap, Beow Y., Peterson, Jennifer, Sakhi, Mustafa, Do, Andrew, Zubiri, Leyre, Stevens, Sara, Vaughn, Jeanne, Goodwin, Kelly, Gavralidis, Alexander, Willers, Henning, Miller, Adam, Farago, Anna, Piotrowska, Zofia, Lin, Jessica J., Dagogo-Jack, Ibiayi, Lennes, Inga T., Sequist, Lecia V., Temel, Jennifer S., Heist, Rebecca S., Digumarthy, Subba, Reynolds, Kerry L., Gainor, Justin F.. Coronavirus Disease 2019 Infection in a Patient Population with Lung Cancer: Incidence, Presentation, and Alternative Diagnostic Considerations. JTO clinical and research reports. 2021; 2(1): 100124. doi: https://dx.doi.org/10.1016/j.jtocrr.2020.100124. Available at: http://ovidsp.ovid.com/ovidweb.cgi?T=JS&PAGE=reference&D=prem&NEWS=N&AN=33205053 |
| 18 | Klingler J., Weiss S., Itri V., Liu X., Oguntuyo K.Y., Stevens C., Ikegame S., Hung C.-T., Enyindah-Asonye G., Amanat F., Baine I., Arinsburg S., Bandres J.C., Kojic E.M., Stoever J., Jurczyszak D., Bermudez-Gonzalez M., Nadas A., Liu S., Lee B., Zolla-Pazner S., Hioe C.E.. Role of IgM and IgA Antibodies in the Neutralization of SARS-CoV-2. The Journal of infectious diseases. 2020; #volume#(#issue#): #pages#. doi: http://dx.doi.org/10.1093/infdis/jiaa784. Available at: http://ovidsp.ovid.com/ovidweb.cgi?T=JS&PAGE=reference&D=emexc&NEWS=N&AN=633884331 |
| 19 | Smane L., Stars I., Pucuka Z., Roge I., Pavare J.. Persistent clinical features in paediatric patients after SARS-CoV-2 virological recovery: A retrospective population-based cohort study from a single centre in Latvia. BMJ Paediatrics Open. 2020; 4(1): e000905. doi: http://dx.doi.org/10.1136/bmjpo-2020-000905. Available at: http://bmjpaedsopen.bmj.com/; http://ovidsp.ovid.com/ovidweb.cgi?T=JS&PAGE=reference&D=emexc&NEWS=N&AN=633774929 |

| | |
|---|---|
| 20 | Bozzani, Antonio, Arici, Vittorio, Tavazzi, Guido, Mojoli, Francesco, Bruno, Raffaele, Sterpetti, Antonio V., Ragni, Franco. Acute Thrombosis of Lower Limbs Arteries in The Acute Phase and after Recovery from COVID19. Annals of Surgery. 2021; Publish Ahead of Print(#issue#): #pages#. doi: https://dx.doi.org/10.1097/SLA.0000000000004700. Available at: http://ovidsp.ovid.com/ovidweb.cgi?T=JS&PAGE=reference&D=medp&NEWS=N&AN=33417332 |
| 21 | A,Rogliani P, Calzetta L, Coppola A, Puxeddu E, Sergiacomi G, D'Amato D, Orlacchio. Are there pulmonary sequelae in patients recovering from COVID-19?; 7598236 CSR - Reports results. Respiratory research. 2020; 21(1): 286. doi: 10.1186/s12931-020-01550-6. Available at: https://pubmed.ncbi.nlm.nih.gov/33126869 |
| 22 | Ruano R, Zorzano-Martinez M, Campos A, Rius F, Hernandez,M.. Subacute thyroiditis might be a complication triggered by SARS-CoV-2; 7553120 CSR - Reports results. Endocrinologia, diabetes y nutricion. 2020; #volume#(#issue#): #pages#. doi: 10.1016/j.endinu.2020.09.002. Available at: https://pubmed.ncbi.nlm.nih.gov/33139217 |
| 23 | Schellekens MMI, Bleeker-Rovers CP, Keurlings PAJ, Mummery CJ, Bloem,B.R.. Reversible Myoclonus-Ataxia as a Postinfectious Manifestation of COVID-19; 7604679 CSR - Reports results. Movement disorders clinical practice. 2020; 7(8): 977. doi: 10.1002/mdc3.13088. Available at: https://pubmed.ncbi.nlm.nih.gov/33163570 |
| 24 | A Leggieri C, Fominskiy E, Scandroglio AM, Colombo S, Tozzi M, Moizo E, Mucci M, Crivellari M, Pieri M, Guzzo F, Piemontese S, De Lorenzo R, Da Prat V, Fedrizzi M, Faustini C, Di Piazza M, Conte F, Lembo R, Esposito A, Dagna L, Landoni G, Zangrillo, Monti G. Two months quality of life of COVID-19 invasively ventilated survivors; an Italian single-center study. Acta Anaesthesiologica Scandinavica. 2021///; #volume#(#issue#): #pages#. doi: 10.1111/aas.13812. Available at: https://pubmed.ncbi.nlm.nih.gov/33655487 |
| 25 | Murad, Haris, Dubberke, Erik, Mattu, Munis, Parikh, Bijal, Wellen, Jason, Alhamad, Tarek. Repeat SARS-CoV-2 testing after recovery. Is a pretransplant PCR necessary?. American journal of transplantation : official journal of the American Society of Transplantation and the American Society of Transplant Surgeons. 2021///; #volume#(#issue#): #pages#. doi: https://dx.doi.org/10.1111/ajt.16506. Available at: http://ovidsp.ovid.com/ovidweb.cgi?T=JS&PAGE=reference&D=medp&NEWS=N&AN=33512073 |
| 26 | A Gottimukkala K, Kumar S, Reddy ES, Edara VV, Kauffman R, Floyd K, Mantus G, Savargaonkar D, Goel PK, Arora S, Rahi M, Davis CW, Linderman S, Wrammert J, Suthar M, Ahmed R, Sharma A, Murali-Krishna K, Chandele, Nayak K. Characterization of neutralizing versus binding antibodies and memory B cells in COVID-19 recovered individuals from India (preprint). Biorxiv. 2020///; #volume#(#issue#): 2020.08.31.276675. doi: 10.1101/2020.08.31.276675. Available at: http://biorxiv.org/content/early/2020/09/01/2020.08.31.276675.abstract |
| 27 | DW Zhao M, Li C, Zhang Y, Wang, Wu J. The SARS-CoV-2 induced targeted amino acid profiling in patients at hospitalized and convalescent stage. Bioscience reports. 2021///; #volume#(#issue#): #pages#. doi: 10.1042/BSR20204201. Available at: https://pubmed.ncbi.nlm.nih.gov/33625490 |
| 28 | Chioh, Florence W J, Wu, Kan-Xing, Siau, Anthony, Chan, Yi-Hao, Carissimo, Guillaume, Renia, Laurent, Ng, Lisa F P, Fong, Siew-Wai, Young, Barnaby E, Krishnan, Shuba, Teo, Louis L Y, Teo, Louis L Y, Gao, Fei, Tan, Ru San, Zhong, Liang, Koh, Angela S, Tan, Seow-Yen, Tambyah, Paul A, Lye, David C, Cheung, Christine. Convalescent covid-19 patients are susceptible to endothelial dysfunction due to persistent immune activation. eLife. 2021///; 10(#issue#): e64909. doi: http://dx.doi.org/10.7554/eLife.64909. Available at: https://elifesciences.org/articles/64909,http://ovidsp.ovid.com/ovidweb.cgi?T=JS&PAGE=reference&D=emexb&NEWS=N&AN=2006866526 |
| 29 | C Campion JY, Dudouet P, Kaphan E, Bregeon F, Tissot-Dupont H, Guis S, Barthelemy F, Habert P, Ceccaldi M, Million M, Raoult D, Cammilleri S, Eldin, Guedj E. 18F-FDG brain PET hypometabolism in patients with long COVID; 7837643 CSR - Reports results. European journal of nuclear medicine and molecular imaging. 2021///; #volume#(#issue#): #pages#. doi: 10.1007/s00259-021-05215-4. Available at: https://pubmed.ncbi.nlm.nih.gov/33501506 |
| 30 | Ciccarelli, Michele, Sollini, Martina, Cecconi, Maurizio, Aghemo, Alessio, Morelli, Paola, Chiola, Silvia, Gelardi, Fabrizia, Chiti, Arturo, Morbelli, Silvia. Long COVID hallmarks on [18F]FDG-PET/CT: a case-control |

| | |
|---|---|
| | study. European Journal of Nuclear Medicine and Molecular Imaging. 2021///; #volume#(#issue#): #pages#. doi: http://dx.doi.org/10.1007/s00259-021-05294-3. Available at: http://link.springer.com/journal/volumesAndIssues/259,http://ovidsp.ovid.com/ovidweb.cgi?T=JS&PAGE=reference&D=emexb&NEWS=N&AN=2010720709 |
| 31 | Ansari, Asgar, Sachan, Shilpa, Jha, Someshwar Nath, Kalia, Anurag, Gupta, Nimesh, Arya, Rakesh, Sharma, Ashok, Lall, Anupam, Coshic, Poonam, Grifoni, Alba, Weiskopf, Daniela, Sette, Alessandro. Immune Memory in Mild COVID-19 Patients and Unexposed Donors Reveals Persistent T Cell Responses After SARS-CoV-2 Infection. Frontiers in Immunology. 2021///; 12(#issue#): 636768. doi: http://dx.doi.org/10.3389/fimmu.2021.636768. Available at: https://www.frontiersin.org/journals/immunology#,http://ovidsp.ovid.com/ovidweb.cgi?T=JS&PAGE=reference&D=emexb&NEWS=N&AN=634567289 |
| 32 | Riou, Marianne, Momas, Cedric, Rouyer, Olivier, Lebourg, Fabienne, Meyer, Alain, Enache, Irina, Pistea, Cristina, Charloux, Anne, Talha, Samy, Geny, Bernard, Oulehri, Walid, Collange, Olivier, Mertes, Michel, Marcot, Christophe, Blay, Frederic de, Andres, Emmanuel. Reduced flow-mediated dilatation is not related to covid-19 severity three months after hospitalization for sars-cov-2 infection. Journal of Clinical Medicine. 2021///; 10(6): 1318. doi: http://dx.doi.org/10.3390/jcm10061318. Available at: https://www.mdpi.com/2077-0383/10/6/1318/pdf,http://ovidsp.ovid.com/ovidweb.cgi?T=JS&PAGE=reference&D=emexb&NEWS=N&AN=2006795239 |
| 33 | Susanto, Agus Dwi, Triyoga, Paulus Arka, Isbaniah, Fathiyah, Fairuz, Aulya, Cendikiawan, Hans, Zaron, Ferdynand, Aryanti, Intan, Irbah, Sarah Naura, Hidayat, Moulid. Lung Fibrosis Sequelae After Recovery from COVID-19 Infection. Journal of infection in developing countries. 2021///; 15(3): 360. doi: https://dx.doi.org/10.3855/jidc.13686. Available at: http://ovidsp.ovid.com/ovidweb.cgi?T=JS&PAGE=reference&D=prem&NEWS=N&AN=33839710 |
| 34 | Kant, Sam, Raman, Gaurav, Damera, Pranav, Antiochos, Brendan, Seo, Philip, Geetha, Duvuru. Characteristics and Outcomes of COVID-19 in Patients With Antineutrophil Cytoplasmic Antibody-Associated Vasculitis. Kidney International Reports. 2021///; 6(3): 806. doi: http://dx.doi.org/10.1016/j.ekir.2020.12.024. Available at: http://www.journals.elsevier.com/kidney-international-reports,http://ovidsp.ovid.com/ovidweb.cgi?T=JS&PAGE=reference&D=emexb&NEWS=N&AN=2011122695 |
| 35 | Mohr, Arno, Dannerbeck, Laura, Lange, Tobias J, Pfeifer, Michael, Blaas, Stefan, Salzberger, Bernd, Hitzenbichler, Florian, Koch, Myriam. Cardiopulmonary exercise pattern in patients with persistent dyspnoea after recovery from COVID-19. Multidisciplinary respiratory medicine. 2021///; 16(1): 732. doi: https://dx.doi.org/10.4081/mrm.2021.732. Available at: http://ovidsp.ovid.com/ovidweb.cgi?T=JS&PAGE=reference&D=pmnm&NEWS=N&AN=33623700 |
| 36 | M Prior M, Meneghetti L, Mercogliano T, Bendini, Priftis K. Alexia without agraphia in a post COVID-19 patient with left-hemisphere ischemic stroke; 7994060 CSR - Reports results. Neurological sciences. 2021///; #volume#(#issue#): #pages#. doi: 10.1007/s10072-021-05211-4. Available at: https://pubmed.ncbi.nlm.nih.gov/33768435 |
| 37 | Gaughan, Maria, Connolly, Sean, O'Riordan, Sean, Tubridy, Niall, McGuigan, Chris, Kinsella, Justin A. Pediatric Parainfectious Encephalitis Associated With COVID-19. Neurology. 2021///; 96(11): 541. doi: http://dx.doi.org/10.1212/WNL.0000000000011476. Available at: http://ovidsp.ovid.com/ovidweb.cgi?T=JS&PAGE=reference&D=emexb&NEWS=N&AN=633933760 |
| 38 | D'Ascanio, Luca, Pandolfini, Manlio, Cingolani, Cristina, Latini, Gino, Gradoni, Paolo, Capalbo, Maria, Frausini, Gabriele, Maranzano, Massimo, Brenner, Michael J, Di Stadio, Arianna. Olfactory Dysfunction in COVID-19 Patients: Prevalence and Prognosis for Recovering Sense of Smell. Otolaryngology--head and neck surgery : official journal of American Academy of Otolaryngology-Head and Neck Surgery. 2021///; 164(1): 82. doi: https://dx.doi.org/10.1177/0194599820943530. Available at: http://ovidsp.ovid.com/ovidweb.cgi?T=JS&PAGE=reference&D=medl&NEWS=N&AN=32662745 |

| | |
|---|---|
| 39 | Newell, Krista L, Clemmer, Deanna C, Cox, Justin B, Kayode, Yetunde I, Zoccoli-Rodriguez, Victoria, Taylor, Harry E, Wilmore, Joel R, Winslow, Gary M, Endy, Timothy P. Switched and unswitched memory B cells detected during SARS-CoV-2 convalescence correlate with limited symptom duration. PLoS ONE. 2021///; 16(1): e0244855. doi: http://dx.doi.org/10.1371/journal.pone.0244855. Available at: https://journals.plos.org/plosone/article/file?id=10.1371/journal.pone.0244855&type=printable,http://ovidsp.ovid.com/ovidweb.cgi?T=JS&PAGE=reference&D=emexb&NEWS=N&AN=2010910457 |
| 40 | LSS Jabeen T, Khader MA, Rao, Mannur S. POST-COVID-19 ASSOCIATED DECLINE IN LONG-TERM MALE FERTILITY AND EMBRYO QUALITY DURING ASSISTED REPRODUCTIVE TECHNOLOGY. QJM : monthly journal of the Association of Physicians. 2021///; #volume#(#issue#): #pages#. doi: 10.1093/qjmed/hcab019. Available at: https://pubmed.ncbi.nlm.nih.gov/33508136 CSR - Reports results |
| 41 | J Yudd M, Dreisbach AW, Michaud, Parpas A. Post COVID 19 multisystem inflammatory syndrome in an older adult. Renal failure. 2021///; 43(1): 530. doi: 10.1080/0886022X.2021.1895839. Available at: https://pubmed.ncbi.nlm.nih.gov/33719897 CSR - Reports results |
| 42 | M Demey L, Bruyneel AV, Roman A, Alard S, De Vos N, Bruyneel, Truffaut L. Post-discharge critical COVID-19 lung function related to severity of radiologic lung involvement at admission; 7819622 CSR - Reports results. Respiratory research. 2021///; 22(1): 29. doi: 10.1186/s12931-021-01625-y. Available at: https://pubmed.ncbi.nlm.nih.gov/33478527 |
| 43 | Jayathilaka, Deshni, Gomes, Laksiri, Jeewandara, Chandima, Wijewickrama, Ananda, Narangoda, Eranga, Idampitiya, Damayanthi, Guruge, Dinuka, Wijayamuni, Ruwan, Manilgama, Suranga, Ogg, Graham S, Malavige, Gathsaurie Neelika, Tan, Chee Wah, Wang, Lin-Fa. SARS-CoV-2 neutralizing antibodies in patients with varying severity of acute COVID-19 illness. Scientific reports. 2021///; 11(1): 2062. doi: http://dx.doi.org/10.1038/s41598-021-81629-2. Available at: http://ovidsp.ovid.com/ovidweb.cgi?T=JS&PAGE=reference&D=emexb&NEWS=N&AN=634091253 |
| 44 | Villanueva, Carolina, Enos, Daniel, Vasquez, Jaime, Sanhueza, Felipe, Melo, Pedro, Labarca, Gonzalo, Llerena, Faryd, Castillo, Daniela, Fraga, Marco, Lamperti, Liliana, Ormazabal, Valeska, Nova-Lamperti, Estefania, Henriquez-Beltran, Mario, Erices, Gustavo, Rubilar, Daniel, Sanhueza, Rocio, Lastra, Jaime, Horta, Gloria, Riffo, Benilde, Dreyse, Jorge, Jorquera, Jorge, Fernandez-Bussy, Sebastian, Gonzalez, Jessica, Barbe, Ferran. Undiagnosed sleep disorder breathing as a risk factor for critical COVID-19 and pulmonary consequences at the midterm follow-up. Sleep medicine. 2021///; #volume#(#issue#): #pages#. doi: http://dx.doi.org/10.1016/j.sleep.2021.02.029. Available at: http://www.elsevier.com/inca/publications/store/6/2/0/2/8/2,http://ovidsp.ovid.com/ovidweb.cgi?T=JS&PAGE=reference&D=emexb&NEWS=N&AN=2011248306 |
| 45 | Sharma, S, Grover, M, Bhargava, S, Samdani, S, Kataria, T. Post coronavirus disease mucormycosis: a deadly addition to the pandemic spectrum. The Journal of laryngology and otology. 2021///; #volume#(#issue#): 1. doi: https://dx.doi.org/10.1017/S0022215121000992. Available at: http://ovidsp.ovid.com/ovidweb.cgi?T=JS&PAGE=reference&D=medp&NEWS=N&AN=33827722 |
| 46 | Evert, Katja, Brochhausen, Christoph, Keil, Felix, Trummer, Matthias, Scheiter, Alexander, Calvisi, Diego F, Evert, Matthias, Dienemann, Thomas, Ritzka, Markus, Lunz, Dirk, Lubnow, Matthias, Salzberger, Bernd, Reischl, Udo, Gessner, Andre, Jantsch, Jonathan, Schmidt, Barbara, Simon, Michaela, Boor, Peter. Autopsy findings after long-term treatment of COVID-19 patients with microbiological correlation. Virchows Archiv. 2021///; #volume#(#issue#): #pages#. doi: http://dx.doi.org/10.1007/s00428-020-03014-0. Available at: http://link.springer.de/link/service/journals/00428/index.htm,http://ovidsp.ovid.com/ovidweb.cgi?T=JS&PAGE=reference&D=emexb&NEWS=N&AN=2010214772 |
| 47 | Lv H., Zhang W., Zhu Z., Xiong Q., Xiang R., Wang Y., Shi W., Deng Z., Xu Y.. Prevalence and recovery time of olfactory and gustatory dysfunction in hospitalized patients with COVID-19 in Wuhan, China. International Journal of Infectious Diseases. 2020; 100(#issue#): 507. doi: http://dx.doi.org/10.1016/j.ijid.2020.09.039. Available at: https://www.journals.elsevier.com/international-journal-of-infectious-diseases; http://ovidsp.ovid.com/ovidweb.cgi?T=JS&PAGE=reference&D=emexc&NEWS=N&AN=2008374139 |

| | |
|---|---|
| 48 | Zhao G., Su Y., Sun X., Cui X., Dang L., Zhao L., Tan X., Wang H., Yang M.. A comparative study of the laboratory features of COVID-19 and other viral pneumonias in the recovery stage. Journal of clinical laboratory analysis. 2020; 34(10): e23483. doi: http://dx.doi.org/10.1002/jcla.23483. Available at: http://onlinelibrary.wiley.com/journal/10.1002/(ISSN)1098-2825; http://ovidsp.ovid.com/ovidweb.cgi?T=JS&PAGE=reference&D=emexc&NEWS=N&AN=2005640800 |
| 49 | An Y.-W., Song S., Li W.-X., Chen Y.-X., Hu X.-P., Zhao J., Li Z.-W., Jiang G.-Y., Wang C., Wang J.-C., Yuan B., Liu H.-Q.. Liver function recovery of COVID-19 patients after discharge, a follow-up study. International Journal of Medical Sciences. 2021; 18(1): 176. doi: http://dx.doi.org/10.7150/ijms.50691. Available at: https://www.medsci.org/v18p0176.pdf; http://ovidsp.ovid.com/ovidweb.cgi?T=JS&PAGE=reference&D=emexc&NEWS=N&AN=2005414507 |
| 50 | Mcloughlin B.C., Miles A., Webb T.E., Knopp P., Eyres C., Fabbri A., Humphries F., Davis D.. Functional and cognitive outcomes after COVID-19 delirium. European Geriatric Medicine. 2020; 11(5): 857. doi: http://dx.doi.org/10.1007/s41999-020-00353-8. Available at: http://www.springer.com/journal/41999; http://ovidsp.ovid.com/ovidweb.cgi?T=JS&PAGE=reference&D=emexc&NEWS=N&AN=2005572908 |
| 51 | Fjaeldstad, A. W.. Prolonged complaint s of chemosensory loss aft er covid-19; 32741438. Danish Medical Journal. 2020; 67(8): 1. doi: #DOI#. Available at: #URL# |
| 52 | Liu, Kai, Zhang, Weitong, Yang, Yadong, Zhang, Jinpeng, Li, Yunqian, Chen, Ying. Respiratory rehabilitation in elderly patients with COVID-19: A randomized controlled study. Complementary Therapies in Clinical Practice. 2020; 39(#issue#): 101166. doi: https://doi.org/10.1016/j.ctcp.2020.101166. Available at: http://www.sciencedirect.com/science/article/pii/S1744388120304278 |
| 53 | Locatello, Luca Giovanni, Bruno, Chiara, Maggiore, Giandomenico, Cilona, Maria, Orlando, Pietro, Fancello, Giuseppe, Piccica, Matteo, Vellere, Iacopo, Lagi, Filippo, Trotta, Michele, Gallo, Oreste. The prognostic role of IL-10 in non-severe COVID-19 with chemosensory dysfunction. Cytokine. 2021///; 141(#issue#): 155456. doi: https://dx.doi.org/10.1016/j.cyto.2021.155456. Available at: http://ovidsp.ovid.com/ovidweb.cgi?T=JS&PAGE=reference&D=medl&NEWS=N&AN=33561690 |
| 54 | Soares, Felipe Henriques Carvalho, Kubota, Gabriel Taricani, Fernandes, Ana Mercia, Hojo, Bruno, Couras, Catarina, Costa, Barbara Venturoti, da Silva Lapa, Jorge Dornellys, Braga, Luiza Mansur, de Almeida, Matheus Merula, da Cunha Henrique Martins, Pedro, Pereira, Vitor Hugo Honorato, de Morais Donizeth Silva, Adriano, Teixeira, Manoel Jacobsen, Ciampi de Andrade, Daniel. Prevalence and characteristics of new-onset pain in COVID-19 survivors, a controlled study. European journal of pain (London, England). 2021///; #volume#(#issue#): #pages#. doi: http://dx.doi.org/10.1002/ejp.1755. Available at: http://ovidsp.ovid.com/ovidweb.cgi?T=JS&PAGE=reference&D=emexb&NEWS=N&AN=634395725 |
| 55 | Ramakrishnan, Aditi, Zreloff, Jennifer, Moore, Miranda A, Bergquist, Sharon H, Cellai, Michele, Higdon, Jason, O'Keefe B., James, Roberts, David, Wu, Henry M. Prolonged Symptoms After COVID-19 Infection in Outpatients. Open forum infectious diseases. 2021///; 8(3): ofab060. doi: https://dx.doi.org/10.1093/ofid/ofab060. Available at: http://ovidsp.ovid.com/ovidweb.cgi?T=JS&PAGE=reference&D=pmnm&NEWS=N&AN=33732751 |
| 56 | Malik, Jahanzeb, Javaid, Muhammad, Zahid, Tayyaba, Malik, Asmara, Ishaq, Uzma, Shoaib, Muhammad. Thyroid function analysis in COVID-19: A retrospective study from a single center. PLoS ONE. 2021///; 16(3): e0249421. doi: http://dx.doi.org/10.1371/journal.pone.0249421. Available at: https://journals.plos.org/plosone/article/file?id=10.1371/journal.pone.0249421&type=printable,http://ovidsp.ovid.com/ovidweb.cgi?T=JS&PAGE=reference&D=emexb&NEWS=N&AN=2011603247 |

**Language not English or French = 32**

| | |
|---|---|
| 1 | Araujo J.M., Branco M., Machado A.. Cognitive defect and covid-19. Sinapse. 2020; 20(2): 17. doi: http://dx.doi.org/10.46531/sinapse/AR/COVID19/AraujoJM/2020. Available at: http://www.spneurologia.com; http://ovidsp.ovid.com/ovidweb.cgi?T=JS&PAGE=reference&D=emexc&NEWS=N&AN=2005631228 |

| | |
|---|---|
| 2 | Nogueira Lopez J., Grasa Lozano C., Ots Ruiz C., Alonso Garcia L., Falces-Romero I., Calvo C., Garcia-Lopez Hortelano M.. Telemedicine follow-ups for COVID-19: Experience in a tertiary hospital. Anales de Pediatria. 2020; #volume#(#issue#): #pages#. doi: http://dx.doi.org/10.1016/j.anpedi.2020.10.017. Available at: http://www.doyma.es/anpediatr/; http://ovidsp.ovid.com/ovidweb.cgi?T=JS&PAGE=reference&D=emexc&NEWS=N&AN=2010332536 |
| 3 | Thede C.. COVID-19-therapeutic options using chinese herbal medicine in the acute phase and convalescence. Deutsche Zeitschrift fur Akupunktur. 2020; 63(4): 208. doi: http://dx.doi.org/10.1007/s42212-020-00316-x. Available at: http://www.springer.com/journal/42212; http://ovidsp.ovid.com/ovidweb.cgi?T=JS&PAGE=reference&D=emexc&NEWS=N&AN=2007020557 |
| 4 | Grannemann J.J., Roper A.. Travelling to High Altitude Destinations after Recovery from COVID-19-infection: New Aspects of Medical Advice in Altitude Medicine. Pneumologie. 2020; #volume#(#issue#): #pages#. doi: http://dx.doi.org/10.1055/a-1239-0334. Available at: http://www.thieme-connect.com/ejournals/toc/pneumologie; http://ovidsp.ovid.com/ovidweb.cgi?T=JS&PAGE=reference&D=emexc&NEWS=N&AN=633397458 |
| 5 | Wan X.-Y., Meng X.-Z., Li J.-C., Gong X.-L., Liang Y.-Q., Gao S.-K., Xu J.-P., Yue X.-Q.. Clinical effect of Guanggu Jisheng decoction in treatment of recovery stage coronavirus disease 2019. Academic Journal of Second Military Medical University. 2020; 41(7): 813. doi: http://dx.doi.org/10.16781/j.0258-879x.2020.07.0697. Available at: http://www.ajsmmu.cn/ajsmmu/ch/reader/view_abstract.aspx?file_no=20200886&flag=1; http://ovidsp.ovid.com/ovidweb.cgi?T=JS&PAGE=reference&D=emexc&NEWS=N&AN=2008018568 |
| 6 | Ros Dopico L., Tung-Chen Y., Pilares Barco M., Munoz Garcia A.. Monitoring of the rehabilitation therapy of COVID-19 effort dyspnea. Enfermedades infecciosas y microbiologia clinica. 2020; #volume#(#issue#): #pages#. doi: http://dx.doi.org/10.1016/j.eimc.2020.08.006. Available at: https://www.journals.elsevier.com/enfermedades-infecciosas-y-microbiologia-clinica; http://ovidsp.ovid.com/ovidweb.cgi?T=JS&PAGE=reference&D=emexc&NEWS=N&AN=2008475924 |
| 7 | Lindholm E., Tonnessen T.I.. Prolonged illness after COVID-19. Tidsskrift for den Norske laegeforening : tidsskrift for praktisk medicin, ny raekke. 2020; 140(14): #pages#. doi: http://dx.doi.org/10.4045/tidsskr.20.0753. Available at: http://ovidsp.ovid.com/ovidweb.cgi?T=JS&PAGE=reference&D=emexc&NEWS=N&AN=633201402 |
| 8 | Burgers J.S., Reijers M.H., Cals J.W.L.. Persisting symptoms after uncomplicated COVID-19: Dealing with uncertainty by general practitioner and patient. Nederlands tijdschrift voor geneeskunde. 2020; 164(40): A27. doi: #DOI#. Available at: https://www.ntvg.nl/artikelen/langdurige-klachten-na-ongecompliceerde-covid-19; http://ovidsp.ovid.com/ovidweb.cgi?T=JS&PAGE=reference&D=emexc&NEWS=N&AN=2008328882 |
| 9 | Tveito K.. #longcovid. Tidsskrift for den Norske laegeforening : tidsskrift for praktisk medicin, ny raekke. 2020; 140(14): #pages#. doi: http://dx.doi.org/10.4045/tidsskr.20.0779. Available at: http://ovidsp.ovid.com/ovidweb.cgi?T=JS&PAGE=reference&D=emexc&NEWS=N&AN=633203465 |
| 10 | Coluccia A.. Covid-19: The implications for patients with haemophilia and congenital haemorrhagic diseases. Blood Transfusion. 2020; 18(#issue#): s315. doi: http://dx.doi.org/10.2450/2020.S3. Available at: http://www.bloodtransfusion.it/scarica.aspx?tipo=A&id=003329&riv=000155; http://ovidsp.ovid.com/ovidweb.cgi?T=JS&PAGE=reference&D=emexc&NEWS=N&AN=633628723 |
| 11 | Holtslag, H. R., van den Borst, B., Reijers, M. H. E., Dettling, D. S.. [Post-COVID-19 rehabilitation; a matter of customisation]. Nederlands tijdschrift voor geneeskunde. 2020; 164(#issue#): #pages#. doi: #DOI#. Available at: http://ovidsp.ovid.com/ovidweb.cgi?T=JS&PAGE=reference&D=prem&NEWS=N&AN=33331730 |
| 12 | Maev, I. V., Shpektor, A. V., Vasilyeva, E. Y., Manchurov, V. N., Andreev, D. N.. [Novel coronavirus infection COVID-19: extrapulmonary manifestations]. Terapevticheskii arkhiv. 2020; 92(8): 4. doi: https://dx.doi.org/10.26442/00403660.2020.08.000767. Available at: http://ovidsp.ovid.com/ovidweb.cgi?T=JS&PAGE=reference&D=medl&NEWS=N&AN=33346454 |
| 13 | Belkind M.A., Gaman S.A., Stukalova O.V., Ternovoy S.K.. Dynamics of changes in the acute period and long-term results of chest CT in patients with COVID-19 pneumonia. Russian Electronic Journal of |

| | |
|---|---|
| | Radiology. 2021; 10(4): 47. doi: http://dx.doi.org/10.21569/2222-7415-2020-10-4-47-59. Available at: http://rejr.ru/volume/40/4.pdf; http://ovidsp.ovid.com/ovidweb.cgi?T=JS&PAGE=reference&D=emexc&NEWS=N&AN=2010542129 |
| 14 | Landsman J.A., Verheij N.P., Alma M.A., van den Boogaard J., Luning-Koster M., Evenboer K.E., van der Mei S.F., Reijneveld S.A.. COVID-19: recovering at home is not easy. Nederlands tijdschrift voor geneeskunde. 2020; 164(#issue#): #pages#. doi: #DOI#. Available at: http://ovidsp.ovid.com/ovidweb.cgi?T=JS&PAGE=reference&D=emexc&NEWS=N&AN=633826594 |
| 15 | Alvarez Martin MC, Del Peso Gilsanz C, Hernandez Lopez,A.. Subacute De Quervain thyroiditis after SARS-CoV-2 infection; 7709717 CSR - Reports results. Endocrinologia, diabetes y nutricion. 2020; #volume#(#issue#): #pages#. doi: 10.1016/j.endinu.2020.10.003. Available at: https://pubmed.ncbi.nlm.nih.gov/33358487 |
| 16 | Skripuletz, Thomas, Mohn, Nora, Franke, Christiana, Prus, Harald. [Neuroimmunology of COVID-19]. Der Nervenarzt. 2021///; #volume#(#issue#): #pages#. doi: https://dx.doi.org/10.1007/s00115-021-01077-1. Available at: http://ovidsp.ovid.com/ovidweb.cgi?T=JS&PAGE=reference&D=medp&NEWS=N&AN=33651117 |
| 17 | Feldmeier, Hermann. Mysterious neuro-COVID: Causes of persistent neurological symptoms are unresolved. Deutsche Apotheker Zeitung. 2021///; 161(10): #pages#. doi: #DOI#. Available at: https://www.deutsche-apotheker-zeitung.de/daz-az/2021/daz-10-2021/raetselhaftes-neuro-covid,http://ovidsp.ovid.com/ovidweb.cgi?T=JS&PAGE=reference&D=emexb&NEWS=N&AN=2011316505 |
| 18 | Schlenger, Ralf. Recovered - but not healthy: Symptoms of "long COVID" burden many patients long after the acute infection. Deutsche Apotheker Zeitung. 2020///; 160(52): A17. doi: #DOI#. Available at: https://www.deutsche-apotheker-zeitung.de/daz-az/2020/daz-52-2020/genesen-aber-nicht-gesund,http://ovidsp.ovid.com/ovidweb.cgi?T=JS&PAGE=reference&D=emexb&NEWS=N&AN=2010497871 |
| 19 | Hernando, Juan Enrique Cimas. Follow-up of patients with non-respiratory sequelae of COVID-19. FMC Formacion Medica Continuada en Atencion Primaria. 2021///; 28(2): 81. doi: http://dx.doi.org/10.1016/j.fmc.2020.11.004. Available at: http://www.doyma.es/fmc/,http://ovidsp.ovid.com/ovidweb.cgi?T=JS&PAGE=reference&D=emexb&NEWS=N&AN=2011154858 |
| 20 | Di Pasquale, Giuseppe, De Palma, Rossana, Fortuna, Daniela, Berti, Elena, Campo, Gianluca, Casella, Gianni. [Indirect effects of the COVID-19 pandemic on cardiovascular mortality]. Giornale italiano di cardiologia (2006). 2021///; 22(3): 188. doi: https://dx.doi.org/10.1714/3557.35336. Available at: http://ovidsp.ovid.com/ovidweb.cgi?T=JS&PAGE=reference&D=medl&NEWS=N&AN=33687369 |
| 21 | Garcia, Jose Carlos Herrera, Montellano, Ek Ixel Arellano, Gonzalez, Luis Ignacio Juarez, Andrade, Rivon Contreras. Persistence of symptoms in patients after coronavirus disease (COVID-19) in a third level hospital of Puebla, Mexico. Medicina Interna de Mexico. 2020///; 36(6): 789. doi: http://dx.doi.org/10.24245/mim.v36i6.4581. Available at: https://www.medigraphic.com/pdfs/medintmex/mim-2020/mim206g.pdf,http://ovidsp.ovid.com/ovidweb.cgi?T=JS&PAGE=reference&D=emexb&NEWS=N&AN=2010725937 |
| 22 | Holtslag, Herman R, Van Den Borst, Bram, Reijers, Monique H E, Dettling, Daniela S. Post-COVID-19 rehabilitation: A matter of customization. Nederlands tijdschrift voor geneeskunde. 2020///; 164(45): D5393. doi: #DOI#. Available at: https://www.ntvg.nl/artikelen/nazorg-voor-covid-19-patienten,http://ovidsp.ovid.com/ovidweb.cgi?T=JS&PAGE=reference&D=emexb&NEWS=N&AN=2008596180 |
| 23 | Kovalchuk, V V. The role of the new coronavirus infection (COVID-19) in the progression and development of cerebrovascular diseases. A competent choice of pathogenic treatment is the key to success in treatment and prevention. An expert's view from the 'red zone'. Nevrologiya, Neiropsikhiatriya, Psikhosomatika. 2021///; 13(1): 57. doi: http://dx.doi.org/10.14412/2074-2711-2021-1-57-66. Available at: https://nnp.ima- |

| | |
|---|---|
| | press.net/nnp/article/view/1508,http://ovidsp.ovid.com/ovidweb.cgi?T=JS&PAGE=reference&D=emexb&NEWS=N&AN=2011311536 |
| 24 | Ostroumova, T M, Chernousov, P A, Kuznetsov, I V. Cognitive impairment in COVID-19 survivors. Nevrologiya, Neiropsikhiatriya, Psikhosomatika. 2021///; 13(1): 126. doi: http://dx.doi.org/10.14412/2074-2711-2021-1-126-130. Available at: https://nnp.ima-press.net/nnp/article/view/1519,http://ovidsp.ovid.com/ovidweb.cgi?T=JS&PAGE=reference&D=emexb&NEWS=N&AN=2011311521 |
| 25 | Karkashadze, Georgiy A, Zelenkova, Irina V, Sergienko, Natalia S, Nesterova, Yuliya V, Gubanova, Svetlana G, Gankovskiy, Viktor A, Gogberashvili, Tinatin Yu, Konstantinidi, Tatiana A, Bushueva, Daria A, Rykunova, Anastasiya I, Gordeeva, Olga B, Dubonosova, Ekaterina Yu, Vishneva, Elena A, Kaytukova, Elena V, Shirdanina, Ekaterina V, Lamasova, Anastasiya D, Leonova, Elizaveta V, Pankova, Alina R, Namazova-Baranova, Leyla, Baranov, Alexander A, Rusinova, Dina S. Non-randomized comparative study of olfaction in post-COVID-19 children. Intermediary results. Pediatricheskaya Farmakologiya. 2020///; 17(6): 502. doi: http://dx.doi.org/10.15690/PF.V17I6.2201. Available at: https://www.pedpharma.ru/jour/article/view/1921,http://ovidsp.ovid.com/ovidweb.cgi?T=JS&PAGE=reference&D=emexb&NEWS=N&AN=2011083989 |
| 26 | .. Post-COVID-19-Functional-Status-Skala misst Einschränkungen nach SARS-CoV-2-Infektion. Pneumologie. 2021/01//; 75(01): 8. doi: 10.1055/a-1263-2042. Available at: http://www.thieme-connect.com/ejournals/toc/pneumologie,http://ovidsp.ovid.com/ovidweb.cgi?T=JS&PAGE=reference&D=emexb&NEWS=N&AN=633948988,http://www.thieme-connect.de/DOI/DOI?10.1055/a-1263-2042 |
| 27 | Golukhova, E Z, Bulaeva, N I, Sokolova, N Y. Cardiovascular diseases and complications in patients with covid-19. Profilakticheskaya Meditsina. 2020///; 23(7): 72. doi: http://dx.doi.org/10.17116/profmed20202307172. Available at: http://mediasphera.ru/journal/profilakticheskaya-meditsina,http://ovidsp.ovid.com/ovidweb.cgi?T=JS&PAGE=reference&D=emexb&NEWS=N&AN=2005963945 |
| 28 | Alarcon-Rodriguez J Urena-Vacas A, Martin-Pinacho JJ, Rigual-Bobillo JA, Jaureguizar-Oriol A, Gorospe-Sarasua,L., Fernandez-Velilla M. Radiological management and follow-up of post-COVID-19 patients; 7914006 CSR - Reports results. Radiologia. 2021///; #volume#(#issue#): #pages#. doi: 10.1016/j.rx.2021.02.003. Available at: https://pubmed.ncbi.nlm.nih.gov/33726915 |
| 29 | Prieto, Manuel Antonio, Prieto, Omar, Castro, Horacio Matias. Long covid: cross sectional study. Revista de la Facultad de Ciencias Medicas (Cordoba, Argentina). 2021///; 78(1): 33. doi: https://dx.doi.org/10.31053/1853.0605.v78.n1.32048. Available at: http://ovidsp.ovid.com/ovidweb.cgi?T=JS&PAGE=reference&D=mesx&NEWS=N&AN=33787016 |
| 30 | Valga, Francisco, Vega-Diaz, Nicanor, Macia, Manuel, Rodriguez-Perez, Jose. [A new inflammatory-microthrombotic syndrome as an explanation for thrombotic complications in patients with COVID-19]. Revista espanola de cardiologia. 2021///; 74(1): 115. doi: https://dx.doi.org/10.1016/j.recesp.2020.08.010. Available at: http://ovidsp.ovid.com/ovidweb.cgi?T=JS&PAGE=reference&D=pmnm&NEWS=N&AN=32901168 |
| 31 | Fillat, Angel Cequier, Gonzalez-Juanatey, Jose. COVID-19. The social, health and cardiovascular consequences. Revista Espanola de Cardiologia Suplementos. 2020///; 20(#issue#): 1. doi: http://dx.doi.org/10.1016/S1131-3587%2820%2930027-3. Available at: http://www.revespcardiol.org,http://ovidsp.ovid.com/ovidweb.cgi?T=JS&PAGE=reference&D=emexb&NEWS=N&AN=2008601133 |
| 32 | Radvan, Martin, Bartecku, Elis, Sykorova, Ubica, Parizkova, Radana, Richter, Svatopluk, Kamenik, Martin, Koc, Lumir, Kala, Petr. Follow-up care after COVID-19 and its related concerns. Vnitrni lekarstvi. 2021///; 67(1): 30. doi: #DOI#. Available at: http://ovidsp.ovid.com/ovidweb.cgi?T=JS&PAGE=reference&D=medl&NEWS=N&AN=33752388 |

### Supplementary File 3 – Supplementary Figures

#### SHORT-TERM

##### One or more symptoms

Persistence or presence of one or more symptoms at follow-up

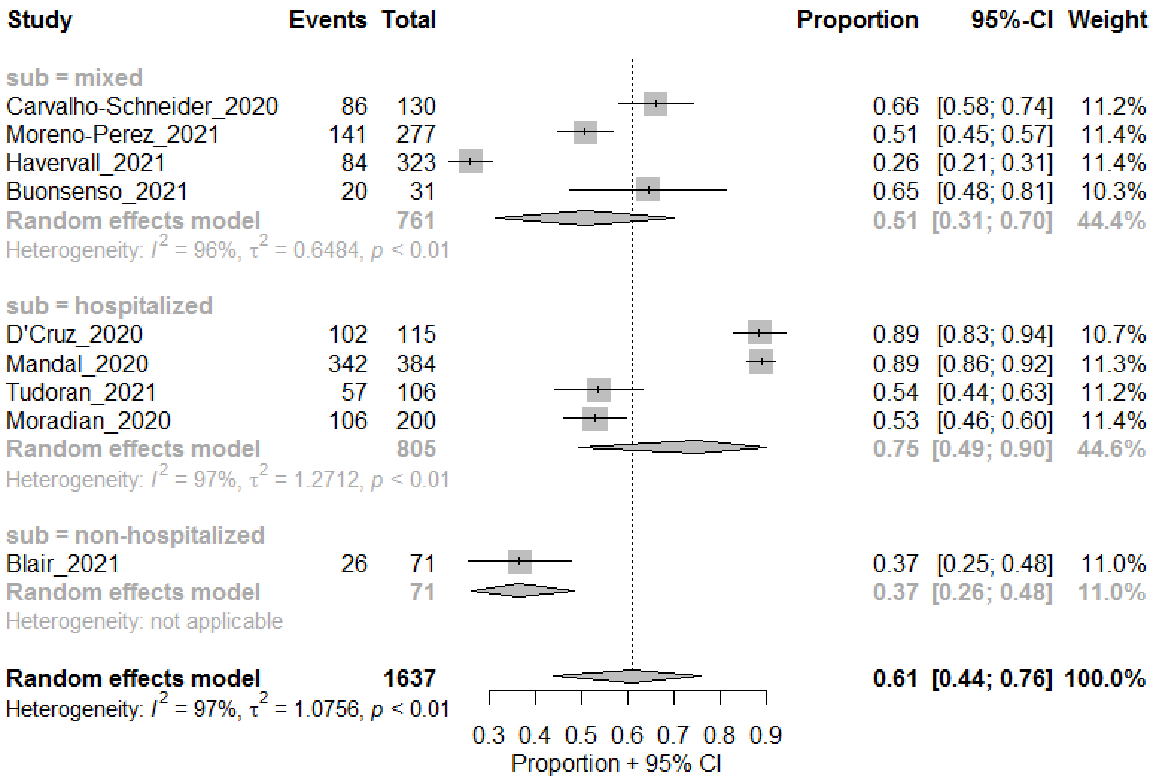

##### Fatigue

*By level of care at acute phase*

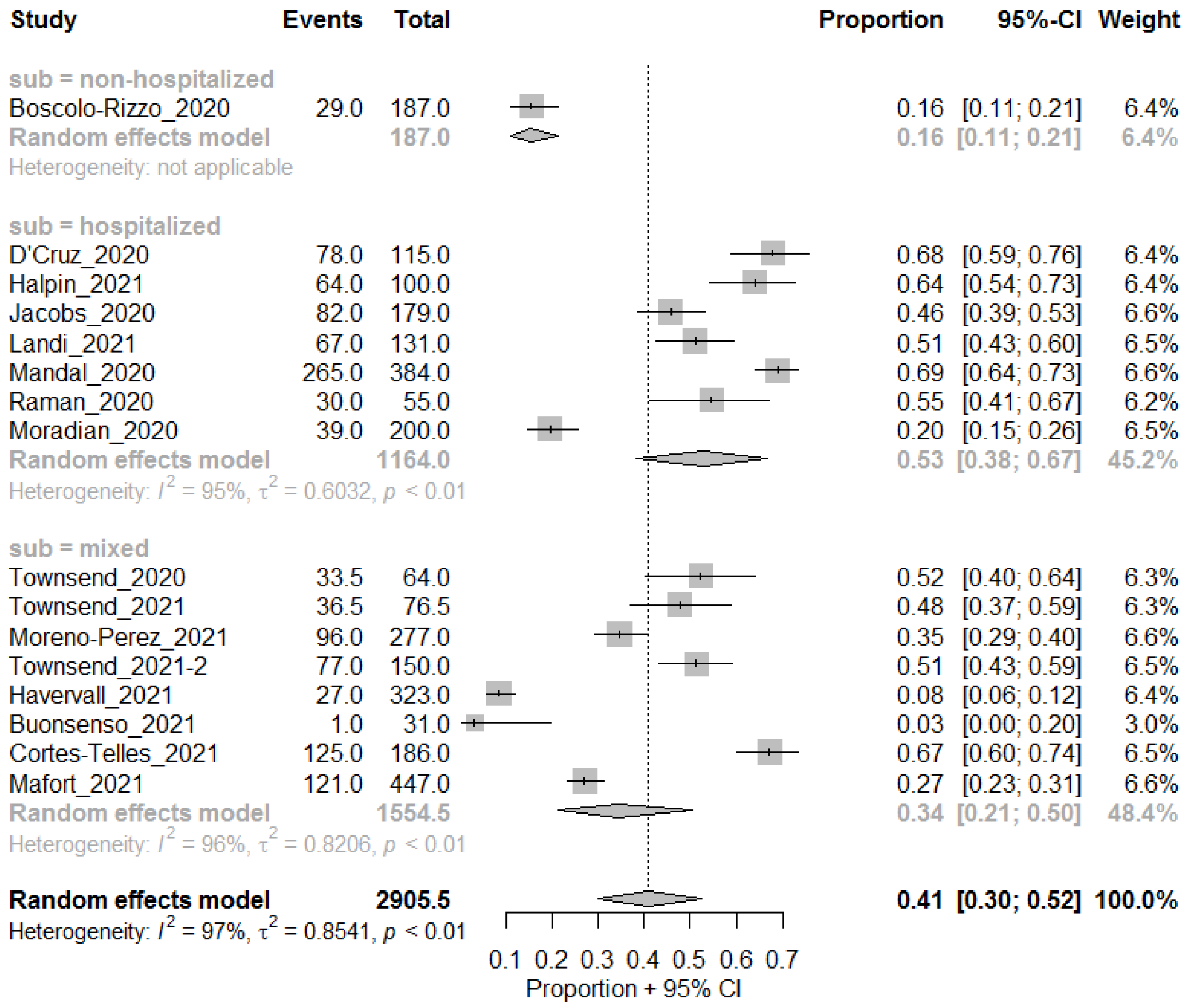

*Adults vs. children*

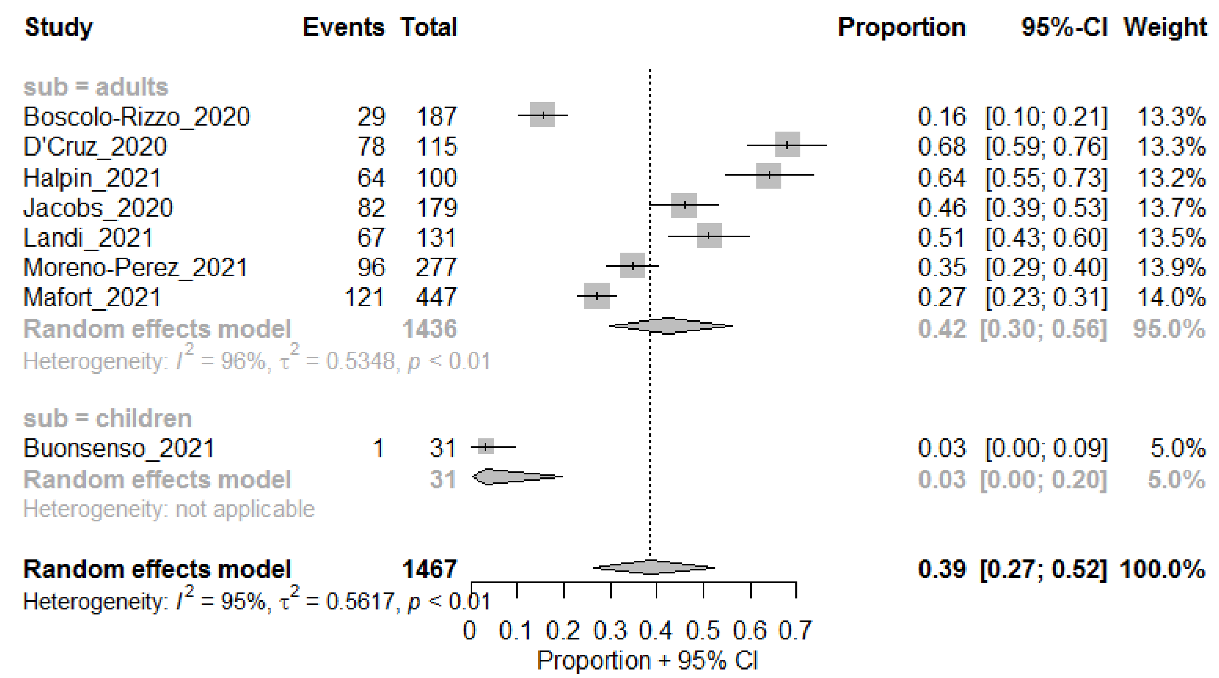

#### Weakness

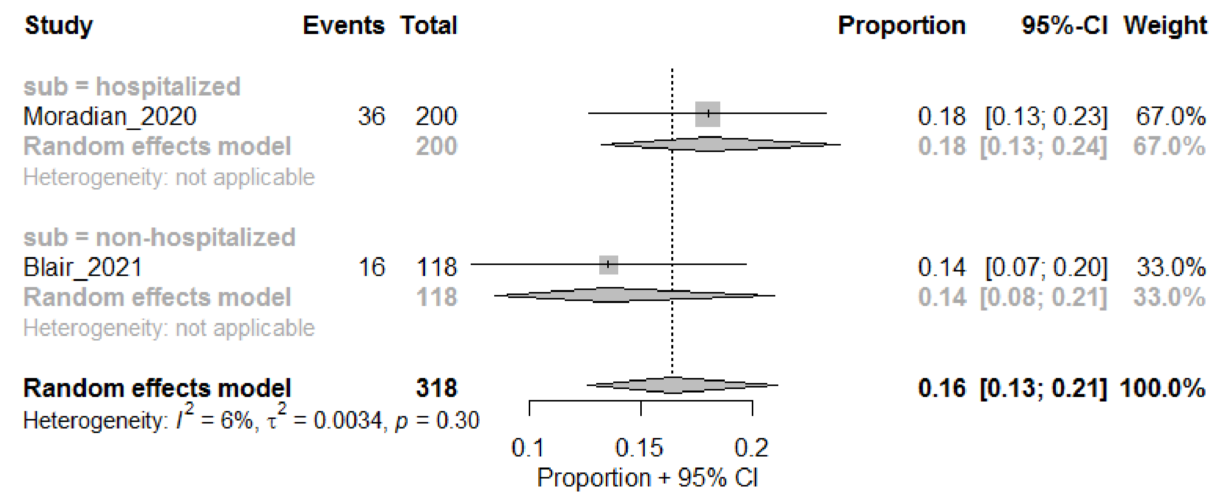

#### Respiratory

##### Shortness of Breath

*2 subgroups of hospitalized*

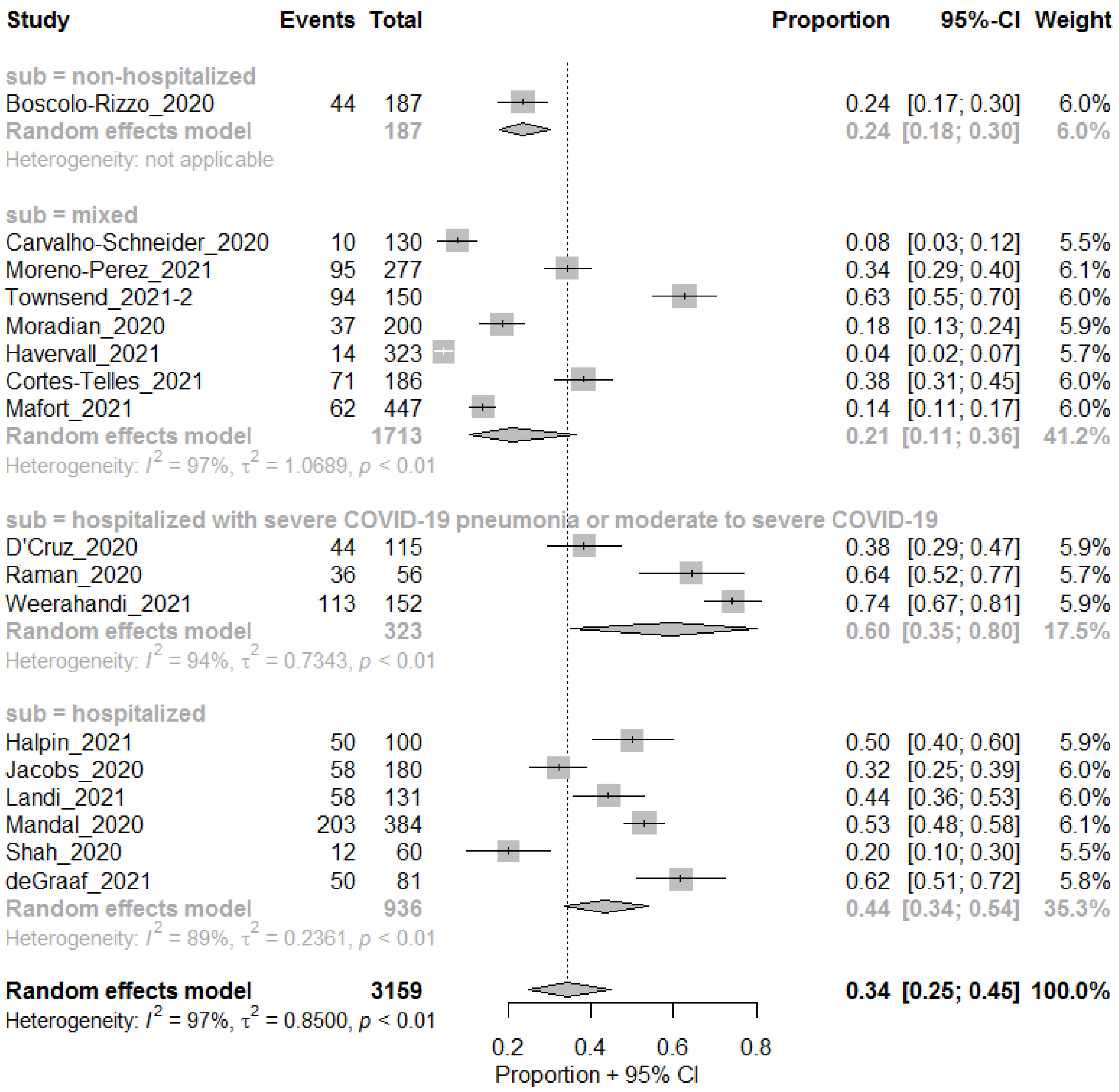

*no subgroups for hospitalized*

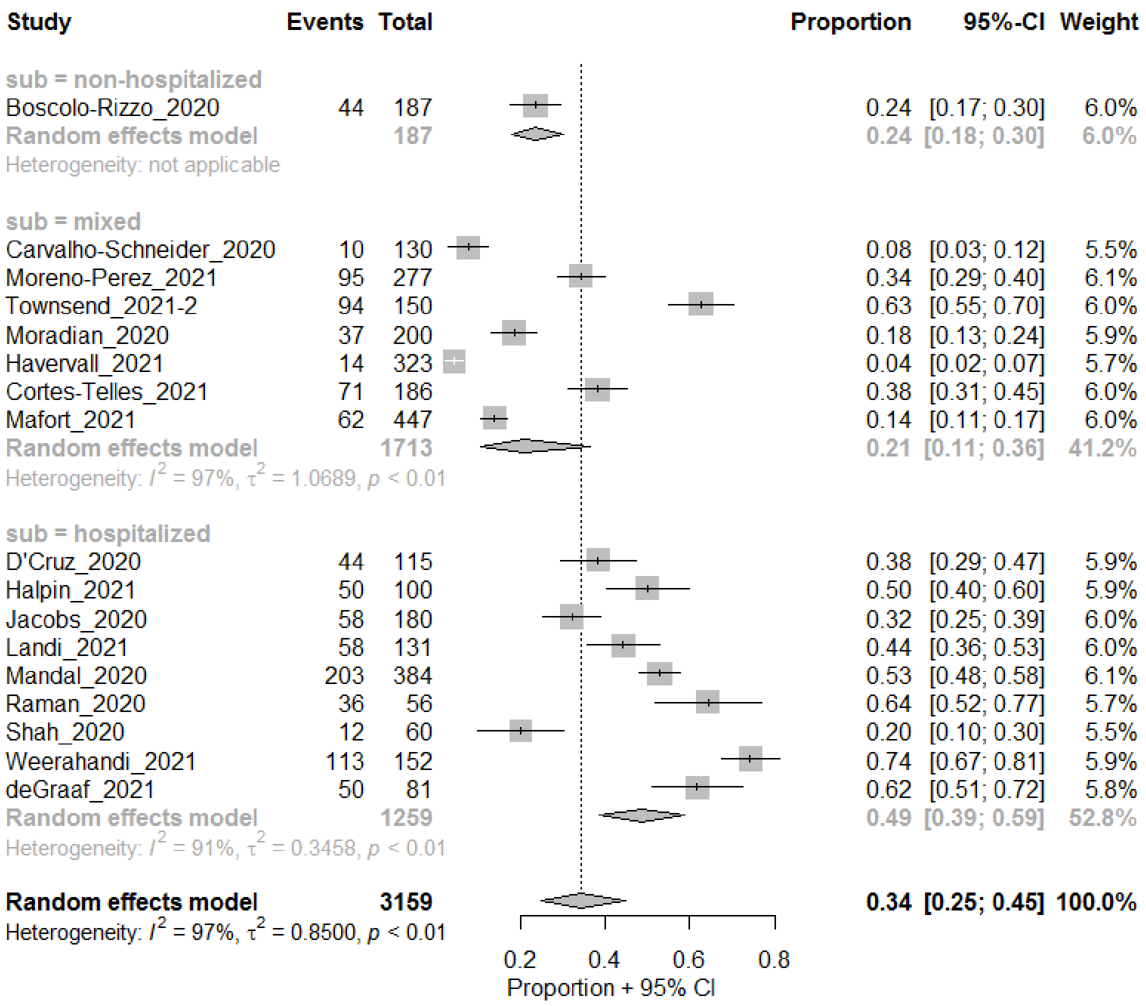
Study Events Total Proportion 95%-CI Weight
sub = non-hospitalized
Boscolo-Rizzo_2020 44 187 0.24 [0.17; 0.30] 6.0%
Random effects model 187 0.24 [0.18; 0.30] 6.0%
Heterogeneity: not applicable
sub = mixed
Carvalho-Schneider_2020 10 130 0.08 [0.03; 0.12] 5.5%
Moreno-Perez_2021 95 277 0.34 [0.29; 0.40] 6.1%
Townsend_2021-2 94 150 0.63 [0.55; 0.70] 6.0%
Moradian_2020 37 200 0.18 [0.13; 0.24] 5.9%
Havervall_2021 14 323 0.04 [0.02; 0.07] 5.7%
Cortes-Telles_2021 71 186 0.38 [0.31; 0.45] 6.0%
Mafort_2021 62 447 0.14 [0.11; 0.17] 6.0%
Random effects model 1713 0.21 [0.11; 0.36] 41.2%
Heterogeneity: $I^2$ = 97%, $\tau^2$ = 1.0689, $p$ < 0.01
sub = hospitalized
D'Cruz_2020 44 115 0.38 [0.29; 0.47] 5.9%
Halpin_2021 50 100 0.50 [0.40; 0.60] 5.9%
Jacobs_2020 58 180 0.32 [0.25; 0.39] 6.0%
Landi_2021 58 131 0.44 [0.36; 0.53] 6.0%
Mandal_2020 203 384 0.53 [0.48; 0.58] 6.1%
Raman_2020 36 56 0.64 [0.52; 0.77] 5.7%
Shah_2020 12 60 0.20 [0.10; 0.30] 5.5%
Weerahandi_2021 113 152 0.74 [0.67; 0.81] 5.9%
deGraaf_2021 50 81 0.62 [0.51; 0.72] 5.8%
Random effects model 1259 0.49 [0.39; 0.59] 52.8%
Heterogeneity: $I^2$ = 91%, $\tau^2$ = 0.3458, $p$ < 0.01
Random effects model 3159 0.34 [0.25; 0.45] 100.0%
Heterogeneity: $I^2$ = 97%, $\tau^2$ = 0.8500, $p$ < 0.01
0.2 0.4 0.6 0.8
Proportion + 95% CI

#### Cough

*Any*

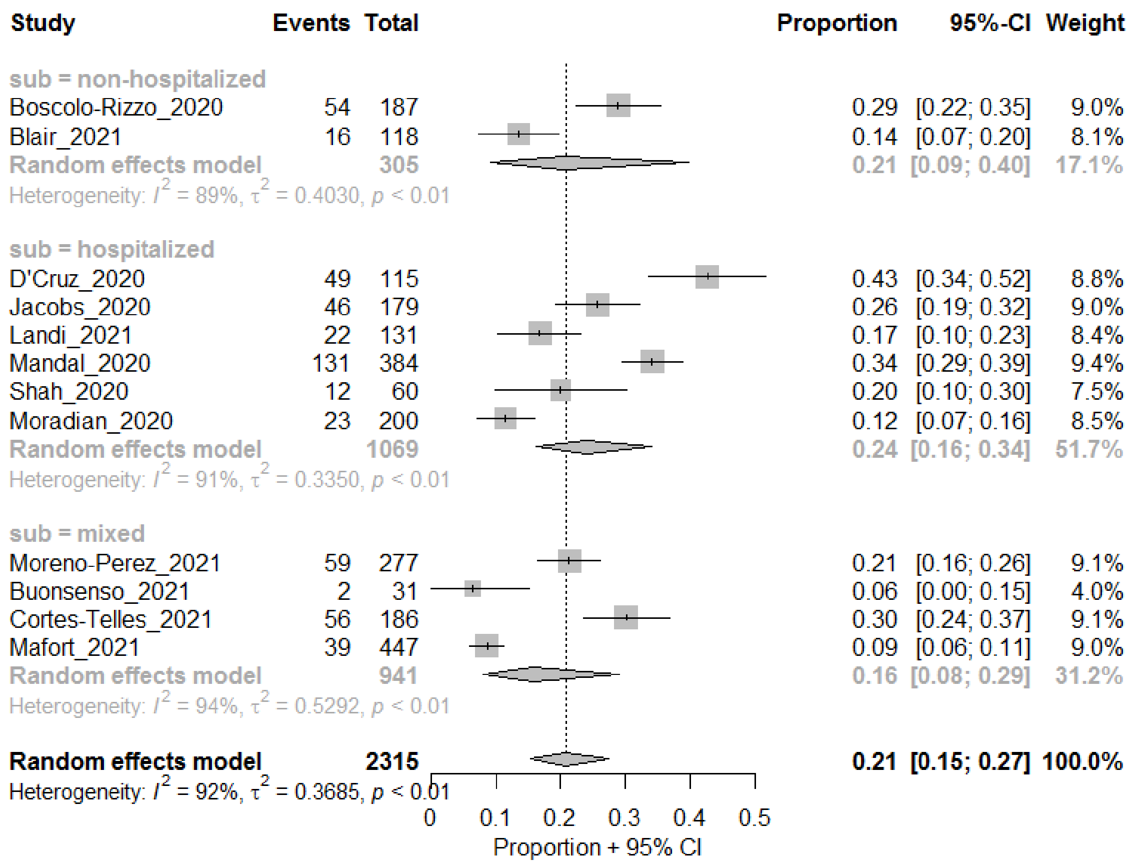

*Dry Cough*

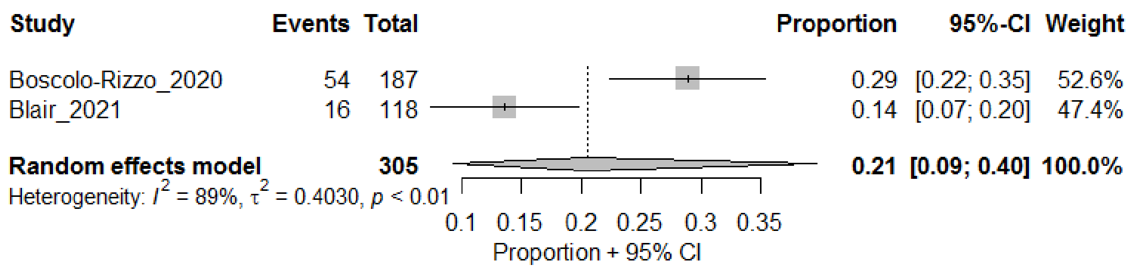

Rhinitis or runny nose

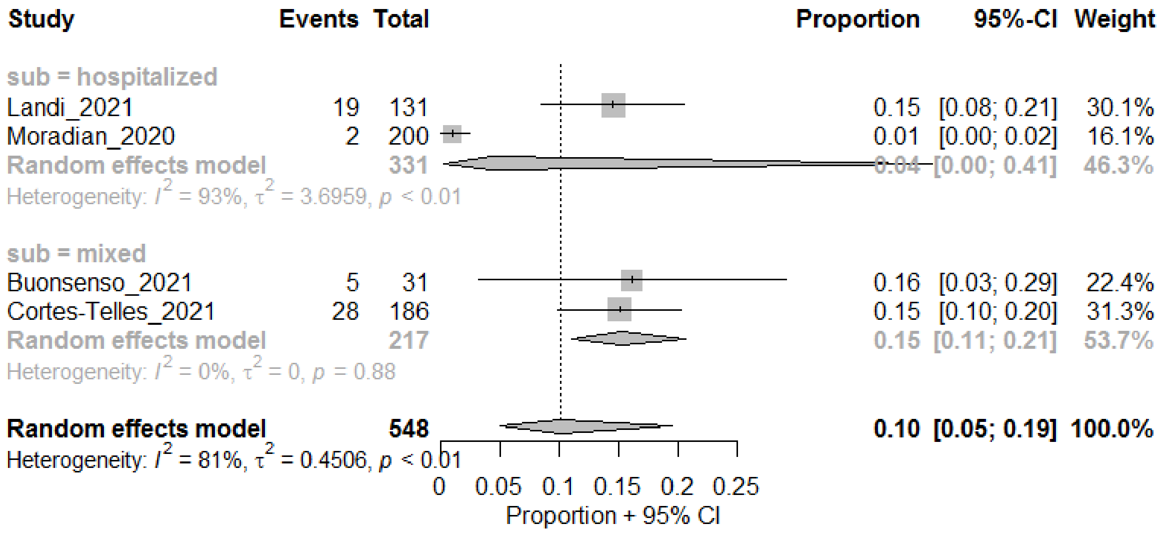

#### Sore throat

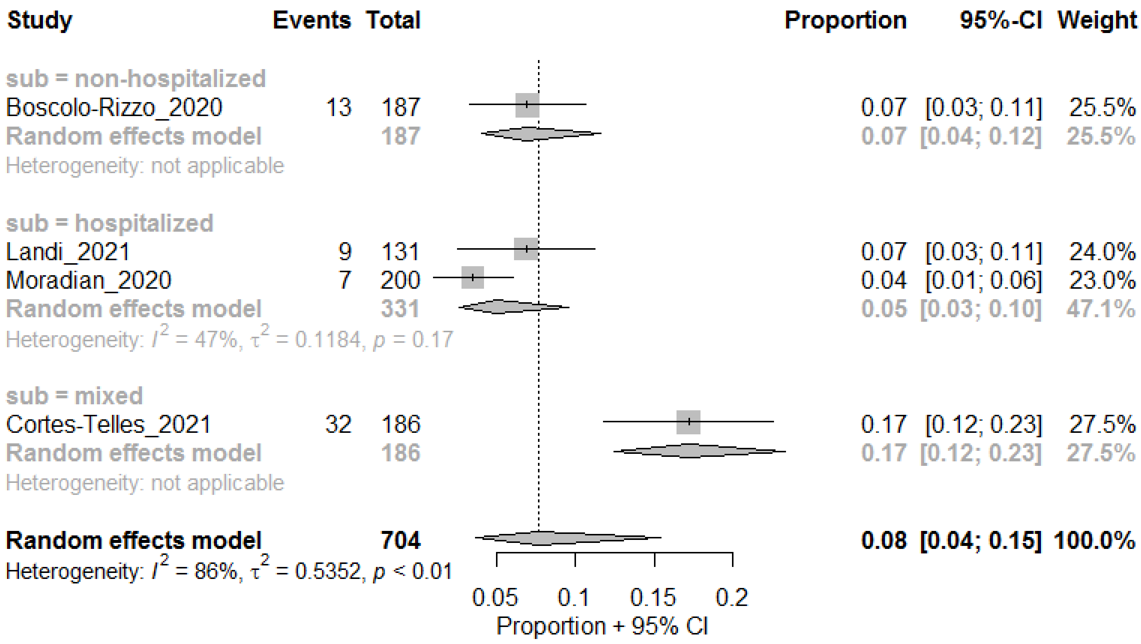

#### Blocked nose

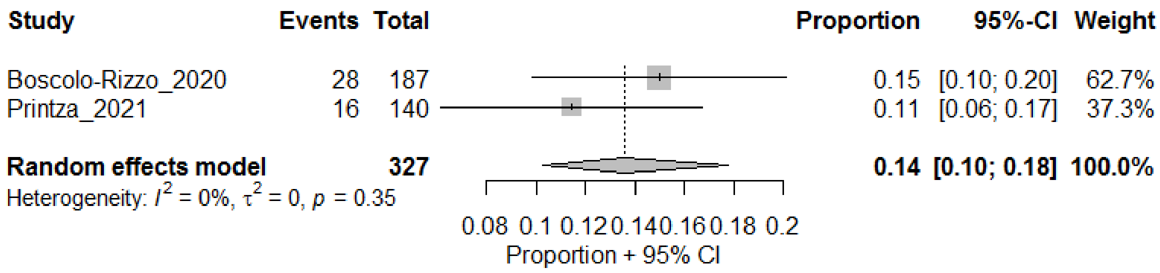

#### Phlegm

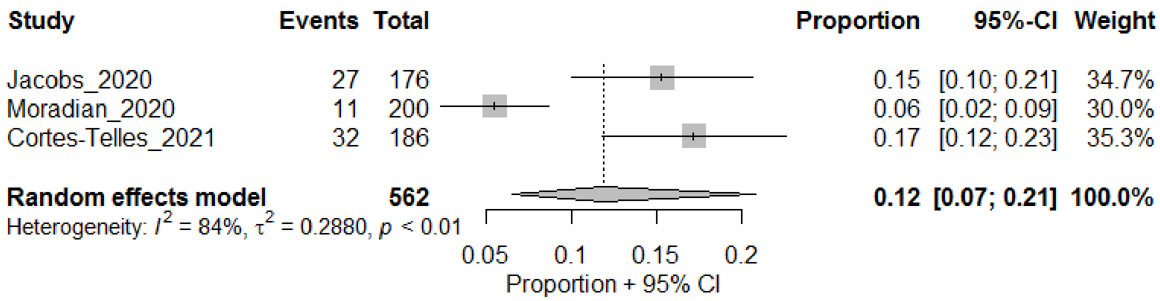

#### Influenza-like illness

##### Headaches

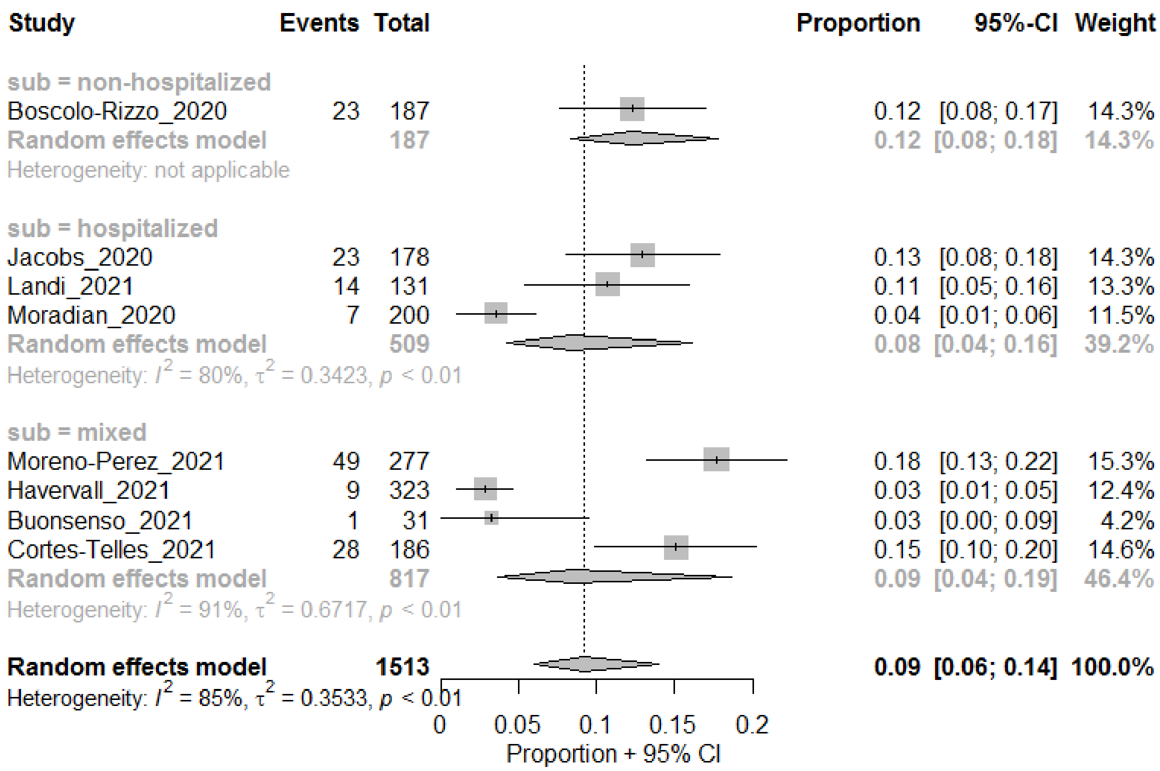
Study Events Total Proportion 95%-CI Weight
sub = non-hospitalized
Boscolo-Rizzo_2020 23 187 0.12 [0.08; 0.17] 14.3%
Random effects model 187 0.12 [0.08; 0.18] 14.3%
Heterogeneity: not applicable
sub = hospitalized
Jacobs_2020 23 178 0.13 [0.08; 0.18] 14.3%
Landi_2021 14 131 0.11 [0.05; 0.16] 13.3%
Moradian_2020 7 200 0.04 [0.01; 0.06] 11.5%
Random effects model 509 0.08 [0.04; 0.16] 39.2%
Heterogeneity: $I^2$ = 80%, $\tau^2$ = 0.3423, $p$ < 0.01
sub = mixed
Moreno-Perez_2021 49 277 0.18 [0.13; 0.22] 15.3%
Havervall_2021 9 323 0.03 [0.01; 0.05] 12.4%
Buonsenso_2021 1 31 0.03 [0.00; 0.09] 4.2%
Cortes-Telles_2021 28 186 0.15 [0.10; 0.20] 14.6%
Random effects model 817 0.09 [0.04; 0.19] 46.4%
Heterogeneity: $I^2$ = 91%, $\tau^2$ = 0.6717, $p$ < 0.01
Random effects model 1513 0.09 [0.06; 0.14] 100.0%
Heterogeneity: $I^2$ = 85%, $\tau^2$ = 0.3533, $p$ < 0.01
0 0.05 0.1 0.15 0.2
Proportion + 95% CI

#### Fever

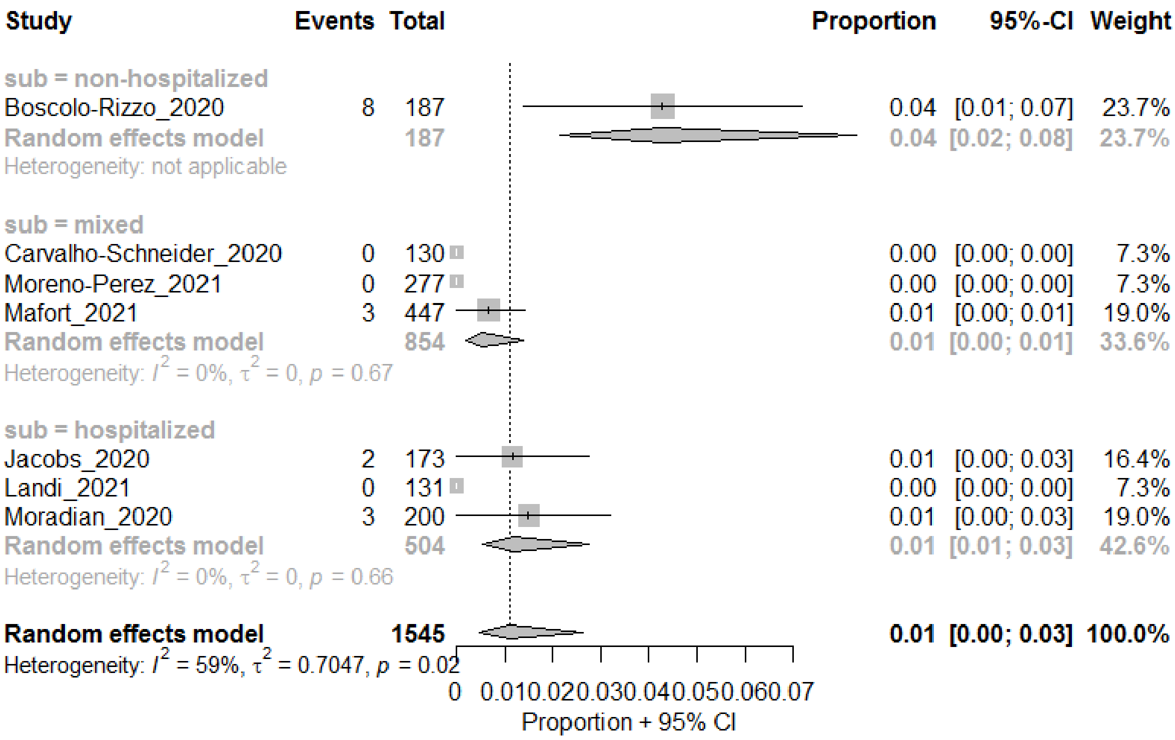
Study Events Total Proportion 95%-CI Weight
sub = non-hospitalized
Boscolo-Rizzo_2020 8 187 0.04 [0.01; 0.07] 23.7%
Random effects model 187 0.04 [0.02; 0.08] 23.7%
Heterogeneity: not applicable
sub = mixed
Carvalho-Schneider_2020 0 130 0.00 [0.00; 0.00] 7.3%
Moreno-Perez_2021 0 277 0.00 [0.00; 0.00] 7.3%
Mafort_2021 3 447 0.01 [0.00; 0.01] 19.0%
Random effects model 854 0.01 [0.00; 0.01] 33.6%
Heterogeneity: $I^2$ = 0%, $\tau^2$ = 0, $p$ = 0.67
sub = hospitalized
Jacobs_2020 2 173 0.01 [0.00; 0.03] 16.4%
Landi_2021 0 131 0.00 [0.00; 0.00] 7.3%
Moradian_2020 3 200 0.01 [0.00; 0.03] 19.0%
Random effects model 504 0.01 [0.01; 0.03] 42.6%
Heterogeneity: $I^2$ = 0%, $\tau^2$ = 0, $p$ = 0.66
Random effects model 1545 0.01 [0.00; 0.03] 100.0%
Heterogeneity: $I^2$ = 59%, $\tau^2$ = 0.7047, $p$ = 0.02
0 0.010.020.030.040.050.060.07
Proportion + 95% CI

#### Olfactory and Taste Impairments

##### Any Smell Dysfunction

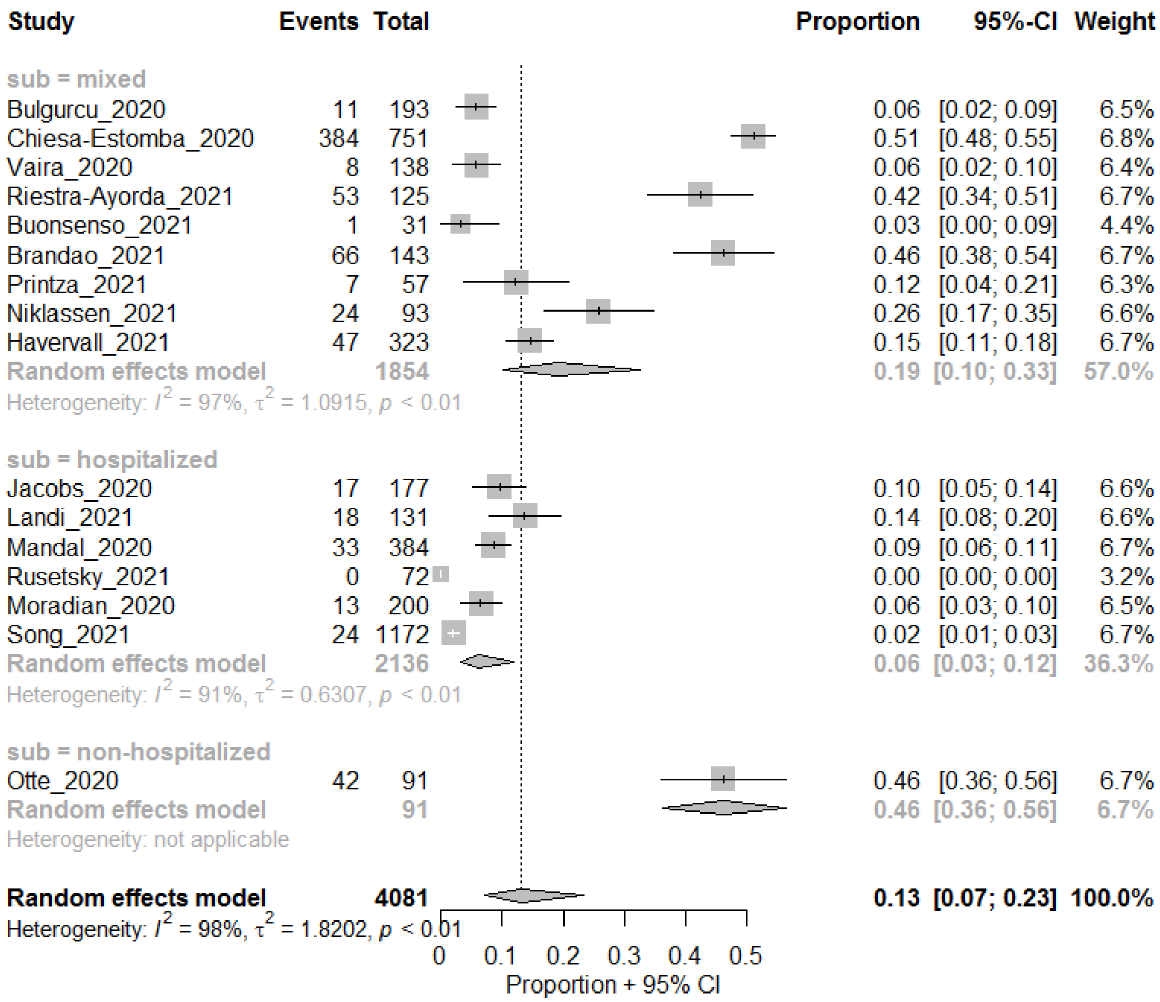

##### Hyposmia

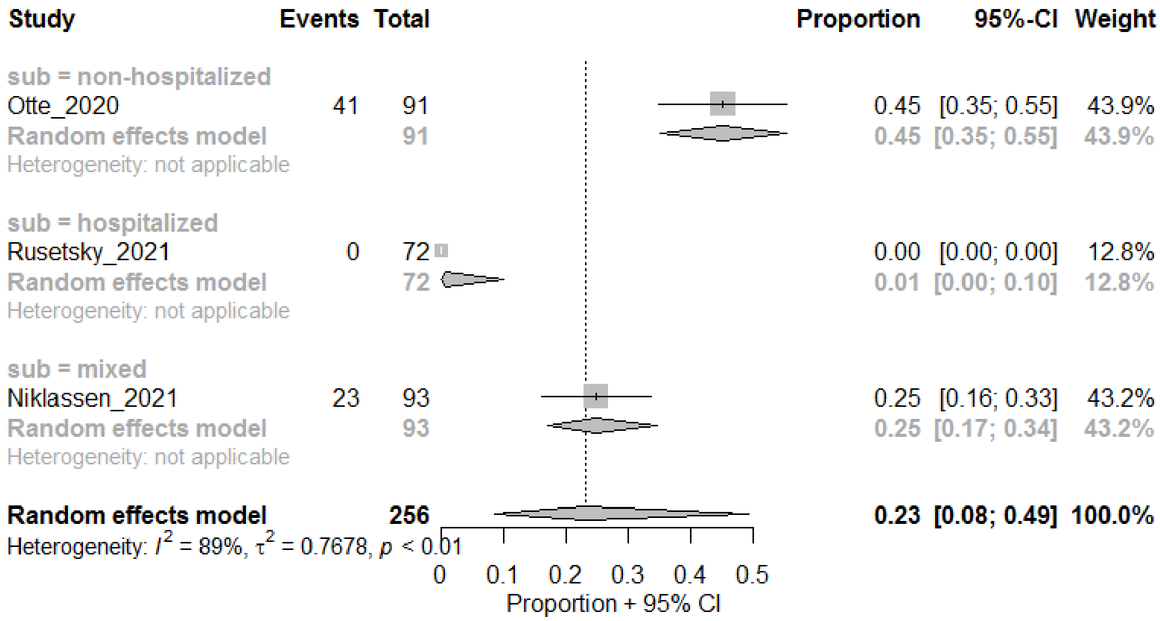

#### Anosmia

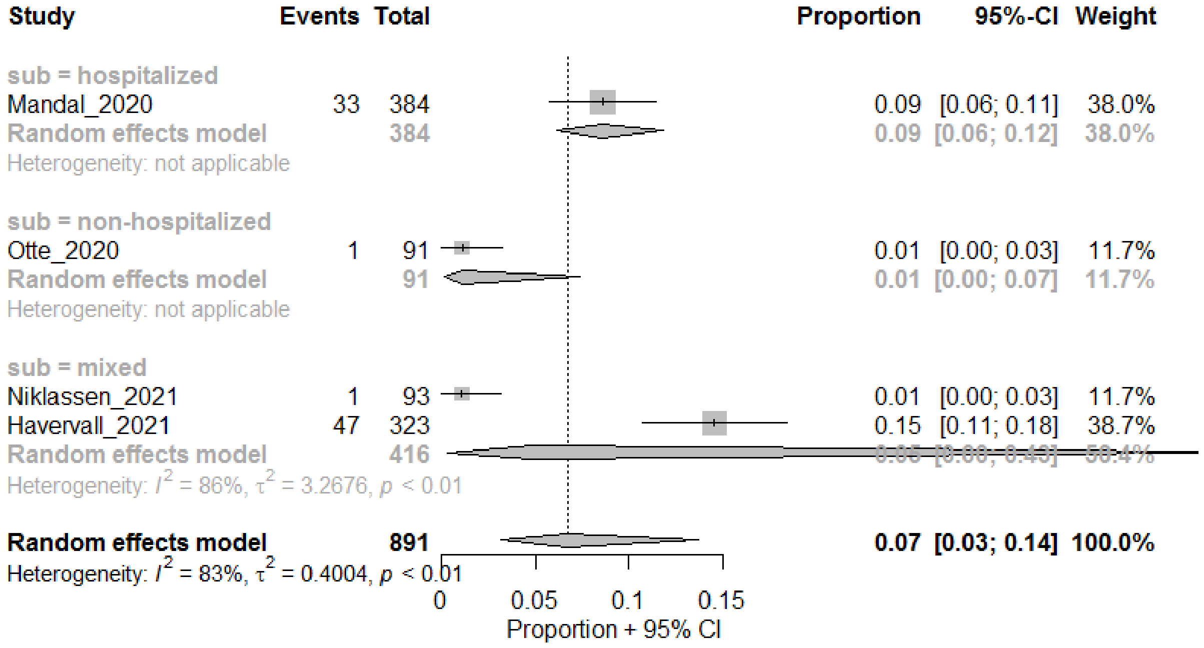

#### Smell or Taste dysfunction

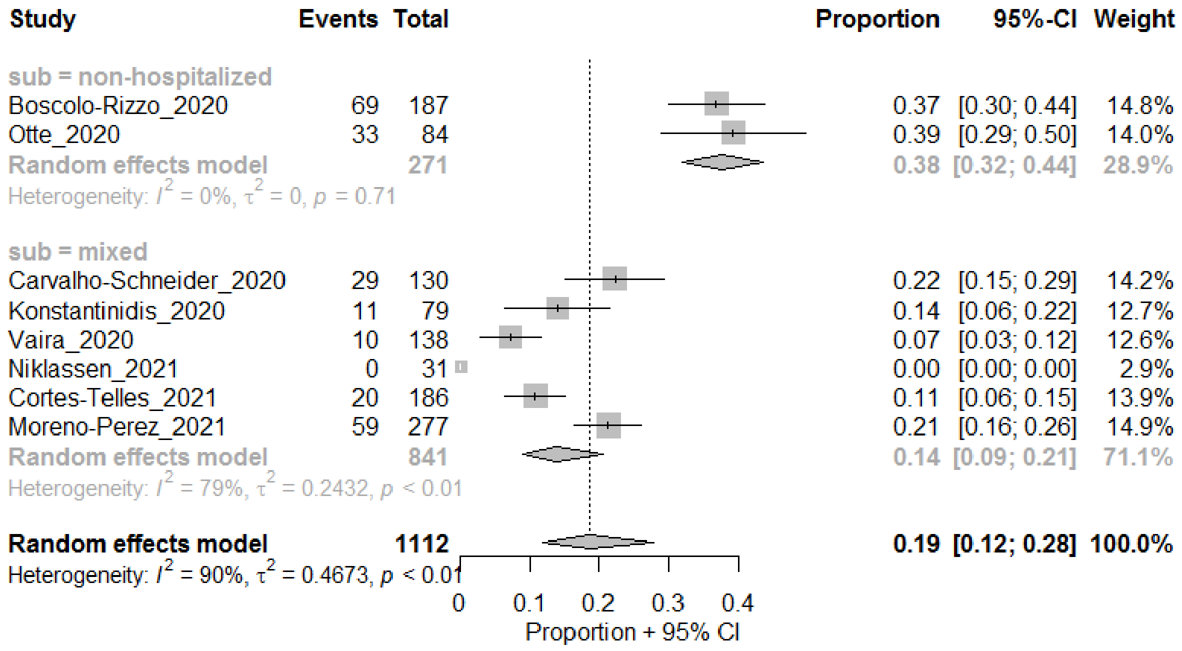

#### Any Taste Dysfunction

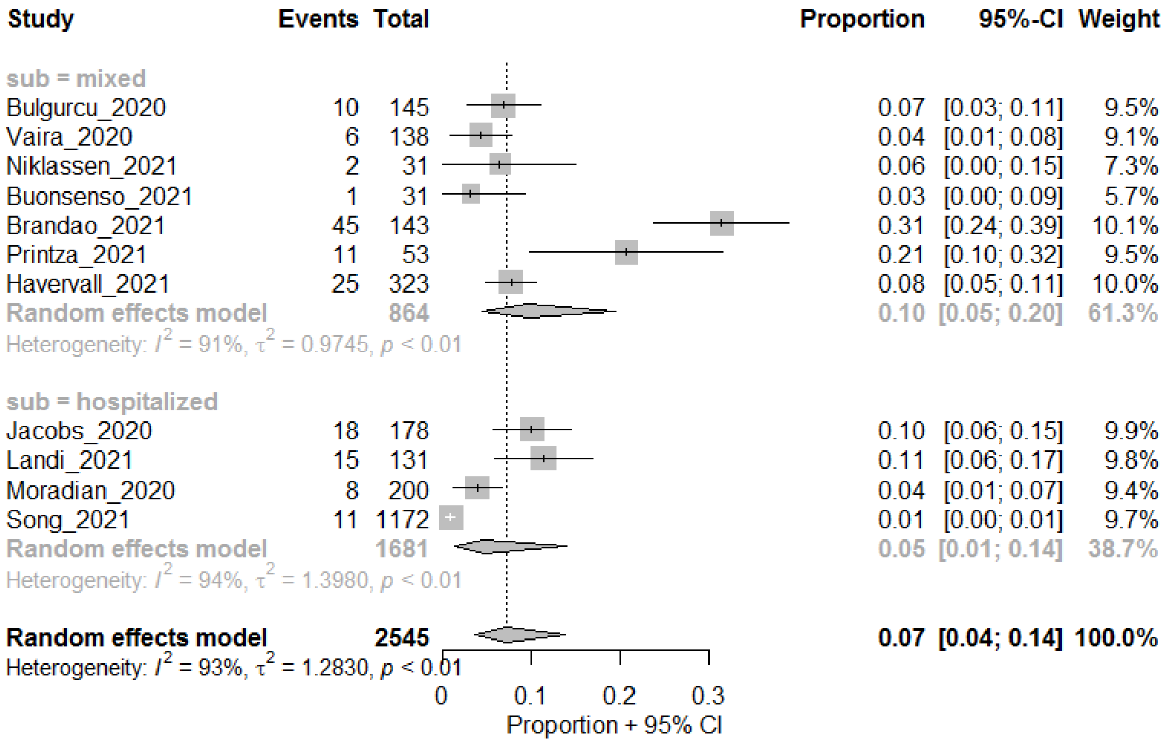

#### Neurocognitive Impairment

#### Cognitive impairment

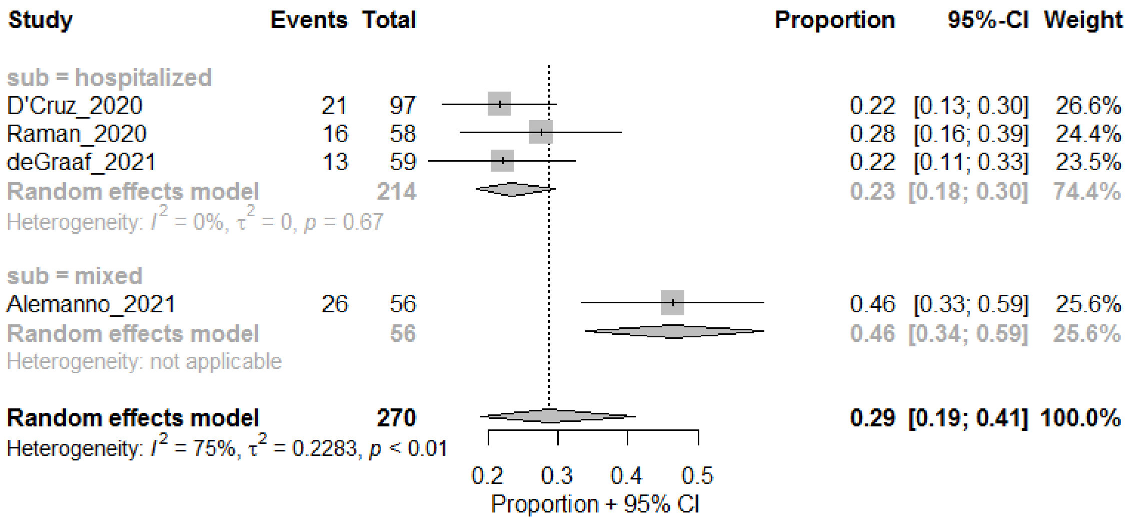

#### Concentration problems

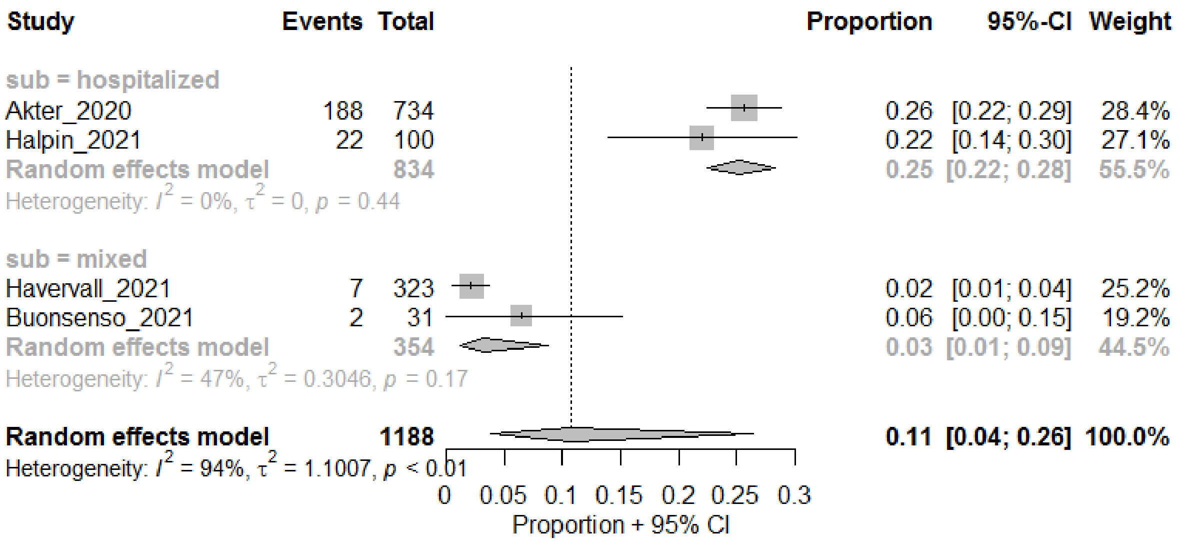

#### Memory problem

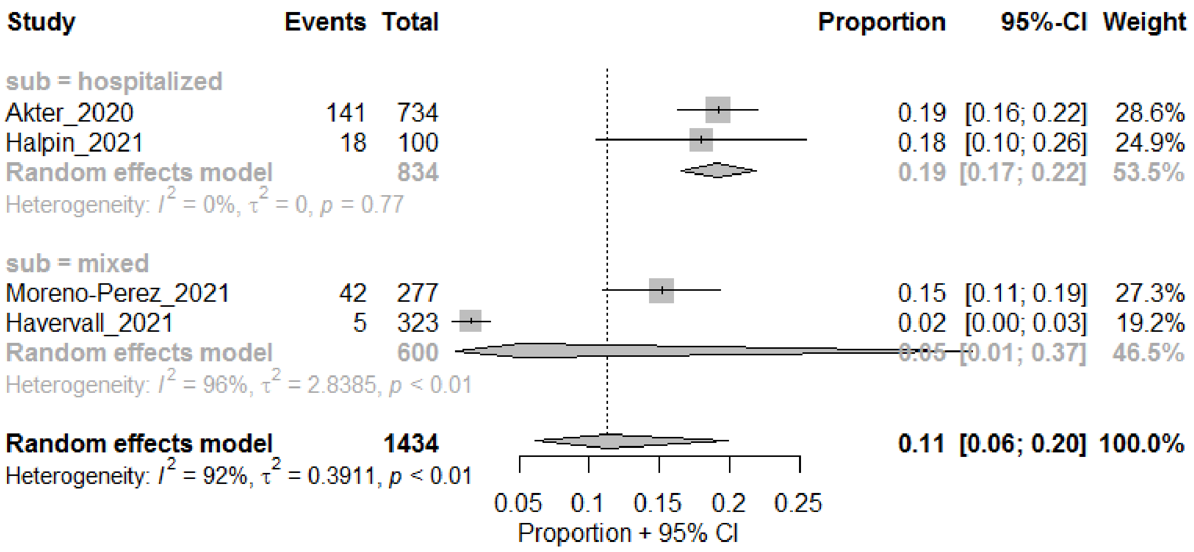

#### Dizziness

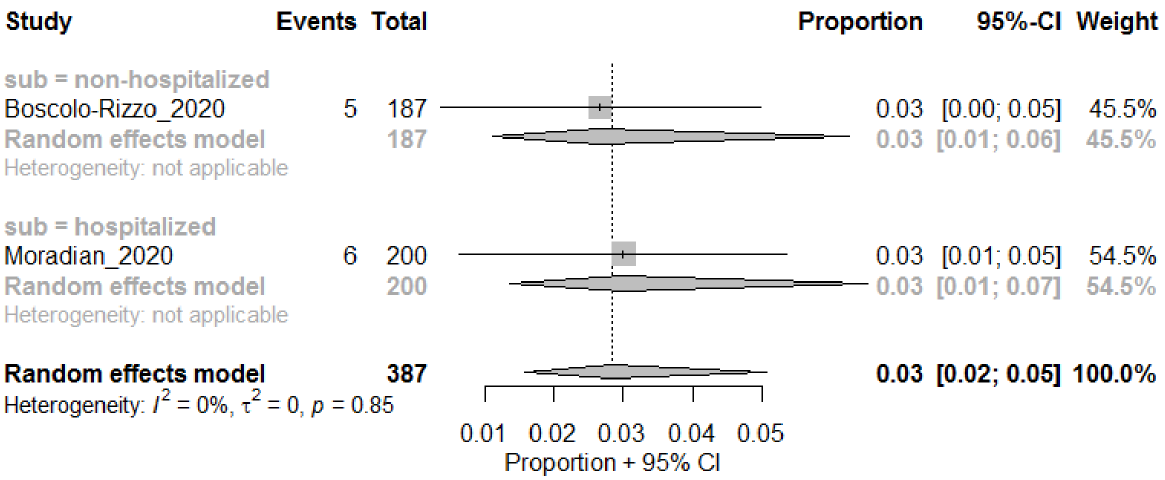

#### Neuromuscular

##### Joint pain

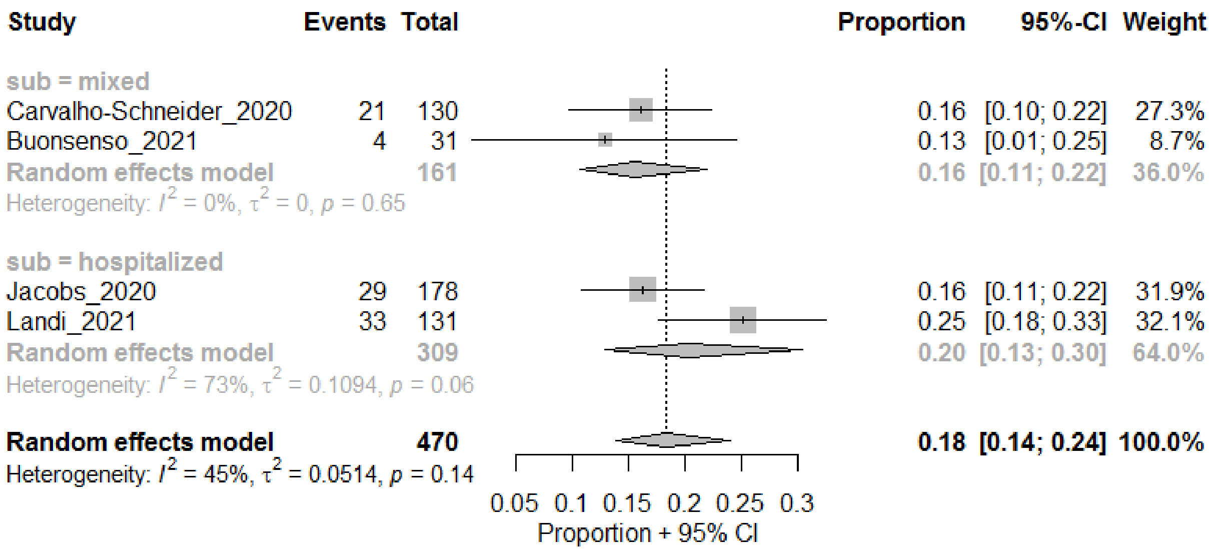

Muscle pain

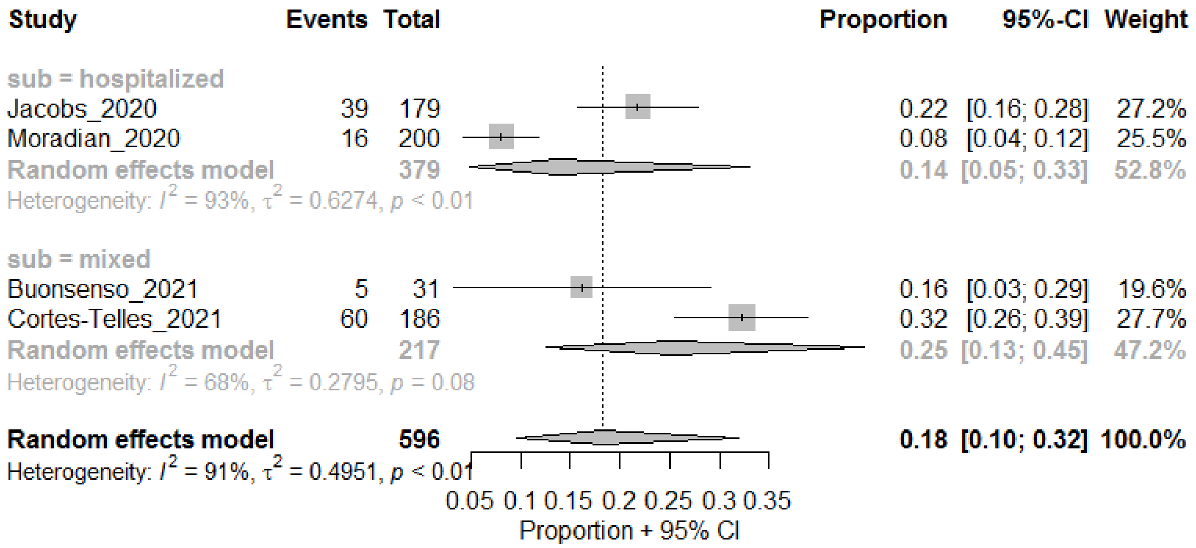

Muscle or joint pain

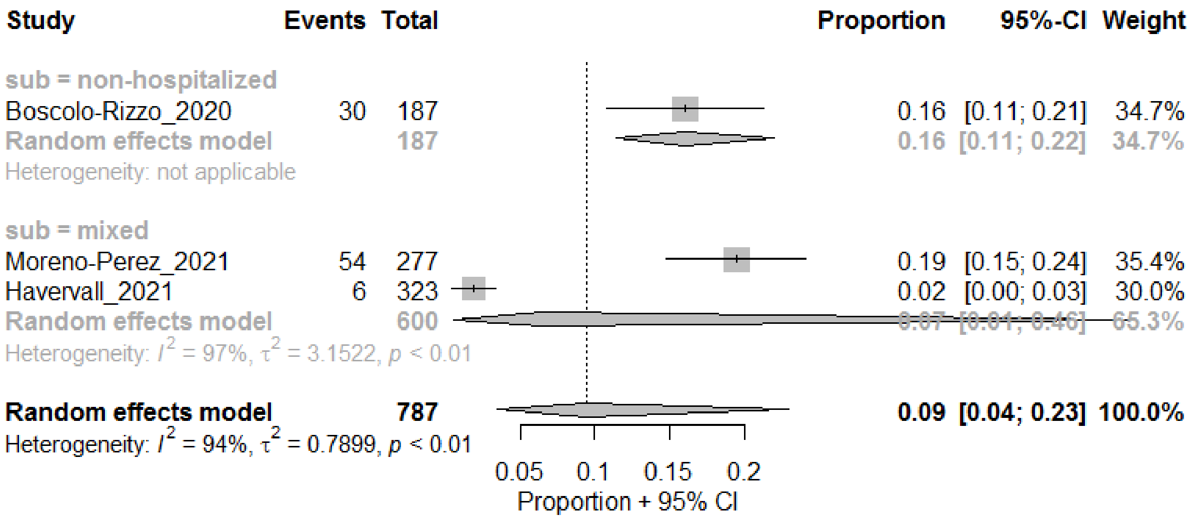

#### Cardiovascular

##### Chest pain

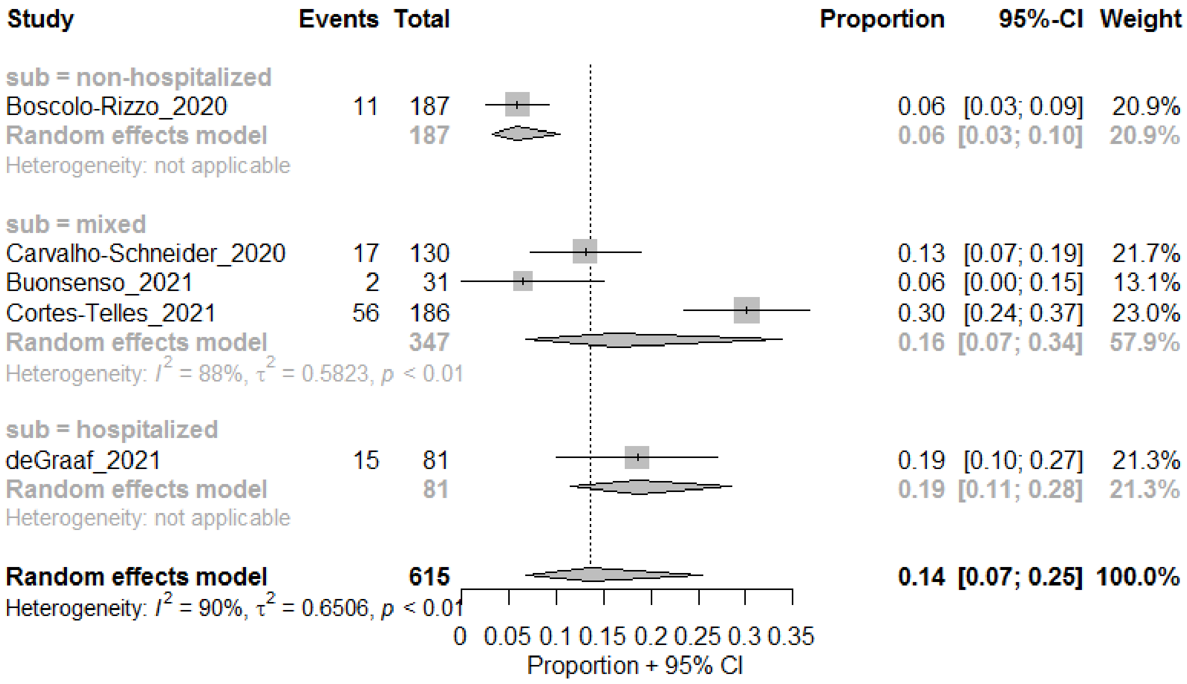

##### Palpitations

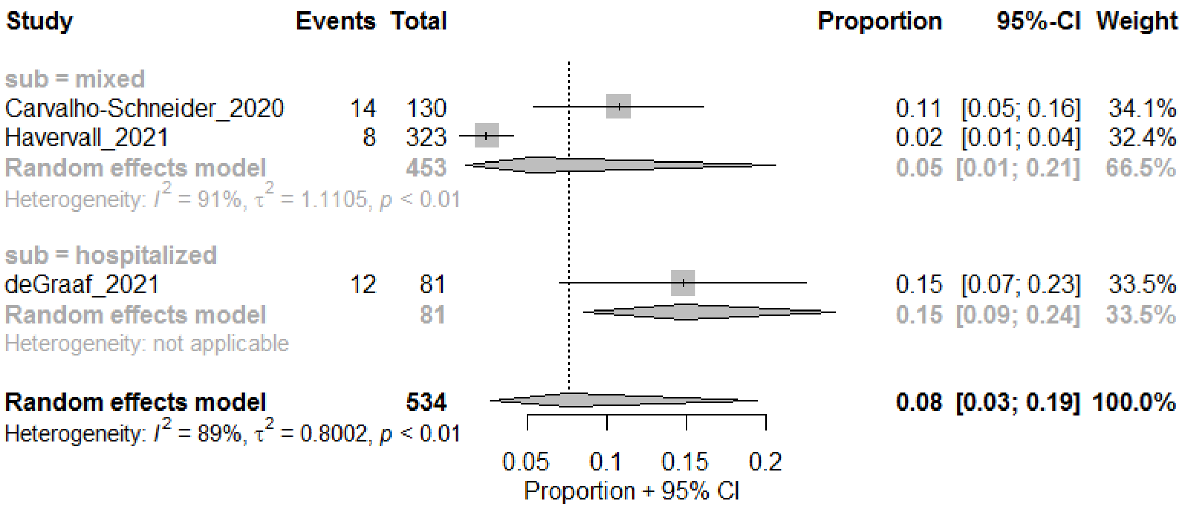

#### Digestive

##### Appetite problem

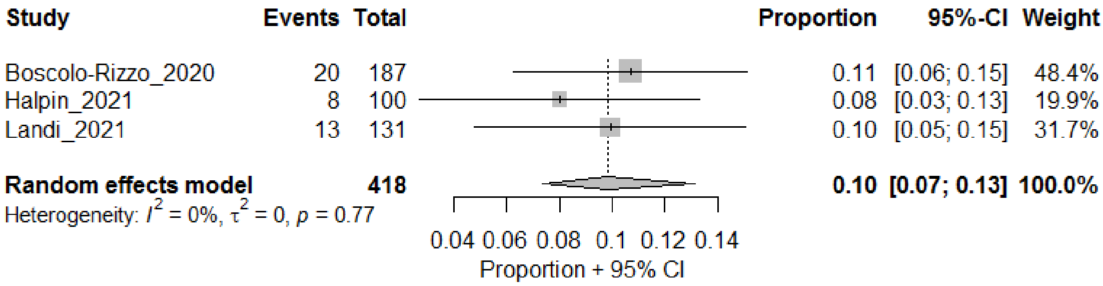

##### Diarrhea

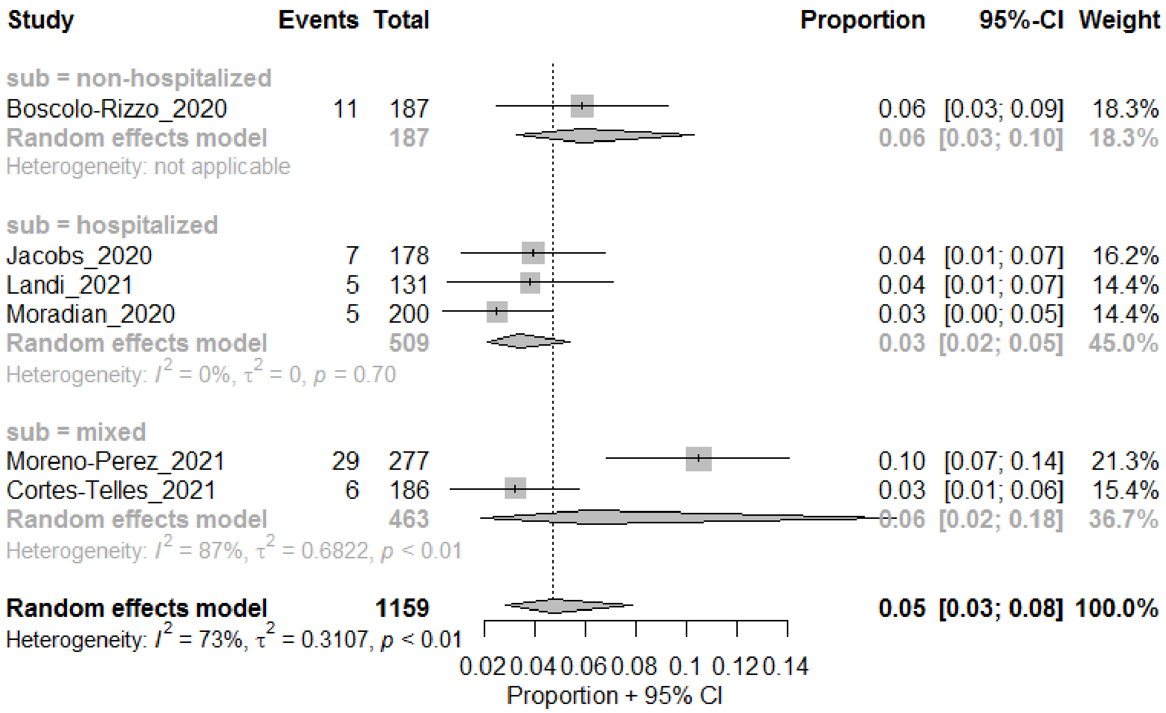

#### Nausea

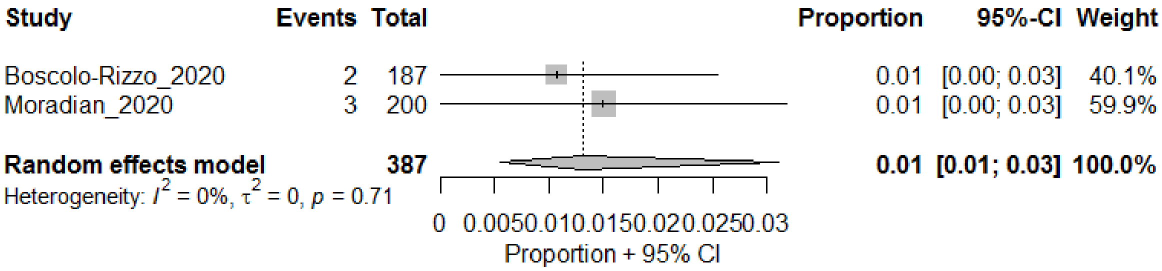

#### Vomiting

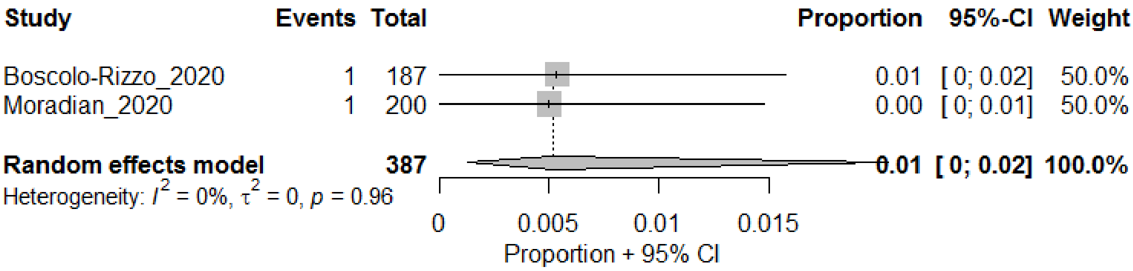

#### Weight loss

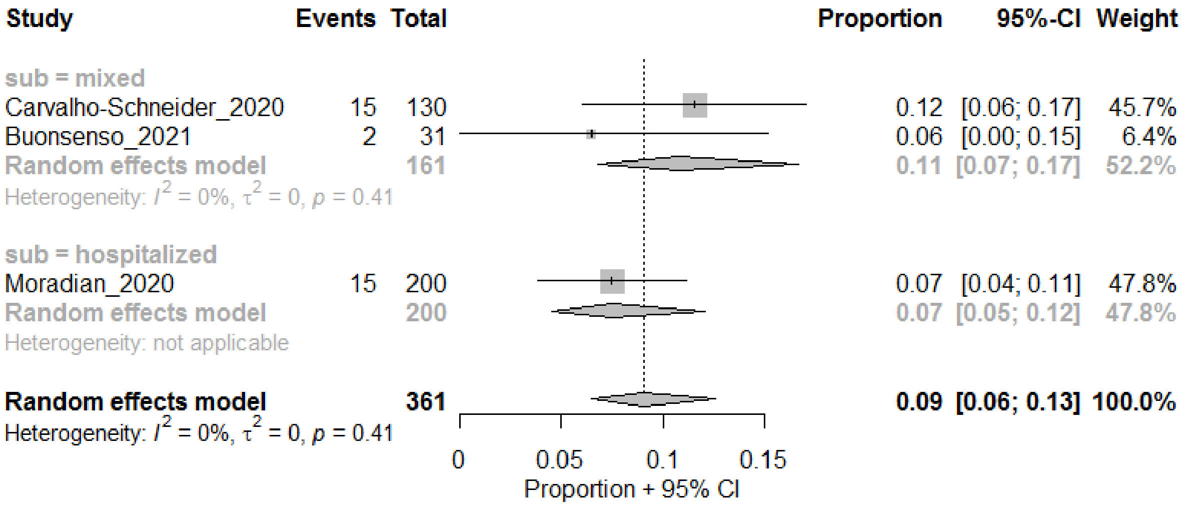

#### Eye-related

##### Eye irritations

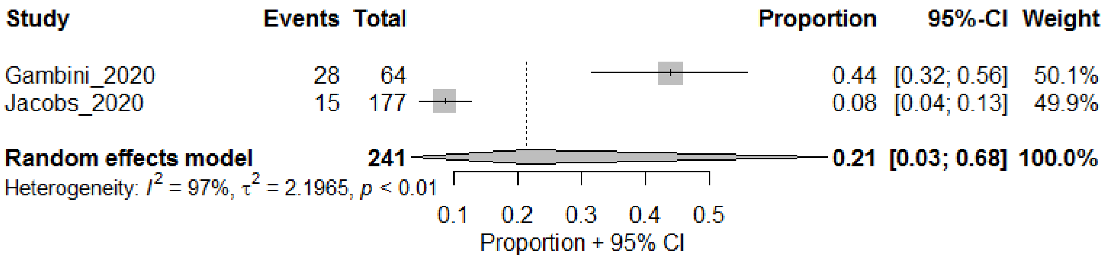

##### Red Eyes

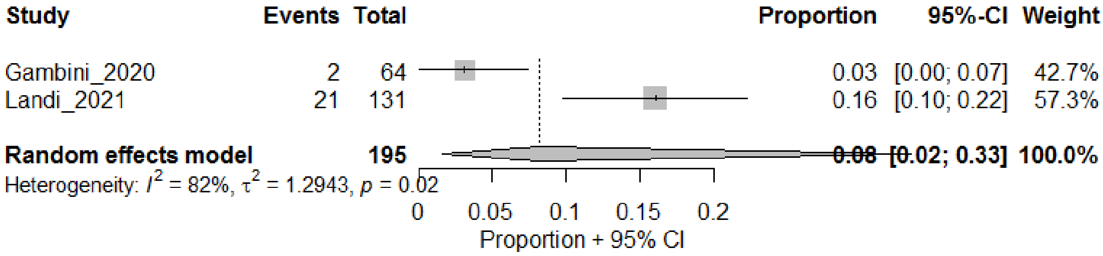

### Mental Health

#### Depression

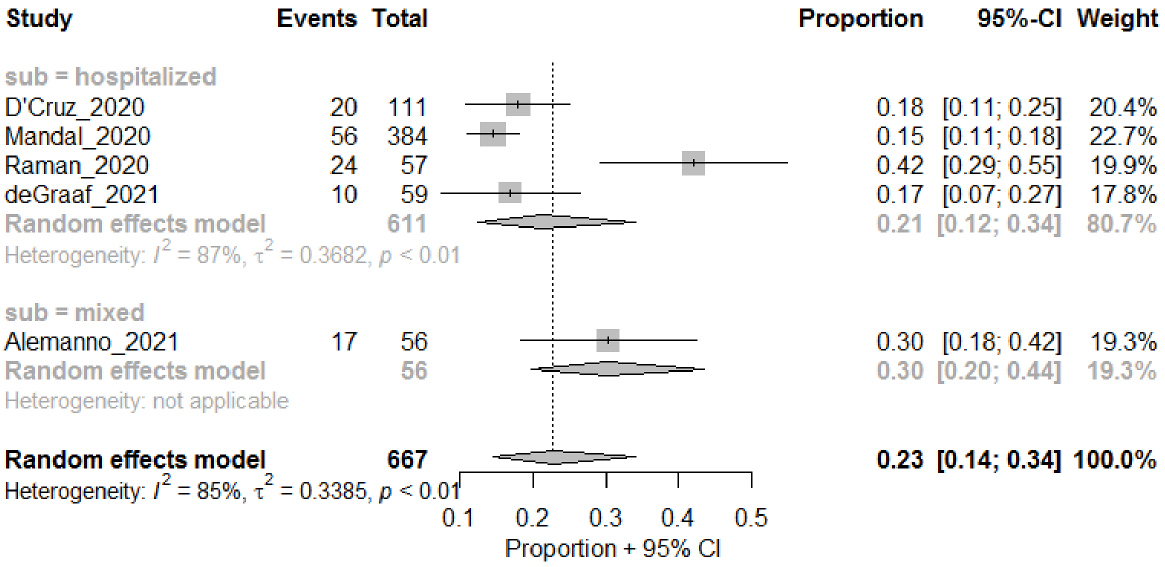

#### Anxiety

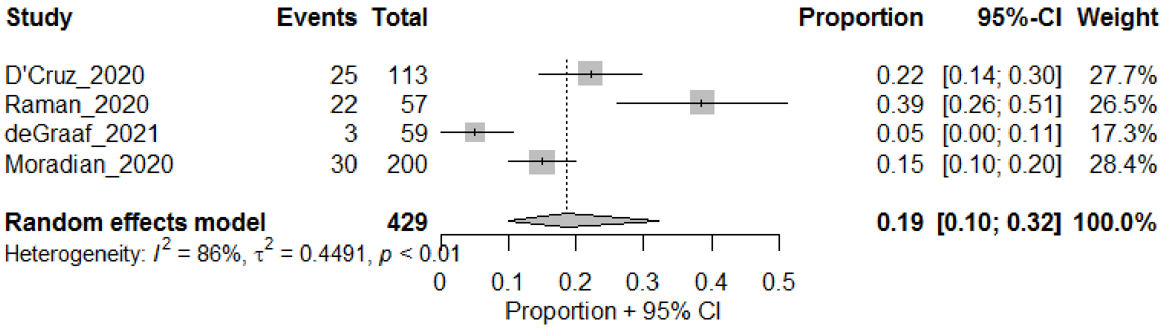

#### PTSD

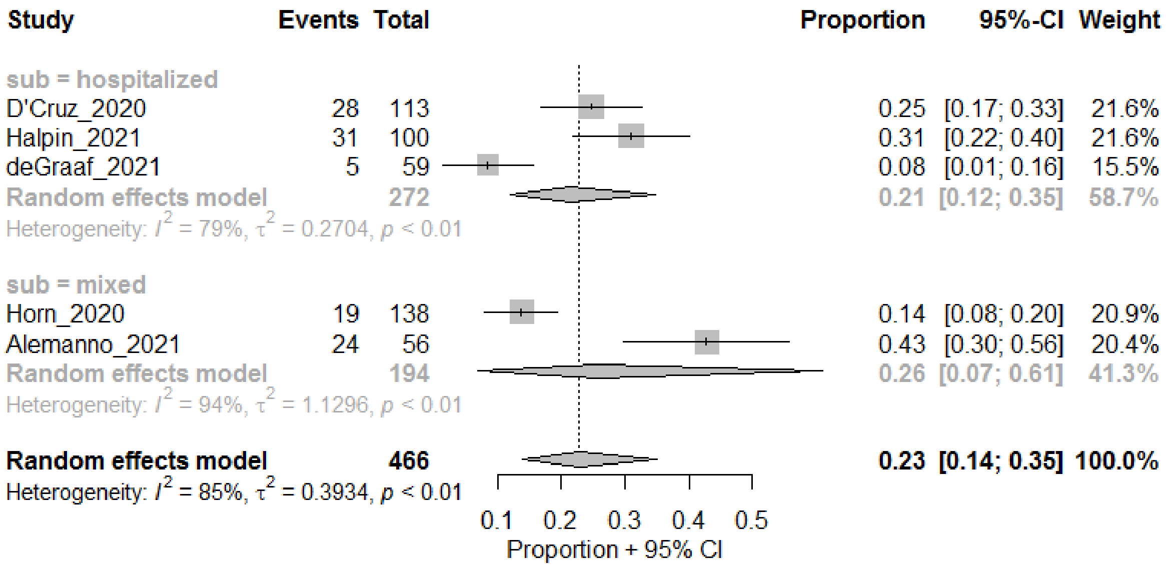

#### Anxiety or Depression

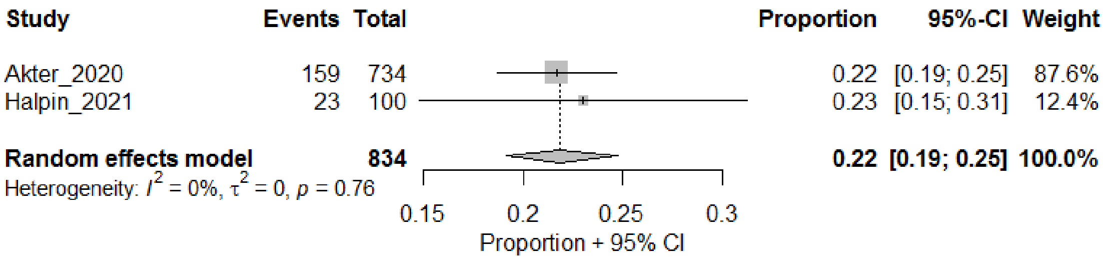

#### Insomnia and Sleep Disturbances

##### Sleep disturbances

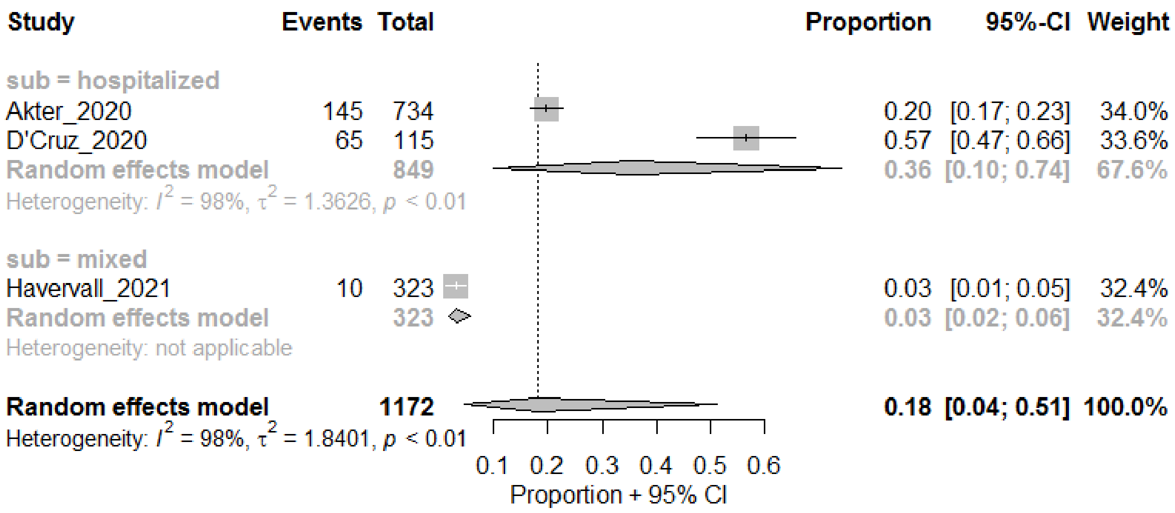
Study
Events
Total
Proportion
95%-CI
Weight
sub = hospitalized
Akter_2020 145 734 0.20 [0.17; 0.23] 34.0%
D'Cruz_2020 65 115 0.57 [0.47; 0.66] 33.6%
Random effects model 849 0.36 [0.10; 0.74] 67.6%
Heterogeneity: $I^2$ = 98%, $\tau^2$ = 1.3626, $p$ < 0.01
sub = mixed
Havervall_2021 10 323 0.03 [0.01; 0.05] 32.4%
Random effects model 323 0.03 [0.02; 0.06] 32.4%
Heterogeneity: not applicable
Random effects model 1172 0.18 [0.04; 0.51] 100.0%
Heterogeneity: $I^2$ = 98%, $\tau^2$ = 1.8401, $p$ < 0.01
0.1 0.2 0.3 0.4 0.5 0.6
Proportion + 95% CI

#### Insomnia

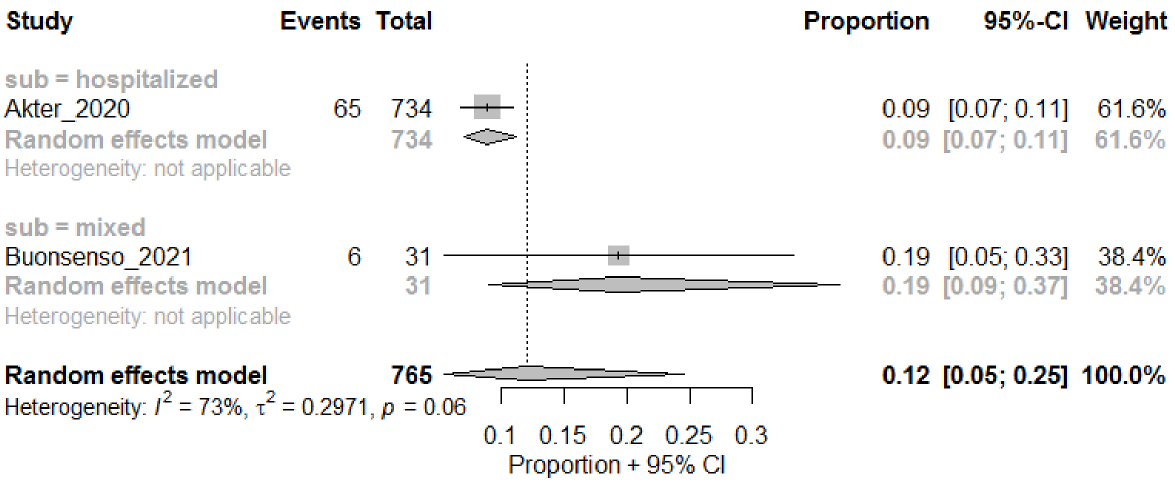
Study
Events
Total
Proportion
95%-CI
Weight
sub = hospitalized
Akter_2020 65 734 0.09 [0.07; 0.11] 61.6%
Random effects model 734 0.09 [0.07; 0.11] 61.6%
Heterogeneity: not applicable
sub = mixed
Buonsenso_2021 6 31 0.19 [0.05; 0.33] 38.4%
Random effects model 31 0.19 [0.09; 0.37] 38.4%
Heterogeneity: not applicable
Random effects model 765 0.12 [0.05; 0.25] 100.0%
Heterogeneity: $I^2$ = 73%, $\tau^2$ = 0.2971, $p$ = 0.06
0.1 0.15 0.2 0.25 0.3
Proportion + 95% CI

#### Overall Functioning

##### General functional impairment

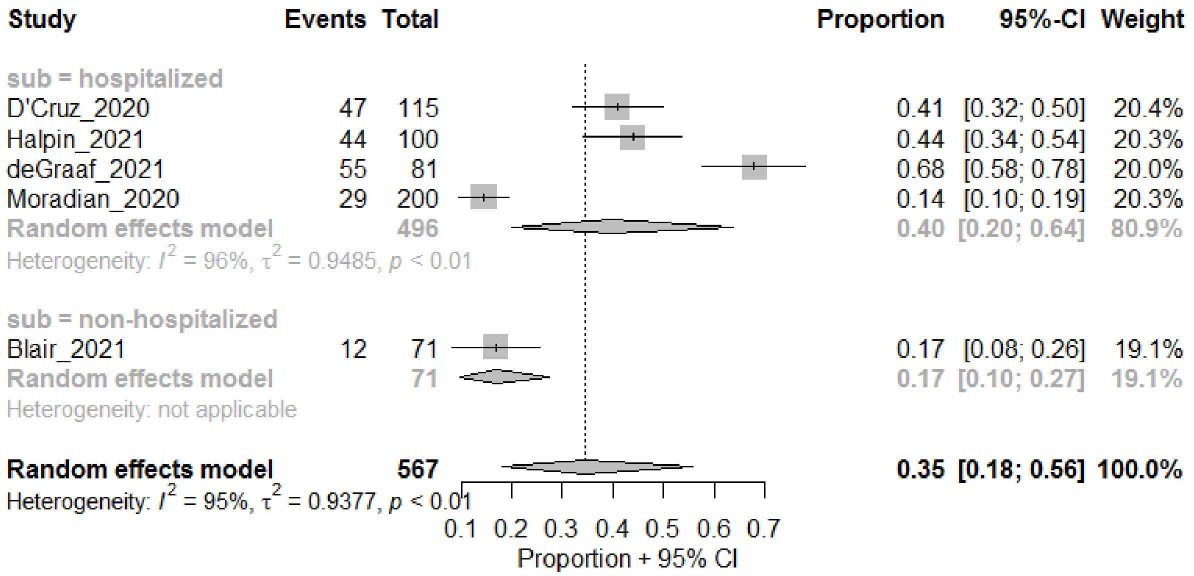

##### Still felt ill or not back to full health

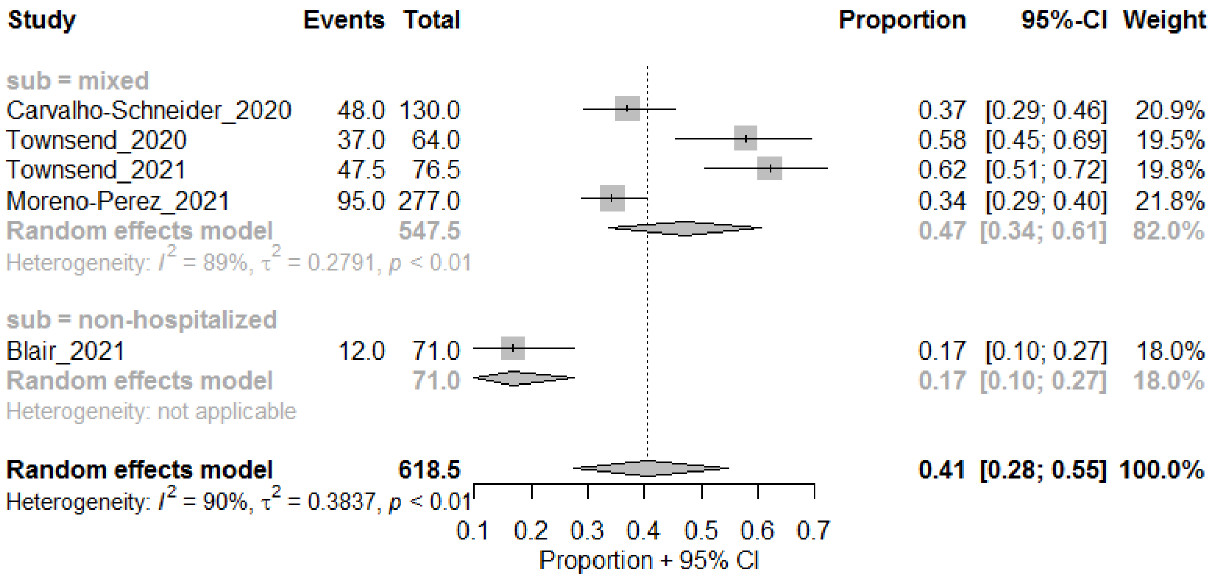

##### General perception of health (poor/fair)

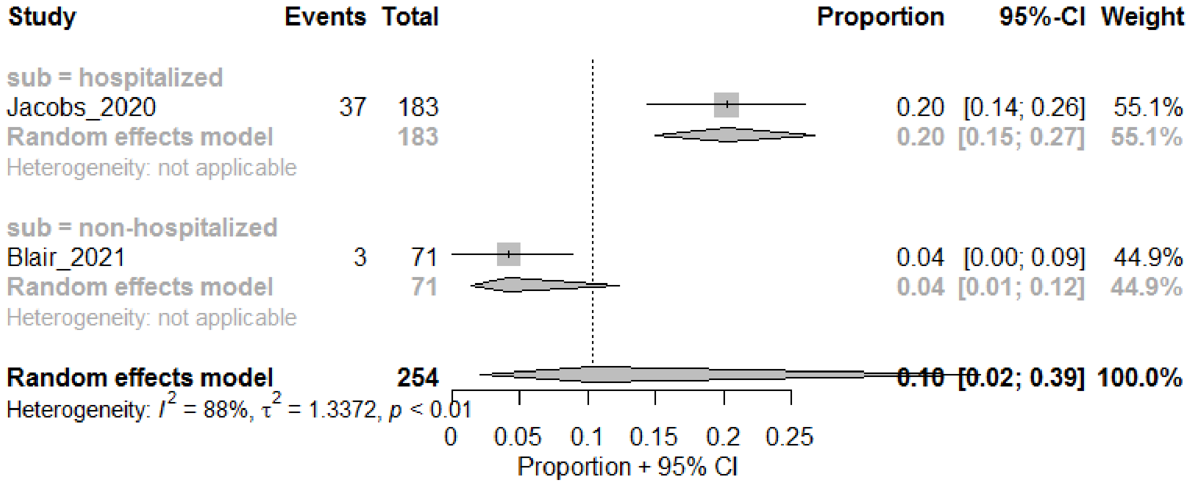

#### Mobility difficulties

##### *Walking*

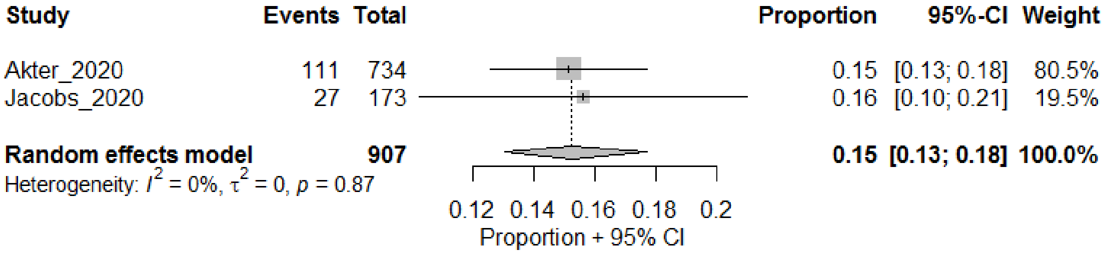

##### *Desaturation or abnormal results at the end of the 6-minute walk test*

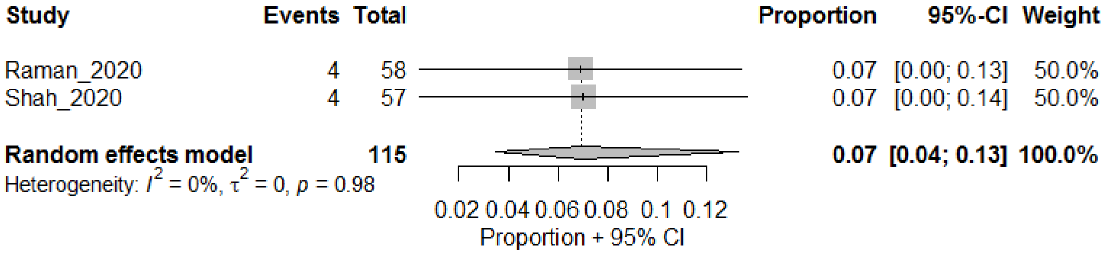

#### Others

##### General pain/discomfort

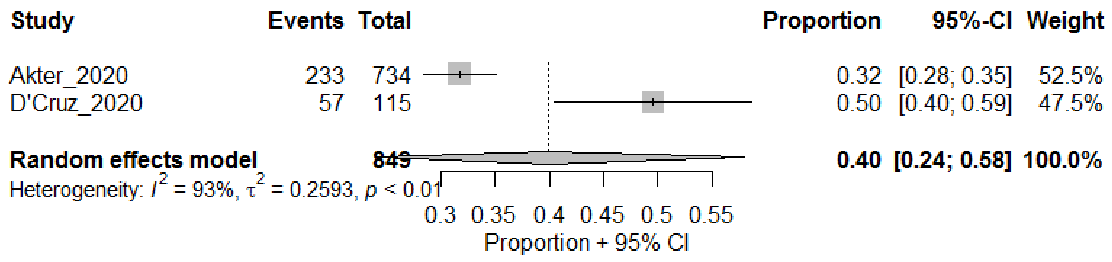

Hair loss

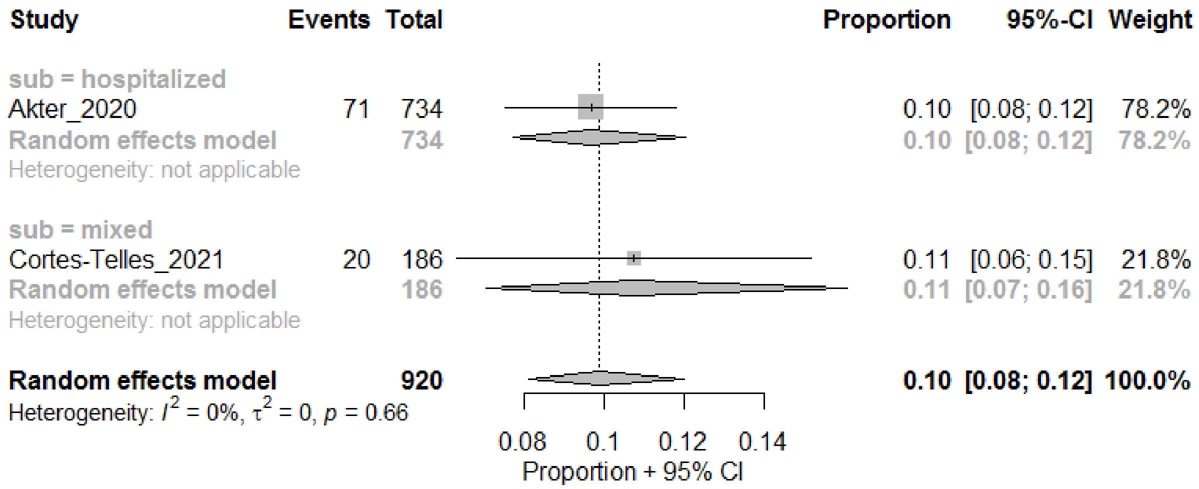

Skin-related

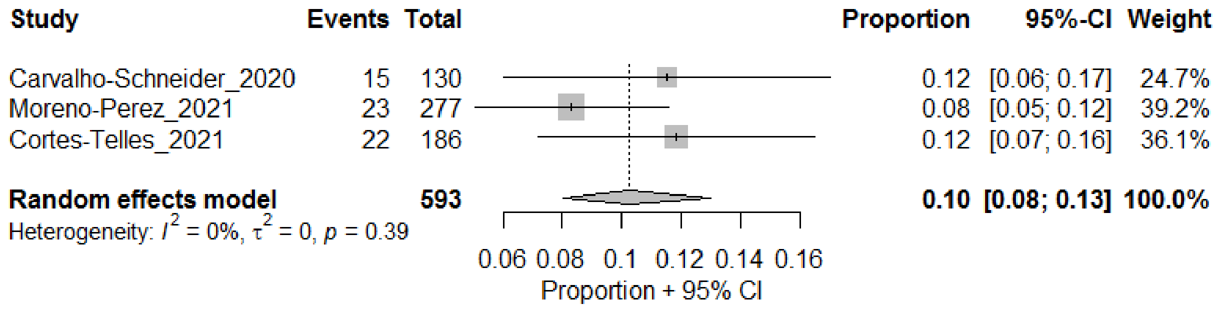

### LONG-TERM

#### One or more symptoms

##### Presence of one or more symptoms at follow-up

| Study | Events | Total | Proportion | 95%-CI | Weight |
|---|---|---|---|---|---|
| **sub = ICU** | | | | | |
| Alharthy_2020 | 62 | 127 | 0.49 | [0.40; 0.58] | 7.1% |
| Gonzalez_2021 | 43 | 59 | 0.73 | [0.62; 0.84] | 6.7% |
| **Random effects model** | | **186** | **0.61** | **[0.36; 0.81]** | **13.8%** |
| Heterogeneity: $I^2 = 89\%$, $\tau^2 = 0.4779$, $p < 0.01$ | | | | | |
| **sub = hospitalized** | | | | | |
| Huang_2020 | 1265 | 1655 | 0.76 | [0.74; 0.78] | 7.4% |
| Qu_2021 | 311 | 540 | 0.58 | [0.53; 0.62] | 7.3% |
| Sykes_2021 | 115 | 134 | 0.86 | [0.80; 0.92] | 6.9% |
| **Random effects model** | | **2329** | **0.74** | **[0.59; 0.86]** | **21.6%** |
| Heterogeneity: $I^2 = 98\%$, $\tau^2 = 0.3842$, $p < 0.01$ | | | | | |
| **sub = mixed** | | | | | |
| Petersen_2020 | 95 | 179 | 0.53 | [0.46; 0.60] | 7.2% |
| Logue_2021 | 53 | 177 | 0.30 | [0.23; 0.37] | 7.2% |
| Havervall_2021 | 48 | 323 | 0.15 | [0.11; 0.19] | 7.2% |
| Buonsenso_2021 | 55 | 98 | 0.56 | [0.46; 0.66] | 7.1% |
| Jacobson_2021 | 79 | 118 | 0.67 | [0.58; 0.75] | 7.1% |
| Klein_2021 | 47 | 103 | 0.46 | [0.36; 0.55] | 7.1% |
| Anastasio_2021 | 260 | 379 | 0.69 | [0.64; 0.73] | 7.3% |
| Trunfio_2021 | 41 | 168 | 0.24 | [0.18; 0.31] | 7.1% |
| **Random effects model** | | **1545** | **0.44** | **[0.28; 0.60]** | **57.2%** |
| Heterogeneity: $I^2 = 97\%$, $\tau^2 = 0.9167$, $p < 0.01$ | | | | | |
| **sub = non-hospitalized** | | | | | |
| Stavem_2020 | 185 | 451 | 0.41 | [0.36; 0.46] | 7.3% |
| **Random effects model** | | **451** | **0.41** | **[0.37; 0.46]** | **7.3%** |
| Heterogeneity: not applicable | | | | | |
| **Random effects model** | | **4511** | **0.53** | **[0.41; 0.65]** | **100.0%** |
| Heterogeneity: $I^2 = 98\%$, $\tau^2 = 0.8118$, $p < 0.01$ | | | | | |

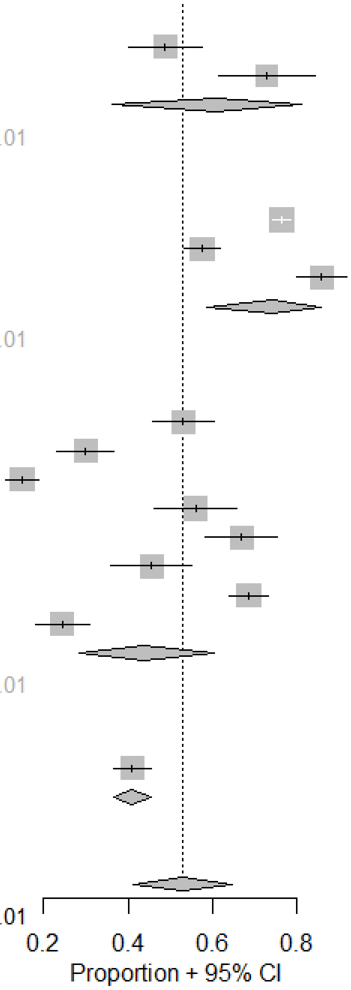

#### Number of symptoms – 1-2

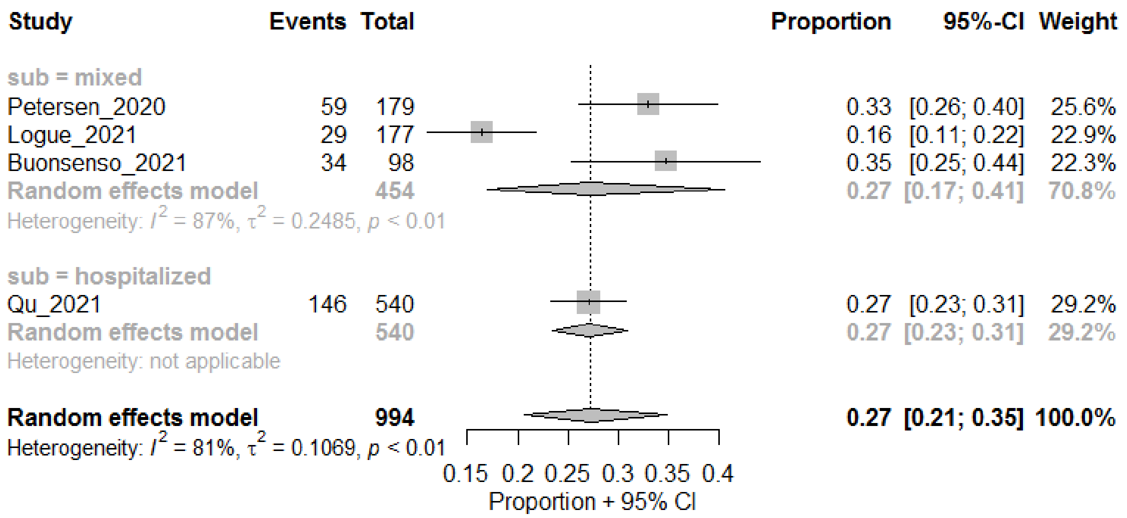

#### Number of symptoms – >=3

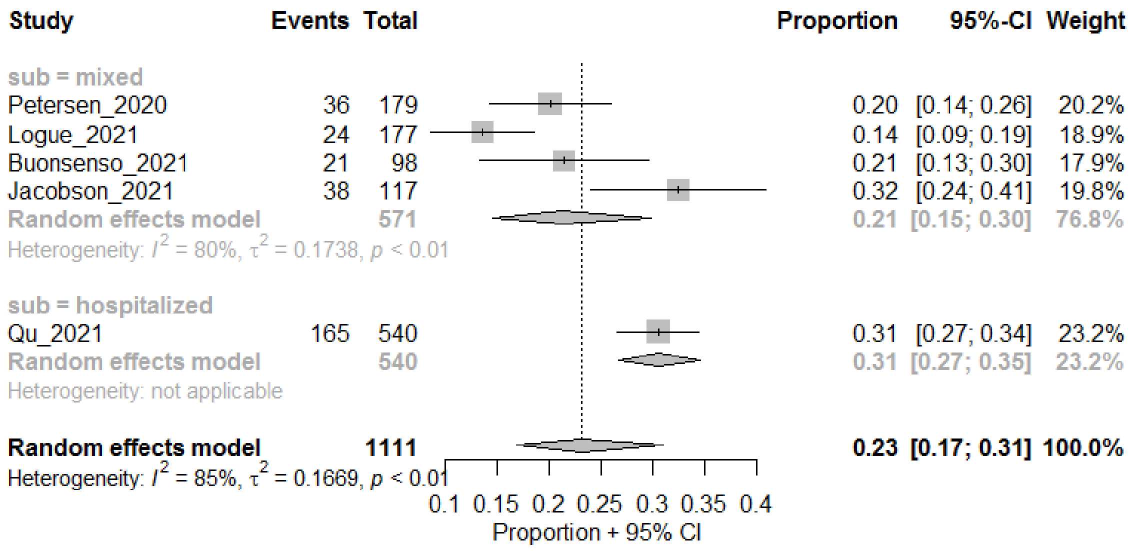

#### Fatigue

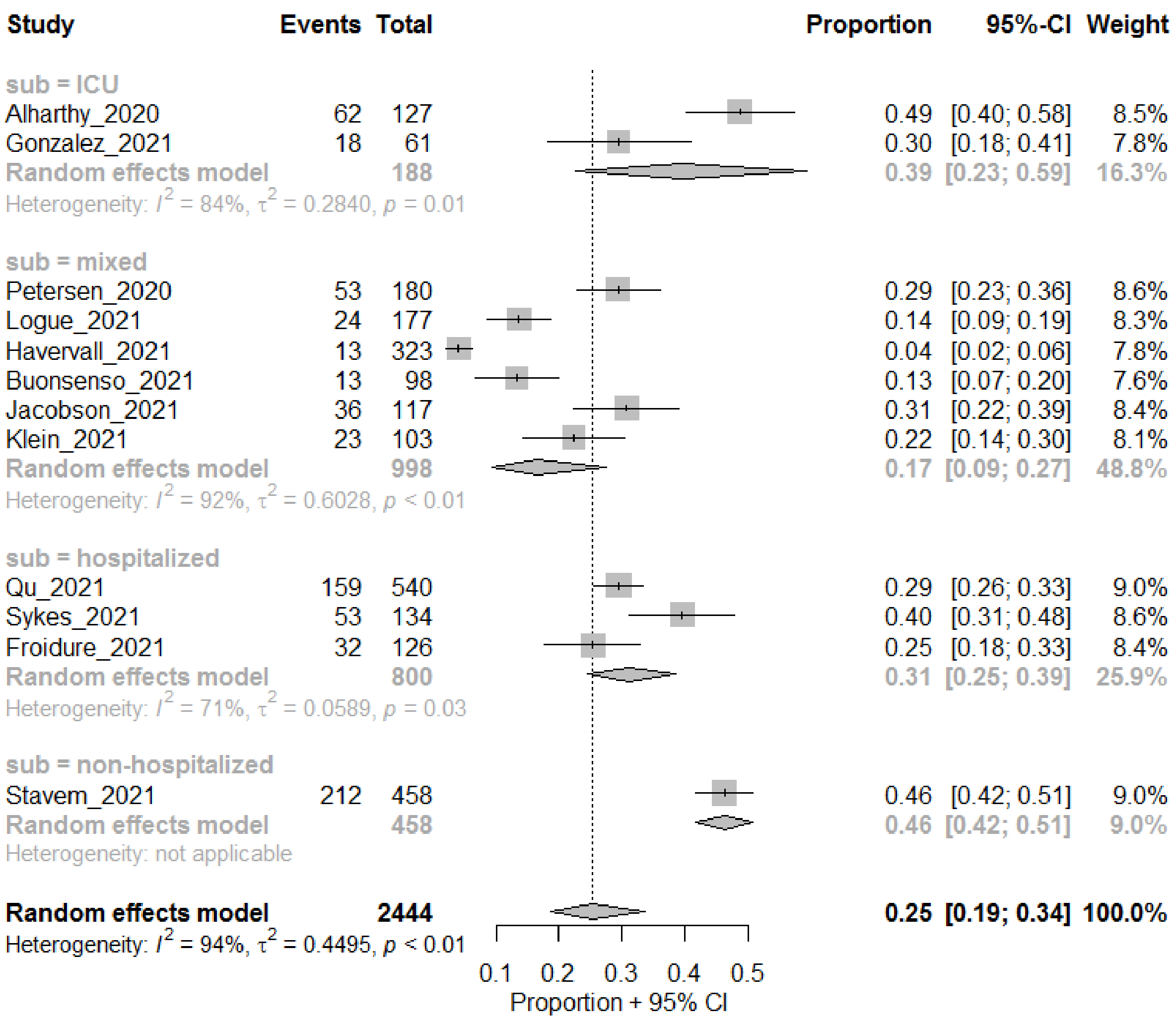

#### Respiratory

##### Shortness of Breath

*Two  subgroups for hospitalized*

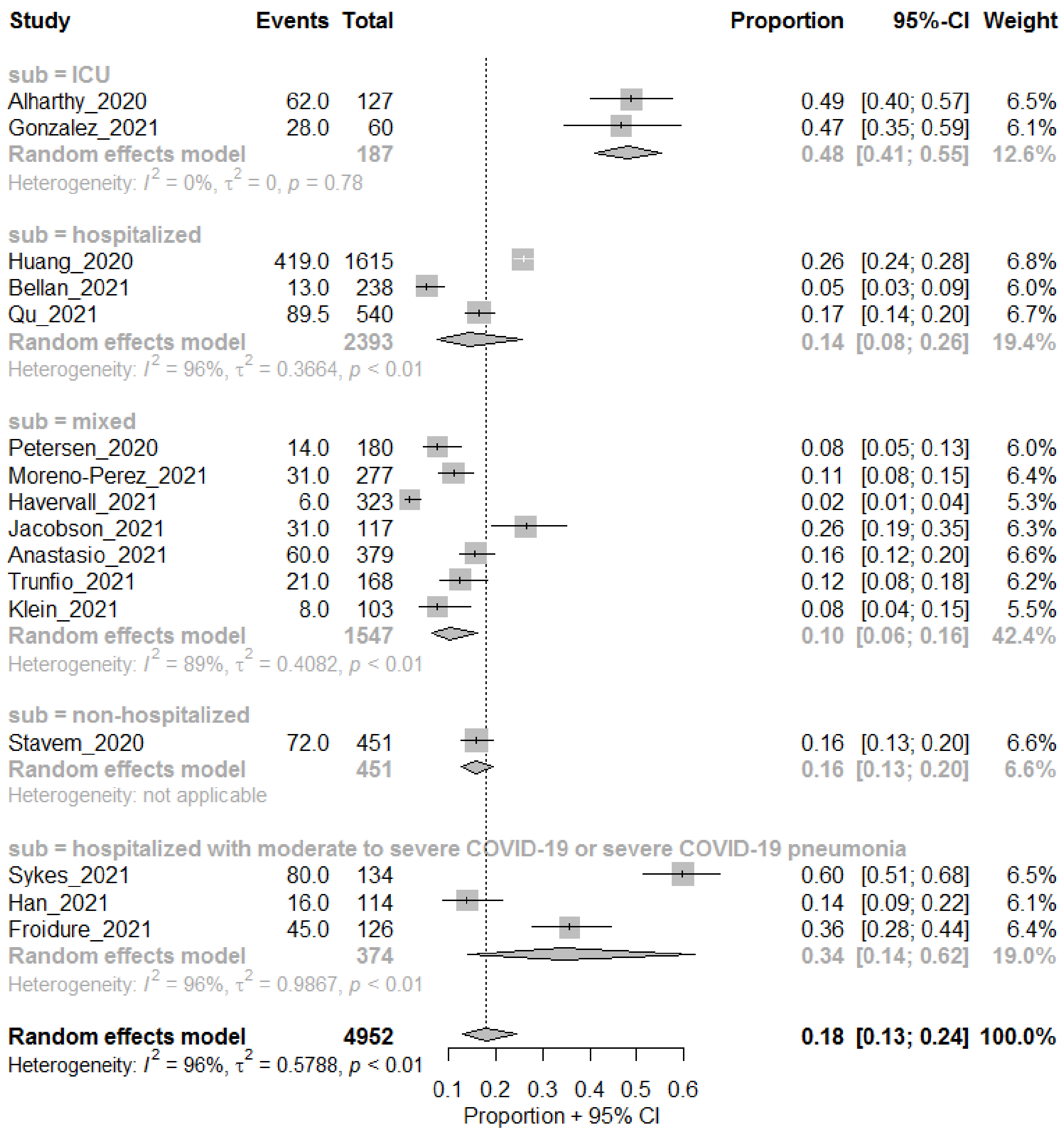

*no subgroups for hospitalized*

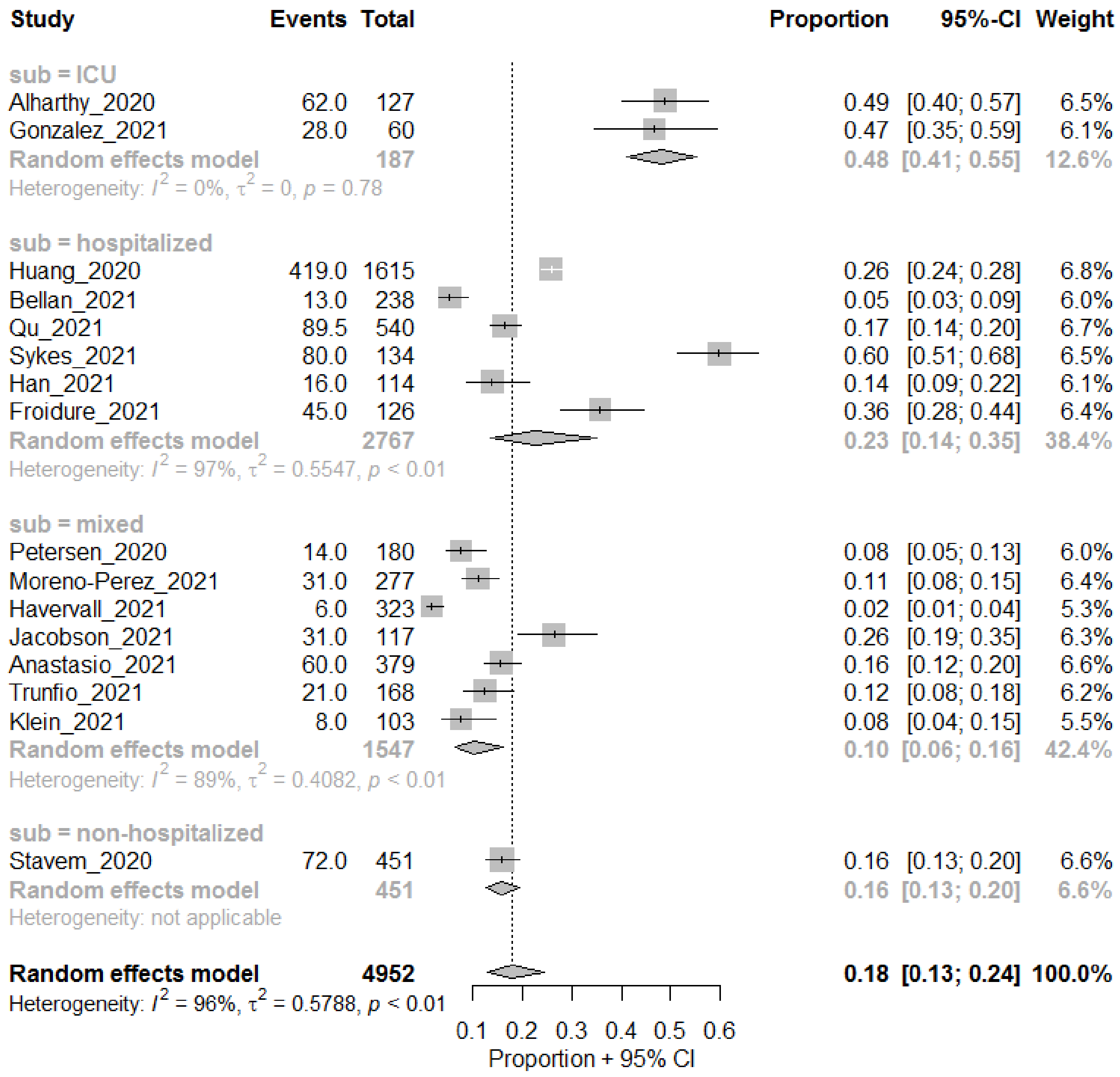
Study Events Total Proportion 95%-CI Weight
sub = ICU
Alharthy_2020 62.0 127 0.49 [0.40; 0.57] 6.5%
Gonzalez_2021 28.0 60 0.47 [0.35; 0.59] 6.1%
Random effects model 187 0.48 [0.41; 0.55] 12.6%
Heterogeneity: $I^2$ = 0%, $\tau^2$ = 0, $p$ = 0.78
sub = hospitalized
Huang_2020 419.0 1615 0.26 [0.24; 0.28] 6.8%
Bellan_2021 13.0 238 0.05 [0.03; 0.09] 6.0%
Qu_2021 89.5 540 0.17 [0.14; 0.20] 6.7%
Sykes_2021 80.0 134 0.60 [0.51; 0.68] 6.5%
Han_2021 16.0 114 0.14 [0.09; 0.22] 6.1%
Froidure_2021 45.0 126 0.36 [0.28; 0.44] 6.4%
Random effects model 2767 0.23 [0.14; 0.35] 38.4%
Heterogeneity: $I^2$ = 97%, $\tau^2$ = 0.5547, $p$ < 0.01
sub = mixed
Petersen_2020 14.0 180 0.08 [0.05; 0.13] 6.0%
Moreno-Perez_2021 31.0 277 0.11 [0.08; 0.15] 6.4%
Havervall_2021 6.0 323 0.02 [0.01; 0.04] 5.3%
Jacobson_2021 31.0 117 0.26 [0.19; 0.35] 6.3%
Anastasio_2021 60.0 379 0.16 [0.12; 0.20] 6.6%
Trunfio_2021 21.0 168 0.12 [0.08; 0.18] 6.2%
Klein_2021 8.0 103 0.08 [0.04; 0.15] 5.5%
Random effects model 1547 0.10 [0.06; 0.16] 42.4%
Heterogeneity: $I^2$ = 89%, $\tau^2$ = 0.4082, $p$ < 0.01
sub = non-hospitalized
Stavem_2020 72.0 451 0.16 [0.13; 0.20] 6.6%
Random effects model 451 0.16 [0.13; 0.20] 6.6%
Heterogeneity: not applicable
Random effects model 4952 0.18 [0.13; 0.24] 100.0%
Heterogeneity: $I^2$ = 96%, $\tau^2$ = 0.5788, $p$ < 0.01
0.1 0.2 0.3 0.4 0.5 0.6
Proportion + 95% CI

#### Runny nose

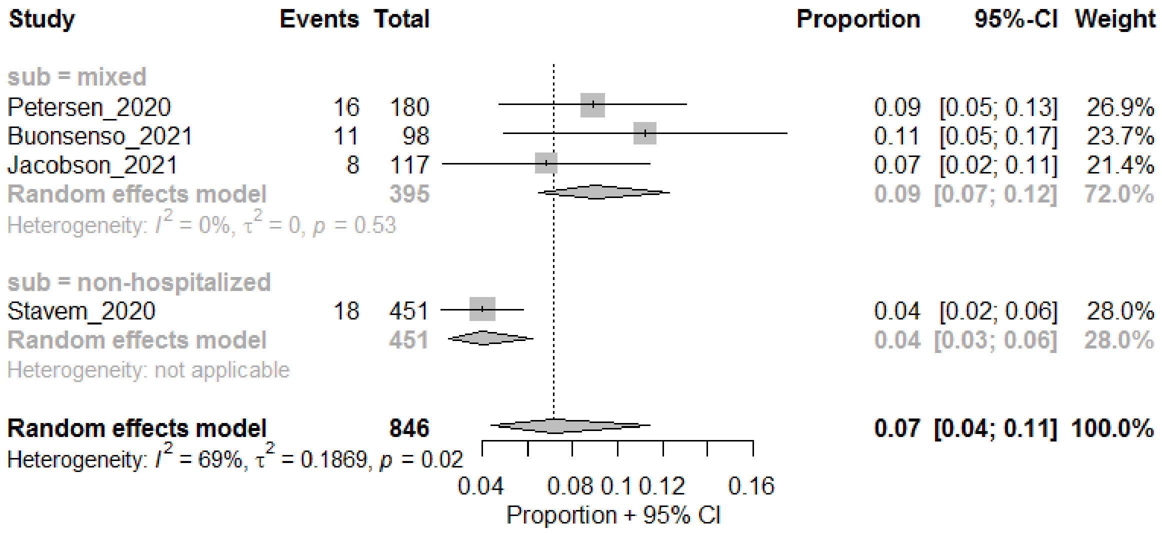
Study Events Total Proportion 95%-CI Weight
sub = mixed
Petersen_2020 16 180 0.09 [0.05; 0.13] 26.9%
Buonsenso_2021 11 98 0.11 [0.05; 0.17] 23.7%
Jacobson_2021 8 117 0.07 [0.02; 0.11] 21.4%
Random effects model 395 0.09 [0.07; 0.12] 72.0%
Heterogeneity: $I^2$ = 0%, $\tau^2$ = 0, $p$ = 0.53
sub = non-hospitalized
Stavem_2020 18 451 0.04 [0.02; 0.06] 28.0%
Random effects model 451 0.04 [0.03; 0.06] 28.0%
Heterogeneity: not applicable
Random effects model 846 0.07 [0.04; 0.11] 100.0%
Heterogeneity: $I^2$ = 69%, $\tau^2$ = 0.1869, $p$ = 0.02
0.04 0.08 0.1 0.12 0.16
Proportion + 95% CI

#### Any cough

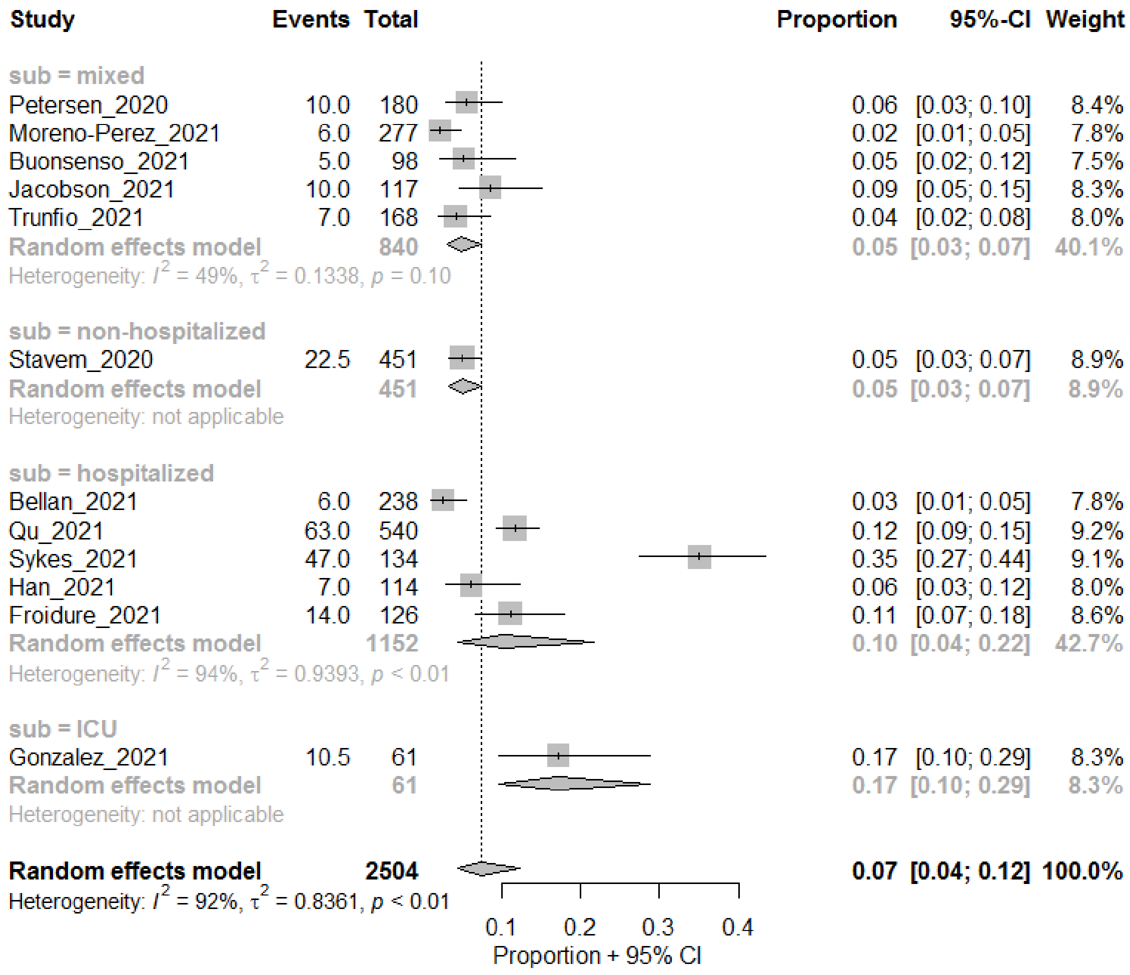
Study Events Total Proportion 95%-CI Weight
sub = mixed
Petersen_2020 10.0 180 0.06 [0.03; 0.10] 8.4%
Moreno-Perez_2021 6.0 277 0.02 [0.01; 0.05] 7.8%
Buonsenso_2021 5.0 98 0.05 [0.02; 0.12] 7.5%
Jacobson_2021 10.0 117 0.09 [0.05; 0.15] 8.3%
Trunfio_2021 7.0 168 0.04 [0.02; 0.08] 8.0%
Random effects model 840 0.05 [0.03; 0.07] 40.1%
Heterogeneity: $I^2$ = 49%, $\tau^2$ = 0.1338, $p$ = 0.10
sub = non-hospitalized
Stavem_2020 22.5 451 0.05 [0.03; 0.07] 8.9%
Random effects model 451 0.05 [0.03; 0.07] 8.9%
Heterogeneity: not applicable
sub = hospitalized
Bellan_2021 6.0 238 0.03 [0.01; 0.05] 7.8%
Qu_2021 63.0 540 0.12 [0.09; 0.15] 9.2%
Sykes_2021 47.0 134 0.35 [0.27; 0.44] 9.1%
Han_2021 7.0 114 0.06 [0.03; 0.12] 8.0%
Froidure_2021 14.0 126 0.11 [0.07; 0.18] 8.6%
Random effects model 1152 0.10 [0.04; 0.22] 42.7%
Heterogeneity: $I^2$ = 94%, $\tau^2$ = 0.9393, $p$ < 0.01
sub = ICU
Gonzalez_2021 10.5 61 0.17 [0.10; 0.29] 8.3%
Random effects model 61 0.17 [0.10; 0.29] 8.3%
Heterogeneity: not applicable
Random effects model 2504 0.07 [0.04; 0.12] 100.0%
Heterogeneity: $I^2$ = 92%, $\tau^2$ = 0.8361, $p$ < 0.01
0.1 0.2 0.3 0.4
Proportion + 95% CI

#### Dry cough

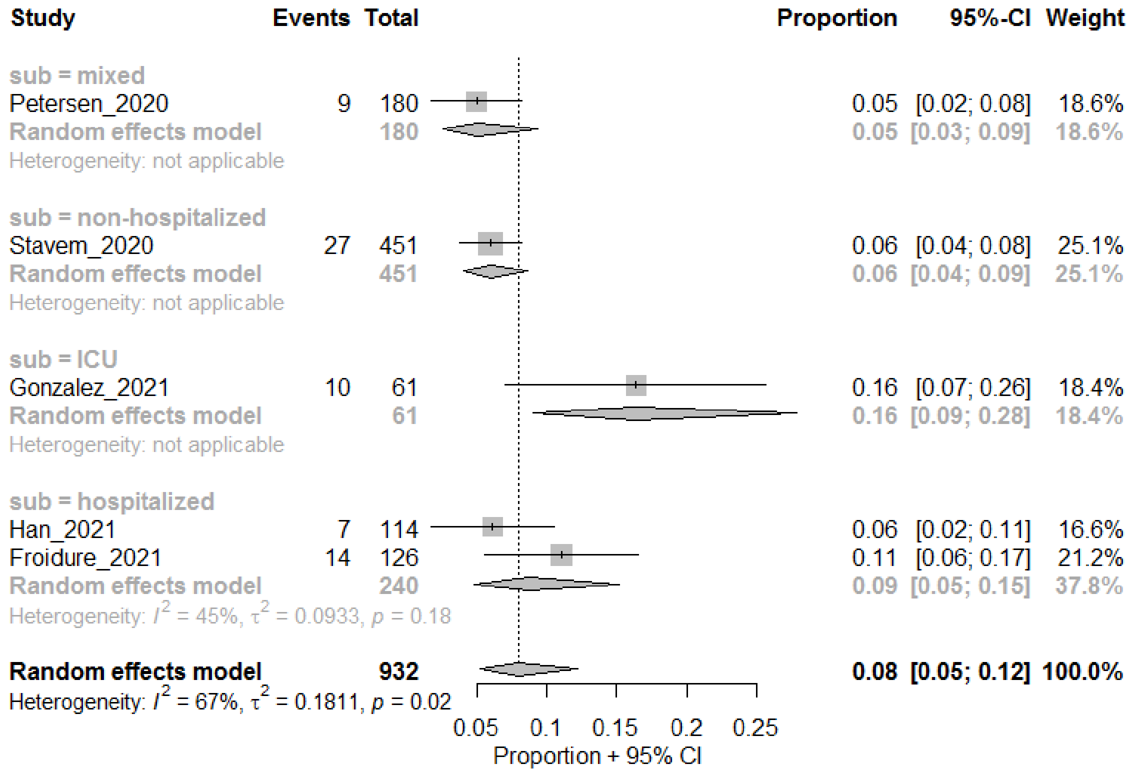

#### Productive cough

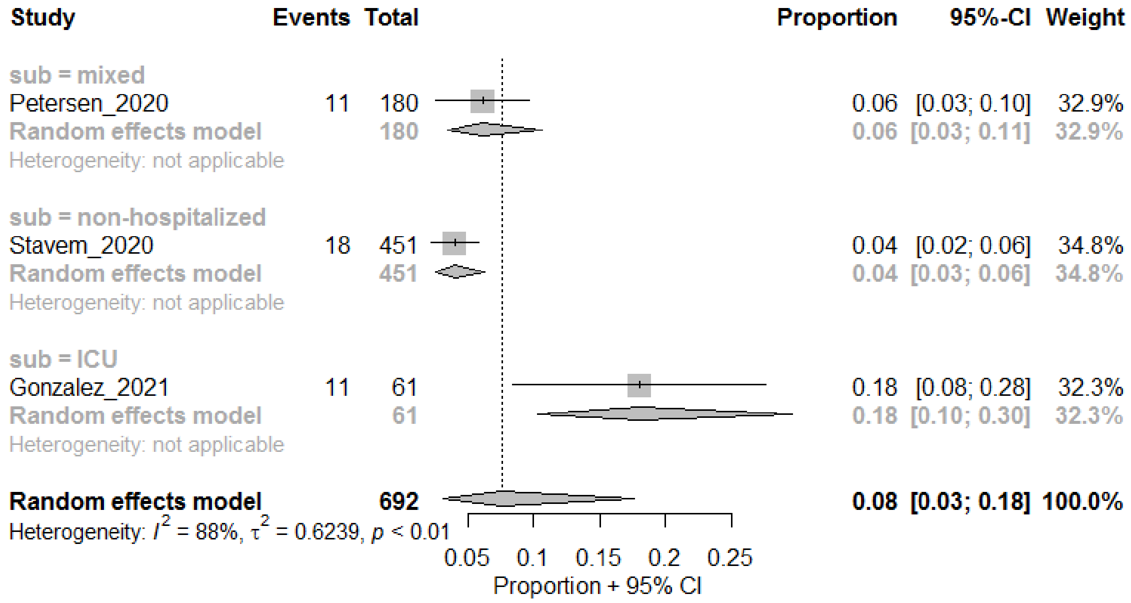

#### Sore throat

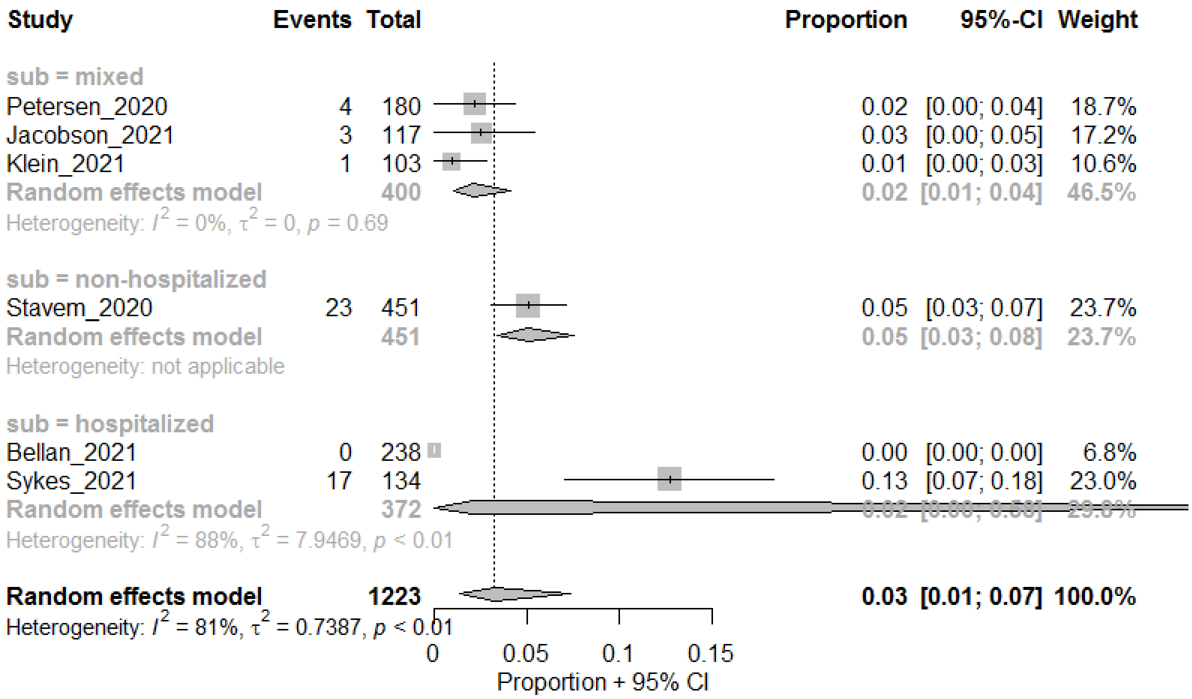

Phlegm

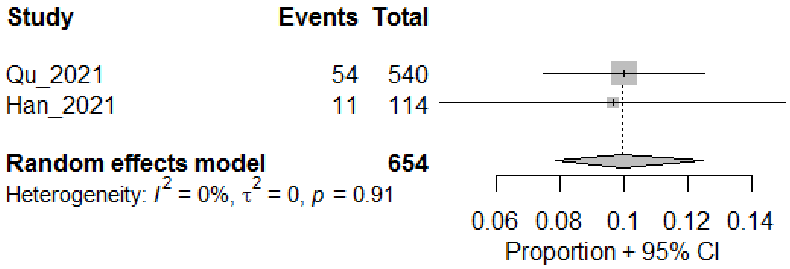

Wheezing

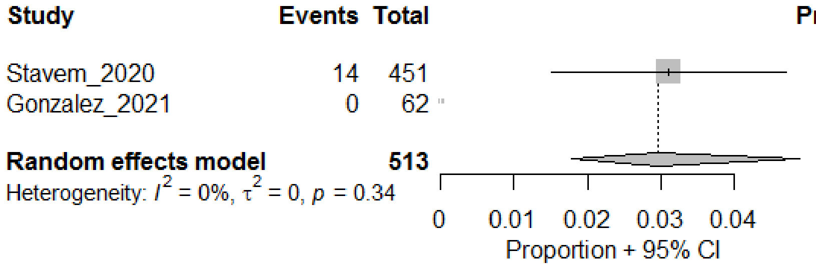

#### Influenza-like illness

##### Headaches

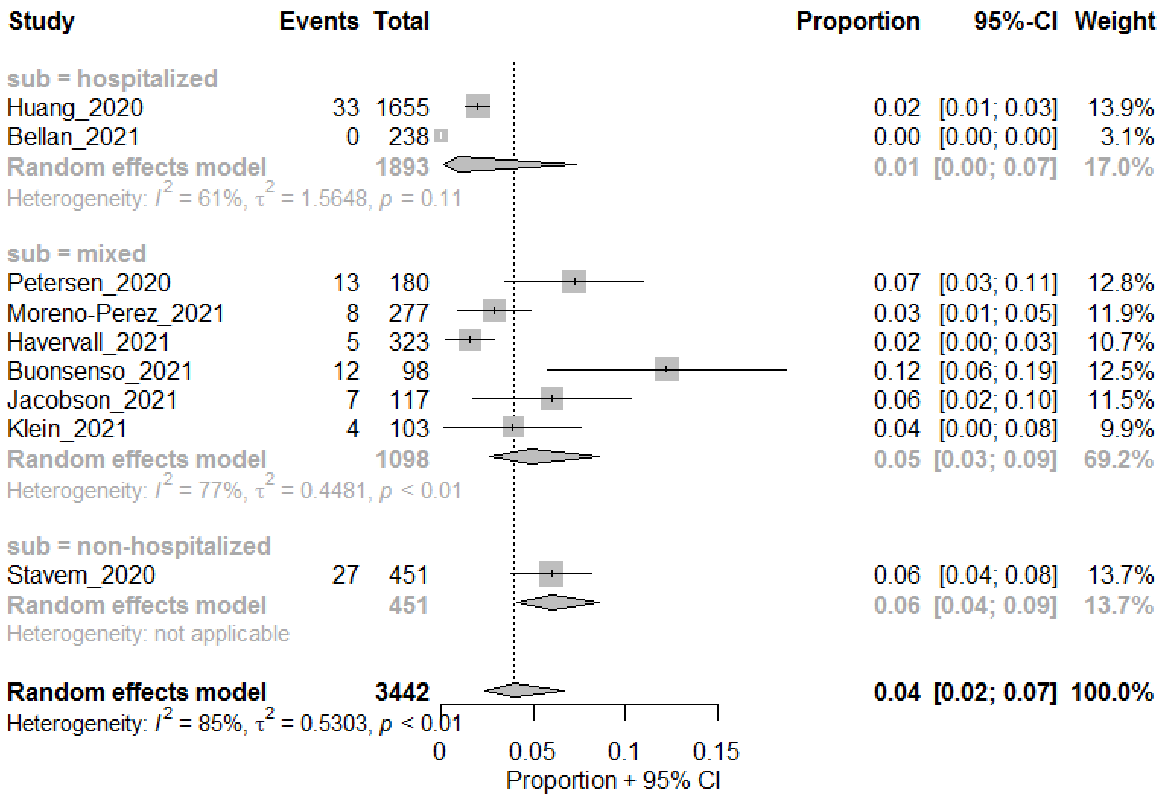

##### Chills

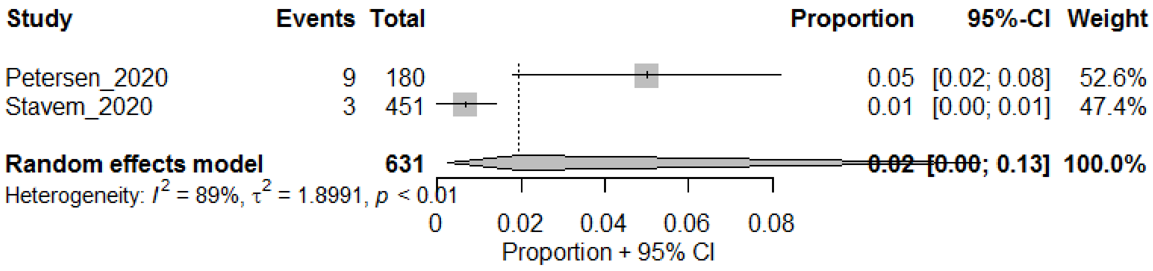

##### Fever

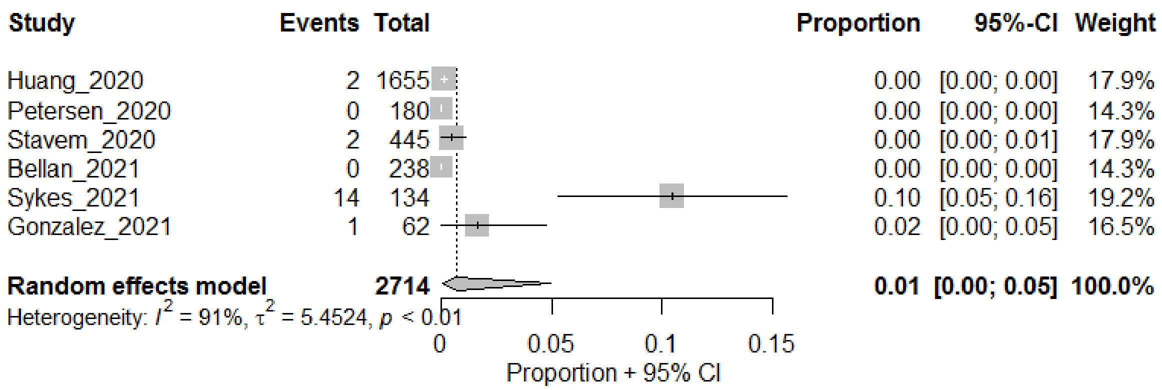

#### Olfactory and Taste Impairments

##### Any Smell dysfunction

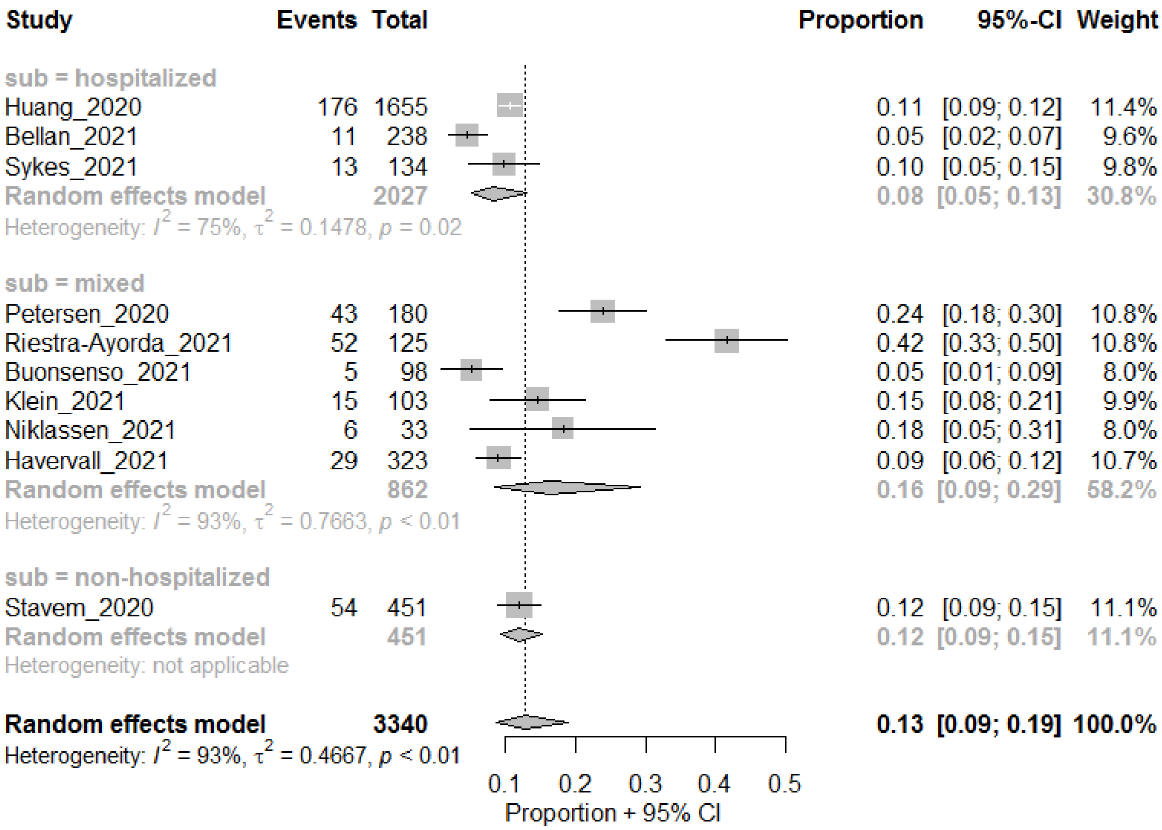

##### Anosmia

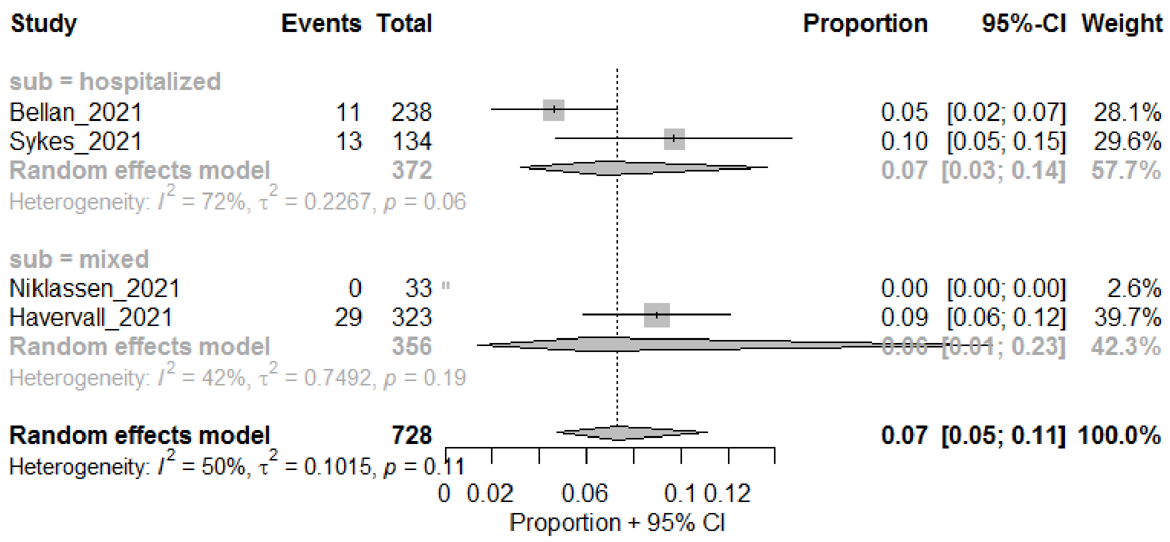

#### Any Taste dysfunction

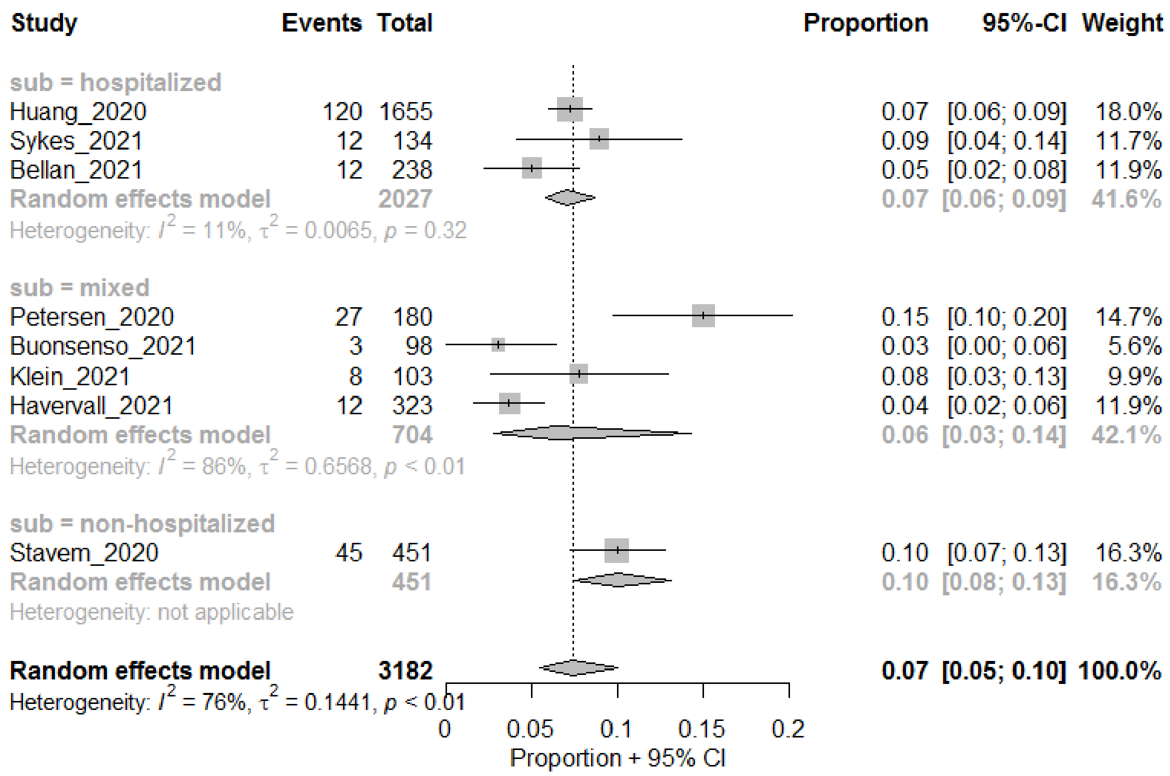

#### Ageusia

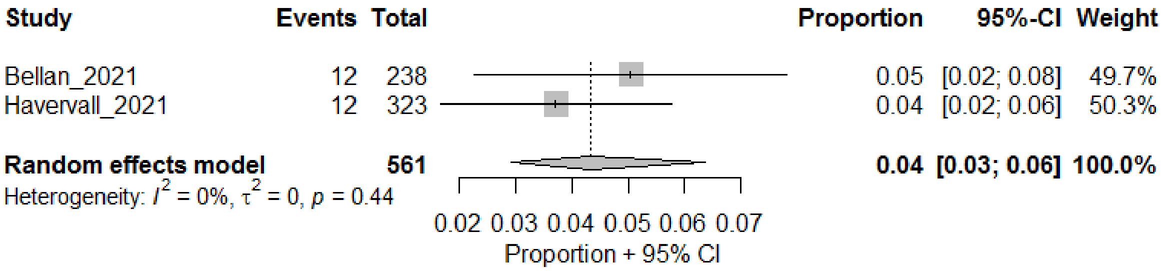

#### Any Smell or Taste dysfunction

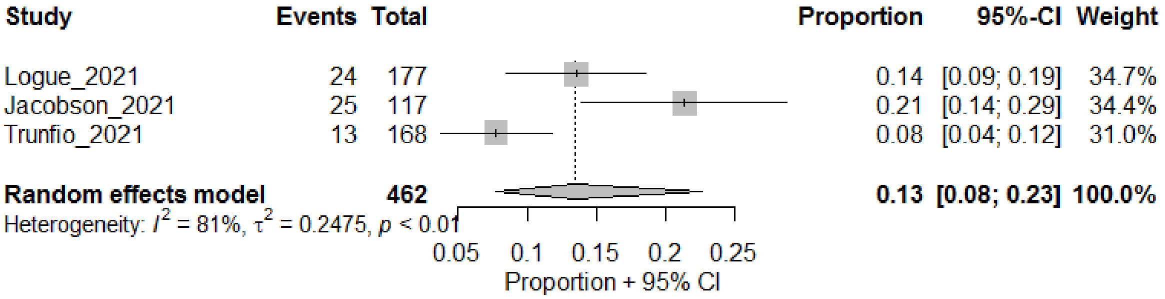

### Neuromuscular

#### Joint pain

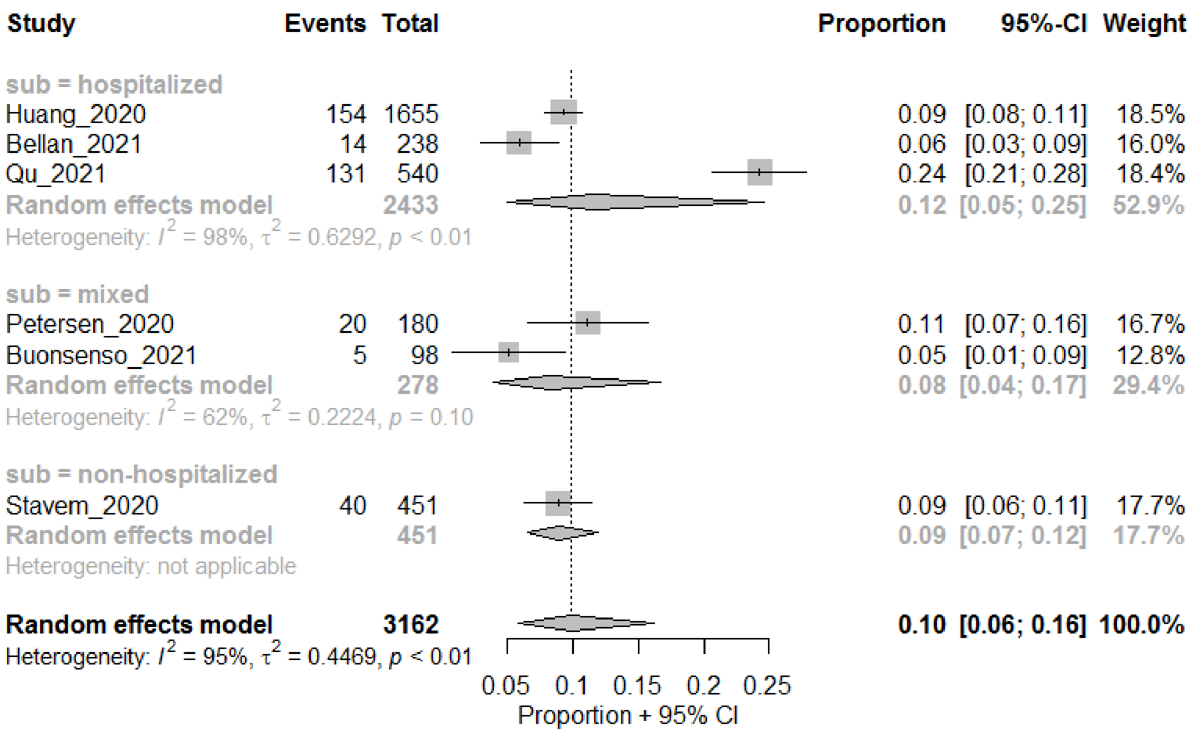
Study Events Total Proportion 95%-CI Weight
sub = hospitalized
Huang_2020 154 1655 0.09 [0.08; 0.11] 18.5%
Bellan_2021 14 238 0.06 [0.03; 0.09] 16.0%
Qu_2021 131 540 0.24 [0.21; 0.28] 18.4%
Random effects model 2433 0.12 [0.05; 0.25] 52.9%
Heterogeneity: $I^2$ = 98%, $\tau^2$ = 0.6292, $p$ < 0.01
sub = mixed
Petersen_2020 20 180 0.11 [0.07; 0.16] 16.7%
Buonsenso_2021 5 98 0.05 [0.01; 0.09] 12.8%
Random effects model 278 0.08 [0.04; 0.17] 29.4%
Heterogeneity: $I^2$ = 62%, $\tau^2$ = 0.2224, $p$ = 0.10
sub = non-hospitalized
Stavem_2020 40 451 0.09 [0.06; 0.11] 17.7%
Random effects model 451 0.09 [0.07; 0.12] 17.7%
Heterogeneity: not applicable
Random effects model 3162 0.10 [0.06; 0.16] 100.0%
Heterogeneity: $I^2$ = 95%, $\tau^2$ = 0.4469, $p$ < 0.01
0.05 0.1 0.15 0.2 0.25
Proportion + 95% CI

#### Muscle pain

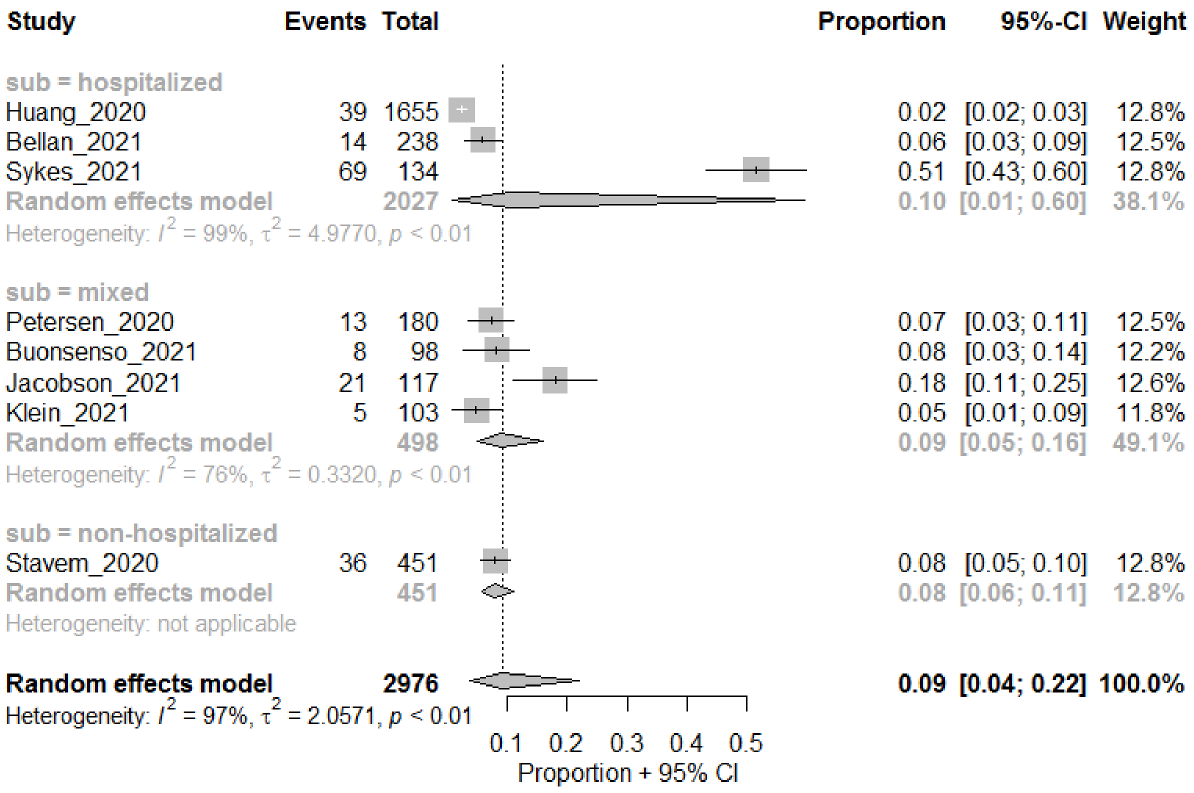
Study Events Total Proportion 95%-CI Weight
sub = hospitalized
Huang_2020 39 1655 0.02 [0.02; 0.03] 12.8%
Bellan_2021 14 238 0.06 [0.03; 0.09] 12.5%
Sykes_2021 69 134 0.51 [0.43; 0.60] 12.8%
Random effects model 2027 0.10 [0.01; 0.60] 38.1%
Heterogeneity: $I^2$ = 99%, $\tau^2$ = 4.9770, $p$ < 0.01
sub = mixed
Petersen_2020 13 180 0.07 [0.03; 0.11] 12.5%
Buonsenso_2021 8 98 0.08 [0.03; 0.14] 12.2%
Jacobson_2021 21 117 0.18 [0.11; 0.25] 12.6%
Klein_2021 5 103 0.05 [0.01; 0.09] 11.8%
Random effects model 498 0.09 [0.05; 0.16] 49.1%
Heterogeneity: $I^2$ = 76%, $\tau^2$ = 0.3320, $p$ < 0.01
sub = non-hospitalized
Stavem_2020 36 451 0.08 [0.05; 0.10] 12.8%
Random effects model 451 0.08 [0.06; 0.11] 12.8%
Heterogeneity: not applicable
Random effects model 2976 0.09 [0.04; 0.22] 100.0%
Heterogeneity: $I^2$ = 97%, $\tau^2$ = 2.0571, $p$ < 0.01
0.1 0.2 0.3 0.4 0.5
Proportion + 95% CI

#### Neurocognitive

##### Cognitive impairment

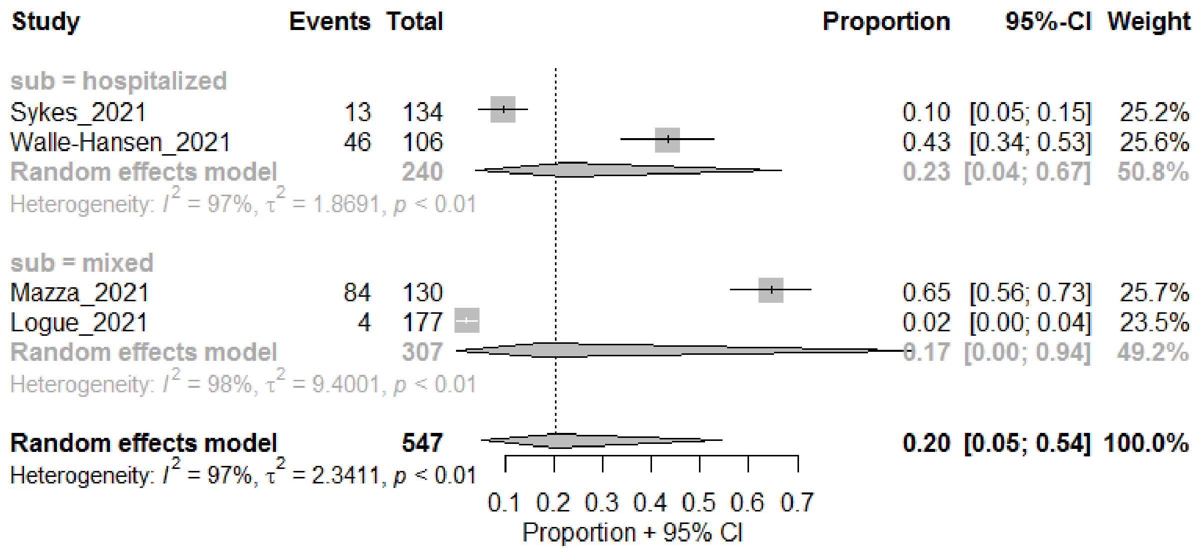

##### Concentration problems

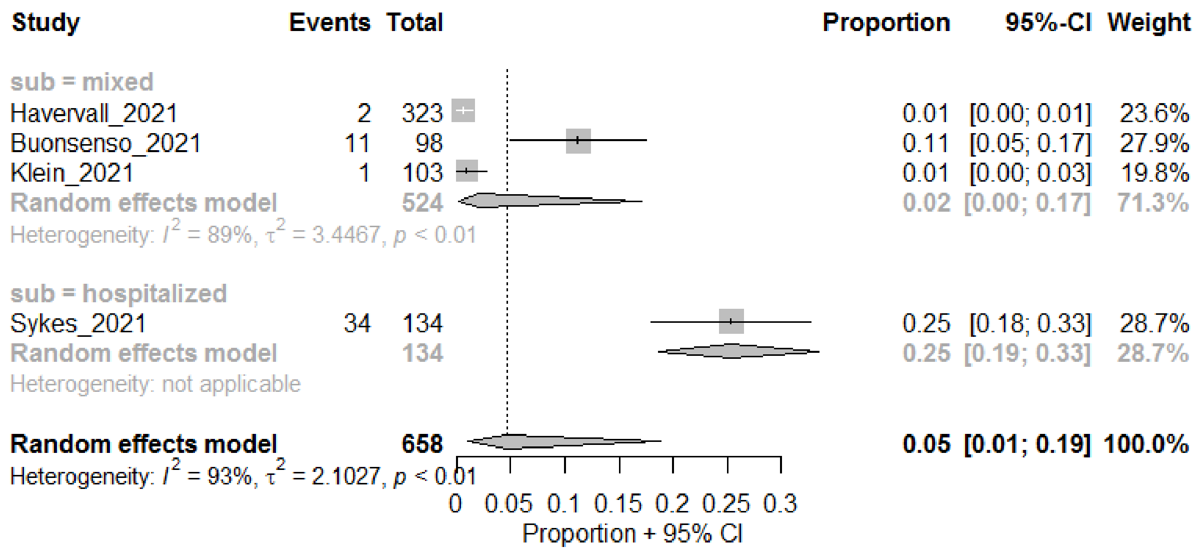

##### Memory problems

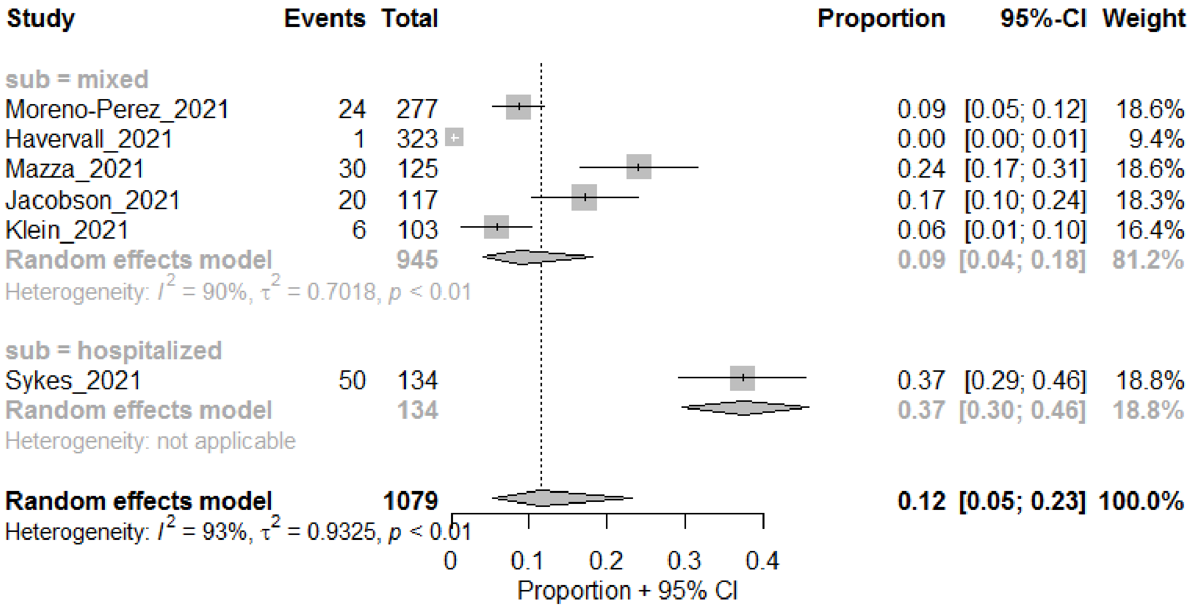

#### Dizziness

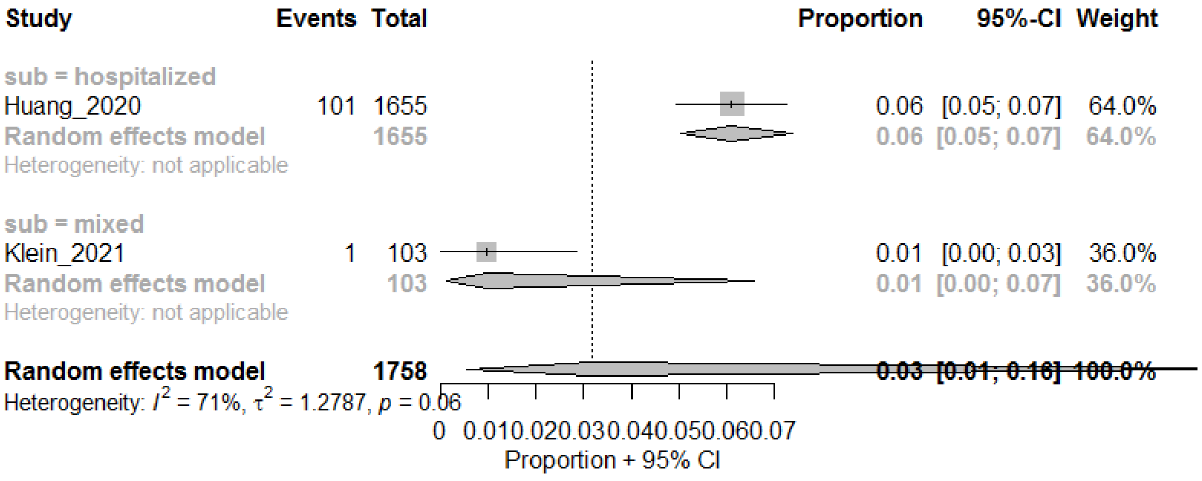

#### Cardiovascular

##### Chest pain

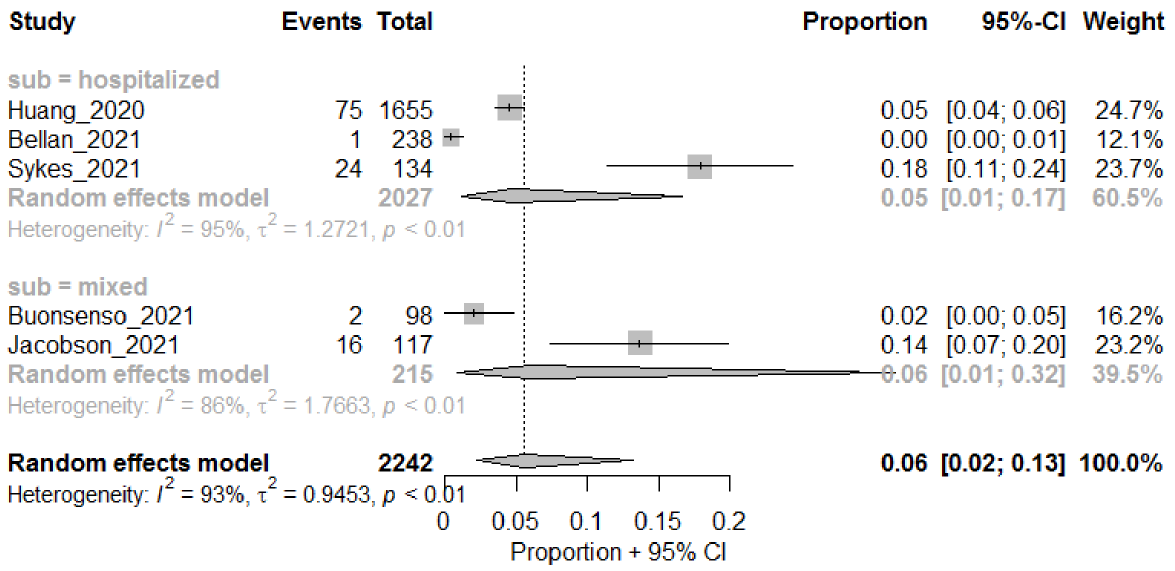

Palpitations

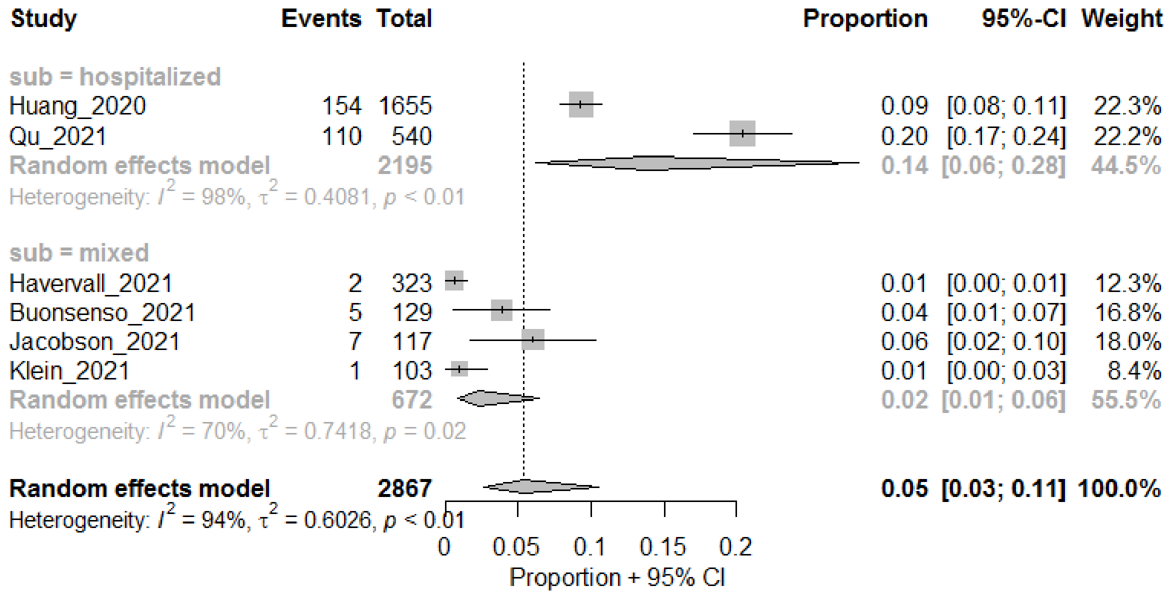

Chest tightness

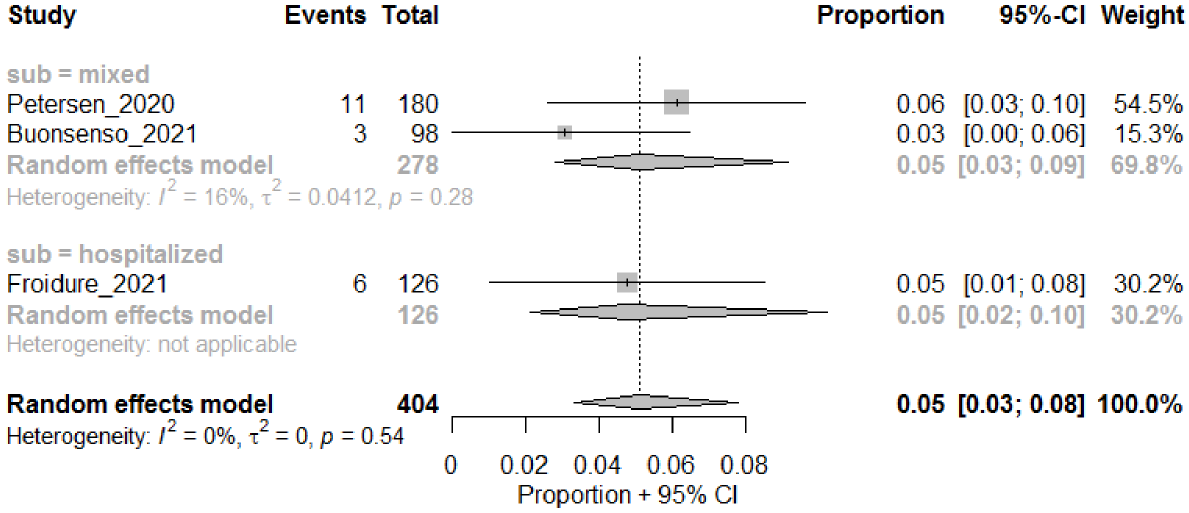

#### Digestive

##### Diarrhea

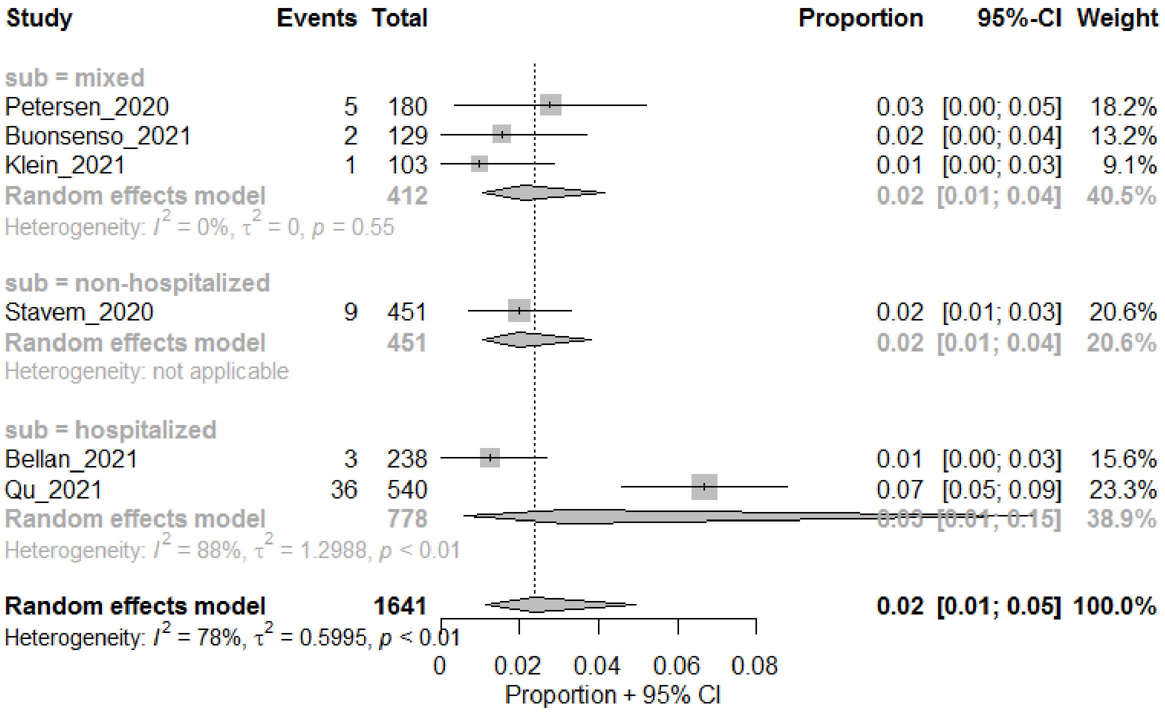

#### Abdominal pain

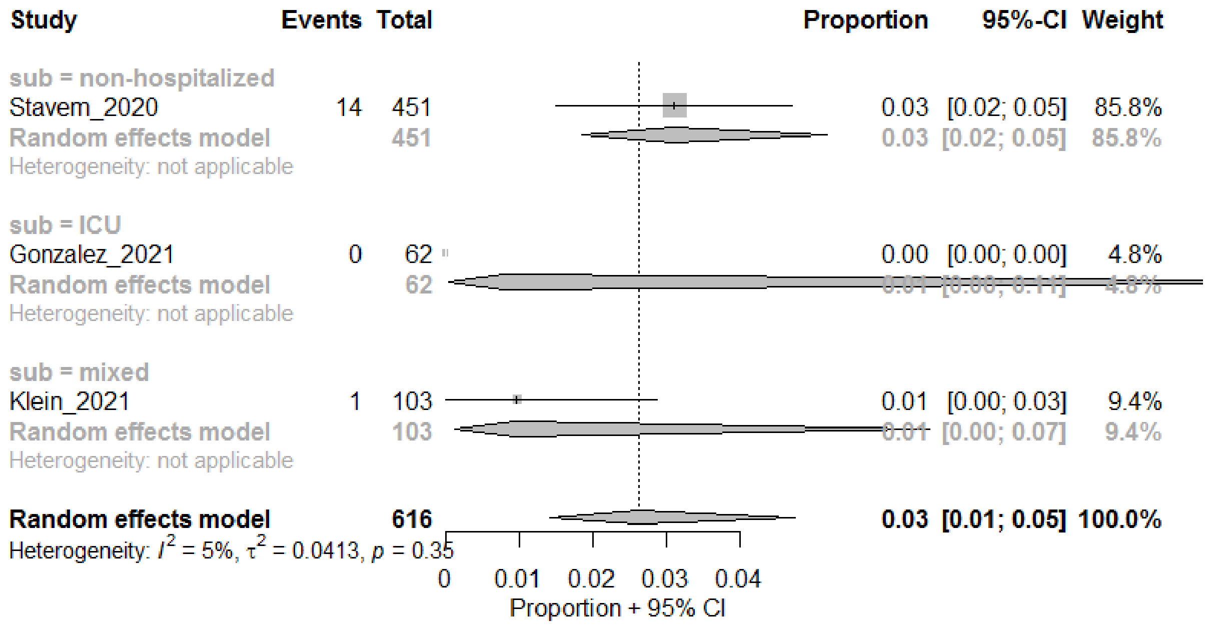

#### Mental

##### Anxiety

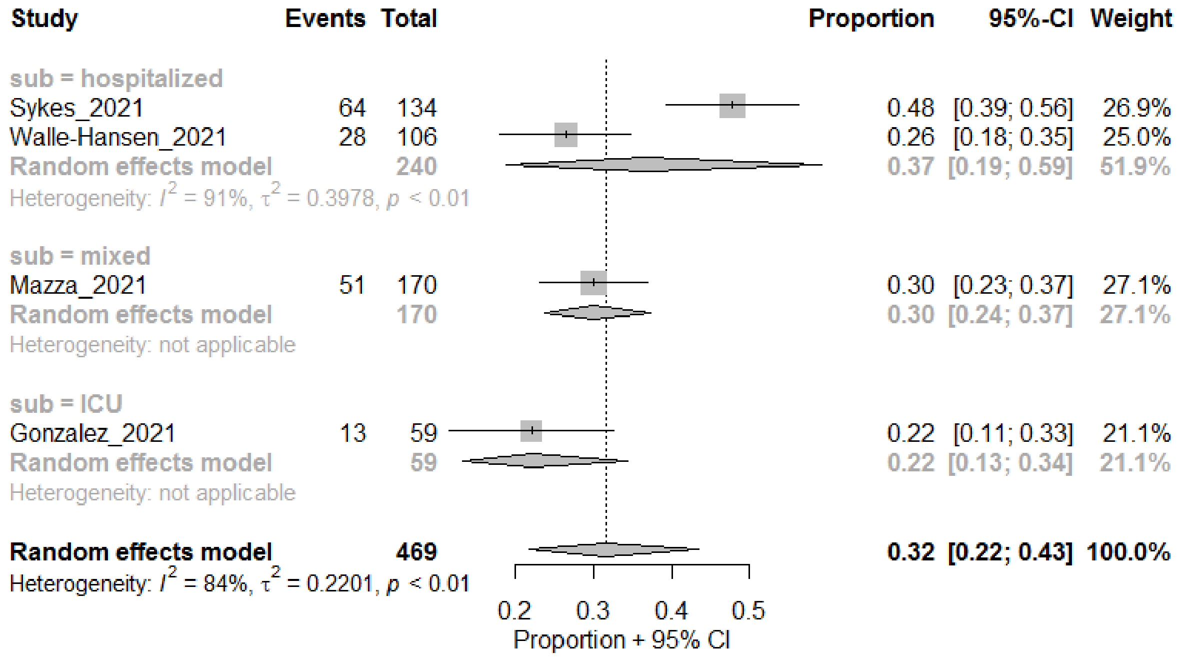

##### Depression

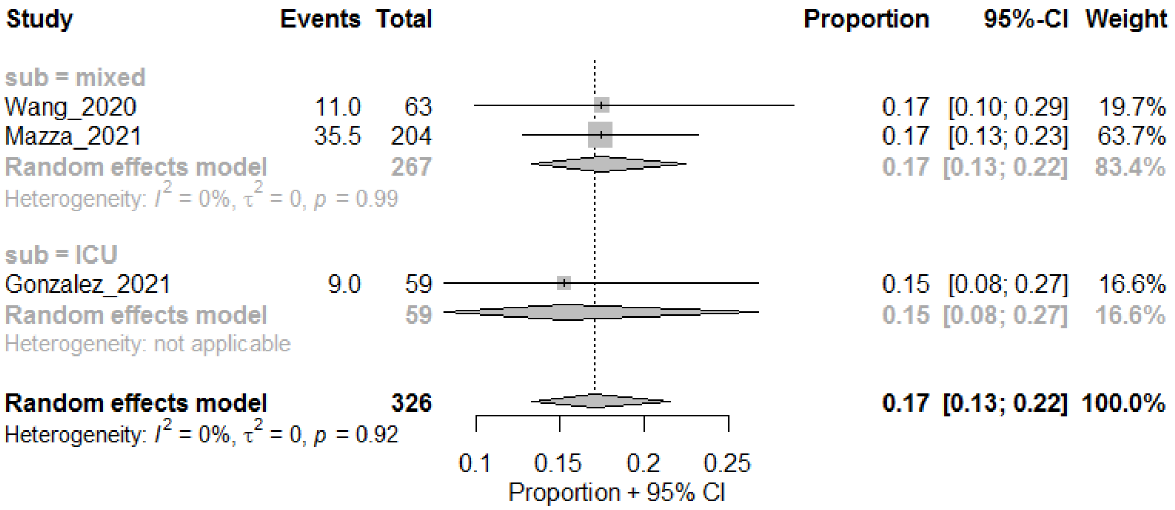

#### PTSD

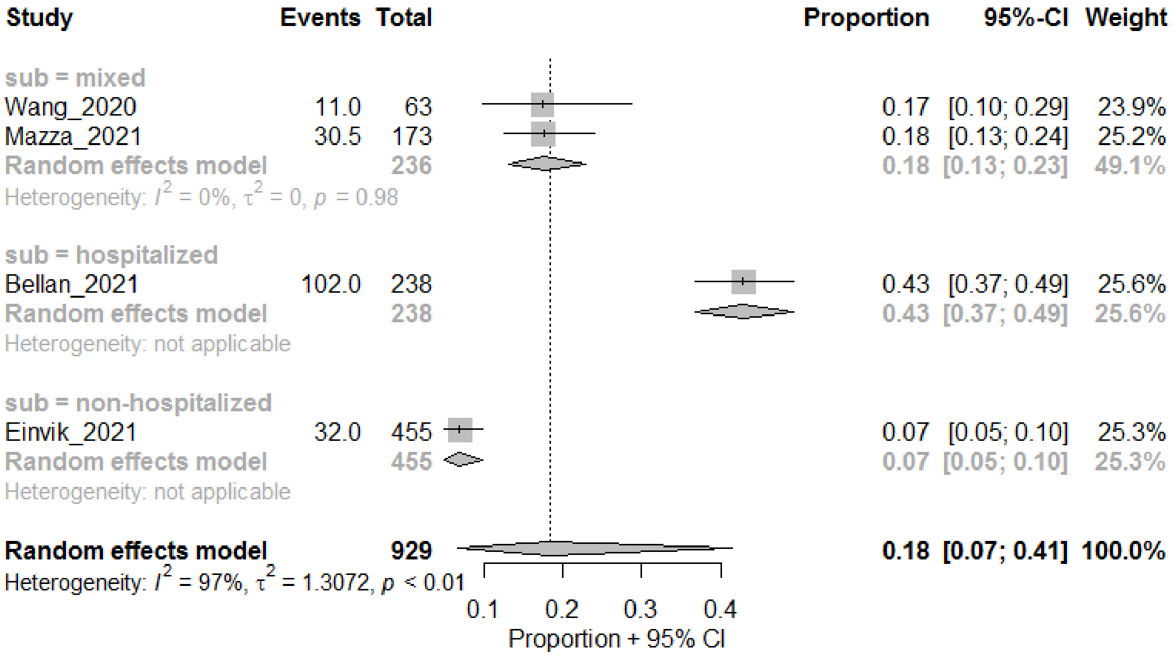

#### Sleep-related

##### Insomnia

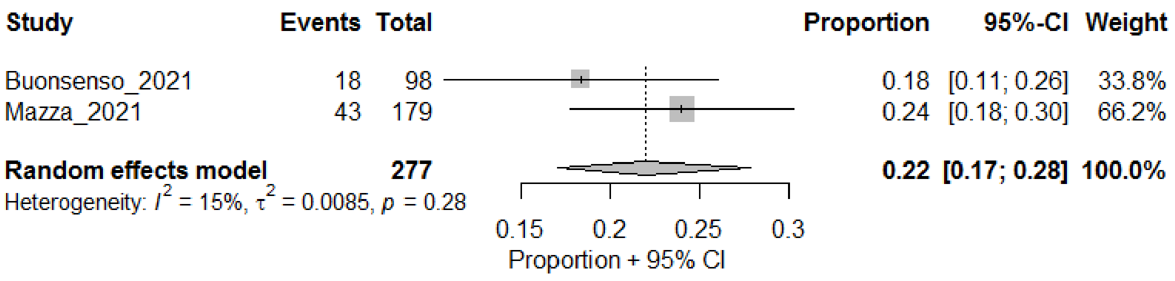

#### Sleep disturbances

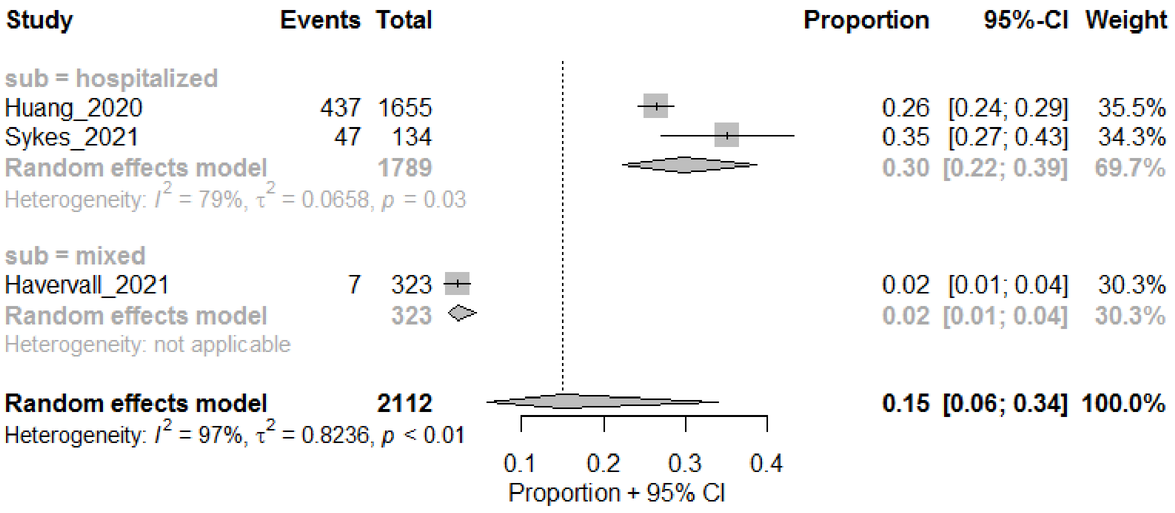

### Overall Functioning

#### General functional impairment

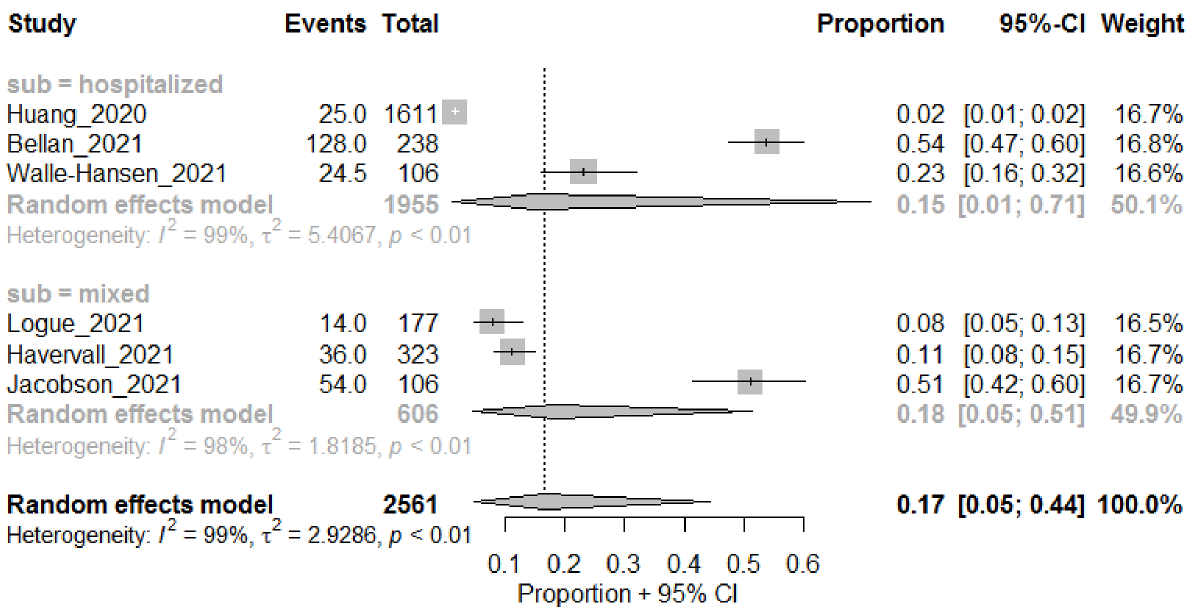

#### Desaturation or abnormal results at the end of the test

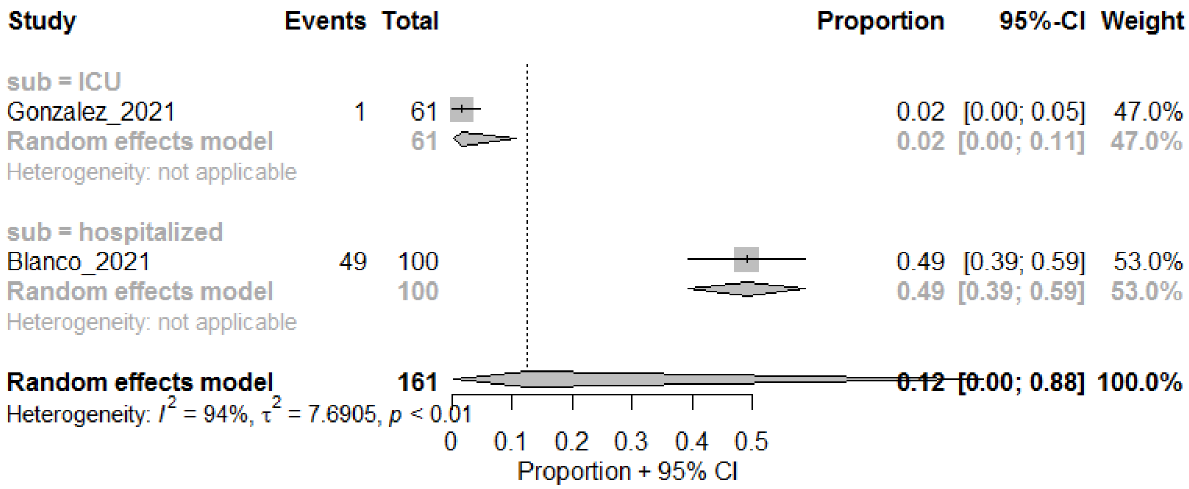

### Others

#### Rashes

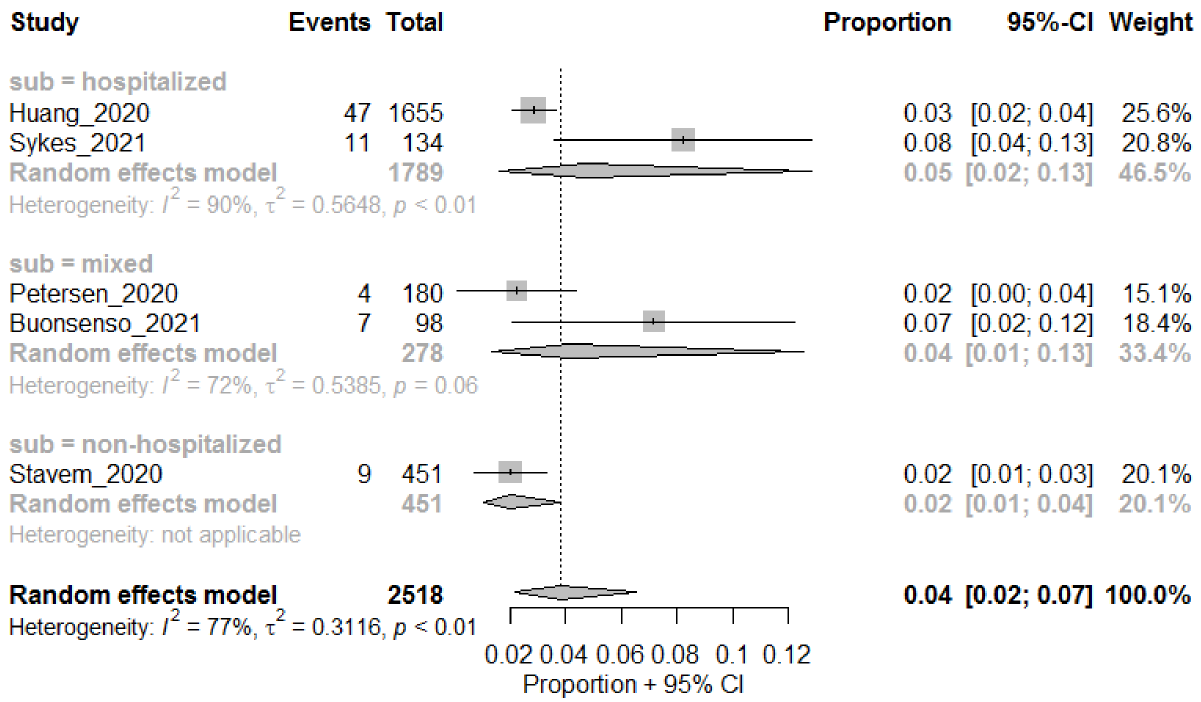

#### General pain

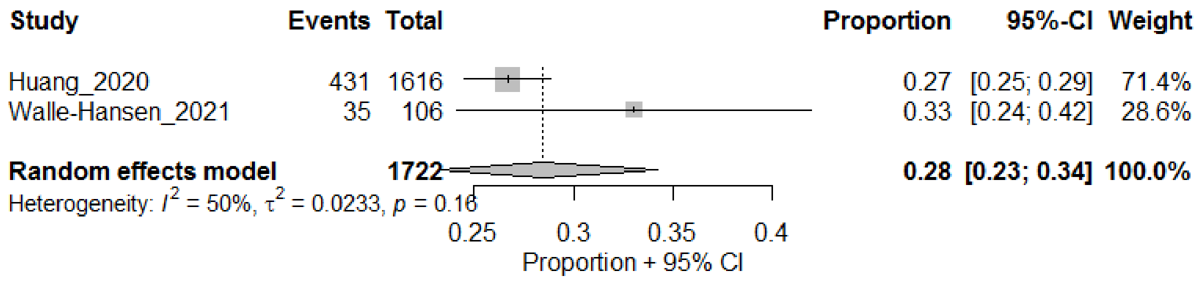

#### Hair loss

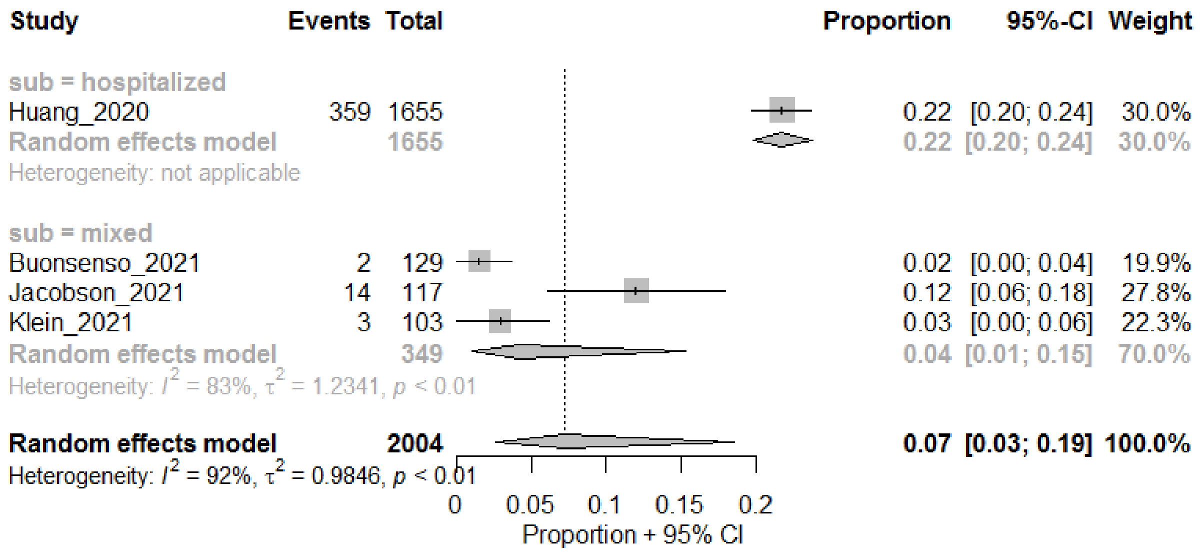

#### Ear pain

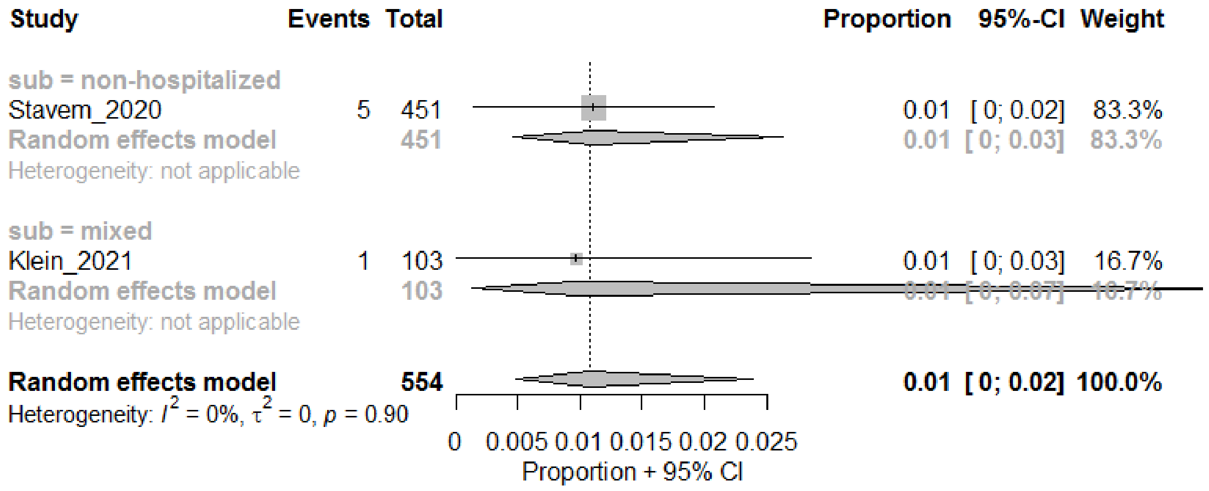

#### Complications

##### Pulmonary embolism

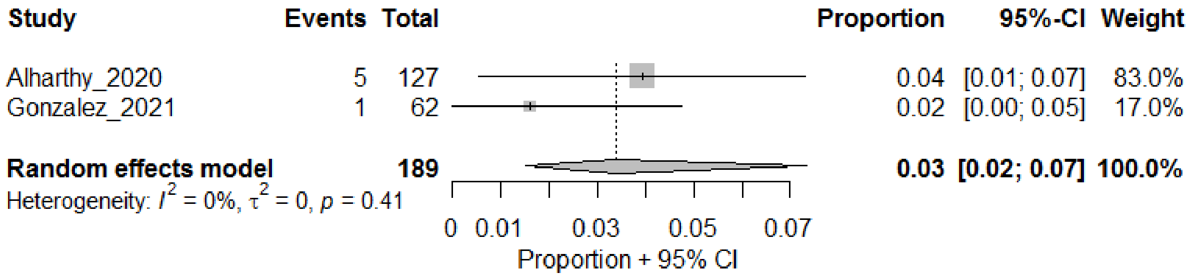

#### Abnormal CT findings

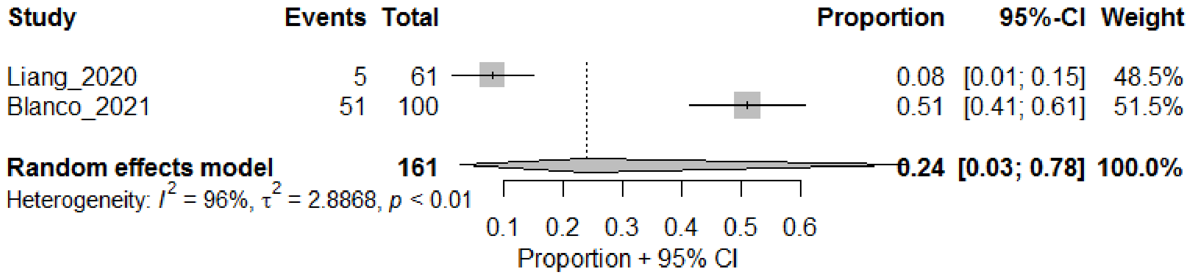

#### Ground glass opacities

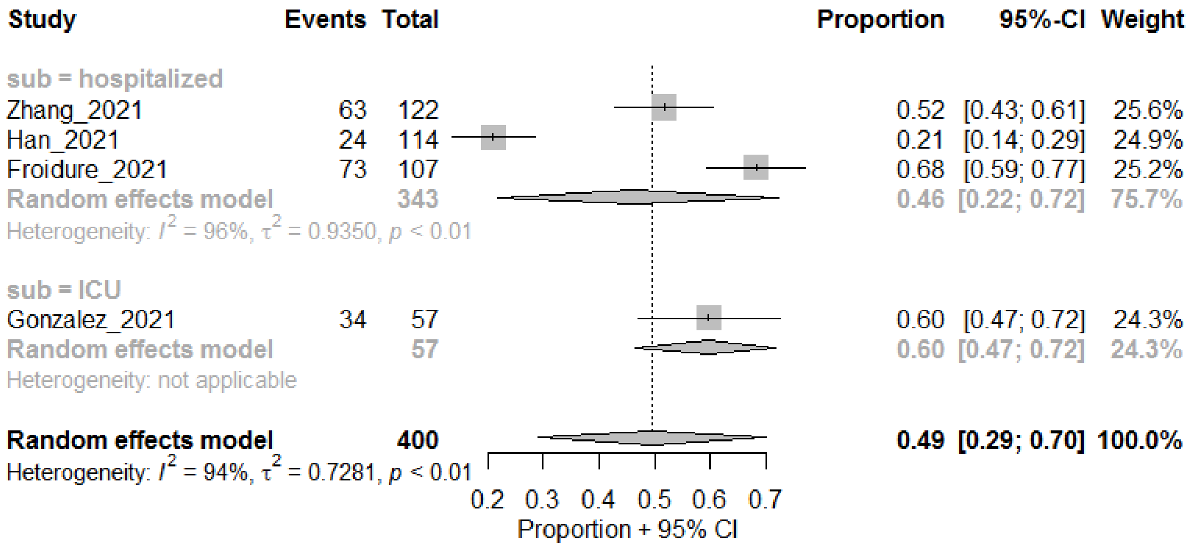

#### Fibrosis-like findings

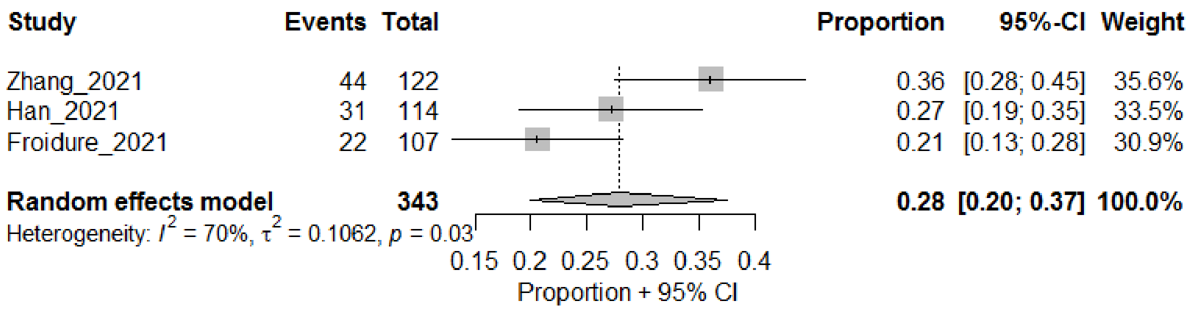

#### Bronchiectasis

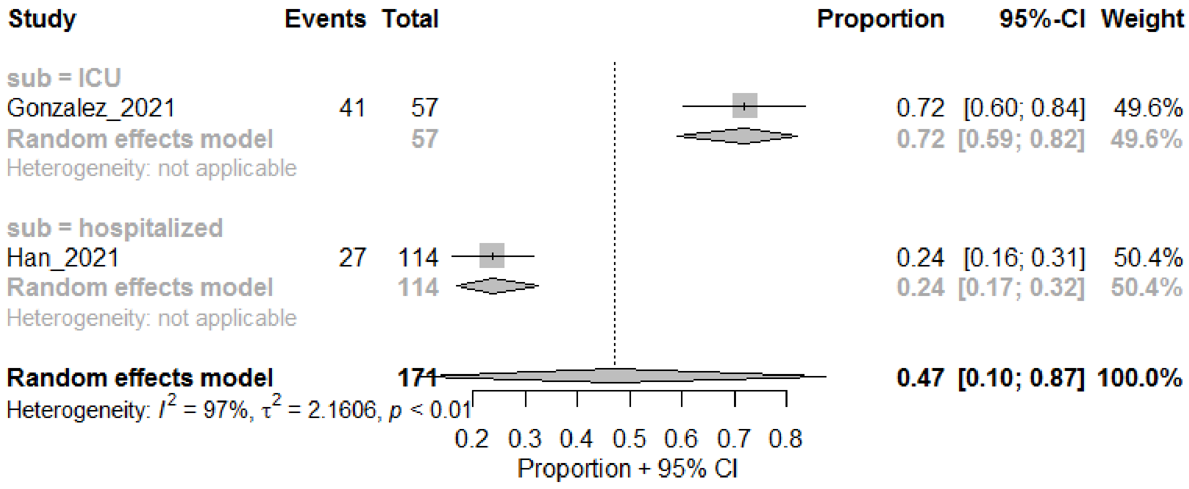

#### Atelectasis

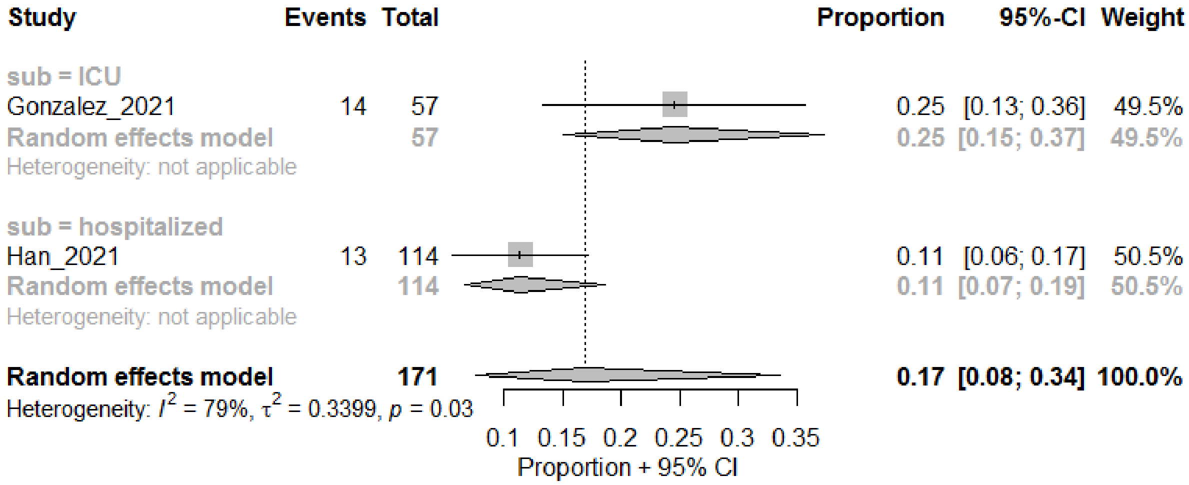

#### Nodules

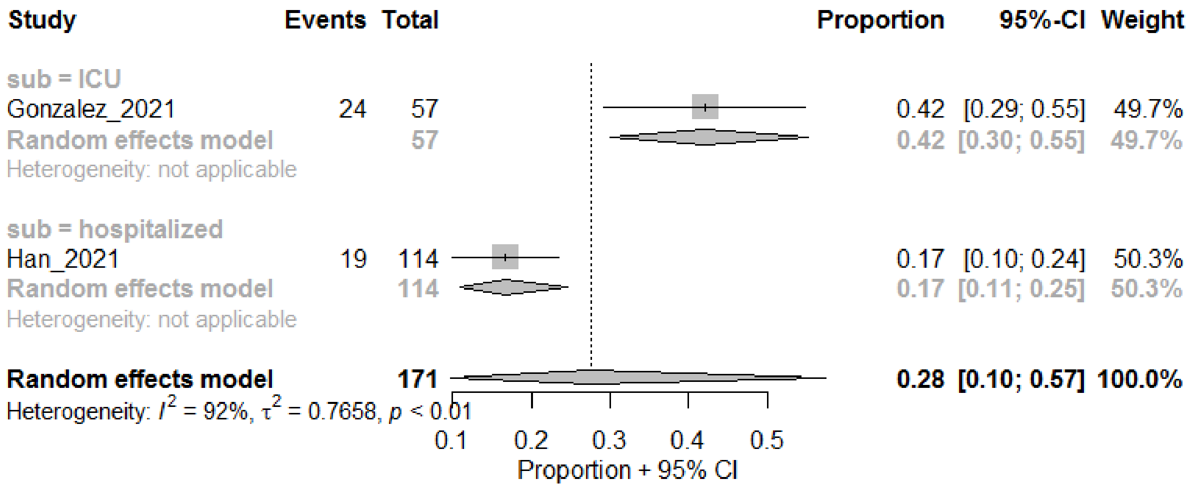

DLCO less than expected or abnormal

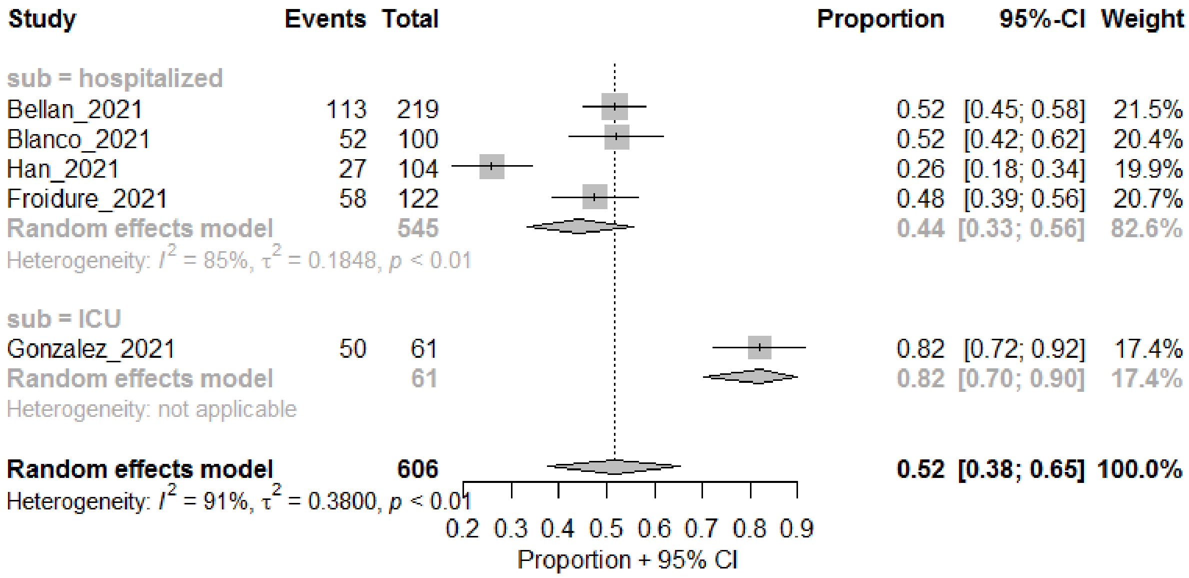

FVC less than expected or abnormal

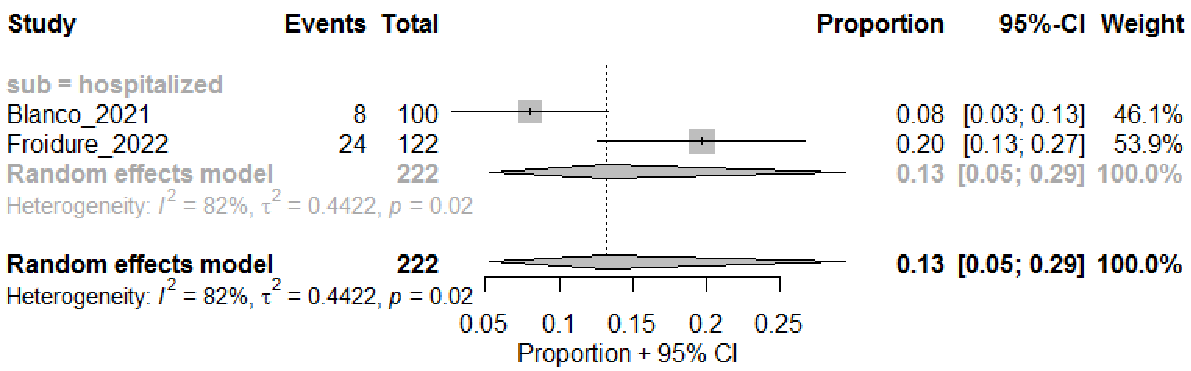

#### FEV1 less than expected or abnormal

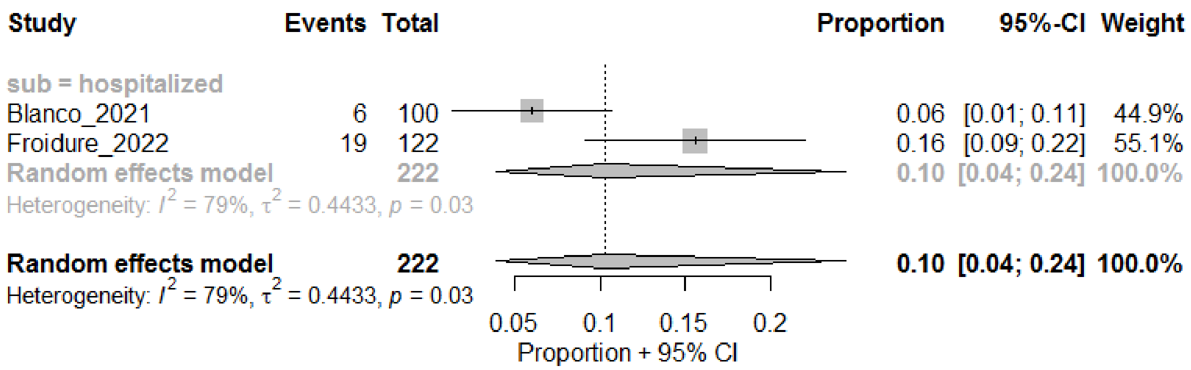

### Supplementary File 4 – PRISMA 2020 Checklist

| Section and Topic | Item # | Checklist item | Location where item is reported |
|---|---|---|---|
| **TITLE** | | | |
| Title | 1 | Identify the report as a systematic review. | Page 1 |
| **ABSTRACT** | | | |
| Abstract | 2 | See the PRISMA 2020 for Abstracts checklist. | Page 2 |
| **INTRODUCTION** | | | |
| Rationale | 3 | Describe the rationale for the review in the context of existing knowledge. | Page 3 |
| Objectives | 4 | Provide an explicit statement of the objective(s) or question(s) the review addresses. | Page 3 |
| **METHODS** | | | |
| Eligibility criteria | 5 | Specify the inclusion and exclusion criteria for the review and how studies were grouped for the syntheses. | Page 3-4, Supplementary File 1 |
| Information sources | 6 | Specify all databases, registers, websites, organisations, reference lists and other sources searched or consulted to identify studies. Specify the date when each source was last searched or consulted. | Page 3, Supplementary File 1 |
| Search strategy | 7 | Present the full search strategies for all databases, registers and websites, including any filters and limits used. | Supplementary File 1 |
| Selection process | 8 | Specify the methods used to decide whether a study met the inclusion criteria of the review, including how many reviewers screened each record and each report retrieved, whether they worked independently, and if applicable, details of automation tools used in the process. | Page 3-4, Supplementary File 1 |
| Data collection process | 9 | Specify the methods used to collect data from reports, including how many reviewers collected data from each report, whether they worked independently, any processes for obtaining or confirming data from study investigators, and if applicable, details of automation tools used in the process. | Page 4, Supplementary File 1 |
| Data items | 10a | List and define all outcomes for which data were sought. Specify whether all results that were compatible with each outcome domain in each study were sought (e.g. for all measures, time points, analyses), and if not, the methods used to decide which results to collect. | Page 4, Supplementary File 1 |
| | 10b | List and define all other variables for which data were sought (e.g. participant and intervention characteristics, funding sources). Describe any assumptions made about any missing or unclear information. | Supplementary File 1 |
| Study risk of bias assessment | 11 | Specify the methods used to assess risk of bias in the included studies, including details of the tool(s) used, how many reviewers assessed each study and whether they worked independently, and if applicable, details of automation tools used in the process. | Pages 5, Supplementary File 1 |
| Effect measures | 12 | Specify for each outcome the effect measure(s) (e.g. risk ratio, mean difference) used in the synthesis or presentation of results. | Page 4, Supplementary File 1 |
| Synthesis methods | 13a | Describe the processes used to decide which studies were eligible for each synthesis (e.g. tabulating the study intervention characteristics and comparing against the planned groups for each synthesis (item #5)). | Page 4, Supplementary |

| Section and Topic | Item # | Checklist item | Location where item is reported |
|---|---|---|---|
| | | | File 1 |
| | 13b | Describe any methods required to prepare the data for presentation or synthesis, such as handling of missing summary statistics, or data conversions. | Supplementary File 1 |
| | 13c | Describe any methods used to tabulate or visually display results of individual studies and syntheses. | Page 4, Supplementary File 1 |
| | 13d | Describe any methods used to synthesize results and provide a rationale for the choice(s). If meta-analysis was performed, describe the model(s), method(s) to identify the presence and extent of statistical heterogeneity, and software package(s) used. | Page 4, Supplementary File 1 |
| | 13e | Describe any methods used to explore possible causes of heterogeneity among study results (e.g. subgroup analysis, meta-regression). | Page 4-5, Supplementary File 1 |
| | 13f | Describe any sensitivity analyses conducted to assess robustness of the synthesized results. | Page 4, Supplementary File 1 |
| Reporting bias assessment | 14 | Describe any methods used to assess risk of bias due to missing results in a synthesis (arising from reporting biases). | Supplementary File 1 |
| Certainty assessment | 15 | Describe any methods used to assess certainty (or confidence) in the body of evidence for an outcome. | Pages 5, Supplementary File 1 |
| **RESULTS** | | | |
| Study selection | 16a | Describe the results of the search and selection process, from the number of records identified in the search to the number of studies included in the review, ideally using a flow diagram. | Page 6 |
| | 16b | Cite studies that might appear to meet the inclusion criteria, but which were excluded, and explain why they were excluded. | Figure 1, Supplementary File 2 |
| Study characteristics | 17 | Cite each included study and present its characteristics. | Table 1, Supplementary Table 1 |
| Risk of bias in studies | 18 | Present assessments of risk of bias for each included study. | Supplementary Table 2 |
| Results of individual studies | 19 | For all outcomes, present, for each study: (a) summary statistics for each group (where appropriate) and (b) an effect estimate and its precision (e.g. confidence/credible interval), ideally using structured tables or plots. | Table 2, Supplementary File 3 |
| Results of syntheses | 20a | For each synthesis, briefly summarise the characteristics and risk of bias among contributing studies. | Table 2, Supplementary Table 4 |
| | 20b | Present results of all statistical syntheses conducted. If meta-analysis was done, present for each the summary estimate and its precision (e.g. confidence/credible interval) and measures of statistical heterogeneity. If comparing groups, describe the direction of the effect. | Table 2, Supplementary |

| Section and Topic | Item # | Checklist item | Location where item is reported |
|---|---|---|---|
| | | | File 3 |
| | 20c | Present results of all investigations of possible causes of heterogeneity among study results. | Supplementary File 3 |
| | 20d | Present results of all sensitivity analyses conducted to assess the robustness of the synthesized results. | Supplementary File 3 |
| Reporting biases | 21 | Present assessments of risk of bias due to missing results (arising from reporting biases) for each synthesis assessed. | Supplementary Table 2 |
| Certainty of evidence | 22 | Present assessments of certainty (or confidence) in the body of evidence for each outcome assessed. | Table 2, Supplementary Table 3 |
| **DISCUSSION** | | | |
| Discussion | 23a | Provide a general interpretation of the results in the context of other evidence. | Page 8 |
| | 23b | Discuss any limitations of the evidence included in the review. | Page 8-9 |
| | 23c | Discuss any limitations of the review processes used. | Page 8-9 |
| | 23d | Discuss implications of the results for practice, policy, and future research. | Pages 9-10 |
| **OTHER INFORMATION** | | | |
| Registration and protocol | 24a | Provide registration information for the review, including register name and registration number, or state that the review was not registered. | Page 3 |
| | 24b | Indicate where the review protocol can be accessed, or state that a protocol was not prepared. | Page 3 |
| | 24c | Describe and explain any amendments to information provided at registration or in the protocol. | |
| Support | 25 | Describe sources of financial or non-financial support for the review, and the role of the funders or sponsors in the review. | Page 1, 9 |
| Competing interests | 26 | Declare any competing interests of review authors. | Page 1 |
| Availability of data, code and other materials | 27 | Report which of the following are publicly available and where they can be found: template data collection forms; data extracted from included studies; data used for all analyses; analytic code; any other materials used in the review. | Supplementary File 1 |

*From:* Page MJ, McKenzie JE, Bossuyt PM, Boutron I, Hoffmann TC, Mulrow CD, et al. The PRISMA 2020 statement: an updated guideline for reporting systematic reviews. BMJ 2021;372:n71. doi: 10.1136/bmj.n71

For more information, visit: http://www.prisma-statement.org/
