## Supplementary Tables for "Prevalence of long-term effects in individuals diagnosed with COVID-19: an updated living systematic review"

#### Supplementary Tables 1 to 4

**Authors:**

Francesca Reyes Domingo MHSc[1], Lisa A Waddell MSc PhD[2], Angela M. Cheung MD FRCPC[3], Curtis L. Cooper MD FRCPC[4ah], Veronica J. Belcourt MSc PhD[1], Alexandra M. E. Zuckermann PhD[1], Tricia Corrin MPH[2], Rukshanda Ahmad MBBS MHA[5], Laura Boland SLP-C PhD[1], Claudie Laprise MSc PhD[6,7], Leanne Idzerda MSc PhD[1], Anam Khan MPH[5], Kate Morissette MSc[1], Alejandra Jaramillo Garcia MSc[1]

**Affiliations:**

[1] Evidence Synthesis and Knowledge Translation Unit, Applied Research Division, Public Health Agency of Canada

[2] Public Health Risk Sciences Division, National Microbiology Laboratory, Public Health Agency of Canada

[3] Department of Medicine, University Health Network and University of Toronto, Canada

[4] Department of Medicine, University of Ottawa; Ottawa Hospital Research Institute

[5] Health Professionals Guidance Unit, Centre for Food-borne, Environmental and Zoonotic Infectious Diseases, Public Health Agency of Canada

[6] Public Health Capacity and Knowledge Management Unit, Quebec Regional Office, Public Health Agency of Canada

[7] Division of Oral Health and Society, Faculty of Dentistry, McGill University

#### Table S1: Characteristics of Included Studies with Clinically-Diagnosed Participants

| First author, publication year, study title & link | Country, study design, study recruitment dates | Study Population (# of participants; mean or median age or range; % female) | COVID-19 disease severity inclusion criteria | Time of outcome follow-up | Main symptoms/sequelae measured at follow-up |
|---|---|---|---|---|---|
| Arnold, 2020<br>Patient outcomes after hospitalisation with COVID-19 and implications for follow-up: results from a prospective UK cohort | UK,<br>Single-centre prospective study,<br>March - June 2020 | 163 adults (≥18 years) with a positive PCR result for SARS-CoV-2 or clinic-radiological diagnosis of COVID-19 disease<br><br>median age 60 years (IQR 46–73)<br><br>44% female | Hospitalized for COVID-19 | 90 days (IQR 80–97 days) after COVID-19 symptom onset. | At least one ongoing symptom<br>Fever<br>Cough<br>Breathlessness<br>Anosmia<br>Excessive fatigue<br>Myalgia<br>Headache<br>Chest pain<br>Arthralgia<br>Diarrhoea<br>Abdominal pain<br>Nausea<br>Insomnia<br>Significant desaturation on STS test |
| Janiri, 2021<br>Posttraumatic Stress Disorder in Patients After Severe COVID-19 Infection | Italy,<br>Cross-Sectional Study,<br>April 21 - October 15, 2020 | 381 patients<br><br>55.26 (14.86; 18-89)<br><br>43.6 % female | Severe COVID-19 | 30 to 120 days after recovery | PTSD<br>Depressive episode<br>Hypomanic episode<br>Generalized anxiety disorder<br>Psychotic disorders |
| Guler, 2021<br>Pulmonary function and radiological features four months after COVID-19: first results from the national prospective observational Swiss COVID-19 lung study | Switzerland,<br>Multicentre,<br>prospective observational cohort study,<br>May –September, 2020 | 113 adults who survived COVID-19<br><br>(mild/moderate 47)<br>52.9 (10.9) years<br>42.5% female<br><br>(severe/critical 66)<br>60.3 (12.0) years | n/a | follow up visit was 128 (108–144) days after symptom onset | Cough |

| First author, publication year, study title & link | Country, study design, study recruitment dates | Study Population (# of participants; mean or median age or range; % female) | COVID-19 disease severity inclusion criteria | Time of outcome follow-up | Main symptoms/sequelae measured at follow-up |
|---|---|---|---|---|---|
| | | 39.3% female | | | |
| Hittesdorf, 2020<br>Mortality and renal outcomes of patients with severe COVID-19 treated in a provisional intensive care unit | US,<br>Retrospective cohort study,<br>March - May 2020 | 116 patients who were treated for severe COVID-19<br><br>61.5 years +/− 12.1<br><br>35.3% female | Individuals admitted to a provisional ICU for severe COVID-19 | 30 to 90 days after admission | Renal replacement therapy - 30 days<br>Renal replacement therapy - at discharge<br>Renal replacement therapy - 90 days<br>Worse serum creatinine |
| Lerum, 2020<br>Dyspnoea, lung function and CT findings three months after hospital admission for COVID-19 | Norway,<br>Multicentre prospective cohort study,<br>March -June 2020 | 103 adults diagnosed with COVID-19<br><br>59 years (49-72)<br><br>48% female | Hospitalized for COVID-19 | median 83 (73-90) days (after admission) | Forced vital capacity < lower limits of normal (FVC < LLN)<br>Forced expiratory volume in one second < lower limits of normal (FEV1 < LLN)<br>Diffusion capacity of carbon monoxide < lower limits of normal (DLCO < LLN)<br>Dyspnoea > 0 in the modified Medical Research Council dyspnea scale (mMRC)<br>Ground-glass chest opacities<br>Parenchymal bands |
| Lucidi, 2021<br>Patient-reported olfactory recovery after SARS-CoV-2 infection: A 6-month follow-up study | Italy,<br>Retrospective Cohort Study,<br>September 2020 | 110 patients<br><br>41.4 ± 12.3<br><br>63.6 % female | Any | 6.1 ± 1.1 months | anosmia/ hyposmia<br>persistent olfactory dysfunction |
| Mallia, 2021 | United Kingdom, | 401 patients | Discharged patients with COVID-19 | Median of 53 days post | Any symptom(s)<br>Breathlessness<br>Fatigue |

| First author, publication year, study title & link | Country, study design, study recruitment dates | Study Population (# of participants; mean or median age or range; % female) | COVID-19 disease severity inclusion criteria | Time of outcome follow-up | Main symptoms/sequelae measured at follow-up |
|---|---|---|---|---|---|
| Symptomatic, biochemical and radiographic recovery in patients with COVID-19 | Retrospective Cohort Study, May 1 and July 21 2020 | 59.0 (21–95)<br><br>40.4 % female | | discharge and 72 days after symptom onset | Cough<br>Chest Pain<br>At least two symptoms among symptomatic patients<br>No return to pre-illness exercise tolerance level<br>Desaturation on sit-to-stand test<br>Psychological distress related to hospital admission (PTSD) |
| Miyazato, 2020<br>Prolonged and Late-Onset Symptoms of Coronavirus Disease 2019 | Japan,<br>Cross-sectional,<br>July –August 2020 | 78 patients who were diagnosed with COVID-19<br><br>48.1years (18.5)<br><br>33.3% female | Hospitalized for COVID-19 | Up to 120 days after symptom onset | Persistent Cough - 60 & 120 days<br>Persistent Fatigue - 60 & 120 days<br>Persistent Dyspnea - 60 & 120 days<br>Persistent dysgeusia - 60 & 120 days<br>Persistent Dysosmia - 60 & 120 days<br>Dysosmia<br>Alopecia |
| Naidu, 2021<br>The high mental health burden of "Long COVID" and its association with on-going physical and respiratory symptoms in all adults discharged from hospital; 8015645 CSR - Reports results | United Kingdom Prospective Cohort Study, May 22, 2020 | 760 patients<br><br>Mean age 60.7 (standard deviation [sd]±16.3 year<br><br>30.8 % female | Any | Median of 9 weeks post discharge | Persisting physical and psychiatric symptoms<br>Depression<br>PTSD<br>any persistent symptoms<br>anorexia<br>Not back to work<br>Confusion or “fuzzy head”<br>myalgia<br>Non-improved breathlessness<br>Non-improved cough<br>Non-improved fatigue<br>Non-improved sleep quality |

| First author, publication year, study title & link | Country, study design, study recruitment dates | Study Population (# of participants; mean or median age or range; % female) | COVID-19 disease severity inclusion criteria | Time of outcome follow-up | Main symptoms/sequelae measured at follow-up |
|---|---|---|---|---|---|
| Nunez-Cortes, 2021<br>Use of sit-to-stand test to assess the physical capacity and exertional desaturation in patients post COVID-19 | Chile, Cross-Sectional Study, August 4 and September 11, 2020 (6 weeks) | 50 patients<br><br>62.7 ± 12.5<br><br>46.0 % female | Hospitalized due to COVID-19 and discharged | 4 weeks since discharged from hospital | Repetitions <2.5th percentile of 1 sit-to-stand-test<br>Repetitions <25th percentile of 1 sit-to-stand-test<br>Change in SpO2≥4<br>SPO2 <90% post test |
| Simani, 2021,<br>Prevalence and correlates of chronic fatigue syndrome and post-traumatic stress disorder after the outbreak of the COVID-19; 7852482 CSR - Reports results | Iran, Cross-Sectional Study, February- April 2020 | 120 patients<br><br>Mean age 54.62 ± 16.94<br><br>33.3 % female | Any hospitalized | 6 months after discharge | Chronic fatigue syndrome/myalgic encephalomyelitis (CFS/ME) *with and without PTSD<br>PTSD<br>CFSLWIFS (CFS-like with insufficient fatigue syndrome)<br>CIF (chronic idiopathic fatigue)<br>CFS (chronic fatigue syndrome) |
| Samimi Ardestani, 2021<br>The coronavirus disease 2019: the prevalence, prognosis, and recovery from olfactory dysfunction (OD) | Iran, Cross-Sectional Study, March - May 2020 | 311 patients<br><br>47.00 ± 12.42<br><br>28.3 % female | Referred to the department of respiratory emergencies | OD patients were followed up later a month after the initial diagnosis | Olfactory dysfunction |
| Writing Committee for the COMEBAC Study Group, 2021<br>Four-Month Clinical Status of a Cohort of Patients After Hospitalization for COVID-19 | France, Prospective Cohort Study, between March 1 and May 29, 2020 | 478 patients<br><br>60.9/16.1<br><br>42.1 % female | Hospitalized due to COVID-19 | 4 months after discharge | at least one symptom<br>at least one cognitive symptom<br>dyspnea (new onset)<br>Anosmia<br>headaches<br>paresthesia<br>anorexia<br>fatigue<br>weight loss >5% baseline weight<br>chest discomfort/ pain |

| First author, publication year, study title & link | Country, study design, study recruitment dates | Study Population (# of participants; mean or median age or range; % female) | COVID-19 disease severity inclusion criteria | Time of outcome follow-up | Main symptoms/sequelae measured at follow-up |
|---|---|---|---|---|---|
| | | | | | cough<br>limb palsy<br>memory difficulties<br>mental slowness<br>concentration problems<br>persistent cough<br>abnormal lung CT scan<br>persistent ground glass opacities<br>lung fibrotic lesions<br>DLCO <70%<br>RV dilation on ECG<br>LVEF 40-50% on ECG<br>Cognitive complaint<br>cognitive impairment<br>anxiety<br>Depression<br>insomnia<br>PTSD<br>Dysfunctional breathing<br>Positive hyperventilation provocation test |
| Shendy, 2021<br>Prevalence of fatigue in patients post Covid-19 | Egypt, Retrospective Cohort Study, September - December 2020 | 81 patients<br><br>Mean age 34.03 ± SD 4.9<br><br>68 % female | Mild or moderate cases | 3-5 months after recovery | Fatigue |
| Sudre, 2021<br>Attributes and predictors of long COVID | Multiple, Prospective Cohort Study, March - September 2020 | 4,182 patients<br><br>Median 42 (IQR 32–53) | Any | 12 weeks after symptom onset | fatigue<br>headache<br>shortness of breath<br>loss of smell<br>persistent cough |

| First author, publication year, study title & link | Country, study design, study recruitment dates | Study Population (# of participants; mean or median age or range; % female) | COVID-19 disease severity inclusion criteria | Time of outcome follow-up | Main symptoms/sequelae measured at follow-up |
|---|---|---|---|---|---|
| | | 71.5 % female | | | sore throat<br>fever<br>unusual muscle pains<br>skipped meals<br>chest pain<br>diarrhoea<br>hoarse voice<br>abdominal pain<br>delirium<br>any symptoms |
| Tomasoni, 2020<br>Anxiety and depression symptoms after virological clearance of COVID-19: A cross-sectional study in Milan, Italy | Italy,<br>Cross-sectional study,<br>April to June 2020 | 105 patients who were diagnosed with COVID-19<br><br>55 (43-65) years<br><br>27% female | Hospitalized for COVID-19 | 1-3 months after clinical & virological recovery from symptomatic COVID-19 disease | Anxiety<br>Depression<br>Persistence of at least one physical symptom<br>Cognitive deficits (memory disorder) - ongoing<br>Anosmia - ongoing<br>Dysgeusia - ongoing<br>Gastro-intenstinal symptoms - ongoing<br>Fever - ongoing<br>Burning pain - ongoing<br>Dyspnea - ongoing<br>Asthenia - ongoing<br>Other symptoms - ongoing |
| van den Borst, 2020<br>Comprehensive health assessment three months after recovery from acute COVID-19 | The Netherlands,<br>Prospective observational study,<br><br>April-July 2020 | 124 patients who were treated for COVID-19<br><br>age 59±14 years,<br><br>40% female | Hospitalized for COVID-19 | ~3 months after onset of COVID-19 | Maximal vital capacity < lower limits of normal (Vcmax<LLN)<br>Forced expiratory volume in one second < lower limits of normal (FEV1 < LLN)<br>Forced expiratory volume over maximal vital capacity < lower limits of normal (FEV1/Vcmax < LLN) |

| First author, publication year, study title & link | Country, study design, study recruitment dates | Study Population (# of participants; mean or median age or range; % female) | COVID-19 disease severity inclusion criteria | Time of outcome follow-up | Main symptoms/sequelae measured at follow-up |
|---|---|---|---|---|---|
| | | | | | diffusion capacity of carbon monoxide < lower limits of normal (DLCO < LLN)<br>total lung capacity < lower limits of normal (TLC < LLN)<br>residual volume < lower limits of normal (RV < LLN)<br>Residual CT abnormality - ground glass opacity<br>Residual CT abnormality - Bronchi(ol)ectasis<br>Residual CT abnormality - lines and bands<br>Residual CT abnormality - fibrosis<br>Number of Residual CT abnormalities – 1 - 4<br>Frail<br>6-minute walking distance (6MWD)<80% predicted<br>Desaturation ≥ 4% upon 6-minute walk test (6MWT)<br>anxiety<br>depression<br>Telephone Interview of Cognitive Status (TICS)<34<br>Cognitive Failure Questionnaire (CFQ)>43<br>Post Traumatic Stress Syndrome (PTSS) Checklist DSM-5 (PCL-5)>33<br>Impact of Event Scale-Revised (IES-R)>33<br>Abnormal scored on mental or cognitive status questionnaires |

| First author, publication year, study title & link | Country, study design, study recruitment dates | Study Population (# of participants; mean or median age or range; % female) | COVID-19 disease severity inclusion criteria | Time of outcome follow-up | Main symptoms/sequelae measured at follow-up |
|---|---|---|---|---|---|
| Venturelli, 2021<br>Surviving COVID-19 in Bergamo Province: A post-Acute outpatient re-evaluation | Italy,<br>Cohort<br>2 May through 31 July | 767 patients<br><br>63 (s.d. 13.6, range 20-92)<br><br>32.9 % female | COVID-19 patients discharged from ED | 81 days | Confusion<br>Asthenia<br>Dyspnoea<br>Fever<br>Myalgia<br>Cough<br>Headache<br>Chest pain<br>Palpitations<br>Syncope<br>Anosmia/Dysgeusia<br>Upper gastro-intestinal symptoms<br>Lower gastro-intestinal symptoms<br>Others<br>⩾3 persistent symptoms<br>% Oxygen saturation on room air<br>% Karnofsky performance status scale<br>Anxiety<br>Depression<br>PTSD<br>Resiliance<br>Cognitive impairment<br>Any symptom(s)<br>Not fully recovered<br>No longer fully independent (after COVID-19; Barthel Index)<br>Newly moderately-severely dependent<br>New onset fatigue<br>Pulmonary function - obstruction<br>Pulmonary function - restrictive pattern |

| First author, publication year, study title & link | Country, study design, study recruitment dates | Study Population (# of participants; mean or median age or range; % female) | COVID-19 disease severity inclusion criteria | Time of outcome follow-up | Main symptoms/sequelae measured at follow-up |
|---|---|---|---|---|---|
| | | | | | Pulmonary function - mixed pattern<br>DLCO - reduced |
| Xiong, 2021<br>Clinical sequelae of COVID-19 survivors in Wuhan, China: a single-centre longitudinal study | China, Longitudinal study based on telephone follow-up survey, March 2020 | 538 adults diagnosed with COVID-19<br><br>52.0 (41.0–62.0) years<br><br>54.5% female | Hospitalized for COVID-19 | 3 months after discharge from hospital | General symptoms<br>Physical decline/fatigue<br>Sweating<br>Myalgia<br>Arthalgia<br>Chills<br>Limb oedema<br>Dizziness<br>Respiratory symptoms<br>Postactivity polypnoea<br>Nonmotor polypnoea<br>Chest distress<br>Chest pain<br>Cough<br>Sputum<br>Throat pain<br>Cardiovascular-related symptoms<br>Resting heart rate increase<br>Discontinuous flushing (palpitations)<br>Newly diagnosed hypertension<br>Psychosocial symptoms<br>Somnipathy<br>Depression<br>Anxiety<br>Dysphoria<br>Feelings of inferiority<br>Alopecia |
| Xu, 2021<br>Plasma metabolomic profiling of patients | China | 130 patients | mild/moderate/severe & critical | 3 months | FEV1 <80% predicted<br>FEV1/FVC <70%<br>TLC < 80% predicted |

| First author, publication year, study title & link | Country, study design, study recruitment dates | Study Population (# of participants; mean or median age or range; % female) | COVID-19 disease severity inclusion criteria | Time of outcome follow-up | Main symptoms/sequelae measured at follow-up |
|---|---|---|---|---|---|
| recovered from COVID-19 with pulmonary sequelae 3 months after discharge | Prospective Cohort<br>March 1 and March 30, 2020 | 56.00 (44.75–63.25)<br>61.00 (55.00–68.00)<br><br>53.6 % female | | | RV (L) <65% predicted<br>DLCO <80% predicted<br>DLCO 60-80% predicted<br>DLCO 40-60% predicted<br>DLCO/VA <80% predicted<br>Abnormal CT |

### Table S2. Summary of Risk of Bias (RoB) assessments (a) by study and (b) by outcome (using a modified Joanna-Briggs Institute's critical appraisal checklist)

#### By Study

| | Participants | | | Participant criterion assessment | Outcome Measures | | Outcome measures criterion assessment | Statistics | Statistics assessment | Risk of Bias for study overall |
|---|---|---|---|---|---|---|---|---|---|---|
| | Was the sample frame appropriate to address the target population? | Were study participants sampled in an appropriate way? | Was the response rate adequate, and if not, was the low response rate managed appropriately? | | Were valid methods used for the identification of the outcome? | Was the outcome measured in a standard, reliable way for all participants? | | Was the data for the outcome reported clearly and completely? | | |
| Laboratory-Confirmed | | | | | | | | | | |
| Akter 2020 | No | No | No | Not Met | No | Yes | Partially Met | Yes | Met | Moderate |
| Alemanno 2021 | Yes | No | No | Not Met | Yes | Yes | Met | Yes | Met | Moderate |
| Alharthy 2020 | No | No | No | Not Met | No | No | Not Met | Yes | Met | High |
| AlShakhs 2021 | No | No | No | Not Met | No | Yes | Partially Met | No | Not Met | High |
| Anastasio 2021 | No | No | No | Not Met | No | No | Not Met | Yes | Met | High |
| Bellan 2021 | No | Yes | No | Not Met | No | No | Not Met | Yes | Met | High |
| Blair 2021 | No | No | No | Not Met | No | Yes | Partially Met | No | Not Met | High |
| Blanco 2021 | No | Yes | No | Not Met | Yes | Yes | Met | Yes | Met | Moderate |
| Boscolo-Rizzo 2020 | Yes | No | Yes | Not Met | No | Yes | Partially Met | Yes | Met | Moderate |
| Brandao 2021 | Yes | No | No | Not Met | No | Yes | Partially Met | No | Not Met | High |
| Bulgurcu 2020 | No | No | No | Not Met | No | No | Not Met | Yes | Met | High |
| Buonsenso 2021 | Yes | No | Yes | Not Met | No | Yes | Partially Met | Yes | Met | Moderate |
| Carvalho-Schneider 2020 | No | No | No | Not Met | No | Yes | Partially Met | Yes | Met | Moderate |
| Chiesa-Estomba 2020 | No | No | No | Not Met | No | Yes | Partially Met | Yes | Met | Moderate |
| Cortes-Telles 2021 | No | No | No | Not Met | No | Yes | Partially Met | Yes | Met | Moderate |
| D'Cruz 2020 | Yes | No | Yes | Not Met | No | Yes | Partially Met | Yes | Met | Moderate |
| deGraaf 2021 | Yes | No | No | Not Met | No | Yes | Partially Met | Yes | Met | Moderate |
| Einvik 2021 | Yes | No | No | Not Met | No | Yes | Partially Met | Yes | Met | Moderate |
| Froidure 2021 | No | No | Yes | Not Met | Yes | Yes | Met | Yes | Met | Moderate |
| Gambini 2020 | No | No | No | Not Met | No | No | Not Met | Yes | Met | High |

| | | | | | | | | | |
|---|---|---|---|---|---|---|---|---|---|
| Gonzalez 2021 | Yes | No | Yes | Not Met | Yes | Yes | Met | Yes | Met | Moderate |
| Halpin 2021 | No | No | Yes | Not Met | No | Yes | Partially Met | Yes | Met | Moderate |
| Han 2021 | No | No | No | Not Met | Yes | Yes | Met | Yes | Met | Moderate |
| Havervall 2021 | Yes | No | Yes | Not Met | No | Yes | Partially Met | Yes | Met | Moderate |
| Horn 2020 | No | No | No | Not Met | No | Yes | Partially Met | Yes | Met | Moderate |
| Huang 2020 | No | No | No | Not Met | No | Yes | Partially Met | Yes | Met | Moderate |
| Jacobs 2020 | No | No | No | Not Met | No | No | Not Met | Yes | Met | High |
| Jacobson 2021 | No | No | No | Not Met | Yes | Yes | Met | Yes | Met | Moderate |
| Khasawneh 2020 | No | No | No | Not Met | No | Yes | Partially Met | Yes | Met | Moderate |
| Klein 2021 | Yes | No | Yes | Not Met | No | Yes | Partially Met | Yes | Met | Moderate |
| Konstantinidis 2020 | Yes | Yes | No | Partially Met | No | No | Not Met | Yes | Met | Moderate |
| Landi 2021 | Yes | No | Yes | Not Met | No | No | Not Met | Yes | Met | High |
| Lascarrou 2021 | No | No | No | Not Met | No | No | Not Met | Yes | Met | High |
| Liang 2020 | Yes | No | No | Not Met | Yes | Yes | Met | Yes | Met | Moderate |
| Logue 2021 | No | No | No | Not Met | No | Yes | Partially Met | Yes | Met | Moderate |
| Mafort 2021 | Yes | No | Yes | Not Met | No | No | Not Met | Yes | Met | High |
| Mandal 2020 | No | No | Yes | Not Met | No | Yes | Partially Met | Yes | Met | Moderate |
| Mazza 2021 | No | No | Yes | Not Met | No | Yes | Partially Met | Yes | Met | Moderate |
| Moradian 2020 | Yes | No | No | Not Met | No | Yes | Partially Met | Yes | Met | Moderate |
| Moreno-Perez 2021 | No | No | No | Not Met | No | No | Not Met | Yes | Met | High |
| Niklassen 2021 | Yes | No | No | Not Met | Yes | No | Partially Met | Yes | Met | Moderate |
| Otte 2020 | No | No | No | Not Met | Yes | Yes | Met | Yes | Met | Moderate |
| Petersen 2020 | No | No | Yes | Not Met | No | No | Not Met | No | Not Met | High |
| Printza 2021 | No | Yes | Yes | Not Met | No | Yes | Partially Met | Yes | Met | Moderate |
| Qu 2021 | No | No | Yes | Not Met | No | Yes | Partially Met | Yes | Met | Moderate |
| Raman 2020 | No | No | No | Not Met | No | Yes | Partially Met | Yes | Met | Moderate |
| Riestra-Ayora 2021 | No | No | Yes | Not Met | No | Yes | Partially Met | Yes | Met | Moderate |
| Rusetsky 2021 | No | No | No | Not Met | No | No | Not Met | Yes | Met | High |
| Shah 2020 | No | No | No | Not Met | No | Yes | Partially Met | Yes | Met | Moderate |
| Song 2021 | No | No | Yes | Not Met | No | Yes | Partially Met | Yes | Met | Moderate |
| Stavem 2020 | No | No | No | Not Met | No | Yes | Partially Met | No | Not Met | High |
| Stavem 2021 | Yes | Yes | No | Partially Met | No | Yes | Partially Met | Yes | Met | Moderate |

| | | | | | | | | | |
|---|---|---|---|---|---|---|---|---|---|
| Sykes 2021 | Yes | No | Yes | Not Met | No | Yes | Partially Met | Yes | Met | Moderate |
| Townsend 2020 | No | No | No | Not Met | No | Yes | Partially Met | Yes | Met | Moderate |
| Townsend 2021 | No | No | No | Not Met | No | Yes | Partially Met | Yes | Met | Moderate |
| Townsend 2021-2 | Yes | No | No | Not Met | No | Yes | Partially Met | Yes | Met | Moderate |
| Trunfio 2021 | Yes | Yes | Yes | Met | No | No | Not Met | Yes | Met | Moderate |
| Tudoran 2021 | No | Yes | No | Not Met | No | No | Not Met | Yes | Met | High |
| Vaira 2020 | No | No | Yes | Not Met | No | No | Not Met | Yes | Met | High |
| Walle-Hansen 2021 | Yes | No | No | Not Met | No | Yes | Partially Met | Yes | Met | Moderate |
| Wang 2020 | Yes | Yes | No | Partially Met | No | No | Not Met | Yes | Met | Moderate |
| Weerahandi 2021 | No | No | No | Not Met | No | Yes | Partially Met | Yes | Met | Moderate |
| Zhang 2021 | No | No | No | Not Met | Yes | No | Partially Met | Yes | Met | Moderate |
| Clinically-diagnosed | | | | | | | | | | |
| Samimi Ardestani 2021 | No | No | No | Not Met | No | No | Not Met | Yes | Met | High |
| Arnold 2020 | No | No | No | Not Met | No | Yes | Partially Met | Yes | Met | Moderate |
| Einvik_2021 | Yes | No | No | Not Met | No | Yes | Partially Met | Yes | Met | Moderate |
| Guler 2021 | Yes | No | No | Not Met | No | Yes | Partially Met | Yes | Met | Moderate |
| Hittesdorf 2021 | yes | no | yes | Partially Met | yes | yes | Met | yes | Met | Moderate |
| Janiri_2021 | Yes | No | Yes | Not Met | No | Yes | Partially Met | Yes | Met | Moderate |
| Lerum 2020 | No | No | No | Not Met | Yes | Yes | Met | Yes | Met | Moderate |
| Lucidi_2021 | No | No | No | Not Met | No | Yes | Partially Met | Yes | Met | Moderate |
| Mallia_2021 | No | No | Yes | Not Met | No | Yes | Partially Met | Yes | Met | Moderate |
| Miyazato 2020 | No | No | Yes | Not Met | No | No | Not Met | Yes | Met | High |
| Naidu 2021 | Yes | No | No | Not Met | No | Yes | Partially Met | Yes | Met | Moderate |
| Nunez-Cortes 2021 | No | No | Yes | Not Met | Yes | Yes | Met | Yes | Met | Moderate |
| Writing Committee for the COMEBAC Study Group 2021 | No | No | No | Not Met | No | Yes | Partially Met | Yes | Met | Moderate |
| Shendy 2021 | Yes | No | No | Not Met | No | Yes | Partially Met | Yes | Met | Moderate |
| Simani 2021 | No | No | Yes | Not Met | No | Yes | Partially Met | Yes | Met | Moderate |
| Sudre 2021 | No | No | No | Not Met | No | Yes | Partially Met | No | Not Met | High |
| Tomasoni 2020 | No | No | No | Not Met | No | Yes | Partially Met | Yes | Met | Moderate |
| van den Borst 2020 | No | No | Yes | Not Met | Yes | Yes | Met | No | Not Met | High |
| Venturelli_2021 | No | No | No | Not Met | No | Yes | Partially Met | Yes | Met | Moderate |

| | | | | | | | | | | |
|---|---|---|---|---|---|---|---|---|---|---|
| Wu_2021 | No | No | No | Not Met | Yes | Yes | Met | Yes | Met | Moderate |
| Xiong 2020 | No | No | No | Not Met | No | No | Not Met | Yes | Met | High |
| Xu_2021 | No | No | Yes | Not Met | Yes | Yes | Met | Yes | Met | Moderate |

#### By Outcome

| | Were valid methods used for the identification of the outcome? | Was the outcome measured in a standard, reliable way for all participants? | Was the data for the outcome reported clearly and completely? |
|---|---|---|---|
| Laboratory-confirmed | | | |
| Akter 2020 | | | |
| Mobility issues: Confined to bed | No | Yes | Yes |
| Mobility issues: Some problems in walking | No | Yes | Yes |
| Self care: Unable to wash or dry myself | No | Yes | Yes |
| Self care: some problem in washing or dressing | No | Yes | Yes |
| Pain/discomfort: Extreme pain or discomfort | No | Yes | Yes |
| Pain/discomfort: Moderate pain or discomfort | No | Yes | Yes |
| Anxiety/depression: extremely anxious or depressed | No | Yes | Yes |
| Anxiety/depression: moderately anxious or depressed | No | Yes | Yes |
| Sleep: Can't sleep | No | Yes | Yes |
| Sleep: Disturbance in sound sleep | No | Yes | Yes |
| Sleep: Nightmare | No | Yes | Yes |
| Panic attack | No | Yes | Yes |
| Loss of concentration | No | Yes | Yes |
| Memory loss: extreme | No | Yes | Yes |
| Memory loss: moderate | No | Yes | Yes |
| Hair fall | No | Yes | Yes |
| Alemanno 2021 | | | |
| cognitive deficit - MoCA test | Yes | Yes | Yes |
| cognitive deficit - MMSE test | Yes | Yes | Yes |
| depression | No | Yes | Yes |
| PTSD symptoms | Yes | Yes | Yes |
| Alharthy 2020 | | | |
| Pulmonary embolism | No | No | Yes |
| Pulmonary hypertension | No | No | Yes |
| Breathing difficulties | No | No | Yes |
| Fatigue | No | No | Yes |
| Walking difficulties | No | No | Yes |
| Deep vein thrombosis | No | No | Yes |
| Interstitial lung disease | No | No | Yes |
| Symptomatic | No | No | Yes |
| AlShakhs 2021 | | | |
| sense of taste | No | Yes | No |
| Anastasio 2021 | | | |
| inactive | No | Yes | No |

| | | | |
|---|---|---|---|
| breathlessness (MMRc ≥ 2) | No | Yes | Yes |
| at least one symptom | No | No | No |
| dyspnea | No | No | Yes |
| weakness | No | No | Yes |
| joint and muscular pain | No | No | Yes |
| thoracic pain | No | No | Yes |
| anosmia and ageusia | No | No | Yes |
| depression | No | No | Yes |
| cough | No | No | Yes |
| heart palpitations | No | No | Yes |
| headache | No | No | Yes |
| sleeping disorders | No | No | Yes |
| heair loss | No | No | Yes |
| memory disorders | No | No | Yes |
| dizziness | No | No | Yes |
| Bellan 2021 | | | |
| Fever | No | No | Yes |
| Cough | No | No | Yes |
| Dyspnea | No | No | Yes |
| Ageusia | No | No | Yes |
| Anosmia | No | No | Yes |
| Diarrhea | No | No | Yes |
| Arthralgia | No | No | Yes |
| Myalgia | No | No | Yes |
| Chest pain | No | No | Yes |
| Sore throat | No | No | Yes |
| Headache | No | No | Yes |
| Pulmanory function tests | Yes | Yes | Yes |
| Inability to complete the assessment of the DLCO (the diffusion capacity of the lung for carbon monoxide) | Yes | Yes | Yes |
| Limited physical performance on SPPB test VB: In the publication this is referred to as "limited mobility" | Yes | Yes | Yes |
| 2-minute walk test | No | No | Yes |
| Some degree of functional impairment | No | No | Yes |
| Tolerance to exercise | No | No | Yes |
| Mild posttraumatic stress symptoms | No | Yes | Yes |
| Moderate posttraumatic stress symptoms | No | Yes | Yes |
| Severe posttraumatic stress symptoms | No | Yes | Yes |
| Blair 2021 | | | |
| not returned to usual health | No | Yes | No |
| not returned to doing usual activities | No | Yes | No |

| | | | |
|---|---|---|---|
| any symptoms | No | Yes | No |
| any symptoms - mild | No | Yes | No |
| any symptoms - moderate | No | Yes | No |
| perception of health - poor | No | Yes | No |
| effect of symptoms on activities - a little bit to very much | No | Yes | No |
| effect of symptoms on activities - a little bit | No | Yes | No |
| effect of symptoms on activities - somewhat | No | Yes | No |
| weakness | No | Yes | No |
| dry cough | No | Yes | No |
| Blanco 2021 | | | |
| Abnormal lung function by forced vital capacity | Yes | Yes | Yes |
| Abnormal lung function by forced expiratory volume | Yes | Yes | Yes |
| Abnormal lung function by Tiffeneau-Pinelli index | Yes | Yes | Yes |
| Abnormal lung function by diffusing capacity for carbon monoxide | Yes | Yes | Yes |
| 6MWT - Pathological | Yes | Yes | Yes |
| Pathological CT | Yes | Yes | Yes |
| Boscolo-Rizzo 2020 | | | |
| Fever | No | Yes | Yes |
| Dry cough or productive cough | No | Yes | Yes |
| Blocked nose | No | Yes | Yes |
| Problems breathing | No | Yes | Yes |
| Headache | No | Yes | Yes |
| Sore throat | No | Yes | Yes |
| Muscle or joint pain | No | Yes | Yes |
| Chest pain | No | Yes | Yes |
| Sinonasal pain | No | Yes | Yes |
| Loss of appetite | No | Yes | Yes |
| Felt tired | No | Yes | Yes |
| Diarrhea | No | Yes | Yes |
| Nausea | No | Yes | Yes |
| Vomiting | No | Yes | Yes |
| Abdominal pain | No | Yes | Yes |
| Dizziness | No | Yes | Yes |
| Altered sense of smell or taste | No | Yes | Yes |
| Brandao 2021 | | | |
| olfactory deficit (partial recovery or no recovery) | No | Yes | No |
| olfactory deficit (partial recovery) | No | Yes | No |
| olfactory deficit (no recovery) | No | Yes | No |
| taste dysfunction (partial recovery or no improvement) | No | Yes | No |

| | | | |
|---|---|---|---|
| taste dysfunction (partial recovery or no improvement) | No | Yes | No |
| taste dysfunction (no improvement) | No | Yes | No |
| Bulgurcu 2020 | | | |
| Non-recovery of smell loss | No | No | Yes |
| Non-recovery of taste loss | No | No | Yes |
| Buonsenso 2021 | | | |
| A bit more fatigue compared to before COVID−19 diagnosis | No | Yes | Yes |
| More fatigue compared to before COVID−19 diagnosis | No | Yes | Yes |
| Insomnia | No | Yes | Yes |
| Nasal congestion/rhinorrea | No | Yes | Yes |
| Persistent muscle pain | No | Yes | Yes |
| Headache | No | Yes | Yes |
| Lack of concentration | No | Yes | Yes |
| Weight loss | No | Yes | Yes |
| Joint pain or swelling | No | Yes | Yes |
| Skin rashes | No | Yes | Yes |
| Chest tightness | No | Yes | Yes |
| Constipation | No | Yes | Yes |
| Persistent cough | No | Yes | Yes |
| Altered smell | No | Yes | Yes |
| Palpitations | No | Yes | Yes |
| Chest pain | No | Yes | Yes |
| Altered taste | No | Yes | Yes |
| Hypersomnia | No | Yes | Yes |
| Stomach/abdominal pain | No | Yes | Yes |
| Diarrhoea | No | Yes | Yes |
| Menstruation | No | Yes | Yes |
| Other: yes | No | Yes | Yes |
| 1–2 persisting symptoms | No | Yes | Yes |
| 3 or more persisting symptoms | No | Yes | Yes |
| Symptoms distress child only a little | No | Yes | Yes |
| Symptoms distress child quite a lot | No | Yes | Yes |
| Symptoms distress child a great deal | No | Yes | Yes |
| Hair loss | No | Yes | Yes |
| Skin peeling | No | Yes | Yes |
| Carvalho-Schneider 2020 | | | |
| Fever | Yes | Yes | Yes |
| Dyspnea | No | Yes | Yes |
| Chest pain | No | Yes | Yes |
| Flulike symptoms | No | Yes | Yes |

| | | | |
|---|---|---|---|
| Digestive disorders | No | Yes | Yes |
| Weight loss >5% | Yes | Yes | Yes |
| Anosmia/ageusia | No | Yes | Yes |
| Palpitations* | No | No | Yes |
| Arthralgia | No | Yes | Yes |
| Cutaneous signs | No | Yes | Yes |
| Sick leave | No | Yes | Yes |
| Presence of one or more symptoms at follow-up | No | Yes | Yes |
| # of patients who still felt ill or were in worse clinical condition that at COVID-19 onset | No | Yes | Yes |
| Chiesa-Estomba 2020 | | | |
| Persistent subjective smell loss | No | Yes | Yes |
| Partial recovery of smell loss | No | Yes | Yes |
| Cortes-Telles 2021 | | | |
| fatigue on effort | No | Yes | Yes |
| dyspnoea | No | Yes | Yes |
| myalgias | No | Yes | Yes |
| cough | No | Yes | Yes |
| chest pain | No | Yes | Yes |
| sore throat | No | Yes | Yes |
| sputum production | No | Yes | Yes |
| diaphoresis | No | Yes | Yes |
| headache | No | Yes | Yes |
| rhinitis | No | Yes | Yes |
| telogen effluvium | No | Yes | Yes |
| anosmia/ageusia | No | Yes | Yes |
| dermatological symptoms | No | Yes | Yes |
| wheezing | No | Yes | Yes |
| conjunctivitis | No | Yes | Yes |
| diarrhea | No | Yes | Yes |
| D'Cruz 2020 | | | |
| Persistent symptoms | No | Yes | Yes |
| Disease-specific functional impairment | Yes | Yes | Yes |
| Burdensome breathlessness | No | Yes | Yes |
| Persistent cough | No | Yes | Yes |
| Burdensome cough | No | Yes | Yes |
| Fatigue | No | Yes | Yes |
| Sleep disturbance | No | Yes | Yes |
| Pain | No | Yes | Yes |
| Depression | No | Yes | Yes |

| | | | |
|---|---|---|---|
| Anxiety | No | Yes | Yes |
| Cognitive impairment | Yes | Yes | Yes |
| Post-Traumatic Stress Disorder | No | Yes | Yes |
| deGraaf 2021 | | | |
| Post COVID functional status score | No | No | Yes |
| chest pain | No | No | Yes |
| dyspnea | No | Yes | Yes |
| palpitations | Yes | Yes | Yes |
| Anxiety | No | Yes | Yes |
| Depression | No | Yes | Yes |
| PTSD | No | Yes | Yes |
| Psychiatric morbidities | No | No | Yes |
| Cognitive functioning | No | Yes | Yes |
| Cognitive functioning by partners | No | Yes | Yes |
| Einvik 2021 | | | |
| PTSD | No | Yes | Yes |
| Froidure 2021 | | | |
| fatigue | No | No | Yes |
| dyspnea | Yes | Yes | Yes |
| requiring oxygen supplementation at exercise | No | No | Yes |
| chronic dry cough | No | No | No |
| chest oppression | No | No | Yes |
| FEF <2 SD | Yes | Yes | Yes |
| impaired DLCO | Yes | Yes | Yes |
| moderate to severe DLCO impairment | Yes | Yes | Yes |
| Patients with impaired FVC (forced vital capacity) | Yes | Yes | Yes |
| Patients with impaired FEV1 (forced expired volume in 1) | Yes | Yes | Yes |
| Patients with impaired FEF25-75 (forced expiratory flow at 25–75% of forced vital capacity) | Yes | Yes | Yes |
| Patients with impaired DLCO (lung diffusion capacity) | Yes | Yes | Yes |
| ground glass opacities | Yes | Yes | Yes |
| consolidations | Yes | Yes | Yes |
| isolated reticulations | Yes | Yes | Yes |
| signs of fibrosis | Yes | Yes | Yes |
| lung parenchyma | Yes | Yes | Yes |
| Gambini 2020 | | | |
| Eye irritation - any symptom | No | No | Yes |
| Eye irritation - watering | No | No | Yes |
| Eye irritation - foreign body sensation | No | No | Yes |
| Eye irritation - red eye | No | No | Yes |

| | | | |
|---|---|---|---|
| Eye irritation - moderate to severe dry eye disease (DED) | No | No | No |
| Eye irritation - burning | No | No | Yes |
| Dry eye disease - mild | No | No | No |
| Dry eye disease - moderate to severe | Yes | Yes | No |
| Dry eye disease - mild | Yes | Yes | No |
| Eye irritation - reduced visual acuity in the near distance | No | No | Yes |
| Gonzalez 2021 | | | |
| symptomatic | Yes | Yes | Yes |
| Dry cough | Yes | Yes | Yes |
| Wet cough | Yes | Yes | Yes |
| Dyspnea - 1 | Yes | Yes | Yes |
| Dyspnea - 2 | Yes | Yes | Yes |
| Dyspnea - 4 | Yes | Yes | Yes |
| Muscular fatigue | Yes | Yes | Yes |
| Depression - borderline abnormal | No | Yes | Yes |
| Depression - abnormal | No | Yes | Yes |
| Anxiety - borderline abnormal | No | Yes | Yes |
| Anxiety - abnormal | No | Yes | Yes |
| Dyspnea - any | Yes | Yes | Yes |
| fever | Yes | Yes | No |
| wheeze | Yes | Yes | No |
| abdominal pain | Yes | Yes | No |
| receiving supplemental oxygen | Yes | Yes | No |
| incidental pulmonary thromboembolism | Yes | Yes | No |
| TLC ≤ 50%-80% | Yes | Yes | Yes |
| TLC < 50% | Yes | Yes | Yes |
| DLCO ≤ 60%-80% | Yes | Yes | Yes |
| DLCO < 60% | Yes | Yes | Yes |
| Chest CT - Density ground glass | Yes | Yes | Yes |
| Chest CT - Density mixed ground glass | Yes | Yes | Yes |
| Chest CT - Density consolidation | Yes | Yes | Yes |
| Chest CT - Internal structures interlobular septal thickening | Yes | Yes | Yes |
| Chest CT - Internal structures bronchiectasis | Yes | Yes | Yes |
| Chest CT - Internal structures atelectasis | Yes | Yes | Yes |
| Chest CT - Internal structures solid nodule | Yes | Yes | Yes |
| Chest CT - Nonsolid nodule | Yes | Yes | Yes |
| Chest CT - Lesions reticular | Yes | Yes | Yes |
| Chest CT - Lesions fibrotic | Yes | Yes | Yes |
| DLCO - abnormal | Yes | Yes | Yes |
| TLC - altered | Yes | Yes | Yes |

| | | | |
|---|---|---|---|
| Severe decrease of oxygen saturation after 6MWT | Yes | Yes | Yes |
| Chest CT - at least one lobe affected by ground glass or consolidative opacities | Yes | Yes | Yes |
| Halpin 2021 | | | |
| Fatigue - Any new fatigue | No | Yes | Yes |
| Fatigue - Mild (0-3) | No | Yes | Yes |
| Fatigue - Moderate (4-6) | No | Yes | Yes |
| Fatigue - Severe (7-10) | No | Yes | Yes |
| Breathlessness - Any new or worsened breathlessness | No | Yes | Yes |
| Breathlessness - Mild (increased by 1-3/10) | No | Yes | Yes |
| Breathlessness - Moderate (increased by 4-6/10) | No | Yes | Yes |
| Breathlessness - Severe (increased by 7-10/10) | No | Yes | Yes |
| Breathlessness - Increased at rest | No | Yes | Yes |
| Breathlessness - Increased on dressing | No | Yes | Yes |
| Breathlessness - Increased on stairs | No | Yes | Yes |
| Neuropsychological - Any PTSD symptoms related to illness | No | Yes | Yes |
| Neuropsychological - Mild symptoms | No | Yes | Yes |
| Neuropsychological - Moderate symptoms | No | Yes | Yes |
| Neuropsychological - Severe symptoms | No | Yes | Yes |
| Neuropsychological - Thoughts of self-harm | No | Yes | Yes |
| Neuropsychological - New or worsened concentration problem | No | Yes | Yes |
| Neuropsychological - New or worsened short-term memory problem | No | Yes | Yes |
| Speech and Swallow - Swallow problem | No | Yes | Yes |
| Speech and Swallow - Laryngeal sensitivity | No | Yes | Yes |
| Speech and Swallow - Voice change | No | Yes | Yes |
| Speech and Swallow - Communication difficulty | No | Yes | Yes |
| Speech and Swallow - SLT referral criteria met (impact rating of 1 or more in any SLT domain) | No | Yes | Yes |
| Nutrition - Concern about weight/nutrition | No | Yes | Yes |
| Nutrition - Appetite problem severity 2 or more | No | Yes | Yes |
| Nutrition - Dietetics referral criteria met (either of the above criteria) | No | Yes | Yes |
| Continence - New bowel control problem | No | Yes | Yes |
| Continence - New bladder control problem | No | Yes | Yes |
| EQ-5D-5L - Decreased by at least 0.05 | No | Yes | Yes |
| EQ-5D-5L - Worsened mobility | No | Yes | Yes |
| EQ-5D-5L - Worsened self-care | No | Yes | Yes |
| EQ-5D-5L - Worsened usual activities | No | Yes | Yes |
| EQ-5D-5L - Worsened pain/discomfort | No | Yes | Yes |
| EQ-5D-5L - Worsened anxiety/depression | No | Yes | Yes |
| Perceived health (self-rated 0-100 scale) - Decrease by more than 7 points | No | Yes | Yes |

| | | | |
|---|---|---|---|
| Han 2021 | | | |
| fibrotic-like changes | Yes | Yes | Yes |
| fibrotic-like abnormalities | Yes | Yes | Yes |
| residual ground glass opacity or interstitial thickening | Yes | Yes | No |
| unilateral lung involvement | Yes | Yes | Yes |
| bilateral lunch involvement | Yes | Yes | Yes |
| ground glass opacities | Yes | Yes | Yes |
| consolidation | Yes | Yes | Yes |
| reticulation | Yes | Yes | Yes |
| Presence of nodule or mass | Yes | Yes | Yes |
| Pleural effusion | Yes | Yes | Yes |
| Emphysema | Yes | Yes | Yes |
| Thickening of the adjacent pleura | Yes | Yes | Yes |
| Interlobar pleural traction | Yes | Yes | Yes |
| Honeycombing | Yes | Yes | Yes |
| Pulmonary atelectasis | Yes | Yes | Yes |
| Bronchiectasis | Yes | Yes | Yes |
| dry cough | No | No | Yes |
| expectoration | No | No | Yes |
| slight dyspnea on exertion | No | No | Yes |
| abnormal pulmonary diffusion | Yes | Yes | Yes |
| Havervall 2021 | | | |
| Any symptom – 2 months | No | Yes | Yes |
| Any symptom – 4 months | No | Yes | Yes |
| Any symptom – 8 months | No | Yes | Yes |
| Anosmia – 2 months | No | Yes | Yes |
| Anosmia – 4 months | No | Yes | Yes |
| Anosmia – 8 months | No | Yes | Yes |
| Fatigue – 2 months | No | Yes | Yes |
| Fatigue – 4 months | No | Yes | Yes |
| Fatigue – 8 months | No | Yes | Yes |
| Ageusia – 2 months | No | Yes | Yes |
| Ageusia – 4 months | No | Yes | Yes |
| Ageusia – 8 months | No | Yes | Yes |
| Dyspnea – 2 months | No | Yes | Yes |
| Dyspnea – 4 months | No | Yes | Yes |
| Dyspnea – 8 months | No | Yes | Yes |
| Sleeping disorder – 2 months | No | Yes | Yes |
| Sleeping disorder – 4 months | No | Yes | Yes |
| Sleeping disorder – 8 months | No | Yes | Yes |

| | | | |
|---|---|---|---|
| Headache – 2 months | No | Yes | Yes |
| Headache – 4 months | No | Yes | Yes |
| Headache – 8 months | No | Yes | Yes |
| Palpitations – 2 months | No | Yes | Yes |
| Palpitations – 4 months | No | Yes | No |
| Palpitations – 8 months | No | Yes | Yes |
| Concentration impairment – 2 months | No | Yes | Yes |
| Concentration impairment – 4 months | No | Yes | Yes |
| Concentration impairment – 8 months | No | Yes | Yes |
| Muscle/ joint pain – 2 months | No | Yes | Yes |
| Muscle/ joint pain – 4 months | No | Yes | Yes |
| Muscle/ joint pain – 8 months | No | Yes | Yes |
| memory impairment – 2 months | No | Yes | Yes |
| memory impairment – 4 months | No | Yes | Yes |
| memory impairment – 8 months | No | Yes | Yes |
| at least 1 moderate to severe symptom – 2 months | No | Yes | Yes |
| at least 1 moderate to severe symptom – 4 months | No | Yes | Yes |
| moderately or markedly disrupted work life – 2 months | No | Yes | Yes |
| moderately or markedly disrupted social life – 2 months | No | Yes | Yes |
| moderately or markedly disrupted home life – 2 months | No | Yes | Yes |
| moderate or marked disruption in any Sheehan Diasbility Scale Category and at least 1 moderate or severe symptom lasting at least 8 months | No | Yes | Yes |
| **Horn 2020** | | | |
| PTSD - probable | No | Yes | Yes |
| PTSD | No | Yes | Yes |
| **Huang 2020** | | | |
| Reported 1 symptom at follow-up | No | Yes | Yes |
| Fatigue or muscle weakness | No | Yes | Yes |
| Sleep difficulties | No | Yes | Yes |
| Hair loss | No | Yes | Yes |
| Smell disorder | No | Yes | Yes |
| Palpitations | No | Yes | Yes |
| Joint pain | No | Yes | Yes |
| Decreased appetite | No | Yes | Yes |
| Taste disorder | No | Yes | Yes |
| Dizziness | No | Yes | Yes |
| Diarrhoea or vomiting | No | Yes | Yes |
| Chest pain | No | Yes | Yes |
| Sore throat or difficult to swallow | No | Yes | Yes |
| Skin rash | No | Yes | Yes |

| | | | |
|---|---|---|---|
| Myalgia | No | Yes | Yes |
| Headache | No | Yes | Yes |
| Low grade fever | No | Yes | Yes |
| Dyspnoea | No | Yes | Yes |
| QoL - Mobility: problems with walking around | No | Yes | Yes |
| QoL -Personal care: problems with washing or dishing | No | Yes | Yes |
| QoL -Usual activity: problems with usual activity* | No | No | Yes |
| QoL -Pain or discomfort | No | Yes | Yes |
| QoL -Anxiety or depression | No | Yes | Yes |
| QoL - distance walked in 6 minutes | No | No | Yes |
| Jacobs 2020 | | | |
| Fatigue | No | No | Yes |
| Shortness of breath | No | No | Yes |
| Cough | No | No | Yes |
| Lack of taste | No | No | Yes |
| Muscular pain | No | No | Yes |
| Diarrhea | No | No | Yes |
| Lack of smell | No | No | Yes |
| Phlegm | No | No | Yes |
| Headache | No | No | Yes |
| Joint pain | No | No | Yes |
| Confusion | No | No | Yes |
| Eye irritation | No | No | Yes |
| Fever | No | No | Yes |
| Ulcer | No | No | Yes |
| General health - poor, fair | No | No | Yes |
| Quality of life - poor, fair | No | No | Yes |
| Physical health - poor, fair | No | No | Yes |
| Mental health - poor, fair | No | No | Yes |
| Social relationships - poor, fair | No | No | Yes |
| Social active role - poor, fair | No | No | Yes |
| Physical activity - not at all, a little | No | No | Yes |
| Emotional - always, often | No | No | Yes |
| Fatigue - severe, very severe | No | No | Yes |
| Fatigue - moderate | No | No | Yes |
| Dyspnea - Dressing - some/much difficulty | No | No | Yes |
| Dyspnea - Walking - some/much difficulty | No | No | Yes |
| Dyspnea - Stairs - some/much difficulty | No | No | Yes |
| Dyspnea - Meal preparation - some/much difficulty | No | No | Yes |
| Dyspnea - Wash dishes - some/much difficulty | No | No | Yes |

| | | | |
|---|---|---|---|
| Dyspnea - Sweep - some/much difficulty | No | No | Yes |
| Dyspnea - Make bed - some/much difficulty | No | No | Yes |
| Dyspnea - Lift - some/much difficulty | No | No | Yes |
| Dyspnea - Lift and carry - some/much difficulty | No | No | Yes |
| Dyspnea - Walk fast - some/much difficulty | No | No | Yes |
| Jacobson 2021 | | | |
| Fatigue | Yes | Yes | Yes |
| Dyspnea | Yes | Yes | Yes |
| Loss of taste/smell | Yes | Yes | Yes |
| Myalgias | Yes | Yes | Yes |
| Memory problems | Yes | Yes | Yes |
| Chest pain | Yes | Yes | Yes |
| Hair loss | Yes | Yes | Yes |
| Cough | Yes | Yes | Yes |
| Headache | Yes | Yes | Yes |
| Congestion/Rhinorrhea | Yes | Yes | Yes |
| Nausea/Vomiting/Diarrhea | Yes | Yes | Yes |
| Palpitations | Yes | Yes | Yes |
| Sore throat | Yes | Yes | Yes |
| Fever/Chills | Yes | Yes | Yes |
| Any Symptom | Yes | Yes | Yes |
| >=2 Symptoms | Yes | Yes | Yes |
| >=3 Symptoms | Yes | Yes | Yes |
| Missed work due to health | No | Yes | Yes |
| Any work impairment due to health | No | Yes | Yes |
| Any activity impairment due to health | No | Yes | Yes |
| Khasawneh 2020 | | | |
| Duration of symptoms | No | Yes | Yes |
| Klein 2021 | | | |
| Any unresolved symptom | No | Yes | Yes |
| Fatigue | No | Yes | Yes |
| Smell changes | No | Yes | Yes |
| breathing difficulty | No | Yes | Yes |
| Taste changes | No | Yes | Yes |
| Memory disoder | No | Yes | Yes |
| Muscle aches | No | Yes | Yes |
| Headache | No | Yes | Yes |
| Musculoskeletal pain | No | Yes | Yes |
| Hair loss | No | Yes | Yes |
| Anxiety and depression | No | Yes | Yes |

| | | | |
|---|---|---|---|
| Abdominal pain | No | Yes | Yes |
| Concentration disorders | No | Yes | Yes |
| Diarrhea | No | Yes | Yes |
| Dizziness | No | Yes | Yes |
| Ear pain | No | Yes | Yes |
| Eye disorders | No | Yes | Yes |
| Hearing disorder | No | Yes | Yes |
| Low physical performance | No | Yes | Yes |
| Mouth sores | No | Yes | Yes |
| Nose blockage | No | Yes | Yes |
| Palpitations | No | Yes | Yes |
| Paresthesia | No | Yes | Yes |
| Throat ache | No | Yes | Yes |
| Vomiting | No | Yes | Yes |
| **Konstantinidis 2020** | | | |
| Persistent chemosensory deficits | No | No | Yes |
| **Landi 2021** | | | |
| Cough | No | No | Yes |
| Fatigue | No | No | Yes |
| Diarrhea | No | No | Yes |
| Headache | No | No | Yes |
| Smell disorders | No | No | Yes |
| Dysgeusia | No | No | Yes |
| Red eyes | No | No | Yes |
| Joint pain | No | No | Yes |
| Short of breath | No | No | Yes |
| Loss of appetite | No | No | Yes |
| Sore throat | No | No | Yes |
| Rhinitis | No | No | Yes |
| Fever | No | No | Yes |
| No clinical improvement | No | No | Yes |
| **Lascarrou 2021** | | | |
| Breathing with assistance | No | No | Yes |
| **Liang 2020** | | | |
| Return to work | No | Yes | Yes |
| Impaired pulmonary function | Yes | Yes | Yes |
| Abnormal Lung HRCT | Yes | Yes | Yes |
| **Logue 2021** | | | |
| 1-2 persistent symptoms | No | Yes | Yes |
| 3+ persistent symptoms | No | Yes | Yes |

| | | | |
|---|---|---|---|
| Fatigue | No | Yes | Yes |
| Loss of smell or taste | No | Yes | Yes |
| Brain fog | No | Yes | Yes |
| Other symptoms | No | Yes | Yes |
| Quality of Life (HRQoL) | No | Yes | Yes |
| Activity of daily living (ADL) | No | Yes | Yes |
| **Mafort 2021** | | | |
| general fatigue | No | No | Yes |
| dyspnoea | No | No | Yes |
| cough | No | No | Yes |
| fever | No | No | Yes |
| **Mandal 2020** | | | |
| Breathlessness | No | Yes | Yes |
| Cough | No | Yes | Yes |
| Fatigue | No | Yes | Yes |
| Depression | No | Yes | Yes |
| Anosmia | No | Yes | Yes |
| One or more of the following persistent symptoms (breathlessness, cough, fatigue & poor sleep quality) | No | Yes | Yes |
| **Mazza 2021** | | | |
| Depression - ZSDS index ≥ 50 Yes | No | Yes | No |
| Depression - BDI-13 ≥ 8 Yes | No | Yes | Yes |
| PTSD - IES-R ≥ 33 Yes | No | Yes | No |
| PTSD - PCL-5 ≥ 33 Yes | No | Yes | No |
| Anxiety - STAI-Y state ≥ 40 Yes | No | Yes | No |
| Obsessive compulsive - OCI ≥ 21 Yes | No | Yes | No |
| Insomnia - WHIIRS ≥ 9 Yes | No | Yes | No |
| Verbal memory poor performance | Yes | Yes | No |
| Verbal fluency poor performance | Yes | Yes | No |
| Working Memory poor performance | Yes | Yes | No |
| Attention and Information Processing poor performance | Yes | Yes | No |
| Psychomotor coordination poor performance | Yes | Yes | No |
| Executive Functions poor performance | Yes | Yes | No |
| Symptoms in at least one psychopathological dimension | No | Yes | Yes |
| At least one current major psychiatric disorder | No | Yes | Yes |
| Major depressive disorder | No | Yes | Yes |
| Anxiety Disorders | No | Yes | Yes |
| Insomnia | No | Yes | Yes |
| Other major psychiatric disorder | No | Yes | Yes |
| Poor performance in one neurocognitive function | Yes | Yes | Yes |

| | | | |
|---|---|---|---|
| Poor performance in two neurocognitive functions | Yes | Yes | Yes |
| Poor performance in three neurocognitive functions | Yes | Yes | Yes |
| Poor performance in four neurocognitive functions | Yes | Yes | Yes |
| Poor performance in five neurocognitive functions | Yes | Yes | Yes |
| Poor performance in all neurocognitive functions | Yes | Yes | Yes |
| Moradian 2020 | | | |
| Fever | No | Yes | Yes |
| Dyspnea | No | Yes | Yes |
| Cough | No | Yes | Yes |
| myalgia | No | Yes | Yes |
| activity intolerance | No | Yes | Yes |
| fatigue | No | Yes | Yes |
| weakness | No | Yes | Yes |
| weight loss | No | Yes | Yes |
| dizziness | No | Yes | Yes |
| headache | No | Yes | Yes |
| shivering | No | Yes | Yes |
| otalgia | No | Yes | Yes |
| sore throat | No | Yes | Yes |
| sputum | No | Yes | Yes |
| sneezing | No | Yes | Yes |
| rainfall | No | Yes | Yes |
| odor disorder | No | Yes | Yes |
| taste disorder | No | Yes | Yes |
| nausea | No | Yes | Yes |
| vomiting | No | Yes | Yes |
| diarrhea | No | Yes | Yes |
| anorexia | No | Yes | Yes |
| dyspepsia | No | Yes | Yes |
| anxiety | No | Yes | Yes |
| More than 5 symptoms | No | Yes | Yes |
| Moreno-Perez 2021 | | | |
| Post-acute COVID-19 syndrome (PCS) | No | No | Yes |
| Fatigue | No | No | Yes |
| Anosmia-dysgeusia | No | No | Yes |
| Myalgias-arthralgias | No | No | Yes |
| Dyspnea | No | No | Yes |
| Cough | No | No | Yes |
| Any headache | No | No | Yes |
| Moderate-severe headache | No | No | Yes |

| | | | |
|---|---|---|---|
| Any headache | No | No | Yes |
| Mnesic complaints | No | No | Yes |
| Diarrhoea | No | No | Yes |
| Skin features | No | No | Yes |
| Visual loss | No | No | Yes |
| Fever | No | No | Yes |
| Any headache - de novo | No | No | Yes |
| Mnesic complaints - clinically relevant | No | No | Yes |
| Mnesic complaints - de novo | No | No | Yes |
| not recovered | No | No | Yes |
| relevant neurological symptoms | No | No | Yes |
| Niklassen 2021 | | | |
| Anosmia | Yes | No | Yes |
| Hyposmia | Yes | No | Yes |
| Normosmia | Yes | No | Yes |
| Gustatory dysfunction | Yes | No | Yes |
| Otte 2020 | | | |
| Self-reported olfactory and tasting impairment | No | Yes | Yes |
| Hyposmia | Yes | Yes | Yes |
| Anosmia | Yes | Yes | Yes |
| Petersen 2020 | | | |
| Persistent Loss of smell | No | No | Yes |
| Loss of smell | No | No | No |
| Persistent Loss of taste | No | No | Yes |
| Loss of taste | No | No | No |
| Persistent Fatigue | No | No | Yes |
| Fatigue | No | No | Yes |
| Persistent Headache | No | No | Yes |
| Had symptoms at last follow-up | No | No | Yes |
| Had 1-2 symptoms at last follow-up | No | No | Yes |
| Had 3-5 symptoms at last follow-up | No | No | Yes |
| Had 6-8 symptoms at last follow-up | No | No | Yes |
| Had 9-12 symptoms at last follow-up | No | No | Yes |
| Had 13+ symptoms at last follow-up | No | No | Yes |
| Fever | No | No | No |
| Headache | No | No | No |
| Chills | No | No | No |
| Myalgia | No | No | No |
| Dry cough | No | No | No |
| Rhinorrhea | No | No | No |

| | | | |
|---|---|---|---|
| Anorexia | No | No | No |
| Sore throat | No | No | No |
| Arthralgia | No | No | No |
| Dyspnea | No | No | No |
| Diarrhea | No | No | No |
| Cough with expectoration | No | No | No |
| Nausea | No | No | No |
| Chest tightness | No | No | No |
| Rashes | No | No | No |
| Printza 2021 | | | |
| Olfactory disorders | No | Yes | Yes |
| Gustatory disorders | No | Yes | Yes |
| Olfactory AND gustatory disorder | No | Yes | Yes |
| Nasal obstruction | No | Yes | Yes |
| Rhinorrhea | No | Yes | Yes |
| Allergic rhinitis | No | Yes | Yes |
| Chronic rhinosinusitis | No | Yes | Yes |
| Smell loss severity - MILD | No | Yes | Yes |
| Smell loss severity - MODERATE | No | Yes | Yes |
| Smell loss severity - SEVERE | No | Yes | Yes |
| Smell loss severity - Extremely severe (anosmia) | No | Yes | Yes |
| Taste loss severity - MILD | No | Yes | Yes |
| Taste loss severity - MODERATE | No | Yes | Yes |
| Taste loss severity - SEVERE | No | Yes | Yes |
| Taste loss severity - Extremely severe | No | Yes | Yes |
| Hyposmia before other symptoms | No | Yes | Yes |
| Qu 2021 | | | |
| One or more uncomfortable symptoms | No | Yes | Yes |
| 1 physical symptom | No | Yes | Yes |
| 2 physical symptoms | No | Yes | Yes |
| 3 physical symptoms | No | Yes | Yes |
| 4 physical symptoms | No | Yes | Yes |
| 5+ physical symptoms | No | Yes | Yes |
| HRQoL - physical component summary (PCS) | Yes | Yes | Yes |
| HRQoL - mental component summary (MCS) | Yes | Yes | Yes |
| Fatigue | No | Yes | Yes |
| Cough | No | Yes | Yes |
| Sputum | No | Yes | Yes |
| Dyspnoea | No | Yes | Yes |
| Diarrhoea | No | Yes | Yes |

| | | | |
|---|---|---|---|
| Shortness of breath | No | Yes | Yes |
| Joint pain | No | Yes | Yes |
| Dysbasia | No | Yes | Yes |
| Palpitations | No | Yes | Yes |
| Other symptoms | No | Yes | Yes |
| Raman 2020 | | | |
| Breathlessness | No | Yes | Yes |
| Fatigue | No | Yes | Yes |
| Desaturation at the end of a 6-minute walk test* | No | No | Yes |
| Cognitive function - abnormal | No | Yes | Yes |
| Minimal anxiety | No | Yes | Yes |
| Mild anxiety | No | Yes | Yes |
| Moderate anxiety | No | Yes | Yes |
| Severe anxiety | No | Yes | Yes |
| Moderate or worse anxiety | No | Yes | Yes |
| Minimal depression | No | Yes | Yes |
| Mild depression | No | Yes | Yes |
| Moderate depression | No | Yes | Yes |
| Moderately severe or severe depression | No | Yes | Yes |
| Moderate or worse mood symptoms (depression) | No | Yes | Yes |
| Riestra-Ayora 2021 | | | |
| headache | No | No | Yes |
| cough | No | No | Yes |
| dysthermia | No | No | Yes |
| dyspnea | No | No | Yes |
| myalgia | No | No | Yes |
| asthenia | No | No | Yes |
| hypogeusia | No | Yes | Yes |
| hyposmia | No | Yes | Yes |
| nasal obstruction | No | No | Yes |
| dysosmia | No | No | Yes |
| olfactory dysfunction | No | Yes | Yes |
| gustatory dysfunction | No | Yes | Yes |
| olfactory symptoms | No | Yes | Yes |
| olfactory symptoms | No | Yes | Yes |
| Rusetsky 2021 | | | |
| Hyposmia - 60 days after discharge | No | No | Yes |
| Persistent Hyposmia - more than 30 days after discharge | No | No | Yes |
| Shah 2020 | | | |
| Dyspnoea | No | Yes | Yes |

| | | | |
|---|---|---|---|
| 6MWT - abnormal | Yes | Yes | Yes |
| Cough | No | Yes | Yes |
| Song 2021 | | | |
| Loss of smell | No | Yes | Yes |
| Loss of taste | No | Yes | Yes |
| persistent Loss of smell | No | Yes | Yes |
| persistent Loss of taste | No | Yes | Yes |
| Stavem 2020 | | | |
| Fever | No | Yes | No |
| Loss/disturbance of taste | No | Yes | No |
| Headache | No | Yes | No |
| Dry cough | No | Yes | No |
| Loss/disturbance of smell | No | Yes | No |
| Myalgia | No | Yes | No |
| Chills | No | Yes | No |
| Dyspnea | No | Yes | No |
| Sore throat | No | Yes | No |
| Arthralgia | No | Yes | No |
| Runny nose | No | Yes | No |
| Diarrhoea | No | Yes | No |
| Abdominal pain | No | Yes | No |
| Productive cough | No | Yes | No |
| Vomiting/nausea | No | Yes | No |
| Wheeze | No | Yes | No |
| Confusion/changed consciousness | No | Yes | No |
| Skin rash | No | Yes | No |
| Vision disturbance/blurring | No | Yes | No |
| Ear pain | No | Yes | No |
| Seizures/cramps | No | Yes | No |
| Conjunctivitis | No | Yes | No |
| Any symptom[#] | No | Yes | Yes |
| Stavem 2021 | | | |
| Fatigue | No | Yes | Yes |
| Sykes 2021 | | | |
| At least one residual symptom | No | Yes | Yes |
| Dyspnoea | No | Yes | Yes |
| Myalgia | No | Yes | Yes |
| Anxiety | No | Yes | Yes |
| Extreme fatigue | No | Yes | Yes |
| Low mood | No | Yes | Yes |

| | | | |
|---|---|---|---|
| Low mood | No | Yes | Yes |
| Memory impairment | No | Yes | Yes |
| Sleep disturbances | No | Yes | Yes |
| Cough | No | Yes | Yes |
| Attention deficit | No | Yes | Yes |
| Pleuritic chest pain | No | Yes | Yes |
| Sore throat | No | Yes | Yes |
| Fever | No | Yes | Yes |
| Anosmia | No | Yes | Yes |
| Cognitive impairment | No | Yes | Yes |
| Taste deficiency | No | Yes | Yes |
| Rash | No | Yes | Yes |
| Symptom cluster A | No | Yes | Yes |
| Symptom cluster B | No | Yes | Yes |
| Symptom cluster C | No | Yes | Yes |
| **Townsend 2020** | | | |
| Fatigue | No | Yes | Yes |
| Fatigue - <8 weeks | No | Yes | Yes |
| Fatigue - 8-10 weeks | No | Yes | Yes |
| Fatigue - 10-12 weeks | No | Yes | Yes |
| Fatigue - 12 weeks | No | Yes | Yes |
| Not feeling back to full health | No | Yes | Yes |
| Not returning to work (among those that had been employed)* | No | No | Yes |
| **Townsend 2021** | | | |
| Significant oxygen desaturation during 6MWT | Yes | Yes | Yes |
| Perceived lack of full health | No | Yes | Yes |
| Fatigue | No | Yes | Yes |
| **Townsend 2021-2** | | | |
| Fatigue | No | Yes | Yes |
| Breathlessness | No | Yes | Yes |
| **Trunfio 2021** | | | |
| any sequelae | No | No | Yes |
| dyspnea | No | No | Yes |
| olfactory/gustatory dysfunction | No | No | Yes |
| chronic cough | No | No | Yes |
| others | No | No | Yes |
| **Tudoran 2021** | | | |
| Symptoms (most frequently fatigue, dyspnea, and palpitations) | No | No | Yes |
| **Vaira 2020** | | | |
| Olfactory dysfunction | No | No | Yes |

| | | | |
|---|---|---|---|
| Taste disorder | No | No | Yes |
| Combined chemosensitive dysfunction | No | No | Yes |
| Isolated smell impairments | No | No | Yes |
| Isolated taste disorders | No | No | Yes |
| Persistent chemosensitive disorders | No | No | Yes |
| Walle-Hansen 2021 | | | |
| Worse health-related quality of life | No | Yes | Yes |
| Decline in mobility | No | Yes | Yes |
| Decline in ability to perform daily activities | No | Yes | Yes |
| More pain or discomfort | No | Yes | Yes |
| Increased anxiety | No | Yes | Yes |
| Decline in self-care ability | No | Yes | Yes |
| Major change in mobility | No | Yes | Yes |
| Major change in usual activities | No | Yes | Yes |
| Negative change in cognitive function | No | No | Yes |
| Wang 2020 | | | |
| PTSD - potential risk | No | No | Yes |
| PTSD | No | No | Yes |
| Postpartum depression - minor | No | No | Yes |
| Postpartum depression - major | No | No | Yes |
| Suffering from PTSD or depression | No | No | Yes |
| Weerahandi 2021 | | | |
| Dyspnea | No | Yes | Yes |
| Dyspnea - severe | No | Yes | Yes |
| Requiring oxygen | No | Yes | Yes |
| Zhang 2021 | | | |
| cumulative values of complete absorption | Yes | No | Yes |
| cumulative values of absorption of fibrosis-like findings | Yes | No | Yes |
| Absorption of lesions | Yes | No | Yes |
| distribution | Yes | No | Yes |
| Density | Yes | No | Yes |
| Fibrosis like findings | Yes | No | Yes |
| Involvement of lobe | Yes | No | Yes |
| Clinically-diagnosed | | | |
| Samimi Ardestani 2021 | | | |
| Olfactory dysfunction - no change | No | No | Yes |
| Olfactory dysfunction - a little change | No | No | Yes |
| Olfactory dysfunction - not recovered | No | No | Yes |
| Arnold 2020 | | | |
| At least one ongoing symptom | No | Yes | Yes |

| | | | |
|---|---|---|---|
| Fever | No | Yes | Yes |
| Cough | No | Yes | Yes |
| Breathlessness | No | Yes | Yes |
| Anosmia | No | Yes | Yes |
| Excessive fatigue | No | Yes | Yes |
| Myalgia | No | Yes | Yes |
| Headache | No | Yes | Yes |
| Chest pain | No | Yes | Yes |
| Arthralgia | No | Yes | Yes |
| Diarrhoea | No | Yes | Yes |
| Abdominal pain | No | Yes | Yes |
| Nausea | No | Yes | Yes |
| Insomnia | No | Yes | Yes |
| Significant desaturation on STS test | Yes | Yes | Yes |
| Einvik_2021 | | | |
| PTSD | No | Yes | Yes |
| Guler 2021 | | | |
| Cough | No | Yes | Yes |
| Hittesdorf 2021 | | | |
| Renal replacement therapy - 30 days | yes | yes | yes |
| Renal replacement therapy - at discharge | yes | yes | yes |
| Renal replacement therapy - 90 days | yes | yes | yes |
| Worse serum creatinine | no | no | yes |
| Janiri_2021 | | | |
| PTSD | No | Yes | Yes |
| Depressive episode | No | Yes | Yes |
| Hypomanic episode | No | Yes | Yes |
| Generalized anxiety disorder | No | Yes | Yes |
| Psychotic disorders | No | Yes | Yes |
| Lerum 2020 | | | |
| FVC < LLN | Yes | Yes | Yes |
| FEV1 < LLN | Yes | Yes | Yes |
| DLCO < LLN | Yes | Yes | Yes |
| Dyspnoea > 0 mMRC | No | Yes | Yes |
| Ground-glass chest opacities | Yes | Yes | Yes |
| Parenchymal bands | Yes | Yes | Yes |
| Lucidi_2021 | | | |
| anosmia/ hyposmia | No | Yes | Yes |
| partial recovery of anosmia/ hyposmia | No | Yes | Yes |

| | | | |
|---|---|---|---|
| persistent OD | No | Yes | Yes |
| Mallia_2021 | | | |
| Any symptom(s) | No | Yes | Yes |
| Breathlessness | No | Yes | Yes |
| Fatigue | No | Yes | Yes |
| Cough | No | Yes | Yes |
| Chest Pain | No | Yes | Yes |
| At least two symptoms among symptomatic patients | No | Yes | Yes |
| No return to pre-illness exercise tolerance level | No | Yes | Yes |
| Desaturation on sit-to-stand test | No | No | Yes |
| Psychological distress related to hospital admission (PTSD) | No | Yes | Yes |
| Abnormal chest radiograph | Yes | Yes | Yes |
| Any symptom(s) | No | Yes | Yes |
| 1 symptom | No | Yes | Yes |
| 2 symptoms | No | Yes | Yes |
| 3 symptoms | No | Yes | Yes |
| 4 symptoms | No | Yes | Yes |
| Cough | No | Yes | Yes |
| breathlessness | No | Yes | Yes |
| Chest Pain | No | Yes | Yes |
| Fatigue | No | Yes | Yes |
| Abnormal chest radiograph | Yes | Yes | Yes |
| CRP (abornmal) | Yes | Yes | Yes |
| Ferritin (abornmal) | Yes | Yes | Yes |
| D-dimer (abornmal) | Yes | Yes | Yes |
| Miyazato 2020 | | | |
| Persistent Cough - 60 days | No | No | Yes |
| Persistent Fatigue - 60 days | No | No | Yes |
| Persistent Dyspnea - 60 days | No | No | Yes |
| Persistent dysgeusia - 60 days | No | No | Yes |
| Persistent Dysosmia - 60 days | No | No | Yes |
| Persistent Cough - 120 days | No | No | Yes |
| Persistent Fatigue - 120 days | No | No | Yes |
| Persistent Dyspnea - 120 days | No | No | Yes |
| Persistent dysgeusia - 120 days | No | No | Yes |
| Persistent Dysosmia - 120 days | No | No | Yes |
| Late-onset Dysosmia - 32 days | No | No | Yes |
| Late-onset Dysosmia - 92 days | No | No | Yes |
| Alopecia | No | No | Yes |
| Naidu_2021 | | | |

| | | | |
|---|---|---|---|
| Persisting physical and psychiatric symptoms | No | No | Yes |
| Depression | No | Yes | Yes |
| PTSD | No | Yes | Yes |
| Any persistent symptoms | No | Yes | Yes |
| Anorexia | No | Yes | Yes |
| Not back to work | No | Yes | Yes |
| Confusion or “fuzzy head” | No | Yes | Yes |
| Myalgia | No | Yes | Yes |
| Non-improved breathlessness | No | Yes | Yes |
| Non-improved cough | No | Yes | Yes |
| Non-improved fatigue | No | Yes | Yes |
| Non-improved sleep quality | No | Yes | Yes |
| Nunez-Cortes_2021 | | | |
| Repetitions <2.5th percentile of 1STST | Yes | Yes | Yes |
| Repetitions <25th percentile of 1STST | Yes | Yes | Yes |
| Change in SpO2≥4 | Yes | Yes | Yes |
| SPO2 <90% post test | Yes | Yes | Yes |
| Writing Committee for the COMEBAC Study Group 2021 | | | |
| dyspnea (new onset) | No | Yes | Yes |
| Anosmia | No | Yes | Yes |
| headaches | No | Yes | Yes |
| paresthesia | No | Yes | Yes |
| anorexia | No | Yes | Yes |
| fatigue | No | Yes | Yes |
| weight loss | No | Yes | Yes |
| chest discomfort/ pain | No | Yes | Yes |
| cough | No | Yes | Yes |
| limb palsy | No | Yes | Yes |
| memory difficulties | No | Yes | Yes |
| mental slowness | No | Yes | Yes |
| concentration problems | No | Yes | Yes |
| persistent cough | No | Yes | Yes |
| abnormal lung CT scan | Yes | Yes | Yes |
| persistent ground glass opacities | Yes | Yes | Yes |
| lung fibrotic lesions | Yes | Yes | Yes |
| DLCO <70% | Yes | Yes | Yes |
| RV dilation on ECG | Yes | Yes | Yes |
| LVEF 40-50% on ECG | Yes | Yes | Yes |
| Cognitive complaint | No | No | Yes |
| cognitive impairment | No | Yes | Yes |

| | | | |
|---|---|---|---|
| anxiety | No | Yes | Yes |
| Depression | No | Yes | Yes |
| insomnia | No | Yes | Yes |
| PTSD | No | Yes | Yes |
| Dysfunctional breathing | No | Yes | Yes |
| Positive hyperventilation provocation test | No | No | Yes |
| Shendy_2021 | | | |
| fatigue | No | Yes | No |
| dyspnea | No | Yes | Yes |
| No Dyspnea | No | Yes | Yes |
| Mild Dyspnea | No | Yes | Yes |
| Moderate Dyspnea | No | Yes | Yes |
| Severe Dyspnea | No | Yes | Yes |
| Simani_2021 | | | |
| Chronic fatigue syndrome/myalgic encephalomyelitis (CFS/ME) *with and without PTSD | No | Yes | Yes |
| PTSD | No | Yes | Yes |
| CFSLWIFS (CFS-like with insufficient fatigue syndrome) | No | Yes | Yes |
| CIF (chronic idiopathic fatigue) | No | Yes | Yes |
| CFS (chronic fatigue syndrome) | No | Yes | Yes |
| Sudre_2021 | | | |
| fatigue | No | Yes | No |
| headache | No | Yes | No |
| shortness of breath | No | Yes | No |
| loss of smell | No | Yes | No |
| persistent cough | No | Yes | No |
| sore throat | No | Yes | No |
| fever | No | No | No |
| unusual muscle pains | No | Yes | No |
| skipped meals | No | Yes | No |
| chest pain | No | Yes | No |
| diarrhoea | No | Yes | No |
| hoarse voice | No | Yes | No |
| abdominal pain | No | Yes | No |
| delirium | No | Yes | No |
| any symptoms at >=2 months | No | Yes | Yes |
| any symptoms at >=3 months | No | Yes | Yes |
| Tomasoni 2020 | | | |
| Anxiety | No | Yes | Yes |
| Depression | No | Yes | Yes |

| | | | |
|---|---|---|---|
| Anxiety and depression | No | Yes | Yes |
| Anxiety, no depression | No | Yes | Yes |
| Depression, no anxiety | No | Yes | Yes |
| Either anxiety or depression | No | Yes | Yes |
| Persistence of at least one physical symptom | No | Yes | Yes |
| Cognitive deficits (memory disorder) - ongoing | Yes | Yes | Yes |
| Anosmia - ongoing | No | Yes | Yes |
| Dysgeusia - ongoing | No | Yes | Yes |
| Gastro-intestinal symptoms - ongoing | No | Yes | Yes |
| Fever - ongoing | Yes | Yes | Yes |
| Burning pain - ongoing | No | Yes | Yes |
| Dyspnea - ongoing | No | Yes | Yes |
| Asthenia - ongoing | No | Yes | Yes |
| Other symptoms - ongoing | No | Yes | Yes |
| van den Borst 2020 | | | |
| Vcmax<LLN | Yes | Yes | No |
| FEV1 < LLN | Yes | Yes | No |
| FEV1/Vcmax < LLN | Yes | Yes | No |
| DLCO < LLN | Yes | Yes | No |
| TLC < LLN | Yes | Yes | No |
| RV < LLN | Yes | Yes | No |
| Residual CT abnormality - ground glass opacity | Yes | Yes | Yes |
| Residual CT abnormality - Bronchi(ol)ectasis | Yes | Yes | Yes |
| Residual CT abnormality - lines and bands | Yes | Yes | Yes |
| Residual CT abnormality - fibrosis | Yes | Yes | Yes |
| Number of Residual CT abnormalities - 1 | Yes | Yes | Yes |
| Number of Residual CT abnormalities - 2 | Yes | Yes | Yes |
| Number of Residual CT abnormalities - 3 | Yes | Yes | Yes |
| Number of Residual CT abnormalities - 4 | Yes | Yes | Yes |
| Frailty - Somewhat | No | No | No |
| Frailty - Frail | No | No | No |
| 6MWD<80% predicted | Yes | Yes | No |
| Desaturation ≥ 4% upon 6MWT | Yes | Yes | No |
| FFMI<LN | Yes | Yes | No |
| Anxiety HADS>10 | No | Yes | No |
| Depression HADS>10 | No | Yes | No |
| TICS<34 | Yes | Yes | No |
| CFQ>43 | No | Yes | No |
| PCL-5>33 | No | Yes | No |
| IES-R>33 | No | Yes | No |

| | | | |
|---|---|---|---|
| Abnormal scored on mental or cognitive status questionnaires | No | Yes | No |
| Venturelli_2021 | | | |
| Confusion | No | Yes | Yes |
| Asthenia | No | Yes | Yes |
| Dyspnoea | No | Yes | Yes |
| Fever | No | No | Yes |
| Myalgia | No | No | Yes |
| Cough | No | No | Yes |
| Headache | No | No | Yes |
| Chest pain | No | No | Yes |
| Palpitations | No | No | Yes |
| Syncope | No | No | Yes |
| Anosmia/Dysgeusia | No | No | Yes |
| Upper gastro-intestinal symptoms | No | No | Yes |
| Lower gastro-intestinal symptoms | No | No | Yes |
| Others | No | No | Yes |
| ⩾3 persistent symptoms | No | No | Yes |
| 93-94% Oxygen saturation on room air | No | No | Yes |
| <93% Oxygen saturation on room air | No | No | Yes |
| Grade 1 – Mild dyspnoea | No | Yes | Yes |
| Grade 2 – Moderate dyspnoea | No | No | Yes |
| Grade 3 – Severe dyspnoea | No | No | Yes |
| Grade 4 – Very severe dyspnoea | No | No | Yes |
| 80-90% Karnofsky performance status scale | No | No | Yes |
| <80% Karnofsky performance status scale | No | No | Yes |
| Mild asthenia | No | Yes | Yes |
| Moderate asthenia | No | No | Yes |
| Severe asthenia | No | No | Yes |
| IES-R Pathologic | No | Yes | Yes |
| HADS-Anxiety Pathologic | No | Yes | Yes |
| HADS-Depression Pathologic | No | Yes | Yes |
| RSA Pathologic | No | Yes | Yes |
| MoCa Pathologic | No | Yes | Yes |
| Any symptom(s) | No | No | Yes |
| Not fully recovered | No | No | Yes |
| Dyspnoea | No | Yes | Yes |
| Dyspnoea - Moderate or Severe | No | Yes | Yes |
| No longer fully independent (after COVID-19; Barthel Index) | No | Yes | Yes |
| Newly moderately-severely dependent | No | Yes | Yes |
| New onset fatigue | No | Yes | Yes |

| | | | |
|---|---|---|---|
| New onset fatigue - moderate or severe | No | Yes | Yes |
| MoCa - reporting related symptoms | No | Yes | Yes |
| Pulmonary function - obstruction | Yes | Yes | Yes |
| Pulmonary function - restrictive pattern | Yes | Yes | Yes |
| Pulmonary function - mixed pattern | Yes | Yes | Yes |
| DLCO - reduced | Yes | Yes | Yes |
| Wu_2021 | | | |
| Fatigue | No | No | Yes |
| Exertional dyspnea | No | No | Yes |
| Smell and taste dysfunction | No | No | Yes |
| Pulmonary dysfunction | Yes | Yes | Yes |
| Abnormal FVC | Yes | Yes | Yes |
| Small airway dysfunction | Yes | Yes | Yes |
| Pulmonary diffusion impairment | Yes | Yes | Yes |
| Abnormal Chest CT | Yes | Yes | Yes |
| Abnormal CT findings | Yes | Yes | Yes |
| Abnormal CT findings of both lungs | Yes | Yes | Yes |
| Abnormal CT findings of the right lung only | Yes | Yes | Yes |
| Abnormal CT findings of the left lung only | Yes | Yes | Yes |
| Affected lung lobes - right upper lobe | Yes | Yes | Yes |
| Affected lung lobes - right middle lobe | Yes | Yes | Yes |
| Affected lung lobes - right lower lobe | Yes | Yes | Yes |
| Affected lung lobes - left upper lobe | Yes | Yes | Yes |
| Affected lung lobes - left lower lobe | Yes | Yes | Yes |
| Ground-glass opacities | Yes | Yes | Yes |
| Consolidation opacities | Yes | Yes | Yes |
| Linear opacities | Yes | Yes | Yes |
| Reticular opacities | Yes | Yes | Yes |
| Traction bronchiectasis | Yes | Yes | Yes |
| Pleural thickening | Yes | Yes | Yes |
| Xiong 2020 | | | |
| General symptoms | No | No | Yes |
| Physical decline/fatigue | No | No | Yes |
| Sweating | No | No | Yes |
| Myalgia | No | No | Yes |
| Arthralgia | No | No | Yes |
| Chills | No | No | Yes |
| Limb oedema | No | No | Yes |
| Dizziness | No | No | Yes |
| Respiratory symptoms | No | No | Yes |

| | | | |
|---|---|---|---|
| Postactivity polypnoea | No | No | Yes |
| Nonmotor polypnoea | No | No | Yes |
| Chest distress | No | No | Yes |
| Chest pain | No | No | Yes |
| Cough | No | No | Yes |
| Sputum | No | No | Yes |
| Throat pain | No | No | Yes |
| Cardiovascular-related symptoms | No | No | Yes |
| Resting heart rate increase | No | No | Yes |
| Discontinuous flushing (palpitations) | No | No | Yes |
| Newly diagnosed hypertension | No | No | Yes |
| Psychosocial symptoms | No | No | Yes |
| Somnipathy | No | No | Yes |
| Depression | No | No | Yes |
| Anxiety | No | No | Yes |
| Dysphoria | No | No | Yes |
| Feelings of inferiority | No | No | Yes |
| Alopecia | No | No | Yes |
| Xu_2021 | | | |
| FEV1 <80% predicted | Yes | Yes | Yes |
| FEV1/FVC <70% | Yes | Yes | Yes |
| TLC < 80% predicted | Yes | Yes | Yes |
| RV (L) <65% predicted | Yes | Yes | Yes |
| DLCO <80% predicted | Yes | Yes | Yes |
| DLCO 60-80% predicted | Yes | Yes | Yes |
| DLCO 40-60% predicted | Yes | Yes | Yes |
| DLCO/VA <80% predicted | Yes | Yes | Yes |
| Abnormal CT | Yes | Yes | Yes |

* The study-level risk of bias assessment changed from high to moderate for these outcomes.
\# The study-level risk of bias assessment changed from moderate to high for this outcome

### Table S3: GRADE assessments for certainty of evidence

Symptom(s) and Fatigue Outcomes (4-12 weeks after COVID-19 Diagnosis)

| Certainty assessment | | | | | | | | Summary of findings |
|---|---|---|---|---|---|---|---|---|
| **Outcomes (weeks after COVID-19 diagnosis)** | **No. of participants (studies)** | **Risk of bias** | **Inconsistency** | **Indirectness** | **Imprecision** | **Publication bias** | **Overall certainty of evidence** | **Prevalence (estimate [95% Confidence Interval])** |
| Persistence or presence of one or more symptoms at follow-up (4-12 weeks) | 1637 (9 studies) | serious[a] | serious[b,c] | not serious[d] | not serious[e,f] | not serious[g] | ⊕⊕◯◯ LOW | *Pooled result:* 61% [95% CI: 44%, 76%] |
| Fatigue (all) (4-12 weeks) | 2905.5 (16 studies) | serious[h] | serious[c, i] | not serious[j] | not serious[e,f] | not serious[g] | ⊕⊕◯◯ LOW | *Pooled result:* 41% [95% CI: 30%, 52%] |
| Fatigue (4-12 weeks) (non-hospitalized) | 187 (1 study) | not serious[k] | serious[l] | not serious[m] | serious[e,n] | not serious[g] | ⊕⊕◯◯ LOW | 16% [95% CI: 11%, 21%] |
| Fatigue (both hospitalized and non-hospitalized subgroup) (4-12 weeks) | 1554.5 (8 studies) | serious[h] | serious[c, o] | not serious[p] | not serious[e, f] | not serious[g] | ⊕⊕◯◯ LOW | *Pooled result:* 34% [95% CI: 21%, 50%] |
| Fatigue (hospitalized group) (4-12 weeks) | 1164 (7 studies) | serious[h] | serious[c, q] | not serious[r] | not serious[e, f] | not serious[g] | ⊕⊕◯◯ LOW | *Pooled result:* 53% [95% CI: 38%, 67%] |

| | | | | | | | | |
|---|---|---|---|---|---|---|---|---|
| | | | Certainty assessment | | | | | Summary of findings |
| **Outcomes (weeks after COVID-19 diagnosis)** | **No. of participants (studies)** | **Risk of bias** | **Inconsistency** | **Indirectness** | **Imprecision** | **Publication bias** | **Overall certainty of evidence** | **Prevalence (estimate [95% Confidence Interval])** |
| Fatigue (ICU) (4-12 weeks) | No studies | | | | | | | |

Explanations:

a. At least one study, but <50% of studies, was at high risk of bias (3 out of 9 studies).

b. $I^2$ was high ($I^2$=97%),

c. In this case, heterogeneity cannot be explained by differences in level of care received during the acute phase (i.e., non-hospitalized vs. hospitalized). Note that level of care may be considered a proxy for severity of COVID-19 (i.e., patients with more severe COVID-19 are more likely to require hospitalized care). We downgraded by 1 point.

d. We did not rate down for indirectness given that the study populations (i.e., ranging from mildly symptomatic individuals to those with severe COVID-19 pneumonia and those admitted to ICU) were reasonably representative of the target population, despite no explicit mention of including asymptomatic individuals.

e. In the judgment of subject matter experts, decision-making (e.g., regarding whether to invest in additional research and/or surveillance for this outcome) would not differ if the upper versus the lower boundary of the confidence interval, or the lowest versus highest point estimate from individual studies, represented the truth.

f. The optimal information size was met.

g. An *a priori* decision was made not to downgrade for risk of publication bias due to the large volume of COVID-related manuscripts being published and the large interest in publishing any findings related to long-COVID.

h. At least one study, but <50% of studies, was at high risk of bias. We downgraded by 1 point.

i. $I^2$ was high ($I^2$=97%).

j. We did not rate down for indirectness given that the study populations (i.e., individuals diagnosed with severe COVID-19 pneumonia and admitted to ICU; all critical COVID-19 survivors 3 months after discharge; Hospitalized with COVID-19; Mild to severe symptoms, hospitalized; any; n/a; any; mild symptoms; Individuals hospitalised & non-hospitalised; Confirmed COVID-19 infection; mild symptoms; any; non-hospitalized) corresponded to the target population of interest for our review.

k. A single study with a moderate risk of bias; we downgraded by 0.5 points for risk of bias (and combined this with the 0.5 points from imprecision).

l. Serious inconsistency because there was only one study; adding additional studies is very likely to increase heterogeneity.

m. We did not rate down for indirectness given that the study population (i.e., mildly symptomatic individuals with no evidence of pneumonia and not requiring hospitalization) was reasonably representative of the target population, despite no mention of including asymptomatic individuals.

n. The optimal information size was not met. We downgraded by 0.5 points; combined with the 0.5 point downgrade from risk of bias, we downgraded by 1 for imprecision.

o. $I^2$ was high ($I^2$=96%).

p. We did not rate down for indirectness given that the study populations corresponded to the target population of interest for our review.

q. $I^2$ was high ($I^2$=95%).

r. We did not rate down for indirectness given that the study population (i.e. hospitalized w/ severe COVID-19 pneumonia; hospitalized w/ moderate to severe COVID-19 infection; 5 studies with "hospitalized") corresponded to the target population of interest for our review.

OLFACTORY AND GUSTATORY

| | | Certainty assessment | | | | | | Summary of findings |
|---|---|---|---|---|---|---|---|---|
| **Outcomes (weeks after COVID-19 diagnosis)** | **No. of participants (studies)** | **Risk of bias** | **Inconsistency** | **Indirectness** | **Imprecision** | **Publication bias** | **Overall certainty of evidence** | **Prevalence (estimate [95% Confidence Interval])** |
| Smell or taste dysfunction (4-12 weeks) | 1112 (8 studies) | serious[a] | serious[b] | not serious[c] | not serious[d] | not serious[e] | ⊕⊕◯◯ LOW | *Pooled result:* 19% [95% CI: 12%, 28%] |
| Smell dysfunction, any (4-12 weeks) | 4081 (16 studies) | serious[f] | serious[g] | not serious[h] | not serious[d] | not serious[e] | ⊕⊕◯◯ LOW | *Pooled result:* 13% [95% CI: 7%, 23%] |
| Smell dysfunction, hyposmia (4-12 weeks) | 256 (3 studies) | serious[a] | not serious[i] | not serious[h] | serious[j] | not serious[e] | ⊕⊕◯◯ LOW | *Pooled result:* 23% [95% CI: 8%, 49%] |
| Smell dysfunction, anosmia (4-12 weeks) | 891 (4 studies) | serious[k] | serious[g] | not serious[h] | not serious[d] | not serious[e] | ⊕⊕◯◯ LOW | *Pooled result:* 7% [95% CI: 3%, 14%] |
| Taste dysfunction, any (4-12 weeks) | 2545 (11 studies) | serious[a] | serious[g] | not serious[l] | not serious[d] | not serious[e] | ⊕⊕◯◯ LOW | *Pooled result:* 7% [95% CI: 4%, 14%] |
| Taste dysfunction, ageusia (4-12 weeks) | 323 (1 study) | not serious[m] | serious[n] | serious[o] | not serious[d] | not serious[e] | ⊕⊕◯◯ LOW | 8% [95% CI: 5%, 11%] |

| | | | | | | | | |
|---|---|---|---|---|---|---|---|---|
| | | Certainty assessment | | | | | | Summary of findings |
| **Outcomes (weeks after COVID-19 diagnosis)** | **No. of participants (studies)** | **Risk of bias** | **Inconsistency** | **Indirectness** | **Imprecision** | **Publication bias** | **Overall certainty of evidence** | **Prevalence (estimate [95% Confidence Interval])** |
| Taste dysfunction, dysgeusia (4-12 weeks) | 131 (1 study) | serious[p] | serious[n] | serious[q] | serious[r] | not serious[e] | ⊕◯◯◯ VERY LOW | 11% [95% CI: 6%, 17%] |

Explanations:

a. At least one study, but <50% of studies, was at high risk of bias. We downgraded by 1 point.

b. $I^2$ was high ($I^2$=90%), and heterogeneity can be partially explained by differences in level of care received during the acute phase (i.e., non-hospitalized vs. hospitalized). Note that level of care may be considered a proxy for severity of COVID-19 (i.e., patients with more severe COVID-19 are more likely to require hospitalized care). We downgraded by 0.5 points because the heterogeneity is only partially explained but rounded up to downgrade by 1 because our overall certainty in the evidence is low (not moderate).

c. We did not rate down for indirectness given that the study population (i.e., non-hospitalized, and mixed) corresponded to the target population of interest for our review.

d. In the judgment of subject matter experts, decision-making (e.g., regarding whether to invest in additional research and/or surveillance for this outcome) would not differ if the upper versus the lower boundary of the confidence interval represented the truth. In addition, the optimal information size was met.

e. An a priori decision was made not to downgrade for risk of publication bias due to the large volume of COVID-related manuscripts being published and the large interest in publishing any findings related to long-COVID.

f. At least one study, but <50% of studies, was at high risk of bias. We downgraded by 1 point.

g. In this case, heterogeneity cannot be explained by differences in level of care received during the acute phase (i.e., non-hospitalized vs. hospitalized). Note that level of care may be considered a proxy for severity of COVID-19 (i.e., patients with more severe COVID-19 are more likely to require hospitalized care). We downgraded by 1 point.

h. We did not rate down for indirectness given that the study population (i.e., non-hospitalized, hospitalized, and mixed) corresponded to the target population of interest for our review.

i. $I^2$ was high ($I^2$=89%), but in this case, heterogeneity can be explained by differences in level of care received during the acute phase (i.e., non-hospitalized vs. hospitalized). Note that level of care may be considered a proxy for severity of COVID-19 (i.e., patients with more severe COVID-19 are more likely to require hospitalized care).

j. In the judgment of subject matter experts, decision-making (e.g., regarding whether to invest in additional research and/or surveillance for this outcome) would differ if the upper versus the lower boundary of the confidence interval, or the lowest versus highest point estimate from individual studies, represented the truth.

k. All studies contributing to this outcome were at moderate risk of bias. We downgraded by 0.5 points, but rounded up to downgrade by 1 because our overall certainty in the evidence is low (not moderate).

l. We did not rate down for indirectness given that the study population (i.e., hospitalized, and mixed) corresponded to the target population of interest for our review.

m. One study at moderate risk of bias. We downgraded by 0.5 points (and combined this with the 0.5 points from indirectness).

n. Serious inconsistency because there was only one study; adding additional studies is very likely to increase heterogeneity.

o. The study population excluded those with severe symptoms. We downgraded by 0.5 (and combined this with 0.5 points from risk of bias).

p. A single study with a high risk of bias; we downgraded by 1.5 points for risk of bias (and combined this with the 0.5 points from indirectness to downgrade by 1 point for each of risk of bias and indirectness).

q. A single study that included only a subset of our target population (i.e., individuals hospitalized with COVID-19). We downgraded by 0.5 points; combined with the 1.5 point downgrade from risk of bias, we downgraded by 1 point for each of risk of bias and indirectness.

r. In the judgment of subject matter experts, decision-making (e.g., regarding whether to invest in additional research and/or surveillance for this outcome) would not differ if the upper versus the lower boundary of the confidence interval represented the truth. However, the optimal information size was <u>not</u> met.

NEUROCOGNITIVE

| | | Certainty assessment | | | | | | Summary of findings |
|---|---|---|---|---|---|---|---|---|
| **Outcomes (weeks after COVID-19 diagnosis)** | **No. of participants (studies)** | **Risk of bias** | **Inconsistency** | **Indirectness** | **Imprecision** | **Publication bias** | **Overall certainty of evidence** | **Prevalence (estimate [95% Confidence Interval])** |
| Cognitive impairment (4-12 weeks) | 270 (4 studies) | not serious[a] | not serious[b] | serious[c] | serious[d] | not serious[e] | ⊕⊕◯◯ LOW | *Pooled result:* 29% [95% CI: 19%, 41%] |
| Concentration problems (4-12 weeks) | 1188 (4 studies) | not serious[a] | not serious[f] | not serious[g] | serious[h] | not serious[e] | ⊕⊕◯◯ LOW | *Pooled result:* 11% [95% CI: 4%, 26%] |
| Memory problem (4-12 weeks) | 1434 (4 studies) | serious[i] | serious[j] | not serious[g] | serious[h] | not serious[e] | ⊕◯◯◯ VERY LOW | *Pooled result:* 11% [95% CI: 6%, 20%] |
| Confusion (4-12 weeks) | 175 (1 study) | serious[k] | serious[l] | serious[m] | serious[n] | not serious[e] | ⊕◯◯◯ VERY LOW | 9% [95% CI: 5%, 13%] |
| Dizziness (4-12 weeks) | 387 (2 studies) | not serious[a] | serious[o] | serious[p] | not serious[q] | not serious[e] | ⊕⊕⊕◯ MODERATE | *Pooled result:* 3% [95% CI: 2%, 5%] |

Explanations:

a. All studies contributing to this outcome were at moderate risk of bias; we downgraded by 0.5 points for risk of bias.

b. . $I^2$ was moderate ($I^2$=75%), but in this case, heterogeneity can be explained by differences in level of care received during the acute phase (i.e., non-hospitalized vs. hospitalized). Note that level of care may be considered a proxy for severity of COVID-19 (i.e., patients with more severe COVID-19 are more likely to require hospitalized care).

c. The studies included only a subset of our target population (i.e., individuals hospitalized with severe COVID-19 pneumonia, hospitalized with moderate-to-severe COVID-19 infection, high/severe symptoms, or patients admitted in COVID-19 rehabilitation unit). We downgraded by 0.5 points; combined with the 0.5 point downgrade from risk of bias, we downgraded by 1 for indirectness.

d. In the judgment of subject matter experts, decision-making (e.g., regarding whether to invest in additional research and/or surveillance for this outcome) would not differ if the upper versus the lower boundary of the confidence interval represented the truth; however, the optimal information size was not met. We downgraded by 0.5 points, but rounded up to downgrade by 1 because our overall certainty in the evidence is low (not moderate).

e. An *a priori* decision was made not to downgrade for risk of publication bias due to the large volume of COVID-related manuscripts being published and the large interest in publishing any findings related to long-COVID.

f. $I^2$ was high ($I^2$=94%), but in this case, heterogeneity can be explained by differences in level of care received during the acute phase (i.e., non-hospitalized vs. hospitalized). Note that level of care may be considered a proxy for severity of COVID-19 (i.e., patients with more severe COVID-19 are more likely to require hospitalized care).

g. We did not rate down for indirectness given that the study population (i.e., mild symptoms, non-hospitalized, hospitalized, and mixed) corresponded to the target population of interest for our review.

h. In the judgment of subject matter experts, decision-making (e.g., regarding whether to invest in additional research and/or surveillance for this outcome) would differ if the upper versus the lower boundary of the confidence interval, or the lowest versus highest point estimate from individual studies, represented the truth. We downgraded by 1 point.

i. At least one study, but <50% of studies, was at high risk of bias. We downgraded by 1 point.

j. In this case, heterogeneity cannot be explained by differences in level of care received during the acute phase (i.e., non-hospitalized vs. hospitalized). Note that level of care may be considered a proxy for severity of COVID-19 (i.e., patients with more severe COVID-19 are more likely to require hospitalized care). We downgraded by 1 point.

k. A single study with a high risk of bias; we downgraded by 1.5 points for risk of bias (and combined this with the 0.5 points from indirectness to downgrade by 1 point for each of risk of bias and indirectness).

l. Serious inconsistency because there was only one study; adding additional studies is very likely to increase heterogeneity.

m. A single study that included only a subset of our target population (i.e., individuals hospitalized with COVID-19). We downgraded by 0.5 points; combined with the 1.5 point downgrade from risk of bias, we downgraded by 1 point for each of risk of bias and indirectness.

n. In the judgment of subject matter experts, decision-making (e.g., regarding whether to invest in additional research and/or surveillance for this outcome) would differ if the upper versus the lower boundary of the confidence interval represented the truth. We downgraded by 1 point, but it is not possible to further downgrade the certainty of the evidence below "very low".

o. $I^2$ was low ($I^2$=0%), we did not downgrade for inconsistency.

p. We did not rate down for indirectness given that the study population (i.e., mildly symptomatic individuals w/ no evidence of pneumonia and not requiring hospitalization; hospitalized due to COVID-19) corresponded to the target population of interest for our review.

q. In the judgment of subject matter experts, decision-making (e.g., regarding whether to invest in additional research and/or surveillance for this outcome) would not differ if the upper versus the lower boundary of the confidence interval represented the truth; additionally, the optimal information size was met.

NEUROMUSCULAR AND OTHER SYMPTOMS/SEQUELA

| | | Certainty assessment | | | | | | Summary of findings |
|---|---|---|---|---|---|---|---|---|
| **Outcomes (weeks after COVID-19 diagnosis)** | **No. of participants (studies)** | **Risk of bias** | **Inconsistency** | **Indirectness** | **Imprecision** | **Publication bias** | **Overall certainty of evidence** | **Prevalence (estimate [95% Confidence Interval])** |
| Muscle or joint pain (4-12 weeks) | 787 (3 studies) | serious[a] | serious[b] | not serious[c] | serious[d] | not serious[e] | ⊕◯◯◯ VERY LOW | *Pooled result:* 9% [95% CI: 4%, 23%] |
| Muscle pain (4-12 weeks) | 596 (4 studies) | serious[a] | serious[b] | serious[c] | serious[f] | not serious[e] | ⊕◯◯◯ VERY LOW | *Pooled result:* 18% [95% CI: 10%, 32%] |
| Joint pain (4-12 weeks) | 470 (4 studies) | serious[g] | serious[h] | serious[c] | not serious[i] | not serious[e] | ⊕⊕◯◯ LOW | *Pooled result:* 18% [95% CI: 14%, 24%] |
| Other symptoms/ sequelae (general pain/discomfort(4-12 weeks) | 849 (2 studies) | serious[j] | not serious[k] | not serious[l] | serious[d] | not serious[e] | ⊕⊕◯◯ LOW | *Pooled result:* 40% [95% CI: 24%, 58%] |

Explanations:

a. At least one study, but <50% of studies, was at high risk of bias. We downgraded by 1 point.

b. In this case, heterogeneity cannot be explained by differences in level of care received during the acute phase (i.e., non-hospitalized vs. hospitalized). Note that level of care may be considered a proxy for severity of COVID-19 (i.e., patients with more severe COVID-19 are more likely to require hospitalized care). We downgraded by 1 point.

c. We did not rate down for indirectness given that the study population (i.e., mild to severe symptoms) corresponded to the target population of interest for our review.

d. In the judgment of subject matter experts, decision-making (e.g., regarding whether to invest in additional research and/or surveillance for this outcome) would differ if the upper versus the lower boundary of the confidence interval, or the lowest versus highest point estimate from individual studies, represented the truth. We downgraded by 1 point.

e. An a priori decision was made not to downgrade for risk of publication bias due to the large volume of COVID-related manuscripts being published and the large interest in publishing any findings related to long-COVID.

f. In the judgment of subject matter experts, decision-making (e.g., regarding whether to invest in additional research and/or surveillance for this outcome) would not differ if the upper versus the lower boundary of the confidence interval represented the truth; however, the optimal information size was not met. We downgraded by 0.5 points, but rounded up to downgrade by 1 because our overall certainty in the evidence is very low (not low).

g. 50% or more of the studies contributing to this outcome were at high risk of bias. We downgraded by 1.5 points.

h. $I^2$ was low to moderate ($I^2$= 45%), we did not downgrade for inconsistency.

i. In the judgment of subject matter experts, decision-making (e.g., regarding whether to invest in additional research and/or surveillance for this outcome) would not differ if the upper versus the lower boundary of the confidence interval represented the truth; additionally, the optimal information size was met.

j. Both studies were at moderate risk of bias; we downgraded by 0.5 points, but rounded up to downgrade by 1 because our overall certainty in the evidence is low (not moderate).

k. $I^2$ was high ($I^2$=93%). In this case, heterogeneity can be explained by differences in the study populations (i.e., hospitalized with severe COVID-19 pneumonia vs. mixed severity).

l. We did not rate down for indirectness given that the study populations (i.e., no COVID-19 disease severity inclusion criteria, and individuals with severe COVID-19 pneumonia) were reasonably representative of the target population, despite no explicit mention of including asymptomatic individuals.

CARDIOVASCULAR AND RESPIRATORY

| | Certainty assessment | | | | | | | Summary of findings |
|---|---|---|---|---|---|---|---|---|
| **Outcomes (weeks after COVID-19 diagnosis)** | **No. of participants (studies)** | **Risk of bias** | **Inconsistency** | **Indirectness** | **Imprecision** | **Publication bias** | **Overall certainty of evidence** | **Prevalence (estimate [95% Confidence Interval])** |
| Palpitations (4-12 weeks) | 534 (3 studies) | serious[a] | serious[b] | not serious[c] | serious[d] | not serious[e] | ⊕◯◯◯ VERY LOW | *Pooled result:* 8% [95% CI: 3%, 19%] |
| Chest pain (4-12 weeks) | 615 (5 studies) | not serious[f] | serious[b] | not serious[g] | serious[d] | not serious[e] | ⊕◯◯◯ VERY LOW | *Pooled result:* 14% [95% CI: 7%, 25%] |
| Chest tightness (4-12 weeks) | 98 (1 study) | not serious[f] | serious[h] | serious[i] | serious[d] | not serious[e] | ⊕◯◯◯ VERY LOW | 16% [95% CI: 5%, 34%] |
| Shortness of breath (4-12 weeks) | 3159 (17 studies) | serious[a] | serious[b] | not serious[j] | not serious[k] | not serious[e] | ⊕⊕◯◯ LOW | *Pooled result:* 34% [95% CI: 25%, 45%] |
| Shortness of breath (non-hospitalized group) (4-12 weeks) | 187 (1 study) | not serious[f] | serious[h] | serious[l] | serious[m] | not serious[e] | ⊕◯◯◯ VERY LOW | 24% [95% CI: 18%, 30%] |
| Shortness of breath (hospitalized and non-hospitalized) (4-12 weeks) | 1713 (7 studies) | serious[a] | serious[b] | not serious[j] | not serious[k] | not serious[e] | ⊕⊕◯◯ LOW | *Pooled result:* 21% [95% CI: 11%, 36%] |

| | | | | | | | | |
|---|---|---|---|---|---|---|---|---|
| | | | Certainty assessment | | | | | Summary of findings |
| **Outcomes (weeks after COVID-19 diagnosis)** | **No. of participants (studies)** | **Risk of bias** | **Inconsistency** | **Indirectness** | **Imprecision** | **Publication bias** | **Overall certainty of evidence** | **Prevalence (estimate [95% Confidence Interval])** |
| Shortness of breath (hospitalized group) (4-12 weeks) | 936 (6 studies) | serious[a] | serious[b] | not serious[n] | not serious[k] | not serious[e] | ⊕⊕◯◯ LOW | *Pooled result:* 44% [95% CI: 34%, 54%] |
| Shortness of breath (hospitalized with moderate to severe COVID-19 or severe COVID-19 pneumonia) (4-12 weeks) | 323 (3 studies) | not serious[f] | serious[b] | not serious[n] | serious[m] | not serious[e] | ⊕⊕◯◯ LOW | *Pooled result:* 60% [95% CI: 35%, 80%] |
| Shortness of breath (ICU) (4-12 weeks) | No studies | | | | | | | |

Explanations:

a. At least one study, but <50% of studies, was at high risk of bias. We downgraded by 1 point.

b. In this case, heterogeneity cannot be explained by differences in level of care received during the acute phase (i.e., non-hospitalized vs. hospitalized). Note that level of care may be considered a proxy for severity of COVID-19 (i.e., patients with more severe COVID-19 are more likely to require hospitalized care). We downgraded by 1 point.

c. We did not rate down for indirectness given that the study population (i.e., non-hospitalized, and mixed) corresponded to the target population of interest for our review.

d. In the judgment of subject matter experts, decision-making (e.g., regarding whether to invest in additional research and/or surveillance for this outcome) would differ if the upper versus the lower boundary of the confidence interval represented the truth. We downgraded by 1 point.

e. An a priori decision was made not to downgrade for risk of publication bias due to the large volume of COVID-related manuscripts being published and the large interest in publishing any findings related to long-COVID.

f. All studies contributing to this outcome were at moderate risk of bias; we downgraded by 0.5 points for risk of bias.

g. We did not rate down for indirectness given that the study population corresponded to the target population of interest for our review.

h. Serious inconsistency because there was only one study; adding additional studies is very likely to increase heterogeneity.

i. A single study that included only a subset of our target population (i.e., pediatric individuals non-hospitalized and hospitalized with COVID-19). We downgraded by 0.5 points; combined with the 0.5 point downgrade from risk of bias, we downgraded by 1 point for indirectness.

j. We did not rate down for indirectness given that the study population (i.e., individuals with mild/severe symptoms, hospitalized and non-hospitalized) corresponded to the target population of interest for our review.

k. In the judgment of subject matter experts, decision-making (e.g., regarding whether to invest in additional research and/or surveillance for this outcome) would not differ if the upper versus the lower boundary of the confidence interval represented the truth; additionally the optimal information size was met.

l. A single study that included only a subset of our target population (i.e., mildly symptomatic individuals with no evidence of pneumonia and not requiring hospitalization). We downgraded by 0.5 points; combined with the 0.5 point downgrade from risk of bias, we downgraded by 1 for indirectness.

m. In the judgment of subject matter experts, decision-making (e.g., regarding whether to invest in additional research and/or surveillance for this outcome) would not differ if the upper versus the lower boundary of the confidence interval represented the truth. However, the optimal information size was not met. We downgraded by 0.5 points for imprecision.

n. We did not rate down for indirectness given that the study population corresponded to the target subgroup population of interest.

| | | Certainty assessment | | | | | | Summary of findings |
|---|---|---|---|---|---|---|---|---|
| **Outcomes (weeks after COVID-19 diagnosis)** | **No. of participants (studies)** | **Risk of bias** | **Inconsistency** | **Indirectness** | **Imprecision** | **Publication bias** | **Overall certainty of evidence** | **Prevalence (estimate [95% Confidence Interval])** |
| Anxiety (4-12 weeks) | 429 (4 studies) | not serious[a] | serious[b] | serious[c] | not serious[d] | not serious[e] | ⊕⊕◯◯ LOW | *Pooled result:* 19% [95% CI: 10%, 32%] |
| Depression (4-12 weeks) | 667 (5 studies) | not serious[a] | serious[b] | serious[c] | not serious[d] | not serious[e] | ⊕⊕◯◯ LOW | *Pooled result:* 23% [95% CI: 14%, 34%] |
| Anxiety or depression (4-12 weeks) | 834 (2 studies) | not serious[a] | not serious[f] | Serious[g] | Serious[h] | not serious[e] | ⊕⊕◯◯ LOW | *Pooled result:* 22% [95% CI: 19%, 25%] |
| Post-traumatic stress disorder (4-12 weeks) | 466 (5 studies) | not serious[a] | serious[b] | not serious[i] | not serious[d] | not serious[e] | ⊕⊕◯◯ LOW | *Pooled result:* 23% [95% CI: 14%, 35%] |
| Depression or post-traumatic stress disorder (4-12 weeks) | No studies | | | | | | | |
| Obsessive compulsive | No studies | | | | | | | |

Explanations:

a. All studies were at moderate risk of bias; we downgraded by 0.5 points for risk of bias.

b. $I^2$ was high ($I^2 \geq 85\%$), and heterogeneity cannot be explained by differences in level of care received during the acute phase (i.e., non-hospitalized vs. hospitalized). Note that level of care may be considered a proxy for severity of COVID-19 (i.e., patients with more severe COVID-19 are more likely to require hospitalized care). We downgraded by 1 point.

c. The studies included only a subset of our target population (i.e., individuals hospitalized with severe COVID-19 pneumonia, or hospitalized with moderate-to-severe COVID-19). We downgraded by 0.5 points; combined with the 0.5 point downgrade from risk of bias, we downgraded by 1 for indirectness.

d. In the judgment of subject matter experts, decision-making (e.g., regarding whether to invest in additional research and/or surveillance for this outcome) would not differ if the upper versus the lower boundary of the confidence interval, or the lowest versus highest point estimate from individual studies, represented the truth. The optimal information size was met.

e. An *a priori* decision was made not to downgrade for risk of publication bias due to the large volume of COVID-related manuscripts being published and the large interest in publishing any findings related to long-COVID.

f. $I^2$ was low ($I^2$=0%), we did not downgrade for inconsistency.

g. The studies included only a subset of our target population (i.e., individuals hospitalized with COVID-19). We downgraded by 0.5 points; combined with the 0.5 point downgrade from risk of bias, we downgraded by 1 for indirectness.

h. In the judgment of subject matter experts, decision-making (e.g., regarding whether to invest in additional research and/or surveillance for this outcome) would differ if the upper versus the lower boundary of the confidence interval represented the truth.

i. We did not rate down for indirectness given that the study populations (i.e., ranging from any severity, rehabilitation and hospitalized individuals) were reasonably representative of the target population.

#### SLEEP-RELATED AND OVERALL FUNCTIONING

| | | | | | | | | |
|---|---|---|---|---|---|---|---|---|
| | | **Certainty assessment** | | | | | | **Summary of findings** |
| **Outcomes (weeks after COVID-19 diagnosis)** | **No. of participants (studies)** | **Risk of bias** | **Inconsistency** | **Indirectness** | **Imprecision** | **Publication bias** | **Overall certainty of evidence** | **Prevalence (estimate [95% Confidence Interval])** |
| Sleep-related (sleep disturbances or difficulties) (4-12 weeks) | 1172 (3 studies) | not serious[a] | serious[b] | not serious[c] | serious[d] | not serious[e] | ⊕◯◯◯ VERY LOW | *Pooled result:* 18% [95% CI: 4%, 51%] |
| Sleep-related (insomnia) (4-12 weeks) | 765 (2 studies) | not serious[a] | serious[f] | serious[g] | not serious[h] | not serious[e] | ⊕⊕◯◯ LOW | *Pooled result:* 12% [95% CI: 5%, 25%] |
| General functioning (i.e., difficulties conducting usual activities) (4-12 weeks) | 567 (5 studies) | serious[i] | serious[b] | not serious[c] | not serious[h] | not serious[e] | ⊕⊕◯◯ LOW | *Pooled result:* 35% [95% CI: 18%, 56%] |
| Still felt ill or not back to full health (4-12 weeks) | 618.5 (5 studies) | serious[i] | serious[b] | not serious[c] | not serious[h] | not serious[e] | ⊕⊕◯◯ LOW | *Pooled result:* 41% [95% CI: 28%, 55%] |
| Employment-related (not returned to work) (4-12 weeks) | 105 (1 study) | not serious[a] | serious[j] | serious[k] | serious[d] | not serious[e] | ⊕◯◯◯ VERY LOW | 31% [95% CI: 23%, 40%] |
| Employment-related (sick-leave) (4-12 weeks) | 130 (1 study) | serious[l] | serious[j] | serious[m] | serious[d] | not serious[e] | ⊕◯◯◯ VERY LOW | 11% [95% CI: 5%, 16%] |

| | | | Certainty assessment | | | | | Summary of findings |
|---|---|---|---|---|---|---|---|---|
| **Outcomes (weeks after COVID-19 diagnosis)** | **No. of participants (studies)** | **Risk of bias** | **Inconsistency** | **Indirectness** | **Imprecision** | **Publication bias** | **Overall certainty of evidence** | **Prevalence (estimate [95% Confidence Interval])** |
| Any work impairment due to health (4-12 weeks) | No studies | | | | | | | |
| Missed work due to health (4-12 weeks) | No studies | | | | | | | |

Explanations:

a. All of the studies were at moderate risk of bias; we downgraded by 0.5 points for risk of bias

b. In this case, heterogeneity cannot be explained by differences in level of care received during the acute phase (i.e., non-hospitalized vs. hospitalized). Note that level of care may be considered a proxy for severity of COVID-19 (i.e., patients with more severe COVID-19 are more likely to require hospitalized care). We downgraded by 1 point.

c. We did not rate down for indirectness given that the study populations (i.e., ranging from mildly symptomatic individuals to those hospitalized with severe COVID-19) were reasonably representative of the target population.

d. In the judgment of subject matter experts, decision-making (e.g., regarding whether to invest in additional research and/or surveillance for this outcome) would differ if the upper versus the lower boundary of the confidence interval, or the lowest versus highest point estimate from individual studies, represented the truth. We downgraded by 1 point.

e. An *a priori* decision was made not to downgrade for risk of publication bias due to the large volume of COVID-related manuscripts being published and the large interest in publishing any findings related to long-COVID.

f. $I^2$ was high ($I^2$=73%), heterogeneity cannot be explained by differences in level of care received during the acute phase (i.e., non-hospitalized vs. hospitalized) since only one study in each subgroup.

g. The studies included only a subset of our target population (i.e., hospitalized patients (no severity criteria) and children). We downgraded by 0.5 points; combined with the 0.5 point downgrade from risk of bias, we downgraded by 1 for indirectness.

h. In the judgment of subject matter experts, decision-making (e.g., regarding whether to invest in additional research and/or surveillance for this outcome) would not differ if the upper versus the lower boundary of the confidence interval, or the lowest versus highest point estimate from individual studies, represented the truth. The optimal information size was met.

i. At least one study, but <50% of studies, was at high risk of bias. We downgraded by 1 point.

j. Serious inconsistency because there was only one study; adding additional studies is very likely to increase heterogeneity.

k. A single study that included only a subset of our target population (i.e., individuals who needed inpatient admission, supplemental oxygen, or critical care, and who had fatigue following COVID-19). We downgraded by 0.5 points; combined with the 0.5 point downgrade from risk of bias, we downgraded by 1 point for indirectness.

l. A single study with a high risk of bias; we downgraded by 1.5 points for risk of bias (and combined this with the 0.5 points from indirectness to downgrade by 1 point for each of risk of bias and indirectness).

m. A single study that included only a subset of our target population (i.e., individuals not requiring ICU admission). We downgraded by 0.5 points; combined with the 1.5 point downgrade from risk of bias, we downgraded by 1 point for each of risk of bias and indirectness.

#### Long-term Outcomes

##### Symptom(s) and Fatigue Outcomes (> 12 weeks after COVID-19 Diagnosis)

| | | Certainty assessment | | | | | | Summary of findings |
|---|---|---|---|---|---|---|---|---|
| **Outcomes (weeks after COVID-19 diagnosis)** | **No. of participants (studies)** | **Risk of bias** | **Inconsistency** | **Indirectness** | **Imprecision** | **Publication bias** | **Overall certainty of evidence** | **Prevalence (estimate [95% Confidence Interval])** |
| Persistence or presence of one or more symptoms at follow-up (>12 weeks) | 4511 (14 studies) | serious[a] | serious[b, c] | not serious[d] | not serious[e, f] | not serious[g] | ⊕⊕◯◯ LOW | *Pooled result:* 53% [95% CI: 41%, 65%] |
| Fatigue (>12 weeks) | 2444 (12 studies) | serious[a] | serious[b, c] | not serious[d] | not serious[e, f] | not serious[g] | ⊕⊕◯◯ LOW | *Pooled result:* 25% [95% CI: 19%, 34%] |
| Fatigue (>12 weeks) (non-hospitalized) | 458 (1 study) | not serious[h] | serious[i] | not serious[j] | not serious[e, f] | not serious[g] | ⊕⊕◯◯ LOW | 46% [95% CI: 42%, 51%] |
| Fatigue (both hospitalized and non-hospitalized subgroup) (>12 weeks) | 998 (6 studies) | serious[a] | serious[b, c] | not serious[k] | not serious[e, f] | not serious[g] | ⊕⊕◯◯ LOW | *Pooled result:* 17% [95% CI: 9%, 27%] |
| Fatigue (hospitalized group) (>12 weeks) | 800 (3 studies) | serious[a] | serious[l] | not serious[m] | not serious[e, f] | not serious[g] | ⊕⊕◯◯ LOW | *Pooled result:* 31% [95% CI: 25%, 39%] |
| Fatigue (ICU) (>12 weeks) | 188 (2 studies) | serious[n] | serious[b, c] | not serious[o] | serious[e, p] | not serious[g] | ⊕◯◯◯ VERY LOW | *Pooled result:* 39% [95% CI: 23%, 59%] |

Explanations:

a. At least one study, but <50% of studies, was at high risk of bias.

b. $I^2$ was high ($I^2$ >84%).

c. In this case, heterogeneity cannot be explained by differences in level of care received during the acute phase (i.e., non-hospitalized vs. hospitalized). Note that level of care may be considered a proxy for severity of COVID-19 (i.e., patients with more severe COVID-19 are more likely to require hospitalized care). We downgraded by 1 point.

d. We did not rate down for indirectness given that the study populations were reasonably representative of the target population, despite no explicit mention of including asymptomatic individuals.

e. In the judgment of subject matter experts, decision-making (e.g., regarding whether to invest in additional research and/or surveillance for this outcome) would not differ if the upper versus the lower boundary of the confidence interval, or the lowest versus highest point estimate from individual studies, represented the truth.

f. The optimal information size was met.

g. An *a priori* decision was made not to downgrade for risk of publication bias due to the large volume of COVID-related manuscripts being published and the large interest in publishing any findings related to long-COVID.

h. All studies contributing to this outcome were at moderate risk of bias. We downgraded by 0.5 points.

i. Serious inconsistency because there was only one study; adding additional studies is very likely to increase heterogeneity.

j. We did not rate down for indirectness given that the study populations (i.e., non-hospitalized individuals) corresponded to the target population of interest for this outcome.

k. We did not rate down for indirectness given that the study population was reasonably representative of the target population, despite no mention of including asymptomatic individuals.

l. Although $I^2$ was moderate ($I^2$ =71%), heterogeneity cannot be explained by differences in level of care received during the acute phase (i.e., non-hospitalized vs. hospitalized). Note that level of care may be considered a proxy for severity of COVID-19 (i.e., patients with more severe COVID-19 are more likely to require hospitalized care). We downgraded by 1 point.

m. We did not rate down for indirectness given that the study population (i.e.,mild to severe symptoms, hospitalized; any; severe and critical COVID-19) was reasonably representative of the target population, despite no mention of including asymptomatic individuals.

n. $\geq$50% of studies were at a high risk of bias; we downgraded by 1.5 points for risk of bias (and combined this with the 0.5 points from imprecision to downgrade by 1 point for each of risk of bias and imprecision).

o. We did not rate down for indirectness given that the study population (i.e.,indiciduals with severe and critical COVID-19 admitted to the ICU) was reasonably representative of the target population, despite no mention of including asymptomatic individuals.

p. The optimatl information size was not met. We downgraded by 0.5 points (and combined this with the 1.5 points from risk of bias to downgrade by 1 point for each of risk of bias and imprecision).

OLFACTORY AND GUSTATORY

| | Certainty assessment | | | | | | | Summary of findings |
|---|---|---|---|---|---|---|---|---|
| **Outcomes (weeks after COVID-19 diagnosis)** | **No. of participants (studies)** | **Risk of bias** | **Inconsistency** | **Indirectness** | **Imprecision** | **Publication bias** | **Overall certainty of evidence** | **Prevalence (estimate [95% Confidence Interval])** |
| Smell or taste dysfunction (>12 weeks) | 462 (3 studies) | serious[a] | serious[b] | not serious[c] | not serious[d] | not serious[e] | ⊕⊕◯◯ LOW | *Pooled result:* 13% [95% CI: 8%, 23%] |
| Smell dysfunction, any (>12 weeks) | 3340 (10 studies) | serious[a] | serious[b] | not serious[f] | not serious[d] | not serious[e] | ⊕⊕◯◯ LOW | *Pooled result:* 13% [95% CI: 9%, 19%] |
| Smell dysfunction, **hyposmia** (>12 weeks) | 33 (1 study) | not serious[g] | serious[h] | not serious[c] | serious[i] | not serious[e] | ⊕◯◯◯ VERY LOW | 18% [95% CI: 5%, 31%] |
| Smell dysfunction, **anosmia** (>12 weeks) | 728 (4 studies) | serious[a] | not serious[j] | not serious[k] | not serious[d] | not serious[e] | ⊕⊕◯◯ LOW | *Pooled result:* 7% [95% CI: 5%, 11%] |
| Taste dysfunction, any (>12 weeks) | 3182 (8 studies) | serious[a] | not serious[j] | not serious[f] | not serious[d] | not serious[e] | ⊕⊕◯◯ LOW | *Pooled result:* 7% [95% CI: 5%, 10%] |
| Ageusia (>12 weeks) | 561 (2 studies) | serious[l] | not serious[m] | not serious[k] | not serious[d] | not serious[e] | ⊕⊕◯◯ LOW | *Pooled result:* 4% [95% CI: 3%, 6%] |

| | | Certainty assessment | | | | | Summary of findings |
|---|---|---|---|---|---|---|---|
| **Outcomes (weeks after COVID-19 diagnosis)** | **No. of participants (studies)** | **Risk of bias** | **Inconsistency** | **Indirectness** | **Imprecision** | **Publication bias** | **Overall certainty of evidence** | **Prevalence (estimate [95% Confidence Interval])** |
| Dysgeusia (>12 weeks) | No studies | | | | | | | |

Explanations:

a. At least one study, but <50% of studies, was at high risk of bias. We downgraded by 1 point.

b. $I^2$ was high ($I^2 > 80\%$), and heterogeneity cannot be explained by differences in level of care received during the acute phase (i.e., non-hospitalized vs. hospitalized). Note that level of care may be considered a proxy for severity of COVID-19 (i.e., patients with more severe COVID-19 are more likely to require hospitalized care). We downgraded by 1 point.

c. We did not rate down for indirectness given that the study population (i.e., any severity, uncomplicated COVID-19) corresponded to the target population of interest for our review.

d. In the judgment of subject matter experts, decision-making (e.g., regarding whether to invest in additional research and/or surveillance for this outcome) would not differ if the upper versus the lower boundary of the confidence interval represented the truth. In addition, the optimal information size was met.

e. An a priori decision was made not to downgrade for risk of publication bias due to the large volume of COVID-related manuscripts being published and the large interest in publishing any findings related to long-COVID.

f. We did not rate down for indirectness given that the study population (i.e., non-hospitalized, hospitalized, and mixed) corresponded to the target population of interest for our review.

g. All studies contributing to this outcome were at moderate risk of bias. We downgraded by 0.5 points.

h. Serious inconsistency because there was only one study; adding additional studies is very likely to increase heterogeneity.

i. In the judgment of subject matter experts, decision-making (e.g., regarding whether to invest in additional research and/or surveillance for this outcome) would differ if the upper versus the lower boundary of the confidence interval, or the lowest versus highest point estimate from individual studies, represented the truth.

j. $I^2$ was moderate ($I^2$ =50-71%), and heterogeneity cannot be explained by differences in level of care received during the acute phase (i.e., non-hospitalized vs. hospitalized). Note that level of care may be considered a proxy for severity of COVID-19 (i.e., patients with more severe COVID-19 are more likely to require hospitalized care). We downgraded by 0.5 points.

k. We did not rate down for indirectness given that the study population (i.e., hospitalized and mixed) corresponded to the target population of interest for our review.

l. >50% of studies were at a high risk of bias; we downgraded by 1.5 points for risk of bias. We rounded up to 2 points because our certainty was low (not moderate).

m. $I^2$ was low ($I^2$=0%), we did not downgrade for inconsistency.

NEUROCOGNITIVE

| | Certainty assessment | | | | | | | Summary of findings |
|---|---|---|---|---|---|---|---|---|
| Outcomes (weeks after COVID-19 diagnosis) | No. of participants (studies) | Risk of bias | Inconsistency | Indirectness | Imprecision | Publication bias | Overall certainty of evidence | Prevalence (estimate [95% Confidence Interval]) |
| Cognitive impairment (>12 weeks) | 547 (4 studies) | serious[a] | serious[b] | not serious[c] | serious[d] | not serious[e] | ⊕◯◯◯ VERY LOW | *Pooled result:* 20% [95% CI: 5%, 54%] |
| Concentration problems (>12 weeks) | 658 (4 studies) | not serious[f] | not serious[g] | not serious[h] | serious[d] | not serious[e] | ⊕⊕◯◯ LOW | *Pooled result:* 5% [95% CI: 1%, 19%] |
| Memory problem (>12 weeks) | 1079 (6 studies) | serious[i] | serious[b] | not serious[j] | serious[d] | not serious[e] | ⊕◯◯◯ VERY LOW | *Pooled result:* 12% [95% CI: 5%, 23%] |
| Confusion (>12 weeks)ok | 451 (1 study) | serious[k] | serious[l] | serious[m] | not serious[n] | not serious[e] | ⊕◯◯◯ VERY LOW | 2% [95% CI: 1%, 4%] |
| Dizziness (>12 weeks) | 1758 (2 studies) | not serious[f] | not serious[o] | not serious[p] | serious[d] | not serious[e] | ⊕⊕◯◯ LOW | *Pooled result:* 3% [95% CI: 1%, 16%] |

Explanations:

a. ≥50% of studies were at a high risk of bias; we downgraded by 1.5 points for risk of bias.

b. $I^2$ was high ($I^2$ >90%), and heterogeneity cannot be explained by differences in level of care received during the acute phase (i.e., non-hospitalized vs. hospitalized). Note that level of care may be considered a proxy for severity of COVID-19 (i.e., patients with more severe COVID-19 are more likely to require hospitalized care).

c. We did not rate down for indirectness given that the study population (i.e., no severity inclusion criteria, and hospitalized) corresponded to the target population of interest for our review.

d. In the judgment of subject matter experts, decision-making (e.g., regarding whether to invest in additional research and/or surveillance for this outcome) would differ if the upper versus the lower boundary of the confidence interval, or the lowest versus highest point estimate from individual studies, represented the truth. We downgraded by 1 point.

e. An *a priori* decision was made not to downgrade for risk of publication bias due to the large volume of COVID-related manuscripts being published and the large interest in publishing any findings related to long-COVID.

f. All studies contributing to this outcome were at moderate risk of bias; we downgraded by 0.5 points for risk of bias.

g. $I^2$ was high ($I^2$=93%), but in this case, heterogeneity could be partially explained by differences in level of care received during the acute phase (i.e., non-hospitalized vs. hospitalized). Note that level of care may be considered a proxy for severity of COVID-19 (i.e., patients with more severe COVID-19 are more likely to require hospitalized care). We downgraded by 0.5 point.

h. We did not rate down for indirectness given that the study population (i.e., no severity inclusion criteria, hospitalized, mild symptoms, individuals hospitalized and non-hospitalized) corresponded to the target population of interest for our review.

i. At least one study, but <50% of studies, was at high risk of bias. We downgraded by 1 point.

j. We did not rate down for indirectness given that the study population (i.e., mild to severe symptoms; mild symptoms; COVID-19 pneumonia survivors after admission to the ED; confirmed COVID-19 infection; mild symptoms; any) corresponded to the target population of interest for our review.

k. A single study with a high risk of bias; we downgraded by 1.5 points for risk of bias (and combined this with the 0.5 points from indirectness to downgrade by 1 point for each of risk of bias and indirectness).

l. Serious inconsistency because there was only one study; adding additional studies is very likely to increase heterogeneity.

m. A single study that included only a subset of our target population (i.e., non-hospitalized patients with COVID-19). We downgraded by 0.5 points; combined with the 1.5 point downgrade from risk of bias, we downgraded by 1 point for each of risk of bias and indirectness.

n. In the judgment of subject matter experts, decision-making (e.g., regarding whether to invest in additional research and/or surveillance for this outcome) would not differ if the upper versus the lower boundary of the confidence interval represented the truth. In addition, the optimal information size was met.

o. $I^2$ was moderate ($I^2$=71%), but in this case, heterogeneity could be explained by differences in level of care received during the acute phase (i.e., non-hospitalized vs. hospitalized). Note that level of care may be considered a proxy for severity of COVID-19 (i.e., patients with more severe COVID-19 are more likely to require hospitalized care). We did not downgrade for inconsistency.

p. We did not rate down for indirectness given that the study population (i.e., mild/mixed severity inclusion criteria, and hospitalized individuals) corresponded to the target population of interest for our review.

NEUROMUSCULAR AND OTHER SYMPTOMS/SEQUELA

| | | | | | | | | |
|---|---|---|---|---|---|---|---|---|
| | | | Certainty assessment | | | | | Summary of findings |
| **Outcomes (weeks after COVID-19 diagnosis)** | **No. of participants (studies)** | **Risk of bias** | **Inconsistency** | **Indirectness** | **Imprecision** | **Publication bias** | **Overall certainty of evidence** | **Prevalence (estimate [95% Confidence Interval])** |
| Muscle or joint pain (>12 weeks) | 323 (1 study) | not serious[a] | serious[b] | serious[c] | not serious[d] | not serious[e] | ⊕⊕◯◯ LOW | 0.6% [95% CI: 0%, 1%] |
| Muscle pain (>12 weeks) | 2976 (8 studies) | serious[f] | serious[g] | not serious[h] | serious[i] | not serious[e] | ⊕◯◯◯ VERY LOW | *Pooled result:* 9% [95% CI: 4%, 22%] |
| Joint pain (>12 weeks) | 3162 (6 studies) | serious[f] | serious[g] | not serious[h] | not serious[d] | not serious[e] | ⊕⊕◯◯ LOW | *Pooled result:* 10% [95% CI: 6%, 16%] |
| Other symptoms/ sequelae (general pain/discomfort (>12 weeks) | 1722 (2 studies) | not serious[a] | not serious[j] | serious[k] | not serious[d] | not serious[e] | ⊕⊕⊕◯ MODERATE | *Pooled result:* 28% [95% CI: 23%, 34%] |

Explanations:

a. All studies contributing to this outcome were at moderate risk of bias; we downgraded by 0.5 points for risk of bias.

b. Serious inconsistency because there was only one study; adding additional studies is very likely to increase heterogeneity.

c. The studies included only a subset of our target population (i.e., mildly symptomatic individuals). We downgraded by 0.5 points; combined with the 0.5 point downgrade from risk of bias, we downgraded by 1 for indirectness.

d. In the judgment of subject matter experts, decision-making (e.g., regarding whether to invest in additional research and/or surveillance for this outcome) would not differ if the upper versus the lower boundary of the confidence interval, or the lowest versus highest point estimate from individual studies, represented the truth. Additionally, the optimal information size was met. We did not downgrade for imprecision.

e. An *a priori* decision was made not to downgrade for risk of publication bias due to the large volume of COVID-related manuscripts being published and the large interest in publishing any findings related to long-COVID.

f. At least one study, but <50% of studies, was at high risk of bias. We downgraded by 1 point.

g. $I^2$ was high ($I^2 \geq 95\%$), and heterogeneity cannot be explained by differences in level of care received during the acute phase (i.e., non-hospitalized vs. hospitalized). Note that level of care may be considered a proxy for severity of COVID-19 (i.e., patients with more severe COVID-19 are more likely to require hospitalized care).

h. We did not rate down for indirectness given that the study populations (i.e., non-hospitalized and hospitalized) were reasonably representative of the target population, despite no explicit mention of including asymptomatic individuals.

i. In the judgment of subject matter experts, decision-making (e.g., regarding whether to invest in additional research and/or surveillance for this outcome) would differ if the upper versus the lower boundary of the confidence interval represented the truth.

j. $I^2$ was moderate ($I^2$= 50%), we did not downgrade for inconsistency.

k. The studies included only a subset of our target population (i.e., hospitalized individuals). We downgraded by 0.5 points; combined with the 0.5 point downgrade from risk of bias, we downgraded by 1 for indirectness.

CARDIOVASCULAR AND RESPIRATORY

| | Certainty assessment | | | | | | | Summary of findings |
|---|---|---|---|---|---|---|---|---|
| Outcomes (weeks after COVID-19 diagnosis) | No. of participants (studies) | Risk of bias | Inconsistency | Indirectness | Imprecision | Publication bias | Overall certainty of evidence | Prevalence (estimate [95% Confidence Interval]) |
| Palpitations (>12 weeks) | 2867 (6 studies) | not serious[a] | serious[b] | not serious[c] | not serious[d] | not serious[e] | ⊕⊕⊕◯ MODERATE | *Pooled result:* 5% [95% CI: 3%, 11%] |
| Chest pain (>12 weeks) | 2242 (5 studies) | serious[f] | serious[g] | not serious[h] | not serious[d] | not serious[e] | ⊕⊕◯◯ LOW | *Pooled result:* 6% [95% CI: 2%, 13%] |
| Chest tightness (>12 weeks) | 404 (3 studies) | serious[f] | not serious[i] | not serious[h] | not serious[d] | not serious[e] | ⊕⊕⊕◯ MODERATE | *Pooled result:* 5% [95% CI: 3%, 8%] |
| Shortness of breath (>12 weeks) | 4952 (16 studies) | serious[f] | serious[g] | not serious[h] | not serious[d] | not serious[e] | ⊕⊕◯◯ LOW | *Pooled result:* 18% [95% CI: 13%, 24%] |
| Shortness of breath (non-hospitalized group) (>12 weeks) | 451 (1 study) | very serious[k] | serious[l] | not serious[j] | not serious[d] | not serious[e] | ⊕◯◯◯ VERY LOW | 16% [95% CI: 13%, 20%] |
| Shortness of breath (hospitalized and non-hospitalized) (>12 weeks) | 1547 (7 studies) | serious[f] | serious[g] | not serious[j] | not serious[d] | not serious[e] | ⊕⊕◯◯ LOW | *Pooled result:* 10% [95% CI: 6%, 16%] |

| | | | | | | | | Summary of findings |
|---|---|---|---|---|---|---|---|---|
| | | | **Certainty assessment** | | | | | **Summary of findings** |
| **Outcomes (weeks after COVID-19 diagnosis)** | **No. of participants (studies)** | **Risk of bias** | **Inconsistency** | **Indirectness** | **Imprecision** | **Publication bias** | **Overall certainty of evidence** | **Prevalence (estimate [95% Confidence Interval])** |
| Shortness of breath (hospitalized group) (>12 weeks) | 2393 (3 studies) | serious[f] | serious[g] | not serious[j] | not serious[d] | not serious[e] | ⨁⨁◯◯ LOW | *Pooled result:* 14% [95% CI: 8%, 26%] |
| Shortness of breath (hosp with moderate to severe COVID-19 or severe COVID-19 pneumonia) (>12 weeks) | 374 (3 studies) | not serious[a] | serious[g] | not serious[j] | not serious[d] | not serious[e] | ⨁⨁◯◯ LOW | *Pooled result:* 34% [95% CI: 14%, 62%] |
| Shortness of breath (ICU) (>12 weeks) | 187 (2 studies) | serious[m] | not serious[i] | not serious[j] | not serious[x] | not serious[e] | ⨁⨁◯◯ LOW | *Pooled result:* 48% [95% CI: 41%, 55%] |

Explanations:

a. All studies were at moderate risk of bias; we downgraded by 0.5 points for risk of bias.

b. I2 was high (I2=94%), but in this case, heterogeneity could be partially explained by differences in level of care received during the acute phase (i.e., non-hospitalized vs. hospitalized). Note that level of care may be considered a proxy for severity of COVID-19 (i.e., patients with more severe COVID-19 are more likely to require hospitalized care). We downgraded by 0.5 point, and combined with 0.5 from risk of bias, to downgrade by 1 for inconsistency.

c. We did not rate down for indirectness given that the study population (i.e., mild symptoms, non-hospitalized, hospitalized, and mixed) corresponded to the target population of interest for our review.

d. In the judgment of subject matter experts, decision-making (e.g., regarding whether to invest in additional research and/or surveillance for this outcome) would not differ if the upper versus the lower boundary of the confidence interval represented the truth. Additionally, the optimal information size was met.

e. An a priori decision was made not to downgrade for risk of publication bias due to the large volume of COVID-related manuscripts being published and the large interest in publishing any findings related to long-COVID.

f. At least one study, but <50% of studies, was at high risk of bias. We downgraded by 1 point.

g. $I^2$ was high ($I^2 \geq 89\%$), and heterogeneity cannot be explained by differences in level of care received during the acute phase (i.e., non-hospitalized vs. hospitalized). Note that level of care may be considered a proxy for severity of COVID-19 (i.e., patients with more severe COVID-19 are more likely to require hospitalized care).

h. We did not rate down for indirectness given that the study population corresponded to the target population of interest for our review.

i. $I^2$ was low ($I^2$=0%), we did not downgrade for inconsistency.

j. We did not rate down for indirectness given that the study population corresponded to the target subgroup population of interest for this outcome.

k. A single study with a high risk of bias; we downgraded by 1.5 points for risk of bias, but rounded up to downgrade by 2 because our overall certainty in the evidence is very low (not low).

l. Serious inconsistency because there was only one study; adding additional studies is very likely to increase heterogeneity.

m. 50% or more of the studies contributing to this outcome were at high risk of bias. We downgraded by 1.5 points.

MENTAL HEALTH

| | Certainty assessment | | | | | | | Summary of findings |
|---|---|---|---|---|---|---|---|---|
| **Outcomes (weeks after COVID-19 diagnosis)** | **No. of participants (studies)** | **Risk of bias** | **Inconsistency** | **Indirectness** | **Imprecision** | **Publication bias** | **Overall certainty of evidence** | **Prevalence (estimate [95% Confidence Interval])** |
| Depression (>12 weeks) | 326 (3 studies) | serious[a] | not serious[b] | not serious[c] | not serious[d] | not serious[e] | ⨁⨁◯◯ LOW | *Pooled result:* 17% [95% CI: 13%, 22%] |
| Anxiety (>12 weeks) | 469 (4 studies) | serious[a] | serious[f] | not serious[g] | not serious[d] | not serious[e] | ⨁◯◯◯ VERY LOW | *Pooled result:* 32% [95% CI: 22%, 43%] |
| Anxiety or depression (>12 weeks) | 1617 (1 study) | not serious[h] | serious[i] | serious[j] | not serious[d] | not serious[e] | ⨁⨁◯◯ LOW | 23% [95% CI: 21%, 25%] |
| Post-traumatic stress disorder (>12 weeks) | 929 (4 studies) | serious[a] | not serious[k] | not serious[l] | serious[m] | not serious[e] | ⨁◯◯◯ VERY LOW | *Pooled result:* 18% [95% CI: 7%, 41%] |
| Depression or post-traumatic stress disorder (>12 weeks) | 63 (1 study) | not serious[h] | serious[i] | serious[n] | serious[m] | not serious[e] | ⨁◯◯◯ VERY LOW | 22% [95% CI: 12%, 32%] |
| Obsessive compulsive (> 12 weeks) | 127 (1 study) | not serious[h] | serious[i] | not serious[g] | not serious[d] | not serious[e] | ⨁⨁◯◯ LOW | 26% [95% CI: 18%, 34%] |

Explanations:

a. At least one study, but <50% of studies, was at high risk of bias. We downgraded by 1 point.

b. $I^2$ was low ($I^2$=0%), we did not downgrade for inconsistency.

c. The studies included only a subset of our target population (i.e., individuals hospitalized with severe COVID-19 pneumonia, or hospitalized with moderate-to-severe COVID-19). We downgraded by 0.5 points.

d. In the judgment of subject matter experts, decision-making (e.g., regarding whether to invest in additional research and/or surveillance for this outcome) would not differ if the upper versus the lower boundary of the confidence interval, or the lowest versus highest point estimate from individual studies, represented the truth. The optimal information size was met.

e. An *a priori* decision was made not to downgrade for risk of publication bias due to the large volume of COVID-related manuscripts being published and the large interest in publishing any findings related to long-COVID.

f. $I^2$ was high ($I^2 \geq$85%), and heterogeneity cannot be explained by differences in level of care received during the acute phase (i.e., non-hospitalized vs. hospitalized). Note that level of care may be considered a proxy for severity of COVID-19 (i.e., patients with more severe COVID-19 are more likely to require hospitalized care). We downgraded by 1 point.

g. The studies included only a subset of our target population (i.e., hospitalized, more severe COVID-19). We downgraded by 0.5 points.

h. All studies were at moderate risk of bias; we downgraded by 0.5 points for risk of bias.

i. Serious inconsistency because there was only one study; adding additional studies is very likely to increase heterogeneity.

j. A single study that included only a subset of our target population (i.e., individuals hospitalized with COVID-19). We downgraded by 0.5 points; combined with the 0.5 point downgrade from risk of bias, we downgraded by 1 for indirectness.

k. $I^2$ was high ($I^2$=97%), but in this case, heterogeneity could be partially explained by differences in level of care received during the acute phase (i.e., non-hospitalized vs. hospitalized). Note that level of care may be considered a proxy for severity of COVID-19 (i.e., patients with more severe COVID-19 are more likely to require hospitalized care). We downgraded by 0.5 points.

l. We did not rate down for indirectness given that the study population (i.e., non-hospitalized, hospitalized, and mixed) corresponded to the target population of interest for our review.

m. In the judgment of subject matter experts, decision-making (e.g., regarding whether to invest in additional research and/or surveillance for this outcome) would differ if the upper versus the lower boundary of the confidence interval represented the truth.

n. A single study that included only a subset of our target population (i.e., 100% female; all pregnant). We downgraded by 0.5 points; combined with the 0.5 point downgrade from risk of bias, we downgraded by 1 for indirectness.

SLEEP-RELATED AND OVERALL FUNCTIONING

| | | Certainty assessment | | | | | | Summary of findings |
|---|---|---|---|---|---|---|---|---|
| **Outcomes (weeks after COVID-19 diagnosis)** | **No. of participants (studies)** | **Risk of bias** | **Inconsistency** | **Indirectness** | **Imprecision** | **Publication bias** | **Overall certainty of evidence** | **Prevalence (estimate [95% Confidence Interval])** |
| Sleep-related (sleep disturbances or difficulties) (>12 weeks) | 2112 (3 studies) | not serious[a] | not serious[b] | not serious[c] | serious[d] | not serious[e] | ⊕◯◯◯ VERY LOW | *Pooled result:* 15% [95% CI: 6%, 34%] |
| Sleep-related (insomnia) (>12 weeks) | 277 (2 studies) | not serious[a] | not serious[f] | serious[g] | not serious[h] | not serious[e] | ⊕⊕◯◯ LOW | *Pooled result:* 22% [95% CI: 17%,281%] |
| General functioning (i.e., difficulties conducting usual activities) (>12 weeks) | 2561 (6 studies) | serious[i] | serious[j] | not serious[c] | serious[d] | not serious[e] | ⊕◯◯◯ VERY LOW | *Pooled result:* 17% [95% CI: 5%, 44%] |
| Still felt ill or not back to full health (>12 weeks) | No studies | | | | | | | |
| Employment-related (not returned to work) (>12 weeks) | 76 (1 study) | serious[k] | serious[l] | serious[m] | serious[d] | not serious[e] | ⊕◯◯◯ VERY LOW | 9% [95% CI: 3%, 16%] |

| | | | | | | | | Summary of findings |
|---|---|---|---|---|---|---|---|---|
| | | | | **Certainty assessment** | | | | **Summary of findings** |
| **Outcomes (weeks after COVID-19 diagnosis)** | **No. of participants (studies)** | **Risk of bias** | **Inconsistency** | **Indirectness** | **Imprecision** | **Publication bias** | **Overall certainty of evidence** | **Prevalence (estimate [95% Confidence Interval])** |
| Employment-related (sick-leave) (>12 weeks) | No studies | | | | | | | |
| Any work impairment due to health (>12 weeks) | 72 (1 study) | not serious[a] | serious[l] | not serious[n] | serious[o] | not serious[e] | ⨁⨁◯◯ LOW | 39% [95% CI: 28%, 50%] |
| Missed work due to health (>12 weeks) | 78 (1 study) | not serious[a] | serious[l] | not serious[n] | serious[d] | not serious[e] | ⨁◯◯◯ VERY LOW | 12% [95% CI: 4%, 19%] |

Explanations:

a. All of the studies were at moderate risk of bias; we downgraded by 0.5 points for risk of bias.

b. We did not downgrade for inconsistency; the heterogeneity can be explained by differences in level of care received during the acute phase (i.e., non-hospitalized vs. hospitalized). Note that level of care may be considered a proxy for severity of COVID-19 (i.e., patients with more severe COVID-19 are more likely to require hospitalized care).

c. We did not rate down for indirectness given that the study populations (i.e., ranging from mildly symptomatic individuals to those hospitalized with COVID-19) were reasonably representative of the target population.

d. In the judgment of subject matter experts, decision-making (e.g., regarding whether to invest in additional research and/or surveillance for this outcome) would differ if the upper versus the lower boundary of the confidence interval, or the lowest versus highest point estimate from individual studies, represented the truth. We downgraded by 1 point.

e. An *a priori* decision was made not to downgrade for risk of publication bias due to the large volume of COVID-related manuscripts being published and the large interest in publishing any findings related to long-COVID.

f. $I^2$ was low ($I^2$=15%), we did not downgrade for heterogeneity.

g. The studies included only a subset of our target population (i.e., hospitalized and non-hospitalized children, and COVID-19 pneumonia survivors after admission to the ED). We downgraded by 0.5 points; combined with the 0.5 point downgrade from risk of bias, we downgraded by 1 for indirectness.

h. In the judgment of subject matter experts, decision-making (e.g., regarding whether to invest in additional research and/or surveillance for this outcome) would not differ if the upper versus the lower boundary of the confidence interval, or the lowest versus highest point estimate from individual studies, represented the truth. The optimal information size was met.

i. At least one study, but <50% of studies, was at high risk of bias. We downgraded by 1 point.

j. $I^2$ was high ($I^2 \geq 99\%$), and heterogeneity cannot be explained by differences in level of care received during the acute phase (i.e., non-hospitalized vs. hospitalized). Note that level of care may be considered a proxy for severity of COVID-19 (i.e., patients with more severe COVID-19 are more likely to require hospitalized care). We downgraded by 1 point.

k. A single study with a high risk of bias; we downgraded by 1.5 points for risk of bias (and combined this with the 0.5 points from indirectness to downgrade by 1 point for each of risk of bias and indirectness).

l. Serious inconsistency because there was only one study; adding additional studies is very likely to increase heterogeneity.

m. A single study that included only a subset of our target population (i.e., individuals hospitalized with COVID-19). We downgraded by 0.5 points; combined with the 1.5 point downgrade from risk of bias, we downgraded by 1 point for each of risk of bias and indirectness.

n. We did not rate down for indirectness given that the study populations (i.e., individuals with COVID-19 infection) were reasonably representative of the target population.

o. In the judgment of subject matter experts, decision-making (e.g., regarding whether to invest in additional research and/or surveillance for this outcome) would not differ if the upper versus the lower boundary of the confidence interval, or the lowest versus highest point estimate from individual studies, represented the truth. The optimal information size was not met. We downgraded by 0.5 points; combined with the 0.5 point downgrade from risk of bias, we downgraded by 1 point for imprecision.

Pediatric Outcomes (>12 weeks after COVID-19 Diagnosis)

| | Certainty assessment | | | | | | | Summary of findings |
|---|---|---|---|---|---|---|---|---|
| **Outcomes** | **No. of participants (studies)** | **Risk of bias** | **Inconsistency** | **Indirectness** | **Imprecision** | **Publication bias** | **Overall certainty of evidence** | **Prevalence (estimate [95% Confidence Interval])** |
| Persistence or presence of one or more symptoms at follow-up (in children) | 129 (1 study) | not serious[a] | serious[b] | not serious[c] | serious[d] | not serious[e] | ⊕⊕◯◯ LOW | 58% [95% CI: 50%, 67%] |
| Fatigue (in children) | 129 (1 study) | not serious[a] | serious[b] | not serious[c] | serious[d] | not serious[e] | ⊕⊕◯◯ LOW | 11% [95% CI: 5%, 16%] |
| Nasal congestion or runny nose (in children) | 129 (1 study) | not serious[a] | serious[b] | not serious[c] | serious[d] | not serious[e] | ⊕⊕◯◯ LOW | 12% [95% CI: 7%, 18%] |
| Headache (in children) | 129 (1 study) | not serious[a] | serious[b] | not serious[c] | serious[d] | not serious[e] | ⊕⊕◯◯ LOW | 10% [95% CI: 5%, 15%] |
| Neurocognitive (Concentration problems) (in children) | 129 (1 study) | not serious[a] | serious[b] | not serious[c] | serious[d] | not serious[e] | ⊕⊕◯◯ LOW | 10% [95% CI: 5%, 15%] |
| Neuromuscular (Muscle pain) (in children) | 129 (1 study) | not serious[a] | serious[b] | not serious[c] | serious[d] | not serious[e] | ⊕⊕◯◯ LOW | 10% [95% CI: 5%, 15%] |
| Digestive (Weight loss) (in children) | 129 (1 study) | not serious[a] | serious[b] | not serious[c] | serious[d] | not serious[e] | ⊕⊕◯◯ LOW | 8% [95% CI: 3%, 12%] |

| | Certainty assessment | | | | | | | Summary of findings |
|---|---|---|---|---|---|---|---|---|
| **Outcomes** | **No. of participants (studies)** | **Risk of bias** | **Inconsistency** | **Indirectness** | **Imprecision** | **Publication bias** | **Overall certainty of evidence** | **Prevalence (estimate [95% Confidence Interval])** |
| Sleep-related (Insomnia) (in children) | 129 (1 study) | not serious[a] | serious[b] | not serious[c] | serious[d] | not serious[e] | ⨁⨁◯◯ LOW | 19% [95% CI: 12%, 25%] |

Explanations:

a. A single study with a moderate risk of bias; we downgraded by 0.5 points for risk of bias.

b. Serious inconsistency because there was only one study; adding additional studies is very likely to increase heterogeneity.

c. Study population is reasonably representative of target population (children $\leq$ 18 years, both hospitalized and non-hospitalized. Note that although the sample is weighted towards non-hospitalized children, this is likely representative of the general pediatric COVID-19 population). We did not downgrade for indirectness.

d. In the judgment of subject matter experts, decision-making (e.g., regarding whether to invest in additional research and/or surveillance for this outcome) would not differ if the upper versus the lower boundary of the confidence interval, or the lowest versus highest point estimate from individual studies, represented the truth. However, the optimal information size was not met. We downgraded by 0.5 points (and combined this with the 0.5 points for risk of bias).

e. An a priori decision was made not to downgrade for risk of publication bias due to the large volume of COVID-related manuscripts being published and the large interest in publishing any findings related to long-COVID.

#### Table S4: Prevalence of various symptoms, sequelae and difficulties conducting usual activities post-COVID-19 infection in laboratory-confirmed and clinically diagnosed individuals

| **Symptoms, sequelae and difficulties conducting usual activities** | **Short-term (4-12 weeks after COVID-19 diagnosis)** (prevalence in %, (number of studies), risk of bias across studies risk of bias across studies [low 🟩, moderate 🟨 or high 🟥]) | **Long-term (>12 weeks after COVID-19 diagnosis)** (prevalence in %, (number of studies), risk of bias across studies risk of bias across studies [low 🟩, moderate 🟨 or high 🟥]) |
|---|---|---|
| **One or more symptoms** | | |
| Persistence or presence of one or more symptoms at follow-up | 4.5-75.1% (4 studies)(1-4) 🟨 | 2.3-73.6% (5 studies)(4-9) 🟨 |
| Number of symptoms:<br>1<br>2<br>3<br>≥3<br>4 | <br>27.2% (1 study)(2) 🟨<br>26.0% (1 study)(2) 🟨<br>16.3% (1 study)(2) 🟨<br>No studies<br>5.1% (1 study)(2) 🟨 | <br>No studies<br>No studies<br>No studies<br>1.0% (1 study)(6) 🟥<br>No studies |
| Abnormal scores on mental or cognitive questionnaires | No studies | 35.1% (1 study)(7) 🟥 |
| **Fatigue** | | |
| All | 3.3-17.9% (3 studies)(3,4,10) 🟥 | 9.5-64.2% (5 studies)(5,6,8,10,11) 🟥 |
| By level of care received during the acute phase:<br>Non-hospitalized<br>Both (hospitalized & non-hospitalized)<br>Hospitalized<br>Admitted to ICU | <br><br>No studies<br>3.3-17.9% (2 studies)(3,4) 🟥<br>15.9% (1 study)(10) 🟥<br>No studies | <br><br>No studies<br>44.1-64.2% (2 studies)(6,11) 🟥<br>9.5-39.1% (3 studies)(5,8,10) 🟥<br>No studies |
| Weakness/asthenia | 31.4% (1 study)(1) 🟨 | 71.1% (1 study)(6) 🟥 |
| Chronic fatigue:<br>Chronic fatigue syndrome/myalgic encephalomyelitis (CFS/ME)<br>CFS-like with insufficient fatigue syndrome<br>Chronic idiopathic fatigue<br>Chronic fatigue syndrome | <br>No studies<br>No studies<br>No studies<br>No studies | <br>17.5% (1 study)(12) 🟨<br>5.0% (1 study)(12) 🟨<br>10.0% (1 study)(12) 🟨<br>2.5% (1 study)(12) 🟨 |
| **Respiratory** | | |
| **Shortness of breath:** | | |
| All | 2.2-46.4% (5 studies)(1-4,10) 🟥 | 11.1-53.6% (6 studies) (5,6,9,10,13,14) 🟥 |
| By level of care received during the acute phase:<br>Non-hospitalized<br>Both (hospitalized & non-hospitalized)<br>Hospitalized | <br><br>No studies<br>2.2-46.4% (3 studies)(2-4) 🟥<br>6.7-17.5% (2 studies)(1,10) 🟥 | <br><br>No studies<br>21.8-29.7% (1 study)(6) 🟨<br>11.1-53.6% (5 studies) (5,9,10,13,14) 🟥 |

| | | |
|---|---|---|
| <br>Hospitalized (with moderate to severe COVID-19 or severe COVID-19 pneumonia)<br>Admitted to ICU | <br>No studies<br>No studies | No studies<br>No studies |
| **Other respiratory symptoms/sequelae:** | | |
| One or more respiratory symptoms | No studies | 39.0% (1 study)(8) |
| Cough | 1.0-30.5% (4 studies) (2-4,10) | 0.0-11.8% (6 studies)(5,6,8-10,15) |
| Polypnoea<br>Post-activity<br>Non-motor | <br>No studies<br>No studies | <br>21.4% (1 study)(8)<br>4.6% (1 study)(8) |
| Chest distress | No studies | 14.1% (1 study)(8) |
| Phlegm | No studies | 3.0% (1 study)(8) |
| Sore throat/throat pain | 1.2% (1 study)(4) | 3.2% (1 study)(8) |
| Dysfunctional breathing/hyperventilation | No studies | 11.9%-20.9% (1 study)(9) |
| Hoarse voice | 1.0% (1 study)(4) | No studies |
| **Flu-like symptoms** | | |
| Headaches | 2.4% (1 study)(4) | 0.5-5.5% (3 studies)(5,6,9) |
| Fever | 0.0-0.7% (2 studies)(1,4) | 0.5-0.9% (2 studies)(5,6) |
| Chills | No studies | 4.6% (1 study)(8) |
| **Olfactory and Gustatory** | | |
| Smell dysfunction<br>Any<br>Anosmia<br>Dysosmia | <br>1.8-16.1% (4 studies)(1,4,10,16)<br>5.7% (1 study)(1)<br>16.1% (1 study)(10) | <br>5.7-36.5% (4 studies)(5,9,10,17)<br>5.7-11.8% (2 studies)(5,9)<br>9.7% (1 study)(10) |
| Taste dysfunction<br>Any<br>Dysgeusia | <br>No studies<br>4.8-5.7% (2 studies)(1,10) | <br>No studies<br>1.6% (1 study)(10) |
| Smell and/or taste dysfunction | No studies | 3.0-9.3% (2 studies)(6,14) |
| **Neurocognitive** | | |
| Cognitive impairment | No studies | 0.7-22.7% (3 studies)(6,7,9) |
| Memory problem | 17.1% (1 study)(1) | 17.5% (1 study)(9) |
| Confusion | 14.3% (1 study)(3) | 3.0% (1 study)(6) |
| Concentration problems | No studies | 10.0% (1 study)(9) |
| Mental slowness | No studies | 10.1% (1 study)(9) |
| Dizziness | No studies | 2.6% (1 study)(8) |
| Delirium | 0.7% (1 study)(4) | No studies |
| Syncope | No studies | 0.1% (1 study)(6) |
| **Neuromuscular** | | |
| Muscle pain | 1.4-21.4% (2 studies)(3,4) | 3.8-22.7% (3 studies)(5,6,8) |
| Joint pain | No studies | 4.5-7.6% (2 studies)(5,8) |
| Paresthesia | No studies | 12.1% ( 1 study)(9) |
| Limb palsy | No studies | 9.6% (1 study)(9) |
| **Cardiovascular** | | |
| One or more cardiovascular-related symptoms | No studies | 13.0% (1 study)(8) |
| Palpitations | No studies | 3.9-4.8% (2 studies)(6,8) |

| | | |
|---|---|---|
| Chest pain | 3.1-23.8% (2 studies) | 3.1-12.7% (4 studies)(5,6,8,9) ■ |
| Increased heart rate | No studies | 11.2% (1 study)(8) ■ |
| Newly-diagnosed hypertension | No studies | 1.3% (1 study)(8) ■ |
| **Digestive** | | |
| Digestive disorders | 1.0% (1 study)(1) ■ | No studies |
| Weight loss | No studies | 9.1% (1 study)(9) ■ |
| Diarrhea | 0.9% (1 study)(4) ■ | 0.9% (1 study)(5) ■ |
| Abdominal pain | 1.0% (1 study) (4) ■ | 1.8% (1 study)(5) ■ |
| Gastrointestinal symptoms | No studies | 1.0% (1 study)(6) ■ |
| Nausea | No studies | 0.0% (1 study)(5) ■ |
| Skipped meals | 0.9% (1 study) | No studies |
| **Organ damage** | | |
| No studies | | |
| **Eye-related** | | |
| No studies | | |
| **Mental Health** | | |
| Any psychosocial symptom (includes somnipathy, depression, anxiety, dysphoria and feelings of inferiority) | No studies | 22.7% (1 study)(8) ■ |
| Anxiety | 7.1-29.0% (2 studies)(1,18) ■ | 6.5-31.4% (4 studies)(6-9) ■ |
| Depression | 11.0-14.5% (2 studies)(1,3) ■ | 4.3-20.6% (4 studies)(6-9) ■ |
| Depressive episode | No studies | 17.3% (1 study)(18) ■ |
| Anxiety or depression | 30.0% (1 study)(1) ■ | No studies |
| Post-traumatic stress disorder | 10.7-30.2% (3 studies)(2,3,18) ■ | 5.8-30.5% (4studies)(6,7,9,12,19) ■ |
| Post-traumatic stress syndrome | No studies | 7.2-10.5% (1 study)(7) ■ |
| Anorexia | 8.3% (1 study)(3) ■ | 7.8% (1 study)(9) ■ |
| Resilience – pathologic | No studies | 4.5% (1 study)(6) ■ |
| Dysphoria | No studies | 1.7% (1 study)(8) ■ |
| Hypomanic episode | No studies | 0.8% (1 study)(18) ■ |
| Feelings of inferiority | No studies | 0.6% (1 study)(8) ■ |
| Psychotic disorders | 0.3% (1 study)(18) ■ | No studies |
| **Quality of life (QoL)** | | |
| No studies | | |
| **Sleep-related** | | |
| Any sleep disturbances or difficulties | No studies | 17.7-53.6% (4 studies)(3,5,8,9) ■ |
| Insomnia | No studies | 23.6-53.6% (2 studies)(5,9) ■ |
| **Overall functioning** | | |
| Not fully recovered | No studies | 33.5% (1 study)(6) ■ |
| Not returned to pre-illness exercise tolerance level | 40.8% (1 study)(2) ■ | No studies |
| Frail | No studies | 16.0% (1 study)(7) ■ |
| No longer fully independent | No studies | 16.0% (1 study)(6) ■ |
| Newly moderately-severely dependent | No studies | 0.8% (1 study)(6) ■ |
| Functional impairment | | 1.8% (1 study)(6) ■ |
| **Difficulties with the following activities related to mobility:** | | |
| Sit-to-stand test results: | | |

| | | |
|---|---|---|
| Desaturation on sit-to-stand test | 13.1% (1 study)(2) | 13.6% (1 study)(5) |
| Repetitions <25th percentile | 90.0% (1 study)(20) | No studies |
| Repetitions <2.5th percentile | 42.0% (1 study)(20) | No studies |
| Pulse oxygen saturation (SpO2) test results: | | |
| Change in SpO2≥4 | 32.0% (1 study)(20) | No studies |
| SpO2 <90% post test | 16.0% (1 study)(20) | No studies |
| 6-minue walk test results: | | |
| Desaturation ≥ 4% | No studies | 22.9% (1 study)(7) |
| Distance walked <80% of predicted | No studies | 25.0% (1 study)(7) |
| **Employment-related** | | |
| Not back to work | 46.1% (1 study)(3) | No studies |
| **Complications from COVID-19+** | | |
| CT image abnormalities: | | |
| Abnormal lung CT scan | Not applicable | 22.9-63.2% (3 studies)(9,14,21) |
| Ground-glass chest opacity | Not applicable | 20.8-85.9% (4 studies) (7,9,13,14) |
| Consolidation opacities | Not applicable | 0.0% (1 study)(14) |
| Linear opacities | Not applicable | 14.6% (1 study)(14) |
| Reticular opacities | Not applicable | 6.3% (1 study)(14) |
| Parenchymal bands | Not applicable | 19.0% (1 study)(13) |
| Lines and bands | Not applicable | 63.5% (1 study)(7) |
| Bronchio(ol)ectasis/traction bronchiectasis | Not applicable | 2.1-60.0% (2 studies)(7,14) |
| Fibrosis | Not applicable | 25.9% (1 study)(7) |
| Lesions | Not applicable | 19.4% (1 study)(9) |
| Pleural thickening | Not applicable | 10.4% (1 study)(14) |
| By number of residual CT abnormalities: | | |
| 1 | Not applicable | 20.0% (1 study)(7) |
| 2 | Not applicable | 20.0% (1 study)(7) |
| 3 | Not applicable | 27.1% (1 study)(7) |
| 4 | Not applicable | 23.5% (1 study)(7) |
| Pulmonary function test abnormalities: | | |
| Pulmonary dysfunction | Not applicable | 41.5% (1 study)(14) |
| Pulmonary dysfunction due to coexistence of obstruction and restrictive pattern | Not applicable | 0.8% (1 study)(6) |
| Pulmonary diffusion impairment | Not applicable | 32.1% (1 study)(14) |
| Small airway dysfunction | Not applicable | 18.9% (1 study)(14) |
| Oxygen saturation on room air | Not applicable | 5.9% (1 study)(6) |
| DLCO<LLN or abnormal | Not applicable | 19.1-44.7% (5 |
| DLCO/VA <LLN or abnormal | Not applicable | studies)(6,7,9,13,21) |
| TLC<LLN or abnormal | Not applicable | 17.0% (1 study)(21) |
| FEV1<LLN or abnormal | Not applicable | 15.2-16.0% (2 studies)(7,21) |
| FEV1/FVC <LLN or abnormal | Not applicable | 6.4-10.7% (3 studies)(7,13,21) |
| FEV1/Vcmax<LLN | Not applicable | 3.7-17.0% (2 studies) (6,21) |
| RV<LLN or abnormal | Not applicable | 10.5% (1 study)(7) |
| Vcmax<LLN or abnormal | Not applicable | 8.7-9.6% (2 studies)(7,21) |
| FVC<LLN or abnormal | Not applicable | 8.2% (1 study)(7) |

| | | |
|---|---|---|
| | | 6.8-11.7% (3 studies)(6,13,14) ■ |
| Cardiac test results:<br>RV dilation on ECG<br>LVEF 40-50% on ECG | <br>Not applicable<br>Not applicable | <br>25.3% (1 study)(9) ■<br>12.0% (1 study)(9) ■ |
| Worse serum creatinine | Not applicable | 25.0% (1 study)(22) ■ |
| **Other symptoms/sequelae** | | |
| Burning pain | 10.5% (1 study)(1) ■ | No studies |
| Hair fall/loss | No studies | 15.5-28.6% (2 studies)(8,10) ■ |
| Sweating | No studies | 23.6% (1 study)(8) ■ |
| Needing renal replacement therapy | 3.2% (1 study) ■ | 3.2% (1 study)(22) ■ |
| Fat-free mass index < LLN | No studies | 17.8% (1 study)(7) ■ |
| Limb oedema | No studies | 2.6% (1 study)(8) ■ |

**Risk of Bias per study or across studies reporting on the outcome(s):** ■ >50% were at low risk of bias; ■ >50% were moderate risk of bias; ■ ≥50% were high risk of bias

[+] Prevalence data in the short-term for these outcomes were out of scope for this review.

(9) Writing Committee for the COMEBAC Study Group, Morin L, Savale L, Pham T, Colle R, Figueiredo S, Harrois A, Gasnier M, Lecoq AL, Meyrignac O, Noel N, Baudry E, Bellin MF, Beurnier A, Choucha W, Corruble E, Dortet L, Hardy-Leger I, Radiguer F, Sportouch S, Verny C, Wyplosz B, Zaidan M, Becquemont L, Montani D, Monnet,X. Four-Month Clinical Status of a Cohort of Patients After Hospitalization for COVID-19. JAMA 2021.

(10) Miyazato Y, Morioka S, Tsuzuki S, Akashi M, Osanai Y, Tanaka K, et al. Prolonged and Late-Onset Symptoms of Coronavirus Disease 2019. Open forum infect dis 2020;7(11):ofaa507.

(11) Shendy W, Elsherif AA, Ezzat MM, ELaidy DA. Prevalence of fatigue in patients post Covid-19. Eur J Mol Clin Med 2021;8(3):1330-1340.

(12) H,Simani L, Ramezani M, Darazam IA, Sagharichi M, Aalipour MA, Ghorbani F, Pakdaman. Prevalence and correlates of chronic fatigue syndrome and post-traumatic stress disorder after the outbreak of the COVID-19; 7852482 CSR - Reports results. J Neurovirol 2021.

(18) Group,Janiri D, Carfi A, Kotzalidis GD, Bernabei R, Landi F, Sani G, Gemelli Against COVID-19 Post-Acute Care Study. Posttraumatic Stress Disorder in Patients After Severe COVID-19 Infection. JAMA psychiatry 2021.

(19) Einvik G, Dammen T, Ghanima W, Heir T, Stavem K. Prevalence and risk factors for post-traumatic stress in hospitalized and non-hospitalized COVID-19 patients. Int J Environ Res Public Health 2021;18(4):1-12.

(20) Nunez-Cortes R, Rivera-Lillo G, Arias-Campoverde M, Soto-Garcia D, Garcia-Palomera R, Torres-Castro,R. Use of sit-to-stand test to assess the physical capacity and exertional desaturation in patients post COVID-19. Chronic respiratory disease 2021;18:1479973121999205.

(21) J,Xu J, Zhou M, Luo P, Yin Z, Wang S, Liao T, Yang F, Wang Z, Yang D, Peng Y, Geng W, Li Y, Zhang H, Yang. Plasma metabolomic profiling of patients recovered from COVID-19 with pulmonary sequelae 3 months after discharge. Clinical infectious diseases 2021.

(22) Hittesdorf E., Panzer O., Wang D., Stevens J.S., Hastie J., Jordan D.A., et al. Mortality and renal outcomes of patients with severe COVID-19 treated in a provisional intensive care unit. J Crit Care 2021;62:172-175.
